# mRNA-1273 Vaccine-elicited Neutralization of SARS-CoV-2 Omicron in Adolescents and Children

**DOI:** 10.1101/2022.01.24.22269666

**Authors:** Bethany Girard, Joanne E Tomassini, Weiping Deng, Maha Maglinao, Honghong Zhou, Amparo Figueroa, Sabine Schnyder Ghamloush, David C Montefiori, Rituparna Das, Rolando Pajon

## Abstract

**Background:** The highly transmissible severe acute respiratory syndrome coronavirus-2 (SARS-CoV-2) Omicron variant is a global concern. This study assessed the neutralization activity of two-dose regimens of mRNA-1273 vaccination against Omicron in adults, adolescents and children.

**Methods:** Neutralizing activity against the Omicron variant was evaluated in serum samples from adults (≥18 years) in the phase 3, Coronavirus Efficacy (COVE) and from adolescents (12-17 years) in the TeenCOVE trials following a two-dose regimen of 100 µg mRNA-1273 and from children (6-<12 years) in the KidCOVE trial administered two doses of 50 µg mRNA-1273. Neutralizing antibody geometric mean ID50 titers (GMT) were measured using a lentivirus-based pseudovirus neutralizing assay at day 1 and 4 weeks (day 57) following the second mRNA-1273 dose, compared with wild-type (D614G).

**Results:** At 4 weeks following a second dose of mRNA-1273 (100 µg), the GMT was reduced 28.8-fold compared with D614G in adults (≥18 years). In adolescents (12-17 years), the GMT was 11.8-fold lower than D614G, 4 weeks after a second dose of mRNA-1273 (100 µg), and compared with adults, were 1.5- and 3.8-fold higher for D614G and the Omicron variant, respectively. In children (6-<12 years), 4 weeks post-second dose of 50 µg mRNA-1273, Omicron GMTs were reduced 22.1-fold versus D614G and were 2.0-fold higher for D614G and 2.5-fold higher for Omicron compared with adults.

**Conclusions:** A two-dose regimen of 100 µg mRNA-1273 in adolescents and of 50 µg in children elicited neutralization responses against the Omicron variant that were reduced compared with the wild-type D614G, and numerically higher than those in adults.

---

The emergence of the highly transmissible SARS-CoV-2 Omicron variant has become a global concern in the coronavirus disease 2019 (COVID-19) pandemic.^1^ A two-dose primary regimen of 100 µg mRNA-1273 in adults is effective against multiple variants of SARS-CoV-2, although there is concern that effectiveness is reduced against the Omicron variant, strengthening the need for a booster dose in adults.^2^ The 100 µg primary series also elicited immunogenicity in adolescents that was comparable to young adults as well as efficacious in protecting against COVID-19 and SARS-CoV-2 infection.^3^ A two-dose primary regimen of 50 µg mRNA-1273 is currently being evaluated in children 6-<12 years of age. A preliminary evaluation has shown that the immune responses to a two-dose primary series of 100 µg of mRNA-1273 in children 6-11 years of age are comparable to higher than those seen in adults.^4^ seen in adults.^4^

In this report, neutralization of the Omicron variant was compared with the prototypic wild-type (D614G) strain using serum samples obtained from twenty participants each in ongoing clinical trials that evaluated 2 doses of 100 µg mRNA-1273 in adults ≥18 years (Coronavirus Efficacy [COVE])^5^ and adolescents 12-17 years (TeenCOVE),^3^ and 2 doses of 50 µg mRNA in children 6-<12 years of age (KidCOVE) (Table S1). Neutralizing titers were measured using a lentivirus-based pseudovirus neutralizing assay (supplementary methods) at day 1 and 4 weeks (day 57) following the second mRNA-1273 dose. Participant characteristics including age, sex and race/ethnicity of the participant samples were generally consistent with those of the larger trial populations (Table S1). Median (range) ages were 57.8 (22-75) for adults, 13.9 (12-17) for adolescents, and 8.8 (6-11) for children. Sex was generally balanced across the groups.

In adults (≥18 years), the primary 2-dose regimen of mRNA-1273 (100 µg) elicited detectable Omicron neutralizing antibodies in 95% of participants, 4 weeks post-second dose. The geometric mean ID50 titer (GMT) was reduced 28.8-fold compared to D614G (Fig 1A). Omicron neutralization was detected in 100% of adolescents, 4 weeks following a second dose of 100 µg of mRNA-1273, and Omicron GMTs were 11.8-fold lower than D614G titers (Fig 1B). Compared with adults, GMTs in adolescents were 1.5- and 3.8-fold higher for D614G and the Omicron variant, respectively. In children, at 4 weeks following a second dose of 50 µg mRNA-1273, Omicron neutralization was observed in 100% of participants and GMTs were reduced 22.1-fold versus D614G (Fig. 1C). Neutralizing titers in children were 2.0-fold higher for D614G and 2.5-fold higher for Omicron compared with those of adults.

**Figure 1.**
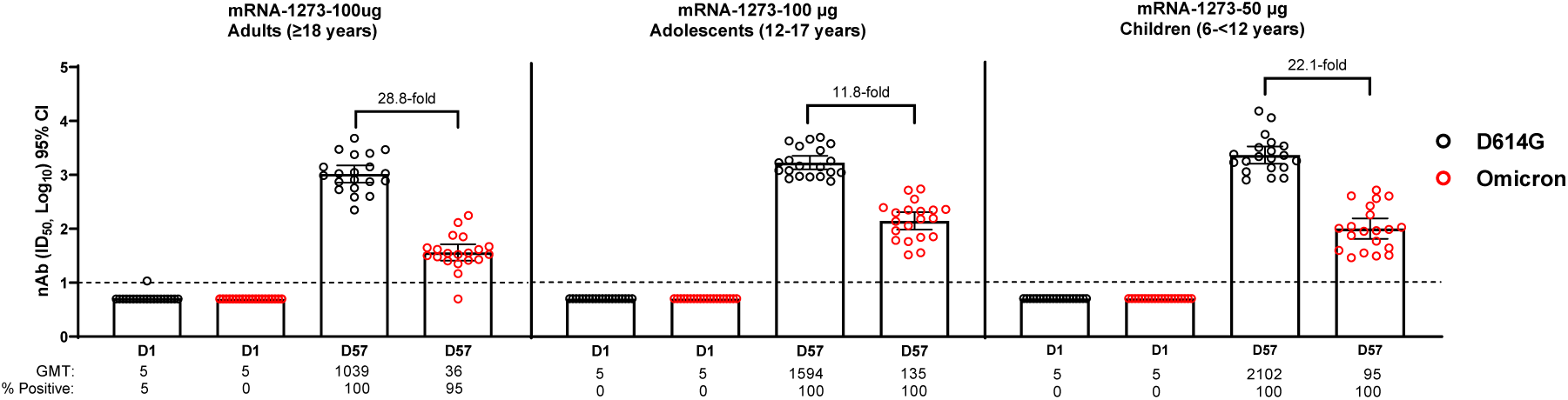
Neutralization of D614G and Omicron SARS-CoV-2 Pseudoviruses by Sera from mRNA-1273 Primary Vaccination Recipients. Pseudovirus neutralizing assay (PsVNA) titers against wild-type (D614G) and Omicron variant pseudoviruses in adults ≥18 years of age (**Panel A**), adolescents 12-17 years of age (**Panel B)** and children 6-<12 years (**Panel C**) who received a 2-dose primary regimen of mRNA-1273 in clinical trials. PsVNA titers against wild-type (D614G) and Omicron pseudovirus were measured prior to dose 1 at day 1 (D1) and 4 weeks after the 2^nd^ dose (D57) of mRNA-1273. Included in each group were 20 participants in clinical trials of adults (≥18 years) and adolescents (12-17 years) who received 100 µg mRNA-1273 and children (6-<12 years) who received 50 µg mRNA-1273. Neutralizing antibody (nAb) titers ID50 were assayed against pseudoviruses containing the spike protein of D614G and the Omicron variant (see supplementary methods). Whisker bars represent 95% confidence intervals (CI). The assay lower limit of detection (LOD) was 10, indicated by the dotted line. Values below the LOD are assigned a value of 5. D=day, GMT=geometric mean titers, NA=not available for testing.

These results indicate that D614G and Omicron neutralizing titers observed were numerically higher in adolescents and children than adults, 4 weeks following the second dose of a two-dose primary vaccination regimen of mRNA-1273. Omicron neutralization titers in adolescents and children were also reduced less compared with D614G neutralization than those in adults. The effectiveness and the durability of protection against the Omicron variant in adolescents and children remains to be determined.^2^ Study limitations include small sample sets that may not reflect neutralization in diverse populations and a lack of statistical testing, efficacy and longer-term data which can be further evaluated in these ongoing trials.

## Supporting information

Consort checklist

Protocol

## Data Availability

As the trials are ongoing, access to patient-level data and supporting clinical documents with qualified external researchers may be available upon request and subject to review once the trial is complete.

## Methods

### Trials and participants

Serum samples from three ongoing clinical trials were assessed for neutralization of Omicron variant compared with the prototypic wild-type D614G strain. Included were the previously reported phase 3 Coronavirus Efficacy (COVE; NCT04470427) trial in healthy adults (≥18 years)^5,6^ and phase2/3 TeenCOVE (NCT04649151) study in adolescents (12-17 years)^3^ both of which evaluated a two-dose vaccination regimen of 100 µg mRNA-1273 compared with placebo, and the phase 2/3 KidCOVE (NCT04796896) study which evaluated a two-dose doses of 50 µg mRNA in children (6-<12 years) (Table S1). The trials are being conducted in accordance with the International Council for Technical Requirements for Registration of Pharmaceuticals for Human Use, Good Clinical Practice Guidance, and applicable government regulations. The central Institutional Review Board/Ethics Committee, Advarra, Inc., 6100 Merriweather Drive, Columbia, MD 21044 approved the protocol and consent forms. All participants provided written informed consent. The details of the study designs for the COVE and TeenCOVE clinical trials have been previously reported^3,5,6^ and are also provided in the online study protocols, as well as those of the KidCOVE trial.

### Selection of Clinical Trial Participant Samples

Serum samples for testing in the immunogenicity subset were randomly selected from healthy adult participants (≥18 years) who completed a primary vaccination series of two doses of 100 µg mRNA-1273 in the blinded Part A of the phase 3 COVE^5,6^ and in adolescents (12-17 years) in the blinded Part A of the phase 2/3 TeenCOVE^3^ trials, and children 6-<12 years who received 2 doses of 50 µg mRNA in the open-label Part 1 of the KidCOVE trial (Table S1). Samples were selected from the trials such that 20 participants would be included in each study vaccine group and were SARS-CoV-2 negative at baseline by RT-PCR and ROCHE ELECSYS serology test. The adult group selection was balanced with 10 participants each in age groups of <65 and ≥65 years to avoid biasing the results for younger vs older age groups. Samples were generally balanced for characteristics as in the larger clinical trials (Tables S1 and S2).

### Assessment of Neutralization Titers Against D614G Wild-type and Omicron Variant

Omicron neutralization titers in serum samples obtained on the day 1 prior to dose 1 and one month after dose 2 in adults and adolescents who received 100 µg mRNA-1273, and children who received 50 µg mRNA-1273 were compared to corresponding neutralization titers of the same serum samples assayed against the prototypic wild-type (D614G) strain. Twenty participants were randomly selected from each trial for evaluation. Geometric mean titers and 95% confidence intervals are provided for each trial group. No formal statistical comparisons were made between Omicron and wild-type (D614G) or age groups.

### Pseudovirus Neutralization Assay (Duke Laboratory)

The pseudovirus neutralization assay performed at Duke has been described in detail^7^ and is a formally validated adaptation of the assay utilized by the VRC; the Duke assay is FDA approved for D614G. For measurements of neutralization, pseudovirus was incubated with 8 serial 5-fold dilutions of serum samples (1:10 starting dilution) in duplicate in a total volume of 150 µl for 1 hr at 37°C in 96-well flat-bottom culture plates. 293T/ACE2-MF cells were detached from T75 culture flasks using TrypLE Select Enzyme solution, suspended in growth medium (100,000 cells/ml) and immediately added to all wells (10,000 cells in 100 µL of growth medium per well). One set of 8 wells received cells + virus (virus control) and another set of 8 wells received cells only (background control). After 66-72 hrs of incubation, medium was removed by gentle aspiration and 30 µl of Promega 1X lysis buffer was added to all wells. After a 10 minute incubation at room temperature, 100 µl of Bright-Glo luciferase reagent was added to all wells. After 1-2 minutes, 110 µl of the cell lysate was transferred to a black/white plate. Luminescence was measured using a GloMax Navigator luminometer (Promega). Neutralization titers are the inhibitory dilution (ID) of serum samples at which RLUs were reduced by 50% (ID50) compared to virus control wells after subtraction of background RLUs. Serum samples were heat-inactivated for 30 minutes at 56°C prior to assay.

### SARS-CoV-2 Spike Variants

This study utilized two SARS-CoV-2 spike mutations in corresponding pseudoviruses. The D614G (B.1) variant contained D614G as the only spike mutation. The Omicron (B.1.1.529) variant contained spike mutations A67V, Δ69-70, T95I, G142D, Δ143-145, Δ211, L212I, +214EPE, G339D, S371L, S373P, S375F, K417N, N440K, G446S, S477N, T478K, E484A, Q493R, G496S, Q498R, N501Y, Y505H, T547K, D614G, H655Y, N679K, P681H, N764K, D796Y, N856K, Q954H, N969K, L9.

## References

1. Classification of Omicron (B.1.1.529): SARS-CoV-2 Variant of Concern. 2021. (Accessed January19, 2021, at https://www.who.int/news/item/26-11-2021-classification-of-omicron-(b.1.1.529)-sars-cov-2-variant-of-concern.)

2. Doria-Rose NA, Shen X, Schmidt SD, et al. Booster of mRNA-1273 Vaccine Reduces SARS-CoV-2 Omicron Escape from Neutralizing Antibodies. medRxiv 2021:2021.12.15.21267805.

3. Ali K, Berman G, Zhou H, et al. Evaluation of mRNA-1273 SARS-CoV-2 Vaccine in Adolescents. N Engl J Med 2021;385:2241–51. 10.1056/NEJMoa2109522

4. Bartsch YC, St Denis KJ, Kaplonek P, et al. Comprehensive antibody profiling of mRNA vaccination in children. bioRxiv 2021:2021.10.07.463592. 10.1101/2021.10.07.463592

5. El Sahly HM, Baden LR, Essink B, et al. Efficacy of the mRNA-1273 SARS-CoV-2 Vaccine at Completion of Blinded Phase. N Engl J Med 2021;385:1774–85. 10.1056/NEJMoa2113017

6. Baden LR, El Sahly HM, Essink B, et al. Efficacy and Safety of the mRNA-1273 SARS-CoV-2 Vaccine. N Engl J Med 2021;384:403–16. 10.1056/NEJMoa2035389

7. Gilbert PB, Montefiori DC, McDermott AB, et al. Immune correlates analysis of the mRNA-1273 COVID-19 vaccine efficacy clinical trial. Science 2021:eab3435. 10.1126/science.abm3425

