## Supplement for "mRNA-1273 Vaccine-elicited Neutralization of SARS-CoV-2 Omicron in Adolescents and Children"

### Table S1. Sample selection for immunogenicity and characteristics

| n (%) | 100 µg mRNA-1273 | 100 µg mRNA-1273 | 50 µg mRNA-1273 |
|---|---|---|---|
| N | 20 | 20 | 20 |
| Study | Phase 3 COVE Part A (blinded)* | Phase 2/3 TeenCOVE Part A (blinded)* | Phase 2/3 KidCOVE Part 1*† |
| Age (years) | | | |
| Mean (range) | 57.8 (22-75) | 13.9 (12-17) | 8.8 (6-11) |
| <65 | 10 (50) | NA | NA |
| ≥65 | 10 (50) | NA | NA |
| ≥6 and <12 | NA | NA | 20 (100) |
| ≥12 and <16 | NA | 17 (85) | NA |
| ≥16 and <18 | NA | 3 (15) | NA |
| Sex | | | |
| Female | 10 (50) | 12 (60) | 8 (40) |
| Male | 10 (50) | 8 (40) | 12 (60) |
| Race | | | |
| Asian | 1 (5) | 1 (5) | 1 (5) |
| Black or African American | 2 (10) | 1 (5) | 3 (15) |
| Native Hawaiian or other Pacific Islander | 1 (5) | 0 (0) | 0 (0) |
| American Indian or Alaska Native | 0 (0) | 0 (0) | 0 (0) |
| Multiple | 0 (0) | 2 (10) | 0 (0) |
| Other | 0 (0) | 1 (5) | 0 (0) |
| White | 16 (80) | 15 (75) | 16 (80) |
| Ethnicity | | | |
| Hispanic or Latino | 6 (30) | - | 3 (15) |
| Not Hispanic or Latino | 14 (70) | 20 (100) | 17 (85) |

*Primary vaccination series; †Part 1 expansion set (open-label). NA=not applicable.

### Table S2. Representativeness of the study sample

| Category | Description |
|---|---|
| Age | Older individuals generally have increased susceptibility to severe disease and weaker immune responses to vaccines than younger individuals. |
| Sex and gender | Males tend to have greater susceptibility to COVID-19 infections than women and women report greater side effects to COVID-19 vaccines than males. |
| Race or ethnicity | COVID-19 affects communities of color disproportionately in the United States. |
| Overall representativeness of this trial | The participants across the 3 trials in this study were generally balanced and demonstrated expected ratios of males to females. Participants (or their legally authorized representative) completed questionnaires regarding their assigned sex at birth and gender identity at study entry in each of the trials. Older adults generally have greater susceptibility to severe disease and weaker immune responses to vaccines, including COVID-19. While the majority of participants were white in this study, the proportions of racial and ethnic representations in the larger study populations were consistent with US demographics. |
