## Supplementary material for "mRNA-1273 Vaccine-elicited Neutralization of SARS-CoV-2 Omicron in Adolescents and Children": Protocol

### CLINICAL STUDY PROTOCOL

| | |
|---|---|
| **Protocol Title:** | A Phase 3, Randomized, Stratified, Observer-Blind, Placebo-Controlled Study to Evaluate the Efficacy, Safety, and Immunogenicity of mRNA-1273 SARS-CoV-2 Vaccine in Adults Aged 18 Years and Older |
| **Protocol Number:** | mRNA-1273-P301 |
| **Sponsor Name:** | ModernaTX, Inc. |
| **Legal Registered Address:** | 200 Technology Square<br>Cambridge, MA 02139 |
| **Sponsor Contact and Medical Monitor:** | Allison August, MD<br>VP, Clinical Development<br>ModernaTX, Inc.<br>200 Technology Square, Cambridge, MA 02139<br>Telephone: 1-617-678-8937<br> |
| **Regulatory Agency Identifier Number(s):** | IND: 19745 |
| **Amendment Number:** | 9 |
| **Date of Amendment 9:** | 10 Sep 2021 |
| **Date of Amendment 8:** | 23 Mar 2021 |
| **Date of Amendment 7:** | 10 Feb 2021 |
| **Date of Amendment 6:** | 23 Dec 2020 |
| **Date of Amendment 5:** | 11 Nov 2020 |
| **Date of Amendment 4:** | 30 Sep 2020 |
| **Date of Amendment 3:** | 20 Aug 2020 |
| **Date of Amendment 2:** | 31 Jul 2020 |
| **Date of Amendment 1:** | 26 Jun 2020 |
| **Date of Original Protocol:** | 15 Jun 2020 |

**CONFIDENTIAL**

All financial and nonfinancial support for this study will be provided by ModernaTX, Inc. The concepts and information contained in this document or generated during the study are considered proprietary and may not be disclosed in whole or in part without the expressed written consent of ModernaTX, Inc. The study will be conducted according to the *International Council for Harmonisation (ICH) Technical Requirements for Registration of Pharmaceuticals for Human Use, E6(R2) Good Clinical Practice (GCP) Guidance.*

#### Protocol Approval – Sponsor Signatory

| | |
|---|---|
| **Study Title:** | A Phase 3, Randomized, Stratified, Observer-Blind, Placebo-Controlled Study to Evaluate the Efficacy, Safety, and Immunogenicity of mRNA-1273 SARS-CoV-2 Vaccine in Adults Aged 18 Years and Older |
| **Protocol Number:** | mRNA-1273-P301 |
| **Protocol Version Date:** | 10 Sep 2021 |

Protocol accepted and approved by:

**Please see eSignature and date in the Last page of the document**

| | |
|---|---|
| Allison August, MD<br>VP, Clinical Development<br>ModernaTX, Inc.<br>200 Technology Square<br>Cambridge, MA 02139<br>Telephone: 1-617-678-8937 | Date |

### DECLARATION OF INVESTIGATOR

I have read and understood all sections of the protocol entitled "A Phase 3, Randomized, Stratified, Observer-Blind, Placebo-Controlled Study to Evaluate the Efficacy, Safety, and Immunogenicity of mRNA-1273 SARS-CoV-2 Vaccine in Adults Aged 18 Years and Older" and the most recent version of the Investigator's Brochure.

I agree to supervise all aspects of the protocol and to conduct the clinical investigation in accordance with the current protocol, the *International Council for Harmonisation (ICH) Technical Requirements for Registration of Pharmaceuticals for Human Use, E6(R2) Good Clinical Practice (GCP) Guidance*, and all applicable government regulations. I will not make changes to the protocol before consulting with ModernaTX, Inc. or implement protocol changes without IRB/IEC approval except to eliminate an immediate risk to participants.

I agree to administer study treatment only to participants under my personal supervision or the supervision of a sub-investigator. I will not supply study treatment to any person not authorized to receive it. I also agree that persons debarred from conducting or working on clinical studies by any court or regulatory agency will not be allowed to conduct or work on studies for the Sponsor or a partnership in which the Sponsor is involved. I will immediately disclose it in writing to the Sponsor if any person who is involved in the study is debarred, or if any proceeding for debarment is pending, or, to the best of my knowledge, threatened.

I will not disclose confidential information contained in this document including participant information, to anyone other than the recipient study staffs and members of the IRB/IEC. I agree to ensure that this information will not be used for any purpose other than the evaluation or conduct of the clinical investigation without the prior written consent from ModernaTX, Inc. I will not disclose information regarding this clinical investigation or publish results of the investigation without authorization from ModernaTX, Inc.

The signature below provides the necessary assurance that this study will be conducted according to all stipulations of the protocol, including statements regarding confidentiality, and according to local legal and regulatory requirements, US federal regulations, and ICH E6(R2) GCP guidelines.

______________________________ ______________

Signature of Principal Investigator Date

______________________________

Printed Name of Principal Investigator

### Protocol Amendment Summary of Changes

| DOCUMENT HISTORY | |
|---|---|
| **Document** | **Date** |
| Amendment 9 | 10 Sep 2021 |
| Amendment 8 | 23 Mar 2021 |
| Amendment 7 | 10 Feb 2021 |
| Amendment 6 | 23 Dec 2020 |
| Amendment 5 | 11 Nov 2020 |
| Amendment 4 | 30 Sep 2020 |
| Amendment 3 | 20 Aug 2020 |
| Amendment 2 | 31 Jul 2020 |
| Amendment 1 | 26 Jun 2020 |
| Original Protocol | 15 Jun 2020 |

**Amendment 9, 10 Sep 2021: Current Amendment**

This amendment is considered to be substantial based on the criteria set forth in Article 10(a) of Directive 2001/20/EC of the European Parliament and the Council of the European Union.

**Main Rationale for the Amendment:**

1. To provide a 50 µg booster dose (BD) of mRNA-1273 to participants. This change is prompted by the announcement by the President of the US that, as of 20 Sep 2021, certain individuals will be eligible to receive a BD in addition to the prior receipt of a primary vaccine regimen with mRNA-1273. The interim results of an ongoing Moderna Phase 2 study (mRNA-1273-P201), in which participants who 6 to 8 months prior received 2 doses of 50µg or 100µg of mRNA-1273 were administered a 50µg booster of mRNA-1273, demonstrated enhanced immune responses compared to pre-boost levels and met the noninferiority criteria stipulated in the FDA Guidance on EUA for Vaccines to Prevent COVID-19. Additionally, no new safety signals emerged upon administration of the booster dose in Study mRNA-1273-P201. Based on cumulative evidence, the benefit-risk profile of a booster dose of mRNA-1273 is favorable, particularly in light of increasing breakthrough disease with the emergence of the Delta variant. The details of eligibility for BDs provided by the US federal government have not been specified as of the time of writing this protocol amendment. Providing the option for a BD to all federally eligible participants currently enrolled in Part B is expected to promote retention of participants in the ongoing study and thereby defend the scientific integrity of the study for the planned 2-year duration of follow-up after the completion of the primary vaccination series.
2. To provide guidance to the investigators regarding assessing and reporting adverse events of special interest, including myocarditis and pericarditis, for this study population.

#### Summary of Major Changes from Protocol Amendment 8 to Protocol Amendment 9:

| Section # and Name | Description of Change | Brief Rationale |
|---|---|---|
| Section 1.2 (Schema); Section 4.1 (General Design); Section 4.1.3 (Part C, the Booster Dose Phase); Section 4.3 (Choice of Dose and Control Product); Section 5.1.1 (Inclusion Criteria); Section 6.2.2 (Administration of Investigational Product); Section 6.2.4 (Packaging and Labeling); Section 6.2.5 (Storage); Section 8.1.2 (Surveillance for COVID-19 Symptoms); Section 8.1.5 (Immunogenicity Assessments); Section 8.2 (Safety Assessments); Section 8.2.1 (Safety Phone Calls); Section 8.2.6 (Blood Sampling); Section 8.3.6 (Recording and Follow-up of Pregnancy); Section 8.3.7 (Recording and Follow-up of an AE and/or SAE); Section 9 (Statistical Considerations); Section 9.5.1.3 (Long-term Efficacy Analysis); Section 9.5.2.1 (Adverse Events); Section 11.1. (APPENDIX 1: Schedules of Events; Table 23) | Text updated to reflect initiation of Part C, providing for administration of an mRNA-1273 booster dose. | To promote retention of participants in the ongoing study and thereby defend the scientific integrity of the study for the planned 2-year duration of follow-up after completion of the primary vaccination series. |
| Section 2.1 (Study Rationale); Section 2.2.2 (Clinical Studies); Section 2.3.2 (Risks from Study Participation) | Updated the status of ongoing clinical research studies | To provide context for Protocol Amendment 9 |
| Section 2.3.2 (Risks from Study Participation) | Updated to include no clinical evidence for enhanced vaccine disease from the Phase 3 clinical trial. Also updated to include very rare reports of myocarditis and pericarditis occurring after vaccination with Moderna COVID-19 vaccine. | To provide updated information to the investigators in line with updates made to the IB |
| Section 3 (Objectives and Endpoints) | Updated to include evaluation of the safety and immunogenicity of a booster dose of mRNA-1273 as exploratory objectives | To identify objectives related to the initiation of Part C. |

| Section # and Name | Description of Change | Brief Rationale |
|---|---|---|
| Section 8.1.1 (Efficacy Assessments Related to COVID-19 and SARS-CoV-2 Infection) | Definition of seroconversion for participants seropositive at Baseline has been deleted. | Seroconversion in participants seropositive at Baseline was not assessed for the Part A Blinded Phase and will not be assessed for Part B or Part C. |
| Section 8.1.2 (Surveillance for COVID-19 Symptoms); Section 11.1 (Appendix 1: Schedules of Events) | Clarified that 2 NP swab samples are collected at Illness Visits | To enhance consistency of data collection at study sites |
| Section 8.3.3 (Adverse Events of Special Interest); Section 10 (References); Section 11.5 (Appendix 5: Adverse Events of Special Interest Terms) | Added CDC case definitions for myocarditis and pericarditis, as well as other terms for other potentially relevant adverse events of special interest | To provide guidance to the investigators regarding assessing and reporting adverse events of special interest, including myocarditis and pericarditis, for this study population. |
| Section 8.4.4 (Independent Cardiac Event Adjudication Committee) | Added Section 8.4.4, Independent Cardiac Event Adjudication Committee. | To describe the proposed mechanism to assess risk of myocarditis and pericarditis in the study population (to address Center for Biologics Evaluation and Research (CBER) request to describe how risk of myocarditis and pericarditis will be assessed in the study population receiving mRNA-1273). |
| Section 8.8 (Exploratory Assessments and Biomarkers) | Text updated to reflect initiation of Part C of the study. | To reflect more comprehensively the current thinking on potential exploratory assessments |
| Section 9.5.3 (Immunogenicity Analyses) | A definition of seroresponse was added. | To more clearly define the relevant antibody response. |
| Section 11.1 (Appendix 1: Schedules of Events; Table 17 and Table 18) | Adjusted the collection of eDiary data to be weekly from Day 64 through Day 759. | To ensure no gap in eDiary data collection, as they run weekly, regardless of scheduled study site visits or safety calls. |

| Section # and Name | Description of Change | Brief Rationale |
|---|---|---|
| Section 11.1 (Appendix 1: Schedules of Events; Table 18) | Reduced the frequency of safety calls from every month after Visit 5 (Month 13) to every 2 months after Visit 5. | To reduce participant burden. COVID-19 and safety surveillance will be maintained through the continued weekly surveillance by eDiary during the second year of the Surveillance Phase. |

### IRB and Regulatory Authority Approval

A copy of this amended protocol will be sent to the institutional review board (IRB) and regulatory authority.

The changes described in this amended protocol require IRB approval prior to implementation. In addition, if the changes herein affect the informed consent, sites are required to update and submit a revised informed consent for approval that incorporates the changes described in this amended protocol.

### 1. PROTOCOL SUMMARY

#### 1.1. Synopsis

| | |
|---|---|
| **Protocol Number:** | **mRNA-1273-P301** |
| **Title:** | A Phase 3, Randomized, Stratified, Observer-Blind, Placebo-Controlled Study to Evaluate the Efficacy, Safety, and Immunogenicity of mRNA-1273 SARS-CoV-2 Vaccine in Adults Aged 18 Years and Older. |
| **Study Phase:** | 3 |
| **Objectives:** | Primary:<br>• To demonstrate the efficacy of mRNA-1273 to prevent COVID-19.<br>• To evaluate the safety and reactogenicity of 2 injections of mRNA-1273 given 28 days apart.<br>Secondary:<br>• To evaluate the efficacy of mRNA-1273 to prevent severe COVID-19.<br>• To evaluate the efficacy of mRNA-1273 to prevent serologically confirmed SARS-CoV-2 infection or COVID-19 regardless of symptomatology or severity.<br>• To evaluate vaccine efficacy (VE) against a secondary definition of COVID-19.<br>• To evaluate VE to prevent death caused by COVID-19.<br>• To evaluate the efficacy of mRNA-1273 to prevent COVID-19 after the first dose of investigational product (IP).<br>• To evaluate the efficacy of mRNA-1273 to prevent COVID-19 in all study participants, regardless of evidence of prior SARS-CoV-2 infection.<br>• To evaluate the efficacy of mRNA-1273 to prevent asymptomatic SARS-CoV-2 infection. |

| | |
|---|---|
| | Exploratory for Part A:<br>• To evaluate the effect of mRNA-1273 on the viral infection kinetics as measured by viral load at SARS-CoV-2 infection diagnosis by RT-PCR and number of days from the estimated date of SARS-CoV-2 infection until undetectable SARS-CoV-2 infection by RT-PCR.<br>• To assess VE to reduce the duration of symptoms of COVID-19.<br>• To evaluate VE against all-cause mortality.<br>• To assess VE against burden of disease (BOD) due to COVID-19.<br>• To evaluate the genetic and/or phenotypic relationships of isolated SARS-CoV-2 strains to the vaccine sequence.<br>• To evaluate immune response markers after dosing with IP as correlates of risk of COVID-19 and as correlates of risk of SARS-CoV-2 infection.<br>• To conduct additional analyses related to furthering the understanding of SARS-CoV-2 infection and COVID-19, including analyses related to the immunology of this or other vaccines, detection of viral infection, and clinical conduct.<br>Exploratory for Part C:<br>• To evaluate the safety of a booster dose (BD) of mRNA-1273<br>• To evaluate the immunogenicity of a BD of mRNA-1273. |
| **Study Design and Methodology:** | This is a 3-part Phase 3 study, comprising Part A, Part B, and Part C (Figure 1). Participants in Part A, the Blinded Phase of this study were blinded to their treatment assignment. Given that the primary efficacy endpoint for mRNA-1273 against COVID-19 was met per the protocol-defined interim analysis (IA), Part B, the Open-Label Observational Phase of this study, was designed to offer participants who received placebo in Part A of this study an option to request open-label mRNA-1273 while investigational vaccine was still available. With regards to eligibility for Part B, the CDC-EUA guidance specifications superseded eligibility criteria in the protocol. Part C is designed to offer eligible participants in Part B the option to request a BD of mRNA-1273. With regards to eligibility for Part C, the CDC-EUA guidance specifications will supersede eligibility criteria in the protocol. |

**Part A, the Blinded Phase**:

The Blinded Phase of this study (Figure 1) is a randomized, stratified, observer-blind, placebo-controlled evaluation of the efficacy, safety, and immunogenicity of mRNA-1273 SARS-CoV-2 vaccine compared to placebo in adults 18 years of age and older who have no known history of SARS-CoV-2 infection but whose locations or circumstances put them at appreciable risk of acquiring COVID-19 and/or SARS-CoV-2 infection.

Participants will be randomly assigned to receive injections of either 100 µg of mRNA-1273 vaccine or a placebo control in a 1:1 randomization ratio. Assignment will be stratified by age and health risk. This is a case-driven study and thus final sample size of the study will depend on the actual attack rate of COVID-19.

All participants will be assessed for efficacy and safety endpoints and provide a nasopharyngeal (NP) swab sample and blood sample before the first and second dose of IP, in addition to a series of post-dose blood samples for immunogenicity through 24 months after the second dose of IP. Efficacy assessments will include surveillance for COVID-19 with RT-PCR confirmation of SARS-CoV-2 infection after the first and second dose of IP.

As noted above, this is a case-driven study: if the prespecified criteria for early efficacy are met at the time of either IA or overall efficacy at the primary analysis, a final study report describing the efficacy and safety of mRNA-1273 will be prepared based on the data available at that time. In the event that success criteria are met either at the time of the IA or when the total number of cases toward the primary endpoint have accrued, participants will continue to be followed in a blinded fashion until Month 25 to enable assessment of long-term safety and durability of VE.

If the study concludes early, all participants will be requested to provide a final blood sample at the time of study conclusion.

Each participant will receive 2 doses of IP by 0.5 mL intramuscular (IM) injection, the first on Day 1 and the second on Day 29. An NP swab sample will be collected prior to the first and second dose of IP, for evaluation by RT-PCR. Participants will be given an electronic diary (eDiary) to report solicited adverse reactions (ARs) for 7 days after each dose of IP (Part A, Blinded Phase only) and to prompt an unscheduled clinic visit for clinical evaluation and NP swab sample if a participant experiences any symptoms of COVID-19. All participants will receive safety calls on Day 8, Day 15, Day 22,

Day 36, and Day 43 that will serve both to monitor for unsolicited adverse events (AEs) and to monitor for symptoms of COVID-19.

Surveillance for COVID-19 will be performed through weekly contacts with the participant via a combination of telephone calls and completion of an eDiary starting at Day 1 through the end of the study, including both the Blinded and Open-Label Observational Phases.

Participants with symptoms of COVID-19 lasting at least 48 hours (except for fever and/or respiratory symptoms) will return to the clinic or will be visited at home by medically qualified site staff within 72 hours (an "Illness Visit") to collect an NP swab sample for RT-PCR testing for SARS-CoV-2 and other respiratory pathogens, or alternatively, if a clinic or home visit is not possible, will submit a saliva sample for SARS-CoV-2 RT-PCR testing. Any confirmed COVID-19 occurring in a participant will be captured as a medically attended adverse event (MAAE) along with relevant concomitant medications and details about severity, seriousness, and outcome. All confirmed serious COVID-19 cases will be reported to the Sponsor or designee within 24 hours.

Starting with the Illness Visit, study participants will be monitored by the study investigator (or appropriately delegated study staff) for a 14-day period after diagnosis or until symptoms resolve, whichever is later. Each participant diagnosed with COVID-19 will monitor their body temperature, oxygen saturation, and symptoms following the diagnosis of COVID-19. In addition to daily follow-up of symptoms, assessments will include the collection of saliva samples during the 28-day period following the diagnosis of SARS-CoV-2 infection. The investigator will determine if medical attention is required due to worsening of COVID-19. Finally, a convalescent visit will be scheduled approximately 28 days after the initial Illness Visit. At this visit, a saliva sample will be collected, and a blood sample will be drawn for immunologic assessment of SARS-CoV-2 infection.

The 28-day period following the Illness Visit is referred to as the Convalescent Period. If during the Convalescent Period the participant has a positive result for SARS-CoV-2 from the Illness Visit, the participant will continue the Convalescent Period. If the participant has a negative result for SARS-CoV-2 from the Illness Visit, the participant will exit the Convalescent Period, including discontinuation of daily telemedicine visits and collection of saliva samples, and will return to their respective study schedule.

At each dosing visit, participants will be instructed (Day 1) or reminded (Day 29) on how to document and report solicited ARs in the eDiary provided. Solicited ARs will be assessed for 7 days after each IP dose (Part A, the Blinded Phase only) and unsolicited AEs will be assessed for 28 days after each IP dose (Part A, the Blinded Phase only); serious adverse events (SAEs), MAAEs, AEs leading to withdrawal, will be assessed throughout the study.

**Part B, the Open-Label Observational Phase**:

Part B, the Open-Label Observational Phase of the study, is prompted by the authorization of a COVID-19 vaccine under EUA. Transitioning the study to Part B permits all ongoing study participants to be informed of the availability and eligibility criteria of any COVID-19 vaccine made available under an EUA and the option to offer all ongoing study participants who request unblinding, an opportunity to schedule a participant decision visit (PDV) at the study site to learn their original treatment assignment (placebo vs. mRNA-1273 vaccine).

Part B, the Open-Label Observational Phase of the study, also provides the opportunity for eligible study participants who previously received placebo to actively request to receive 2 doses of mRNA-1273 vaccine (subject to investigational vaccine availability). All study participants will receive a notification letter and will be asked to schedule a PDV. Principal Investigators should consider current local public health guidance for administration of COVID-19 vaccines under EUA when determining the scheduling priority of participants.

All participants will proceed to Part B, the Open-Label Observational Phase of the study, starting with a PDV (Figure 2).

At the PDV, all participants will:

- Be encouraged to remain in the ongoing study,
- Be given the option to be unblinded as to their original group assignment (placebo vs. mRNA-1273 vaccine),
- Be counselled about the importance of continuing other public health measures to limit the spread of disease including social distancing, wearing a mask, and hand-washing,
- Sign a revised informed consent form,
- Provide a NP swab for RT-PCR for SARS-CoV-2 and a blood sample for serology prior to unblinding.

The following participants will continue with the original study Schedule of Events (SoEs) (Figure 2):

- Participants who request to not be unblinded,
- Participants who request to be unblinded and received mRNA-1273, and
- Participants who request to be unblinded and received placebo and choose to remain on placebo.

Participants who are unblinded, received placebo, are eligible, and request to receive mRNA-1273 (Figure 2) will have the following clinic visits:

- Open-label Dose 1 (OL-D1): To occur at the PDV or at a scheduled subsequent visit.
- Open-label Dose 2 (OL-D29): To occur 28 days after Dose 1 on OL-D1.
- Open-label Clinic Visit (OL-D57): To occur 1-month after Dose 2 on OL-D29.
- In addition, participants will continue to comply with the Original Study SoEs, as applicable.

Participants who are unblinded, received ONLY 1 dose of mRNA-1273, are eligible, and request to receive mRNA-1273 (Figure 2) will have the following clinic visits:

- Their second dose on OL-D1: To occur at the PDV or at a scheduled subsequent visit
- And return on OL-D29: To occur 1-month after Dose 2 on OL-D1.

- In addition, participants will continue to comply with the Original Study SoEs, as applicable.

All participants remain on their original SoE to complete 24 months of follow-up after Dose 2; study participants who enter the Supplemental/Modified Supplemental SoE as described above do so in addition to their original SoE. Accordingly, all study participants will complete the full study follow-up to 24 months after the second dose of their original inoculation (mRNA-1273 or placebo).

At a point when the mRNA-1273 vaccine is no longer available for study use, any participant who schedules a PDV after this time point will be unblinded (with no option to stay blinded) but will not be offered to receive mRNA-1273 study vaccine. These participants will have all of the same procedures performed outlined in the SoE table, with the exception of being offered to receive mRNA-1273 vaccine.

After the Biologics License Application (BLA) database lock (on 04 May 2021), any remaining participants who had not been unblinded were unblinded by the Moderna study team and received their unblinding treatment assignment (placebo vs. mRNA vaccine) via certified letter from the study site.

**Part C, the Booster Dose Phase**

Part C, providing a BD for all eligible participants who chose to receive one, is prompted by the announcement by the President of the US that, as of 20 Sep 2021, certain individuals will be eligible to receive a BD in addition to their primary vaccination series with mRNA-1273. The details of eligibility for BDs provided by the US federal government have not been specified as of the time of writing this protocol amendment. Providing the option for a BD to all federally eligible participants currently enrolled in Part B is expected to promote retention of participants in the ongoing study and thereby defend the scientific integrity of the study for the planned 2-year duration of follow-up after the completion of the primary vaccination series regimen.

Each study participant will receive a notification letter and will be asked to schedule a BD-1 visit at their study site. Principal Investigators should consider current local public health guidance for administration of COVID-19 vaccines under EUA and marketing authorization (if any) when determining the scheduling priority of participants.

At the BD-1 visit, each participant will:

| | |
|---|---|
| | • Be encouraged to remain in the ongoing study,<br>• Sign a revised informed consent form (ICF) that includes both updated safety information relevant to the ongoing study and a BD, and the option to receive a BD<br>• Be given the option to receive a BD consisting of a 50 µg dose of mRNA-1273,<br>• Be counselled about the importance of continuing other public health measures to limit the spread of disease including social distancing, wearing a mask, and hand-washing.<br>After the BD-1 visit, participants who do not receive a BD will continue with the Part C Supplemental SoEs with the exception of the BD-1a visit and blood draw and the D8, D15, and D22 safety calls.<br>At the BD-1 visit, participants who request a BD and are eligible will have the following study site visits and complete scheduled activities (subject to investigational vaccine availability) according to the Part C Supplemental SoE:<br>• BD-1 visit: Participants will receive a single 50 µg dose of mRNA-1273<br>• BD-1a visit: Optional, and subject to blood sample kit availability, Day 4, 3 days after BD on Day 1<br>• BD-2 visit: Day 29, 28 days after the BD on Day 1<br>• BD-3 visit: Day 181, 180 days after the BD on Day 1<br>The investigator is responsible for conducting all assessments as specified in the Part C Supplemental SoE, according to the schedule. As this Supplemental SoE is intended to occur in addition to the original SoEs being followed by all participants in Part B, there is a possibility for study visits to overlap. If visits overlap according to respective visit windows, a single visit may be done with the combined study procedures completed once.<br>This study will be conducted in compliance with the protocol, Good Clinical Practice (GCP), and all applicable regulatory requirements. |
| **Randomization:** | In Part A, the Blinded Phase of the study, approximately 30,000 participants will be randomly assigned in 1:1 ratio to receive either mRNA-1273 100 µg or placebo. The randomization will be in a blinded manner using a centralized Interactive Response |

| | |
|---|---|
| | Technology, in accordance with pre-generated randomization schedules. |
| **Stratification:** | Randomization in Part A, the Blinded Phase of the study, will be stratified based on age and, if they are < 65 years of age, based on the presence or absence of risk factors for severe illness from COVID-19 based on CDC recommendation as of March 2020. There will be 3 strata for randomization: ≥ 65 years, < 65 years and categorized to be at increased risk ("at risk") for the complications of COVID-19, and < 65 years "not at risk". Risk will be defined based on the study participants' relevant past and current medical history. At least 25% of enrolled participants, up to 50%, will be either ≥ 65 years of age or < 65 years of age and "at risk" at Screening.<br><br>Participants who are < 65 years old will be categorized as at risk for severe COVID-19 illness if they have at least 1 of the following risk factors at Screening:<br>• Chronic lung disease (eg, emphysema and chronic bronchitis, idiopathic pulmonary fibrosis, and cystic fibrosis) or moderate to severe asthma<br>• Significant cardiac disease (eg, heart failure, coronary artery disease, congenital heart disease, cardiomyopathies, and pulmonary hypertension)<br>• Severe obesity (body mass index ≥ 40 kg/m$^2$)<br>• Diabetes (Type 1, Type 2 or gestational)<br>• Liver disease<br>• Human Immunodeficiency Virus (HIV) infection |
| **Study Population:** | Participants (males and females 18 years of age or older at time of consent), who are at risk of SARS-CoV-2 infection with no known history of SARS-CoV-2 infection, are a subset of the planned target population. Additionally, potential study participants at increased risk of complications from COVID-19 will be included, since it is hypothesized that these participants might derive the greatest benefit from a vaccine. Participants ≥ 65 years of age will be eligible for enrollment with or without underlying medical conditions further increasing their risk of severe COVID-19.<br>Study sites may be selected based on SARS-COV-2 infection risk of the local population. Approximately 30,000 participants will be enrolled. |

The full lists of inclusion and exclusion criteria are provided in the body of the protocol.

**Efficacy Assessments:**

**Primary Efficacy Assessment:**

To be considered as a case of COVID-19 for the evaluation of the Primary Efficacy Endpoint, the following criteria must be met:

- The participant must have experienced at least TWO of the following systemic symptoms: Fever (≥ 38ºC), chills, myalgia, headache, sore throat, new olfactory and taste disorder(s), OR
- The participant must have experienced at least ONE of the following respiratory signs/symptoms: cough, shortness of breath or difficulty breathing, OR clinical or radiographical evidence of pneumonia; AND
- The participant must have at least 1 NP swab, or saliva sample (or respiratory sample, if hospitalized) positive for SARS-CoV-2 by RT-PCR.

**Secondary Efficacy Assessments:**

To be considered a severe COVID-19, the following criteria must be met: a confirmed COVID-19 as per the Primary Efficacy Endpoint case definition, plus any of the following:

- Clinical signs indicative of severe systemic illness, Respiratory Rate ≥ 30 per minute, Heart Rate ≥ 125 beats per minute, $SpO_2$ ≤ 93% on room air at sea level or $PaO_2/FIO_2$ < 300 mm Hg, OR
- Respiratory failure or Acute Respiratory Distress Syndrome (ARDS), (defined as needing high-flow oxygen, non-invasive or mechanical ventilation, or ECMO), evidence of shock (systolic blood pressure < 90 mmHg, diastolic BP < 60 mmHg or requiring vasopressors), OR
- Significant acute renal, hepatic or neurologic dysfunction, OR
- Admission to an intensive care unit or death.

The secondary case definition of COVID-19 is defined as the following systemic symptoms: fever (temperature ≥ 38ºC) or chills, cough, shortness of breath or difficulty breathing, fatigue, muscle aches or body aches, headache, new loss of taste or smell, sore throat, nasal congestion or rhinorrhea, nausea or vomiting or diarrhea AND a positive NP swab or saliva sample (or respiratory sample, if hospitalized) for SARS-CoV-2 by RT-PCR.

| | |
|---|---|
| | Death attributed to COVID-19 is defined as any participant who dies during the study with a cause directly attributed to a complication of COVID-19.<br><br>Asymptomatic SARS-CoV-2 infection is determined by seroconversion due to infection assessed by binding antibody (bAb) levels against SARS-CoV-2 as measured by a ligand-binding assay specific to the SARS-CoV-2 nucleocapsid protein and a negative NP swab sample for SARS-CoV-2 at Day 1. |
| **Immunogenicity Assessments:** | Immunogenicity assessments will include the following:<br>• Serum bAb levels against SARS-CoV-2 as measured by ligand-binding assay specific to the SARS-CoV-2 S protein.<br>• Serum bAb levels against SARS-CoV-2 as measured by ligand-binding assay specific to the SARS-CoV-2 nucleocapsid protein.<br>• Serum neutralizing antibody (nAb) titer against SARS-CoV-2 as measured by pseudovirus and/or live virus neutralization assays. |
| **Safety Assessments:** | Safety assessments will include monitoring and recording of the following for each participant:<br>• Solicited local and systemic ARs that occur during the 7 days following each injection (ie, the day of injection and 6 subsequent days). Solicited ARs will be recorded daily using eDiaries. (Part A, Blinded Phase Only)<br>• Unsolicited AEs observed or reported during the 28 days following each injection (ie, the day of injection and 27 subsequent days). (Part A and Part C)<br>• AEs leading to discontinuation from dosing and/or study participation from Day 1 through Day 759 or withdrawal from the study.<br>• MAAEs from Day 1 through Day 759 or withdrawal from the study.<br>• SAEs from Day 1 through Day 759 or withdrawal from the study.<br>• Abnormal vital sign measurements.<br>• Physical examination findings.<br>• Pregnancy and accompanying outcomes.<br>• Concomitant medications and non-study vaccinations. |

| | |
|---|---|
| **Investigational Product, Dosage, and Route of Administration:** | The mRNA-1273 IP is an LNP dispersion of an mRNA encoding the prefusion stabilized S protein of SARS-CoV-2 formulated in LNPs composed of 4 lipids (1 proprietary and 3 commercially available): the proprietary ionizable lipid SM-102; cholesterol; 1,2-distearoyl-sn-glycero-3 phosphocholine (DSPC); and 1 monomethoxypolyethyleneglycol-2,3-dimyristylglycerol with polyethylene glycol of average molecular weight 2000 (PEG2000-DMG). The mRNA-1273 vaccine is provided as a sterile liquid for injection and is a white to off- white dispersion in appearance, at a concentration of 0.2 mg/mL in 20 mM Tris buffer containing 87 mg/mL sucrose and 10.7 mM sodium acetate at pH 7.5.<br><br>The placebo is 0.9% sodium chloride (normal saline) injection, which meets the criteria of the United States Pharmacopeia (USP).<br><br>In Part A and Part B as applicable, investigational product will be administered as an IM injection into the deltoid muscle on a 2-dose injection schedule on Day 1 and Day 29. For injections administered for Part A and Part B, each injection will have a volume of 0.5 mL and contain mRNA-1273 100 µg or saline placebo. For Part C, each injection will have a volume of 0.25 mL and contain mRNA-1273 50 µg. Preferably, vaccine should be administered into the nondominant arm. The second dose of IP should be administered in the same arm as the first dose (Part A and Part B, as applicable).<br><br>The IP will be prepared for injection as a single 0.5 mL (Part A and Part B) or 0.25 mL (Part C) dose for each participant per protocol, as detailed in the Pharmacy Manual. Unblinded personnel who will not participate in any other aspect of the study during Part A, the Blinded Phase, will perform IP accountability, dose preparation, and IP administration. Study site personnel who were blinded during the Blinded Phase, will be unblinded at the participant level at the PDV. |
| **Sample Size:** | The sample size is driven by the total number of cases to demonstrate VE (mRNA-1273 vs. placebo) to prevent COVID-19 in Part A. Under the assumption of proportional hazards over time and with 1:1 randomization of mRNA-1273 and placebo, a total of 151 COVID-19 cases will provide 90% power to detect a 60% reduction in hazard rate (60% VE), rejecting the null hypothesis H0: VE ≤ 30%, with 2 IAs at 35% and 70% of the target total number of cases using a 1-sided O'Brien-Fleming boundary for efficacy and a log-rank test statistic with a 1-sided false positive error rate of 0.025. The total number of cases pertains to the Per-protocol (PP) Set accruing at least 14 days after the second dose. There are 2 planned IAs in this study, |

which will be performed when approximately 35% and 70% of the target total number of cases have been observed. Approximately 30,000 participants will be randomized with the following assumptions:

- The target VE against COVID-19 is 60% (with 95% confidence interval (CI) lower bound ruling out 30%, rejecting the null hypothesis H0: VE ≤ 30%).
- A 6-month COVID-19 incidence rate of 0.75% in the placebo arm.
- An annual dropout rate of 2% (loss of evaluable participants).
- Two IAs at 35% and 70% of total target cases across the 2 treatment groups with O’Brien-Fleming boundaries for efficacy monitoring.
- 3-month uniform accrual.
- Approximately 15% of participants will be excluded from the PP population, and participants are at risk for COVID-19 starting 14 days after the second dose.

**Power for Selected Secondary Efficacy Endpoints:**

For the secondary objective on VE against virologically confirmed SARS-CoV-2 infection or COVID-19 regardless of symptomology or severity (COV-INF), the study will have ≥ 90% power to demonstrate the VE is above 30% (to reject null hypothesis VE ≤ 30%) at 1-sided alpha of 2.5% if the true VE to prevent COV-INF is 60%, because every COVID-19 endpoint is necessarily a COV-INF endpoint.

For the secondary objective on VE against severe COVID-19, the power of demonstrating VE based on a total of 30 and 60 events under different scenarios of true VE and VE criteria has been calculated.

| | |
|---|---|
| **Statistical Methods:** | **Statistical Hypotheses:** For the primary efficacy objective, the null hypothesis of this study is that the VE of mRNA-1273 to prevent first occurrence of COVID-19 is ≤ 30% (ie, $H_0^{efficacy}$: VE ≤ 0.3).<br><br>The study will be considered to meet the primary efficacy objective if the corresponding CI of VE rules out 30% at either one of the IAs or at the primary analysis. In the primary analysis of VE of COVID-19, cases will be counted starting 14 days after the second dose of IP.<br><br>Vaccine efficacy is defined as the percent reduction in the hazard of the primary endpoint (mRNA-1273 vs placebo). Equivalently, the null hypothesis is:<br><br>• $H_0^{efficacy}$: hazard ratio (HR) ≥ 0.7 (equivalently, proportional hazards VE ≤ 0.3).<br><br>A stratified Cox proportional hazard model will be used to assess the magnitude of the treatment group difference (ie, HR) between mRNA-1273 and placebo at a 1-sided 0.025 significance level.<br><br>**Analysis Populations:** Analysis populations for statistical analyses are Randomization Set, Full Analysis Set (FAS), Modified Intent-to-Treat (mITT) Set, PP Set, Immunogenicity Subset, Solicited Safety Set, and Safety Set, as shown below:<br><br>• Randomization Set: All participants who are randomized, regardless of the participants' treatment status in the study.<br>• FAS: All randomized participants who received at least one dose of IP. Participants will be analyzed according to the group to which they were randomized. |

- mITT Set: All participants in the FAS who had no immunologic or virologic evidence of prior COVID-19 (ie, negative NP swab test at Day 1 and/or bAb against SARS-CoV-2 nucleocapsid below limit of detection or lower limit of quantification [LLOQ]) at Day 1 before the first dose of IP. Participants will be analyzed according to the group to which they were randomized.
- PP Set: All participants in the mITT Set who received planned doses of IP per schedule and have no major protocol deviations, as determined and documented by Sponsor prior to database lock and unblinding, that impact critical or key study data. Participants will be analyzed according to the group to which they were randomized.
- Immunogenicity Subset: All participants in the FAS who were sampled into subset for characterizing mRNA-1273 immunogenicity and had a valid immunogenicity test result prior to the first dose of IP and at least 1 valid result after the first dose of IP. Participants in the subset who had major protocol deviations that impact critical or key immunogenicity or study data may be excluded from the Immunogenicity Subset. The details of the Immunogenicity Subset will be documented prior to analysis of immunogenicity data.
- Solicited Safety Set: The Solicited Safety Set consists of all randomized participants who received at least one dose of IP and contributed any solicited AR data. The Solicited Safety Set will be used for the analyses of solicited ARs and participants will be included in the treatment group corresponding to the IP that they actually received.
- Safety Set: All randomized participants who received at least one dose of IP. The Safety Set will be used for all analyses of safety except for the solicited ARs. Participants will be included in the treatment group corresponding to the IP that they actually received.

**Efficacy Analyses:** Efficacy analyses will be performed using the FAS, mITT and PP populations, and participants will be included in the treatment group to which they are randomized. The primary analysis population for efficacy will be the PP Set.

The table below summarizes the analysis approach for primary and secondary efficacy endpoints for Part A, the randomized, observer-blind, and placebo-controlled phase of the study. Sensitivity analysis methods are described for each endpoint as applicable.

| Endpoint | Statistical Analysis Methods |
|---|---|
| **Primary endpoint:**<br>VE of mRNA-1273 to prevent COVID-19 | • Primary analysis: VE will be estimated with 1 - HR (mRNA-1273 vs placebo) using a Cox proportional hazard regression model with treatment group as a fixed effect and adjust for stratification factor based on the PP Set, with cases counted starting 14 days after the second dose of IP.<br>• Analysis using the same model based on the mITT Set.<br>• Sensitivity analysis using the same model based on the PP Set, with cases counted starting either immediately after the second dose of IP or immediately after the first dose of IP.<br>• Subgroup analysis of the primary efficacy endpoint will be performed to assess consistency of VE, such as in the age groups ≥ 18 and < 65 years and ≥ 65 years.<br>• Supportive analysis of VE to be estimated with 1 - ratio of incidence rates with 95% CI using the exact method conditional upon the total number of cases.<br>• Supportive analysis of cumulative incidence VE. |
| **Secondary endpoints:**<br>• Vaccine efficacy of mRNA-1273 to prevent severe COVID-19<br>• Vaccine efficacy of mRNA-1273 to prevent serologically confirmed SARS-CoV-2 infection or COVID-19 regardless of symptomatology or severity.<br>• Vaccine efficacy of mRNA-1273 to prevent COVID-19 using a secondary definition of symptoms | Similar analysis method as for the primary endpoint analysis.<br>For each of the secondary endpoints:<br>• Primary analysis: VE will be estimated with 1 - HR (mRNA-1273 vs placebo) using a Cox proportional hazard regression model with treatment group as a fixed effect and adjusting for stratification factor based on the PP Set, with cases counted starting 14 days after the second dose of IP.<br>• Analysis using the same model based on the mITT Set. |

| | |
|---|---|
| • Vaccine efficacy of mRNA-1273 to prevent death caused by COVID-19<br>• Vaccine efficacy of mRNA-1273 to prevent COVID-19 after the first dose of IP<br>• Vaccine efficacy of mRNA-1273 to prevent asymptomatic SARS-CoV-2 infection | • Sensitivity analyses with cases counted starting immediately after the second dose of IP, 14 days after the first dose of IP, immediately after the first dose of IP, and immediately after randomization.<br>• Vaccine efficacy and 95% CI based on the case incidence will be estimated with 1 - ratio of incidence rates using the exact method conditional upon the total number of cases. |
| Vaccine efficacy of mRNA-1273 to prevent COVID-19 in all study participants, regardless of evidence of prior SARS-CoV-2 infection | The FAS population will be used for this secondary objective, using similar analysis methods as for the primary endpoint analysis.<br>• Primary analysis: VE will be estimated with 1 - HR (mRNA-1273 vs placebo) using a Cox proportional hazard regression model with treatment group as a fixed effect and adjusting for stratification factor based on the FAS, with cases counted starting 14 days after the second dose of IP.<br>• Sensitivity analyses with cases counted starting immediately after the second dose of IP, 14 days after the first dose of IP, immediately after the first dose of IP, and immediately after randomization. |

The table below summarizes the analysis approach for long-term endpoints.

<table>
<tr><th>Endpoint</th><th>Statistical Analysis Methods</th></tr>
<tr><td>Long-term endpoints:<br>Cases starting 14 days after the second injection of IP for participants in the mRNA-1273 Cohort and the Placebo Cohort, or after the second injection of mRNA-1273 for participants in the Placebo-mRNA-1273 Cohort<ul><li>COVID-19</li><li>Severe COVID-19</li><li>Either COVID-19 or SARS-CoV-2 infection</li><li>Secondary definition of COVID-19</li><li>Death caused by COVID-19</li><li>COVID-19 regardless of evidence of prior SARS-CoV-2 infection determined by serologic titer against SARS-CoV-2 nucleocapsid</li></ul>Cases starting 14 days after the first injection of IP for participants in the mRNA-1273 Cohort and the Placebo Cohort, or after the first injection of mRNA-1273 for participants in the Placebo-mRNA-1273 Cohort<ul><li>COVID-19</li></ul>SARS-CoV-2 infection in the absence of symptoms defining COVID-19 starting 14-days after the second injection of:<ul><li>IP for participants in the mRNA-1273 Cohort and the Placebo Cohort, or mRNA-1273 for participants in the</li></ul></td><td><ul><li>Long-term efficacy will be evaluated after the primary analysis of the primary vaccination series with mRNA-1273 by including data collected in the Open-Label Phase.</li><li>Long-term efficacy data will be summarized descriptively by treatment cohort without cohort comparison.</li><li>In the primary approach, cases will be counted starting 14 days after the second dose of IP for participants in treatment cohorts of mRNA-1273 and Placebo or starting 14 days after the second dose of mRNA-1273 for participants in the Placebo-mRNA-1273 Cohort.</li><li>Sensitivity analyses with cases starting from the second dose, 14 days after the first dose, or first dose of IP for participants in the cohorts of mRNA-1273 and placebo, or mRNA-1273 for participants in the Placebo-mRNA-1273 Cohort will be provided.</li><li>Incidences of cases assessed by numbers, rates, and 2-sided 95% CI based on the exact method adjusting for person-time will be summarized by treatment cohort.</li><li>The Kaplan-Meier analysis will be used to estimate cumulative incidences of time to first cases by treatment cohort.</li><li>Long-term efficacy analysis will be performed using the PP Set and mITT Set.</li><li>Efficacy analyses of unblinded/open-label phase data collected in Part B will be provided, as will the incidence rate of COVID-19 after the BD of mRNA-1273. The details of</li></ul></td></tr>
</table>

| | |
|---|---|
| Placebo-mRNA-1273 Cohort | analysis of long-term efficacy and open-label phase data (Part B and Part C) will be provided in the statistical analysis plan (SAP). |

**Safety:** Safety and reactogenicity will be assessed by clinical review of all relevant parameters. Safety analyses will be provided for Parts A, B, and C separately unless specified otherwise.

All safety analyses will be based on the Safety Set, except summaries of solicited ARs, which will be based on the Solicited Safety Set. All safety analyses will be provided by treatment group, and by treatment cohort as applicable, unless otherwise specified.

The number and percentage of participants with any solicited local AR, with any solicited systemic AR, and with any solicited AR during the 7-day follow-up period after each injection will be provided only for the Blinded Phase (Part A). A 2-sided 95% exact CI using the Clopper-Pearson method will be also provided for the percentage of participants with any solicited AR for each treatment group.

The number and percentage of participants with solicited ARs, unsolicited AEs, SAEs, MAAEs, Grade 3 or higher ARs and AEs, and AEs leading to discontinuation from study vaccine or participation in the study will be summarized. Unsolicited AE will be presented by MedDRA preferred term and system organ class.

For all other safety parameters, descriptive summary statistics will be provided.

Further details will be described in the statistical analysis plan (SAP).

**Immunogenicity:** The secondary immunogenicity endpoints will be analyzed using the Immunogenicity Subset by treatment group, by treatment cohort as applicable, and by baseline SARS-CoV-2 serostatus, unless otherwise specified.

The SAP will describe the complete set of immunogenicity analyses, including the approach to sample individuals into an Immunogenicity Subset for characterizing vaccine immunogenicity and assessing immunological correlates of risk and protection.

Data from quantitative immunogenicity assays will be summarized for each treatment group using positive response rates and geometric means with 95% CIs, for each timepoint for which an assessment is performed. Data from qualitative (ie, yielding a positive or negative result) assays will be summarized by tabulating the frequency of positive responses for each assay by group at each timepoint that an assessment is performed. Analyses will focus on the 2 key immunogenicity time points and the change in marker response between them: Day 1 before the first dose of IP and Day 57 (28 days after the second dose of IP). The SAP will describe the complete set of immunogenicity analyses.

Quantitative levels or geometric mean titer (GMT) of specific bAb with corresponding 95% CI at each timepoint and geometric mean fold rise (GMFR) of specific bAb with corresponding 95% CI at each postbaseline timepoint over pre-dose baseline at Day 1 will be provided by study arm. Descriptive summary statistics including median, minimum, and maximum will also be provided.

GMT of specific nAb with corresponding 95% CI at each timepoint and GMFR of specific nAb with corresponding 95% CI at each post-baseline timepoint over pre-dose baseline at Day 1 will be provided by study arm. Descriptive summary statistics including median, minimum, and maximum will also be provided. For summarizations of group variables values, antibody values reported as below the LLOQ will be replaced by 0.5 × LLOQ. Values that are reported as greater than the upper limit of quantification (ULOQ) without the actual values will be converted to the ULOQ.

The number and percentage of participants with a fold rise ≥ 2, ≥ 3, and ≥ 4 of serum SARS-CoV-2-specific nAb titers and participants with seroresponse from baseline will be provided with 2-sided 95% CI using the Clopper-Pearson method at each post-baseline timepoint. Seroresponse at a participant level may be defined as a change from below the LLOQ to at least 4 times the LLOQ, or at least a 4-fold rise in nAb or vaccine antigen-specific bAb in participants with pre-existing nAb or bAb of at least the LLOQ at baseline. Seroresponse may also be defined for each specific assay assessing nAb or bAb. The definition of seroresponse will be finalized and documented in the SAP.

The GMT of specific nAb for each group and the geometric mean ratio (GMR) of mRNA-1273 versus placebo in Part A with corresponding 2-sided 95% CI will be estimated at each study timepoint using an analysis of covariance (ANCOVA) model with the treatment group and baseline values, if applicable, as explanatory variables, the analysis may adjust for the stratification factor.

#### 1.2. Schema

**Figure 1: Study Flow Diagram: Part A, the Blinded Phase followed by Part B, the Open-Label Phase Including Part C, the Booster Dose Phase**

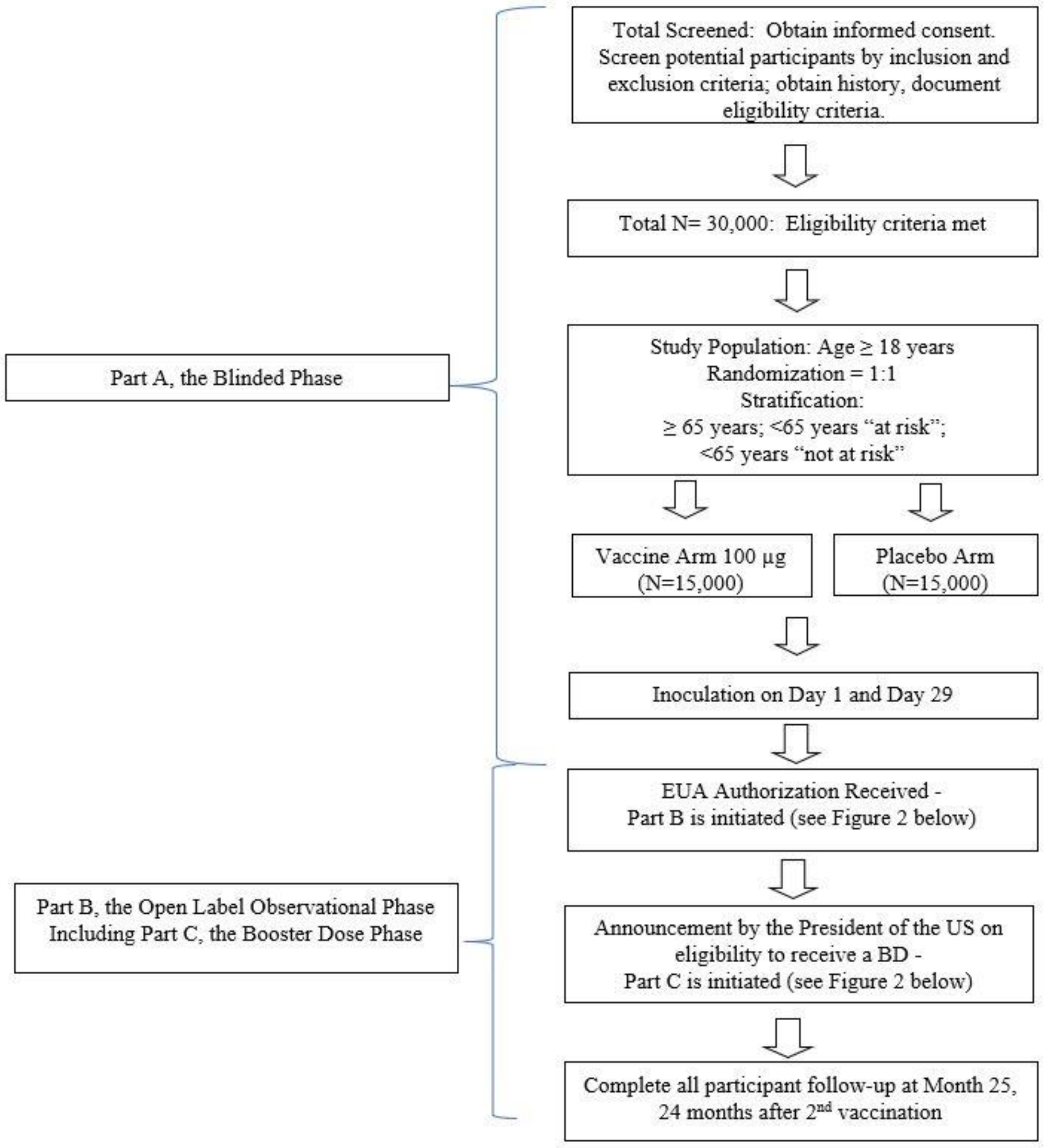

##### Figure 2: Part B, the Open-Label Observational Phase

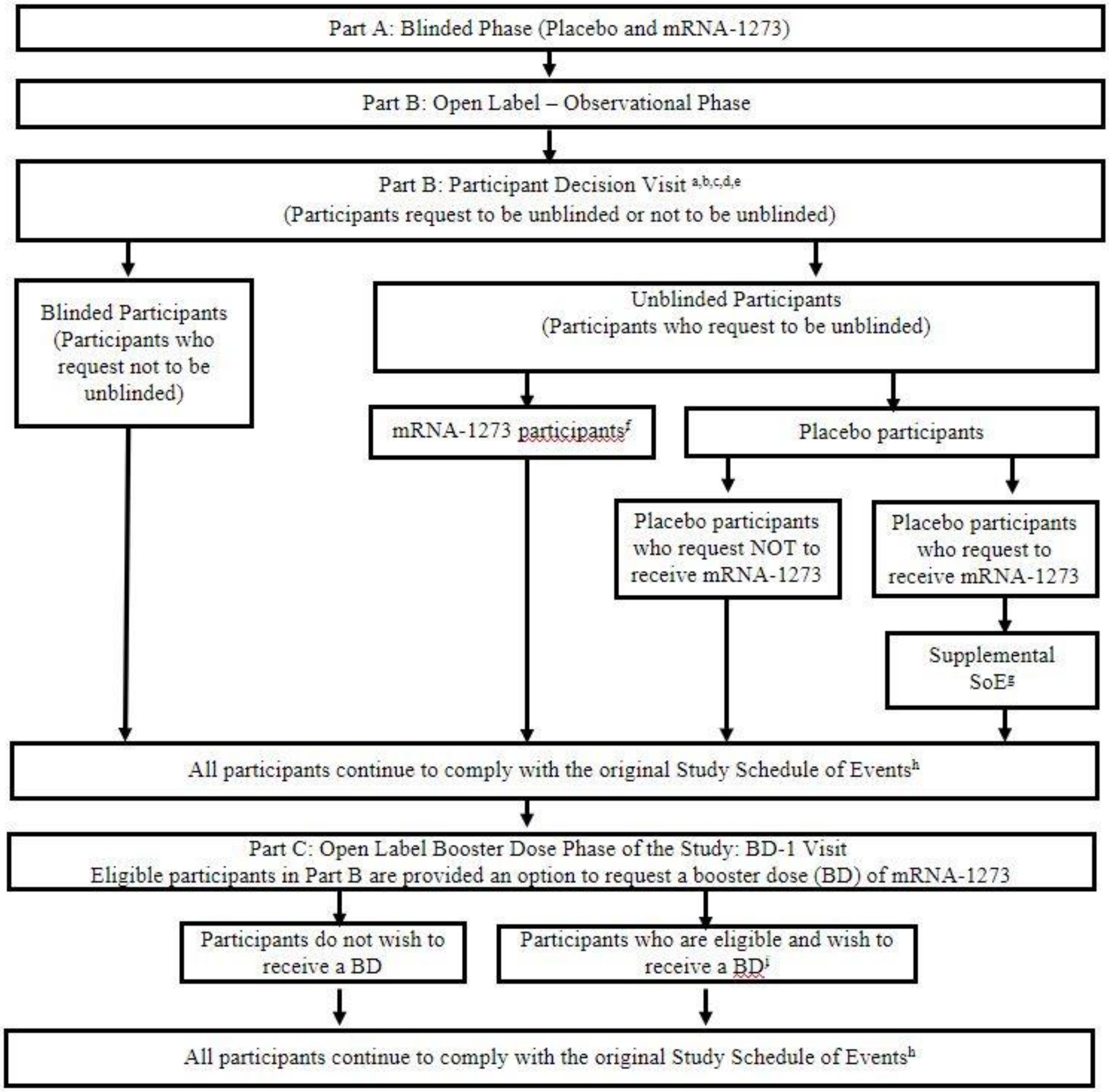

[a] All participants are encouraged to remain in the study.
[b] All participants are given the option to be unblinded to treatment received in Part A: Blinded Phase.
[c] All participants are counselled about the importance of continuing other public health measures to limit the spread of disease including physical-social distancing, wearing a mask, and hand-washing.
[d] All participants sign a revised ICF.
[e] All participants consent to provide a nasopharyngeal swab and a blood sample for immunologic analysis.
[f] mRNA-1273 recipients who only received 1 dose of the mRNA-1273 vaccine will receive the second dose of mRNA-1273 vaccine. Additional details provided in the Supplemental Schedule of Events (Table 22).
[g] Placebo recipients who request to receive mRNA-1273 and meet eligibility, will comply with a Supplemental Schedule of Events (Table 21) in addition to the Original Study Schedule of Events.
[h] Original Study Schedule of Events (Table 16, Table 17, Table 18, and Table 19).
[i] Participants who opt to receive a BD and meet eligibility will comply with the Part C supplemental SoE (Table 23).

5.3. Screen Failures (Part A: Blinded Phase Only) ... 64
6. Study Treatment ... 65
6.1. Investigational Product ... 65
6.2. Dosing and Management of Investigational Product ... 65
6.2.1. Method of Randomly Assigning Participants to Treatment Groups (Part A, Blinded Phase Only) ... 65
6.2.2. Administration of Investigational Product ... 66
6.2.3. Delivery and Receipt ... 67
6.2.4. Packaging and Labeling ... 67
6.2.5. Storage ... 68
6.2.6. Investigational Product Accountability ... 68
6.2.7. Handling and Disposal ... 68
6.2.8. Unblinding ... 69
6.3. Study Treatment Compliance ... 71
6.4. Prior and Concomitant Therapy ... 72
6.4.1. Prior Medications and Therapies ... 72
6.4.2. Concomitant Medications and Therapies ... 72
6.4.3. Concomitant Medications and Vaccines that May Lead to the Elimination of a Participant from Per-protocol Analyses ... 73
7. Delaying or Discontinuing Study Treatment and Participant Withdrawal from the Study ... 74
7.1. Criteria for Delay of Study Treatment ... 74
7.2. Discontinuation of Study Treatment ... 74
7.3. Participant Withdrawal from the Study ... 76
7.3.1. Participant Withdrawal ... 76
7.4. Lost to Follow-up ... 77
8. Study Assessments and Procedures ... 78
8.1. Efficacy and Immunogenicity Assessments and Procedures ... 79
8.1.1. Efficacy Assessments Related to COVID-19 and SARS-CoV-2 Infection ... 79
8.1.2. Surveillance for COVID-19 Symptoms ... 80
8.1.3. Convalescent Period Starting with the Illness Visit ... 83
8.1.4. Ancillary Supplies for Participant Use ... 85
8.1.5. Immunogenicity Assessments ... 85
8.2. Safety Assessments ... 86
8.2.1. Safety Phone Calls ... 87

9.5. Statistical Analyses ... 112
9.5.1. Efficacy Analyses ... 112
9.5.2. Safety Analyses ... 118
9.5.3. Immunogenicity Analyses ... 119
9.5.4. Exploratory Analyses ... 121
9.5.5. Subgroup Analyses ... 121
9.6. Interim Analyses ... 122
10. References ... 124
11. Supporting Documentation and Operational Considerations ... 128
11.1. APPENDIX 1: Schedules of Events ... 129
11.2. APPENDIX 2: Study Governance Considerations ... 151
11.2.1. Regulatory and Ethical Considerations ... 151
11.2.2. Study Monitoring ... 151
11.2.3. Audits and Inspections ... 153
11.2.4. Financial Disclosure ... 153
11.2.5. Recruitment Procedures ... 153
11.2.6. Informed Consent Process ... 153
11.2.7. Protocol Amendments ... 155
11.2.8. Protocol Deviations ... 155
11.2.9. Data Protection ... 155
11.2.10. Sample Retention and Future Biomedical Research ... 156
11.2.11. Data Quality Assurance and Quality Control ... 157
11.2.12. Data Collection and Management ... 158
11.2.13. Source Documents ... 158
11.2.14. Retention of Records ... 159
11.2.15. Study and Site Closure ... 159
11.2.16. Publication Policy ... 160
11.3. APPENDIX 3: Contraceptive Guidance ... 161
11.4. APPENDIX 4: Statistical Appendices ... 162
11.4.1. Estimands and Estimand Specifications ... 162
11.4.2. Statistical Methods and Sensitivity Analyses ... 164
11.5. APPENDIX 5: Adverse Events of Special Interest Terms ... 165
11.6. APPENDIX 6: Protocol Amendment History ... 167

### LIST OF ABBREVIATIONS

The following abbreviations and terms are used in this study protocol.

| Abbreviation | Definition |
|---|---|
| AC | adjudication committee |
| AE | adverse event |
| AESI | adverse event of special interest |
| AR | adverse reaction |
| ARDS | acute respiratory distress syndrome |
| bAb | binding antibody |
| BARDA | Biomedical Advance Research and Development Authority |
| BD | booster dose |
| BLA | Biologics License Application |
| BOD | burden of disease |
| CBER | Center for Biologics Evaluation and Research |
| CDC | US Centers for Disease Control and Prevention |
| CEAC | Cardiac Event Adjudication Committee |
| CFR | Code of Federal Regulations |
| CI | confidence interval |
| CLIA | Clinical Laboratory Improvement Amendments |
| cMRI | cardiac magnetic resonance imaging |
| CMV | cytomegalovirus |
| CONSORT | Consolidated Standards of Reporting Trials |
| CoV | coronavirus |
| COV-INF | virologically confirmed SARS-CoV-2 infection regardless of symptomology or severity |
| CRO | contract research organization |
| CSR | clinical study report |
| DHHS | Department of Health and Human Services |
| DMID | Division of Microbiology and Infectious Diseases |
| DSMB | Data and Safety Monitoring Board |
| DSPC | 1,2-distearoyl-sn-glycero-3-phosphocholine |
| eCRF | electronic case report form |
| eDiary | electronic diary |
| EUA | Emergency Use Authorization |
| ART | antiretroviral therapy |
| FAS | full analysis set |

| Abbreviation | Definition |
|---|---|
| FDA | Food and Drug Administration |
| FSH | follicle-stimulating hormone |
| GCP | Good Clinical Practice |
| GMFR | geometric mean fold rise |
| GMP | Good Manufacturing Practice |
| GMT | geometric mean titer |
| HCP | healthcare practitioner |
| HIV | Human Immunodeficiency Virus |
| hMPV | human metapneumovirus |
| HR | hazard ratio |
| HRT | hormonal replacement therapy |
| IA | interim analysis |
| IB | investigator's brochure |
| ICF | informed consent form |
| ICH | International Council for Harmonisation |
| IM | intramuscular |
| IP | investigational product |
| IRB | institutional review board |
| IRT | interactive response technology |
| LB | lower boundary |
| LLOQ | lower limit of quantification |
| LNP | lipid nanoparticle |
| LOD | limit of detection |
| LTFU | lost to follow-up |
| MAAE | medically attended adverse event |
| MedDRA | Medical Dictionary for Regulatory Activities |
| MERS-CoV | Middle East Respiratory Syndrome coronavirus |
| mRNA | messenger RNA |
| nAb | neutralizing antibody |
| NIAID | National Institute of Allergy and Infectious Diseases |
| NP | nasopharyngeal |
| PCR | polymerase chain reaction |
| PDV | participant decision visit |
| PEG2000-DMG | 1-monomethoxypolyethyleneglycol-2,3-dimyristylglycerol with polyethylene glycol of average molecular weight 2000 |

| Abbreviation | Definition |
|---|---|
| PIV3 | parainfluenza virus type 3 |
| PP | per-protocol |
| RT-PCR | reverse transcriptase polymerase chain reaction |
| S | spike |
| S-2P | spike protein with 2 proline residues introduced for stability in a prefusion conformation |
| SAE | serious adverse event |
| SAP | statistical analysis plan |
| SARS-CoV | Severe Acute Respiratory Syndrome coronavirus |
| SM-102 | heptadecan-9-yl 8-((2-hydroxyethyl) (6-oxo-6-(undecyloxy) hexyl) amino) octanoate |
| SoE | Schedule of Events |
| TEAE | treatment-emergent adverse event |
| ULOQ | upper limit of quantification |
| USP | United States Pharmacopeia |
| VE | vaccine efficacy |
| WHO | World Health Organization |

### 2. INTRODUCTION

#### 2.1. Study Rationale

Coronaviruses (CoVs) are a large family of viruses that cause illness ranging from the common cold to more severe diseases, such as Middle East Respiratory Syndrome (MERS-CoV) and Severe Acute Respiratory Syndrome (SARS-CoV).

An outbreak of the CoV disease (COVID-19) caused by SARS-CoV-2 began in Wuhan, Hubei Province, China in December 2019 and has spread throughout China and to over 216 other countries and territories, including the United States (WHO 2020). On 11 March 2020, the World Health Organization (WHO) officially declared COVID-19 a pandemic. As of 28 May 2020, the WHO reported more than 5,593,631 confirmed cases and 353,334 deaths globally and the US Centers for Disease Control and Prevention (CDC) reported 1,698,523 confirmed and probable cases of COVID-19, with 100,446 deaths in the United States (CDC 2020a). The CDC have reported that the highest risk of disease burden is in older adults (≥ 65 years old) and people of any age who have serious underlying medical conditions, such as chronic lung disease or moderate to severe asthma; serious heart conditions; severe obesity; diabetes; chronic kidney disease requiring dialysis; liver disease; and those who are immunocompromised (CDC 2020b).

At the time the present Phase 3 study was initiated, no vaccine against SARS-CoV-2 had been authorized or approved. Global efforts to evaluate novel antivirals and therapeutic strategies to treat severe SARS-CoV-2 infections have intensified, but no proven therapeutic currently exists. Therefore, there is an urgent public health need for rapid development of novel interventions to prevent the spread of this disease. The primary goal of this Phase 3 study is to evaluate the vaccine efficacy (VE) of mRNA-1273 to prevent COVID-19, compared to placebo.

#### 2.2. Background and Overview

ModernaTX, Inc. (the Sponsor) has developed a rapid response, proprietary vaccine platform based on a messenger RNA (mRNA) delivery system. The platform is based on the principle and observations that cells in vivo can take up mRNA, translate it, and then express protein viral antigen(s) on the cell surface. The delivered mRNA does not enter the cellular nucleus or interact with the genome, is nonreplicating, and is expressed transiently. mRNA vaccines have been used to induce immune responses against infectious pathogens such as cytomegalovirus (CMV) (NCT03382405), human metapneumovirus (hMPV) and parainfluenza virus type 3 (PIV3) (NCT03392389), Zika virus (NCT03325075), and influenza virus (NCT03076385 and NCT03345043).

The Sponsor is using its mRNA-based platform to develop a novel lipid nanoparticle (LNP)-encapsulated mRNA-based vaccine against SARS-CoV-2 (mRNA-1273). mRNA-1273 encodes for the full-length spike (S) protein of SARS-CoV-2, modified to introduce 2 proline residues to stabilize the S protein (S2P) in a prefusion conformation. The CoV S protein mediates attachment and entry of the virus into host cells (by fusion), making it a primary target for neutralizing antibodies that prevent infection (Johnson et al 2016; Wang et al 2015; Wang et al 2018; Chen et al 2017; Corti et al 2015; Yu et al 2015; Kim et al 2019; Widjaja et al 2019). It has been confirmed that the stabilized SARS-CoV-2 S2P antigen presents in the correct prefusion conformation (Wrapp et al 2020).

The development of the mRNA-1273 vaccine is being accelerated to address the current SARS-CoV-2 outbreak as a result of the uniquely rapid and scalable manufacturing process for mRNA-1273. The primary goal of this Phase 3 study is to evaluate the VE of mRNA-1273 to prevent COVID-19, compared to placebo.

##### 2.2.1. Nonclinical Studies

Nonclinical studies have demonstrated that CoV S proteins are immunogenic and S protein-based vaccines, including those based on mRNA delivery platforms, are protective in animals. Prior clinical studies of vaccines targeting related CoVs and other viruses have demonstrated that mRNA-based vaccines are safe and immunogenic. It is therefore anticipated that mRNA-1273 will generate robust immune responses to the SARS-CoV-2 S protein and will be well tolerated. In addition, mRNA-1273 has shown preliminary evidence of protection against SARS-CoV-2 in a murine model of infection (data on file).

In support of development of mRNA-1273 for prophylaxis against SARS-CoV-2 infection, nonclinical immunogenicity, biodistribution, and safety studies have been completed with similar mRNA-based vaccines formulated in LNPs containing SM-102 (heptadecan-9-yl 8 ((2 hydroxyethyl)(6 oxo 6-(undecyloxy)hexyl)amino)octanoate), the novel proprietary lipid used in the mRNA-1273 LNP formulation.

A detailed review of nonclinical experience with mRNA-1273 vaccine is provided in the Investigator's Brochure (IB).

##### 2.2.2. Clinical Studies

The mRNA-1273 vaccine is currently being evaluated for safety and immunogenicity in a dose-ranging Phase 1 study (NCT04283461) sponsored and conducted by the Division of Microbiology and Infectious Diseases (DMID) of the National Institute of Allergy and Infectious Diseases (NIAID). The Phase 1 DMID study is an open-label dose-ranging study of mRNA-1273 in healthy adult male and non-pregnant female participants in 3 age groups: age 18 to 55 years, inclusive (45 participants); age 56 to 70 years, inclusive (30 participants); and ≥ 71 years (30 participants). Each participant received an IM injection (0.5 mL) of mRNA-1273 on Days 1 and 29 in the deltoid muscle and will be followed for 13 months after the second injection. Sera obtained from 41 participants who were convalescing from COVID-19 were included as a comparative control.

A total of 120 participants were enrolled, and 116 participants received the second injection. Three mRNA-1273 dose levels (25, 50, and 100 µg) administered 28 days apart have been assessed in participants 18 to 55 years, 56 to 70 years, and ≥ 71 years of age. The mRNA-1273 250-µg dose was not evaluated in participants 56 to 70 years and ≥ 71 years of age due to reactogenicity observed in 4 participants in the 250 µg (18 to 55 years) dose cohort. Across all age groups, solicited adverse reactions (ARs) were predominantly mild or moderate in severity and most commonly included fatigue, chills, headache, myalgia, and pain at the injection site. The incidence and severity of solicited ARs were generally dose dependent and increases in incidence and severity were generally observed after the second injection. The incidence of systemic reactions increased after the second injection, particularly at the highest (250 µg) dose. Four (27%) participants in the 250 µg dose cohort reported at least 1 severe solicited AR after the second injection, including feverishness, fatigue, fever, headache, myalgia, nausea, and erythema/redness. No serious adverse events (SAEs) were reported through Day 119, and no pause rules were triggered during the study. mRNA-1273 induced robust binding antibody (bAb) responses after 2 injections. Notably, 100 µg of mRNA-1273 resulted in numerically higher S-2P specific binding titers than 25 µg and 50 µg of mRNA-1273 in all age groups. Two injections of mRNA-1273 also induced robust neutralizing antibody (nAb) responses. The nAb response was marginal after the first injection but increased substantially after the second injection. The nAb response assessed by a pseudovirus neutralization assay at the 100 µg mRNA-1273 dose level was similar to that at the 250 µg mRNA-1273 dose level in the 18 to 55 years of age group and numerically higher than that observed at the 25 µg and 50 µg mRNA-1273 dose levels in the older age groups. The neutralizing antibody response at the 100 µg dose level was similar across all age groups.

mRNA-1273-P201 (Study 201) is an ongoing Phase 2a, safety, reactogenicity, and immunogenicity study in healthy adults that provided confirmation of the immunogenicity of both the 100 µg and 50 µg doses. The study was designed as a randomized, observer-blind, placebo-controlled dose confirmation study (Part A). Two mRNA-1273 dose levels, 50 µg and 100 µg, and placebo were evaluated in 2 age cohorts: Cohort 1 enrolled participants ≥ 18 to < 55 years old (300 participants), and Cohort 2 enrolled participants ≥ 55 years old (300 participants). A total of 600 participants received either mRNA-1273 or placebo according to a 1:1:1 randomization ratio; ie, within each age cohort, 100 participants each received mRNA-1273 50 µg, mRNA-1273 100 µg, or placebo.

An amendment to the Study 201 protocol adapted the study design to include open-label interventional phases (Part B and Part C). Part B allowed unblinding of participants and offered 2 injections of mRNA-1273 in an open-label manner, 28 days apart, to all participants who received placebo in Part A. Part B also offered a single booster dose (BD) of mRNA-1273 (50 µg) to participants who received 1 or 2 doses of mRNA-1273 (50 µg or 100 µg) in Part A. Part C was prompted by the need to proactively prepare for vaccination strategies that induce broader protection, including against emerging variants of SARS-CoV-2 such as B.1.351. Part C enrolled participants from Study 301 who received 2 doses of mRNA-1273 100 µg at least 6 months prior. Part C participants received a single injection of mRNA-1273.351 (20 µg or 50 µg) or mRNA-1273.351 mixture (50 µg total – 25 µg of mRNA-1273 and 25 µg of mRNA-1273.351). mRNA-1273 demonstrated an acceptable safety profile in the participant population enrolled in this study at both dose levels and both age cohorts, as observed through 6 months after the second injection.

Vaccination with mRNA-1273 in Study 201 resulted in robust immune responses to SARS-CoV-2 in participants 18 years and older at both dose levels, and persistence of immune response was observed up to 6 months after the second injection in Part A of the study. In Part A of Study 201, the time course and magnitude of antibody (both bAb and nAb) responses to mRNA-1273 was similar between 100 µg and 50 µg dose levels at each postbaseline time point (Days 29, 43, 57, and 209), although the 100 µg dose group had numerically greater responses. In Part B of Study 201, administration of a 50 µg BD of mRNA-1273 6 months or more after the primary series improved the immune responses to 1.7-fold the peak achieved after the primary vaccination series in the current mRNA-1273-P301 study, where efficacy of mRNA-1273 against COVID-19 was demonstrated.

This present ongoing Phase 3 study (mRNA-1273-P301) is a pivotal Phase 3, efficacy, safety, and immunogenicity study that has provided the primary clinical evidence of VE and safety (see Section 4 for details of the study design). The study was designed as a randomized, observer and participant blind, placebo-controlled study of the efficacy, safety, and immunogenicity of mRNA 1273 compared to placebo. More than 30,000 participants 18 years of age and older were randomized 1:1 to mRNA-1273 100 µg or placebo based on 3 strata: ≥ 65 years of age, 18 to < 65 years of age and at increased risk for complications of COVID-19, and 18 to < 65 years of age and not at risk. The initial mRNA-1273 100 µg dose was followed by a second 100 µg dose 28 days later. Participants are being followed for efficacy and safety until 24 months after the second dose. Vaccine efficacy was demonstrated based on the prespecified efficacy success criterion at the interim analysis (11 Nov 2020 dataset), based on a total of 95 adjudicated COVID-19 cases. The subsequent primary analysis of efficacy was performed with a total of 196 adjudicated COVID 19 cases (25 Nov 2020 dataset) and was consistent with the IA. mRNA-1273 was subsequently granted emergency use authorization (EUA) in the US and conditional approvals worldwide.

After EUA in the US was granted for mRNA-1273 and another mRNA COVID-19 vaccine, Part B, the Open-Label Observational Phase of the study was initiated. All participants in Part A were invited to proceed to Part B, starting with a participant decision visit (PDV) at the study site, at which participants were given the option to be unblinded to their original group assignment or remain blinded. Unblinded participants who had received placebo in Part A had the choice to be vaccinated with mRNA-1273 in Part B.

On 25 Aug 2021, Moderna announced that it had completed the rolling submission process for its Biologics License Application (BLA) to the US Food and Drug Administration (FDA) for the full licensure of the Moderna COVID-19 vaccine (mRNA-1273) for active immunization to prevent COVID-19 in individuals 18 years of age and older. mRNA-1273 has been showing durable efficacy of 93% through 6 months after the second dose. mRNA-1273 was highly immunogenic as measured by both bAb and nAb in both SARS-CoV-2 baseline-negative and baseline-positive individuals, as indicated by increased bAb and nAb levels 1 month after first injection (Day 29) and 1 month after second injection (Day 57). mRNA-1273 demonstrated an acceptable safety profile in the participant population enrolled in this study, having a reactogenicity profile consistent with parenteral vaccination and being generally well tolerated. No unexpected findings were identified in the final assessment of the randomized, blinded phase of the study (Part A).

The present version of protocol mRNA-1273-P301 (Amendment 9) initiates Part C of the study (Section 4.1.3), providing for administration of a BD of mRNA-1273 in Part B, the Open-Label Observational Phase of the study (Section 4.1.2)

#### 2.3. Benefit/Risk Assessment

##### 2.3.1. Potential Benefits of Study Participation

The target study population for this study is adults with no known history of SARS-CoV-2 infection but whose locations or circumstances put them at high risk of COVID-19. The following benefits may accrue to participants:

- The mRNA-1273 vaccine may be an effective vaccine against COVID-19.
- A baseline (Day 1) evaluation for SARS-CoV-2 infection and ongoing surveillance for COVID-19 throughout the study.
- Contributing to the development of a vaccine against COVID-19, a current pandemic disease.

##### 2.3.2. Risks from Study Participation

Immediate systemic allergic reactions (eg, anaphylaxis) can occur following any vaccination. These reactions are very rare and are estimated to occur once per 450,000 vaccinations for vaccines that do not contain allergens such as gelatin or egg protein (Zent et al 2002). As a precaution, all participants will remain under observation at the study site for at least 30 minutes after vaccination.

Vasovagal syncope (fainting) can occur before or after any vaccination, is usually triggered by the pain or anxiety caused by the injection and is not related to the substance injected. Therefore, it is important that standard precautions and procedures be followed to avoid injury from fainting.

Intramuscular injection with other mRNA vaccines manufactured by the Sponsor containing the SM-102 lipid formulation commonly results in a transient and self-limiting local inflammatory reaction. This typically includes pain, erythema (redness), or swelling (hardness) at the injection site, which are mostly mild to moderate in severity and usually occur within 24 hours of vaccination. More severe, but self-limited, local reactions, erythema and induration, have been observed at dose of mRNA-1273 exceeding the dose proposed in this study.

Most systemic adverse events (AEs) observed after vaccination do not exceed mild to moderate severity. The most commonly reported systemic ARs are anticipated to be fever, fatigue, chills, headache, myalgias and arthralgias. More severe reactions, including erythema, induration, fever, headache and nausea, were reported after receiving doses of mRNA-1273 that were greater than the dose proposed for use in this study. In all cases, the reactions resolved spontaneously.

Laboratory abnormalities (including increases in liver functional tests and serum lipase levels) following vaccination were observed in clinical studies with similar mRNA-based vaccines. These abnormalities were without clinical symptoms or signs and returned toward baseline (Day 1) values over time. The clinical significance of these observations is unknown. Further details are provided in the current IB.

There is a theoretical risk that active vaccination to prevent the novel viral infection caused by SARS-CoV-2 may cause a paradoxical increase in the risk of disease. This possibility is based on the rare phenomenon of vaccine-associated disease enhancement which was first seen in the 1960s with 2 vaccines made in the same way (formalin-inactivated whole virus) and designed to protect children against infection with RSV (Chin et al 1969) or measles (Fulginiti et al 1967). Disease enhancement has also been proposed as a possible explanation for cases of more serious disease associated with dengue vaccination (Thomas and Yoon 2019; WHO 2018).

To monitor the risk of enhanced disease in this study, an independent Data and Safety Monitoring Board (DSMB) will review unblinded cases of COVID-19 to assess for inefficacy and also for numerical imbalance in cases of both COVID-19 and severe COVID-19 with the purpose of providing a non-binding recommendation to the Sponsor (Section 8.4.2). To date, clinical immunogenicity data from the DMID Phase 1 study of mRNA-1273 demonstrated high levels of nAbs and Th1 polarized CD4+ T-cell responses (Jackson et al 2020). In addition, in Study P301, after a median follow-up of 2 months after the second dose of vaccine, the overwhelming majority of COVID-19 cases occurred in participants who received placebo rather than mRNA-1273 (Baden et al 2020), consistent with a low risk of ERD following vaccination with mRNA-1273.

These data suggest that a paradoxical increase in the risk of disease, while not eliminated, is likely to be low. Further details are provided in the current investigator's brochure (IB).

In the context of the EUA for individuals 18 years and older for mRNA-1273, there have been very rare reports of myocarditis and pericarditis occurring after vaccination with Moderna COVID-19 vaccine. Although causality has not been established, the majority of the cases have been reported in young males shortly after the second dose of the vaccine. These are typically mild cases and individuals tend to recover within a short time following standard treatment and rest (Gargano et al 2021).

The AEs after getting a third dose of mRNA-1273 are currently being studied. It is unknown if the AEs after getting a third dose of mRNA-1273 are different from getting 2 doses of mRNA-1273.

##### 2.3.3. Overall Benefit/Risk Conclusion

All participants will be included based on their increased risk of SARS-CoV-2 infection. Accordingly, all will benefit from baseline and ongoing evaluations for SARS-CoV-2 infection.

Since this is a placebo-controlled study (Section 4), half the participants will have the potential to receive mRNA-1273 vaccine, the efficacy of which is unknown at present. Vaccination with mRNA-1273 may not prevent COVID-19 in all vaccines.

Participants who receive placebo as part of this study may have an opportunity cost of not being treated with another investigational vaccine against COVID-19.

The placebo for this study is a saline solution, without any LNP. Thus, participants receiving saline may be at lower risk of AEs related to injection than participants receiving mRNA-1273.

Safety findings will be monitored and periodically reviewed by the DSMB to evaluate the safety and treatment status of all participants. The DSMB will review and assess the safety data as described in Section 8.4.2.

Considering the lack of approved vaccines for COVID-19, the participants' risk of COVID-19 outside the study, and the nonclinical and clinical data to date, the Sponsor considers the potential benefits of participation to exceed the risks.

Based on the interim results from the pivotal Phase 3 study, mRNA-1273 prevents COVID-19 and severe COVID-19. The demonstrated clinical benefit of mRNA-1273 is supported by evidence of a robust immune response both in terms of bAbs and nAbs as well as the induction of CD4+ T-cells with a Th-1 dominant phenotype. Based on administration of mRNA-1273 to approximately 15,693 adults across all 3 clinical studies to date, there have been no emergent safety concerns and the AE profile is manifested largely by mild to moderate reactogenicity lasting 2 to 3 days.

### 3. OBJECTIVES AND ENDPOINTS

#### Table 1: Objectives and Endpoints

| **Objectives and Endpoints** | |
|---|---|
| **Primary Objective** | **Primary Endpoints** |
| **Efficacy Objective (Primary):**<br>To demonstrate the efficacy of mRNA-1273 to prevent COVID-19. | **Efficacy Endpoints (Primary):** Vaccine efficacy of mRNA-1273 to prevent the first occurrence of COVID-19 starting 14 days after the second dose of investigational product (IP), where COVID-19 is defined as symptomatic disease based on the following criteria:<br>• The participant must have experienced at least TWO of the following systemic symptoms: Fever (≥ 38ºC), chills, myalgia, headache, sore throat, new olfactory and taste disorder(s), OR<br>• The participant must have experienced at least ONE of the following respiratory signs/symptoms: cough, shortness of breath or difficulty breathing, OR clinical or radiographical evidence of pneumonia; AND<br>• The participant must have at least 1 NP swab or saliva sample (or respiratory sample, if hospitalized) positive for SARS-CoV-2 by RT-PCR |
| **Safety Objective (Primary):**<br>To evaluate the safety and reactogenicity of 2 injections of the mRNA-1273 vaccine given 28 days apart. | **Safety Endpoint (Primary):**<br>• Solicited local and systemic ARs through 7 days after each dose of IP (Part A: Blinded Phase only).<br>• Unsolicited AEs through 28 days after each dose of IP (Part A: Blinded Phase only).<br>• MAAEs or AEs leading to withdrawal through the entire study period.<br>• SAEs throughout the entire study period. |

| **Objectives and Endpoints** | |
|---|---|
| **Efficacy Objectives (Secondary)** | **Efficacy Endpoints (Secondary)** |
| To evaluate the efficacy of mRNA-1273 to prevent severe COVID-19 | Vaccine efficacy of mRNA-1273 to prevent severe COVID-19, defined as first occurrence of COVID-19 starting 14 days after the second dose of IP, (as per the primary endpoint) AND any of the following:<br>• Clinical signs indicative of severe systemic illness, Respiratory Rate $\geq$ 30 per minute, Heart Rate $\geq$ 125 beats per minute, $SpO_2 \leq 93\%$ on room air at sea level or $PaO_2/FIO_2$ < 300 mm Hg, OR<br>• Respiratory failure or Acute Respiratory Distress Syndrome (ARDS), (defined as needing high-flow oxygen, non-invasive or mechanical ventilation, or ECMO), evidence of shock (systolic blood pressure < 90 mmHg, diastolic BP < 60 mmHg or requiring vasopressors), OR<br>• Significant acute renal, hepatic or neurologic dysfunction, OR<br>• Admission to an intensive care unit or death |
| To evaluate the efficacy of mRNA-1273 to prevent serologically confirmed SARS-CoV-2 infection or COVID-19 regardless of symptomatology or severity. | Vaccine efficacy of mRNA-1273 to prevent the first occurrence of either COVID-19 or SARS-CoV-2 infection starting 14 days after the second IP dose.<br>This endpoint is a combination of COVID-19, defined as for the primary endpoint, and asymptomatic SARS-CoV-2 infection, determined by seroconversion assessed by bAb levels against SARS-CoV-2 as measured by a ligand-binding assay specific to the SARS-CoV-2 nucleocapsid protein and with a negative nasopharyngeal (NP) swab sample for SARS-CoV-2 at Day 1 (Section 8.1.1). |

| **Objectives and Endpoints** | |
|---|---|
| To evaluate VE against a secondary definition of COVID-19. | Vaccine efficacy of mRNA-1273 to prevent the secondary case definition of COVID-19 starting 14 days after the second IP dose.<br>The secondary case definition of COVID-19 is defined as the following systemic symptoms: fever (temperature ≥ 38ºC), or chills, cough, shortness of breath or difficulty breathing, fatigue, muscle aches or body aches, headache, new loss of taste or smell, sore throat, nasal congestion or rhinorrhea, nausea or vomiting, or diarrhea AND a positive NP swab or saliva sample (or respiratory sample, if hospitalized) for SARS-CoV-2 by RT-PCR |
| To evaluate VE to prevent death caused by COVID-19. | Vaccine efficacy of mRNA-1273 to prevent death due to a cause directly attributed to a complication of COVID-19, starting 14 days after the second IP dose. |
| To evaluate the efficacy of mRNA-1273 to prevent COVID-19 after the first dose of IP. | Vaccine efficacy of mRNA-1273 to prevent the first occurrence of COVID-19 starting 14 days after the first dose of IP. |
| To evaluate the efficacy of mRNA-1273 to prevent COVID-19 in all study participants, regardless of evidence of prior SARS-CoV-2 infection. | Vaccine efficacy of mRNA-1273 to prevent the first occurrence of COVID-19 starting 14 days after the second dose of IP regardless of evidence of prior SARS-CoV-2 infection determined by serologic titer against SARS-CoV-2 nucleocapsid (FAS analysis population, see Section 9.4). |
| To evaluate the efficacy of mRNA-1273 to prevent asymptomatic SARS-CoV-2 infection. | Vaccine efficacy to prevent the first occurrence of SARS-CoV-2 infection in the absence of symptoms defining COVID-19 starting 14 days after the second IP dose.<br>SARS-CoV-2 infection determined by seroconversion assessed by bAb levels against SARS-CoV-2 as measured by a ligand-binding assay specific to the SARS-CoV-2 nucleocapsid protein and with a negative NP swab sample for SARS-CoV-2 at Day 1 (Section 8.1.1). |

| **Objectives and Endpoints** | |
|---|---|
| **Immunogenicity Objective (Secondary):** | **Immunogenicity Endpoints (Secondary):** |
| To evaluate the immunogenicity of 2 doses of mRNA-1273 given 28 days apart. | • Geometric mean titer (GMT) of SARS-CoV-2 -specific neutralizing antibody (nAb) on Day 1, Day 29, Day 57, Day 209, Day 394, and Day 759.<br>• Geometric mean fold rise (GMFR) of SARS-CoV-2-specific nAb relative to Day 1 on Day 29, Day 57, Day 209, Day 394, and Day 759.<br>• Quantified levels or GMT of S protein-specific binding antibody (bAb) on Day 1, Day 29, Day 57, Day 209, Day 394, and Day 759.<br>• GMFR of S protein -specific bAb relative to Day 1 on Day 29, Day 57, Day 209, Day 394, and Day 759. |
| **Exploratory Objectives for Part A** | |
| To evaluate the effect of mRNA-1273 on the viral infection kinetics as measured by viral load at SARS-CoV-2 infection diagnosis by RT-PCR and number of days from the estimated date of SARS-CoV-2 infection until undetectable SARS-CoV-2 infection by RT-PCR. | |
| To assess VE to reduce the duration of symptoms of COVID-19. | |
| To evaluate VE against all-cause mortality. | |
| To assess VE against burden of disease (BOD) due to COVID-19. | |
| To evaluate the genetic and/or phenotypic relationships of isolated SARS-CoV-2 strains to the vaccine sequence. | |
| To evaluate immune response markers after dosing with IP as correlates of risk of COVID-19 and as correlates of risk of SARS-CoV-2 infection. | |
| To conduct additional analyses related to furthering the understanding of SARS-CoV-2 infection and COVID-19, including analyses related to the immunology of this or other vaccines, detection of viral infection, and clinical conduct. | |
| **Exploratory Objectives for Part C** | |
| To evaluate the safety of a booster dose of mRNA-1273. | |
| To evaluate the immunogenicity of a booster dose of mRNA-1273. | |

Abbreviations: AE = adverse event; AR = adverse reaction; bAb = binding antibody; FAS = full analysis set; GMFR = geometric mean fold rise; GMT = geometric mean titer; ICU = intensive care unit; MAAE = medically attended adverse event; nAb = neutralizing antibody; NP = nasopharyngeal; SAE = serious adverse event; VE = vaccine efficacy.

### 4. STUDY DESIGN

#### 4.1. General Design

This is a 3-part Phase 3 study, comprising Part A, Part B, and Part C. Participants in Part A, the Blinded Phase of this study were blinded to their treatment assignment. Given that the primary efficacy endpoint for mRNA-1273 against COVID-19 was met per the protocol-defined IA, Part B, the Open-Label Observational Phase of this study, was designed to offer eligible participants who received placebo in Part A of this study an option to request open-label mRNA-1273, subject to availability of investigational vaccine (Figure 1). Part C, providing for an mRNA-1273 booster dose (BD) in Part B, is designed to offer eligible participants in Part B the option to request a booster dose of mRNA-1273.

Upon entry into Part A, all participants had 8 scheduled clinic visits, including Screening, Day 1, Day 29, Day 57, Day 209, Day 394, Day 759, as specified in the schedules of events (SoEs) (Table 16, Table 17, Table 18Table 19). With the initiation of Part B, a PDV was scheduled for all participants currently enrolled in Part A at that time (Table 20) and participants eligible for Part B dosing may have had up to 2 additional scheduled clinic visits as specified in SoEs (Table 21 and Table 22). Part C will begin with a BD-1 visit (see SoE; Table 23) scheduled for all participants currently enrolled in Part B, and participants deciding to receive a BD may have up to 3 additional clinic visits scheduled (Section 4.1.3;Table 23).

This study will be conducted in compliance with the protocol, Good Clinical Practice (GCP), and all applicable regulatory requirements.

##### 4.1.1. Part A, the Blinded Phase

The Blinded Phase of this study is a randomized, stratified, observer-blind, placebo-controlled evaluation of the efficacy, safety, and immunogenicity of mRNA-1273 SARS-CoV-2 vaccine compared to placebo in adults 18 years of age and older who have no known history of SARS-CoV-2 infection but whose locations or circumstances put them at appreciable risk of acquiring COVID-19 and/or SARS-CoV-2 infection. Figure 1 shows the study flow and Appendix 1 (Table 16, Table 17, Table 18, and Table 19) presents the planned SoEs.

Approximately 30,000 participants will be randomly assigned to receive doses of either 100 µg of mRNA-1273 vaccine or a placebo control in a 1:1 randomization ratio. Assignment will be stratified by age and health risk (Section 6.2.1.1). This is a case-driven study and thus final sample size of the study will depend on the actual attack rate of COVID-19.

All participants will be assessed for efficacy and safety endpoints and provide an NP swab sample and blood sample before the first and second dose of IP, in addition to a series of post-dose blood samples for immunogenicity through 24 months after the second dose of IP. Efficacy assessments will include surveillance for COVID-19 with RT-PCR confirmation of SARS-CoV-2 infection after the first and second dose of IP. As noted above, this is a case-driven study: if the prespecified criteria for early efficacy are met at the time of either IA or overall efficacy at the primary analysis, a final study report describing the efficacy and safety of mRNA-1273 will be prepared based on the data available at that time. In the event that success criteria are met either at the time of the IA or when the total number of cases toward the primary endpoint have accrued, participants will continue to be followed in a blinded fashion until Month 25 to enable assessment of long-term safety and durability of VE. If the study concludes early, all participants will be requested to provide a final blood sample at the time of study conclusion.

Each participant will receive 2 doses of IP by 0.5 mL intramuscular (IM) injection, the first on Day 1 and the second on Day 29. An NP swab sample will be collected prior to the first and second dose of IP, for evaluation by RT-PCR. To preserve observer blinding, only delegated unblinded study personnel responsible for study vaccine preparation, administration and/or accountability will have knowledge of study treatment assignment (Section 6.2.8.1).

Participants will be given an electronic diary (eDiary) to report solicited ARs for 7 days after each dose of IP (Part A, Blinded Phase only) and to prompt an unscheduled clinic visit for clinical evaluation and NP swab sample if a participant experiences any symptoms of COVID-19. Participants will use the eDiary to report solicited ARs for 7 days after each dose of IP (Part A, Blinded Phase only) and weekly eDiary prompts (every 7 days) to elicit an unscheduled Illness Visit if the participant is experiencing COVID-19 symptoms. All participants will receive safety calls on Day 8, Day 15, Day 22, Day 36, and Day 43 that will serve both to monitor for unsolicited AEs and to monitor for symptoms of COVID-19.

Safety telephone calls and eDiary safety prompts will be performed in conjunction with surveillance for COVID-19 according to the SoEs (Table 16, Table 17, Table 18, and Table 19) and are intended to capture SAEs, MAAEs, AEs leading to withdrawal, concomitant medications associated with these events, receipt of non-study vaccinations, and pregnancy (Section 8.2.1). If an eDiary prompt results in identification of a relevant safety event, a follow-up safety call will be triggered.

Surveillance for COVID-19 will be performed through weekly contacts with the participant via a combination of telephone calls and completion of an eDiary starting at Day 1 through the end of the study, including both the Blinded and Open-Label Observational Phases (Section 8.1.2). Participants with symptoms of COVID-19 lasting at least 48 hours (except for fever and/or respiratory symptoms) will return to the clinic or will be visited at home by medically qualified site staff within 72 hours to collect an NP swab sample for RT-PCR testing for SARS-CoV-2 and other respiratory pathogens, or alternatively, if a clinic or home visit is not possible, will submit a saliva sample for SARS-CoV-2 RT-PCR testing (Section 8.1.1).

All study participants who experience COVID-19 symptoms and subsequently present for an Illness Visit (in-clinic or at home) will be given an instruction card listing symptoms and severity grading system along with a thermometer, an oxygen saturation monitor, and saliva collection tubes. The list of symptoms is presented in Section 8.1.2 and the severity scoring system is presented in Section 8.1.3. Study participants will be contacted by the investigator (or appropriately delegated study staff) daily with telemedicine visits through Day 14 or until symptoms have resolved, whichever is later. During the telemedicine visit (preferably done in the evening), the participant will be asked to verbally report the severity of each symptom, their highest body temperature and lowest oxygen saturation for that day, and the investigator will determine if medical attention is required due to worsening of COVID-19 symptoms (Table 19). Study participants will collect their own saliva sample on 3, 5, 7, 9, 14, and 21 days after the initial Illness Visit meeting criteria for COVID-19 (defined as the date of onset of symptoms and positive virologic test). Finally, a convalescent visit will be scheduled approximately 28 days after the initial Illness Visit. At this visit, a saliva sample will be collected and a blood sample will be drawn for immunologic assessment of SARS-CoV-2 infection.

At each dosing visit, participants will be instructed (Day 1) or reminded (Day 29) on how to document and report solicited ARs in the eDiary provided. Solicited ARs will be assessed for 7 days after each IP dose (Part A, Blinded Phase only) and unsolicited AEs will be assessed for 28 days after each IP dose (Part A, Blinded Phase and Part C, Open-Label Observational Booster Dose Phases only); SAEs, MAAEs, AEs leading to withdrawal, will be assessed throughout the study.

Participants may experience AEs that necessitate an unscheduled visit. There may also be situations in which the investigator asks a participant to report for an unscheduled visit following the report of an AE. Additional examinations may be conducted at these visits as necessary to ensure the safety and well-being of participants during the study. Electronic case report forms (eCRFs) should be completed for each unscheduled visit.

##### 4.1.2. Part B, the Open-label Observational Phase

The Part B, the Open-Label Observational Phase of the study is prompted by the authorization of a COVID-19 vaccine under EUA. Transitioning the study to Part B permits all ongoing study participants to be informed of the availability and eligibility criteria of any COVID-19 vaccine made available under an EUA and the option to offer all ongoing study participants who request unblinding, an opportunity to schedule a PDV to learn their original treatment assignment (placebo vs. mRNA-1273 vaccine).

Part B, the Open-Label Observational Phase of the study, also provides the opportunity for eligible study participants who previously received placebo, to actively request to receive 2 doses of mRNA-1273 vaccine (as long as investigational vaccine is available). All study participants will receive a notification letter and will be asked to schedule a PDV. Principal Investigators should consider current local public health guidance for administration of COVID-19 vaccines under EUA when determining the scheduling priority of participants. With regards to eligibility in Part B, the CDC-EUA guidance specifications will supersede eligibility criteria in the protocol.

All participants will proceed to Part B, the Open-Label Observational Phase of the study, starting with a PDV (Figure 2).

At the PDV, all participants will:

- Be encouraged to remain in the ongoing study,
- Be given the option to be unblinded as to their original group assignment (placebo vs. mRNA-1273 vaccine),
- Be counselled about the importance of continuing other public health measures to limit the spread of disease including social distancing, wearing a mask, and hand-washing,
- Sign a revised informed consent form (ICF),
- Provide an NP swab sample for RT-PCR for SARS-CoV-2 and a blood sample for serology prior to unblinding.

Figure 2 shows the Part B Study Flow schematic.

The following participants will continue with the original study SoEs presented in Table 16, Table 17, Table 18, and Table 19 (Figure 2):

- Participants who request to not be unblinded,
- Participants who request to be unblinded and received mRNA-1273, and
- Participants who request to be unblinded and received placebo and choose to remain on placebo.

Participants who are unblinded, received placebo, are eligible, and request to receive mRNA-1273 (as long as investigational vaccine is available) (Figure 2), will have the following clinic visits as shown in the Supplemental SoE (Table 21):

- Open-label Dose 1 (OL-D1): To occur at the PDV or at a scheduled subsequent visit.
- Open-label Dose 2 (OL-D29): To occur 28 days after Dose 1 on OL-D1.
- Open-label Clinic Visit (OL-D57): To occur 1-month after Dose 2 on OL-D29.
- And in addition, continue to comply with the Original Study SoEs (Table 16, Table 17, Table 18, and Table 19) as applicable.

- Participants who are unblinded, and received ONLY 1 dose of mRNA-1273 may be eligible to receive 1 more dose of mRNA-1273 if: They had a dosing error in Part A of the study that resulted in 1 dose of mRNA-1273 and 1 dose of placebo being administered.
- They did not have an AE that contraindicated the second dose of mRNA-1273 in Part A of the study.
- They did not withdraw consent in Part A of the study.
- They did not complete their second dose in Part A of the study for reasons other than the above.

If participants are eligible, and request to receive mRNA-1273 (Figure 2) will proceed to have the following clinic visits as shown in the Modified Supplemental SoEs (Table 22):

- Their second dose on OL-D1: To occur at the PDV or at a scheduled subsequent visit.
- Open-label Clinic Visit (OL-D29): To occur 1-month after Dose 2 on OL-D1.
- And in addition, continue to comply with the Original Study SoEs (Table 16, Table 17, Table 18, and Table 19) as applicable.

All participants remain on their original SoE to complete 24 months of follow-up after Dose 2; study participants who enter the Supplemental/Modified Supplemental SoE as described above do so in addition to their original SoE. Accordingly, all study participants will complete the full study follow-up to 24 months after the second dose of their original inoculation (mRNA-1273 or placebo).

At the point when the mRNA-1273 vaccine is no longer available for study use, any participant who schedules a PDV after this time point will be unblinded (with no option to stay blinded) but will not be offered to receive mRNA-1273 study vaccine. These participants will have all the same procedures performed outlined in Table 20 with the exception of being offered to receive mRNA-1273 vaccine. There will be no scientific metric or ethical advantage for participants to remain blinded when most of the study has already been unblinded. If the participant wants to receive EUA vaccine outside of the study after being unblinded, they will need to be withdrawn from the study.

After the BLA database lock (on 04 May 2021), any remaining participants who had not been unblinded were unblinded by the Moderna study team and received their unblinding treatment assignment (placebo vs. mRNA vaccine) via certified letter from the study site. These participants were not required to return to the site for an NP swab sample or blood draw.

##### 4.1.3. Part C, the Booster Dose Phase

The Part C, the Booster Dose (BD) Phase providing a BD for all eligible participants who chose to receive one, is prompted by the announcement by the President of the US that, as of 20 Sep 2021, certain individuals will be eligible to receive a BD in addition to the prior receipt of a primary vaccine regimen with mRNA-1273. The interim results of an ongoing Moderna Phase 2 study (mRNA-1273 P201), in which participants who 6-8 months prior received 2 doses of 50µg or 100µg of mRNA-1273 were administered a 50µg booster of mRNA-1273, demonstrated enhanced immune responses compared to pre-boost levels and met the noninferiority criteria stipulated in the FDA Guidance on EUA for Vaccines to Prevent COVID-19. Additionally, no new safety signals emerged upon administration of the booster dose in Study mRNA-1273-P201. Based on cumulative evidence, the benefit-risk profile of a booster dose of mRNA-1273 is favorable, particularly in light of increasing breakthrough disease with the emergence of the Delta variant. The details of eligibility for BDs provided by the US federal government have not been specified as of the time of writing this protocol amendment. Providing the option for a BD to all federally eligible participants currently enrolled in Part B is expected to promote retention of participants in the ongoing study and thereby defend the scientific integrity of the study for the planned 2-year duration of follow-up after the completion of the primary vaccination series regimen.

Each study participant will receive a notification letter and will be asked to schedule BD-1 visit at their study site (Table 23). Principal Investigators should consider current local public health guidance for administration of COVID-19 vaccines under EUA and marketing authorization (if any) when determining the scheduling priority of participants. At BD-1, each participant will:

- Be encouraged to remain in the ongoing study,
- Sign a revised ICF that includes both updated safety information relevant to the ongoing study and a BD, and the option to receive a BD
- Be given the option to receive a BD consisting of a 50 µg dose of mRNA-1273,
- Be counselled about the importance of continuing other public health measures to limit the spread of disease including social distancing, wearing a mask, and hand-washing.

After the BD-1 visit, participants who do not receive a BD will continue with the Part C SoEs with the exception of the BD-1a visit and blood draw and the D8, D15, and D22 safety calls (Table 23).

At the BD-1 visit, participants who request a BD, are eligible and have no contraindications to further dosing (see Section 7.1) will have the following study site visits and complete scheduled activities (subject to investigational vaccine availability) according to the Part C Supplemental SoE (Table 23):

- BD-1 visit: Participants will receive a single 50 µg dose of mRNA-1273
- BD-1a visit: Optional, and subject to blood sample kit availability, Day 4, 3 days after BD on Day 1
- BD-2 visit: Day 29, 28 days after the BD on Day 1
- BD-3 visit: Day 181, 180 days after the BD on Day 1

The investigator is responsible for conducting all assessments as specified in the Part C Supplemental SoE, according to the schedule. As this Supplemental SoE is intended to occur in addition to the original SoEs (Table 16, Table 17, Table 18, and Table 19) being followed by all participants in Part B, there is a possibility for study visits to overlap. If visits overlap according to respective visit windows, a single visit may be done with the combined study procedures completed once (refer to Table 4 footnotes for more detailed instructions and exceptions).

#### 4.2. Scientific Rationale for Study Design

The mRNA-1273 vaccine is being developed to prevent COVID-19, the disease resulting from SARS-CoV-2 infection. The present study has been designed to primarily evaluate the clinical efficacy and safety of mRNA-1273 to prevent COVID-19 for up to 2 years after the planned second dose of mRNA-1273. The immunogenicity endpoints and detection of SARS-CoV-2 infection are secondary objectives.

The design and focus of the study as designed for Part A were dependent on the then current COVID-19 pandemic, requiring identification of participant candidates at high risk of SARS-CoV-2 infection. This Phase 3 study was designed to be a randomized, stratified, observer-blind, placebo-controlled study to evaluate the efficacy, safety, and immunogenicity of mRNA-1273 SARS-CoV-2 vaccine compared to placebo in adults 18 years of age and older who had no known history of SARS-CoV-2 infection but whose locations or circumstances put them at appreciable risk of acquiring COVID-19 and/or SARS-CoV-2 infection.

Following authorization of a COVID-19 vaccine under EUA, the study design was amended to include a transition to Part B, the Open-Label Observational Phase (Section 2.2.2). The demonstration of efficacy against COVID-19 with satisfactory safety data, at a time when the COVID-19 pandemic remained critical, warranted allowing those study participants who actively requested it to know their original assignment (placebo or mRNA-1273 vaccine) on study. Transitioning the study to the Open-Label Observational Phase permitted all ongoing study participants (a) to be informed of the availability and eligibility criteria of any COVID-19 vaccine made available under an EUA and (b) to schedule a study visit to know their original group assignment (placebo vs. mRNA-1273 vaccine). Part B also provided the opportunity for eligible study participants who previously received placebo to receive 2 doses of mRNA-1273 vaccine under open-label conditions. Prompted by the announcement by the President of the US and the anticipated recommendations from the FDA, and CDC, this study design was further amended to include a transition to Part C, the BD Phase. The interim results of an ongoing Moderna Phase 2 study (mRNA-1273 P201), in which participants who 6-8 months prior received 2 doses of 50μg or 100μg of mRNA-1273 were administered a 50μg booster of mRNA-1273, demonstrated enhanced immune responses compared to pre-boost levels and met the noninferiority criteria stipulated in the FDA Guidance on EUA for Vaccines to Prevent COVID-19. Additionally, no new safety signals emerged upon administration of the booster dose in Study mRNA-1273-P201. Based on cumulative evidence, the benefit-risk profile of a booster dose of mRNA-1273 is favorable, particularly in light of increasing breakthrough disease with the emergence of the Delta variant. The details of eligibility for BDs provided by the US federal government have not been specified as of the time of writing this protocol amendment. This study design provides the option for a BD to all participants currently enrolled in Part B and as such is expected to promote retention of participants in the ongoing study and thereby defend the scientific integrity of the study for the planned 2-year duration of follow-up after the completion of the primary vaccination series.

#### 4.3. Choice of Dose and Control Product

The primary vaccination series originally selected for Part A of this study, 2 doses of 100 µg mRNA-1273 administered 28 days apart, was based on assessment of available safety and immunogenicity data from Phase 1 studies of mRNA-1647 (NCT03382405), the DMID study 20-0003 entitled "Phase I, Open-Label, Dose-Ranging Study of the Safety and Immunogenicity of 2019-nCoV Vaccine (mRNA-1273) in Healthy Adults" (NCT04283461) and Study 201 (Section 2.2.2). The primary vaccination series (at the 100 µg dose level) used in this study is being used in these studies, with evidence of safety and immunogenicity. At the time Part A was initiated, there was no licensed SARS-CoV-2 vaccine currently available to serve as a reference control; accordingly, 0.9% sodium chloride (normal saline) injection (USP) was used as a placebo control during Part A.

For Part C, provision of the option of a single BD for eligible participants during the Open-Label Observational Phase, the BD selected is 50 µg, based primarily on data from Study mRNA-1273-P201. In Part A of Study mRNA-1273-P201, the time course and magnitude of antibody (both bAb and nAb) responses to mRNA-1273 was similar between 100 µg and 50 µg dose levels at each postbaseline time point (Days 29, 43, 57, and 209), although the 100 µg dose group had numerically greater responses. In Part B of Study mRNA-1273-P201, administration of a 50 µg BD of mRNA-1273 6 months or more after the primary series, improved the immune responses to 1.7-fold the peak achieved after the primary vaccination series in the current mRNA-1273-P301 study, where efficacy of mRNA-1273 against COVID-19 was demonstrated.

#### 4.4. End of Study Definition

Participants are considered to have completed the study if they complete the final visit at Day 759 (Month 25), 24 months following their receipt of the original second dose of IP (where original refers to the IP received following randomization).

The study duration will be approximately 26 months for each participant. This includes a screening period of up to 1 month and a study period of 25 months that includes the first dose of IP on Day 1 and the second dose on Day 29 (Part A). The participant's final scheduled visit will be on Day 759 (Month 25), 24 months after the second dose of IP on Day 29 (Month 1).

The end of study will be the final participant's final scheduled visit at Day 759 (Month 25).

### 5. STUDY POPULATION

The study population, adults at risk of SARS-CoV-2 infection who have no known history of SARS-CoV-2 infection, is a subset of the planned target population. Additionally, potential study participants at increased risk of complications from COVID-19 will be included since it is hypothesized that these participants might derive the greatest benefit from a vaccine. Participants ≥ 65 years of age will be eligible for enrollment with or without underlying medical conditions further increasing their risk of severe COVID-19.

Given the disproportionate disease burden of COVID-19 in racial and ethnic minorities, the study will also aim to enroll a representative sample of participants from these minority population and adjust site selection and enrollment accordingly, per FDA Draft Guidance "Enhancing the Diversity of Clinical Trial Populations - Eligibility Criteria, Enrollment Practices, and Trial Designs" (FDA 2019). Study sites may be selected on the basis of SARS-COV-2 infection risk of the local population as well. Approximately 30,000 participants will be enrolled.

There will be no prospective approval of protocol deviations to recruitment and enrollment criteria (also known as protocol waivers or exemptions).

#### 5.1. Inclusion and Exclusion Criteria (Part A, Part B, and Part C)

##### 5.1.1. Inclusion Criteria

Participants are eligible to be included in the study, or further dosing (Part B and Part C), only if all the following criteria apply:

1. (Part A only) Adults, ≥ 18 years of age at time of consent, who are at high risk of SARS-CoV-2 infection, defined as adults whose locations or circumstances put them at appreciable risk of exposure to SARS-CoV-2 and COVID-19
2. Understands and agrees to comply with the study procedures and provides written informed consent.
3. Able to comply with study procedures based on the assessment of the investigator.
4. Female participants of nonchildbearing potential may be enrolled in the study. Nonchildbearing potential is defined as surgically sterile (history of bilateral tubal ligation, bilateral oophorectomy, hysterectomy) or postmenopausal (defined as amenorrhea for ≥ 12 consecutive months prior to Screening without an alternative medical cause). A follicle-stimulating hormone (FSH) level may be measured at the discretion of the investigator to confirm postmenopausal status (see additional information in Appendix 11.3).

5. Female participants of childbearing potential may be enrolled in the study if the participant fulfills all the following criteria:
   - Has a negative pregnancy test at Screening and on the day of the first dose (Day 1, OL-D1 and BD-1).
   - Has practiced adequate contraception or has abstained from all activities that could result in pregnancy for at least 28 days prior to the first dose (Day 1, OL-D1 and BD-1).
   - Has agreed to continue adequate contraception through 3 months following the last dose (Day 29, OL-D29 and BD-1).
   - Is not currently breastfeeding.

   Adequate female contraception is defined as consistent and correct use of an FDA approved contraceptive method in accordance with the product label. For example:
   - Barrier method (such as condoms, diaphragm, or cervical cap) used in conjunction with spermicide
   - Intrauterine device
   - Prescription hormonal contraceptive taken or administered via oral (pill), transdermal (patch), subdermal, or IM route
   - Sterilization of a female participant’s monogamous male partner prior to entry into the study

   Note: periodic abstinence (eg, calendar, ovulation, symptothermal, post-ovulation methods) and withdrawal are not acceptable methods of contraception.
6. Healthy adults or adults with pre-existing medical conditions who are in stable condition. A stable medical condition is defined as disease not requiring significant change in therapy or hospitalization for worsening disease during the 3 months before enrollment.
7. (Part C Only) Is currently enrolled in Part B of the current study (mRNA-1273-P301)
8. (Part C Only) Has received at least 1 dose of mRNA-1273 in the current study (mRNA-1273-P301)

##### 5.1.2. Exclusion Criteria

Participants are excluded from the study or further dosing (Part B and Part C) if any of the following criteria apply:

1. Is acutely ill or febrile 72 hours prior to or at Screening or dosing (Part B and Part C). Fever is defined as a body temperature ≥ 38.0°C/100.4°F. Participants meeting this criterion may be rescheduled within the relevant window periods. Afebrile participants with minor illnesses can be enrolled/dosed at the discretion of the investigator.
2. Is pregnant or breastfeeding.
3. (Part A Only) Known history of SARS-CoV-2 infection.
4. (Part A only) Prior administration of an investigational coronavirus (SARS-CoV, MERS-CoV) vaccine or current/planned simultaneous participation in another interventional study to prevent or treat COVID-19
5. (Part A Only) Demonstrated inability to comply with the study procedures.
6. (Part A Only) An immediate family member or household member of this study's personnel.
7. Known or suspected allergy or history of anaphylaxis, urticaria, or other significant AR to the vaccine or its excipients.
8. Bleeding disorder considered a contraindication to intramuscular injection or phlebotomy.
9. Has received or plans to receive a non-study vaccine within 28 days prior to or after any dose of IP (except for seasonal influenza vaccine which is not permitted within 14 days before or after any dose of IP, see Section 6.4.3).
10. Has participated in an interventional clinical study within 28 days prior to the day of enrollment.
11. (Part A only) Immunosuppressive or immunodeficient state, asplenia, recurrent severe infections (HIV-positive participants with CD4 count ≥350 cells/$mm^3$ and an undetectable HIV viral load within the past year [low level variations from 50-500 viral copies which do not lead to changes in antiretroviral therapy [ART] are permitted]).
12. Has received systemic immunosuppressants or immune-modifying drugs for >14 days in total within 6 months prior to Screening (for corticosteroids ≥ 20 mg/day of prednisone equivalent).
13. (Part A only) Has received systemic immunoglobulins or blood products within 3 months prior to the day of Screening.

14. (Part A only) Has donated ≥ 450 mL of blood products within 28 days prior to Screening.

#### 5.2. Participant Restrictions

Participants must not eat or drink anything hot or cold within 10 minutes before oral temperature is taken.

#### 5.3. Screen Failures (Part A: Blinded Phase Only)

Screen failures are defined as participants who consent to participate in the clinical study but are not subsequently randomly assigned to treatment. A minimum set of screen failure information is required to ensure transparent reporting of screen failures to meet the Consolidated Standards of Reporting Trials (CONSORT) publishing requirements and to respond to queries from regulatory authorities. Minimum information includes date of informed consent, demography, screen failure details, eligibility criteria, and information on any SAE that may have occurred from the time informed consent was obtained to the time of withdrawal.

Participants meeting the exclusion criterion #1, acutely ill or febrile prior to or at the Screening Visit (exclusion criterion #1, Section 5.1.2), may be rescheduled within the relevant window periods and will retain their initially assigned participant number.

### 6. STUDY TREATMENT

#### 6.1. Investigational Product

The term “investigational product” refers to both mRNA-1273 vaccine (blinded and open-label) and placebo administered in this study.

The mRNA-1273 IP is an LNP dispersion of an mRNA encoding the prefusion stabilized S protein of SARS-CoV-2 formulated in LNPs composed of 4 lipids (1 proprietary and 3 commercially available): the proprietary ionizable lipid SM-102; cholesterol; 1,2-distearoyl-sn-glycero-3-phosphocholine (DSPC); and 1-monomethoxypolyethyleneglycol-2,3-dimyristylglycerol with polyethylene glycol of average molecular weight 2000 (PEG2000-DMG). The mRNA-1273 vaccine is provided as a sterile liquid for injection and is a white to off-white dispersion in appearance, at a concentration of 0.2 mg/mL in 20 mM Tris buffer containing 87 mg/mL sucrose and 10.7 mM sodium acetate at pH 7.5.

The placebo is 0.9% sodium chloride (normal saline) injection, which meets the criteria of the United States Pharmacopeia (USP).

#### 6.2. Dosing and Management of Investigational Product

##### 6.2.1. Method of Randomly Assigning Participants to Treatment Groups (Part A, Blinded Phase Only)

Approximately 30,000 participants will be randomly assigned in 1:1 ratio to receive either mRNA-1273 100 µg or placebo. The randomization will be in a blinded manner using a centralized Interactive Response Technology (IRT), in accordance with pre-generated randomization schedules. Only the unblinded personnel (Section 6.2.8.1) will have controlled access to which arm the participant is randomly assigned.

Dose group assignment in is summarized in Table 2.

**Table 2: Summary of Treatment Groups**

| Treatment Groups | Investigational Product | Age (years) | Estimated Total Participants |
|---|---|---|---|
| mRNA-1273 | mRNA-1273 100 µg | ≥ 18 | 15,000 |
| Placebo | Placebo | ≥ 18 | 15,000 |
| **Total** | | | **30,000** |

###### 6.2.1.1. Stratification

Randomization in Part A, Blinded Phase of the study will be stratified based on age and, if they are < 65 years of age, based on the presence or absence of risk factors for severe illness from COVID-19 based on CDC recommendation as of March 2020 (CDC 2020b). There will be 3 strata for randomization: ≥ 65 years, < 65 years and categorized to be at increased risk ("at risk") for the complications of COVID-19, and < 65 years "not at risk." Risk will be defined based on the study participants' relevant past and current medical history. At least 25% of enrolled participants, up to 50%, will be either ≥ 65 years of age or < 65 years of age and at risk at Screening.

Participants who are < 65 years old will be categorized as at risk for severe COVID-19 illness if they have at least 1 of the following risk factors at Screening:

- Chronic lung disease (eg, emphysema and chronic bronchitis), idiopathic pulmonary fibrosis and cystic fibrosis) or moderate to severe asthma
- Significant cardiac disease (eg, heart failure, coronary artery disease, congenital heart disease, cardiomyopathies, and pulmonary hypertension)
- Severe obesity (body mass index ≥ 40 kg/m$^2$)
- Diabetes (Type 1, Type 2 or gestational)
- Liver disease
- HIV infection

##### 6.2.2. Administration of Investigational Product

In Part A and Part B as applicable, investigational product will be administered as an IM injection into the deltoid muscle on a 2-dose injection schedule on Day 1 and Day 29. For injections administered for Part A and Part B, each injection will have a volume of 0.5 mL and contain mRNA-1273 100 µg or saline placebo. For Part C, each injection will have a volume of 0.25 mL and contain mRNA-1273 50 µg. Preferably, vaccine should be administered into the nondominant arm. The second dose of IP should be administered in the same arm as the first dose (Part A and Part B, as applicable).

The IP will be prepared for injection as a single 0.5 mL (Part A and Part B) or 0.25 mL (Part C) dose for each participant per protocol, as detailed in the Pharmacy Manual. Unblinded personnel who will not participate in any other aspect of the study during Part A, the Blinded Phase, will perform IP accountability, dose preparation, and IP administration. The investigator will designate unblinded medically qualified personnel (not involved in assessments of study endpoints) to administer the IP according to the procedures stipulated in this study protocol (Part A, the Blinded Phase), and Pharmacy Manual. Study-specific training will be provided. Study site personnel who were blinded during the Blinded Phase, will be unblinded at the participant level at the PDV.

At each visit when IP is administered, participants will be monitored for a minimum of 30 minutes after administration. Assessments will include vital sign measurements and monitoring for local or systemic reactions (Table 16, Table 21, Table 22 and Table 23).

Eligibility for a subsequent dose of IP is determined by following the criteria outlined in Section 7.

The study site will be appropriately staffed with individuals with basic cardiopulmonary resuscitation training/certification. Either on-site resuscitation equipment and personnel or appropriate protocols for the rapid transport of participant to a resuscitation area/facility are required.

##### 6.2.3. Delivery and Receipt

The Sponsor or designee is responsible for the following:

- Supplying the IP
- Confirming the appropriate labeling of mRNA-1273 IP, so that it complies with the legal requirements of the US

The investigator is responsible for acknowledging the receipt of the IP by a designated staff member at the site, including the following:

- Confirming that the IP was received in good condition
- Confirmation that the temperature during shipment from the Sponsor to the investigator's designated storage location was appropriate
- Confirming whether the Sponsor has authorized the IP for use
- Ensuring the appropriate dose level of mRNA-1273 is properly prepared using aseptic technique

Further description of the IP and instructions for the receipt, storage, preparation, administration, accountability, and destruction of the IP are described in the mRNA-1273-P201 Pharmacy Manual.

##### 6.2.4. Packaging and Labeling

The Sponsor will provide the investigator and study site with adequate quantities of mRNA-1273. The sterile vaccine product is packaged in a 10R glass vial with a 5.0-mL or 6.3-mL fill volume (Part A and Part B) or a 10R glass vial with an 8.0 mL fill volume (Part C). mRNA-1273 vaccine will have all required labeling per regulations and will be supplied to the pharmacy in an unblinded manner. Each vial will be individually labeled for future participant identification purposes.

mRNA-1273 will be packaged and labeled in accordance with the standard operating procedures of the Sponsor or of its designee, Code of Federal Regulations Title 21 (CFR), Good Manufacturing Practice (GMP) guidelines, International Council for Harmonisation (ICH) GCP guidelines, guidelines for Quality System Regulations, and applicable regulations.

The Sponsor or Sponsor's designee will supply the 0.9% sodium chloride injection for use as both a placebo and a diluent to mRNA-1273. The 0.9% sodium chloride bears a commercial label and does not contain study-specific identification (Part A only).

##### 6.2.5. Storage

The vials of mRNA-1273 with 5.0 ml fill volume must be stored at 2°C to 8°C in a secure area with limited access (unblinded personnel only) and protected from moisture and light until it is prepared for administration (Section 6.2.2). The vials of mRNA-1273 with 6.3 mL or 8.0 mL fill volume must be stored at -20°C in a secure area with limited access and protected from moisture and light until it is prepared for administration. The vials of mRNA-1273 with 6.3 mL fill volume can be stored at 2°C to 8°C for up to 30 days, but site personnel must account for this manually as it will not be managed in the IRT system. The freezer or refrigerator should have automated temperature recording and a 24-hour alert system in place that allows for rapid response in case of freezer or refrigerator malfunction. There must be an available backup freezer or refrigerator. The freezers or refrigerators must be connected to a backup generator. In addition, vaccine accountability study staff (eg, the unblinded personnel) are required to keep a temperature log to establish a record of compliance with these storage conditions. The site is responsible for reporting any mRNA-1273 that was not temperature controlled during shipment or during storage to the unblinded site monitor. Such mRNA-1273 will be retained for inspection by the unblinded monitor and disposed of according to approved methods.

The 0.9% sodium chloride injection (USP) should be stored at 20ºC to 25ºC (68ºF to 77ºF) in a restricted access area (applicable to Part A only).

##### 6.2.6. Investigational Product Accountability

It is the investigator's responsibility that the unblinded personnel maintain accurate records in an IP accountability log of receipt of all IP, inventory at the site, dispensing of mRNA-1273 and placebo, IP injections, and return to the Sponsor or alternative disposition of used/unused products.

An unblinded site monitor will review the inventory and accountability log during site visits and at the completion of the study. Additional details are found in the mRNA-1273-P301 Pharmacy Manual.

##### 6.2.7. Handling and Disposal

An unblinded site monitor will reconcile the IP during the conduct and at the end of the study for compliance. Once fully reconciled at the site at the end of the study, the IP can be destroyed at the investigational site or at a Sponsor-selected third party, as appropriate.

Investigational product may be destroyed at the study site only if permitted by local regulations and authorized by the Sponsor. A Certificate of Destruction must be completed and sent to the Sponsor or designee.

##### 6.2.8. Unblinding

###### 6.2.8.1. Planned Unblinding

See Section 4.1.2 regarding the PDV.

- Part A, the Blinded Phase of this study is observer-blind. The investigator, study staff, study participants, site monitors, and Sponsor personnel (or its designees) will be blinded to the IP administered until study end, with the following exceptions:
- Unblinded personnel (of limited number) will be assigned to vaccine accountability procedures and will prepare IP for all participants. These personnel will have no study functions other than study vaccine management, documentation, accountability, preparation, and administration. They will not be involved in participant evaluations and will not reveal the identity of IP to either the participant or the blinded study site personnel involved in the conduct of the study unless this information is necessary in the case of an emergency.
- Unblinded medically qualified study site personnel will administer the IP. They will not be involved in assessments of any study endpoints.
- The dosing assignment will be concealed by having the unblinded personnel prepare the IP in a secure location that is not accessible or visible to other study staff. An opaque sleeve over the syringe used for injection will maintain the blind at the time of injection, as the doses containing mRNA-1273 will look different than placebo. Only delegated unblinded site staff will conduct the injection procedure. Once the injection is completed, only the blinded study staff will perform further assessments and interact with the participants. Access to the randomization code will be strictly controlled at the pharmacy.
- Unblinded site monitors, not involved in other aspects of monitoring, will be assigned as the IP accountability monitors. They will have responsibilities to ensure that sites are following all proper IP accountability, preparation, and administration procedures.
- An unblinded statistical and programming team will perform the pre-planned IAs (Section 9.6).
- An independent DSMB will review the interim data to safeguard the interests of clinical study participants and to help ensure the integrity of the study. The DSMB will review

unblinded statistical outputs and IA results, provided by the independent unblinded statistician, and make recommendations to the Sponsor (Section 8.4.2).

- At the initiation of Part B, the Open-Label Observational Phase of this study (Section 4.1.3), study site personnel who were blinded during the Blinded Phase will be unblinded at the participant level at the PDV.

If prespecified criteria for early efficacy are met by an IA or if the primary efficacy analysis is completed based on accrual of prespecified COVID-19 cases, pre-identified Sponsor and CRO team members responsible for the analysis and reporting will be unblinded to treatment assignments in order to prepare a final study report. In order to maintain an observer-blind design, investigators, site staff, participants, and Sponsor and CRO staff with oversight of study conduct will remain blinded to treatment allocation for the study duration. All study participants will be followed for efficacy and safety endpoints through the remainder of planned study period and results will be summarized in an end of study report (Sections 4.1 and 9.1).

###### 6.2.8.2. Unplanned Unblinding

A participant or participants may be unblinded in the event of an SAE or other severe event, or if there is a medical emergency requiring the identity of the product to be known to properly treat a participant. If a participant becomes seriously ill or pregnant during the study, the study investigator may request that the blind will be broken if knowledge of the administered vaccine will affect that participant's dosing options. In this situation or in the event of a medical emergency requiring identification of the IP administered to an individual participant, the investigator will make every attempt to contact the Sponsor medical lead to explain the need for opening the code within 24 hours of opening the code. The investigator will be responsible for documenting the time, date, reason for the code break, and the names of the personnel involved.

In addition to the situations described above, where the blind may be broken, the data will also be unblinded to a statistical team at specified time points for IAs as outlined in Section 9.6.

In December 2020, COVID-19 vaccines have started to become available under EUA as an alternative option to some participants based on evolving recommended populations by the CDC and local supply chain distribution. While all participants should be encouraged to stay blinded in the study for as long as possible, their participation should not otherwise deny them the opportunity to receive a COVID-19 vaccine under EUA. Clinical investigators can exercise discretion as to whether individual participants should be unblinded upon request to allow them to make an informed decision regarding receipt of a COVID-19 vaccine outside of this study. Investigator judgment should consider a participant's risk status under CDC recommendations, any current local public health guidance, and their access to imminently receive a COVID-19 vaccine under an EUA.

If a decision is made to unblind a participant to support an informed decision to receive a COVID-19 vaccine under an EUA outside of this study, investigators are asked to take the following steps:

- Obtain a final assessment of safety from the participant to collect and resolve any outstanding safety experience. Verbal interview is sufficient.
- Unblind the participant in the IRT and inform the participant of their treatment assignment.
- Inform the Sponsor Medical Lead within 24 hours to confirm rationale for unblinding.

Participants who are confirmed to have received placebo upon unblinding and intend to receive a COVID-19 vaccine under an EUA outside of this study will be withdrawn from the study at the point of unblinding (Section 7.3.1).

Note: Participants who requested unblinding because of a stated imminent access to a COVID-19 vaccine under an EUA outside of this study, were withdrawn from study, and then were unable to in fact receive a COVID-19 vaccine, will be permitted to re-enter this study upon re-consent to receive the mRNA-1273 vaccine, once they are eligible (as long as investigational vaccine is available).

#### 6.3. Study Treatment Compliance

All doses of IP will be administered at the study site under direct observation of unblinded medically qualified study personnel and appropriately recorded (date and time) in the eCRF. Unblinded personnel will confirm that the participant has received the entire dose of vaccine. If a participant does not receive vaccine or does not receive all of the planned doses, the reason for the missed dose will be recorded. Data will be reconciled with site accountability records to assess compliance.

Participants who miss the second dose of IP due to noncompliance with the visit schedule and not due to a safety pause will still be required to follow the original visit and testing schedule as described in the protocol. Unless consent is withdrawn, a participant who withdraws or is withheld from receiving the second dose of study vaccine will remain in the study and complete all efficacy, safety and immunogenicity assessments required through the participant's scheduled end of study.

The study site is responsible for ensuring that participants comply with the study windows allowed. If a participant misses a visit, every effort should be made to contact the participant and complete a visit within the defined visit window (SoE Tables, Section 11.1). If a participant does not complete a visit within the time window, that visit will be classified as a missed visit (with the exception of Dose 2 visits in Part A and Part B, as applicable) and the participant will continue with subsequent scheduled study visits. All safety requirements of the missed visit will be captured and included in the subsequent visit (eg, clinical laboratory testing, eDiary review for reactogenicity, immunologic testing, as applicable).

#### 6.4. Prior and Concomitant Therapy

##### 6.4.1. Prior Medications and Therapies

Information about prior medications (including any prescription or over-the-counter medications, vaccines, or blood products) taken by the participant within the 28 days before providing informed consent (or as designated in the inclusion/exclusion requirements) will be recorded in the participant's eCRF.

##### 6.4.2. Concomitant Medications and Therapies

Study site staff must question the participant regarding any medications taken and vaccinations received by the participant and record the following information in the eCRF:

- All non-study vaccinations administered within the period starting 28 days before the first dose of IP.
- Seasonal influenza vaccine administered for the current influenza season (typically October through April in the Northern Hemisphere).
- Part A only: All concomitant medications and non-study vaccinations taken through 28 days after each dose of IP. Antipyretics and analgesics taken prophylactically (ie, taken in the absence of any symptoms in anticipation of an injection reaction) will be recorded as such.
- Any concomitant medications used to prevent or treat COVID-19.
- Any concomitant medications relevant to or for the treatment of an SAE or a MAAE.
- Part A only: Participant will be asked in the eDiary if they have taken any antipyretic or analgesic to treat or prevent fever or pain within 7 days after each IP injection, including the day of dosing. Reported antipyretic or analgesic medications should be recorded in the source document by the site staff during the post-injection study visits or via other participant interactions (eg, phone calls).

##### 6.4.3. Concomitant Medications and Vaccines that May Lead to the Elimination of a Participant from Per-protocol Analyses

The use of the following concomitant medications and/or vaccines will not require withdrawal of the participant from the study (with the exception of planned or actual receipt of a non-study COVID-19 vaccine either under EUA or licensed, Section 7.3.1) but may determine a participant's eligibility to receive a second dose or evaluability in the per-protocol (PP) analysis (Analysis Sets are described in Section 9.4):

- Any investigational or nonregistered product (drug or vaccine) other than the study vaccine used during the study period.
- A non-study vaccine administered during the period from 28 days before through 28 days after each dose of IP or any seasonal influenza vaccine that was administered within 14 days before or after any dose of IP.
- Immunoglobulins and/or any blood products administered during the study period (except for treatment of COVID-19).
- Medications that suppress the immune system (except for treatment of COVID-19).

If a participant takes a prohibited drug therapy, the investigator and the contract research organization's (CRO's) medical monitor will make a joint decision about continuing or withholding further dosing from the participant based on the time the medication was administered, the drug's pharmacology and pharmacokinetics, and whether use of the medication will compromise the participant's safety or the interpretation of data. It is the investigator's responsibility to ensure that details regarding the concomitant medications are adequately recorded in the eCRF.

All medication and interventions necessary for the appropriate care for the study participant, particularly to treat COVID-19, should be administered and appropriately documented along with the AE.

### 7. DELAYING OR DISCONTINUING STUDY TREATMENT AND PARTICIPANT WITHDRAWAL FROM THE STUDY

#### 7.1. Criteria for Delay of Study Treatment

Body temperature must be measured at the Day 1, Day 29, OL-D1, OL-D29, and BD-1 visits prior to any study treatment administration. The following events constitute criteria for delay of study treatment, and if either of these events occur at the time scheduled for dosing, the participant may be injected at a later date within the time window specified in Table 16, Table 21, Table 22 and Table 23, or the participant may be discontinued from dosing at the discretion of the investigator (Section 7.2):

- Acute moderate or severe infection with or without fever at the time of dosing
- Fever, defined as body temperature ≥ 38.0°C (100.4°F) at the time of dosing

Participants with a minor illness without fever, as assessed by the investigator, can be administered IP. Participants with a fever of 38.0°C (100.4°F) or higher will be contacted within the time window acceptable for participation and reevaluated for eligibility. If the investigator determines that the participant's health on the day of administration temporarily precludes dosing with IP, the visit should be rescheduled within the allowed interval for that visit.

#### 7.2. Discontinuation of Study Treatment

Every reasonable attempt will be made to follow up with participants for safety throughout the entire study period, even if further dosing is discontinued or the participant misses one or more visits. Unless consent is withdrawn, a participant who withdraws or is withheld from receiving the second dose of study vaccine will remain in the study and complete all scheduled visits and assessments (Section 11.1).

The investigator, in consultation with the Sponsor's medical monitor, may withhold a participant from further dosing if the participant experiences any of the following:

- Becomes pregnant (Section 8.3.6)
- Develops, during the course of the study, symptoms or conditions listed in the exclusion criteria
- Experiences an AE (other than reactogenicity) after dosing that is considered by the investigator to be related to IP (Section 8.3.5) and is of Grade 3 (severe) or greater intensity (Section 8.3.9)
- Experiences an AE or SAE that, in the judgment of the investigator, requires study IP withdrawal due to its nature, severity, or required treatment, regardless of the causal relationship to vaccine

- Experiences a clinically abnormal vital sign measurement or finding on physical examination, or general condition that, in the judgment of the investigator, requires IP withdrawal

The reason(s) for discontinuation from further dosing will be recorded in the eCRF.

Prior to receiving a second dose of IP, participants will be reassessed to ensure that they continue to meet eligibility requirements as outlined below.

The following events in a participant constitute absolute contraindications to any further dosing of the IP to that participant. If any of these events occur during the study, the participant must not receive additional doses of vaccine but will be encouraged to continue study participation for safety through 24 months following last dose.

- Part A only: Diagnosed COVID-19 by detection of SARS-CoV-2 in a Day 1 NP swab sample or COVID-19 diagnosed prior to Day 29. If COVID-19 is suspected on or prior to Day 29, further administration of IP must be withheld until COVID-19 test results are available.
- Anaphylaxis or systemic hypersensitivity reaction following the administration of vaccine.
- Any SAE judged by investigator or Sponsor to be related to study vaccine.
- Any clinically significant medical condition that, in the opinion of the investigator, poses an additional risk to the participant if he/she continues to participate in the study.

The following events constitute contraindications to administration of study vaccine at certain points in time, and if any of these events occur at the time scheduled for dosing, the participant may be injected at a later date, within the time window specified in the SoE (Table 16), or the participant may be withdrawn from dosing at the discretion of the investigator (Section 7.3):

- Acute moderate or severe infection with or without fever at the time of dosing
- Fever, defined as body temperature ≥ 38.0°C (100.4°F) at the time of dosing
- Part B only: Diagnosed COVID-19 by detection of SARS-CoV-2 by RT-PCR and accompanying symptoms. Participant must be asymptomatic at the time of dosing although may still be RT-PCR positive, with the exception of loss of sense of smell or taste and fatigue, if by the investigator's judgement these are deemed to be mild chronic sequelae.

Participants with a minor illness without fever, as assessed by the investigator, can be administered IP. Participants with a fever of 38.0°C (100.4°F) or higher will be contacted within the time window acceptable for participation and reevaluated for eligibility.

#### 7.3. Participant Withdrawal from the Study

##### 7.3.1. Participant Withdrawal

Participants who withdraw from the study will not be replaced. A “withdrawal” from the study refers to a situation wherein a participant does not return for the final visit planned in the protocol. The statistical management of participant withdrawals is discussed in Section 9.

Participants can withdraw consent and withdraw from the study at any time, for any reason, without prejudice to further treatment the participant may need to receive. The investigator will request that the study participant complete all study procedures pending at the time of withdrawal.

If participant desires to withdraw from the study because of an AE, the investigator will try to obtain agreement to follow up with the participant until the event is considered resolved or stable and will then complete the end of study eCRF.

Information related to the withdrawal will be documented in the eCRF. The investigator will document whether the decision to withdraw a participant from the study was made by the participant or by the investigator, as well as which of the following possible reasons was responsible for withdrawal:

- AE (specify)
- SAE (specify)
- Death
- Lost to follow-up
- Physician decision (specify)
- Pregnancy
- Protocol deviation
- Study terminated by Sponsor
- Withdrawal of consent by participant (specify); this includes participants who, at the PDV, withdraw consent from continuing in the study.
- Other (specify)

Participants who are withdrawn from the study because of AEs (including SAEs) must be clearly distinguished from participants who are withdrawn for other reasons. Investigators will follow up with participants who are withdrawn from the study as result of an SAE or AE until resolution of the event.

Participants who are confirmed to have received placebo upon unblinding and intend to receive a COVID-19 vaccine either licensed or under an EUA outside of this study will be withdrawn from the study, as will participants who have been confirmed to have received an outside COVID-19 vaccine whilst on study.

Participants who are confirmed to have received or plan to receive a COVID-19 booster vaccine, either licensed or under EUA, outside of this study will be withdrawn from the study.

A participant withdrawing from the study may request destruction of any samples taken and not tested, and the investigator must document this in the site study records.

If the participant withdraws consent for disclosure of future information, the Sponsor may retain and continue to use any data collected before such a withdrawal of consent (see Section 11.2.10).

The Sponsor will continue to retain and use all research results that have already been collected for the study evaluation, unless the participant has requested destruction of these samples. All biological samples that have already been collected may be retained and analyzed at a later date (or as permitted by local regulations).

#### 7.4. Lost to Follow-up

Participants will be considered lost to follow-up (LTFU) if they repeatedly fail to return for scheduled visits without stating an intention to withdraw consent and they cannot be contacted by the study site. The following actions must be taken if a participant fails to return to the clinic for a required study visit:

- The site must attempt to contact the participant and reschedule the missed visit as soon as possible, counsel the participant on the importance of maintaining the assigned visit schedule and ascertain whether the participant wishes to and/or should continue in the study.
- Before a participant is deemed lost to follow-up, the investigator or designee must make every effort to regain contact with the participant (where possible, 3 telephone calls and, if necessary, a certified letter to the participant's last known mailing address or local equivalent methods). These contact attempts (eg, dates of telephone calls and registered letters) should be documented in the participant's medical record. A participant should not be considered LTFU until these efforts have been made.
- Should the participant continue to be unreachable, he/she will be considered to have withdrawn from the study.

### 8. STUDY ASSESSMENTS AND PROCEDURES

Before performing any study procedures, all potential participants will sign an ICF (as detailed in Section 11.2.6). Participants will undergo study procedures at the time points specified in the SoEs (Section 11.1).

A participant also can be seen for an unscheduled visit at any time during the study. An unscheduled visit may be prompted by reactogenicity issues, Illness Visit criteria for COVID-19, or new or ongoing AEs. The site also has the discretion to make reminder phone calls or send text messages to inform the participant about visits, review eDiary requirements, or follow-up on ongoing or outstanding issues.

In accordance with "FDA Guidance on Conduct of Clinical Trials of Medical Products during the COVID-19 Pandemic" (DHHS 2020), investigators may convert study site visits to telemedicine visits with the approval of the Sponsor. Such action should be taken to protect the safety and well-being of study participants and study site staff or to comply with state or municipal mandates.

General considerations for study assessments and procedures include the following:

- Protocol waivers or exemptions are not allowed. The study procedures and their timing must be followed as presented in Section 11.1. Adherence to the study design requirements is essential and required for study conduct.
- Immediate safety concerns should be discussed with the Sponsor immediately upon occurrence or awareness to determine if the participant should continue study treatment or participation in the study.
- All screening evaluations must be completed and reviewed to confirm that potential participants meet all eligibility criteria. The investigator will maintain a screening log to record details of all participants screened and to confirm eligibility or record reasons for screening failure, as applicable.
- Procedures conducted as part of the participant's routine clinical management (eg, blood count) and obtained before signing of the ICF may be utilized for screening or baseline purposes provided the procedures met the protocol-specified criteria and were performed within the time frame defined in the SoE.

#### 8.1. Efficacy and Immunogenicity Assessments and Procedures

##### 8.1.1. Efficacy Assessments Related to COVID-19 and SARS-CoV-2 Infection

Each study participant will have an NP swab sample collected for SARS-CoV-2 testing by RT-PCR on Day 1 and Day 29, prior to receiving a dose of the IP as specified in the SoE (Section 11.1).

**COVID-19:**

To be considered as a case of COVID-19 for the evaluation of the Primary Efficacy Endpoint, the following case definition must be met:

- The participant must have experienced at least TWO of the following systemic symptoms: Fever (≥ 38ºC), chills, myalgia, headache, sore throat, new olfactory and taste disorder(s), OR
- The participant must have experienced at least ONE of the following respiratory signs/symptoms: cough, shortness of breath or difficulty breathing, OR clinical or radiographical evidence of pneumonia; AND
- The participant must have at least 1 NP swab or saliva sample (or respiratory sample, if hospitalized) positive for SARS-CoV-2 by RT-PCR.

**Severe COVID-19:**

To be considered severe COVID-19, the following criteria must be met:

- Confirmed COVID-19 as per the Primary Efficacy Endpoint case definition, plus any of the following:
    - Clinical signs indicative of severe systemic illness, Respiratory Rate ≥ 30 per minute, Heart Rate ≥ 125 beats per minute, $SpO_2$ ≤ 93% on room air at sea level or $PaO_2/FIO_2$ < 300 mm Hg, OR
    - Respiratory failure or Acute Respiratory Distress Syndrome (ARDS), (defined as needing high-flow oxygen, non-invasive or mechanical ventilation, or ECMO), evidence of shock (systolic blood pressure < 90 mmHg, diastolic BP < 60 mmHg or requiring vasopressors), OR
    - Significant acute renal, hepatic or neurologic dysfunction, OR
    - Admission to an intensive care unit or death.

The secondary case definition of COVID-19 is defined as the following systemic symptoms: fever (temperature ≥ 38ºC), or chills, cough, shortness of breath or difficulty breathing, fatigue, muscle aches, or body aches, headache, new loss of taste or smell, sore throat, nasal congestion or rhinorrhea, nausea, or vomiting or diarrhea AND a positive NP swab or saliva sample (or respiratory sample, if hospitalized) for SARS-CoV-2 by RT-PCR.

Death attributed to COVID-19 is defined as any participant who dies during the study with a cause directly attributed to a complication of COVID-19.

**SARS-CoV-2 Infection:**

- SARS-CoV-2 infection is defined by seroconversion due to infection measured by bAb against SARS-CoV-2 nucleocapsid. Seroconversion is defined as follows for participants who are seronegative at Baseline:
    - Binding antibody levels against SARS-CoV-2 nucleocapsid either below the limit of detection (LOD) or lower limit of quantification (LLOQ) at Study Day 1 that increase to above or equal to LOD or LLOQ starting at Study Day 57 or later.

##### 8.1.2. Surveillance for COVID-19 Symptoms

Surveillance for COVID-19 symptoms will be conducted by a combination of safety phone calls and eDiary completion as presented in the SoEs in Section 11.1, in addition to study site visits (Figure 3). If there is no response to an eDiary prompt for 2 days, the site staff will contact the study participant by phone.

According to the CDC as of 10 June 2020 (CDC 2020c), patients with COVID-19 have reported a wide range of symptoms ranging from mild symptoms to severe illness. Throughout the study, to surveil for COVID-19, the following prespecified symptoms that meet the criteria for suspicion of COVID-19 will be elicited weekly from the participant and the presence of any one of these symptoms lasting at least 48 hours (except for fever and/or respiratory symptoms) will result in the site arranging an Illness Visit to collect an NP swab sample within 72 hours:

- Fever (temperature $\geq$ 38ºC) or chills (of any duration, including $\leq$ 48 hours)
- Shortness of breath or difficulty breathing (of any duration, including $\leq$ 48 hours)
- Cough (of any duration, including $\leq$ 48 hours)
- Fatigue
- Muscle or body aches
- Headache
- New loss of taste or smell
- Sore throat
- Congestion or runny nose

- Nausea or vomiting
- Diarrhea

It is important to note that some of the symptoms of COVID-19 overlap with solicited systemic ARs that are expected after vaccination with mRNA-1273 (eg, myalgia, headache, fever, and chills). During the first 7 days after vaccination, when these solicited ARs are common, investigators should use their clinical judgement to decide if an NP swab sample should be collected (Part A, the Blinded Phase only). The collection of an NP swab sample prior to the Day 1 and Day 29 vaccination can help ensure that cases of COVID-19 are not overlooked. Any study participant reporting respiratory symptoms during the 7-day period after vaccination should be evaluated for COVID-19.

**Figure 3: Surveillance for COVID-19 Symptoms and the Corresponding Clinical Data Pathways**

Symptoms that meet the criteria for suspicion of COVID-19 infection

- Fever (temperature ≥ 38°C) or chills
- Cough
- Shortness of breath or difficulty breathing
- Fatigue
- Muscle or body aches
- Headache
- New loss of taste or smell
- Sore throat
- Congestion or runny nose
- Nausea or vomiting
- Diarrhea

*Elicited through interaction with the (in person or phone) and/or clinical evaluation by investigator*

- Clinical signs indicative of severe systemic illness, Respiratory Rates ≥ 30 per minute, Heart Rate ≥ 125 beats per minute, SpO2 ≤ 93% on room air at sea level or PaO2/FIO2 < 300 mm Hg, OR
- Respiratory failure or Acute Respiratory Distress Syndrome (ARDS), (defined as needing high-flow oxygen, non-invasive or mechanical ventilation, or ECMO), evidence of shock (systolic blood pressure < 90 mmHg, diastolic BP < 60 mmHg or requiring vasopressors), OR
- Significant acute renal, hepatic or neurologic dysfunction, OR
- Admission to an intensive care unit or death.

*From Medically-Attended HCP Visit (site or external)*

- Positive virologic result by RT-PCR for SARS-CoV-2 infection

*PCR results on NP Swab, Nasal Swab, or Saliva Sample*

During the course of the study, participants with symptoms of COVID-19 will be asked to return within 72 hours or as soon as possible to the study site or medically qualified staff from the study site will conduct a home visit as soon as possible to collect an NP swab sample (for RT-PCR), collect a blood sample for immunologic analysis of SARS-CoV-2 infection, and to evaluate for COVID-19. Both study site visits and home visits are referred to as Illness Visits. An additional NP swab sample will be collected and tested for the presence of other respiratory pathogens.

In addition, the study site may collect an additional (non-study/local) respiratory sample for SARS-CoV-2 testing to be able to render appropriate medical care for the study participant as determined by local standards of care. If neither a study site visit or home visit is possible, participants will be sent a saliva kit via courier or other Sponsor-approved method. The study site will arrange to retrieve a saliva sample by local courier or other Sponsor-approved method, and the sample will be tested by RT-PCR for SARS-CoV-2. If participants are confirmed to have SARS-CoV-2 infection, the investigator will notify the participant, and the participant's primary care physician, of the diagnosis. If the study participant does not have a primary care physician, the investigator will assist them to obtain one. The participant will also be instructed on infection prevention measures consistent with local public health guidance.

If scheduled, a study site Illness Visit includes assessments such as medical history, physical examination, blood sampling for clinical laboratory testing, and 2 NP swabs, 1 for viral PCR SARS-CoV-2 testing and 1 for multiplex PCR testing for respiratory viruses, to evaluate the severity of the clinical case. Radiologic imaging studies may be conducted. Blood samples will be collected for potential future immunologic assessment of SARS-CoV-2 infection. Every effort to complete an Illness Visit that includes both the NP swabs and blood draw should be made by the site, even in the presence of a local positive or negative Clinical Laboratory Improvement Amendments (CLIA) certified or CLIA-certified waiver laboratory result for the participant.

Cases are defined as participants meeting clinical criteria based both on symptoms for COVID-19 and on RT-PCR detection of SARS-CoV-2 from samples collected within 72 hours of the study participant reporting symptoms meeting the definition of COVID-19. Participants who are hospitalized for COVID-19 without the opportunity for a clinic or home visit will also be considered cases, assuming that the symptomology criteria for COVID-19 are met and a respiratory sample is positive for SARS-CoV-2 by PCR at a CLIA-certified or CLIA-certified waiver laboratory. Investigators are encouraged to try to obtain a respiratory sample during the course of hospitalization for submission to the study central laboratory, if feasible. The investigator should determine if the criteria for severe COVID-19 has been met.

Evidence of severe COVID-19 is defined as in Section 8.1.1.

All clinical findings will be recorded in the eCRF. All confirmed cases of COVID-19 will be captured as MAAEs, along with relevant concomitant medications and details about severity, seriousness, and outcome.

##### 8.1.3. Convalescent Period Starting with the Illness Visit

All study participants who experience COVID-19 symptoms and subsequently present for an Illness Visit (in-clinic or at home) will be given an instruction card listing symptoms and severity grading system, a thermometer, an oxygen saturation monitor, and saliva collection tubes. Participants will be trained on the use of the oxygen saturation monitor and how to take saliva specimens. The list of symptoms is presented in Section 8.1.2 and the severity scoring system is presented in Table 3.

**Table 3: Grading of COVID-19 Symptoms**

| Grading | All Symptoms | For Nausea/Vomiting ONLY | For Sense of Smell/Taste ONLY |
|---|---|---|---|
| None | No symptom | | |
| Mild | I had the symptom, but I could still do my normal activities | I was able to eat and drink normally | I had the symptom, but I retained some taste/smell |
| Moderate | The symptom really bothered me. It was hard to do my normal activities | It bothered me enough that I did not eat or drink normally | My taste/smell was significantly affected |
| Severe | The symptom was very bad. I was not able to do activities that I usually do | I could not eat or drink | I have no taste or smell |

The initial Illness Visit is considered Day 1 for the Convalescent Period. Starting on Day 2 of the Convalescent Period, the investigator (or medically qualified staff appropriately delegated by the investigator) will contact participants daily with telemedicine visits through Day 14 or until symptoms have resolved, whichever is later. Telemedicine visits may be conducted by videoconference or by audio only (telephone). During the telemedicine visit (preferably done in the evening), the participant will be asked to verbally report the severity of each symptom, their highest body temperature and lowest oxygen saturation for that day, and the investigator will determine if medical attention is required due to worsening of COVID-19 symptoms. The presence and severity of each symptom reported by the participant will be noted in the appropriate source document (Table 19).

During the telemedicine visits, participants will be reminded both to collect their own saliva sample on 3, 5, 7, 9, 14, and 21 days after the initial Illness Visit and to return the sample to the study site. Immediately upon receipt of a saliva sample, the study site will send it for testing to the study central virology laboratory.

During the telemedicine visits, if the participant has a positive result for SARS-CoV-2 from the Day 1 Illness Visit, the participant will continue the Convalescent Period. If the participant has a negative result for SARS-CoV-2 from the Day 1 Illness Visit, the participant will exit the Convalescent Period, including discontinuation of daily telemedicine visits and collection of saliva samples, and will return to their respective study schedule (Table 16, Table 17, Table 18, or Table 23). Participants who are pending a central laboratory PCR may exit from the Convalescent Period based on a negative PCR result from a CLIA-certified or CLIA-certified waiver local laboratory, at the investigator’s discretion. However, if the central laboratory PCR results are positive after exit from the Convalescent Period, the Convalescent Period must resume.

All participants confirmed to be COVID-19 cases will be scheduled for a convalescent visit (study site or home visit) 28 days after the initial Illness Visit. At this visit, a saliva sample will be collected and a blood sample will be drawn for immunologic assessment of SARS-CoV-2 infection (Table 19).

If the participant is hospitalized, medically qualified site personnel will try to obtain medical records and SARS-CoV-2 diagnostic results and document if the criteria for COVID-19 or severe COVID-19 have been met. If the participant is later discharged from the hospital during the 28-day period following diagnosis of COVID-19, the study site personnel will arrange for a resumption of a schedule for telemedicine visits and sampling for the remainder of the Convalescent Period, followed by a return to their respective study schedule (Table 16, Table 17, Table 18, or Table 23).

##### 8.1.4. Ancillary Supplies for Participant Use

Study sites will distribute Sponsor-provided oral thermometers and rulers for use by participants in assessing body temperature and injection site reactions for recording solicited ARs in eDiaries (Section 8.2.2). Based on availability, smartphone devices may be provided to those participants who do not have their own device to use for eDiary activities.

Participants will also receive the following Sponsor-provided supplies at Illness Visits where COVID-19 is suspected:

- An instruction card listing symptoms and severity grading system
- A pulse oximeter for measuring oxygen saturation
- Saliva collection tubes and instructions/means for returning saliva samples collected at home to the study site
- Additional oral thermometer, if required.

##### 8.1.5. Immunogenicity Assessments

Blood samples for immunogenicity assessments will be collected at the time points indicated in the SoEs (Section 11.1). On Day 1 and Day 29, as specified in Table 16, on OL-D1 and OL-D29 as specified in Table 21, on OL-D1 as specified in Table 22, and on BD-1 as specified in Table 23, blood samples for immunogenicity assessment will be collected before administration of IP. The following analytes will be measured:

- Serum bAb levels against SARS-CoV-2 as measured by ligand-binding assay specific to the SARS-CoV-2 S protein

- Serum bAb levels against SARS-CoV-2 as measured by ligand-binding assay specific to the SARS-CoV-2 nucleocapsid protein
- Serum nAb titer against SARS-CoV-2 as measured by pseudovirus and/or live virus neutralization assays

Serum will be tested using the ligand-binding assay specific to the SARS-CoV-2 nucleocapsid to determine the immunologic status of study participants at baseline and assess for seroconversion due to infection during the course of the study. Serum from a subset of participants will be tested in the other assays. The selection of the subset and timepoints to be tested will be described in the statistical analysis plan (SAP). Sample aliquots will be designed to ensure that backup samples are available and that adequate vial volumes may allow for further testing. The actual time and date of each sample collected will be recorded in the eCRF, and unique sample identification will be used to maintain the blind at the laboratory at all times and to allow for automated sample tracking and storage. Handling and preparation of the samples for analysis, as well as shipping and storage requirements, will be provided in a separate study manual.

The ligand-binding assay and measurement of nAb titers will be performed in laboratories designated by the Sponsor.

#### 8.2. Safety Assessments

Safety assessments will include monitoring and recording of the following for each participant:

- Solicited local and systemic ARs (Section 8.3.5) that occur during the 7 days following each injection (ie, the day of dosing and 6 subsequent days). Solicited ARs will be recorded daily using eDiaries (Section 8.2.2); (Part A, the Blinded Phase only).
- Unsolicited AEs observed or reported during the 28 days following each dose of IP (ie, the day of dosing and 27 subsequent days (Part A and Part C). Unsolicited AEs are AEs that are not included in the protocol-defined solicited ARs (Section 8.3.5
- AEs leading to discontinuation from dosing and/or study participation from Day 1 through Day 759 or withdrawal from the study.
- MAAEs from Day 1 through Day 759 or withdrawal from the study.
- SAEs from Day 1 through Day 759 or withdrawal from the study.
- Abnormal vital sign measurements.
- Physical examination findings.
- Pregnancy and accompanying outcomes.
- Concomitant medications and non-study vaccinations.

##### 8.2.1. Safety Phone Calls

A safety phone call is a telephone call made to the participant by medically qualified study staff. Medically qualified staff are those appropriately delegated individuals who are permitted to elicit verbal medical history from participants based on local regulations and local licensing requirements.

This call will follow a script, which will facilitate the collection of relevant safety information. The participant will be interviewed according to the script about occurrence of unsolicited AEs, MAAEs, SAEs, or AEs leading to study withdrawal and concomitant medications associated with those events, receipt of any non-study vaccinations, and pregnancy. Occurrence of AEs will only be collected by safety phone call during the Vaccination Phase (Table 16, Table 21, and Table 23).

The timing of the safety phone calls and the relevant safety information collected is provided in the SoEs (Section 11.1).

All safety information described by the participant must be documented in source documents and not documented on the script used for the safety telephone contact. All AEs, MAAEs, SAEs, and AEs leading to study withdrawal must be recorded in the eCRF as specified in Section 8.3.7.

##### 8.2.2. Use of Electronic Diaries

In Part A, the Blinded Phase of the study, at the time of consent, the participants must confirm they will be willing to complete an eDiary (for 7-day reactogenicity and to surveil weekly for COVID-19 symptoms) using either an application downloaded to their smartphone or using a device that is provided at the time of enrollment. This study will utilize the Medidata Patient Cloud Application as the eDiary for both collection of 7-day reactogenicity and weekly eDiary prompts to elicit an unscheduled Illness Visit if the participant is experiencing COVID-19 symptoms. This application allows for real-time data collection on a 21 CFR Part 11 compliant system directly from participants.

In Part A, the Blinded Phase of the study, before enrollment on Day 1, the participant will be instructed to download the eDiary application on their personal smartphone or will be provided a Sponsor-provisioned device to record solicited ARs (Section 8.3.5) and also to be utilized for eDiary prompts through the COVID-19 surveillance period.

In Part A, the Blinded Phase of the study, at each dosing visit, participants will record data into the eDiary starting approximately 30 minutes after dosing under supervision of the study site staff to ensure successful entry of assessments. The 30-minute assessment is an opportunity for site staff to train the participant. The site staff will perform any retraining as necessary. Study participants will continue to record data in an eDiary after they leave the study site, preferably in the evening and at the same time each day, on the day of dosing and for 6 days following dosing.

In Part A, the Blinded Phase of the study, the following local ARs will be solicited by the eDiary: pain at injection site, erythema (redness) at injection site, swelling/induration (hardness) at injection site, and localized axillary swelling or tenderness ipsilateral to the injection arm.

The following systemic ARs will be solicited by the eDiary: headache, fatigue, myalgia (muscle aches all over the body), arthralgia (aching in several joints), nausea/vomiting, body temperature (potentially fever), and chills.

In Part A, the Blinded Phase of the study, solicited local and systemic reactogenicity ARs, as defined in Section 8.3.5, will be collected on the day of each IP injection and during the 7 days after IP injection (ie, the day of dosing and 6 subsequent days). Any solicited AR that is ongoing beyond Day 7 will be reported in an eDiary until resolution. Adverse reactions recorded in diaries beyond Day 7 should be reviewed by study site staff either during the next scheduled phone call or at the next study site visit (Table 16).

If eDiary prompts result in identification of relevant safety events according to the study period-or symptoms of COVID-19, a follow-up safety call will be triggered.

In Part A, the Blinded Phase of the study, at each dosing visit, participants will be instructed or reminded on thermometer usage to measure body temperature, ruler usage to measure injection site erythema and swelling/induration (hardness), and self-assessment for localized axillary swelling or tenderness on the same side as the injection arm.

In Part A, the Blinded Phase of the study, daily oral body temperature measurement should be performed at approximately the same time each day using the thermometer provided by the study site.

Surveillance for signs and symptoms of COVID-19 will start at Part A Day 1 and continue through the entire Surveillance Phase of the study (Table 17 and Table 18). After participants complete Part A Vaccination Phase of the Study (Table 16 and Table 21), the weekly eDiary Safety Follow-up prompts will be triggered on Day 61 (Table 17). The weekly eDiary prompts will utilize the same Medidata Patient Cloud Application. Each week (ie, every 7 days) the participant will receive a prompt on their smartphone device to regularly surveil for signs and symptoms of COVID-19 and any other changes in health status. The participant will be trained on how to complete the weekly eDiary prompts at the Part A Day 57 clinic visit and also reminded to call the site immediately if they experience any COVID-19 symptoms. The weekly eDiary prompt will inquire about the following:

- Changes in health since the last time completing the weekly eDiary prompt or the last contact with the study staff
- Any known exposure to someone with SARS-CoV-2 infection or COVID-19 since the last time completing the weekly eDiary prompt or the last contact with the study staff
- Capture of any COVID-19 symptoms currently being experienced or experienced since the last time completing the weekly check-in as defined in Section 8.1.2
- Any contact with a healthcare provider that may indicate a MAAE.

A positive response by the participant to these prompts will result in a notification to both the participant and study staff to arrange a call. A follow-up safety call will be performed to the participant to determine if an unscheduled Illness Visit for the participant should be arranged as defined in Section 8.1.2. The results of the safety call should be recorded in the appropriate source documentation.

If a participant does not respond to the weekly eDiary within a 2-day window around the scheduled timepoint, study staff will follow-up directly with the participant via phone call or text to confirm their health status and to remind the participant of the importance of maintaining weekly contact via the eDiary prompt.

In addition, there will be an eDiary prompt to solicit the collection of information regarding participant's history of facial injections or dermal fillers, for cosmetic or medical indications such as migraine headaches.

##### 8.2.3. Demographics/Medical History

Demographic information relating to the participant's sex, age, and race will be recorded at Screening on the appropriate eCRF page.

Additionally, information regarding participant occupational circumstances (eg, essential worker status) will be collected at Screening.

Medical history of each participant, including risk factors for severe COVID-19 as defined in Section 6.2.1.1, will be collected and recorded on the Medical History eCRF page. Significant findings that were present prior to the signature of the informed consent will also be included in the Medical History eCRF page.

Study participants will also be asked to report history of receipt of seasonal influenza vaccine during the current influenza season (typically October through April in the Northern Hemisphere) as a concomitant medication.

##### 8.2.4. Physical Examination

A full physical examination, including vital signs, height, and weight, will be performed at Screening and on Day 1, and symptom-directed physical examinations at other scheduled time points as indicated in the SoEs (Section 11.1). The full examination will include assessment of skin, head, ears, eyes, nose, throat, neck, thyroid, lungs, heart, cardiovascular, abdomen, lymph nodes, and musculoskeletal system/extremities. Symptom-directed physical examinations may be performed at other timepoints at the discretion of the investigator.

On each dosing day before injection, the arm receiving the injection should be examined and the associated lymph nodes should be evaluated.

Body mass index will be calculated at the Screening Visit (Day 0).

Any clinically significant finding identified during a study visit after the first dose should be reported as a MAAE.

Significant new findings that begin or worsen after informed consent must be recorded on the AE eCRF page.

##### 8.2.5. Vital Sign Measurements

Vital signs will be measured at the time points indicated in the SoEs (Section 11.1). The participant will be seated for at least 5 minutes before all measurements are taken. On Day 1 and Day 29, vital sign measurements will be collected once before IP injection and at least 30 minutes after IP injection (before participants are discharged from the study site). When applicable, vital sign measurements should be performed before blood collection.

Febrile participants at Day 1 and Day 29 visits (fever is defined as a body temperature ≥ 38.0°C/100.4°F) may be rescheduled within the relevant window periods. Criteria for delay of study treatment are provided in Section 7.1. Afebrile participants with minor illnesses may be injected at the discretion of the investigator.

If any of the vital sign measurements meet the toxicity grading criteria for clinical abnormalities of Grade 3 or greater, the abnormal value and Grade will be documented on the AE page of the eCRF (unless there is another known cause of the abnormality that would result in an AE classification). The investigator will continue to monitor the participant with additional assessments until the vital sign value has reached the reference range, returns to the vital sign value at baseline, is considered stable, or until the investigator determines that follow-up is no longer medically necessary.

##### 8.2.6. Blood Sampling

The maximum planned volumes of blood sampled for immunogenicity analyses per participant are approximately 50 mL for 1 day, 100 mL for 28 days (SoE Table 16), and 300 to 420 mL (dependent on participation in Part B and Part C) for the total study volume (Table 4).

**Table 4: Maximum Planned Immunogenicity Blood Sampling Volumes per Participant by Visit**

| Study Visit Day | **Table 16 D1** | **Table 16 D29** | **Table 16 D57** | **Table 17 D209** | **Table 17 D394** | **Table 18 D759** | **Total** |
|---|---|---|---|---|---|---|---|
| Immunogenicity blood samples [a] | 50 mL | 50 mL | 50 mL | 50 mL | 50 mL | 50 mL | 300 mL |
| Study Visit Day | **Part B Day 1[b]** | **Part B Day 29[b]** | **Part B Day 57[b]** | NA | NA | NA | NA |
| Immunogenicity blood samples [a] | 20 mL | 20 mL | 20 mL | NA | NA | NA | 60 mL[c] |
| Study Visit Day | **Part C Day 1[d]** | **Part C Day 4[d]** | **Part C Day 29[d]** | **Part C Day 181[d]** | NA | NA | NA |
| Immunogenicity blood samples [a] | 24 mL | 8 mL | 24 mL | 24 mL | NA | NA | 80 mL |

Abbreviations: D = Day; NA = not applicable; RT-PCR = reverse transcriptase polymerase chain reaction.

[a] Additional blood samples (16 mL) will be taken at Illness Day 1 and Convalescent Visit Day 28 for those participants who report symptoms of COVID-19 and who have RT-PCR confirmed symptomatic COVID-19, respectively.

[b] If a visit from SoE Table 16 and 17 and a Part B visit overlap, only the Part B sample will be taken (using the Part B kit). The exception would be if an Illness Day 1 or Convalescent Visit Day 28 overlapped with a Part B visit. In this case the Illness and Convalescent Visit Day 28 samples would be taken and the corresponding kits used.

[c] For participants who received placebo during the Part A Blinded Phase and receive mRNA-1273 during Part B. Note that immunogenicity blood samples are collected only on OL-D1 and OL-D57 (Table 21) or OL-D1 and OL-D29 (Table 22) in Part B.

[d] If a visit from SoE Tables 17 and 18 and a Part C visit overlap, only the Part C sample will be taken (using the Part C kit). Exceptions would be if an Illness Day 1 or Convalescent Visit Day 28 overlapped with a Part C visit, in this case the Illness and Convalescent Visit Day 28 samples would be taken and corresponding kits used or in the case of a Part C Day 4 visit, both sets of samples would need to be taken using both kits.

#### 8.3. Safety Definitions and Procedures

##### 8.3.1. Adverse Event

An AE is defined as any untoward medical occurrence associated with the use of a drug in humans, whether or not considered drug related.

A TEAE is defined as any event not present before exposure to IP or any event already present that worsens in intensity or frequency after exposure.

**Events Meeting the Adverse Event Definition**

- Exacerbation of a chronic or intermittent pre-existing condition including either an increase in frequency and/or intensity of the condition.
- New conditions detected or diagnosed after the first dose of IP even though they may have been present before the start of the study.

**Events NOT Meeting the Adverse Event Definition**

- Medical or surgical procedure (eg, endoscopy, appendectomy): the condition that leads to the procedure should be the AE.
- Situations in which an untoward medical occurrence did not occur (social and/or convenience admission to a hospital).

An AR is any AE for which there is a reasonable possibility that the IP caused the AE (Section 8.3.5). For the purposes of investigational new drug safety reporting, "reasonable possibility" means that there is evidence to suggest a causal relationship between the IP and the AE.

An unsolicited AE is any AE reported by the participant that is not specified as a solicited AR in the protocol; or is specified as a solicited AR in the protocol, but starts outside the protocol-defined period for reporting solicited ARs (ie, for the 7 days after each dose of IP, Part A, Blinded Phase only).

##### 8.3.2. Medically Attended Adverse Events

An MAAE is an AE that leads to an unscheduled visit (including a telemedicine visit) to a healthcare practitioner (HCP). This would include visits to a study site for unscheduled assessments (eg, rash assessment, abnormal laboratory follow-up, COVID-19 [Section 8.1.1]) and visits to HCPs external to the study site (eg, urgent care, primary care physician). Investigators will review unsolicited AEs for the occurrence of any MAAEs. All MAAEs must be fully reported on the MAAE page of the eCRF.

###### 8.3.2.1. Anaphylaxis

All suspected cases of anaphylaxis should be recorded as MAAEs and reported as an SAE, based on criteria for a medically important event, unless the event meets other serious criteria. As an SAE, the event should be reported to the Sponsor or designee immediately and in all circumstances within 24 hours as per Section 8.3.12 (Reporting SAEs). The investigator will submit any updated anaphylaxis case data to the Sponsor within 24 hours of it being available. For reporting purposes, a participant who displays signs/symptoms consistent with anaphylaxis as shown below should be reported as a potential case of anaphylaxis. This is provided as general guidance for investigators and is based on the Brighton Collaboration case definition (Rüggeberg et al 2007).

Anaphylaxis is an acute hypersensitivity reaction with multi-organ system involvement that can present as, or rapidly progress to, a severe life-threatening reaction. It may occur following exposure to allergens from a variety of sources. Anaphylaxis is a clinical syndrome characterized by:

- Sudden onset AND
- Rapid progression of signs and symptoms AND
- Involving 2 or more organ systems, as follows:
    - **Skin/mucosal**: urticaria (hives), generalized erythema, angioedema, generalized pruritus with skin rash, generalized prickle sensation, red and itchy eyes
    - **Cardiovascular**: measured hypotension, clinical diagnosis of uncompensated shock, loss of consciousness or decreased level of consciousness, evidence of reduced peripheral circulation
    - **Respiratory**: bilateral wheeze (bronchospasm), difficulty breathing, stridor, upper airway swelling (lip, tongue, throat, uvula, or larynx), respiratory distress, persistent dry cough, hoarse voice, sensation of throat closure, sneezing, rhinorrhea
    - **Gastrointestinal**: diarrhea, abdominal pain, nausea, vomiting

##### 8.3.3. Adverse Events of Special Interest

An adverse event of special interest (AESI) is a serious or nonserious AE of scientific and medical concern specific to the Sponsor's product or program, for which ongoing monitoring and immediate notification by the investigator to the Sponsor is required and documentation is in the form of a case narrative. Such events may require further investigation to characterize and understand them. Refer to Section 11.5 for a list of AESIs pertinent to this study. All AESIs will be collected through the entire study period and must be reported to the Sponsor or designee immediately and in all circumstances within 24 hours of becoming aware of the event via the electronic data capture system. If a site receives a report of a new AESI from a study participant or receives updated data on a previously reported AESI, and the eCRF has been taken offline, then the site can report this information on a paper AESI form using the SAE Mailbox, the SAE Hotline, or the SAE Fax line (Section 8.3.11).

**Acute Myocarditis and/or Pericarditis**

These definitions are intended to serve as a guide to help in the reporting of suspected cases of myocarditis and/or pericarditis; however, the diagnosis of suspected cases is left to the investigator's clinical judgement.

All suspected cases of probable and confirmed myocarditis, pericarditis, or myopericarditis should be recorded as an AESI, and reported as an SAE, if the event meets seriousness criteria. As an SAE, the event should be reported to the Sponsor or designee immediately and in all circumstances within 24 hours as per Section 8.3.11. The investigator will submit any updated myocarditis, pericarditis, or myopericarditis case data to the Sponsor within 24 hours of it being available. For reporting purposes, a participant who displays signs/symptoms consistent with the CDC case definitions as described below (Gargano et al 2021), should be reported as a potential case of confirmed or probable myocarditis, pericarditis, or myopericarditis.

**Acute Myocarditis Case Definition**

Presence of ≥ 1 new or worsening of the following clinical symptoms (persons who lack the listed symptoms but who meet other criteria may be classified as subclinical myocarditis [probable or confirmed]):

- Chest pain/pressure/discomfort
- Dyspnea/shortness of breath/pain with breathing
- Palpitations
- Syncope

AND

For PROBABLE CASE:

Presence of ≥ 1 new finding of the following:

- Troponin level above upper limit of normal (any type of troponin)
- Abnormal electrocardiogram (ECG or EKG) or rhythm monitoring findings consistent with myocarditis
    - To meet the ECG or rhythm monitoring criterion, a probable case must include at least 1 of the following:
        - ST segment or T-wave abnormalities
        - Paroxysmal or sustained atrial, supraventricular, or ventricular arrhythmias
        - AV nodal conduction delays or intraventricular conduction defects
- Abnormal cardiac function or wall motion abnormalities on echocardiogram
- Cardiac magnetic resonance imaging (cMRI) finding consistent with myocarditis (Ferreira et al 2018)

AND

- No other identifiable cause of the symptoms and findings

For CONFIRMED CASE:

- Histopathologic confirmation of myocarditis (using Dallas criteria [Aretz et al 1987])

OR

- cMRI findings consistent with myocarditis in the presence of troponin level above upper limit of normal (any type of troponin)

AND

- No other identifiable cause of the symptoms and findings

**Acute Pericarditis Case Definition**

Presence of $\geq 2$ new or worsening of the following clinical features (Adler et al 2015):

- Acute chest pain (Typically described as pain made worse by lying down, deep inspiration, or cough; and relieved by sitting up or leaning forward, although other types of chest pain may occur)
- Pericardial rub on examination
- New ST-elevation or PR-depression on EKG
- New or worsening pericardial effusion on echocardiogram or magnetic resonance imaging

**Myopericarditis Case Definition**

Participants who meet criteria for both myocarditis and pericarditis may be described under myopericarditis.

##### 8.3.4. Serious Adverse Events

An AE (including an AR) is considered an SAE if, in the view of either the investigator or Sponsor, it results in any of the following outcomes:

- **Death**
  A death that occurs during the study or that comes to the attention of the investigator during the protocol-defined follow-up period must be reported to the Sponsor, whether or not it is considered related to IP.
- **Is life-threatening**
  An AE is considered life-threatening if, in the view of either the investigator or the

Sponsor, its occurrence places the participant at immediate risk of death. It does not include an AE that, had it occurred in a more severe form, might have caused death.

- **Inpatient hospitalization or prolongation of existing hospitalization**
  In general, inpatient hospitalization indicates the participant was admitted to the hospital or emergency ward for at least one overnight stay for observation and/or treatment that would not have been appropriate in the physician's office or outpatient setting. The hospital or emergency ward admission should be considered an SAE regardless of whether opinions differ as to the necessity of the admission. Complications that occur during inpatient hospitalization will be recorded as an AE; however, if a complication/AE prolongs hospitalization or otherwise fulfills SAE criteria, the complication/AE will be recorded as a separate SAE.

- **Persistent or significant incapacity or substantial disruption of the ability to conduct normal life functions**
  This definition is not intended to include experiences of relatively minor medical significance such as uncomplicated headache, nausea/vomiting, diarrhea, influenza, and accidental trauma (eg, sprained ankle) which may interfere with or prevent everyday life functions but do not constitute a substantial disruption.

- **Congenital anomaly or birth defect**

- **Medically important event**
  Medical judgment should be exercised in deciding whether SAE reporting is appropriate in other situations, such as important medical events that may not be immediately life-threatening or result in death or hospitalization but may jeopardize the participant or require medical or surgical intervention to prevent one of the other outcomes listed in the above definition. These events should usually be considered serious. Examples of such medical events include allergic bronchospasm requiring intensive treatment in an emergency room or at home, blood dyscrasias or convulsions that do not result in inpatient hospitalization, or the development of drug dependency or drug abuse.

##### 8.3.5. Solicited Adverse Reactions (Part A, Blinded Phase Only)

The term "reactogenicity" refers to the occurrence and intensity of selected signs and symptoms (ARs) occurring after IP injection. The eDiary will solicit daily participant reporting of ARs using a structured checklist (Section 8.2.2). Participants will record such occurrences in an eDiary on the day of each IP injection and for the 6 days after the day of dosing.

Severity grading of reactogenicity will occur automatically based on participant entry into the eDiary according to the grading scales presented in Table 5 modified from the Toxicity Grading

Scale for Healthy Adult and Adolescent Volunteers Enrolled in Preventative Vaccine Clinical Trials (DHHS 2007).

If a solicited local or systemic AR continues beyond 7 days after dosing, the participant will be prompted daily to capture solicited local or systemic AR in the eDiary until resolution. Adverse reactions recorded in eDiaries beyond Day 7 should be reviewed by the investigator either via phone call or at the following study visit. All solicited ARs (local and systemic) will be considered causally related to dosing.

#### Table 5: Solicited Adverse Reactions and Grades

| Reaction | Grade 0 | Grade 1 | Grade 2 | Grade 3 | Grade 4 |
|---|---|---|---|---|---|
| Injection site pain | None | Does not interfere with activity | Repeated use of over-the-counter pain reliever > 24 hours or interferes with activity | Any use of prescription pain reliever or prevents daily activity | Requires emergency room visit or hospitalization |
| Injection site erythema (redness) | < 25 mm/<br>< 2.5 cm | 25 - 50 mm/<br>2.5 - 5 cm | 51 - 100 mm/<br>5.1 - 10 cm | > 100 mm/<br>> 10 cm | Necrosis or exfoliative dermatitis |
| Injection site swelling/induration (hardness) | < 25 mm/<br>< 2.5 cm | 25 - 50 mm/<br>2.5 - 5 cm | 51 - 100 mm/<br>5.1 - 10 cm | > 100 mm/<br>> 10 cm | Necrosis |
| Axillary (underarm) swelling or tenderness ipsilateral to the side of injection | None | No interference with activity | Repeated use of over-the-counter (non-narcotic) pain reliever > 24 hours or some interference with activity | Any use of prescription (narcotic) pain reliever or prevents daily activity | Emergency room visit or hospitalization |
| Headache | None | No interference with activity | Repeated use of over-the-counter pain reliever > 24 hours or some interference with activity | Significant; any use of prescription pain reliever or prevents daily activity | Requires emergency room visit or hospitalization |
| Fatigue | None | No interference with activity | Some interference with activity | Significant; prevents daily activity | Requires emergency room visit or hospitalization |
| Myalgia (muscle aches all over body) | None | No interference with activity | Some interference with activity | Significant; prevents daily activity | Requires emergency room visit or hospitalization |
| Arthralgia (joint aches in several joints) | None | No interference with activity | Some interference with activity | Significant; prevents daily activity | Requires emergency room visit or hospitalization |

| **Reaction** | **Grade 0** | **Grade 1** | **Grade 2** | **Grade 3** | **Grade 4** |
|---|---|---|---|---|---|
| Nausea/vomiting | None | No interference with activity or 1-2 episodes/ 24 hours | Some interference with activity or > 2 episodes/24 hours | Prevents daily activity, requires outpatient intravenous hydration | Requires emergency room visit or hospitalization for hypotensive shock |
| Chills | None | No interference with activity | Some interference with activity not requiring medical intervention | Prevents daily activity and requires medical intervention | Requires emergency room visit or hospitalization |
| Fever (oral) | < 38.0°C<br>< 100.4°F | 38.0 – 38.4°C<br>100.4 – 101.1°F | 38.5 – 38.9°C<br>101.2 – 102.0°F | 39.0 – 40.0°C<br>102.1 – 104.0°F | > 40.0°C<br>> 104.0°F |

Any solicited AR that meets any of the following criteria must be entered into the participant's source document and must also be recorded as an AE in the participant's Adverse Event eCRF:

- Solicited local or systemic AR that results in a visit to an HCP (MAAE)
- Solicited local or systemic AR leading to the participant withdrawing from the study or the participant being withdrawn from the study by the investigator (AE leading to withdrawal)
- Solicited local or systemic AR lasting beyond 7 days post injection
- Solicited local or systemic AR that leads to participant withdrawal from IP
- Solicited local or systemic AR that otherwise meets the definition of an SAE

###### 8.3.6. Recording and Follow-up of Pregnancy

Female participants who have a positive pregnancy test at Screening should not be enrolled; participants who have a positive pregnancy test any time during the study should receive no further dosing with IP but should be asked to remain in the study and be followed-up for safety. Pregnancy testing is scheduled to occur at Screening, Blinded and Open-label Day 1, Blinded and Open-label Day 29, and Part C BD-1 (Table 16, Table 21, Table 22, and Table 23).

Details of all pregnancies in female participants will be collected after the start of study treatment and until Day 759.

- If a pregnancy is reported, the investigator should inform the Sponsor within 24 hours of learning of the pregnancy and should follow the procedures outlined in this section.
- Abnormal pregnancy outcomes (eg, spontaneous abortion, fetal death, stillbirth, congenital anomalies, ectopic pregnancy) are considered SAEs.

Pregnancies occurring in participants after enrollment must be reported to Sponsor or designee within 24 hours of the site learning of its occurrence. If the participant agrees to submit this information, the pregnancy must be followed to determine the outcome, including spontaneous or voluntary termination, details of the birth, and the presence or absence of any birth defects, congenital abnormalities, or maternal and/or newborn complications. This follow-up should occur even if intended duration of the safety follow-up for the study has ended. Pregnancy report forms will be distributed to the study site to be used for this purpose. The investigator must immediately (within 24 hours of awareness) report to the Sponsor any pregnancy resulting in an abnormal outcome according to the procedures described for SAEs.

##### 8.3.7. Recording and Follow-up of an AE and/or SAE

The investigator is responsible for ensuring that all AEs and SAEs are recorded in the eCRF and reported to the Sponsor.

Solicited ARs will be collected from Day 1 through 7 days after each dose (Part A: Blinded Phase only). Other (unsolicited) AEs will be collected from Day 1 through 28 days after each dose in Part A and Part C only.

Both MAAEs and SAEs will be collected from Blinded and Open-label Day 1 throughout entire study duration (Day 759 for all participants), as specified in the SoEs (Table 16, Table 17, Table 18, Table 19, Table 21, Table 22, and Table 23). Any AEs occurring before receipt of IP will be analyzed separately from TEAEs.

At every study site visit or telephone contact, participants will be asked a standard question to elicit any medically related changes, including surveillance for COVID-19 symptoms, in their well-being according to the scripts provided. Participants will also be asked if they have been hospitalized, had any accidents, used any new medications, changed concomitant medication regimens (both prescription and over-the-counter medications), or had any non-study vaccinations.

In addition to participant observations, physical examination findings or other documents relevant to participant safety classified as an AE will be documented on the AE page of the eCRF.

After the initial AE/SAE report, the investigator is required to proactively follow each participant at subsequent visits/contacts. All AEs and SAEs will be treated as medically appropriate and followed until resolution, stabilization, the event is otherwise explained, or the participant is LTFU (as defined in Section 7.4).

##### 8.3.8. Time Period and Frequency for Collecting AE and SAE Information

All confirmed serious COVID-19 cases and SAEs will be recorded and reported to the Sponsor or designee immediately and under no circumstance should this exceed 24 hours, as indicated in Section 8.3.11. The investigator will submit any updated serious COVID-19 cases and SAE data to the Sponsor within 24 hours of it being available. COVID-19 cases are defined in Section 8.1.1.

Investigators are not obligated to actively seek AEs or SAEs after conclusion of the study participation (Day 759). However, if an investigator learns of any SAE, including a death, at any time after a participant has been discharged from the study and considers the event to be reasonably related to the study IP or study participation, the investigator must promptly notify the Sponsor.

##### 8.3.9. Assessment of Intensity

An event is defined as “serious” when it meets at least one of the predefined outcomes as described in the definition of an SAE (Section 8.3.3), NOT when it is rated as severe.

The severity (or intensity) of an AR or AE refers to the extent to which it affects the participant’s daily activities. The Toxicity Grading Scale for Healthy Adult and Adolescent Volunteers Enrolled in Preventative Vaccine Clinical Trials (DHHS 2007) will be used to categorize local and systemic reactogenicity events (solicited ARs), clinical laboratory test results, and vital sign measurements observed during this study. Specific criteria for local and systemic reactogenicity events are presented in Section 8.3.5.

The determination of severity for all unsolicited AEs should be made by the investigator based upon medical judgment and the definitions of severity as follows:

- Mild: These events do not interfere with the participant’s daily activities.
- Moderate: These events cause some interference with the participant’s daily activities and require limited or no medical intervention.
- Severe: These events prevent the participant’s daily activity and require intensive therapeutic intervention.

Study staff should elicit from the participant the impact of AEs on the participant’s activities of daily living to assess severity and document appropriately in the participant’s source documentation. Changes in the severity of an AE should be documented in the participant’s source documentation to allow an assessment of the duration of the event at each level of intensity to be performed. An AE characterized as intermittent requires documentation of onset and duration of each episode. An AE that fluctuates in severity during the course of the event is reported once in the eCRF at the highest severity observed.

##### 8.3.10. Assessment of Causality

The investigator's assessment of an AE's relationship to IP is part of the documentation process but is not a factor in determining what is or is not reported in the study.

The investigator will assess causality (ie, whether there is a reasonable possibility that the IP caused the event) for all AEs and SAEs. The relationship will be characterized using the following classification:

**Not related:** There is not a reasonable possibility of a relationship to the IP. Participant did not receive the IP OR temporal sequence of the AE onset relative to administration of the IP is not reasonable OR the AE is more likely explained by another cause than the IP.

**Related:** There is a reasonable possibility of a relationship to the IP. There is evidence of exposure to the IP. The temporal sequence of the AE onset relative to the administration of the IP is reasonable. The AE is more likely explained by the IP than by another cause.

##### 8.3.11. Reporting Adverse Events

The investigator is responsible for reporting all AEs that are observed or reported during the study, regardless of their relationship to IP or their clinical significance. If there is any doubt as to whether a clinical observation is an AE, the event should be reported.

All unsolicited AEs reported or observed during the study will be recorded on the AE page of the eCRF. Information to be collected includes, type of event, time of onset, investigator -specified assessment of severity (impact on activities of daily living) and relationship to IP, time of resolution of the event, seriousness, as well as any required treatment or evaluations, and outcome. The unsolicited AEs resulting from concurrent illnesses, reactions to concurrent illnesses, reactions to concurrent medications, or progression of disease states must also be reported. All AEs will be followed until they are resolved or stable or judged by the investigator to be not clinically significant. The Medical Dictionary for Regulatory Activities (MedDRA) will be used to code all unsolicited AEs.

Any medical condition that is present at the time that the participant is screened but does not deteriorate should not be reported as an unsolicited AE. However, if it deteriorates at any time during the study, it should be recorded as an unsolicited AE.

Any AE or COVID-19 case considered serious by the investigator or that meets SAE criteria (Section 8.3.4) must be reported to the Sponsor immediately (within 24 hours of becoming aware of the SAE or COVID-19 case). The investigator will assess whether there is a reasonable possibility that the IP caused the SAE. The Sponsor will be responsible for notifying the relevant regulatory authorities of any SAE as outlined in the 21 US CFR Parts 312 and 320. The investigator is responsible for notifying the institutional review board (IRB) directly.

If the eCRF is unavailable at the time of the SAE, the following contact information is to be used for SAE reporting:

- SAE Mailbox:
- SAE Hotline (USA and Canada): +1-866-599-1341
- SAE Fax line (USA and Canada): +1-866-599-1342

##### 8.3.12. Regulatory Reporting Requirements for SAEs

Prompt notification by the investigator to the Sponsor of a SAE is essential so that legal obligations and ethical responsibilities towards the safety of participants and the safety of a study intervention under clinical investigation are met.

The Sponsor has a legal responsibility to notify both the local regulatory authority and other regulatory agencies about the safety of a study intervention under clinical investigation. The Sponsor will comply with country-specific regulatory requirements relating to safety reporting to the regulatory authority, IRBs, and investigators.

Investigator safety reports must be prepared for suspected unexpected serious ARs according to local regulatory requirements and Sponsor policy and forwarded to investigators as necessary.

An investigator who receives an investigator safety report describing a SAE or other specific safety information (eg, summary or listing of SAEs) from the Sponsor will review and then file it along with the IB and will notify the IRB/IEC, if appropriate according to local requirements.

#### 8.4. Monitoring Committees

##### 8.4.1. Protocol Safety Review Team

A Protocol Safety Review Team will be formed to review interim and cumulative blinded safety data on a regular basis with a remit to escalate concerns to the DSMB.

##### 8.4.2. Data and Safety Monitoring Board

An independent DSMB will periodically review blinded and unblinded data, including both safety and cases of COVID-19 at scheduled data review meetings and at 2 planned IAs.

- In addition to blinded and unblinded review of safety data, at each data review meeting the DSMB will review the numbers and rate of COVID-19 cases, including rate of severe COVID disease with prespecified thresholds for imbalance in the treatment groups which would trigger halting rules.
- At the IA, the DSMB will review the IA results and make recommendations to an Oversight Group in terms of study results reporting and unblinding based on the boundaries of early efficacy as described in Section 9.6 of the protocol. The Oversight Group will be comprised of a voting member each from the Sponsor, Biomedical Advance Research and Development Authority (BARDA), and NIAID.
- The DSMB will monitor the study for non-efficacy at the IA. The boundary for non-efficacy is non-binding and will be provided in the DSMB analysis plan.

- The DSMB will also monitor the study for vaccine harm based on severe COVID-19. Continuous harm monitoring will be provided for COVID-19 and severe COVID-19 separately. For harm monitoring, cases will be counted starting after the first dose of study vaccination. Boundaries will be provided based on the exact 1-sided binomial tests conditional on the total number of cases under the assumption of VE=0%. If the prespecified stopping boundary is reached for either COVID-19 or severe COVID-19, then the unblinded statisticians will immediately inform the DSMB that the harm rules have been met. Details will be provided in the DSMB analysis plan.
- The boundaries are considered guidelines, ie, a recommendation to modify the study would not be based solely on statistical rules, as many other factors (ie, totality of the data from the study including additional efficacy, safety and immunogenicity endpoints as well as data external to the study) may be part of the decision process. In the case of a recommendation to continue the trial regardless of crossing the boundaries for efficacy or inefficacy, the reason for disregarding the boundary must be documented in the meeting minutes and communicated to the Sponsor.

After each data review meeting or IA, the DSMB will make a recommendation to the Sponsor through an Oversight Group to take one of the following courses of action:

- Stop further enrollment due to meeting criteria for early efficacy or due to a safety concern.
- Pause enrollment and consider a change in study design.
- Continue enrollment and/or study conduct as planned.

The Sponsor may also request that the DSMB conduct ad hoc reviews of safety events from this study or other data, including new nonclinical or clinical information related to mRNA-1273 external to this study. The DSMB will review all available study data to adjudicate such events in accordance with the DSMB charter.

The DSMB composition, its remit, and frequency of data review will be further defined in the DSMB charter and analysis plan.

##### 8.4.3. Adjudication Committee

An Adjudication Committee (AC) will be assembled for the purpose of reviewing potential cases to determine if the criteria for the primary and secondary endpoints have been met. The AC will remain blinded to treatment assignment when adjudicating cases for participants whose treatment group is still blinded. The AC composition, its remit, and frequency of data review will be further defined in a charter.

###### 8.4.4. Independent Cardiac Event Adjudication Committee

An independent Cardiac Event Adjudication Committee (CEAC) that includes pediatric and adult cardiologists will review suspected cases of myocarditis and pericarditis to determine if they meet CDC criteria of “probable” or “confirmed” events, and to assess severity (Gargano et al 2021). Any cases that the CEAC assesses as representing probable or confirmed cases of myocarditis or pericarditis will be referred to the Sponsor, who will then make a final decision on whether to suspend further enrollment and/or study dosing based on an assessment of the overall potential risk to study participants.

The CEAC will operate under the rules of an approved charter that will be written and reviewed at the organizational meeting of the CEAC. Details regarding the CEAC composition, responsibilities, procedures, and frequency of data review will be defined in its charter.

#### 8.5. Management of Overdose

As the study treatment is to be administered by a healthcare professional, it is unlikely that an overdose will occur. Dose deviations will be tracked as protocol deviations (Section 11.2.8).

#### 8.6. Pharmacokinetics

Pharmacokinetic parameters are not evaluated in this study.

#### 8.7. Pharmacodynamics

Pharmacodynamic parameters are not evaluated in this study.

#### 8.8. Exploratory Assessments and Biomarkers

Exploratory assessments may include assessment of biomarkers for safety, reactogenicity, inflammatory and cardiac function. Serologic markers of disease severity, immune response to SARS-CoV-2, RT-PCR of NP swab or saliva samples, and genetic sequences of SARS-CoV-2 strains isolated from participants’ samples may also be measured.

#### 8.9. Medical Resource Utilization and Health Economics

Medical resource utilization and health economics parameters are not evaluated in this study.

### 9. STATISTICAL CONSIDERATIONS

This section summarizes the planned statistical analysis strategy and procedures for the study, focusing on the Blinded Phase of the study (Part A). The details of statistical analysis will be provided in the SAP, which will be finalized before the clinical database lock for the study and treatment unblinding. If, after the study has begun, but prior to any unblinding, changes are made to primary and/or key secondary objectives/hypotheses, or the statistical methods related to those hypotheses, then the protocol will be amended (consistent with ICH Guideline E9). Changes to other secondary or exploratory analyses made after the protocol has been finalized, along with an explanation as to when and why they occurred, will be listed in the SAP or Clinical Study Report (CSR) for the study. Ad hoc exploratory analyses, if any, will be clearly identified in the CSR. Statistical analysis strategy incorporating the Open-label Phase (Part B), and to assess the BD of mRNA-1273 (Part C) will be provided in a SAP.

#### 9.1. Blinding and Responsibility for Analyses

Blinding during the Part A, the Blinded Phase of the study will be conducted as described in Section 6.2.8. The Sponsor Biostatistics department or designee will generate the randomized allocation schedule(s) for study treatment assignment. Randomization will be implemented via an IRT.

Planned interim and primary analyses are described in Section 9.6. Participant-level unblinding will be restricted to an independent unblinded statistician and, as needed, a statistical programmer performing the IAs, who will have no other responsibilities associated with the study.

In addition to the routine study monitoring outlined in this protocol, an external DSMB will review interim data to safeguard the interests of clinical study participants and to enhancing the integrity of the study. The DSMB will review treatment-level results of the IAs, provided by the independent unblinded statistician. Limited additional Sponsor personnel may be unblinded to the treatment-level results of the IAs, if required, in order to act on the recommendations of the DSMB. The extent to which individuals are unblinded with respect to results of IAs will be documented. Depending on the recommendation of the DSMB, the Sponsor may prepare a regulatory submission after an IA. In this case, pre-identified Sponsor members including the analysis and reporting team will be unblinded to treatment assignments and remain unblinded for the remainder of the study. Participants and investigators will remain blinded.

#### 9.2. Statistical Hypotheses

For the primary efficacy objective, the null hypothesis of this study is that the VE of mRNA-1273 to prevent first occurrence of COVID-19 is ≤ 30% (ie, $H_0^{efficacy}$: VE ≤ 0.3). The study will be considered to meet the primary efficacy objective if the corresponding CI of VE rules out 30% at either one of the IAs or at the primary analysis.

Vaccine efficacy is defined as the percent reduction in the hazard of the primary endpoint (mRNA-1273 vs. placebo). Equivalently, the null hypothesis is:

$H_0^{efficacy}$: HR ≥ 0.7 (equivalently, proportional hazards VE ≤ 0.3).

A stratified Cox proportional hazard model will be used to assess the magnitude of the treatment group difference (ie, HR) between mRNA-1273 and placebo at a 1-sided 0.025 significance level.

The primary analysis population for efficacy will be the PP Set, defined in Section 9.4. In the primary analysis of efficacy, cases will be counted starting 14 days after the second dose of IP.

#### 9.3. Sample Size Determination

The sample size is driven by the total number of cases to demonstrate VE (mRNA-1273 vs. placebo) to prevent COVID-19 in Part A. Under the assumption of proportional hazards over time and with 1:1 randomization of mRNA-1273 and placebo, a total of 151 COVID-19 cases will provide 90% power to detect a 60% reduction in hazard rate (60% VE), rejecting the null hypothesis $H_0$: VE ≤ 30%, with 2 IAs at 35% and 70% of the target total number of cases using a 1-sided O'Brien-Fleming boundary for efficacy and a log-rank test statistic with a 1-sided false positive error rate of 0.025. The total number of cases pertains to the PP Set accruing at least 14 days after the second dose. There are 2 planned IAs in this study, which will be performed when approximately 35% and 70% of the target total number of cases have been observed. Approximately 30,000 participants will be randomized with the following assumptions:

- The target VE against COVID-19 is 60% (with 95% CI lower bound ruling out 30%, rejecting the null hypothesis $H_0$: VE ≤ 30%)
- A 6-month COVID-19 incidence rate of 0.75% in the placebo arm
- An annual dropout rate of 2% (loss of evaluable participants)
- Two IAs at 35% and 70% of total target cases across the 2 treatment groups with O'Brien-Fleming boundaries for efficacy monitoring
- 3-month uniform accrual
- Approximately 15% of participants will be excluded from the PP population, and participants are at risk for COVID-19 starting 14 days after the second dose

Table 6 provides sample size with 90% power to demonstrate VE on COVID-19.

**Table 6: Conditions and Sample Size to Demonstrate Vaccine Efficacy**

| Target VE | Lower Bound | Randomization Ratio | Total # of Cases | 6-Month Incidence Rate | | Total Sample Size[a] |
|---|---|---|---|---|---|---|
| -- | -- | -- | -- | Placebo | mRNA-1273 | -- |
| 60% | 30% | 1:1 | 151 | 0.75% | 0.30% | 30,000 |

a Sample size to account for 15% participants to be excluded from the PP Set (eg, seropositive at baseline, have not received planned IP).

The sample size is calculated using R package gsDesign (Anderson 2020).

Under these above assumptions including 6-month incidence rate of 0.75% on placebo, with 30,000 participants, it will take approximately 5, 8, and 10 months from study start (first subject first dose), respectively, to accrue 35% (approximately 53), 70% (approximately 106) and 100% (151) of the target number of cases in the PP Set.

Figure 4 shows the power of the primary efficacy endpoint under true VE at the 2 planned IAs and the primary efficacy analysis assuming a total of 151 events.

**Figure 4: Boundary Crossing Probabilities by Effect Size**

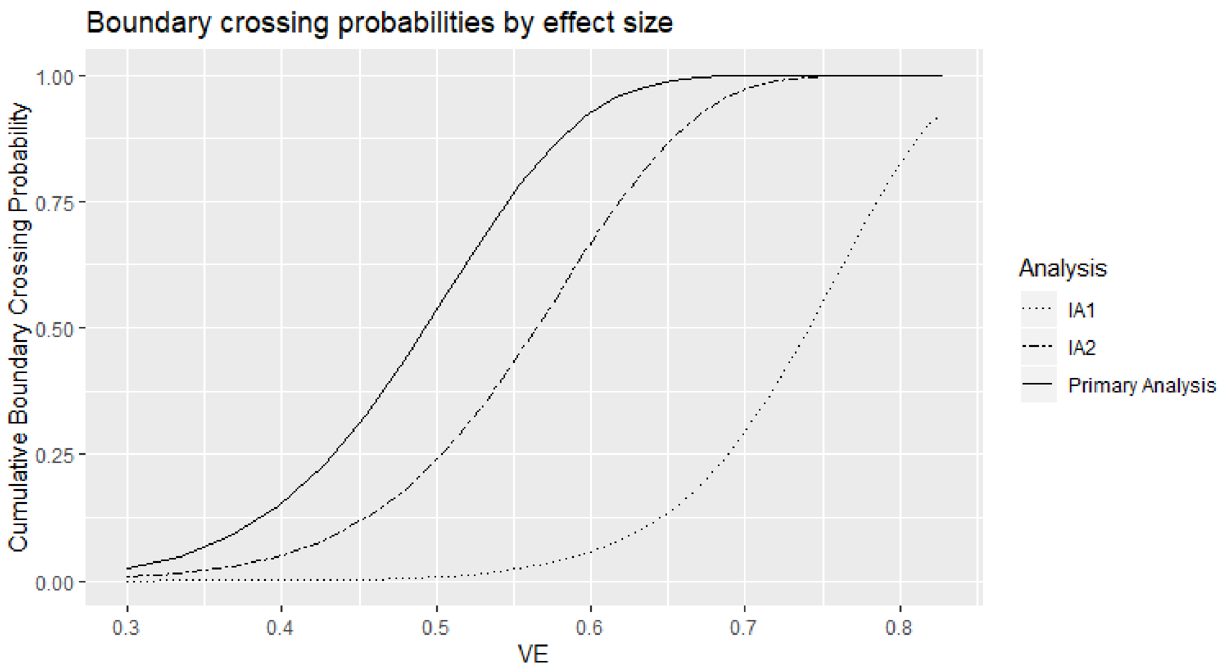

The Sponsor may adjust the size of the study or duration of follow-up based on the blinded review of the total number of cases of COVID-19 accrued during the study, in addition to estimated percentages of study participants with serologic evidence of SARS-CoV-2 infection at baseline.

##### 9.3.1. Power for Selected Secondary Efficacy Endpoints

For the secondary objective on VE against virologically confirmed SARS-CoV-2 infection or COVID-19 regardless of symptomology or severity (COV-INF), the study will have ≥ 90% power to demonstrate the VE is above 30% (to reject null hypothesis VE ≤ 30%) at 1-sided alpha of 2.5% if the true VE to prevent COV-INF is 60% because every COVID-19 endpoint is necessarily a COV-INF endpoint.

For the secondary objective on VE against severe COVID-19, Table 7 provides power of demonstrating VE based on a total of 30 and 60 events under different scenarios of true VE and VE criteria.

**Table 7: Power of Demonstrating Vaccine Efficacy Against Severe COVID-19**

| True VE | Total No. of Severe COVID-19 Cases | Severe COVID-19 (Secondary Objective) | |
|---|---|---|---|
| | | To Reject Null Hypothesis | Power |
| 60% | 30 | VE≤ 0% | 70.90% |
| | | VE≤ 10% | 60.30% |
| | | VE≤ 20% | 47.50% |
| 70% | | VE≤ 0% | > 90% |
| | | VE≤ 10% | 85.30% |
| | | VE≤ 20% | 76.60% |
| 80% | | VE≤ 0% | > 90% |
| | | VE≤ 10% | > 90% |
| | | VE≤ 20% | > 90% |
| 60% | 60 | VE≤ 0% | > 90% |
| | | VE≤ 10% | 88.10% |
| | | VE≤ 20% | 76.60% |
| 70% | | VE≤ 0% | > 90% |
| | | VE≤ 10% | > 90% |
| | | VE≤ 20% | > 90% |
| 80% | | VE≤ 0% | > 90% |
| | | VE≤ 10% | > 90% |
| | | VE≤ 20% | > 90% |

Abbreviation: VE = vaccine efficacy.

#### 9.4. Analysis Populations

Analysis populations for statistical analyses are Randomization Set, Full Analysis Set (FAS), modified intent-to-treat (mITT) set, PP Set, Immunogenicity Subset, Solicited Safety Set, and Safety Set, as shown in Table 8.

**Table 8: Populations for Analyses**

| Population | Description |
|---|---|
| Randomization Set | All participants who are randomized, regardless of the participants' treatment status in the study. |
| Full Analysis Set (FAS) | All randomized participants who received at least one dose of IP.<br>Participants will be analyzed according to the group to which they were randomized. |
| Modified Intent-to-Treat (mITT) Set | All participants in the FAS who had no immunologic or virologic evidence of prior COVID-19 (ie, negative NP swab test at Day 1, and/or bAb against SARS-CoV-2 nucleocapsid below LOD or LLOQ) at Day 1 before the first dose of IP.<br>Participants will be analyzed according to the group to which they were randomized. |
| Per-protocol (PP) Set | All participants in the mITT Set who received planned doses of IP per schedule and have no major protocol deviations, as determined and documented by Sponsor prior to database lock and unblinding, that impact critical or key study data.<br>Participants will be analyzed according to the group to which they were randomized. |
| Immunogenicity Subset | All participants in the FAS who were sampled into a subset for characterizing mRNA-1273 immunogenicity and had a valid immunogenicity test result prior to the first dose of IP and at least 1 valid result after the first dose of IP. Participants in the subset who had major protocol deviations that impact critical or key immunogenicity or study data may be excluded from the Immunogenicity Subset. The details of the Immunogenicity Subset will be documented prior to analysis of immunogenicity data. |
| Solicited Safety Set | The Solicited Safety Set consists of all randomized participants who received at least one dose of IP and contributed any solicited AR data.<br>The Solicited Safety Set will be used for the analyses of solicited ARs and participants will be included in the treatment group corresponding to the IP that they actually received. |
| Safety Set | All randomized participants who received at least one dose of IP. The Safety Set will be used for all analyses of safety except for the solicited ARs.<br>Participants will be included in the treatment group corresponding to the IP that they actually received. |

#### 9.5. Statistical Analyses

This section provides a summary of the planned statistical analyses of the primary and secondary endpoints.

The overall Type I error rate for the primary endpoint at the IAs and the primary analysis is strictly controlled at 2.5% (1-sided) based on the Lan-DeMets O'Brien-Fleming approximation spending function (see Section 9.6 for details). The primary efficacy results that will be considered statistically significant after consideration of the strategy for controlling the Type I error as described in Section 9.6. Statistical significance of the primary efficacy endpoint can be achieved at either one of the IAs or at the primary analysis. A sequential/hierarchical testing procedure will be used to control Type 1 error rate over the primary efficacy endpoint and the secondary efficacy endpoints. Secondary efficacy endpoints will only be tested when the primary efficacy endpoint achieves statistical significance. Multiplicity adjustments among the secondary efficacy endpoints may be performed for secondary efficacy endpoints, in that case, will be specified in the SAP.

No formal multiple comparison adjustments will be employed for multiple safety endpoints or multiple efficacy endpoints. Nominal p-values and confidence intervals (CIs) may be computed for other efficacy analyses without controlling for multiplicity as a measure of VE.

##### 9.5.1. Efficacy Analyses

Efficacy analyses will be performed using the FAS, mITT and PP populations, and participants will be included in the treatment group to which they are randomized. The primary analysis population will be the PP Set.

Table 9 summarizes the analysis approach for primary and secondary efficacy endpoints for Part A, the randomized, observer-blind, and placebo-controlled phase of the study. Sensitivity analysis methods are described for each endpoint as applicable.

**Table 9: Statistical Analysis Methods of Efficacy Endpoints**

| Endpoint | Statistical Analysis Methods |
|---|---|
| **Primary endpoint:**<br>Vaccine Efficacy (VE) of mRNA-1273 to prevent COVID-19 | • Primary analysis: VE will be estimated with 1 - HR (mRNA-1273 vs placebo) using a Cox proportional hazard regression model with treatment group as a fixed effect and adjust for stratification factor based on the PP Set, with cases counted starting 14 days after the second dose of IP.<br>• Analysis using the same model based on the mITT Set.<br>• Sensitivity analysis using the same model based on the PP Set, with cases counted starting either immediately after the second dose of IP or immediately after the first dose of IP. |

| Endpoint | Statistical Analysis Methods |
|---|---|
| | • Subgroup analysis of the primary efficacy endpoint will be performed to assess consistency of VE, such as in the age groups ≥ 18 and < 65 years and ≥ 65 years.<br>• Supportive analysis of VE to be estimated with 1 - ratio of incidence rates with 95% CI using the exact method conditional upon the total number of cases.<br>• Supportive analysis of cumulative incidence VE . |
| **Secondary endpoints:**<br>• Vaccine efficacy of mRNA-1273 to prevent severe COVID-19<br>• Vaccine efficacy of mRNA-1273 to prevent serologically confirmed SARS-CoV-2 infection or COVID-19 regardless of symptomatology or severity<br>• Vaccine efficacy of mRNA-1273 to prevent COVID-19 using a secondary definition of symptoms<br>• Vaccine efficacy of mRNA-1273 to prevent death due to COVID-19<br>• Vaccine efficacy of mRNA-1273 to prevent COVID-19 after the first dose of IP<br>• Vaccine efficacy of mRNA-1273 to prevent asymptomatic SARS-CoV-2 infection | Similar analysis method as for the primary endpoint analysis.<br>For each of the secondary endpoints:<br>• Primary analysis: VE will be estimated with 1 - HR (mRNA-1273 vs placebo) using a Cox proportional hazard regression model with treatment group as a fixed effect and adjusting for stratification factor based on the PP Set, with cases counted starting 14 days after the second dose of IP.<br>• Analysis using the same model based on the mITT Set.<br>• Sensitivity analyses with cases counted starting immediately after the second dose of IP, 14 days after the first dose of IP, immediately after the first dose of IP, and immediately after randomization.<br>• Vaccine efficacy and 95% CI based on the case incidence will be estimated with 1 - ratio of incidence rates using the exact method conditional upon the total number of cases. |
| • Vaccine efficacy of mRNA-1273 to prevent COVID-19 in all study participants, regardless of evidence of prior SARS-CoV-2 infection | The FAS population will be used for this secondary objective, using similar analysis methods as for the primary endpoint analysis.<br>• Primary analysis: VE will be estimated with 1 - HR (mRNA-1273 vs placebo) using a Cox proportional hazard regression model with treatment group as a fixed effect and adjusting for stratification factor based on the FAS, with cases counted starting 14 days after the second dose of IP.<br>• Sensitivity analyses with cases counted starting immediately after the second dose of IP, 14 days after the first dose of IP, immediately after the first dose of IP, and immediately after randomization. |

###### 9.5.1.1. Efficacy Analysis on Primary Endpoint

To assess the primary efficacy endpoint of VE of mRNA-1273 in preventing the first occurrence of COVID-19 from 14 days after second dose of IP, Cox proportional hazards regression will be used to estimate proportional hazards VE (PH VE), measured by one minus the HR (mRNA-1273 vs. placebo), with a 2-sided score-based 95% CI and 2-sided p-value for testing $H_0$: VE ≤ 30%.

Vaccine efficacy is defined as the percent reduction in the hazard of the primary endpoint (mRNA-1273 vs placebo). The VE will be estimated using one minus the HR (mRNA-1273 vs placebo) estimand. A stratified Cox proportional hazard model will be used to assess the magnitude of the treatment group difference (ie, HR) between mRNA-1273 and placebo. The HR and its 95% CI from the stratified Cox model with Efron's method of tie handling and with treatment group as covariate will be reported. The same stratification factors used for randomization will be applied to the stratified Cox model.

For the primary efficacy endpoints, participants without documented COVID-19 will be censored at the last study assessment date. Potential intercurrent events may include: 1) death unrelated to COVID-19 and 2) early COVID-19 up to 14 days after second study dose.

In the estimand of the primary analysis on the primary endpoint, a hypothetical strategy will be used for death unrelated to COVID-19; a hypothetical strategy will also be used for early COVID-19, where the time to COVID-19 will be censored at the onset day of early case. The details of intercurrent event description and estimand strategies are presented in Section 11.4.1.

For the primary efficacy analysis, cases will be counted starting 14 days after the second vaccination. Sensitivity analyses with cases counted immediately after the second vaccination, and after randomization will also be carried out.

Analyses of the primary endpoint will be also performed based on the mITT Set using the same methods described above.

For the primary efficacy analysis, cases will be counted starting 14 days after the second dose of IP. Sensitivity analyses with cases counted starting immediately after the second dose of IP and starting immediately after randomization will also be carried out.

Subgroup analysis of the primary efficacy endpoint will be performed in selected subgroups, such as age groups ≥18 and <65 year and ≥65 years to assess consistency of VE as described in Section 9.5.5.

As a supportive analysis, VE will also be estimated by one minus the infection rate ratio, where the number of cases (ie, participants with first occurrence of COVID-19) will be used and the CI will be computed using the exact method conditional upon the total number of cases. Cumulative incidence VE, one minus the ratio of cumulative incidences (mRNA-1273 vs placebo) of COVID--19, may also be assessed, the cumulative incidence for each arm will be estimated using a covariate adjustment method based on Zeng (2004) that makes use of baseline characteristics.

Additional analysis to evaluate VE against COVID-19 incorporating duration and presence/severity of symptoms will also be performed; the details will be provided in the SAP.

###### 9.5.1.2. Efficacy Analysis on Secondary Endpoint

For each of the below secondary objectives:

- Vaccine efficacy to prevent severe COVID-19
- Vaccine efficacy to prevent serologically confirmed SARS-CoV-2 infection or COVID-19 regardless of symptomology or severity
- Vaccine efficacy to prevent COVID-19 using a broad definition of symptoms
- Vaccine efficacy to prevent death caused by COVID-19
- Vaccine efficacy to prevent asymptomatic SARS-CoV-2 infection

For each of the above secondary objectives, the same Cox proportional hazard model described above for the primary objective will be applied using the PP Set, with cases counted starting 14 days after the second dose of IP. Sensitivity analyses with cases counted starting after the second dose of IP, 14 days after the first dose of IP, after the first dose of IP, and after randomization will also be performed.

The same model will be applied using the mITT population with cases counted starting 14 days after the second dose of IP.

Vaccine efficacy will be estimated with 1- ratio of incidence rates with the 95% CI using the exact method conditional upon the total number of cases.

Vaccine efficacy to prevent COVID-19 after the first dose of IP

The same Cox proportional hazard model described above for the primary objective will be applied using the PP Set, with cases counted starting 14 days after the first dose of IP.

Vaccine efficacy to prevent COVID-19 regardless of prior SARS-CoV-2 infection

The FAS will be used for analysis to evaluate VE to prevent COVID-19 regardless of prior SARS-CoV-2 infection. The same methods described above for the primary objective will be applied with cases counted starting 14 days after the second dose of IP. Sensitivity analyses with cases counted starting after the second dose of IP, 14 days after the first dose of IP, the first dose of IP, and randomization will also be performed.

Vaccine efficacy will be estimated with 1- ratio of incidence rates with the 95% CI using the exact method conditional upon the total number of cases.

###### 9.5.1.3. Long-term Efficacy Analysis

Long-term efficacy of the primary vaccination series with mRNA-1273 will be evaluated after the primary analysis by including data collected in the Open-Label Observational Phase (Part B). Long-term efficacy data will be summarized descriptively by treatment cohort described in Table 10 without cohort comparison. The long-term efficacy endpoints are listed inTable 11with the same case definitions specified in Table 1. In the primary approach, cases will be counted starting 14 days after the second dose of IP for participants in treatment cohorts of mRNA-1273 and Placebo or starting 14 days after the second dose of mRNA-1273 for participants in the Placebo-mRNA-1273 Cohort. Sensitivity analyses with cases starting from the second dose, 14 days after the first dose, or first dose of IP for participants in the cohorts of mRNA-1273 and placebo, or mRNA-1273 for participants in the Placebo-mRNA-1273 Cohort will be provided. Incidences of cases assessed by numbers, rates, and 2-sided 95% CI based on the exact method adjusting for person-time will be summarized by treatment cohort. The Kaplan-Meier analysis will be used to estimate cumulative incidences of time to first cases by treatment cohort. Long-term efficacy analysis will be performed using the PP Set and mITT Set.

Efficacy analyses of unblinded/open-label phase data collected in Part B will be provided, as will the incidence rate of COVID-19 after the BD of mRNA-1273. The details of analysis of long-term efficacy and open-label phase data (Part B and Part C) will be provided in the SAP.

**Table 10: Treatment Cohorts for the Long-term Efficacy Analyses**

| Treatment Cohort | Description |
|---|---|
| **mRNA-1273** | Participants randomized to mRNA-1273 in the Blinded Phase. |
| **Placebo** | Participants randomized to Placebo in the Blinded Phase and did not crossover to mRNA-1273 in the Open-Label Observational Phase. |
| **Placebo-mRNA-1273** | Participants randomized to Placebo in the Blinded Phase and crossed over to mRNA-1273 in the Open-Label Observational Phase. |

**Table 11: Long-term Efficacy Endpoints**

| Long-term Efficacy Endpoint |
|---|
| Cases of COVID-19 starting 14-days after the second injection of:<br>• IP for participants in the mRNA-1273 Cohort and the Placebo Cohort,<br>or<br>• mRNA-1273 for participants in the Placebo-mRNA-1273 Cohort. |

| **Long-term Efficacy Endpoint** |
|---|
| Cases of severe COVID-19 starting 14-days after the second injection of:<br>• IP for participants in the mRNA-1273 Cohort and the Placebo Cohort,<br>or<br>• mRNA-1273 for participants in the Placebo-mRNA-1273 Cohort. |
| Cases of either COVID-19 or SARS-CoV-2 infection starting 14-days after the second injection of:<br>• IP for participants in the mRNA-1273 Cohort and the Placebo Cohort,<br>or<br>• mRNA-1273 for participants in the Placebo-mRNA-1273 Cohort. |
| Cases with a secondary (less restrictive) definition of COVID-19 starting 14-days after the second injection of:<br>• IP for participants in the mRNA-1273 Cohort and the Placebo Cohort,<br>or<br>• mRNA-1273 for participants in the Placebo-mRNA-1273 Cohort. |
| Cases of death due to a cause directly attributed to a complication of COVID-19, starting 14-days after the second injection of:<br>• IP for participants in the mRNA-1273 Cohort and Placebo Cohort,<br>or<br>• mRNA-1273 for participants in the Placebo-mRNA-1273 Cohort. |
| Cases of COVID-19 starting 14-days after the first injection of:<br>• IP for participants in the mRNA-1273 Cohort and the Placebo Cohort,<br>or<br>• mRNA-1273 for participants in the Placebo-mRNA-1273 Cohort. |
| Regardless of evidence of prior SARS-CoV-2 infection determined by serologic titer against SARS-CoV-2 nucleocapsid, cases of COVID-19 starting 14-days after the second injection of:<br>• IP for participants in the mRNA-1273 Cohort and the Placebo Cohort,<br>or<br>• mRNA-1273 for participants in the Placebo-mRNA-1273 Cohort. |
| Cases of SARS-CoV-2 infection in the absence of symptoms defining COVID-19 starting 14-days after the second injection of:<br>• IP for participants in the mRNA-1273 Cohort and the Placebo Cohort,<br>or<br>• mRNA-1273 for participants in the Placebo-mRNA-1273 Cohort. |

##### 9.5.2. Safety Analyses

All safety analyses will be based on the Safety Set, except summaries of solicited ARs, which will be based on the Solicited Safety Set. All safety analyses will be provided by treatment group, and by treatment cohort as applicable, unless otherwise specified. Pregnancies and their known outcomes will be summarized (Section 8.3.6).

###### 9.5.2.1. Adverse Events

Safety and reactogenicity will be assessed by clinical review of all relevant parameters. Safety analyses will be provided for Parts A, B, and C separately unless specified otherwise.

The number and percentage of participants with any solicited local AR, with any solicited systemic AR, and with any solicited AR during the 7-day follow-up period after each dose will be provided only for the Blinded Phase (Part A). A 2-sided 95% exact CI using the Clopper-Pearson method will be also provided for the percentage of participants with any solicited AR for each treatment group.

Number and percentage of participants with unsolicited AEs, SAEs, MAAEs, Grade 3 or higher ARs and AEs, and AEs leading to discontinuation from IP or withdrawal from the study will be summarized. Unsolicited AE will be presented by MedDRA preferred term and system organ class.

Number of events of solicited ARs, unsolicited AEs/SAEs, and MAAEs will be reported in summarization tables accordingly.

For all other safety parameters, descriptive summary statistics will be provided, and Table 12 summarizes analysis strategy for safety parameters. Further details will be described in the SAP.

**Table 12: Analysis Strategy for Safety Parameters**

| Safety Endpoint | Number and Percentage of Participants, Number of Events | 95% CI |
|---|---|---|
| Any Solicited AR (Part A only; overall and by local, systemic) | X | X |
| Any Unsolicited AE | X | |
| Any SAE | X | |
| Any Unsolicited MAAE | X | |
| Any Unsolicited Treatment-Related AE | X | |
| Any Treatment-Related SAE | X | |
| Discontinuation due to AE | X | |
| Any Grade 3 and above AE | X | |
| Any Treatment-Related Grade 3 and above AE | X | |

Notes: 95% CI using the Clopper-Pearson method, X = results will be provided. Unsolicited AEs will be summarized by System Organ Class and Preferred Term coded by MedDRA.

###### 9.5.2.2. Baseline Descriptive Statistics

Demographic variables and baseline characteristics will be summarized by treatment group, and by treatment cohort as applicable, by descriptive statistics (mean, standard deviation for continuous variable, and number and percentage for categorical variables).

##### 9.5.3. Immunogenicity Analyses

The secondary immunogenicity endpoints will be analyzed using the Immunogenicity Subset by treatment group, by treatment cohort as applicable, and by baseline SARS-CoV-2 serostatus, unless otherwise specified. Details for immunogenicity analyses on data collected in the Open-Label Observational Phase will be provided in the SAP.

The SAP will describe the complete set of immunogenicity analyses, including the approach to sample individuals into an Immunogenicity Subset for characterizing mRNA-1273 immunogenicity and assessing immunological correlates of risk and protection.

Data from quantitative immunogenicity assays will be summarized for each treatment group using positive response rates and geometric means with 95% CIs, for each timepoint for which an assessment is performed. Data from qualitative (ie, yielding a positive or negative result) assays will be summarized by tabulating the frequency of positive responses for each assay by group at each timepoint that an assessment is performed. Analyses will focus on the 2 key immunogenicity time points and the change in marker response between them: Day 1 before the first dose of IP and Day 57 (28 days after the second dose of IP). The SAP will describe the complete set of immunogenicity analyses.

Quantitative levels or GMT of specific bAb with corresponding 95% CI at each timepoint and GMFR of specific bAb with corresponding 95% CI at each postbaseline timepoint over pre-dose baseline at Day 1 will be provided by study arm. Descriptive summary statistics including median, minimum, and maximum will also be provided.

GMT of specific nAb with corresponding 95% CI at each timepoint and GMFR of specific nAb with corresponding 95% CI at each postbaseline timepoint over pre-dose baseline at Day 1 will be provided by study arm. Descriptive summary statistics including median, minimum, and maximum will also be provided. For summarizations of group values, antibody values reported as below the LLOQ will be replaced by 0.5 × LLOQ. Values that are reported as greater than the ULOQ without the actual values will be converted to the ULOQ.

The number and percentage of participants with a fold rise ≥ 2, ≥ 3, and ≥ 4 of serum SARS-CoV-2-specific nAb titers and participants with seroresponse from baseline will be provided with 2-sided 95% CI using the Clopper-Pearson method at each post-baseline timepoint.

Seroresponse at a participant level may be defined as a change from below the LLOQ to at least 4 times the LLOQ, or at least a 4-fold rise in neutralizing antibody or vaccine antigen-specific bAb in participants with pre-existing nAb or bAb of at least the LLOQ at baseline. Seroresponse may also be defined for each specific assay assessing nAb or bAb. The definition of seroresponse will be finalized and documented in the SAP.

The GMT of specific nAb for each group and the geometric mean ratio (GMR) of mRNA-1273 versus placebo in Part A with corresponding 2-sided 95% CI will be estimated at each study timepoint using an analysis of covariance (ANCOVA) model with the treatment group and baseline values, if applicable, as explanatory variables, the analysis may adjust for the stratification factor (Table 13).

**Table 13: Immunogenicity Endpoints and Statistical Methods**

| Endpoint | Statistical Analysis Methods |
|---|---|
| Specific bAb and nAb titers/values | • GMT of each group, GMR (mRNA-1273 vs. Placebo)<br>• GMT estimated by the ANCOVA model |
| Fold rise | • GMFR - descriptive statistics<br>• Binomial endpoints of fold rise ≥2, 3, and 4, and seroconversion due to vaccination - the Clopper-Pearson method |

##### 9.5.4. Exploratory Analyses

The endpoint of viral infection kinetics will be assessed by determining the number of days until testing of saliva samples becomes negative after COVID-19 is established.

The endpoint of duration of symptoms will be assessed by determining the total number of days that a study participant with COVID-19 remains symptomatic through daily assessments after diagnosis.

Vaccine efficacy to prevent all-cause mortality will be assessed by similar analysis methods as used for the primary endpoint analysis, using PP Set, mITT Set, and FAS. All deaths, regardless of cause from the time of randomization, will be included. If the number of deaths becomes large enough to warrant analysis, the same Cox proportional hazard model described above for the primary objective will be applied using the PP Set, the mITT Set, and the FAS. Death, regardless of cause, from randomization will be included.

This endpoint of BOD is defined based on the post SARS-CoV-2 infection follow-up. A BOD score will be used to reflect the severity of symptoms with maximum score at COVID-19 death (Table 14).

These exploratory analyses will be based on the Blinded Phase of data (Part A). All other analyses of exploratory endpoints will be described in the SAP before database lock.

**Table 14: Burden of Disease Score**

| Patient State | BOD Score |
|---|---|
| Uninfected/Asymptomatic infection | 0 |
| Symptomatic without hospitalization | 1 |
| Hospitalization | 2 |
| Death | 3 |

##### 9.5.5. Subgroup Analyses

To determine whether the VE is consistent across various subgroups, the VE and its 95% CI may be estimated using the similar model within each category of the following classification variables.

- Age groups: ≥ 18, < 65, and ≥ 65
- Age and health risk for severe disease: ≥ 18 and < 65 and not at risk; ≥ 18 and < 65 and at risk, and ≥ 65
- Sex (female, male)
- Race
- At risk for severe COVID-19 illness (Section 6.2.1.1)

Subgroup analysis for the long-term efficacy may be performed descriptively by treatment cohort within above categories.

#### 9.6. Interim Analyses

In Part A, there are 2 planned IAs at 35% and 70% of total target cases across the 2 treatment groups. The primary objective of the IAs is for early detection of reliable evidence that VE is above 30%. The Lan-DeMets O'Brien-Fleming approximation spending function is used for calculating efficacy bounds and to preserve the (1-sided) 0.025 false positive error rate over the IAs and the primary analysis (when the target number of cases have been observed), relative to the hypothesis:

$H_0^{efficacy}$: HR $\geq$ 0.7 (equivalently, proportional hazards VE $\leq$ 0.3).

There is no intention to stop the study early if the efficacy has been demonstrated at any of the IAs. If efficacy is demonstrated at an IA, the subsequent IA or primary analysis will be considered supportive in nature. The DSMB will review the IA results and make recommendations to the Sponsor in terms of study results reporting and unblinding based on the boundaries of early efficacy as described in this section, safety data, and data external to this study. In addition to possible early efficacy at IAs, the DSMB will monitor for non-efficacy and vaccine harm; the guiding principles (non-binding) is provided in Section 8.4.2, and the details will be provided in the SAP.

Table 15 summarizes the timing, number of cases and decision guidance at each IA and primary analysis.

The first IA will occur when approximately 35% of the total cases have been observed (across both treatment groups). The study will be considered positive at the first IA if the p-value for rejecting HR $\geq$ 0.7 is less than 0.0002 based on the Lan-DeMets O'Brien-Fleming approximation spending function. This corresponds to an observed HR of approximately 0.259, or an observed VE approximately 0.741.

The second IA will occur when approximately 70% of the total cases have been observed. The study will be considered positive (VE has been demonstrate) if the p-value for rejecting HR $\geq$ 0.7 is less than 0.0073 based on the Lan-DeMets O'Brien-Fleming approximation spending function. This corresponds to an observed HR of approximately 0.435, or an observed VE of approximately 0.565.

The primary analysis will be performed when approximately 151 cases have been observed in the study. The study will be considered positive at the primary analysis if the 1-sided p-value for rejecting HR $\geq$ 0.7 is less than 0.0227. This corresponds to an observed HR of approximately 0.505 or observed VE of approximately 0.495.

**Table 15: Interim Boundaries Using O'Brien-Fleming Spending Function, Calculation Based on the PP Set for the Primary Efficacy Endpoint**

| Information Fraction (% of total #cases) | Number of Cases | Nominal Alpha | Efficacy Boundary Rejecting H0: VE ≤ 30% | Cum Prob (crossing efficacy boundary if the true VE = 60%) |
|---|---|---|---|---|
| IA1 35% | 53 | 0.0002 | VE ≥ 0.741<br>(HR ≤ 0.259) | 4.6% |
| IA2 70% | 106 | 0.0073 | VE ≥ 0.565<br>(HR ≤ 0.435) | 61.5% |
| Primary analysis 100% | 151 | 0.0227 | VE ≥ 0.495<br>(HR ≤ 0.505) | 90.0% |

Abbreviations: HR = hazard ratio; IA: interim analysis; LB = lower boundary; PP = per-protocol; VE = vaccine efficacy.

### 10. REFERENCES

Adler Y, Charron P, Imazio M, Badano L, Barón-Esquivias G, Bogaert J, et al. 2015 ESC guidelines for the diagnosis and management of pericardial diseases: the task force for the diagnosis and management of pericardial diseases of the european society of cardiology (ESC) endorsed by: The European Association for Cardio-Thoracic Surgery (EACTS). Eur Heart J. 2015;36(42):2921-64.

Anderson K. gsDesign, version 3.1.1. 2020. Available from The Comprehensive R Archive Network at https://cran.r-project.org/web/packages/gsDesign/.

Aretz HT. Myocarditis: the Dallas criteria. Hum Pathol. 1987 ;18(6):619-24.

Baden LR, El Sahly HM, Essink B, Kotloff K, Frey S, Novak R, et al. Efficacy and safety of the mRNA-1273 SARS-CoV-2 vaccine. N Engl J Med. 2020. doi: 10.1056/NEJMoa2035389. Online ahead of print.

Centers for Disease Control and Prevention (CDC). 2020a. Coronavirus Disease 2019 (COVID-19). Retrieved from: https://www.cdc.gov/coronavirus/2019-ncov/cases-updates/cases-in-us.html. (accessed 2020 May 29).

Centers for Disease Control and Prevention (CDC). 2020b. Coronavirus Disease 2019 (COVID-19). Retrieved from: https://www.cdc.gov/coronavirus/2019-ncov/need-extra-precautions/people-at-higher-risk.html (accessed 2020 May 29).

Centers for Disease Control and Prevention (CDC). 2020c. Coronavirus Disease 2019 (COVID-19). Retrieved from: https://www.cdc.gov/coronavirus/2019-ncov/symptoms-testing/symptoms.html (accessed 2020 Jun 11).

Centers for Disease Control and Prevention (CDC). 2020d. Coronavirus Disease 2019 (COVID-19). Retrieved from: https://www.cdc.gov/coronavirus/2019-ncov/need-extra-precautions/people-with-medical-conditions.html. (accessed 2020 July 28).

Chen Y, Lu S, Jia H, Deng Y, Zhou J, Huang B, et al. A novel neutralizing monoclonal antibody targeting the N-terminal domain of the MERS-CoV spike protein. Emerg Microbes Infect. 2017 May 24;6(5):e37.

Chin J, Magoffin RL, Shearer LA, Schieble JH, Lennette EH. Field evaluation of a respiratory syncytial virus vaccine and a trivalent parainfluenza virus vaccine in a pediatric population. Am J Epidemiol 1969;89:449–63

ClinicalTrials.gov [Internet]. Bethesda (MD): National Library of Medicine (US). - Identifier NCT03076385, Safety, tolerability, and immunogenicity of VAL-506440 in healthy adult subjects. Retrieved from: https://clinicaltrials.gov/ct2/keydates/NCT03076385. (accessed 2020 Jun 01).

ClinicalTrials.gov [Internet]. Bethesda (MD): National Library of Medicine (US). - Identifier NCT03325075, Safety, tolerability, and immunogenicity of VAL-181388 in healthy subjects. Retrieved from: https://clinicaltrials.gov/ct2/keydates/NCT03325075. (accessed 2020 Jun 01).

ClinicalTrials.gov [Internet]. Bethesda (MD): National Library of Medicine (US). - Identifier NCT03345043, Safety, tolerability, and immunogenicity of VAL-339851 in healthy adult subjects. Retrieved from: https://clinicaltrials.gov/ct2/keydates/NCT03345043. (accessed 2020 Jun 01).

ClinicalTrials.gov [Internet]. Bethesda (MD): National Library of Medicine (US). - Identifier NCT03382405, Safety, reactogenicity, and immunogenicity of cytomegalovirus vaccines mRNA-1647 and mRNA-1443 in healthy adults. Retrieved from: https://clinicaltrials.gov/ct2/keydates/NCT03382405. (accessed 2020 Jun 01).

ClinicalTrials.gov [Internet]. Bethesda (MD): National Library of Medicine (US). - Identifier NCT03392389, Safety, reactogenicity, and immunogenicity of mRNA-1653 in healthy adults. Retrieved from: https://clinicaltrials.gov/ct2/keydates/NCT03392389. (accessed 2020 Jun 01).

ClinicalTrials.gov [Internet]. Bethesda (MD): National Library of Medicine (US). - Identifier NCT04283461, Safety and immunogenicity study of SARS-CoV-2 vaccine (mRNA-1273) to treat novel coronavirus; 2020 Feb 22. Retrieved from: https://clinicaltrials.gov/ct2/keydates/NCT04283461. (accessed 2020 Jun 01).

See draft guidance for industry Enhancing the Diversity of Clinical Trial Populations - Eligibility Criteria, Enrollment Practices, and Trial Designs Guidance for Industry (June 2019), available at https://www.fda.gov/media/127712/download.

Corti D, Zhao J, Pedotti M, Simonelli L, Agnihothram S, Fett C, et al. Prophylactic and postexposure efficacy of a potent human monoclonal antibody against MERS coronavirus. Proc Natl Acad Sci U S A. 2015 Aug 18;112(33):10473-8.

Department of Health and Human Services (DHHS), Food and Drug Administration, Center for Biologics Evaluation and Research (US). Guidance Document. FDA guidance on conduct of clinical trials of medical products during COVID-19 public health emergency. March 2020. Updated on April 16, 2020. [cited 2020 Apr 17]. Available from: https://www.fda.gov/media/136238/download.

Department of Health and Human Services (DHHS), Food and Drug Administration, Center for Biologics Evaluation and Research (US). Guidance for industry: Toxicity grading scale for healthy adult and adolescent volunteers enrolled in preventative vaccine clinical trials. September 2007 [cited 2019 Apr 10] [10 screens]. Available from:

https://www.fda.gov/downloads/BiologicsBloodVaccines/GuidanceComplianceRegulatoryInformation/Guidances/Vaccines/ucm091977.pdf.

Feldman RA, Fuhr R, Smolenov I, Mick Ribeiro A, Panther L, Watson M, et al. mRNA vaccines against H10N8 and H7N9 influenza viruses of pandemic potential are immunogenic and well tolerated in healthy adults in phase 1 randomized clinical trials. Vaccine. 2019 May 31;37(25):3326-3334.

Ferreira VM, Schulz-Menger J, Holmvang G, Kramer CM, Carbone I, Sechtem U, et al. Cardiovascular magnetic resonance in nonischemic myocardial inflammation: expert recommendations. J Am Coll Cardiol. 2018;72(24):3158-76.

Fulginiti VA, Eller JJ, Downie AW, Kempe CH. Altered reactivity to measles virus. Atypical measles in children previously immunized with inactivated measles virus vaccines. JAMA. 1967 202:1075-80.

Gargano JW, Wallace M, Hadler SC, Langley G, Su JR, Oster ME, et al. Use of mRNA COVID-19 vaccine after reports of myocarditis among vaccine recipients: update from the advisory committee on immunization practices - United States, June 2021. MMWR Morb Mortal Wkly Rep. 2021;70(27):977-82.

Jackson LA, Anderson EJ, Rouphael NG, Roberts PC, Makhene M, Coler RN, et al. An mRNA vaccine against SARS-CoV-2 – preliminary report. N Engl J Med. 2020;383(20):1920-31.

Johnson RF, Bagci U, Keith L, Tang X, Mollura DJ, Zeitlin L, et al. 3B11-N, a monoclonal antibody against MERS-CoV, reduces lung pathology in rhesus monkeys following intratracheal inoculation of MERS-CoV Jordan-n3/2012. Virology. 2016 Mar;490:49-58.

Kim Y, Lee H, Park K, Park S, Lim JH, So MK, et al. Selection and characterization of monoclonal antibodies targeting middle east respiratory syndrome coronavirus through a human synthetic fab phage display library panning. Antibodies (Basel). 2019 Jul 31;8(3).

Rüggeberg JU, Gold MS, Bayas JM, Blum MD, Bonhoeffer J, Friedlander S, et al. Anaphylaxis: case definition and guidelines for data collection, analysis, and presentation of immunization safety data. Vaccine. 2007;25(31):5675-84.

Thomas SJ and Yoon IK. A review of Dengvaxia®: development to deployment. Hum Vaccin Immunother. 2019;15(10), 2295-2314.

Wang L, Shi W, Joyce MG, Modjarrad K, Zhang Y, Leung K, et al. Evaluation of candidate vaccine approaches for MERS-CoV. Nat Commun. 2015 Jul 28;6:7712.

Wang L, Shi W, Chappell JD, Joyce MG, Zhang Y, Kanekiyo M, et al. Importance of neutralizing monoclonal antibodies targeting multiple antigenic sites on the middle east respiratory syndrome coronavirus spike glycoprotein to avoid neutralization escape. J Virol. 2018 Apr 27;92(10).

Widjaja I, Wang C, van Haperen R, Gutiérrez-Álvarez J, van Dieren B, Okba NMA, et al. Towards a solution to MERS: protective human monoclonal antibodies targeting different domains and functions of the MERS-coronavirus spike glycoprotein. Emerg Microbes Infect. 2019;8(1):516-30.

World Health Organization (WHO). Dengue vaccine: WHO position paper, September 2018 – recommendations. Vaccine. 2019 Aug;37(35):4848-4849

World Health Organization (WHO). 2020 (Data reported as of 2020, May 28). Coronavirus disease 2019 (COVID-19) Situation Report – 129. Retrieved from: https://www.who.int/docs/default-source/coronaviruse/situation-reports/20200528-covid-19-sitrep-129.pdf?sfvrsn=5b154880_2. (accessed 2020 May 29).

Wrapp D, Wang N, Corbett KS, Goldsmith JA, Hseih CL, Abiona O, et al. Cryo-EM structure of the 2019-nCoV spike in the prefusion conformation. Science. 2020;367:1260-3.

Yu X, Zhang S, Jiang L, Cui Y, Li D, Wang D, et al. Structural basis for the neutralization of MERS-CoV by a human monoclonal antibody MERS-27. Sci Rep. 2015 Aug 18;5:13133.

Zeng D. 2004. Estimating marginal survival function by adjusting for dependent censoring using many covariates. The Annals of Statistics, 32(4), 1533-1555.

Zent O, Arras-Reiter C, Broeker M, Hennig R. Immediate allergic reactions after vaccinations – a post-marketing surveillance review. Eur J Pediatr. 2002;161(1):21-25.

### 11. SUPPORTING DOCUMENTATION AND OPERATIONAL CONSIDERATIONS

#### 11.1. APPENDIX 1: Schedules of Events

If a participant cannot attend a study site visit (scheduled or unscheduled, with the exception of Screening, Day 1, Day 29, PDV, OL-D1, OL-D29, and BD-1), a home visit is acceptable if performed by appropriately delegated study site staff or a home healthcare service provided by the Sponsor. If neither a participant visit to the study site nor a home visit to the participant is possible (with the exception of Screening, Day 1, Day 29, PDV, OL-D1, OL-D29, and BD-1), a safety phone call should be performed that includes the assessments scheduled for the biweekly safety phone calls (Table 16).

The Supplemental SoEs (Table 21 and Table 23) and the Modified Supplemental SoE (Table 22) are intended to occur in addition to the original SoEs, as applicable (Table 16, Table 17Table 18Table 19), and therefore there is a possibility for study visits to overlap. If visits overlap according to respective visit windows, a single visit may be completed with any duplicated study procedures each completed once (refer to Table 4 footnotes for more detailed instructions and exceptions).

**Table 16: Schedule of Events (Vaccination Phase, Day 1 – Day 57)**

| **Visit Number** | 0 | 1 | | | | 2 | | | | 3 |
|---|---|---|---|---|---|---|---|---|---|---|
| **Type of Visit** | C | C | SC | SC | SC | C | SC | SC | SC | C |
| **Month/Weekly Timepoint** | | M0 | | | | M1 | | | | M2 |
| **Study Visit Day** | D0[1] (Screening) | D1 (Baseline) | 7 days after D1 D8 | 14 days after D1 D15 | 21 days after D1 D22 | D29 | 7 days after D29 D36 | 14 days after D29 D43 | 21 days after D29 D50 | D57 |
| **Window Allowance (Days)** | -28 | | +3 | +3 | +3 | -3/+7 | +3 | +3 | +3 | -3/+7 |
| **Days Since Most Recent Vaccination** | - | 0 | 7 | 14 | 21 | 28/0 | 7 | 14 | 21 | 28 |
| ICF, demographics, concomitant medications, medical history | X | | | | | | | | | |
| Confirm participant meets inclusion and exclusion criteria | X | X | | | | | | | | |
| Physical examination[2] | X | X | | | | X | | | | X |
| Pregnancy testing[3] | X | X | | | | X | | | | |
| Randomization | | X | | | | | | | | |
| **Dosing** | | | | | | | | | | |
| Study injection (including 30-minute post-dosing observation period) | | X | | | | X | | | | |
| **Efficacy Assessment** | | | | | | | | | | |
| Surveillance for COVID-19/Unscheduled Visit[4] | | X | X | X | X | X | X | X | X | X |
| Nasopharyngeal swab[5] | | X | | | | X | | | | |
| **Immunogenicity Assessment** | | | | | | | | | | |
| Blood for immunologic analysis[5] | | X | | | | X | | | | X |

| Visit Number | 0 | 1 | | | | 2 | | | | 3 |
|---|---|---|---|---|---|---|---|---|---|---|
| Type of Visit | C | C | SC | SC | SC | C | SC | SC | SC | C |
| Month/Weekly Timepoint | | M0 | | | | M1 | | | | M2 |
| **Study Visit Day** | D0[1] (Screening) | D1 (Baseline) | 7 days after D1 D8 | 14 days after D1 D15 | 21 days after D1 D22 | D29 | 7 days after D29 D36 | 14 days after D29 D43 | 21 days after D29 D50 | D57 |
| **Window Allowance (Days)** | -28 | | +3 | +3 | +3 | -3/+7 | +3 | +3 | +3 | -3/+7 |
| **Days Since Most Recent Vaccination** | - | 0 | 7 | 14 | 21 | 28/0 | 7 | 14 | 21 | 28 |
| **Safety Assessments** | | | | | | | | | | |
| eDiary activation for recording solicited adverse reactions (7 days)[6] | | X | | | | X | | | | |
| Review of eDiary | | | X | | | | X | | | |
| Follow-up Safety[7] | | | X | X | X | | X | X | X | |
| Recording of Unsolicited AEs | | X | X | X | X | X | X | X | X | X |
| Recording of MAAEs, AESIs, AE leading to withdrawal and concomitant medications relevant to or for the treatment of the MAAE[8] | | X | X | X | X | X | X | X | X | X |
| Recording of SAEs and concomitant medications relevant to or for the treatment of the SAE[8] | X | X | X | X | X | X | X | X | X | X |
| Recording of concomitant medications and non-study vaccinations[8] | | X | X | X | X | X | X | X | X | X |

Abbreviations: AE = adverse event; C = clinic visit; D = day; eDiary= electronic diary; ICF = informed consent form; M = month; MAAE = medically attended AE; SAE = serious adverse event; SC = safety (phone) call.

Note: In accordance with FDA Guidance on Conduct of Clinical Trials of Medical Products during the COVID-19 Pandemic (DHHS 2020), investigators may convert study site visits to telemedicine visits with the approval of the Sponsor. All scheduled study visits should be completed within the respective visit windows. If the participant is not able to come on-site for a study site visit as a result of the COVID-19 Pandemic (self-quarantine or disruption of study site activities following business continuity plans and/or local government mandates for "stay at home" or "shelter in place"), a safety call to the participant should be made in place of the study site visit. The safety call should encompass all scheduled visit assessments that can be completed remotely, such as assessment for AEs and concomitant medications (eg, as defined in scheduled biweekly safety phone calls). Home visits will be permitted for all non-dosing visits except for Screening if a participant cannot come to the study site as a result of the COVID19 pandemic.

1. Day 0 and Day 1 may be combined the same day. Additionally, the Day 0 visit may be performed over multiple visits if within the 28-day screening window.
2. Physical examination: a full physical examination, including vital signs, height, and weight, will be performed at Screening and Day 1. Body mass index will be calculated at the Screening Visit (Day 0) only. Symptom-directed physical examinations will be performed on Day 29 and Day 57. On each dosing day before injection, the arm receiving the injection should be examined and the associated lymph nodes should be evaluated. Any clinically significant finding identified during a study visit should be reported as a MAAE. Vital signs are to be collected pre- and post-dosing on days of injection (Day 1 and Day 29). When applicable, vital sign measurements should be performed before blood collection. Participants who are febrile (body temperature ≥ 38.0°C/100.4°F) before dosing on Day 1 or Day 29 must be rescheduled within the relevant window period to receive the injection. Afebrile participants with minor illnesses can be enrolled at the discretion of the investigator.
3. Pregnancy test at Screening and Day 1 and before the second vaccination will be a point-of-care urine test. At the discretion of the investigator a pregnancy test either via blood or point-of-care urine test can be performed. Follicle-stimulating hormone level may be measured to confirm menopausal status at the discretion of the investigator.
4. Participants with symptoms of COVID-19 will be asked to return within 72 hours or as soon as possible to the study site for an unscheduled visit, to include an NP swab sample (for RT-PCR testing) and other clinical evaluations. If a study site visit is not possible, a home visit may be arranged to collect the NP swab sample and conduct clinical evaluations. If a home visit is not possible, the participant will be asked to submit a saliva sample to the study site by a Sponsor-approved method. At this visit, an NP swab and blood sample will be collected to evaluate for the presence of SARS-CoV-2 infection. An additional NP swab sample will be collected to test for the presence of other respiratory pathogens. Additionally, clinical information will be carefully collected to evaluate the severity of the clinical case. It is important to note that some of the symptoms of COVID-19 overlap with solicited systemic ARs that are expected after vaccination with mRNA-1273 (eg, myalgia, headache, fever, and chills). During the first 7 days after vaccination, when these solicited ARs are common, investigators should use their clinical judgement to decide if an NP swab should be collected. The collection of an NP swab prior to the Day 1 and Day 29 vaccination can help ensure that cases of COVID-19 are not overlooked. Any study participant reporting respiratory symptoms during the 7-day period after vaccination should be evaluated for COVID-19.
5. Sample must be collected prior to dosing on days of injection (Day 1 and Day 29).
6. The participant will record entries in the eDiary approximately 30 minutes after dosing while at the study site, with instruction provided by study staff. Study participants will continue to record in the eDiary each day after they leave the study site, preferably in the evening, on the day of dosing and for 6 days following. Any solicited AR that is ongoing beyond Day 7 will be reported until resolution. Adverse reactions recorded in eDiaries beyond Day 7 should be reviewed either via phone call or at the following study visit. Participants will be given thermometers to record their temperatures and rulers to measure any injection site reactions.
7. Trained study personnel will call all participants to collect information relating to any unsolicited AEs through Day 57 (including any signs and symptoms of COVID-19), MAAEs, AESIs, AEs leading to withdrawal, SAEs, information on concomitant medications associated with those events, and any non-study vaccinations.
8. All concomitant medications and non-study vaccinations will be recorded through 28 days after each injection; all concomitant medications relevant to or for the treatment of an SAE or MAAE will be recorded from Screening through the final visit (Day 759).

**Table 17: Schedule of Events (Surveillance Phase, Day 64 – Day 394)**

| **Visit Number** | | | | | | 4 | | | | | | 5 |
|---|---|---|---|---|---|---|---|---|---|---|---|---|
| **Type of Visit** | eD | SC | SC | SC | SC | C | SC | SC | SC | SC | SC | C |
| **Month (M)/Weekly (W) Timepoint** | W | M3 | M4 | M5 | M6 | M7 | M8 | M9 | M10 | M11 | M12 | M13 |
| **Study Visit Day** | D64 [1] | D85[1] | D119 [1] 90 days after D29 | D149[1] | D179[1] | D209[1] 180 days after D29 | D239[1] | D269[1] | D299[1] | D329[1] | D359[1] | D394[1] 365 days after D29 |
| **Window Allowance (Days)** | ±2 | ±3 | ±3 | ±3 | ±3 | ±14 | ±3 | ±3 | ±3 | ±3 | ±3 | ±14 |
| **Days Since Most Recent Vaccination** | - | 56 | 90 | 120 | 150 | 180 | 210 | 240 | 270 | 300 | 330 | 365 |
| Physical examination[2] | | | | | | X | | | | | | X |
| **Efficacy Assessments** | | | | | | | | | | | | |
| eDiary activation for surveillance for COVID-19/Unscheduled Visit | X | | | | | | | | | | | |
| eDiary Weekly prompts for surveillance for COVID-19 | □-------------------Weekly eDiary prompts (Day 64 through Day 392) -----------□ |  |  |  |  |  |  |  |  |  |  |  |
| Surveillance for COVID-19/Unscheduled Visit[3] | | X | X | X | X | X | X | X | X | X | X | X |
| **Immunogenicity Assessment** | | | | | | | | | | | | |
| Blood for immunologic analysis | | | | | | X | | | | | | X |
| **Safety Assessments** | | | | | | | | | | | | |
| eDiary activation for safety follow-up | X[4] | | | | | | | | | | | |
| eDiary Weekly prompts for safety follow-up | □--------------Weekly eDiary prompts (Day 64 through Day 387) ----------------□ |  |  |  |  |  |  |  |  |  |  |  |

| Visit Number | | | | | | 4 | | | | | | 5 |
|---|---|---|---|---|---|---|---|---|---|---|---|---|
| **Type of Visit** | eD | SC | SC | SC | SC | C | SC | SC | SC | SC | SC | C |
| **Month (M)/Weekly (W) Timepoint** | W | M3 | M4 | M5 | M6 | M7 | M8 | M9 | M10 | M11 | M12 | M13 |
| **Study Visit Day** | D64 [1] | D85[1] | D119 [1] 90 days after D29 | D149[1] | D179[1] | D209[1] 180 days after D29 | D239[1] | D269[1] | D299[1] | D329[1] | D359[1] | D394[1] 365 days after D29 |
| **Window Allowance (Days)** | ±2 | ±3 | ±3 | ±3 | ±3 | ±14 | ±3 | ±3 | ±3 | ±3 | ±3 | ±14 |
| **Days Since Most Recent Vaccination** | - | 56 | 90 | 120 | 150 | 180 | 210 | 240 | 270 | 300 | 330 | 365 |
| Follow-up safety[5] | | X | X | X | X | X | X | X | X | X | X | X |
| Recording of MAAEs, AESIs, AE leading to withdrawal and concomitant medications relevant to or for the treatment of the MAAE[6] | | X | X | X | X | X | X | X | X | X | X | X |
| Recording of SAEs and concomitant medications relevant to or for the treatment of the SAE[6] | | X | X | X | X | X | X | X | X | X | X | X |
| Recording of concomitant medications and non-study vaccinations[6,7] | | X | X | X | X | X | X | X | X | X | X | X |

Abbreviations: AE = adverse event; C = clinic visit; D = day; eD = electronic diary; IRB = institutional review board; M = month; MAAE = medically attended AE; SAE = serious adverse event; SC = safety (phone) call; W = week.

Note: In accordance with FDA Guidance on Conduct of Clinical Trials of Medical Products during the COVID-19 Pandemic (DHHS 2020), investigators may convert study site visits to telemedicine visits with the approval of the Sponsor.

1. All scheduled study visits should be completed within the respective visit windows. If the participant is not able to come on-site for a study site visit as a result of the COVID-19 pandemic (self-quarantine or disruption of study site activities following business continuity plans and/or local government mandates for “stay at home” or “shelter in place”), a safety phone call to the participant should be made in place of the study site visit. The safety call should encompass all scheduled visit assessments that can be completed remotely, such as assessment for AEs and concomitant medications (eg, as defined in scheduled safety phone calls). Home visits will be permitted for all non-dosing visits except for Screening if a participant cannot come to the study site as a result of the COVID-19 pandemic. Home visits must be permitted by the site IRB and the participant via informed consent and have prior approval from the Sponsor (or its designee).
2. Symptom-directed physical examinations may be performed at the discretion of the investigator.

3. Participants with symptoms of COVID-19 will be asked to return within 72 hours or as soon as possible to the study site for an unscheduled visit, to include an NP swab sample (for RT-PCR testing) and other clinical evaluations. If a study site visit is not possible, a home visit may be arranged to collect the NP swab sample and conduct clinical evaluations. If a home visit is not possible, the participant will be asked to submit a saliva sample to the study site by a Sponsor-approved method. At this visit, an NP swab and blood sample will be collected to evaluate for the presence of SARS-CoV-2 infection. An additional NP swab sample will be collected to test for the presence of other respiratory pathogens. In addition, the study site may collect an additional (local/non-central) respiratory sample for SARS-CoV-2 testing to be able to render appropriate medical care for the study participant as determined by local standards of care. Additionally, clinical information will be carefully collected to evaluate the severity of the clinical case.
4. The eDiary Safety Follow-up prompts will be triggering off of Day 61 to take into consideration the (-3 day) window allowance at Day 29. This first Safety Follow-Up prompt at D61 will follow the same 7-day surveillance per protocol.
5. Trained study personnel will call all participants to collect information relating to any sign/symptoms of COVID-19, MAAEs, AESIs, AEs leading to withdrawal, SAEs, information on concomitant medications associated with those events, and any non-study vaccinations.
6. All concomitant medications relevant to or for the treatment of an SAE or MAAE will be recorded from Screening through the final visit (Day 759).
7. One additional ePRO prompt will be sent to participants specifically to solicit the collection of information regarding participant’s history of facial injections or dermal fillers, for cosmetic or medical indications such as migraine headaches.

##### Table 18: Schedule of Events (Surveillance Phase, Day 401 – Day 759)

| Visit Number | | | | | | 6 |
|---|---|---|---|---|---|---|
| **Type of Visit** | SC | SC | SC | SC | SC | C |
| **Month (M)/Weekly (W) Timepoint** | M15 | M17 | M19 | M21 | M23 | M25 |
| **Study Visit Day** | D454[1] | D514[1] | D574[1] | D634[1] | D694[1] | D759[1] 365 days after Year 1 |
| **Window Allowance (Days)** | ±3 | ±3 | ±3 | ±3 | ±3 | ±14 |
| **Days Since Most Recent Vaccination** | | | | | | 730 |
| Physical examination[2] | | | | | | X |
| eDiary Weekly prompts for surveillance for COVID-19 | Weekly eDiary prompts (Day 401 through Day 759) | | | | | |
| Surveillance for COVID-19/Unscheduled Visit[3] | X | X | X | X | X | X |
| Blood for immunologic analysis | | | | | | X |
| eDiary Weekly prompts for safety follow-up | Weekly eDiary prompts (Day 401 through Day 752) | | | | | |
| Follow-up Safety[4] | X | X | X | X | X | X |
| Recording of MAAEs, AESIs, AE leading to withdrawal and concomitant medications relevant to or for the treatment of the MAAE[5] | X | X | X | X | X | X |
| Recording of SAEs and concomitant medications relevant to or for the treatment of the SAE[5] | X | X | X | X | X | X |
| Recording of concomitant medications and non-study vaccinations[5] | X | X | X | X | X | X |

Abbreviations: AE = adverse event; C = clinic visit; D = day; eD = electronic diary; IRB = institutional review board; M = month; MAAE = medically attended AE; SAE = serious adverse event; SC = safety (phone) call.

Note: In accordance with FDA Guidance on Conduct of Clinical Trials of Medical Products during the COVID-19 Pandemic (DHHS 2020), investigators may convert study site visits to telemedicine visits with the approval of the Sponsor.

1. All scheduled study visits should be completed within the respective visit windows. If the participant is not able to come on-site for a study site visit as a result of the COVID-19 Pandemic (self-quarantine or disruption of study site activities following business continuity plans and/or local government mandates for "stay at home" or "shelter in place"), a safety phone call to the participant should be made in place of the study site visit. The safety phone call should encompass all scheduled visit assessments that can be completed remotely, such as assessment for AEs and concomitant medications (eg, as defined in scheduled safety phone calls). Home visits will be permitted for all non-dosing visits except for Screening if a participant cannot come to the study site as a result of the COVID-19 pandemic. Home visits must be permitted by the site IRB and the participant via informed consent and have prior approval from the Sponsor (or its designee).
2. Symptom-directed physical examinations may be performed at the discretion of the investigator.
3. Participants with symptoms of COVID-19 will be asked to return within 72 hours or as soon as possible to the study site for an unscheduled visit, to include an NP swab sample (for RT-PCR testing) and other clinical evaluations. If a study site visit is not possible, a home visit may be arranged to collect the NP swab sample and conduct clinical evaluations. If a home visit is not possible, the participant will be asked to submit a saliva sample to the study site by a Sponsor-approved method. At this visit, an NP swab and blood sample will be collected to evaluate for the presence of SARS-CoV-2 infection. An additional NP swab sample will be collected to test for the presence of other respiratory pathogens. In addition, the study site may collect an additional (local/non-central) respiratory sample for SARS-CoV-2 testing to be able to render appropriate medical care for the study participant as determined by local standards of care. Additionally, clinical information will be carefully collected to evaluate the severity of the clinical case.
4. Trained study personnel will call all participants to collect information relating to any MAAEs, AESIs, AEs leading to withdrawal, SAEs, information on concomitant medications associated with those events, and any non-study vaccinations.
5. All concomitant medications relevant to or for the treatment of an SAE or MAAE will be recorded from Screening through the final visit (Day 759).

##### Table 19: Schedule of Events (Convalescent Period, Starting with the Illness Visit)

| **Unscheduled Visit** | 1 | | | | | | | | 2 |
|---|---|---|---|---|---|---|---|---|---|
| **Type of Visit** | C/H | | | | | | | | C/H |
| **Daily Timepoint** | D1 | D2- D6 | D7 | D8-D13 | D14 | D15-D20 | D21 | D22-D27 | D28 |
| **Window Allowance (Days)** | - | ±1 | ±1 | ±1 | ±1 | ±1 | ±1 | ±1 | +7 |
| **Safety Assessments** | | | | | | | | | |
| Symptom-directed physical examination[1] | X | | | | | | | | X |
| Follow-up safety[2] | X | | | | | | | | X |
| Recording of MAAEs, AESIs, AE leading to withdrawal and concomitant medications relevant to or for the treatment of the MAAE[3] | X | | | | | | | | X |
| Recording of SAEs and concomitant medications relevant to or for the treatment of the SAE[3] | X | | | | | | | | X |
| Recording of concomitant medications and non-study vaccinations[3] | X | | | | | | | | X |
| **Efficacy Assessments** | | | | | | | | | |
| Daily Telemedicine visit[4] |  | ←----------Daily----------→ |  |  |  |  |  |  |  |
| Respiratory illness sample[5] | X | | | | | | | | |
| Blood sample for immunologic assessment of SARS-CoV-2 infection[6] | X | | | | | | | | X |
| Saliva sample | | D3[7], D5[7] | D7[7] | D9[7] | D14[7] | | D21[7] | | X[8] |

Abbreviations: C = clinic visit; CLIA = Clinical Laboratory Improvement Amendments; D = day; H = home visit; MAAE = medically attended AE; SAE = serious adverse event

Note: In accordance with FDA Guidance on Conduct of Clinical Trials of Medical Products during the COVID-19 Pandemic (DHHS 2020), investigators may convert study site visits to telemedicine visits with the approval of the Sponsor.

1. Physical examination: a symptom-directed physical examination, including vital signs will be performed at the initial visit to confirm the diagnosis (denoted as D1 in this table) and at the Convalescent Visit (28 days after diagnosis [D28 in this table]).
2. Trained study personnel will call all participants to collect information relating to any AEs, MAAEs, AEs leading to withdrawal, SAEs, information on concomitant medications associated with those events, and any non-study vaccinations. All safety events will be followed until resolution.

3. All concomitant medications relevant to or for the treatment of an SAE or MAAE will be recorded from Screening through the final visit (Day 759).
4. Participants will have daily telemedicine visits (via video or phone) for 14 days (or until symptoms resolve, whichever is longer). Telemedicine visits may be performed by medically qualified staff appropriately delegated by the investigator. During the telemedicine visit (preferably done in the evening) the participant will be asked to verbally report the severity of each symptom, their highest body temperature and lowest oxygen saturation for that day and the investigator will determine if medical attention is required due to worsening of COVID-19. The participant will also be reminded to collect a saliva sample and return it to the study site, on the appropriate days.
5. Participants with symptoms of COVID-19 will be asked to return within 72 hours or as soon as possible to the study site or medically qualified staff from the site will conduct a home visit as soon as possible to collect an NP swab sample (for RT-PCR) and collect a blood sample for immunologic assessment of SARS-CoV-2 infection and evaluate for COVID-19. An additional NP swab sample will also be tested for the presence of other respiratory pathogens. In addition, the study site may collect an additional (local/non-central) respiratory sample for SARS-CoV-2 testing to be able to render appropriate medical care for the study participant as determined by local standards of care. Additionally, clinical information will be carefully collected to evaluate the severity of the clinical case. If the RT-PCR test from the NP swab sample from the Illness Visit is negative for SARS-CoV-2, the participant will exit the Convalescent Period and resume the study schedule (Table 16, Table 17, and Table 18).
6. This can be a home visit if necessary.
7. Participants will collect their own saliva sample using the saliva collection tubes provided on 3, 5, 7, 9, 14, and 21 days after the initial Illness Visit, and return them to the study site according to Sponsor instructions. Participants who are pending a central laboratory PCR may exit from the Convalescent Period based on a negative PCR result from a CLIA-certified or CLIA-certified waiver local laboratory at the investigator's discretion.
8. At this visit, a saliva sample will be collected.

**Table 20: Part B: Participant Decision Visit**

| | All Participants | | | | |
|---|---|---|---|---|---|
| Sign revised informed consent form | X | | | | |
| Confirm participant's request to be unblinded or not to be unblinded[a] | X | | | | |
| Nasopharyngeal swab | X | | | | |
| Blood for immunologic analysis | X | | | | |
| Counselling the importance of public health measures[b] | X | | | | |
| | | Open-label Cohort | | | |
| **Participant Status after PDV** | **Blinded Cohort** | **Previously Receiving mRNA-1273** | **Remaining on Placebo** | **Placebo Requesting mRNA-1273** | **mRNA-1273 who received 1 dose ONLY** |
| Continue with original schedules of events, as applicable (Table 16, Table 17, Table 18, and Table 19) | X | X | X | X | X |
| Supplemental Schedule of Events: Open-Label Days 1-57 (Table 21)[c] | -- | -- | -- | X | -- |
| Modified Supplemental Schedule of Events: Open-Label Days 1-57 (Table 22)[c] | -- | -- | -- | -- | X |

Abbreviation: PDV = participant decision visit.

[a] At the point when the mRNA-1273 vaccine is no longer available for study use, any participant who schedules a PDV after this time point will be unblinded (with no option to stay blinded), but will not be offered mRNA-1273 study vaccine.

[b] All participants are counselled about the importance of continuing other public health measures to limit the spread of disease including physical-social distancing, wearing a mask, and hand-washing.

[c] The Supplemental and the Modified Supplemental Schedule of Events are intended to occur in addition to the original schedules of events, as applicable (Table 16, Table 17, Table 18, and Table 19) and therefore there is a possibility for study visits to overlap. If visits overlap according to respective visit windows, a single visit may be done with the combined study procedures completed once.

##### Table 21: Supplemental Schedule of Events: Open-Label Observational Phase - Placebo Participants who Request to Receive mRNA-1273

**NOTE: Supplemental Schedule of Events: Open-Label Observational Phase is to be used ONLY for participants who received placebo during the Blinded Phase of this study (Part A), and request to receive mRNA-1273. These participants will comply with this Supplemental SoE (Table 21) in addition to the original SoEs as applicable (Table 16, Table 17, Table 18, and Table 19). As this Supplemental SoE is intended to occur in addition to the original schedules of events, there is a possibility for study visits to overlap. If visits overlap according to respective visit windows, a single visit may be done with the combined study procedures completed once.**

| **Visit Number** | 0 | OL-1 | | OL-2 | | OL-3 |
|---|---|---|---|---|---|---|
| **Type of Visit** | C | C | SC | C | SC | C |
| **Study Visit Day** | PDV[1] and OL-D1[1] | | 7 days after OL-D1 (OL-D8) | OL-D29 | 7 days after OL-D29 (OL-D36) | OL-D57 |
| **Window Allowance (Days)** | - | | +3 | -3/+14 | +3 | -3/+14 |
| **Days Since Most Recent Vaccination** | 0 | | 7 | 28/0 | 7 | 28 |
| Confirm signing of ICF, concomitant medications, medical history | X | | | | | |
| Confirm participant meets inclusion and exclusion criteria | X | | | | | |
| Physical examination[2] | X | | | X | | X |
| Pregnancy testing[3] | X | | | X | | |
| **Immunogenicity Assessment** | | | | | | |
| Blood for immunologic analysis[5] | X | | | | | X |
| **Dosing** | | | | | | |
| Study injection (including 30-minute post-dosing observation period[6]) | X | | | X | | |
| **Efficacy Assessment** | | | | | | |
| Surveillance for COVID-19/Unscheduled Visit[4] | X | | X | X | X | X |

| **NOTE: Supplemental Schedule of Events: Open-Label Observational Phase is to be used ONLY for participants who received placebo during the Blinded Phase of this study (Part A), and request to receive mRNA-1273. These participants will comply with this Supplemental SoE (Table 21) in addition to the original SoEs as applicable (Table 16, Table 17, Table 18, and Table 19). As this Supplemental SoE is intended to occur in addition to the original schedules of events, there is a possibility for study visits to overlap. If visits overlap according to respective visit windows, a single visit may be done with the combined study procedures completed once.** | | | | | | |
|---|---|---|---|---|---|---|
| **Visit Number** | 0 | OL-1 | | OL-2 | | OL-3 |
| **Type of Visit** | C | C | SC | C | SC | C |
| **Study Visit Day** | PDV[1] and OL-D1[1] | | 7 days after OL-D1 (OL-D8) | OL-D29 | 7 days after OL-D29 (OL-D36) | OL-D57 |
| **Window Allowance (Days)** | - | | +3 | -3/+14 | +3 | -3/+14 |
| **Days Since Most Recent Vaccination** | 0 | | 7 | 28/0 | 7 | 28 |
| Nasopharyngeal swab[5] | X | | | | | |
| **Safety Assessments** | | | | | | |
| Follow-up Safety[7] | | | X | | X | |
| Recording of MAAEs, AESIs, AE leading to withdrawal and concomitant medications relevant to or for the treatment of the MAAE[8] | X | | X | X | X | X |
| Recording of SAEs and concomitant medications relevant to or for the treatment of the SAE[8] | X | | X | X | X | X |
| Recording of concomitant medications and non-study vaccinations[8] | X | | X | X | X | X |

Abbreviations: AE = adverse event; C = clinic visit; D = day; eDiary = electronic diary; ICF = informed consent form; OL = open-label; MAAE = medically attended AE; PDV = participant decision visit; SAE = serious adverse event; SC = safety (phone) call.

Note: In accordance with FDA Guidance on Conduct of Clinical Trials of Medical Products during the COVID-19 Pandemic (DHHS 2020), investigators may convert study site visits to telemedicine visits with the approval of the Sponsor. All scheduled study visits should be completed within the respective visit windows. If the participant is not able to come on-site for a study site visit as a result of the COVID-19 Pandemic (self-quarantine or disruption of study site activities following business continuity plans and/or local government mandates for “stay at home” or “shelter in place”), a safety call to the participant should be made in place of the study site visit. The safety call should encompass all scheduled visit assessments that can be completed remotely, such as assessment for AEs and concomitant medications (eg, as defined in scheduled biweekly safety phone calls). Home visits will be permitted for all non-dosing visits except for Screening if a participant cannot come to the study site as a result of the COVID19 pandemic.

1. The Day 0 and OL-D1 visit may be performed on 2 separate visits.
2. Symptom-directed physical examination will be performed at the PDV and on OL-D29. On each dosing day before injection, the arm receiving the injection should be examined and the associated lymph nodes should be evaluated. Any clinically significant finding identified during a study visit should be reported as a MAAE. Vital signs are to be collected pre- and post-dosing on days of injection (OL-D1 and OL-D29). Participants who are febrile (body temperature ≥ 38.0°C/100.4°F) before dosing (OL-D1 and OL-D29) must be rescheduled within the relevant window period to receive the injection. Afebrile participants with minor illnesses can be enrolled at the discretion of the investigator.
3. Pregnancy test at the PDV and OL-D29 will be a point-of-care urine test. At the discretion of the investigator, a pregnancy test either via blood or point-of-care urine test can be performed. Follicle-stimulating hormone level may be measured to confirm menopausal status at the discretion of the investigator.
4. Participants with symptoms of COVID-19 will be asked to return within 72 hours or as soon as possible to the study site for an unscheduled visit, to include an NP swab sample (for RT-PCR testing) and other clinical evaluations. If a study site visit is not possible, a home visit may be arranged to collect the NP sample and conduct clinical evaluations. If a home visit is not possible, the participant will be asked to submit a saliva sample to the study site by a Sponsor-approved method. At this visit, an NP swab and blood sample will be collected to evaluate for the presence of SARS-CoV-2 infection. An additional NP swab sample will be collected to test for the presence of other respiratory pathogens. In addition, the study site may collect an additional (local/non-central) respiratory sample for SARS-CoV-2 testing to be able to render appropriate medical care for the study participant as determined by local standards of care. Additionally, clinical information will be carefully collected to evaluate the severity of the clinical case. It is important to note that some of the symptoms of COVID-19 overlap with solicited systemic ARs that are expected after vaccination with mRNA-1273 (eg, myalgia, headache, fever, and chills). During the first 7 days after vaccination, when these solicited ARs are common, investigators should use their clinical judgement to decide if an NP swab should be collected. The collection of an NP swab prior to the PDV can help ensure that cases of COVID-19 are not overlooked. Any study participant reporting respiratory symptoms during the 7-day period after vaccination should be evaluated for COVID-19.
5. Sample must be collected prior to unblinding and injection at the PDV.
6. Post-dosing, participants will have a 30-minute observation period.
7. Trained study personnel will call all participants to collect information relating to any MAAEs (including any signs and symptoms of COVID-19), AESIs, AEs leading to withdrawal, SAEs, information on concomitant medications associated with those events, and any non-study vaccinations.
8. All concomitant medications relevant to or for the treatment of an SAE or MAAE will be recorded from Screening through the final visit (Day 759).

**Table 22: Modified Supplemental Schedule of Events: Open-Label Observational Phase – mRNA-1273 Participants who ONLY received 1 dose of mRNA-1273**

| **NOTE: Modified Supplemental Schedule of Events: Open-Label Observational Phase is to be used ONLY for participants who ONLY received 1 dose of mRNA-1273 during the Blinded Phase of this study (Part A) and consented to receive mRNA-1273 (see Section 4.1.2 for details). These participants will comply with this SoE (Table 22) in addition to the original SoEs as applicable (Table 16, Table 17, Table 18, and Table 19). As this Modified Supplemental SoE is intended to occur in addition to the original schedules of events, there is a possibility for study visits to overlap. If visits overlap according to respective visit windows, a single visit may be done with the combined study procedures completed once.** | | | | |
|---|---|---|---|---|
| **Visit Number** | 0 | OL-1 | | OL-2 |
| **Type of Visit** | C | C | SC | C |
| **Study Visit Day** | PDV[1] and OL-D1[1] | | 7 days after OL-D1<br>OL-D8 | OL-D29 |
| **Window Allowance (Days)** | - | | +3 | -3/+14 |
| **Days Since Most Recent Vaccination** | 0 | | 7 | 28/0 |
| Confirm signing of ICF, concomitant medications, medical history | X | | | |
| Confirm participant meets inclusion and exclusion criteria | X | | | |
| Physical examination[2] | X | | | X |
| Pregnancy testing[3] | X | | | |
| **Immunogenicity Assessment** | | | | |
| Blood for immunologic analysis[5] | X | | | X |
| **Dosing** | | | | |
| Study injection (including 30-minute post-dosing observation period[6]) | X | | | |

**NOTE: Modified Supplemental Schedule of Events: Open-Label Observational Phase is to be used ONLY for participants who ONLY received 1 dose of mRNA-1273 during the Blinded Phase of this study (Part A) and consented to receive mRNA-1273 (see Section 4.1.2 for details). These participants will comply with this SoE (Table 22) in addition to the original SoEs as applicable (Table 16, Table 17, Table 18, and Table 19). As this Modified Supplemental SoE is intended to occur in addition to the original schedules of events, there is a possibility for study visits to overlap. If visits overlap according to respective visit windows, a single visit may be done with the combined study procedures completed once.**

| **Visit Number** | 0 | OL-1 | | OL-2 |
|---|---|---|---|---|
| **Type of Visit** | C | C | SC | C |
| **Study Visit Day** | PDV[1] and OL-D1[1] | | 7 days after OL-D1 OL-D8 | OL-D29 |
| **Window Allowance (Days)** | - | | +3 | -3/+14 |
| **Days Since Most Recent Vaccination** | 0 | | 7 | 28/0 |
| **Efficacy Assessment** | | | | |
| Surveillance for COVID-19/Unscheduled Visit[4] | X | | X | X |
| Nasopharyngeal swab[5] | X | | | |
| **Safety Assessments** | | | | |
| Follow-up Safety[7] | | | X | |
| Recording of MAAEs, AESIs, AE leading to withdrawal and concomitant medications relevant to or for the treatment of the MAAE[8] | X | | X | X |
| Recording of SAEs and concomitant medications relevant to or for the treatment of the SAE[8] | X | | X | X |
| Recording of concomitant medications and non-study vaccinations[8] | X | | X | X |

Abbreviations: AE = adverse event; C = clinic visit; D = day; eDiary = electronic diary; ICF = informed consent form; OL = open-label; MAAE = medically attended AE; PDV = participant decision visit; SAE = serious adverse event; SC = safety (phone) call.

Note: In accordance with FDA Guidance on Conduct of Clinical Trials of Medical Products during the COVID-19 Pandemic (DHHS 2020), investigators may convert study site visits to telemedicine visits with the approval of the Sponsor. All scheduled study visits should be completed within the respective visit windows. If the participant is not able to come on-site for a study site visit as a result of the COVID-19 Pandemic (self-quarantine or disruption of study site activities following business continuity plans and/or local government mandates for “stay at home” or “shelter in place”), a safety call to the participant should be made in place of the study site visit. The safety call should encompass all scheduled visit assessments that can be completed remotely, such as assessment for AEs and concomitant medications (eg, as defined in scheduled biweekly safety phone calls). Home visits will be permitted for all non-dosing visits except for Screening if a participant cannot come to the study site as a result of the COVID-19 pandemic.

1. Day 0 and OL-D1 visit may be performed on 2 separate visits.
2. Symptom-directed physical examination will be performed at the PDV. On dosing day before injection, the arm receiving the injection should be examined and the associated lymph nodes should be evaluated. Any clinically significant finding identified during a study visit should be reported as a MAAE. Vital signs are to be collected pre- and post-dosing on the days of injection (OL-D1). Participants who are febrile (body temperature ≥ 38.0°C/100.4°F) before dosing (OL-D1) must be rescheduled within the relevant window period to receive the injection. Afebrile participants with minor illnesses can be enrolled at the discretion of the investigator.
3. Pregnancy test at the PDV will be a point-of-care urine test. At the discretion of the investigator, a pregnancy test either via blood or point-of-care urine test can be performed. Follicle-stimulating hormone level may be measured to confirm menopausal status at the discretion of the investigator.
4. Participants with symptoms of COVID-19 will be asked to return within 72 hours or as soon as possible to the study site for an unscheduled visit, to include an NP swab sample (for RT-PCR testing) and other clinical evaluations. If a study site visit is not possible, a home visit may be arranged to collect the NP sample and conduct clinical evaluations. If a home visit is not possible, the participant will be asked to submit a saliva sample to the study site by a Sponsor-approved method. At this visit, an NP swab and blood sample will be collected to evaluate for the presence of SARS-CoV-2 infection. An additional NP swab sample will be collected to test for the presence of other respiratory pathogens. In addition, the study site may collect an additional (local/non-central) respiratory sample for SARS-CoV-2 testing to be able to render appropriate medical care for the study participant as determined by local standards of care. Additionally, clinical information will be carefully collected to evaluate the severity of the clinical case. It is important to note that some of the symptoms of COVID-19 overlap with solicited systemic ARs that are expected after vaccination with mRNA-1273 (eg, myalgia, headache, fever, and chills). During the first 7 days after vaccination, when these solicited ARs are common, investigators should use their clinical judgement to decide if an NP swab should be collected. The collection of an NP swab prior to the PDV can help ensure that cases of COVID-19 are not overlooked. Any study participant reporting respiratory symptoms during the 7-day period after vaccination should be evaluated for COVID-19.
5. Sample must be collected prior to unblinding and injection at the PDV.
6. Post-dosing, participants will have a 30-minute observation period.
7. Trained study personnel will call all participants to collect information relating to any MAAEs (including any signs and symptoms of COVID-19), AESIs, AEs leading to withdrawal, SAEs, information on concomitant medications associated with those events, and any non-study vaccinations.
8. All concomitant medications relevant to or for the treatment of an SAE or MAAE will be recorded from Screening through the final visit (Day 759).

**Table 23: Part C Supplemental Schedule of Events: Booster Dose Phase**

**NOTE: The Part C Supplemental Schedule of Events is to be used for all participants currently enrolled in Part B and who are eligible for Part C. As this Supplemental SoE is intended to occur in addition to and in parallel with the original SoEs (Table 16, Table 17, Table 18, and Table 19), there is a possibility for study visits to overlap. If visits overlap according to respective visit windows, a single visit may be done with the combined study procedures completed once.**

| **Visit Number** | BD-1 | BD-1a | | BD-2 | BD-3 |
|---|---|---|---|---|---|
| **Type of Visit** | C | C | SC[10] | C | C |
| **Study Visit Day** | BD-D1[1] | BD-D4 | 7, 14, and 21 days after BD-D1: (BD-D8, BD-D15, BD-D22) | BD-D29 | BD-D181 |
| **Window Allowance (Days)** | - | -2 | +3 | -3/+14 | -3/+14 |
| **Days Since Most Recent Vaccination (in Part C)** | 0 | 3 | 7, 14, 21 | 28 | 180 |
| Confirm signing of ICF, concomitant medications, medical history | X | | | | |
| Confirm participant meets inclusion and exclusion criteria | X | | | | |
| Physical examination[2] | X | | | X | X |
| Pregnancy testing[3] | X | | | | |
| **Immunogenicity Assessment** | | | | | |
| Blood for immunologic analysis[5] | X | | | X | X |
| **Biomarker Assessment** | | | | | |
| Blood sample for potential biomarker analysis[9] | | X | | | |

| NOTE: The Part C Supplemental Schedule of Events is to be used for all participants currently enrolled in Part B and who are eligible for Part C. As this Supplemental SoE is intended to occur in addition to and in parallel with the original SoEs (Table 16, Table 17, Table 18, and Table 19), there is a possibility for study visits to overlap. If visits overlap according to respective visit windows, a single visit may be done with the combined study procedures completed once. | | | | | |
|---|---|---|---|---|---|
| **Visit Number** | BD-1 | BD-1a | | BD-2 | BD-3 |
| **Type of Visit** | C | C | SC[10] | C | C |
| **Study Visit Day** | BD-D1[1] | BD-D4 | 7, 14, and 21 days after BD-D1: (BD-D8, BD-D15, BD-D22) | BD-D29 | BD-D181 |
| **Window Allowance (Days)** | - | -2 | +3 | -3/+14 | -3/+14 |
| **Days Since Most Recent Vaccination (in Part C)** | 0 | 3 | 7, 14, 21 | 28 | 180 |
| **Dosing** | | | | | |
| Study injection (including 30-minute post-dosing observation period[6]) | X | | | | |
| **Efficacy Assessment** | | | | | |
| Surveillance for COVID-19/Unscheduled Visit[4] | X | | X | X | X |
| Nasopharyngeal swab[5] | X | | | | |
| **Safety Assessments** | | | | | |
| Follow-up safety[7] | | | X | | |
| Recording of unsolicited AEs[10] | X | X | X | X | |
| Recording of MAAEs, AESIs, AE leading to withdrawal and concomitant medications relevant to or for the treatment of the MAAE[8] | X | X | X | X | X |
| Recording of SAEs and concomitant medications relevant to or for the treatment of the SAE[8] | X | X | X | X | X |

| **NOTE: The Part C Supplemental Schedule of Events is to be used for all participants currently enrolled in Part B and who are eligible for Part C. As this Supplemental SoE is intended to occur in addition to and in parallel with the original SoEs (Table 16, Table 17, Table 18, and Table 19), there is a possibility for study visits to overlap. If visits overlap according to respective visit windows, a single visit may be done with the combined study procedures completed once.** | | | | | |
|---|---|---|---|---|---|
| **Visit Number** | BD-1 | BD-1a | | BD-2 | BD-3 |
| **Type of Visit** | C | C | SC[10] | C | C |
| **Study Visit Day** | BD-D1[1] | BD-D4 | 7, 14, and 21 days after BD-D1: (BD-D8, BD-D15, BD-D22) | BD-D29 | BD-D181 |
| **Window Allowance (Days)** | - | -2 | +3 | -3/+14 | -3/+14 |
| **Days Since Most Recent Vaccination (in Part C)** | 0 | 3 | 7, 14, 21 | 28 | 180 |
| Recording of concomitant medications and non-study vaccinations[8] | X | X | X | X | X |

Abbreviations: AE = adverse event; BD = booster dose; C = clinic visit; D = day; eDiary = electronic diary; ICF = informed consent form; OL = open-label; MAAE = medically attended AE; PDV = participant decision visit; SAE = serious adverse event; SC = safety (phone) call.

Note: In accordance with FDA Guidance on Conduct of Clinical Trials of Medical Products during the COVID-19 Pandemic (DHHS 2020), investigators may convert study site visits to telemedicine visits with the approval of the Sponsor. All scheduled study visits should be completed within the respective visit windows. If the participant is not able to come on-site for a study site visit as a result of the COVID-19 Pandemic (self-quarantine or disruption of study site activities following business continuity plans and/or local government mandates for "stay at home" or "shelter in place"), a safety call to the participant should be made in place of the study site visit. The safety call should encompass all scheduled visit assessments that can be completed remotely, such as assessment for AEs and concomitant medications (eg, as defined in scheduled biweekly safety phone calls). Home visits will be permitted for all non-dosing visits except for Screening if a participant cannot come to the study site as a result of the COVID-19 pandemic.

1. A BD may be administered to all participants who are currently enrolled in Part B, and have received at least 1 dose of mRNA-1273, , provided there are no current contraindications for further dosing (Section 7). A participant who is currently in the Convalescent Period may come in for a BD-1 visit and receive a BD as long as they are no longer symptomatic, and this includes the possibility for a Convalescent Visit Day 28 to overlap and be combined with a BD-1 visit.
2. Symptom-directed physical examination will be performed at the BD Day 1. On dosing day before injection, the arm receiving the injection should be examined and the associated lymph nodes should be evaluated. At visits BD-2 (BD-D29) and BD-3 (BD-D181), a symptom-directed physical examination

may be performed at the discretion of the investigator. Any clinically significant finding identified during a study visit should be reported as a MAAE. Vital signs are to be collected pre- and post-dosing (participant will be seated for at least 5 minutes before all measurements are taken per Section 8.2.5) on the day of injection (BD-D1). Participants who are febrile (body temperature ≥ 38.0°C/100.4°F) before dosing (BD-D1) must be rescheduled to receive the injection. Afebrile participants with minor illnesses can be vaccinated at the discretion of the investigator.

3. The pregnancy test at the BD-1 visit will be a point-of-care urine test. At the discretion of the investigator, a pregnancy test either via blood or point-of-care urine test can be performed. Follicle-stimulating hormone level may be measured to confirm menopausal status at the discretion of the investigator.
4. Participants with symptoms of COVID-19 will be asked to return within 72 hours or as soon as possible to the study site for an unscheduled visit, to collect an NP swab sample (for RT-PCR testing) and other clinical evaluations. If a study site visit is not possible, a home visit may be arranged to collect the NP sample and conduct clinical evaluations. If a home visit is not possible, the participant will be asked to submit a saliva sample to the study site by a Sponsor-approved method. At this visit, an NP swab and blood sample will be collected to evaluate for the presence of SARS-CoV-2 infection. An additional NP swab sample will be collected to test for the presence of other respiratory pathogens. In addition, the study site may collect an additional respiratory sample for SARS-CoV-2 testing to be able to render appropriate medical care for the study participant as determined by local standards of care. Additionally, clinical information will be carefully collected to evaluate the severity of the clinical case. It is important to note that some of the symptoms of COVID-19 overlap with solicited systemic ARs that are expected after vaccination with mRNA-1273 (eg, myalgia, headache, fever, and chills). During the first 7 days after vaccination, when these solicited ARs are common, investigators should use their clinical judgement to decide if an NP swab should be collected. Any study participant reporting respiratory symptoms during the 7-day period after vaccination should be evaluated for COVID-19.
5. The nasopharyngeal swab sample is strongly encouraged but ultimately optional for the participant's. If collected, it must be collected prior to injection at the BD-1 visit.
6. Post-dosing, participants will have a 30-minute observation period.
7. Trained study personnel will call all participants to collect information relating to any unsolicited AEs, MAAEs (including any signs and symptoms of COVID-19), AESIs, AEs leading to withdrawal, SAEs, information on concomitant medications associated with those events, and any non-study vaccinations.
8. All concomitant medications relevant to or for the treatment of an SAE or MAAE will be recorded from Screening through the final visit (Day 759).
9. Only for participants who chose to receive a BD. Serum sample from two ~4 mL optional blood draws, subject to availability of blood draw kits, and will be confirmed with participant at time of re-consenting for Part C. Biomarker plasma and biomarker serum samples will be stored for potential future biomarker assessment.
10. Only for participants who chose to receive a BD.

#### 11.2. APPENDIX 2: Study Governance Considerations

##### 11.2.1. Regulatory and Ethical Considerations

This study will be conducted in accordance with the protocol and with the following:

- Consensus ethical principles derived from international guidelines including the Declaration of Helsinki and Council for International Organizations of Medical Sciences (CIOMS) International Ethical Guidelines.
- Applicable ICH GCP Guidelines.
- Applicable laws and regulatory requirements.
- The protocol, protocol amendments, ICF, Investigator Brochure, and other relevant documents (eg, advertisements) must be submitted to an IRB/IEC by the investigator and reviewed and approved by the IRB/IEC before the study is initiated.
- Any amendments to the protocol will require IRB/IEC approval before implementation of changes made to the study design, except for changes necessary to eliminate an immediate hazard to study participants.
- The investigator will be responsible for the following:

– Providing written summaries of the status of the study to the IRB/IEC annually or more frequently in accordance with the requirements, policies, and procedures established by the IRB/IEC

– Notifying the IRB/IEC of SAEs or other significant safety findings as required by IRB/IEC procedures

– Providing oversight of the conduct of the study at the site and adherence to requirements of 21 CFR, ICH guidelines, the IRB/IEC, European regulation 536/2014 for clinical studies (if applicable), and all other applicable local regulations.

##### 11.2.2. Study Monitoring

Before an investigational site can enter a participant into the study, a representative of Moderna or its representatives will visit the investigational study site to:

- Determine the adequacy of the facilities.
- Discuss with the investigator(s) and other personnel their responsibilities with regard to protocol adherence, and the responsibilities of Moderna or its representatives. This will be documented in a Clinical Study Agreement between Moderna, designated CRO, and the investigator.

According to ICH GCP guideline, the Sponsor of the study is responsible for ensuring the proper conduct of the study with regard to protocol adherence and validity of data recorded on the eCRFs. The study monitor's duties are to aid the investigator and Moderna in the maintenance of complete, accurate, legible, well-organized, and easily retrievable data. The study monitor will advise the investigator of the regulatory necessity for study-related monitoring, audits, IRB/IEC review, and inspection by providing direct access to the source data/documents. In addition, the study monitor will explain to and interpret for the investigator all regulations applicable to the clinical evaluation of an IP as documented in ICH guidelines.

It is the study monitor's responsibility to inspect the eCRFs and source documentation throughout the study to protect the rights of the participants; to verify adherence to the protocol; to verify completeness, accuracy, and consistency of the data; and to confirm adherence of study conduct to any local regulations. Details will be outlined in the Clinical Monitoring Plan. During the study, a monitor from Moderna or a representative will have regular contacts with the investigational site, for the following:

- Provide information and support to the investigator(s).
- Confirm that facilities remain acceptable.
- Confirm that the investigational team is adhering to the protocol, that the data are being accurately recorded in the eCRFs, and that IP accountability checks are being performed.
- Perform source data verification. This includes a comparison of the data in the eCRFs with the participant's medical records at the hospital or practice, and other records relevant to the study. This will require direct access to all original records for each participant (eg, clinical charts or electronic medical record system).
- Record and report any protocol deviations not previously sent.
- Confirm AEs and SAEs have been properly documented on eCRFs and confirm any SAEs have been forwarded to the SAE Hotline, and those SAEs that met criteria for reporting have been forwarded to the IRB/IEC.

The monitor will be available between visits if the investigator(s) or other staff needs information or advice.

In accordance with FDA Guidance on Conduct of Clinical Trials of Medical Products during the COVID-19 Pandemic (DHHS 2020), investigators may convert study site visits to telemedicine visits with the approval of the Sponsor (Section 8 for Procedures).

The DSMB will also have responsibility for safety monitoring (Section 8.4.1).

##### 11.2.3. Audits and Inspections

Moderna, their designee(s), the IRB/IEC, or regulatory authorities will be allowed to conduct site visits to the investigational facilities for the purpose of monitoring or inspecting any aspect of the study. The investigator agrees to allow Moderna, their designee(s), the IRB/IEC, or regulatory authorities to inspect the IP storage area, IP stocks, IP records, participant charts and study source documents, and other records relative to study conduct.

Authorized representatives of Moderna, a regulatory authority, and any IRB/IEC may visit the site to perform audits or inspections, including source data verification. The purpose of a Sponsor audit or inspection is to systematically and independently examine all study-related activities and documents to determine whether these activities were conducted, and data were recorded, analyzed, and accurately reported according to the protocol, ICH GCP E6R2, and any applicable regulatory requirements. The investigator should contact Moderna immediately if contacted by a regulatory agency about an inspection.

The Principal Investigator must obtain IRB approval for the investigation. Initial IRB approval, and all materials approved by the IRB for this study including the participant consent form and recruitment materials must be maintained by the investigator and made available for inspection.

##### 11.2.4. Financial Disclosure

The investigator is required to provide financial disclosure information to allow the Sponsor to submit the complete and accurate certification or disclosure statements required under 21 CFR 54. In addition, the investigator must provide the Sponsor with a commitment to promptly update this information if any relevant changes occur during the course of the investigation and for 1 year following the completion of the study.

The Sponsor, the CRO, and the study site are not financially responsible for further testing or treatment of any medical condition that may be detected during the screening process. In addition, in the absence of specific arrangements, the Sponsor, the CRO, and the study site are not financially responsible for further treatment of the disease under study.

##### 11.2.5. Recruitment Procedures

Advertisements to be used for the recruitment of study participants, and any other written information regarding this study to be provided to the participant should be submitted to the Sponsor for approval. All documents must be approved by the IRB.

##### 11.2.6. Informed Consent Process

The informed consent document(s) must meet the requirements of 21 CFR 50, local regulations, ICH guidelines, Health Insurance Portability and Accountability Act (HIPAA) requirements, where applicable, and the IRB/IEC or study center. All consent documents will be approved by the appropriate IRB/IEC. The actual ICF used at each center may differ, depending on local regulations and IEC / IRB requirements. However, all versions must contain the standard information found in the sample ICF provided by the Sponsor. Any change to the content of the ICF must be approved by the Sponsor and the IEC / IRB prior to the form being used.

If new information becomes available that may be relevant to the participant's willingness to continue participation in the study, this will be communicated to him / her in a timely manner. Such information will be provided via a revised ICF or an addendum to the original ICF.

The investigator or his/her representative will explain the nature of the study to the participant and answer all questions regarding the study.

The investigator is responsible for ensuring that the participant fully understands the nature and purpose of the study. Information should be given in both oral and written form whenever possible.

No participant should be obliged to participate in the study. The participant must be informed that participation is voluntary. Participants, their relatives, guardians, or (if applicable) legal representatives must be given ample opportunity to inquire about details of the study. The information must make clear that refusal to participate in the study or withdrawal from the study at any stage is without any prejudice to the participant's subsequent care.

The participant must be allowed sufficient time to decide whether they wish to participate. The participant must be made aware of and give consent to direct access to his/her source medical records by study monitors, auditors, the IRB/IEC, and regulatory authorities. The participant should be informed that such access will not violate participant confidentiality or any applicable regulations. The participant should also be informed that he/she is authorizing such access by signing the ICF.

A copy of the ICF(s) must be provided to the participant.

A participant who is rescreened is not required to sign another ICF if the rescreening occurs within 28 days from the previous ICF signature date.

The ICF will also explain that excess serum from immunogenicity testing may be used for future research, which may be performed at the discretion of the Sponsor to further characterize the immune response to SARS-CoV-2, additional assay development, and the immune response across CoV.

##### 11.2.7. Protocol Amendments

No change or amendment to this protocol may be made by the investigator or the Sponsor after the protocol has been agreed to and signed by all parties unless such change(s) or amendment(s) has (have) been agreed upon by the investigator or the Sponsor. Any change agreed upon will be recorded in writing, and the written amendment will be signed by the investigator and the Sponsor. Institutional review board approval is required prior to the implementation of an amendment, unless overriding safety reasons warrant immediate action, in which case the IRB(s)/IEC(s) will be promptly notified.

Any modifications to the protocol or the ICF, which may impact the conduct of the study, potential benefit of the study, or may affect participant safety, including changes of study objectives, study design, participant population, sample sizes, study procedures, or significant administrative aspects will require a formal amendment to the protocol. Such amendment will be released by the Sponsor, agreed by the investigator(s), and approved by the relevant IRB(s)/IEC(s) prior to implementation. A signed and dated statement that the protocol, any subsequent relevant amended documents and the ICF have been approved by relevant IRB(s)/IEC(s) must be provided to the Sponsor before the study is initiated.

Administrative changes of the protocol are minor corrections and/or clarifications that have no effect on the way the study is to be conducted. These administrative changes will be released by the Sponsor, agreed by the investigator(s), and notified to the IRB(s)/IEC(s).

##### 11.2.8. Protocol Deviations

The noncompliance may be either on the part of the participant, the investigator, or the study site staff. As a result of deviations, corrective actions are to be developed by the site and implemented promptly.

It is the responsibility of the site investigator to use continuous vigilance to identify and report deviations to the Sponsor or its designee. All deviations must be addressed in study source documents, reported to study monitor. Protocol deviations must be sent to the reviewing IRB/IEC per their policies. The site investigator is responsible for knowing and adhering to the reviewing IRB/IEC requirements.

##### 11.2.9. Data Protection

Participants will be assigned a unique identifier by the Sponsor. Any participant records or datasets that are transferred to the Sponsor will contain the identifier only; participant names or any information which would make the participant identifiable will not be transferred.

The participant must be informed that his/her personal study-related data will be used by the Sponsor in accordance with local data protection law. The level of disclosure must also be explained to the participant.

The participant must be informed that his/her medical records may be examined by Clinical Quality Assurance auditors or other authorized personnel appointed by the Sponsor, by appropriate IRB/IEC members, and by inspectors from regulatory authorities.

Individual participant medical information obtained as a result of this study is considered confidential, and disclosure to third parties is prohibited. Information will be accessible to authorized parties or personnel only. Medical information may be given to the participant's physician or to other appropriate medical personnel responsible for the participant's well-being. Each participant will be asked to complete a form allowing the investigator to notify the participant's primary health care provider of his/her participation in this study.

All laboratory specimens, evaluation forms, reports, and other records will be identified in a manner designed to maintain participant confidentiality. All records will be kept in a secure storage area with limited access. Clinical information will not be released without the written permission of the participant, except as necessary for monitoring and auditing by the Sponsor, its designee, relevant regulatory authority, or the IRB.

The investigator and all employees and coworkers involved with this study may not disclose or use for any purpose other than performance of the study, any data, record, or other unpublished, confidential information disclosed to those individuals for the purpose of the study. Prior written agreement from the Sponsor or its designee must be obtained for the disclosure of any confidential information to other parties.

##### 11.2.10. Sample Retention and Future Biomedical Research

The retention period of laboratory samples will be 15 years, or as permitted by local regulations, to address further scientific questions related to mRNA-1273 or anti-respiratory virus immune response. In addition, identifiable samples can be destroyed at any time at the request of the participant. During the study, or during the retention period, in addition to the analysis outlined in the study endpoints, exploratory analysis may be conducted, using other antibody-based methodologies, on any remaining blood or serum samples, including participants who provide samples for screening, but are not subsequently enrolled. These analyses would extend the search for other potentially relevant biomarkers to investigate the effect of mRNA-1273, as well as to determine how changes in the markers may relate to exposure and clinical outcomes. A decision to perform such exploratory research may arise from new scientific findings related to the drug class or disease, as well as reagent and assay availability.

##### 11.2.11. Data Quality Assurance and Quality Control

Data collection is the responsibility of the clinical study staff at the site under the supervision of the site investigator. The investigator is responsible for ensuring the accuracy, completeness, legibility, and timeliness of the data reported.

- All participant data relating to the study will be recorded eCRF unless transmitted to the Sponsor or designee electronically (eg, laboratory data). The investigator is responsible for verifying that data entries are accurate and correct by physically or electronically signing the eCRF.
- The investigator must maintain accurate documentation (source data) that supports the information entered in the eCRF.
- The investigator must permit study-related monitoring, audits, IRB/IEC review, and regulatory agency inspections and provide direct access to source data documents.
- Monitoring details describing strategy (eg, risk-based initiatives in operations and quality such as Risk Management and Mitigation Strategies and Analytical Risk-Based Monitoring), methods, responsibilities and requirements, including handling of noncompliance issues and monitoring techniques (central, remote, or on-site monitoring) are provided in the study Monitoring Plan, Centralized Monitoring Plan, and Risk Management Plan.
- The Sponsor or designee is responsible for the data management of this study including quality checking of the data.
- The Sponsor assumes accountability for actions delegated to other individuals (eg, Contract Research Organizations).
- Study monitors will perform ongoing source data verification to confirm that data entered into the CRF by authorized site personnel are accurate, complete, and verifiable from source documents; that the safety and rights of participants are being protected; and that the study is being conducted in accordance with the currently approved protocol and any other study agreements, ICH GCP, and all applicable regulatory requirements.
- Records and documents, including signed ICFs, pertaining to the conduct of this study must be retained by the investigator for 15 years after study completion unless local regulations or institutional policies require a longer retention period. No records may be destroyed during the retention period without the written approval of the Sponsor. No records may be transferred to another location or party without written notification to the Sponsor.

Quality assurance (QA) includes all the planned and systematic actions that are established to ensure that the clinical study is performed, and the data are generated, documented (recorded), and reported according to ICH GCP and local/regional regulatory standards.

A QA representative from the Sponsor or qualified designee, who is independent of and separated from routine monitoring, may periodically arrange inspections/audits of the clinical study by reviewing the data obtained and procedural aspects. These inspections may include on-site inspections/audits and source data checks. Direct access to source documents is required for the purpose of these periodic inspections/audits.

##### 11.2.12. Data Collection and Management

This study will be conducted in compliance with ICH CGP guidelines. This study will also be conducted in accordance with the most recent version of the Declaration of Helsinki.

This study will use electronic data collection (to collect data directly from the investigational site using eCRFs). The investigator is responsible for ensuring that all sections of each eCRF are completed promptly and correctly and that entries can be verified against any source data.

Study monitors will perform source document verification to identify inconsistencies between the eCRFs and source documents. Discrepancies will be resolved in accordance with the principles of GCP. Detailed study monitoring procedures are provided in the Clinical Monitoring Plan.

AEs will be coded with the most current available version of MedDRA. Concomitant medications will be coded using WHO – Drug Reference List.

##### 11.2.13. Source Documents

Source documents are original documents or certified copies, and include, but are not limited to, diaries, medical and hospital records, screening logs, informed consent / assent forms, telephone contact logs, and worksheets. Source documents provide evidence for the existence of the participant and substantiate the integrity of the data collected. Source documents are filed at the investigator’s site.

Data reported on the CRF or entered in the eCRF that are transcribed from source documents must be consistent with the source documents or the discrepancies must be explained. The investigator may need to request previous medical records or transfer records, depending on the study. Also, current medical records must be available.

Moderna or its designee requires that the investigator prepare and maintain adequate and accurate records for each participant treated with the IP. Source documents such as any hospital, clinic, or office charts and the signed ICFs are to be included in the investigator’s files with the participant’s study records.

##### 11.2.14. Retention of Records

The Principal Investigator must maintain all documentation relating to the study for a period of at least 2 years after the last marketing application approval or, if not approved, 2 years following the discontinuance of the test article for investigation. If this requirement differs from any local regulations, the local regulations will take precedence unless the local retention policy is less than 2 years.

If it becomes necessary for Moderna or the regulatory authority to review any documentation relating to the study, the investigator must permit access to such records. No records will be destroyed without the written consent of the Sponsor, if applicable. It is the responsibility of the Sponsor to inform the investigator when these documents no longer need to be retained.

##### 11.2.15. Study and Site Closure

If the study is prematurely terminated or suspended, the Sponsor shall promptly inform the investigators, the IECs/IRBs, the regulatory authorities, and any CRO(s) used in the study of the reason for termination or suspension, as specified by the applicable regulatory requirements. The investigator shall promptly inform the participant and should assure appropriate participant therapy and/or follow-up.

The Sponsor or designee reserves the right to close the study site or terminate the study at any time for any reason at the sole discretion of the Sponsor.

The investigator may initiate study-site closure at any time, provided there is reasonable cause and sufficient notice is given in advance of the intended termination.

Reasons for the early closure of a study site by the Sponsor or investigator may include but are not limited to:

- Continuation of the study represents a significant medical risk to participants.
- Failure of the investigator to comply with the protocol, the requirements of the IRB/IEC or local health authorities, the Sponsor's procedures, or GCP guidelines.
- Inadequate recruitment of participants by the investigator.
- Discontinuation of further mRNA-1273 development.

Study sites will be closed upon study completion. A study site is considered closed when all required documents and study supplies have been collected and a study-site closure visit has been performed.

##### 11.2.16. Publication Policy

The results of this study may be published or presented at scientific meetings. If this is foreseen, the investigator agrees to submit all manuscripts or abstracts to the Sponsor before submission. This allows the Sponsor to protect proprietary information and to provide comments.

The Sponsor will comply with the requirements for publication of study results. In accordance with standard editorial and ethical practice, the Sponsor will generally support publication of multicenter studies only in their entirety and not as individual site data. In this case, a coordinating investigator will be designated by mutual agreement.

Authorship will be determined by mutual agreement and in line with International Committee of Medical Journal Editors authorship requirements.

The clinical study plan and the results of the study will be published on www.ClinicalTrials.gov in accordance with 21 CFR 50.25(c). The results of and data from this study belong to Moderna.

#### 11.3. APPENDIX 3: Contraceptive Guidance

**Woman of Childbearing Potential (WOCBP)**

Females of childbearing potential are those who are considered fertile following menarche and until becoming postmenopausal unless permanently sterile (see below). If fertility is unclear (eg, amenorrhea in adolescents or athletes) and a menstrual cycle cannot be confirmed before first dose of study treatment, additional evaluation should be considered.

Women in the following categories are not considered WOCBP:

1. Premenarchal
2. Premenopausal, surgically sterile female with 1 of the following:
    a. Documented complete hysterectomy
    b. Documented bilateral salpingectomy
    c. Documented bilateral oophorectomy

For individuals with permanent infertility due to an alternate medical cause other than the above, (eg, mullerian agenesis, androgen insensitivity), investigator discretion should be applied in determining study entry.

Note: Documentation can come from the site personnel’s: review of the participant’s medical records, medical examination, or medical history interview.

3. Postmenopausal female

- A postmenopausal state is defined as no menses for 12 months without an alternative medical cause. The following age-specific requirements apply:
    - Women <50 years of age would be considered postmenopausal if they have been amenorrheic for 12 months or more following cessation of exogenous hormonal treatments and if they have luteinizing hormone and FSH levels in the postmenopausal range for the institution.
    - Women ≥ 50 years of age would be considered postmenopausal if they have been amenorrheic for 12 months or more, had radiation-induced menopause with last menses >1 year ago or had chemotherapy-induced menopause with last menses > 1 year ago.
- A high FSH level in the postmenopausal range may be used to confirm a postmenopausal state in women not using hormonal replacement therapy (HRT).
- Females on HRT and whose menopausal status is in doubt will be required to use one of the non-estrogen hormonal highly effective contraception methods if they wish to continue their HRT during the study. Otherwise, they must discontinue HRT to allow confirmation of postmenopausal status before study enrollment.

### 11.4. APPENDIX 4: Statistical Appendices

#### 11.4.1. Estimands and Estimand Specifications

**Table 24: Intercurrent Event Types**

| Label | Intercurrent Event Type | Comment |
|---|---|---|
| IcEv1 (death without confirmation of cases, ie, unrelated death) | Unrelated death without documented confirmed COVID-19 | Participants in PP Set who die due to reasons unrelated to COVID-19 without confirmation of being a case will all be included in statistical analysis. |
| IcEv2 (early COVID-19) | COVID-19 starting up to 14 days after the second dose of IP | Participants in PP Set who experience early COVID-19 up to 14 days after the second dose of IP will all be included in statistical analysis. |

Abbreviations: IcEv: intercurrent event; IP = investigational product; PP: per-protocol.

##### Table 25: Primary Objective and Estimands with Rationale for Strategies to Address Intercurrent Events for Per-Protocol Analysis

| **Objective: To demonstrate the efficacy of mRNA-1273 to prevent COVID-19** | |
|---|---|
| **Estimand Description** | Vaccine efficacy will be measured using 1 – HR (mRNA-1273/Placebo) of COVID-19 from 14 days after second dose of IP in adults. A hypothetical strategy will be used for deaths unrelated to COVID-19, and a hypothetical strategy will also be used for early COVID-19 in participants in the PP Set. |
| **Target Population** | Adults aged 18 years and older in circumstances at a high risk of SARS-CoV-2 infection but without medical conditions that pose additional risk of developing severe disease.<br>The population excludes those previously infected or vaccinated for SARS-CoV-2 or with a medical condition, on treatment that poses additional risks (including those requiring immunosuppressants or immune-modifying drugs), or SARS-CoV-2 pre-positive. |
| **Variable/Endpoint** | Time to COVID-19 disease, censoring at early discontinuation, early COVID-19, or last assessment for an event not being observed, whichever comes earlier. |
| **Treatment Condition(s)** | **Test:** mRNA-1273<br>**Reference:** Placebo |
| **Estimand Label** | **Estimand 1** |
| **Population-Level Summary** | Vaccine efficacy defined as 1 - HR of mRNA-1273/Placebo |
| **Intercurrent Event Strategy** | |
| **IcEv1 (Unrelated death):** | Hypothetical |
| **IcEv2 (early infection):** | Hypothetical |
| **Rationale for Strategy(s)** | Hypothetical: unrelated death without confirmation of COVID-19 will be censored at the time of death as if there is no event, handled with independent censoring.<br>Hypothetical: early case in PP Set will be censored at the time of case onset if it is not a case, handled with independent censoring. |

##### 11.4.2. Statistical Methods and Sensitivity Analyses

**Table 26: Summary of Statistical Methods and Sensitivity Analyses**

| Estimand Label | Estimand Description | Main Estimation | | | Sensitivity Analysis |
|---|---|---|---|---|---|
| | | Analysis Set | Imputation/Data/Censoring Rules | Analysis Model/Method | |
| Estimand 1 | See Table 25 | PP | Participants who did not develop COVID-19, ongoing or complete the study will be censored at the last assessment.<br>Participants who discontinue early or die without COVID-19 will be censored at the date of discontinuation or death respectively.<br>Participants who is a confirmed case prior to 14 days after the second dose of IP will be censored on the date of confirmation. | The (1 - HR) with 95% CI will be estimated using Cox proportional hazard regression analysis with treatment arm as a fixed effect stratified by randomization stratification factors. And the null hypothesis will be tested using Wald Chi-square method. | As described in Section 9.5.1 with cases counted starting after the second dose of IP |

#### 11.5. APPENDIX 5: Adverse Events of Special Interest Terms

Investigators should report all events which fall into the following categories as an AESI per the reporting processes specified in Section 8.3.3. The AESIs listed in Table 27 are medical concepts that may be related to COVID-19 or are of interest in COVID-19 vaccine safety surveillance. Even if any of these events occur in the setting of a COVID infection, the event should still be reported as an AESI if it is one of the medical concepts in **Error! Reference source not found.**.

**Table 27: Adverse Events of Special Interest**

| Medical Concept | Additional Notes |
|---|---|
| **Anosmia, Ageusia** | • New onset COVID associated or idiopathic events without other etiology excluding congenital etiologies or trauma |
| **Subacute thyroiditis** | • Including but not limited to events of atrophic thyroiditis, autoimmune thyroiditis, immune-mediated thyroiditis, silent thyroiditis, thyrotoxicosis, and thyroiditis |
| **Acute pancreatitis** | • Including but not limited to events of autoimmune pancreatitis, immune-mediated pancreatitis, ischemic pancreatitis, edematous pancreatitis, pancreatitis, acute pancreatitis, hemorrhagic pancreatitis, necrotizing pancreatitis, viral pancreatitis, and subacute pancreatitis<br>• Excluding known etiologic causes of pancreatitis (alcohol, gallstones, trauma, recent invasive procedures) |
| **Appendicitis** | • Include any event of appendicitis |
| **Rhabdomyolysis** | • New onset rhabdomyolysis without known etiology such as excessive exercise or trauma |
| **Acute respiratory distress syndrome (ARDS)** | • Including but not limited to new events of ARDS and respiratory failure. |
| **Coagulation disorders** | • Including but not limited to thromboembolic and bleeding disorders, disseminated intravascular coagulation, pulmonary embolism, and deep vein thrombosis |
| **Acute cardiovascular injury** | • Including but not limited to myocarditis, pericarditis, microangiopathy, coronary artery disease, arrhythmia, stress cardiomyopathy, heart failure, or acute myocardial infarction |

| Medical Concept | Additional Notes |
|---|---|
| **Acute kidney injury** | • Include events with idiopathic or autoimmune etiologies<br>• Exclude events with clear alternate etiology (trauma, infection, tumor, or iatrogenic causes such as medications or radiocontrast etc)<br>• Include all cases that meet the following criteria<br>o Increase in serum creatinine by ≥ 0.3 mg/dL (≥ 26.5 µmol/L) within 48 hours;<br>OR<br>o Increase in serum creatinine to ≥ 1.5 times baseline, known or presumed to have occurred within prior 7 days<br>OR<br>o Urine volume ≤ 0.5 mL/kg/hour for 6 hours |
| **Acute liver injury** | • Include events with idiopathic or autoimmune etiologies<br>• Exclude events with clear alternate etiology (trauma, infection, tumor, etc)<br>• Include all cases that meet the following criteria:<br>o > 3-fold elevation above the upper normal limit for ALT or AST<br>OR<br>o > 2-fold elevation above the upper normal limit for total serum bilirubin or gamma glutamyltransferase or ALP |
| **Dermatologic findings** | • Chilblain-like lesions<br>• Single organ cutaneous vasculitis<br>• Erythema multiforme<br>• Bullous rashes<br>• Severe cutaneous adverse reactions including but not limited to: Stevens-Johnson Syndrome (SJS), Toxic Epidermal Necrolysis (TEN), Drug Reaction with Eosinophilia and Systemic Symptoms (DRESS) and fixed-drug eruptions |
| **Multisystem inflammatory disorders** | • Multisystem inflammatory syndrome in adults (MIS-A)<br>• Multisystem inflammatory syndrome in children (MIS-C)<br>• Kawasaki's disease |

| Medical Concept | Additional Notes |
|---|---|
| **Thrombocytopenia** | • Platelet counts < 150 × $10^9$<br>• Including but not limited to immune thrombocytopenia, platelet production decreased, thrombocytopenia, thrombocytopenic purpura, thrombotic thrombocytopenic purpura, or hemolysis, elevated liver enzymes, low platelet count syndrome |
| **Acute aseptic arthritis** | • New onset aseptic arthritis without clear alternate etiology (eg, gout, osteoarthritis, and trauma) |
| **New onset of or worsening of neurologic disease** | • Including but not limited to:<br>o Guillain-Barre Syndrome<br>o Acute disseminated encephalomyelitis<br>o Peripheral facial nerve palsy (Bell's palsy)<br>o Transverse myelitis<br>o Encephalitis/Encephalomyelitis<br>o Aseptic meningitis<br>o Febrile seizures<br>o Generalized seizures/convulsions<br>o Stroke (hemorrhagic and nonhemorrhagic)<br>o Narcolepsy |
| **Anaphylaxis** | • Anaphylaxis as defined per protocol<br>• Follow reporting procedures per protocol |
| **Other syndromes** | • Fibromyalgia<br>• Postural Orthostatic Tachycardia Syndrome<br>• Chronic Fatigue Syndrome (includes myalgic encephalomyelitis and Post-Viral Fatigue Syndrome)<br>• Myasthenia gravis |

#### 11.6. APPENDIX 6: Protocol Amendment History

The document history table for this protocol and the Protocol Amendment Summary of Changes Table for the current Amendment 9 is located directly before the Table of Contents.

A description of Amendment 8, Amendment 7, Amendment 6, Amendment 5, Amendment 4, Amendment 3, Amendment 2, and Amendment 1 is presented in this appendix.

**Amendment 8, 23 Mar 2021**

**Main Rationale for the Amendment:**

The purpose of this amendment is to update language around unblinding and open-label dosing in the context of the majority of participants already having completed their blinded follow-up. In addition, the amendment updates the reporting of serious COVID-19 cases.

The summary of changes table provided here describes the major changes made in Amendment 8 relative to Amendment 7, including the sections modified and the corresponding rationales. As applicable, the synopsis of Amendment 8 has been modified to correspond to changes in the body of the protocol.

##### Summary of Major Changes from Protocol Amendment 7 to Protocol Amendment 8:

| Section # and Name | Description of Change | Brief Rationale |
|---|---|---|
| Section 4.1.2 (Part B, the Open-Label Observational Phase) | States that, due to ethical and scientific concerns, all participants will be unblinded once investigational product is no longer available. In addition, the text indicates that if participants want to receive emergency use authorization (EUA) vaccine outside of the study, participants will need to be withdrawn from the study.<br><br>The amendment also provides directions for unblinding after Biologics License Application (BLA) database lock. | At the time of this amendment, more than 95% of the enrolled participants have completed blinded follow-up, either through being unblinded or study discontinuation. Therefore, there is no scientific value for participants to remain blinded.<br>In addition, non-study EUA vaccines are widely available, so ethically, participants should be unblinded to ensure they can make an informed choice if they wish to receive EUA COVID-19 vaccine outside of the study. |
| Section 8.3.7 (Time Period and Frequency for collecting Adverse Events and Serious Adverse Event Information) and Section 8.3.10 (Reporting Adverse Events) | Reporting of only serious COVID-19 cases within 24 hours to Sponsor or it's designee. | Nonserious COVID-19 cases are an efficacy endpoint, so reporting to Pharmacovigilance is no longer warranted. |

##### Amendment 7, 10 Feb 2021

##### Main Rationale for the Amendment:

The purpose of this amendment is to collect safety information on suspected cases of anaphylaxis and to include participant history of facial injections or dermal fillers in the eDiary.

The summary of changes table provided here describes the major changes made in Amendment 7 relative to Amendment 6, including the sections modified and the corresponding rationales. The synopsis of Amendment 7 has been modified to correspond to changes in the body of the protocol.

#### Summary of Major Changes from Protocol Amendment 6 to Protocol Amendment 7:

| Section # and Name | Description of Change | Brief Rationale |
|---|---|---|
| Title Page, Protocol Approval Page, Headers, Protocol Amendment Summary of Changes | Updated the protocol version and date. | To reflect the new version and date of the protocol. |
| Sections 1.1 (Synopsis),<br>6.2.2 (Administration of Investigational Product),<br>6.2.8.1 (Planned Unblinding) | Clarified that study site personnel who were blinded during the Blinded Phase will be unblinded **at the participant level** at the Participant Decision clinic visit. | To clarify that the site personnel is being unblinded only at the Participant Decision clinic visit. |
| Sections 1.1 (Synopsis), 4.1.2 (Part B, the Open-Label Observational Phase) | Clarified that with regards to EUA eligibility in Part B, the CDC-EUA guidance specifications will supersede eligibility criteria in the protocol. | To clarify that the CDC-EUA guidance specifications supersede protocol-specified criteria for EUA eligibility in Part B. |
| Section 4.1 (General Design) | Corrected the number of scheduled clinic visits and added mention of the participant decision visit. | To clarify the number of scheduled visits. |
| Section 4.1.2 (Part B, the Open-Label Observational Phase) | Updated text to clarify requirements for Part B subjects who are unblinded and received only 1 dose of mRNA-1273 during Part A. | Clarification of requirements for Part B subjects who received only 1 dose of mRNA-1273 during Part A. |
| Section 6.2.4 (Packaging and Labeling) | Updated to indicate that vaccine product is packaged in a 10R glass vial with a 6-dose, 5.0-mL or 10-dose, 6.3-mL fill volume. | To include IP presentation for additional supplies that may be used in Part B of the study. |
| Section 6.2.5 (Storage) | Updated to specify storage requirements for the 6-dose and 10-dose mRNA vaccine. | To include IP storage conditions for additional supplies that may be used in Part B of the study. |
| Sections 6.4.2 (Concomitant Medications and Therapies),<br>7.2 (Discontinuation of Study Treatment), 8.2.2 (Use of Electronic Diaries), 8.2.6 (Blood Sampling Volumes, Table 4) | Clarified requirements for Part A and Part B. | Clarification of concomitant medication, eDiary, and blood sampling procedures during Part A and Part B. |

| Section # and Name | Description of Change | Brief Rationale |
|---|---|---|
| Section 8.2.2 (Use of Electronic Diaries), Appendix 1 (Schedule of Events, Table 17 footnote 7). | Clarified that eDiary prompts to surveil for weekly COVID-19 symptoms will be performed for all participants in Part A and Part B.<br><br>Added eDiary prompt to solicit collection of participant history of facial injections or dermal fillers. | Clarification.<br><br>To assure all concomitant medications are recorded. |
| Section 8.2.4 (Physical Examination) | Removed body mass index from the participant decision visit. | Clarification. |
| Section 8.2.6 (Blood Sampling Volumes) | Updated maximum planned volume of blood collection for the complete study to include a range. | Clarification. |
| Section 8.3.2 (Medically Attended Adverse Events) | Added suspected cases of anaphylaxis as an MAAE and requirement to report as an SAE. | Sponsor collecting anaphylaxis information. |
| Section 9.4 (Analysis Populations, Table 8); Synopsis | Clarified Immunogenicity Subset | Clarification |
| Section 9.5.1.1 (Efficacy Analysis on Primary Endpoint), Appendix 4 (Statistical Appendices, Table 23 and Table 24) | Updated intercurrent events and estimands. | Clarification of estimand. |
| Section 9.5.1.3 (Long-term Efficacy Analysis); Synopsis | Updated case count parameters. | Provide long-term efficacy analysis strategy relevant to change in study design. |
| Appendix 1 (Schedule of Events, Table 21 footnote 2, and Table 22 footnote 2) | Updated physical examination to symptom-directed physical examination and removed height and weight requirements from the Open-Label Clinic Visits. | Clarification |

#### Amendment 6, 23 Dec 2020

#### Main Rationale for the Amendment:

The purpose of this amendment is to inform all ongoing study participants of the availability of and eligibility criteria of any COVID-19 vaccine made available under an EUA and to offer participants who originally received placebo in this study the potential benefit of vaccination against COVID-19, given that the primary efficacy endpoint for mRNA-1273 against COVID-19 was met per the protocol-defined IA.

The summary of changes table provided here describes the major changes made in Amendment 6 relative to Amendment 5, including the sections modified and the corresponding rationales. The synopsis of Amendment 6 has been modified to correspond to changes in the body of the protocol.

**Summary of Major Changes from Protocol Amendment 5 to Protocol Amendment 6:**

| Section # and Name | Description of Change | Brief Rationale |
|---|---|---|
| Title Page, Protocol Approval Page, Headers, Protocol Amendment Summary of Changes | Updated the protocol version and date | To reflect the new version and date of the protocol |
| Sections 1.1 (Synopsis), 1.2 (Schema), 4.1.2 (Part B, the Open-Label Observational Phase), 6.2.8.1 (Planned Unblinding), 7.3.1 (Participant Withdrawal), and 11.1 (Schedules of Events) | Added a “Participant Decision clinic visit”. | This visit provides the opportunity for study site personnel to discuss with and offer to participants, the choice to be unblinded, as well as offering to participants who originally received placebo, the choice to receive active vaccination with mRNA-1273 and possible vaccination against COVID-19. Participants will also sign a revised informed consent form at this visit. |
| Sections 1.1 (Synopsis), 1.2 (Schema), 4.1 (General Design), 4.2 (Scientific Rationale for Study), 6.2 (Method of Randomly Assigning Participants to Treatment Groups [Part A], Blinded Phase Only), and 11.1 (Schedules of Events) | Changes to study design that split the study into a Blinded Phase and an Open-Label Observational Phase. | These changes accommodate the breaking of the blind and offering of open-label vaccination to all participants. This change also distinguishes the Open-Label Supplemental Schedule of Events (Vaccination Phase) for participants who received placebo, and who meet EUA eligibility, and request to receive active vaccine. |

| Section # and Name | Description of Change | Brief Rationale |
|---|---|---|
| Sections 1.1 (Synopsis), 1.2 (Schema), 4.1.2 (Part B, the Open-Label Observational Phase), and 11.1 (Schedules of Events) | Added a Supplemental SoE and a Modified Supplemental SoE | These provide paths for participants who choose to be unblinded and receive mRNA-1273, and for participants who previously received only 1 dose of blinded mRNA-1273 during the study (Blinded Phase), to transition into the Open-label Phase of the study. |
| Section 2.2.2 (Clinical Studies) | Updated status of ongoing clinical studies, including this study (mRNA-1273-P301). | The status of the 3 clinical studies (one Phase 1, one Phase 2a, and this Phase 3 study) have changed since Amendment 5.<br>In addition, the results of the interim analyses in this study of the primary efficacy endpoint (prevention of COVID-19 infection), a major secondary endpoint (prevention of severe COVID-19), and safety and reactogenicity endpoints are now available and are provided here. These results provide the justification for offering participants the opportunity to receive active IP (mRNA-1273) and the potential benefit of vaccination against COVID-19. |

| Section # and Name | Description of Change | Brief Rationale |
|---|---|---|
| Section 1.1 (Synopsis) and 9.5.1.3 (Long-term Efficacy Analysis) | Addition of long-term efficacy analyses | Allows for the analysis of the long-term efficacy of mRNA-1273 in the following treatment cohorts:<br>• mRNA-1273 Cohort: Participants randomized to mRNA-1273 in the Blinded Phase.<br>• Placebo Cohort: Participants randomized to Placebo in the Blinded Phase and did not crossover to mRNA-1273 in the Open-label Phase.<br>• Placebo-mRNA-1273 Cohort: Participants randomized to Placebo in the Blinded Phase and crossed over to mRNA-1273 in the Open-label Phase. |
| Section 2.3.3 (Overall Benefit/Risk Conclusion) | Updated the Benefit/Risk Assessment | Based on the interim results from this pivotal Phase 3 study, mRNA-1273 prevents COVID-19 and severe COVID-19. The demonstrated clinical benefit of mRNA-1273 is supported by evidence of a robust immune response both in terms of binding antibodies (bAbs) and neutralizing antibodies (nAbs) as well as the induction of CD4+ T-cells with a Th-1 dominant phenotype. |
| Section 11.1 | Added footnote to Table 16 | To clarify that the eDiary Safety prompts will be triggering off on Day 61 to take into consideration the (-3 day) window allowance on Day 29. |

**Amendment 5, 11 Nov 2020**

**Main Rationale for the Amendment:**

The main purpose of this amendment is to clarify that the eDiary prompts for safety surveillance will be weekly and to add Month 19 safety call.

The summary of changes table provided here describes the major changes made in Amendment 5 relative to Amendment 4, including the sections modified and the corresponding rationales. The synopsis of Amendment 5 has been modified to correspond to changes in the body of the protocol.

**Summary of Major Changes from Protocol Amendment 4 to Protocol Amendment 5:**

| Section # and Name | Description of Change | Brief Rationale |
|---|---|---|
| Title Page, Protocol Approval Page, Headers, Protocol Amendment Summary of Changes | Updated the protocol version and date | To reflect the new version and date of the protocol |
| Section 11.1 | Updated Appendix 1 (Tables 16 and 17) | To clarify the weekly schedule for eDiary prompts through Year 1 and Year 2 of follow-up |
| Section 11.1 | Added Month 19 Safety Call | To clarify that participants will have continuous weekly (eDiary prompts) and monthly (safety calls) through Year 1 and Year 2 of follow-up |

**Amendment 4, 30 Sep 2020**

**Main Rationale for the Amendment:**

The main purpose of this amendment is to increase the upper limit for stratification of enrolled participants considered “at risk” at Screening to 50%.

The summary of changes table provided here describes the major changes made in Amendment 4 relative to Amendment 3, including the sections modified and the corresponding rationales. The synopsis of Amendment 4 has been modified to correspond to changes in the body of the protocol.

**Summary of Major Changes from Protocol Amendment 3 to Protocol Amendment 4:**

| Section # and Name | Description of Change | Brief Rationale |
|---|---|---|
| Title Page, Protocol Approval Page, Headers, Protocol Amendment Summary of Changes | Updated the protocol version and date | To reflect the new version and date of the protocol |
| Section 6.2.1.1 (Stratification) | Increased the upper limit for stratification of enrolled participants considered "at risk" at Screening to up to 50%, from 40%. | To enhance the diversity of the study population by increasing the number of racial and ethnic minority participants in the study. as participants from these communities often have higher rates of comorbidities. |
| Section 9.5.1.1 (Efficacy Analysis on Primary Endpoint) and Section 11.4.1 (Estimands and Estimand Specifications) | Updated the description of an intercurrent event (unrelated death) and its strategy | To align with the description in the Statistical Analysis Plan. |

**Amendment 3, 20 Aug 2020**

**Main Rationale for the Amendment:**

The main purpose of this amendment is to make changes to the protocol in response to feedback from CBER.

The summary of changes table provided here describes the major changes made in Amendment 3 relative to Amendment 2, including the sections modified and the corresponding rationales. Minor editorial or formatting changes are not included in this summary table. The synopsis of Amendment 3 has been modified to correspond to changes in the body of the protocol.

**Summary of Major Changes from Protocol Amendment 2 to Protocol Amendment 3:**

| Section # and Name | Description of Change | Brief Rationale |
|---|---|---|
| Title Page, Protocol Approval Page, Headers, Protocol Amendment Summary of Changes | Updated the protocol version and date | To reflect the new version and date of the protocol |
| Section 5 (Study Population) | Added a sentence to describe the intent to enroll a representative sample of racial and ethnic minority participants in the study | To enhance the diversity of the study population |
| Section 5.2 (Exclusion Criteria) | Added clarification to exclusion criterion #11 to define the parameters based on screening CD4 count and viral load for exclusion of study participants | To clarify the definition of controlled HIV disease in the exclusion criterion such that only participants with well-controlled HIV disease are enrolled in the study |

| Section # and Name | Description of Change | Brief Rationale |
|---|---|---|
| Section 5.2 (Exclusion Criteria) | Removed “topical tacrolimus” from exclusion criterion #12 | No evidence to support any systemic effect of topical tacrolimus to warrant excluding them |
| Section 6.2.1.1 (Stratification) | Added HIV infection to the risk factors at Screening | To stratify participants based on certain risk factors |
| Section 8.2.3 (Demographics/Medical History) | Added collection of risk factors for complications of COVID-19 | To document the diagnosis of any risk factor for complications of COVID19 used for stratification |
| Section 9.3 (Sample Size Determination) | Removed redundant bullet | To remove redundancy in the assumptions listed |
| Section 9.5.2 (Safety Analyses) | Removed safety analysis by serostatus | No added value for this analysis. Other subgroup analyses may be specified in the SAP, as needed. |
| Section 9.5.5 (Subgroup Analyses) | Removed the categories (white, non-white) from the Race Variable | To allow for more refined Race categorization being collected in the eCRF |
| Appendix 11.3 | Removed “cessation of exogenous hormonal therapy” | To allow postmenopausal women to take these medications if needed for the treatment of the symptoms of menopause |
| Appendix 11.3 | Removed “using hormonal contraception” from postmenopausal female with high FSH levels | Not a standard of care for women for treatment of menopausal symptoms |

**Amendment 2, 31 Jul 2020:**

**Main Rationale for the Amendment:**

The main purpose of this amendment is to provide more intensive surveillance of symptoms and severity of cases of COVID-19 after the first dose of investigational product.

The summary of changes table provided here describes the major changes made in Amendment 2 relative to Amendment 1, including the sections modified and the corresponding rationales. Minor editorial or formatting changes are not included in this summary table. The synopsis of Amendment 2 has been modified to correspond to changes in the body of the protocol.

**Summary of Major Changes from Protocol Amendment 1 to Protocol Amendment 2:**

| Section # and Name | Description of Change | Brief Rationale |
|---|---|---|
| Title Page, Protocol Approval Page, Headers, Protocol Amendment Summary of Changes | Updated the protocol version and date | Reflect the new version and date of the protocol |

| Section # and Name | Description of Change | Brief Rationale |
|---|---|---|
| Section 4.1 (General Design), Section 7.2 (Discontinuation of Study Treatment), Section 8.1.1 (Efficacy Assessments Related to COVID-19 and SARS-CoV-2 Infection), Section 8.1.2 (Surveillance for COVID-19 Symptoms),Table 14 (Schedule of Events [Vaccination Phase, Day 1 – Day 57]) | Added a Day 29 NP swab prior to Dose 2 | Improve surveillance for asymptomatic infection prior to Dose 2 to assist in discriminating COVID-19 symptoms from solicited systemic reactions after vaccination |
| Section 4.1 (General Design), Table 15 (Schedule of Events [Surveillance Phase, Day 64 – Day 394]) | Changed the clinic visit at Month 4 to a safety call | Improve participant safety and adherence to protocol |
| Section 4.1 (General Design), Section 8.1.2 (Surveillance for COVID-19 Symptoms), Table 14 Footnote 5 (Schedule of Events [Vaccination Phase, Day 1 – Day 57]) | Clarified symptom duration (> 48 hours) to trigger NP swab collection and investigator judgement whether to obtain an NP swab in the 7 days following vaccination due to overlap of solicited systemic symptoms and COVID-19 | Improve surveillance for cases of COVID-19 |
| Section 4.1 (General Design), Section 8.1.3 (Convalescent Period Starting with the Illness Visit), Section 8.1.4 (Ancillary Supplies for Participant Use) | Removed mention of continuous biometric monitoring | Simplify data management and clinical operations |
| Section 5.1 (Inclusion Criteria), Section | Removed the inclusion criterion regarding male contraception | Requirement not generally applicable for a Phase 3 vaccine study |
| Section 5.2 (Exclusion Criteria), Section 6.4.3 (Concomitant Medications and Vaccines that May Lead to the Elimination of a Participant from Per-Protocol Analyses) | Clarified language around influenza vaccination | Make restrictions on influenza vaccination less restrictive than for other licensed vaccines relative to administration of investigational product |
| Section 5.2 (Exclusion Criteria) | Removed restriction on enrollment of participants with human immunodeficiency virus (HIV) infection | Participants on stable antiretroviral therapy are not excluded, which diversifies the participant group |
| Section 8.3.7 (Time Period and Frequency for Collecting AE and SAE Information), Section 8.3.10 (Reporting Adverse Events) | Added expedited reporting of confirmed COVID-19 cases | Improve surveillance for cases of COVID-19 |

| Section # and Name | Description of Change | Brief Rationale |
|---|---|---|
| Section 8.4.2 (Data and Safety Monitoring Board) | Added an Oversight Group | Clarify responsibility for declaring early efficacy or for taking action to stop, pause, or continue the study |
| Section 8.4.2 (Data and Safety Monitoring Board) | Updated description of monitoring for potential harm, including details on case counting for the purpose of harm monitoring to align with a DSMB analysis plan | Clarify description for harm monitoring, based on FDA feedback |

**Amendment 1, 26 Jun 2020:**

**Main Rationale for the Amendment:**

The main purpose of this amendment is to provide more intensive surveillance of symptoms and severity of cases of COVID-19 after the first dose of investigational product.

The summary of changes table provided here describes the major changes made in Amendment 1 relative to the original protocol, including the sections modified and the corresponding rationales. Minor editorial or formatting changes are not included in this summary table.

**Summary of Major Changes in Protocol Amendment 1:**

| Section # and Name | Description of Change | Brief Rationale |
|---|---|---|
| Title page, Signature page, and header | Updated the protocol version and date | Reflect the new version and date of the protocol |
| Synopsis; Section 4.1, General Design; Section 8.1.3, Convalescent Period; Section 11.1, schedules of events, Table 17 | Increased the frequency of telemedicine contacts during the Convalescent Period following the Initial Illness Visit | Improve the amount and quality of COVID-19 symptom data collected during the Convalescent Period |
| Synopsis; Section 4.1, General Design; Section 8.1.3, Convalescent Period; Section 8.1.4, Ancillary Supplies for Participant Use; Section 11.1, schedules of events, Table 17 | Added monitoring of oxygen saturation to the Convalescent Period | Improve surveillance for incidence of COVID-19 during the study |
| Synopsis; Section 8.1.3, Convalescent Period; Section 8.1.4, Ancillary Supplies for Participant Use; Section 11.1, schedules of events, Table 17 | Increased the frequency of monitoring for SARS-CoV-2, using saliva as the preferred sample matrix after the Illness Visit during the Convalescent Period | Improve the sensitivity of monitoring the time course of viral shedding during COVID-19 |

| Section # and Name | Description of Change | Brief Rationale |
|---|---|---|
| Synopsis; Section 3, Objectives and Endpoints | Added a respiratory sample for hospitalized participants as a matrix for confirming the presence of SARS-CoV-2 | Increase the potential number of evaluable COVID-19 cases |
| Synopsis; Section 3, Objectives and Endpoints | Broadened the definition for seroconversion at a participant level | Included neutralizing antibody (nAb) in addition to binding antibody (bAb) |
| Section 8, Study Assessments and Procedures; Section 8.1.3, Convalescent Period; Section 8.1.4, Ancillary Supplies for Participant Use; Section 8.2.2, Use of Electronic Diaries; Section 11.1, schedules of events, Table 17 | Eliminated paper diaries, substituting an instruction card listing symptoms and a severity grading system to enhance the quality of data obtained by telemedicine contacts | Reduce fomite transmission of SARS-CoV-2 and increase frequency of investigative staff interactions with participants during the Convalescent Period |
| Section 8.1.2, Surveillance for COVID-19 Symptoms | Decreased the number of symptoms (to one of the 11 CDC symptoms) that would result in an Illness Visit | Increase the likelihood of capturing all COVID-19 cases in the earliest stage of disease |
| Section 8.2.2, Use of Electronic Diaries | Expanded the scope of eDiary prompts and data collected during the Surveillance Phase | Increase the likelihood of capturing all COVID-19 cases in the earliest stage of disease |

Signature Page for VV-CLIN-003185 v2.0

| | |
|---|---|
| Approval | Allison August<br>Medical<br>13-Sep-2021 16:26:58 GMT+0000 |

Signature Page for VV-CLIN-003185 v2.0

**CLINICAL STUDY PROTOCOL**

| | |
|---|---|
| **Protocol Title:** | A Phase 2/3, Randomized, Observer-Blind, Placebo-Controlled Study to Evaluate the Safety, Reactogenicity, and Effectiveness of mRNA-1273 SARS-CoV-2 Vaccine in Healthy Adolescents 12 to < 18 Years of Age |
| **Protocol Number:** | mRNA-1273-P203 |
| **Sponsor Name:** | ModernaTX, Inc. |
| **Legal Registered Address:** | 200 Technology Square<br>Cambridge, MA 02139 |
| **Sponsor Contact and Medical Monitor:** | Amparo Figueroa, MD, MPH<br>ModernaTX, Inc.<br>200 Technology Square<br>Cambridge, MA 02139<br>Telephone: 1-617-301-1186<br> |
| **Regulatory Agency Identifier Number(s):** | IND: 019745 |
| **Amendment Number:** | 3 |
| **Date of Amendment 3:** | 04 Nov 2021 |
| **Date of Amendment 2:** | 27 Jul 2021 |
| **Date of Amendment 1:** | 23 Mar 2021 |
| **Date of Original Protocol:** | 04 Nov 2020 |

**CONFIDENTIAL**

All financial and nonfinancial support for this study will be provided by ModernaTX, Inc. The concepts and information contained in this document or generated during the study are considered proprietary and may not be disclosed in whole or in part without the expressed written consent of ModernaTX, Inc. The study will be conducted according to the *International Council for Harmonisation (ICH) of Technical Requirements for Registration of Pharmaceuticals for Human Use, E6(R2) Good Clinical Practice (GCP) Guidance.*

#### PROTOCOL APPROVAL – SPONSOR SIGNATORIES

**Study Title:** A Phase 2/3, Randomized, Observer-Blind, Placebo-Controlled Study to Evaluate the Safety, Reactogenicity, and Effectiveness of mRNA-1273 SARS-CoV-2 Vaccine in Healthy Adolescents 12 to < 18 Years of Age

**Protocol Number:** mRNA-1273-P203

**Amendment Number:** 3

**Amendment 3 Date:** 04 Nov 2021

Protocol accepted and approved by:

**See esignature and date signed on the last page of the document.**

| | |
|---|---|
| Rituparna Das, MD, PhD<br>Vice President Clinical Development, Infectious Diseases<br>ModernaTX, Inc.<br>200 Technology Square<br>Cambridge, MA 02139<br>Telephone: 1-617-710-9794 | Date |

### DECLARATION OF INVESTIGATOR

I have read and understood all sections of the protocol entitled "A Phase 2/3, Randomized, Observer-Blind, Placebo-Controlled Study to Evaluate the Safety, Reactogenicity, and Effectiveness of mRNA-1273 SARS-CoV-2 Vaccine in Healthy Adolescents 12 to < 18 Years of Age" and the most recent version of the investigator's brochure.

I agree to supervise all aspects of the protocol and to conduct the clinical investigation in accordance with the current Protocol, the *International Council for Harmonisation (ICH) of Technical Requirements for Registration of Pharmaceuticals for Human Use, E6(R2) Good Clinical Practice (GCP) Guidance*, and all applicable government regulations. I will not make changes to the protocol before consulting with ModernaTX, Inc. or implement protocol changes without Institutional Review Board (IRB) approval except to eliminate an immediate risk to participants.

I agree to administer study treatment only to participants under my personal supervision or the supervision of a sub-investigator. I will not supply study treatment to any person not authorized to receive it. I also agree that persons debarred from conducting or working on clinical studies by any court or regulatory agency will not be allowed to conduct or work on studies for the Sponsor or a partnership in which the Sponsor is involved. I will immediately disclose it in writing to the Sponsor if any person who is involved in the study is debarred, or if any proceeding for debarment is pending, or, to the best of my knowledge, threatened.

I will not disclose confidential information contained in this document including participant information, to anyone other than the recipient study staffs and members of the IRB. I agree to ensure that this information will not be used for any purpose other than the evaluation or conduct of the clinical investigation without the prior written consent from ModernaTX, Inc. I will not disclose information regarding this clinical investigation or publish results of the investigation without authorization from ModernaTX, Inc.

The signature below provides the necessary assurance that this study will be conducted according to all stipulations of the protocol, including statements regarding confidentiality, and according to local legal and regulatory requirements, US federal regulations, and ICH E6(R2) GCP guidelines.

____________________________________ ____________________

Signature of principal investigator Date

____________________________________

Printed name of principal investigator

### Protocol Amendment Summary of Changes

| DOCUMENT HISTORY | |
|---|---|
| **Document** | **Date** |
| Amendment 3 | 04 Nov 2021 |
| Amendment 2 | 27 Jul 2021 |
| Amendment 1 | 23 Mar 2021 |
| Original Protocol | 04 Nov 2020 |

**Amendment 3, 04 Nov 2021: Current Amendment**

This amendment is considered to be substantial based on the criteria set forth in Article 10(a) of Directive 2001/20/EC of the European Parliament and the Council of the European Union.

**Main Rationale for the Amendment:**

To provide a 50 µg booster dose (BD) of mRNA-1273 to participants. This change is prompted by new interim data from an ongoing Moderna Phase 2 study, mRNA-1273-P201 (Study P201). Adult participants in Study mRNA-1273-P201 received 2 doses of either 50 µg or 100 µg of mRNA-1273 and were administered a 50 µg booster of mRNA-1273 6 to 8 months after the second dose. Participants in mRNA-1273-P201 who received the BD, demonstrated enhanced immune responses to SARS-CoV-2 compared to pre-boost levels and met the noninferiority criteria stipulated in the US Food and Drug Administration Guidance on Emergency Use Authorization for Vaccines to Prevent COVID-19. Additionally, no new safety signals emerged upon administration of the BD in Study P201. Based on cumulative evidence, the benefit-risk profile of a BD of mRNA-1273 is favorable, particularly in light of increasing breakthrough disease with the emergence of the Delta variant. Providing the option for a BD to all eligible participants currently enrolled is expected to generate valuable booster data in the adolescent population.

**Summary of Major Changes in Protocol Amendment 3:**

The Summary of Changes table provided below describes the major changes made in Protocol Amendment 3 relative to Protocol Amendment 2, including the sections modified and the corresponding rationales. The synopsis of Protocol Amendment 3 has been modified to correspond to changes in the body of the protocol. Minor grammar and formatting corrections were made throughout the document to enhance clarity and readability (which did not affect the conduct of the study).

| Section # and Name | Description of Change | Brief Rationale |
|---|---|---|
| Title Page, Signature page, Protocol Amendment Summary of Changes, and Header | Updated the protocol version and date | Updated to reflect the new version and date. |
| Synopsis, Section 4 (Study Population [Part A: Blinded Phase and Part B: Open Label Observational Phase]) | Updated the estimated date last participant completed and total number of sites | Changes made to reflect the confirmation of study sites and updated participant timeline. |
| Synopsis, Section 3.1 (General Design) | Extended the study duration | Changes made to reflect the addition of a new study part (Part C). |
| Synopsis, Section 2 (Objectives and Endpoints) | Added exploratory objectives for Part B | Changes made to reflect the planned analyses |
| | Added primary, key secondary, and exploratory objectives for Part C | Changes made to reflect the addition of a new study part (Part C). |
| Synopsis, Section 1.3.2 (Risks to Study Participants and Risk Mitigation), Section 1.3.3 (Overall Benefit/Risk Conclusion), Section 2 (Objectives and Endpoints), Section 3.1 (General Design), Section 3.1.3 (Part C, Booster Dose Phase), Section 3.2 (Scientific Rationale for Study Design), Section 3.3 (Justification for Dose, Control Product, and Choice of Study Population), Section 4.3 (Study Eligibility Criteria [Part C]), Section 5.1 (Investigational Product Administered), Section 5.3.1 (Preparation of Study Vaccine for Injection), Section 5.3.2 (Administration of Study Vaccine), Section 5.3.4 (Study Vaccine Packaging and Labeling), Section 5.4 (Study Treatment Compliance), Section 6.4 (Study Pause Rules), Section 7 (Study Assessments and Procedures), Section 7.1 (Safety Assessments and Procedures), Section 7.1.1 (Use of Electronic | Part C is being added to Study P203.<br>Part C evaluates the safety and immunogenicity of a third booster dose of 50 µg of mRNA-1273 in adolescents who have previously received 2 doses of mRNA-1273 as a primary series. | Changes made to reflect the addition of a new study part (Part C) to continue to evaluate the benefit-risk profile of a booster dose of mRNA-1273. |

| | | |
|---|---|---|
| Diaries), Section 8.1 (Blinding and Responsibility for Analyses), Section 8.2 (Statistical Hypothesis), Section 8.3 (Power and Sample Size), Section 8.5 (Statistical Methods), Section 10.1 (APPENDIX 1: Schedule of Events; Table 10), Section 10.2.8 (Protocol Deviations) | | |
| Synopsis, Section 3.1.1 (Part A, the Blinded Phase), Section 3.1.2 (Part B, the Open-label Observational Phase), Section 3.2 (Scientific Rationale for Study Design), Section 7.1.6 (Assessment for SARS-CoV-2 Infection), Section 7.3 (Exploratory Assessments and Biomarkers), Section 7.4.1 (Vaccine Effectiveness Assessments), Section 7.4.2 (Surveillance for COVID-19 Symptoms), Section 7.4.3 (Follow up/Convalescent Period After Diagnosis with COVID-19), Section 10.1 (APPENDIX 1: Schedule of Assessments, Table 7, Table 8, Table 9, Table 10) | Add nasal swab to nasopharyngeal at all time points | To align with the original unblinding plan. |
| Synopsis, Section 5.3.1 (Preparation of Study Vaccine for Injection), Section 5.3.2 (Administration of Study Vaccine), Section 5.3.4 (Study Vaccine Packaging and Labeling) | Part C language was added to discuss the difference in volume | Changes made to reflect different vaccine volumes given during the primary series and the boost. |
| Synopsis, Section 7.3 (Exploratory Assessments and Biomarkers) | Addition of Exploratory Assessments and Biomarkers | To align language with other clinical studies. |
| Synopsis, Section 8.5.5 (Long-term Analysis [including Part B]), Section 8.5.6 (Booster Phase Analysis [Part C]) | Updated long-term analysis and booster phase analysis | Changes made to reflect the addition of a booster dose. |
| Synopsis, Section 8.6.1 (Interim Analyses) | Addition of booster interim analyses | Changes made to reflect the addition of a booster dose. |
| Section 1.1 (Study Rationale), Section 1.2.2 (Clinical Studies) | Added updated clinical information of mRNA-1273 | Changes made to reflect new and ongoing clinical |

| | | |
|---|---|---|
| | | data in the current and other studies. |
| Section (5.3.5) Study Vaccine Storage | Updated storage conditions to follow the investigational product label | Changes made to reflect differences in storage conditions between different vial volumes. |
| Section 5.5.4 (Concomitant Medications and Vaccines that May Lead to the Elimination of a Participant from Per-Protocol Analyses) | Added receipt or planned receipt of a non-study COVID-19 would require withdrawal from the study | To clarify that participants could not get other COVID-19 vaccines. |
| Section 6.1.1 (Individual Participant Criteria for Delay of Study Vaccination) | Added investigator judgement to rescheduling of visits | To allow for flexibility of visits. |
| Section 7.1.6 (Assessment for SARS-CoV-2 Infection), Section 7.4.2 (Surveillance for COVID-19 Symptoms), Section 7.4.3 (Follow up/Convalescent Period After Diagnosis with COVID-19), Section 10.1 (APPENDIX 1 Schedule of Events; Table 9). | Added nasal swab for samples for SARS-CoV-2 | To allow more flexibility for participants. |
| Section 8.5.3 (Immunogenicity Analysis) | Added the primary immunogenicity analysis is performed in Part A and Part C | To clarify that only Part A and Part C will have primary immunogenicity analyses. |

**IRB and Regulatory Authority Approval**

A copy of this amended protocol will be sent to the institutional review board (IRB) and regulatory authority.

The changes described in this amended protocol require IRB approval prior to implementation. In addition, if the changes herein affect the informed consent, sites are required to update and submit a revised informed consent for approval that incorporates the changes described in this amended protocol.

### PROTOCOL SYNOPSIS

**Name of Sponsor/Company:** ModernaTX, Inc.

**Name of Investigational Product:** mRNA-1273 for injection

**Name of Active Ingredient:** mRNA-1273

**Protocol Title:** A Phase 2/3, Randomized, Observer-Blind, Placebo-Controlled Study to Evaluate the Safety, Reactogenicity, and Effectiveness of mRNA-1273 SARS-CoV-2 Vaccine in Healthy Adolescents 12 to < 18 Years of Age

**Protocol Number:** mRNA-1273-P203

**Study Period (months):** Approximately 25 months

**Phase of Development:** Phase 2/3

**Estimated date first participant enrolled:** 30 Nov 2020

**Estimated date last participant completed:** 23 Dec 2022

**Total Number of Sites:** 25 study sites in the United States or its territories.

**Objectives and Endpoints**

| Objectives | Endpoints |
|---|---|
| **Primary Objectives** | **Primary Endpoints** |
| • To evaluate the safety and reactogenicity of 100 µg of mRNA-1273 vaccine administered in 2 doses 28 days apart | • Solicited local and systemic ARs through 7 days after each injection<br>• Unsolicited AEs through 28 days after each injection<br>• MAAEs through the entire study period<br>• SAEs through the entire study period<br>• AESIs through the entire study period<br>• Vital sign measurements<br>• Physical examination findings |

| | |
|---|---|
| • To infer efficacy of mRNA-1273 (100 µg, 2 doses 28 days apart), serum Ab responses obtained 28 days after the second injection of mRNA-1273 (Day 57) will be either:<br>– Evaluated against an accepted Ab threshold of protection against COVID-19 (if established in Study P301)<br>– Compared in primary vaccine response as measured by GM values of serum Ab and seroresponse rate in Study P203 with those obtained from young adult recipients (18-25 years of age) of mRNA-1273 in the clinical endpoint efficacy trial (Study P301) | • The proportion of participants with a serum Ab level at Day 57 ≥ an Ab threshold of protection[1]<br>• The primary vaccine response as measured by GM value of serum Ab level and seroresponse rate from Study P203 vaccine recipients at Day 57 compared with those obtained from young adult recipients (18-25 years of age) at Day 57 in the clinical endpoint efficacy trial (Study P301)[2]<br>1. If an accepted serum Ab threshold of vaccine protection against COVID-19 is available, this analysis will form the basis to infer efficacy<br>2. If a threshold is not available, efficacy will be inferred based on establishing noninferiority of adolescent (12 to < 18 years; this clinical study) to adult GM values of serum Ab and seroresponse rate obtained in Study P301 (GM value 12 to < 18 years / GM value 18-25 years). |
| **Secondary Objectives** | **Secondary Endpoints** |
| • To evaluate the persistence of the immune response of mRNA-1273 vaccine (100 µg) administered in 2 doses 28 days apart, as assessed by the level of SARS-CoV-2 S2P-specific bAb through 1 year after Dose 2 | • The GM value of SARS-CoV-2 S2P-specific bAb on Day 1, Day 57 (1 month after Dose 2), Day 209 (6 months after Dose 2), and Day 394 (1 year after Dose 2) |
| • To evaluate the persistence of the immune response of mRNA-1273 vaccine (100 µg) administered in 2 doses 28 days apart, as assessed by the level of nAb through 1 year after Dose 2 | • The GM values of SARS-CoV-2-specific nAb on Day 1, Day 57 (1 month after Dose 2), Day 209 (6 months after Dose 2), and Day 394 (1 year after Dose 2) |
| • To evaluate the effect of mRNA-1273 on the incidence of SARS-CoV-2 | • The incidence of SARS-CoV-2 infection (symptomatic or asymptomatic infection) |

| | |
|---|---|
| infection compared with the incidence among placebo recipients | counted starting 14 days after the second dose of IP<br>• SARS-CoV-2 infection will be defined in participants with negative SARS-CoV-2 at baseline:<br>– bAb level against SARS-CoV-2 nucleocapsid protein negative at Day 1 that becomes positive (as measured by Roche Elecsys) at Day 57 or later, OR<br>– Positive RT-PCR counted starting 14 days after the second dose of IP |
| • To evaluate the incidence of asymptomatic SARS-CoV-2 infection after vaccination with mRNA-1273 or placebo | • The incidence of asymptomatic SARS-CoV-2 infection measured by RT-PCR and/or bAb levels against SARS-CoV-2 nucleocapsid protein (by Roche Elecsys) counted starting 14 days after the second dose of IP in participants with negative SARS-CoV-2 at baseline |
| • To evaluate the incidence of COVID-19 after vaccination with mRNA-1273 or placebo. COVID-19 is defined as clinical symptoms consistent with SARS-CoV-2 infection AND positive RT-PCR for SARS-CoV-2 | • The incidence of the first occurrence of COVID-19 starting 14 days after the second dose of IP, where COVID-19 is defined as symptomatic disease based on the following criteria:<br>– The participant must have experienced at least TWO of the following systemic symptoms: Fever ($\geq$ 38ºC/$\geq$ 100.4ºF), chills, myalgia, headache, sore throat, new olfactory and taste disorder(s), OR<br>– The participant must have experienced at least ONE of the following respiratory signs/symptoms: cough, shortness of breath or difficulty breathing, OR clinical or radiographical evidence of pneumonia; AND<br>– The participant must have at least 1 NP swab, nasal swab, or saliva sample (or respiratory sample, if |

| | |
|---|---|
| | hospitalized) positive for SARS-CoV-2 by RT-PCR |
| **Exploratory Objectives** | **Exploratory Endpoints** |
| • To evaluate the genetic and/or phenotypic relationships of isolated SARS-CoV-2 strains to the vaccine sequence | • Alignment of genetic sequence of viral isolates with that of the vaccine sequence and comparison of bAb and nAb titers against isolated strain relative to prototype vaccine strain |
| • To describe the ratio or profile of specific bAb relative to nAb in serum | • Relative amounts or profiles of S protein-specific bAb and specific nAb levels/titers in serum |
| • To characterize the clinical profile and immune responses of participants with COVID-19 or with SARS-CoV-2 infection | • Description of clinical severity and immune responses of participants who are identified as infected by SARS-CoV-2 (COVID-19) |
| • To evaluate the incidence of asymptomatic SARS-CoV-2 infection after vaccination with mRNA-1273 or placebo in participants with serologic evidence of infection at baseline | • GM and GMFR of bAb levels against SARS-CoV-2 nucleocapsid protein (quantitative IgG) and % of participants with 2x, 3x, and 4x rise of bAb relative to baseline |
| **Exploratory Objectives for Part B:** | **Exploratory Endpoints for Part B:** |
| • To evaluate the safety of 2 doses of mRNA-1273 | • MAAEs through the entire study period<br>• SAEs through the entire study period<br>• AESIs through the entire study period<br>• AEs leading to discontinuation from study participation through the last day of study participation |
| • To evaluate the incidence of SARS-CoV-2 infection or COVID-19 after vaccination with mRNA-1273 | • The incidence of SARS-CoV-2 infection (symptomatic or asymptomatic infection) counted starting 14 days after the second dose of mRNA-1273<br>• To evaluate the incidence of asymptomatic SARS-CoV-2 infection after vaccination with mRNA-1273 measured by RT-PCR and/or bAb levels against SARS-CoV-2 nucleocapsid protein (by Roche Elecsys) counted starting 14 days after the second |

| | |
|---|---|
| | dose in participants with negative SARS-CoV-2 at baseline<br>• The incidence of the first occurrence of symptomatic COVID-19 starting 14 days after the second dose of mRNA-1273 |
| **Objectives for Part C** | **Endpoints for Part C** |
| **Primary Objectives** | **Primary Endpoints** |
| • To evaluate the safety of the 50 µg BD of mRNA-1273 | • Solicited local and systemic ARs through 7 days after BD<br>• Unsolicited AEs through 28 days after BD injection<br>• MAAEs through the entire study period<br>• SAEs through the entire study period<br>• AESIs through the entire study period<br>• AEs leading to discontinuation from study participation post BD through the last day of study participation |
| • To infer effectiveness of the 50 µg booster of mRNA-1273 by establishing noninferiority of Ab response after the booster dose compared to the primary series of mRNA-1273. GM values of serum Ab and seroresponse rate of post-booster in Study P203 compared with primary series from young adult (18-25 years of age) recipients of mRNA-1273 in the clinical endpoint efficacy trial (Study P301) | • Co-Primary endpoint(s):<br>– GM of post-booster (post-Dose 3) Ab against original strain in Study P203 as compared to post-primary series (post-Dose 2) against original strain in the young adults in Study P301<br>– Seroresponse rate of post-booster/Dose 3 from baseline (pre-Dose 1) in Study P203 as compared to post-Dose 2 from baseline (pre-Dose1) against original strain in the young adults in Study P301, using 4-fold rise definition<br>○ Seroresponse is defined as a titer change from baseline (pre-Dose 1) below the LLOQ to ≥ 4 × LLOQ, |

| | |
|---|---|
| | or at least a 4-fold rise if baseline is ≥ LLOQ |
| **Key Secondary Objective** | **Key Secondary Endpoint** |
| • To evaluate immune response elicited by the 50 µg prototype booster of mRNA-1273 against variant(s) of interest | • GM of post-booster (post-Dose 3) Ab against circulating strain/Delta as compared to post-primary series (post-Dose 2) against circulating strain/Delta<br>• Seroresponse rate of post-booster/Dose 3 from baseline (pre-Dose 1) as compared to post-Dose 2 from baseline (pre-Dose1) against circulating strain/Delta using 4-fold rise definition |
| **Exploratory Objectives** | **Exploratory Endpoints** |
| • To evaluate the persistence of the immune response of the BD of mRNA-1273 vaccine (50 µg) as assessed by the level of SARS-CoV-2 S2P specific bAb through 1 year after BD | • The GM value of SARS-CoV-2 S2P specific bAb on BD-Day 1, BD-Day 22 (1 month after BD), BD-Day 181 (6 months after BD), and BD-Day 361 (1 year after BD) |
| • To evaluate the persistence of the immune response of the BD of mRNA-1273 vaccine (50 µg) as assessed by the level of nAb through 1 year after BD | • The GM values of SARS-CoV-2-specific nAb on BD-Day 1, BD-Day 22 (1 month after BD), BD-Day 181 (6 months after BD), and BD-Day 361 (1 year after BD) |
| • To evaluate the incidence of SARS-CoV-2 infection or COVID-19 after vaccination with mRNA-1273 | • The incidence of SARS-CoV-2 infection (symptomatic or asymptomatic infection) counted starting 14 days after BD of mRNA-1273<br>• To evaluate the incidence of asymptomatic SARS-CoV-2 infection after vaccination with mRNA-1273 measured by RT-PCR and/or bAb levels against SARS-CoV-2 nucleocapsid protein (by Roche Elecsys) counted starting 14 days after BD in participants with negative SARS-CoV-2 at baseline or pre-booster |

| | |
|---|---|
| | • The incidence of the first occurrence of symptomatic COVID-19 starting 14 days after BD of mRNA-1273 |

Abbreviations: Ab = antibody; AE = adverse event; AESI = adverse event of special interest; AR = adverse reaction; bAb = binding antibody; BD = booster dose; COVID-19 = coronavirus disease 2019; GM = geometric mean; GMFR = geometric mean fold-rise; IP = investigational product; LLOQ = lower limit of quantification; LOD = limit of detection; MAAE = medically attended adverse event; nAb = neutralizing antibody; NP = nasopharyngeal; RT-PCR = reverse transcriptase polymerase chain reaction; S = spike; S2P = S protein; SAE = severe adverse event; SARS-CoV-2 = Severe Acute Respiratory Syndrome coronavirus 2.

#### Overall Study Design

This is a three-part, Phase 2/3, study: Part A, Part B, and Part C. The study will evaluate the safety, reactogenicity, and effectiveness of mRNA-1273 Severe Acute Respiratory Syndrome coronavirus 2 (SARS-CoV-2) vaccine in healthy adolescents 12 to < 18 years of age.

Participants in Part A, the Blinded Phase of the study, will be randomly assigned to receive injections of either 100 µg of mRNA-1273 vaccine or a placebo control in a 2:1 randomization ratio. Part B, the Open-label Observational Phase of this study, is designed to offer participants who received placebo in Part A of this study, and who meet Emergency Use Authorization (EUA) eligibility criteria, an option to receive mRNA-1273 in an open-label fashion. Part C, the Booster Dose Phase, is designed to offer participants in Part A and Part B who are at least 6 months from the last dose, the option to request a booster dose (BD) (50 µg) of mRNA-1273.

The goal of the study is to seek an indication for use of mRNA-1273 (100 µg intramuscular [IM], given as 2 injections, 28 days apart) in the 12 to < 18 years age group. The basis for demonstrating vaccine effectiveness is proposed to be met by serum antibody (Ab) response measured in this adolescent age group. The approach to inferring vaccine effectiveness will depend on whether an accepted serum Ab threshold conferring protection against coronavirus disease 2019 (COVID-19) has been established. If an Ab threshold of protection has been established, effectiveness will be inferred based on the proportion of adolescent study participants with serum Ab levels (on Day 57) that meet or exceed the Ab threshold. If an Ab threshold of protection has not been established, effectiveness will be inferred by demonstrating noninferiority of both the (i) geometric mean (GM) value of serum neutralizing antibody (nAb) and (ii) the seroresponse rate from adolescent participants compared with those from young adults (18-25 years of age) enrolled in the ongoing clinical endpoint efficacy trial (Study P301).

<u>Part A, the Blinded Phase</u>

This study in adolescents will monitor all participants for a total of 12 months following the second dose of vaccine or placebo. Safety assessments will include solicited adverse reactions (ARs; 7 days after each injection), unsolicited adverse events (AEs); 28 days after each injection), medically attended adverse events (MAAEs), serious adverse events (SAEs), and adverse events of special interest (AESIs) (including multisystem inflammatory syndrome in children [MIS-C]) throughout the study period.

Blood samples will be collected from all participants at baseline (Day 1), Day 57 (28 days after Dose 2), Day 209 (6 months after Dose 2), and Day 394 for measurement of SARS-CoV-2 specific binding and nAb responses. Blood samples will also be tested for the development of Ab directed against nonvaccine antigen (eg, Ab against the nucleocapsid protein), which will signify infection with SARS-CoV-2.

The incidence of SARS-CoV-2 infection among vaccine recipients and placebo recipients will be compared to assess the potential for mRNA-1273 to reduce the rate of infection in vaccine recipients.

Part A comprises 8 scheduled visits including a screening visit and 7 scheduled visits, of which Visit 2 and Visit 4 will be virtual/telephone visits and the other visits will be in-clinic visits. This study will be conducted in compliance with the protocol, Good Clinical Practice (GCP), and all applicable regulatory requirements.

Part B, the Open-label Observational Phase

Part B, the Open-label Observational Phase of the study, will be prompted by the authorization of a COVID-19 vaccine under an EUA for any persons under the age of 18 years. Participants will be transitioned to Part B of the study as their age group becomes EUA-eligible. This transition permits all ongoing study participants to eventually be informed of the availability and eligibility criteria of any COVID-19 vaccine made available under an EUA and the option to offer all ongoing study participants an opportunity to schedule a Participant Decision Visit to know their original treatment assignment (placebo vs. mRNA-1273 100 µg vaccine).

Part B provides the opportunity for study participants to be informed regarding the EUA, be unblinded to their original assignment (mRNA-1273 or placebo), and for those who previously received placebo to actively request to receive 2 doses of mRNA-1273 (100 µg) vaccine. EUA-eligible study participants will receive a Notification Letter summarizing the basis for EUA of a COVID-19 vaccine to receive an EUA and will be asked to schedule a Participant Decision Clinic Visit.

At the Participant Decision Clinic Visit, EUA-eligible participants will:

- Be given the option to be unblinded as to their original group assignment (placebo vs. mRNA-1273 vaccine [100 µg]),
- Be counseled about the importance of continuing other public health measures to limit the spread of disease including social distancing, wearing a mask, and hand-washing,

- Sign a revised informed consent form and assent, and
- Provide a nasopharyngeal (NP) or nasal swab for reverse transcriptase polymerase chain reaction (RT-PCR) for SARS-CoV-2 and a blood sample for serology and immunogenicity.

Participants that decline unblinding will remain in Part A and follow the Part A Schedule of Assessments (SoA).

After the Participant Decision Clinic Visit, participants that consent to unblinding will follow the Part A SoA or Part B SoA as follows:

- Participants received placebo in Part A and consent to receiving 2 doses of mRNA-1273 in Part B: These participants will proceed to Part B and follow the Part B SoA until BD is offered.
- Participants received 2 doses of mRNA-1273 in Part A: Due to statistical considerations, these participants will be considered in the Open-label Observational Phase but will continue to follow the Part A SoA until BD is offered.

Part C, the Booster Dose (BD) Phase

Part C is designed to offer participants in Part A and Part B who are at least 6 months from the last dose, the option to request a BD (50 µg) of mRNA-1273.

If eligible, each study participant will receive a notification letter and will be asked to schedule a BD-1 visit at their study site. Principal Investigators should consider current local public health guidance for administration of COVID-19 vaccines under EUA and marketing authorization (if any) when determining the scheduling priority of participants.

At the BD-1 visit, each participant will:

- Be encouraged to remain in the ongoing study,
- Sign a revised informed consent form (ICF) that includes both updated safety information relevant to the ongoing study and a BD, and the option to receive a BD,
- Be given the option to receive a BD consisting of a 50 µg dose of mRNA-1273,
- Be counselled about the importance of continuing other public health measures to limit the spread of disease including social distancing, wearing a mask, and hand-washing.

At the BD-1 visit, participants who request a BD and are eligible will have the following study site visits and complete scheduled activities (subject to investigational vaccine availability) according to the Part C Supplemental SoA:

- BD-1 visit: Participants will receive a single 50 µg dose of mRNA-1273

- BD-1a visit: All participants who chose to receive BD, Day 4, 3 days after BD on Day 1
- BD-2 visit: Day 29, 28 days after the BD on Day 1
- BD-3 visit: Day 181, 180 days after the BD on Day 1
- BD-4 visit: Day 361, 360 days after the BD on Day 1

Participants that crossed over from placebo to mRNA-1273 and decline BD will continue with Part B SoA.

Participants that received mRNA-1273 in Part A and decline BD will continue with in Part A SoA.

The investigator is responsible for conducting all assessments as specified in the Part C Supplemental SoA, according to the schedule. As this Supplemental SoA is intended to occur in addition to the original SoAs being followed by all participants in Part A or Part B, there is a possibility for study visits to overlap. If visits overlap according to respective visit windows, a single visit may be done with the combined study procedures completed once.

**Safety Oversight:**

Safety oversight will be under the direction of a Data Safety Monitoring Board (DSMB) composed of external independent consultants with relevant expertise.

An independent cardiac event adjudication committee (CEAC) that includes pediatric and adult cardiologists will review suspected cases of myocarditis and pericarditis to determine if they meet Center for Disease Control and Prevention criteria of "probable" or "confirmed" events, and assess severity (Gargano et al 2021). Any cases that the CEAC assesses as representing probable or confirmed cases of myocarditis or pericarditis will be referred to the Sponsor, who will then make a final decision on whether to suspend further enrollment and/or study dosing based on an assessment of the overall potential risk to study participants.

The CEAC will operate under the rules of an approved charter that will be written and reviewed at the organizational meeting of the CEAC. Details regarding the CEAC composition, responsibilities, procedures, and frequency of data review will be defined in its charter.

The contract research organization's medical monitor, the Sponsor's medical monitor, and the individual study site investigators will monitor safety throughout the study.

**Study Duration:**

Participants who received mRNA-1273 in Part A will be in the study approximately 25 months total , which includes 1 month for screening (Day -28 to Day 1), up to 12 months for dosing (on Day 1, Day 29, and Day 209 for BD), and 12 months for follow-up.

Participants who received placebo in Part A will be in the study for approximately 25 months total, which includes approximately 9 months in Part A, approximately 4 months of follow-up following their second dose of mRNA-1273 in Part B before entering Part C or before BD is received; or approximately 12 months of follow up.

Participants that decline to unblinding or decline to receive a BD will be in the study approximately 14 months total, which includes 1 month for screening (Day -28 to Day 1), 1 month for dosing (on Day 1 and Day 29), and 12 months for follow-up.

**Number of Participants:** Approximately 3,000 participants will be enrolled.

**Study Eligibility Criteria (Part A):**

**Inclusion Criteria:**

Each participant must meet all of the following criteria at the Screening Visit (Day 0) or at Day 1, unless noted otherwise, to be enrolled in this study:

1. Male or female, 12 to < 18 years of age at the time of consent (Screening Visit, Day 0) who, in the opinion of the investigator, is in good general health based on review of medical history and screening physical examination.
2. Investigator assessment that the participant, in the case of an emancipated minor, or parent(s)/legally acceptable representative(s) understand and are willing and physically able to comply with protocol-mandated follow-up, including all procedures and provides written informed consent/assent.
3. Body mass index (BMI) at or above the third percentile according to World Health Organization (WHO) Child Growth Standards at the Screening Visit (Day 0).
4. Female participants of nonchildbearing potential may be enrolled in the study. Nonchildbearing potential is defined as premenarche or surgically sterile (history of bilateral tubal ligation, bilateral oophorectomy, hysterectomy).
5. Female participants of childbearing potential may be enrolled in the study if the participant fulfills all the following criteria:

- Has a negative pregnancy test at Screening (Day 0), on the day of the first injection (Day 1), and on the day of the second injection (Day 29)
- Has practiced adequate contraception or has abstained from all activities that could result in pregnancy for at least 28 days prior to the first injection (Day 1)
- Has agreed to continue adequate contraception or abstinence through 3 months following the second injection (Day 29)

**Exclusion Criteria:**

Participants who meet any of the following criteria at the Screening Visit (Day 0) or at Day 1, unless noted otherwise, will be excluded from the study:

1. Travel outside of the United States in the 28 days prior to the Screening Visit (Day 0).
2. Pregnant or breastfeeding.
3. Is acutely ill or febrile 24 hours prior to or at the Screening Visit (Day 0). Fever is defined as a body temperature ≥ 38.0°C/≥ 100.4°F. Participants who meet this criterion may have visits rescheduled within the relevant study visit windows. Afebrile participants with minor illnesses can be enrolled at the discretion of the investigator.
4. Prior administration of an investigational CoV (eg, SARS-CoV-2, Severe Acute Respiratory Syndrome coronavirus [SARS-CoV], Middle East Respiratory Syndrome coronavirus [MERS-CoV]) vaccine.
5. Current treatment with investigational agents for prophylaxis against COVID-19.
6. Has a medical, psychiatric, or occupational condition that may pose additional risk as a result of participation, or that could interfere with safety assessments or interpretation of results according to the investigator’s judgment.
7. Current use of any inhaled substance (eg, tobacco or cannabis smoke, nicotine vapors).
8. History of chronic smoking (≥ 1 cigarette a day) within 1 year of the Screening Visit (Day 0).
9. History of illegal substance use or alcohol abuse within the past 2 years. This exclusion does not apply to historical cannabis use that was formerly illegal in the participant’s state but is legal at the time of screening.
10. History of a diagnosis or condition that, in the judgment of the investigator, may affect study endpoint assessment or compromise participant safety, specifically:

- Congenital or acquired immunodeficiency, including human immunodeficiency virus (HIV) infection
- Suspected active hepatitis
- Has a bleeding disorder that is considered a contraindication to intramuscular (IM) injection or phlebotomy
- Dermatologic conditions that could affect local solicited adverse reaction (AR)2 n
- assessments
- History of anaphylaxis, urticaria, or other significant AR requiring medical intervention after receipt of a vaccine
- Diagnosis of malignancy within the previous 10 years (excluding nonmelanoma skin cancer)
- Febrile seizures

11. Receipt of:
    - Any licensed vaccine within 28 days before the first dose of investigational product (IP) or plans for receipt of any licensed vaccine through 28 days following the last dose of IP.
    - Systemic immunosuppressants or immune-modifying drugs for > 14 days in total within 6 months prior to the day of enrollment (for corticosteroids, ≥ 20 mg/day prednisone equivalent). Topical tacrolimus is allowed if not used within 14 days prior to the day of enrollment. Participants may have visits rescheduled for enrollment if they no longer meet this criterion within the Screening Visit window. Inhaled, nasal, and topical steroids are allowed.
    - Intravenous blood products (red cells, platelets, immunoglobulins) within 3 months prior to enrollment.
12. Has donated ≥ 450 mL of blood products within 28 days prior to the Screening Visit (Day 0) or plans to donate blood products during the study.
13. Participated in an interventional clinical study within 28 days prior to the Screening Visit (Day 0) or plans to do so while participating in this study.
14. Is an immediate family member or has a household contact who is an employee of the research center or otherwise involved with the conduct of the study.

**Study Eligibility Criteria (Part B):**

1. Participants must have been previously enrolled in the mRNA-1273-P203 study.

2. Female participants of childbearing potential may be enrolled in the study if the participant has a negative pregnancy test on the day of the first injection (OL-Day 1) and on the day of the second injection (OL-Day 29).

**Study Eligibility Criteria (Part C):**

**Inclusion Criteria:**

1. Participants must have been previously enrolled in the mRNA-1273-P203 study, are actively participating in Part A or Part B, and are at least 6 months from the last dose.
2. Female participants of childbearing potential may be enrolled in the study if the participant has a negative pregnancy test on the day of the first injection (BD-Day 1).

**Exclusion Criteria:**

1. Pregnant or breastfeeding.
2. Is acutely ill or febrile 24 hours prior to or at the Screening Visit (Day 0). Fever is defined as a body temperature ≥ 38.0°C/≥ 100.4°F. Participants who meet this criterion may have visits rescheduled within the relevant study visit windows. Afebrile participants with minor illnesses can be enrolled at the discretion of the investigator.
3. Has a medical, psychiatric, or occupational condition that may pose additional risk as a result of participation, or that could interfere with safety assessments or interpretation of results according to the investigator's judgment.
4. History of a diagnosis or condition (after enrolment in Part A) that, in the judgment of the investigator, may affect study endpoint assessment or compromise participant safety:
    - Suspected active hepatitis
    - Has a bleeding disorder that is considered a contraindication to IM injection or phlebotomy
    - Dermatologic conditions that could affect local solicited AR assessments
    - History of anaphylaxis, urticaria, or other significant AR requiring medical intervention after receipt of a vaccine
    - Diagnosis of malignancy (excluding nonmelanoma skin cancer)
5. Receipt of:
    - Any authorized or licensed vaccine within 28 days before the first dose of IP or plans for receipt of any licensed vaccine through 28 days following the last dose of IP or any seasonal influenza vaccine within 14 days before the first dose of IP or

plans for receipt of any seasonal influenza vaccine 14 days following the last dose of IP.

6. Participated in an interventional clinical study, other than mRNA-1273-P203 study, within 28 days prior to the Screening Visit (Day 0) or plans to do so while participating in this study.

**Study Treatment:**

**Investigational Product:**

The IP (mRNA-1273 vaccine) is a lipid nanoparticle (LNP) dispersion of a messenger RNA (mRNA) encoding the prefusion stabilized spike (S) protein of SARS-CoV-2 formulated in LNPs composed of 4 lipids (1 proprietary and 3 commercially available): the proprietary ionizable lipid SM-102; cholesterol; 1,2-distearoyl-sn-glycero-3-phosphocholine (DSPC); and 1-monomethoxypolyethyleneglycol-2,3-dimyristylglycerol with polyethylene glycol of average molecular weight 2000 (PEG2000-DMG). mRNA-1273 injection is provided as a sterile liquid for injection, white to off-white dispersion in appearance, at a concentration of 0.5 mg/mL in 20 mM Tris buffer containing 87 mg/mL sucrose and 10.7 mM sodium acetate at pH 7.5 For Parts A and B, each injection will have a volume of 0.5 mL and contain mRNA-1273 100 µg. For Part C, each injection will have a volume of 0.25 mL and contain mRNA-1273 50 µg.

**Mode of Administration:**

Doses will be administered by IM injection into the deltoid muscle according to the procedures specified in the mRNA-1273-P203 Pharmacy Manual. Preferably, all doses should be administered into the nondominant arm.

**Procedures and Assessments:**

**Safety Assessments:**

Safety assessments will include monitoring and recording of the following for each participant:

- Solicited local and systemic ARs that occur during the 7 days following each injection (ie, the day of injection and 6 subsequent days) in **Part A and Part C**. Solicited ARs will be recorded daily using electronic diaries (eDiaries).
- Unsolicited AEs observed or reported during the 28 days following each injection (ie, the day of injection and 27 subsequent days) in Parts A and C
- AEs leading to discontinuation from dosing and/or withdrawal from study participation from Day 1 through the last day of study participation (ie, Day 394 per Part A SoA, OL-Day 178 in Part B, and BD-Day 361 in Part C)

- MAAEs from first dose on Day 1 through the entire study period (ie, Day 394 per Part A SoA, OL-Day 178 in Part B, and BD-Day 361 in Part C)
- SAEs from first dose on Day 1 through the entire study period (ie, Day 394 per Part A SoA, OL-Day 178 in Part B, and BD-Day 361 in Part C)
- AESIs through the entire study period (ie, Day 394 per Part A SoA, OL-Day 178 in Part B, and BD-Day 361 in Part C)
- Vital sign measurements
- Physical examination findings
- Assessments for SARS-CoV-2 infection from Day 1 through study completion
- Details of all pregnancies in female participants will be collected after the start of study treatment and until the end of their participation in the study

**Immunogenicity Assessments:**

The following analytes will be measured in blood samples for immunogenicity assessments:

- Serum nAb level against SARS-CoV-2 as measured by pseudovirus and/or live virus neutralization assays
- Serum binding antibody (bAb) levels against SARS-CoV-2 as measured by ligand-binding assay specific to the SARS-CoV-2 S protein
- Serum bAb levels against SARS-CoV-2 nucleocapsid protein as measured by ligand-binding assay specific to the SARSCoV2 nucleocapsid protein

Serum collected from all participants will be tested for bAb against SARS-CoV-2 nucleocapsid protein at specified time points. In addition, serum samples from a selected subset of study participants who received mRNA-1273 will be selected for testing of nAb and bAb against the SARS-CoV-2 S protein.

**Exploratory Assessments and Biomarkers:**

Exploratory assessments may include assessment of biomarkers for safety, reactogenicity, inflammatory and cardiac function. Serologic markers of disease severity, immune response to SARS-CoV-2, RT-PCR of NP or nasal swab or saliva samples, and genetic sequences of SARS-CoV-2 strains isolated from participants' samples may also be measured.

**Efficacy Assessments:**

Vaccine effectiveness for adolescents of ages of 12 to < 18 years will be inferred based on serum Ab responses obtained on Day 57 (28 days after the second injection of mRNA-1273). Inference will be based on assessing the adolescent Ab responses against the following:

1. *If available at the time of analysis*, adolescent Ab responses will be assessed against an accepted serum Ab threshold conferring protection against COVID-19.
2. *If an accepted threshold of protection is not available*, adolescent Ab responses will be assessed by establishing noninferiority of the GM value and seroresponse rate of serum nAb from adolescent participants compared with those from young adults enrolled in the ongoing clinical endpoint efficacy trial (Study P301).

**Statistical Methods:**

**Hypothesis Testing (Part A):**

If an accepted serum Ab threshold of protection against COVID-19 is established for the primary immunogenicity objective, the null hypothesis is that the percentage of participants on mRNA-1273 with serum Ab equal to or above the established threshold at Day 57 is ≤ 70% (ie, $H_0$: percentage of participants on mRNA-1273 ≤ 70% with serum Ab at Day 57 equal to or above the established threshold).

The study would be considered as meeting the immunogenicity objective if the 95% confidence interval (CI) of percentage of participants on mRNA-1273 rules out 70% (lower bound of the 95% CI > 70%).

If an accepted serum Ab threshold of protection against COVID-19 is not available for the primary immunogenicity objective, the immunogenicity analysis of primary vaccine response will be performed using the noninferiority tests of the two null hypotheses based on the two coprimary endpoints, respectively.

<u>Coprimary endpoint 1: Ab GM at Day 57</u>

$H^1_0$: immunogenicity response to mRNA-1273 as measured by Ab GM at Day 57 is inferior in adolescents (12 to < 18 years of age) compared with that in young adults (18-25 years of age) using mRNA-1273 Study P301 data.

The noninferiority in Ab GM in adolescents compared with that in young adults (18-25 years of age) is demonstrated by that the lower bound of the 95% CI of the GM ratio (GMR) rules out 0.67 (lower bound > 0.67) using a noninferiority margin of 1.5. The GMR is the ratio of the GM value of adolescents on mRNA-1273 in this study, Study P203, at Day 57 compared with the GM value of young adults (18-25 years of age) on mRNA-1273 in Study P301.

Coprimary endpoint 2: Ab seroresponse rate at Day 57

A definition of seroresponse will be provided in the statistical analysis plan (SAP) based on forthcoming information about assay performance.

The null hypothesis:

$H^2_0$: immunogenicity response to mRNA-1273 as measured by seroresponse rate at Day 57 is inferior in adolescents (12 to < 18 years of age) compared with that in young adults (18-25 years of age) using mRNA-1273 Study P301 data.

The noninferiority in seroresponse rate in adolescents compared with that in young adults (18-25 years of age) is demonstrated by that the lower bound of the 95% CI of the seroresponse rate difference rules out -10% (ie, lower bound > -10%) using the noninferiority margin of 10%. The seroresponse rate difference is defined as the rate in adolescents receiving mRNA-1273 minus the rate in young adults (18-25 years of age) receiving mRNA-1273 from Study P301.

The study would be considered as meeting the primary immunogenicity objective if noninferiority is demonstrated based on both coprimary endpoints.

Details regarding the assay to be used to assess noninferiority will be provided in the SAP.

**Power and Sample Size:**

The sample size of this study is driven by safety. Approximately 3,000 participants will be randomly assigned in a 2:1 ratio to receive mRNA-1273 or placebo. With 2,000 participants exposed to mRNA-1273, the study has at least 90% probability to observe at least 1 participant with an AE at a true 0.25% AE rate.

Serum samples from all participants will be collected and banked, a subset of participants will be selected, and their samples will be processed for immunogenicity testing (the Immunogenicity Subset).

Approximately 362 participants who receive mRNA-1273 will be selected for the Immunogenicity Subset, with a target of 289 participants receiving mRNA-1273 in the Per-Protocol (PP) Immunogenicity Subset (adjusting for approximately 20% of participants who may be excluded from the PP Immunogenicity Subset, as they may not have immunogenicity results due to any reason). The sample size of the Immunogenicity Subset may be updated with data from other mRNA-1273 studies or external data especially regarding a threshold of protection. In such a situation, the final sample size of the Immunogenicity Subset will be documented in the SAP.

For the primary immunogenicity objective, with approximately 289 participants in the per protocol (PP) Immunogenicity Subset, the study will have > 90% power to rule out 70% with a 2-sided 95% CI for the percentage of mRNA-1273 participants exceeding the acceptable threshold if the true rate of participants exceeding the acceptable threshold is 80%.

If an acceptable Ab threshold of protection against COVID-19 is not available at the time of analysis, for the primary immunogenicity objective, noninferiority tests of two null hypotheses based on two coprimary endpoints, respectively, will be performed. The sample size calculation for each of the two noninferiority tests was performed, and the larger sample size was chosen for the study.

- With approximately 289 participants in the PP Immunogenicity Subset in Study P203 and 289 participants in the PP Immunogenicity Subset in young adults (18-25 years of age) from Study P301, there will be 90% power to demonstrate noninferiority of the immune response as measured by Ab GM in adolescents in Study P203 at a 2-sided alpha of 0.05, compared with that in young adults (18-25 years of age) from Study P301 receiving mRNA-1273, assuming an underlying GMR value of 1 and a noninferiority margin of 1.5. The standard deviation (SD) of the log-transformed levels is assumed to be 1.5.
- With approximately 289 participants in the PP Immunogenicity Subset in Study P203 and 289 participants in the PP Immunogenicity Subset in young adults (18-25 years of age) from Study P301, there will be at least 90% power to demonstrate noninferiority of the immune response as measured by seroresponse rate in adolescents in Study P203 at a 2-sided alpha of 0.05, compared with that in young adults of (18-25 years of age) from Study P301 receiving mRNA-1273, assuming true seroresponse rate of 85% in young adults (18-25 years of age) from Study P301, and a true seroresponse rate of 85% in adolescents in Study P203 (ie, true rate difference is 0 compared to young adults [18-25 years of age] from Study P301), and a noninferiority margin of 10%.

**Analysis Sets:**

The analysis sets are defined in the following table:

| Analysis Set | Description |
|---|---|
| Randomization Set | All participants who are randomized, regardless of the participants' treatment status in the study. |
| Full Analysis Set (FAS) | All randomized participants who received at least 1 injection of IP. |
| Immunogenicity Subset | A subset of participants in the FAS will be selected for immunogenicity testing. |

| | |
|---|---|
| Per-protocol (PP) Immunogenicity Subset | A subset of participants in the FAS will be selected for immunogenicity testing. The PP Immunogenicity Subset includes participants selected for the Immunogenicity Subset who received planned doses of study vaccination per schedule, complied with immunogenicity testing schedule, and have no major protocol deviations that impact key or critical data. Participants who are seropositive at baseline will be excluded from the PP Immunogenicity Subset. The PP Immunogenicity Subset will be used for analyses of immunogenicity unless specified otherwise. |
| PP Set for Efficacy | All participants in the FAS who received planned doses of study vaccination, had no immunologic or virologic evidence of prior COVID-19, and have no major protocol deviations that impact key or critical efficacy data. |
| Solicited Safety Set | The Solicited Safety Set consists of FAS participants who contributed any solicited AR data.<br>The Solicited Safety Set will be used for the analyses of solicited ARs. |
| Safety Set | All randomized participants who receive at least 1 dose of IP. The Safety Set will be used for all analyses of safety except for the solicited ARs. |
| Modified Intent-to-Treat (mITT) Set | All participants in the FAS who have no serologic or virologic evidence of prior SARS-CoV-2 infection before the first dose of IP (both negative RT-PCR test for SARS-CoV-2 and negative serology test based on bAb specific to SARS-CoV-2 nucleocapsid) at baseline |
| Modified Intent-to-Treat-1 (mITT1) Set | All participants in the mITT Set excluding those who received the wrong treatment (ie, at least 1 dose received in Part A is not as randomized). |

Abbreviations: AR = adverse reaction; bAb = binding antibody; COVID-19 = coronavirus disease 2019; FAS = full analysis set; IP = investigational product; mITT = modified intent-to-treat; PP = per protocol; RT-PCR = reverse transcription polymerase chain reaction.

**Safety Analyses:**

All safety analyses will be based on the Safety Set, except summaries of solicited ARs, which will be based on the Solicited Safety Set. All safety analyses will be provided by treatment group.

Safety and reactogenicity will be assessed by clinical review of all relevant parameters including solicited ARs (local and systemic events), unsolicited AEs, SAEs, MAAEs, AESI, AEs leading to withdrawal, vital sign measurements, and physical examination findings. The number and percentage of participants with any solicited local AR, with any solicited systemic AR, and with any solicited AR during the 7-day follow-up period after each injection (except Part B) will be summarized.

The number and percentage of participants with any solicited local AR, with any solicited systemic AR, and with any solicited AR during the 7-day follow-up period after each dose (except Part B) will be provided. A 2-sided 95% exact CI using the Clopper-Pearson method will also be provided for the percentage of participants with any solicited AR.

The number and percentage of participants with unsolicited AEs, SAEs, MAAEs, Grade 3 or higher ARs and AEs, and AEs leading to discontinuation from IP or withdrawal from the study will be summarized. Unsolicited AEs will be presented by the Medical Dictionary for Regulatory Activities (MedDRA) preferred term and system organ class.

The number of events of solicited ARs, unsolicited AEs/SAEs, and MAAEs will be reported in summary tables accordingly.

**Immunogenicity Analyses:**

The SAP will describe the complete set of immunogenicity analyses, including the approach to sample participants into an Immunogenicity Subset for analysis of immunogenicity. The PP Immunogenicity Subset is the primary analysis set for immunogenicity unless otherwise specified. The primary immunogenicity objective of this study is to use the immunogenicity response to infer efficacy in adolescents (12 to < 18 years in this study). The primary immunogenicity analysis is performed in Part A.

If an accepted serum Ab threshold of protection against COVID-19 is available based on data from other mRNA-1273 studies or external data, the number and percentage of participants with Ab greater than or equal to the threshold at Day 57 will be provided with a 2-sided 95% CI using the Clopper-Pearson method. If the lower bound of the 95% CI on the mRNA-1273 group is > 70%, the primary immunogenicity objective of this study will be considered to be met.

The percentage of participants with serum Ab greater than or equal to the threshold with 95% CI will be provided at each postbaseline time point. The CI will be calculated using the Clopper-Pearson method.

If an accepted serum Ab threshold of protection against COVID-19 is not established, the non-inferiority of primary vaccine response as measured by Ab GM and seroresponse rate in adolescents compared with those in young adults (18-25 years of age) receiving mRNA-1273 will be assessed. The study is considered as meeting the primary immunogenicity objective if the noninferiority of the immune response to mRNA-1273 as measured by both GM and seroresponse rate at Day 57 is demonstrated in adolescents in this study at a 2-sided alpha of

0.05, compared with that in young adults (18-25 years of age) in Study P301 receiving mRNA-1273.

An analysis of covariance model will be carried out with Ab value at Day 57 as a dependent variable and a group variable (adolescents in Study P203 and young adults [18-25 years of age] in Study P301) as the fixed variable. The GM values of the adolescents at Day 57 will be estimated by the geometric least squares mean (GLSM) from the model. The GMR (ratio of GM values) will be estimated by the ratio of GLSM from the model. A corresponding 2-sided 95% CI will be provided to assess the difference in immune response for the adolescents in Study P203 compared to the young adults (18-25 years of age) in Study P301 at Day 57. The noninferiority of immune response to mRNA-1273 as measured by GM will be considered demonstrated if the lower bound of the 95% CI of the GMR is > 0.67 based on the noninferiority margin of 1.5.

The number and percentage (rate) of participants achieving Ab seroresponse at Day 57 will be summarized. The difference of seroresponse rates between adolescents receiving mRNA-1273 in Study P203 and young adults (18-25 years of age) receiving mRNA-1273 in Study P301 will be calculated with 95% CI. The noninferiority in seroresponse rate of adolescents in Study P203 compared to young adults (18-25 years of age) in Study P301 will be considered demonstrated if the lower bound of the 95% of the seroresponse rate difference is > -10%, based on the noninferiority margin of 10%.

In addition, the GM level of specific nAb and bAb with corresponding 95% CI will be provided at each time point. The 95% CIs will be calculated based on the t-distribution of the log transformed values then back transformed to the original scale. The GM fold-rise of nAb and bAb with corresponding 95% CI will be provided at each time point with Day 57 as the primary time point of interest. Descriptive summary statistics including median, minimum, and maximum will also be provided.

**Efficacy Analyses:**

To evaluate the incidence of COVID-19 after vaccination with mRNA-1273 or placebo, the incidence rate will be provided by vaccination group, calculated as the number of cases divided by the total person-time. The incidence rate ratio of mRNA-1273 versus placebo will be provided with 95% CI computed using the exact method conditional upon the total number of cases adjusted by the total person-time.

For SARS-CoV-2 infection (serologically confirmed SARS-CoV-2 infection or COVID-19), regardless of symptomatology or severity, infection rate will be provided by vaccination group. The infection rate ratio of mRNA-1273 versus placebo may be provided with its 95% CI using the exact method conditional upon the total number of cases adjusted by the total person-time. The incidence rate of asymptomatic SARS-CoV-2 infection will also be provided.

The secondary efficacy analyses will be performed in the PP set, with sensitivity analyses in the FAS, mITT Set, and mITT1 Set.

**Long-Term Analysis (including Part B):**

Long-term analysis will be performed including data collected in the Open-label Observational Phase (Part B), and prior to BD if a BD is received. The analysis will include participants who are randomized to mRNA-1273 in Part A and remained in the study with or without unblinding, and participants who are originally randomized to placebo and later crossed over to receive mRNA-1273 in Part B after unblinding. Long-term analysis of applicable safety, efficacy, and immunogenicity endpoints will be summarized descriptively by treatment cohort without treatment group comparison.

In the long-term safety analysis, unsolicited AEs will be summarized.

In the long-term immunogenicity analysis, nAb and bAb values will be summarized at specified timepoints.

In the long-term efficacy analysis, the incidence rates of COVID-19 and of SARS-CoV-2 infection cases will be counted starting 14 days after the second dose of IP for participants in treatment cohorts of mRNA-1273 and Placebo or starting 14 days after the second dose of mRNA-1273 for participants in the Placebo-mRNA-1273 cohort. Incidence rate with 95% CI adjusting for person-time will be provided. The incidence rate of asymptomatic SARS-CoV-2 infection will also be provided.

**Hypothesis Testing (Part C):**

The immunogenicity analysis of BD vaccine response against the original strain will be performed using the noninferiority tests of the two null hypotheses based on the two coprimary endpoints, respectively.

**Coprimary Endpoint 1: Ab GM at BD-Day 29**

Null hypothesis

$H^1_0$: immunogenicity response to mRNA-1273 BD as measured by Ab GM at BD-Day 29 in adolescents (12-<18 years of age) in Study P203 Part C is inferior compared with Ab GM at Day 57 (28 days after Dose 2) in the primary series of mRNA-1273 in young adults (18-25 years of age) in Study P301.

The noninferiority in Ab GM at BD-Day 29 in Study P203 Part C compared with Ab GM at Day 57 in the primary series in young adults (18-25 years of age) in P301 will be demonstrated by the GMR 95% CI lower bound >0.67 using a noninferiority margin of 1.5. The GMR is defined as the GM of Ab at BD-D29 in Study P203 Part C compared with Ab GM at Day 57 (28 days after Dose 2) following the primary series of mRNA-1273 in Study P301.

**Coprimary Endpoint 2: Ab Seroresponse Rate (SRR) at BD-Day 29**

Null hypothesis

$H^2_0$: immunogenicity response to mRNA-1273 BD as measured by SRR at BD-Day 29 in adolescents (12-<18 years of age) in Study P203 Part C is inferior compared with SRR at Day 57 (28 days after Dose 2) following the primary series of mRNA-1273 in young adults (18-25 years of age) in Study P301.

The noninferiority in SRR at BD-D29 compared with SRR at Day 57 (28 days after Dose 2) following the primary series of mRNA-1273 will be demonstrated by the SRR 95% CI lower bound > -10% using the noninferiority margin of 10%. The SRR difference is defined as the SRR at BD-Day 29 Study P203 Part C minus the rate at Day 57 (28 days after Dose 2) following the primary series of mRNA-1273 in young adults in Study P301. The SRR is defined as a titer change from baseline (pre-Dose 1) below the LLOQ to ≥ 4 × LLOQ, or at least a 4-fold rise if baseline is ≥ LLOQ.

The primary immunogenicity objective in Part C is met if the noninferiority is demonstrated based on both coprimary endpoints.

**Key Secondary Endpoint 1: GM of Ab Against Circulating Strain/Delta at BD-Day 29**

Null hypothesis

$H^1_0$: immunogenicity response to mRNA-1273 BD as measured by GM of Ab against circulating strain/Delta at BD-Day 29 in adolescents (12-<18 years of age) in Study P203 Part C is inferior compared with GM of Ab against circulating strain/Delta at Day 57 (28 days after Dose 2) in the primary series of mRNA-1273 in young adults (18-25 years of age) in Study P301.

The noninferiority in Ab GM at BD-Day 29 in Study P203 Part C compared with Ab GM at Day 57 in the primary series in young adults (18-25 years of age) in Study P301 will be demonstrated by the GMR 95% CI lower bound >0.67 using a noninferiority margin of 1.5. The GMR is defined as the GM of Ab at BD-Day 29 in Study P203 Part C compared with Ab GM at Day 57 (28 days after Dose 2) following the primary series of mRNA-1273 in Study P301.

**Key Secondary Endpoint 2: Seroresponse Rate (SRR) of Ab Against Circulating Strain/Delta at BD-Day 29**

Null hypothesis

$H^2_0$: immunogenicity response to mRNA-1273 BD as measured by SRR of Ab against circulating strain/Delta at BD-Day 29 in adolescents (12-<18 years of age) in Study P203 Part C is inferior compared with SRR of Ab against circulating strain/Delta at Day 57 (28 days after Dose 2) following the primary series of mRNA-1273 in young adults (18-25 years of age) in Study P301.

The noninferiority in SRR at BD-D29 compared with SRR at Day 57 (28 days after Dose 2) following the primary series of mRNA-1273 will be demonstrated by the SRR 95% CI lower bound > -10% using the noninferiority margin of 10%. The SRR difference is defined as the SRR at BD-Day 29 Study P203 Part C minus the rate at Day 57 (28 days after Dose 2) following the primary series of mRNA-1273 in young adults in Study P301. The SRR is defined as a titer change from baseline (pre-Dose 1) below the LLOQ to ≥ 4 × LLOQ, or at least a 4-fold rise if baseline is ≥ LLOQ.

The key secondary immunogenicity objective in Part C is met if the noninferiority is demonstrated based on both key secondary endpoints.

**Multiplicity Adjustment**

A sequential hypothesis testing (fixed-sequence) method will be used to adjust multiplicity to preserve the family-wise Type I error rate (alpha = 0.05). The hypothesis testing for the two coprimary endpoints (geometric mean titer [GMT] and SRR) for the primary series of mRNA-1273 in Part A was completed and statistically significant based on data snapshot dated 08 May 2021, and thus the alpha level of 0.05 can be passed to Part C hypothesis testing. In Part C, the hypothesis testing for the two coprimary endpoints (GMT and SRR against the original strain) after BD of mRNA-1273 will be tested first at alpha level of 0.05. If the testing of both the coprimary endpoints in Part C is statistically significant (meeting the noninferiority success criteria of the coprimary endpoints), the alpha level of 0.05 will be passed to the hypothesis testing of the key secondary endpoints (GMT and SRR against circulating strain/Delta). The testing will continue through the sequence only until an endpoint is not statistically significant (did not meet specified noninferiority success criteria), in which case the testing will stop.

**Power and Sample Size (Part C):**

All participants enrolled in Part A or Part B who meet the eligibility criteria for BD will be offered a BD of mRNA-1273 50 µg. With more than 1,000 participants expected to receive mRNA-1273 BD, the study Part C has a 90% probability to observe at least 1 participant with an AE at a true AE rate of 0.25%.

Serum samples from all participants will be collected and banked, a subset of participants will be selected, and their samples will be processed for immunogenicity testing (the Immunogenicity Subset) at specified timepoints. .

Approximately 362 participants who receive mRNA-1273 BD will be selected for the Immunogenicity Subset for Part C, with a target of 289 participants receiving mRNA-1273 BD in the PP Immunogenicity Subset for Part C (adjusting for approximately 20% of participants who may be excluded from the PP Immunogenicity Subset, as they may not have immunogenicity results due to any reason or may have protocol deviations impacting critical data).

For the primary immunogenicity objective in Part C, noninferiority tests of two null hypotheses based on two coprimary endpoints, respectively, will be performed.

- With approximately 289 participants receiving mRNA-1273 BD in the PP Immunogenicity Subset in Study P203 Part C and 289 participants in the PP Immunogenicity Subset in young adults (18-25 years of age) in Study P301, there will be 90% power to demonstrate noninferiority of the immune response post BD as measured by Ab GM in adolescents in Study P203 Part C compared with Ab GM in young adults (18-25 years of age) following primary series of mRNA-1273 in Study P301, at a 2-sided alpha of 0.05, assuming an underlying GMR value of 1 and a noninferiority margin of 1.5. The SD of the log-transformed levels is assumed to be 1.5.
- With approximately 289 participants receiving mRNA-1273 BD in the PP Immunogenicity Subset in Study P203 Part C and 289 participants in the PP Immunogenicity Subset in young adults (18-25 years of age) in Study P301, there will be at least 90% power to demonstrate noninferiority of the immune response post BD as measured by SRR in adolescents in Study P203 Part C compared with SRR in young adults of (18-25 years of age) following primary series of mRNA 1273 in Study P301, at a 2-sided alpha of 0.05, assuming true SRR of 90% in young adults (18-25 years of age) following primary series of mRNA-1273 in Study P301, and a true SRR of 90% post BD in adolescents in Study P203 Part C (ie, true rate difference is 0 compared to young adults [18-25 years of age] from Study P301), and a noninferiority margin of 10%.

**Booster Phase Analysis (Part C)**

Booster phase analysis will be performed in participants who receive a BD in Part C. Safety, immunogenicity endpoints, and incidence of COVID-19 and SARS-CoV-2 infection will be summarized descriptively, and by pre-booster SARS-CoV-2 status if applicable.

In the booster phase safety analysis, the number and percentage of participants with unsolicited AEs, severe AEs, treatment-related AEs, SAEs, MAAEs, AESIs, and AEs leading to discontinuation from the study participation in Part C will be summarized.

The number and percentage of participants with any solicited local AR, with any solicited systemic AR, and with any solicited AR during the 7-day follow-up period after the BD will be provided.

In the Part C booster phase, for the coprimary endpoints (Ab GM and SRR against the original strain) and key secondary endpoints (Ab GM and SRR against the circulating strain/Delta), the following analyses will be performed for the respective hypothesis testing:

- The GM titers with 95% CI will be summarized using t-distribution of the log transferred values and then back transformed to the original scale. The GMR with 95% CI to compare post-booster GM at BD-Day 29 in adolescents in Study P203 with the primary series GM at Day 57 (28 days after Dose 2) in young adults (18-25 years of age) in Study P301 will be computed based on the t-distribution of mean difference in the log transferred values and then back transformed to the original scale.
- The SRR with 95% CI (using Clopper-Pearson method) will be summarized. The SRR difference with 95% CI (using Miettinen-Nurminen score method) to compare post-booster SRR at BD-Day 29 in adolescents in Study P203 with the primary series SRR at Day 57 (28 days after dose 2) in young adults in Study P301 will be computed. The SRR is defined as a titer change from baseline (pre-Dose 1) below the LLOQ to $\geq 4 \times$ LLOQ, or at least a 4-fold rise if baseline is $\geq$ LLOQ.

In the booster phase immunogenicity analyses, the GM titers or levels for specific nAb and bAb, with corresponding 95% CI will be provided at BD-Day 1, BD-Day 29, BD-Day181, and BD-Day 361(Part C). Geometic mean fold-rise relative to pre-Dose 1, BD-Day 1 and prior Day 57 in the phase of primary 2-dose series of mRNA-1273 will be summarized with 95% CIs at BD-Day 29, and BD-Day 181, and BD-Day 361 (Part C). The 95% CIs will be calculated based on the t-distribution of the log transformed values then back transformed to the original scale. Descriptive summary statistics including median, minimum, and maximum will also be provided. The immunogenicity descriptive summaries will be provided in the participants who have available nAb and bAb assay results for the specified time points.

In the booster phase, the incidence rate of the first occurrence of symptomatic COVID-19 starting 14 days after the BD will be summarized by pre-booster SARS-CoV-2 status. The incidence rate of SARS-CoV-2 infection (symptomatic or asymptomatic infection) starting 14 days after the BD, and the incidence rate of asymptomatic SARS-CoV-2 infection starting

14 days after the BD will be summarized. The incidence rates with 95% CI will be computed adjusting for person-years starting from BD-Day 1 in Part C.

**Study Analyses:**

**Interim Analyses:**

More than one interim analysis (IA) may be performed.

- The IA of immunogenicity, safety, and efficacy will be performed after Day 57 immunogenicity data are available for the immunogenicity subset and at least 1,500 participants (1,000 participants receiving mRNA-1273) have completed Day 57 (1 month after Dose 2, Part A). This IA will be considered the primary analysis of immunogenicity.
- An IA of immunogenicity and safety may be performed after all or subset of participants who receive booster dose have completed BD-Day 29 after the booster dose.
- At the Sponsor's discretion, a clinical study report (CSR) may be developed for an IA.

**Final Analysis:**

The final analysis of all applicable endpoints will be performed after all participants have completed all planned study procedures. Results of this analysis will be presented in a final CSR, including individual listings.

### LIST OF ABBREVIATIONS AND TERMS

The following abbreviations and terms are used in this study protocol.

| Abbreviation or Specialist Term | Definition |
|---|---|
| Ab | antibody |
| AE | adverse event |
| AESI | adverse events of special interest |
| AR | adverse reaction |
| bAb | binding antibody |
| BD | booster dose |
| BMI | body mass index |
| CBER | Center for Biologics Evaluation and Research |
| CD | cluster of differentiation |
| CDC | US Centers for Disease Control and Prevention |
| CEAC | cardiac event adjudication committee |
| CFR | Code of Federal Regulations |
| CI | confidence interval |
| cMRI | cardiac magnetic resonance imaging |
| CoV | coronavirus |
| COVID-19 | coronavirus disease 2019 |
| CRO | contract research organization |
| CRP | C-reactive protein |
| CSR | clinical study report |
| DMID | Division of Microbiology and Infectious Diseases |
| DSMB | Data Safety Monitoring Board |
| DSPC | 1,2-distearoyl-sn-glycero-3-phosphocholine |
| ECG (or EKG) | electrocardiogram |
| ECMO | extracorporeal membrane oxygenation |
| eCRF | electronic case report form |
| EDC | electronic data capture |

| Abbreviation or Specialist Term | Definition |
|---|---|
| eDiary | electronic diary |
| EOS | end of study |
| ERD | enhanced respiratory disease |
| ESR | erythrocyte sedimentation rate |
| EUA | Emergency Use Authorization |
| FAS | full analysis set |
| FDA | US Food and Drug Administration |
| FIO2 | fraction of inspired oxygen |
| GCP | Good Clinical Practice |
| GLSM | geometric least squares mean |
| GM | geometric mean |
| GMFR | geometric mean fold-rise |
| GMR | geometric mean ratio |
| GMT | geometric mean titer |
| HCP | healthcare practitioner |
| HIV | human immunodeficiency virus |
| IA | interim analysis |
| IB | investigator's brochure |
| ICF | informed consent form |
| ICH | International Council for Harmonisation |
| ICU | intensive care unit |
| IL-6 | interleukin 6 |
| IM | intramuscular(ly) |
| IP | investigational product |
| IRB | institutional review board |
| LAR | legally acceptable representative |
| LDH | lactic acid dehydrogenase |
| LNP | lipid nanoparticle |

| Abbreviation or Specialist Term | Definition |
|---|---|
| LTFU | lost to follow-up |
| MAAE | medically attended adverse event |
| MedDRA | Medical Dictionary for Regulatory Activities |
| MERS | Middle East Respiratory Syndrome |
| MIS-C | multisystem inflammatory syndrome in children |
| MN | microneutralization |
| mRNA | messenger RNA |
| nAb | neutralizing antibody |
| NHP | nonhuman primate |
| NIAID | National Institute of Allergy and Infectious Diseases |
| NP | nasopharyngeal |
| PaO2 | partial pressure of oxygen |
| PEG2000-DMG | 1-monomethoxypolyethyleneglycol-2,3-dimyristylglycerol polyethylene glycol 2000 |
| PP | per protocol |
| QA | quality assurance |
| RT-PCR | reverse transcriptase polymerase chain reaction |
| S | spike |
| S2P | S protein |
| SAE | serious adverse event |
| SAP | statistical analysis plan |
| SARS | Severe Acute Respiratory Syndrome |
| SARS-CoV-2 | Severe Acute Respiratory Syndrome coronavirus 2 |
| SD | standard deviation |
| SM-102 | heptadecane-9-yl 8-((2-hydroxyethyl)(6-oxo-6-(undecyloxy)hexyl)amino)octanoate |
| SMC | Safety Monitoring Committee |
| SoA | schedule of assessments |

| Abbreviation or Specialist Term | Definition |
|---|---|
| SpO2 | oxygen saturation |
| SRR | seroresponse rate |
| Study P301 | Study mRNA-1273-P301; NCT04470427 |
| Th1 | T helper cell 1 |
| Th2 | T helper cell 2 |
| VE | vaccine efficacy |
| WHO | World Health Organization |
| WOCBP | woman of childbearing potential |

### 1. INTRODUCTION

#### 1.1. Study Rationale

Coronaviruses (CoVs) are a large family of viruses that cause illness ranging from the common cold to more severe diseases, such as Middle East Respiratory Syndrome (MERS) and Severe Acute Respiratory Syndrome (SARS). Coronaviruses are zoonotic, meaning they are transmitted between animals and people. An outbreak of the CoV disease 2019 (COVID-19) caused by Severe Acute Respiratory Syndrome coronavirus 2 (SARS-CoV-2) began in Wuhan, Hubei Province, China in December 2019 and has spread throughout China and to over 215 other countries, territories, and areas including the United States (WHO 2020a). On 11 Mar 2020, the World Health Organization (WHO) officially declared COVID-19 a pandemic. As of 28 Sep 2020, the WHO dashboard (WHO 2020b) reported there have been nearly 1 million COVID-19 deaths worldwide and more than 200,000 deaths in the United States.

As of 21 Sep 2020, the US Centers for Disease Control and Prevention (CDC) reported over 6.7 million confirmed and probable cases of COVID-19 in all 50 states and 5 jurisdictions, with over 199,000 attributed and probable deaths (CDC 2020a). While the CDC have reported that the highest risk of disease burden is in older adults and populations with certain underlying comorbid conditions such as heart disease, diabetes, and lung disease, the burden in the pediatric population is not negligible. Rather, evidence is emerging (described below) to suggest that children < 18 years of age, particularly adolescents, may be disproportionately contributing to the number of new cases as schools re-open for varying degrees of in-person learning. As of 21 Sep 2020, the CDC reported over 408,000 cases of COVID-19 in children less than 18 years of age (8.1% of all US cases) and 88 deaths (< 0.1% of all US deaths; CDC 2020b).

During the incubation period, those infected can also transmit the virus (Chen et al 2020). Person-to-person transmission occurs primarily via direct contact or through droplets spread by coughing or sneezing from an infected individual, whether symptomatic or not (Rothan and Byrareddy 2020; Chen et al 2020; Licciardi et al 2020; Shen et al 2020). SARS-CoV-2 can also be transmitted via the fecal-oral pathway (Cruz and Zeichner 2020).

During this COVID-19 pandemic, children throughout much of the world have had school attendance limited in an attempt to control infection. Therefore, the main source of infection for SARS-CoV-2 in children, with or without clinical symptoms, is infected household contacts. Indeed, a retrospective cohort study of high school students, parents and siblings of students, and school staff conducted in France in early April 2020 suggests that there was little to no transmission from infected students to other students or school staff. Rather, a high prevalence of antibodies (Abs) against SARS-CoV-2 among families suggests familial clustering of COVID-19 cases (Fontanet et al 2020).

A recent report of COVID-19 trends in school-aged children in the United States from 01 Mar 2020 to 19 Sep 2020 indicates that 37% of laboratory-confirmed cases of COVID-19 in school-aged children occurred in children 5 to 11 years of age while 63% occurred in adolescents 12 to 17 years of age (Leeb et al 2020). During this time period, the average weekly incidence among adolescents was 37.4 cases per 100,000 compared with 19.0 per 100,000 for younger children. Among school-aged children with laboratory-confirmed COVID-19, 58% reported at least one symptom and 5% reported no symptoms; although information on symptoms was missing or unknown for 37%. Overall, in this study, 1.2% of school-aged children with COVID-19 were hospitalized, 0.1% required intensive care unit (ICU) admission and < 0.01% died of COVID-19. Furthermore, at least one underlying condition was reported in 3% of adolescents and 2% of younger children. Chronic lung disease, including asthma, was most commonly reported (55%), followed by disability (neurologic or neurodevelopmental disorders, intellectual or physical disability, and vision or hearing impairment; 9%), immunosuppressive conditions (7%), diabetes (6%), psychological conditions (6%), cardiovascular disease (5%), and severe obesity (4%) (Leeb et al 2020).

Another study examined the age distribution of COVID-19 in the United States from May to August 2020 based on 3 indicators: COVID-19-like illness-related emergency department visits, positive reverse transcriptase polymerase chain reaction (RT-PCR) results for SARS-CoV-2, and confirmed COVID-19 cases (Boehmer et al 2020). These authors report an estimated mean COVID-19 incidence during this time period of 179.3 cases per 100,000 in individuals 10 to 19 years of age. Generally, the largest increase in incidence during this time period was observed in persons < 30 years of age. Finally, a recent report describes an adolescent (13-year-old female), whose only symptom was nasal congestion, yet who was the index case in an outbreak of COVID-19 across 4 states (Schwartz et al 2020). Infection of this primary individual led to 11 subsequent cases, during July and August 2020, in 5 households all linked to a family gathering suggesting that adolescents can serve as the source of COVID-19 outbreaks within families, even when their symptoms are mild as in this case.

Taken together, the above evidence suggests that the burden of COVID-19 has begun to increase in younger age groups, particularly as schools in the United States have started to reopen for some in-person instruction. Adolescents, who are often mobile and may demonstrate lower compliance with nonpharmaceutical interventions such as mask-wearing and social distancing, also likely represent a segment of the population contributing toward sustained community transmission of SARS-CoV-2 and may spread SARS-CoV-2 within households. A vaccine that prevents COVID-19 and SARS-CoV-2 transmission in adolescents would be a crucial public health tool to help curb the pandemic.

At the time the present study was initiated, no vaccine had been authorized or licensed to prevent SARS-CoV-2, and there was an urgent public health need to develop one, there being no proven therapy. ModernaTX, Inc (the Sponsor) initiated an accelerated development program for mRNA-1273 vaccine against SARS-CoV-2 infection (Section 1.2), and started a Phase 3 clinical study in the United States involving administration of mRNA-1273 vaccine 100 µg as both an initial dose and a second dose 28 days later (Section 1.2.2). Success criteria for early efficacy were met at first interim analysis (IA) based on 95 adjudicated cases with a vaccine efficacy (VE) of 94.5% (95% CI: 86.5%, 97.8%; one-sided p-value < 0.0001), and EUA was granted for mRNA-1273 in the adult population.

The primary objective for this Phase 2/3 study is to evaluate the safety and reactogenicity of a single dose level (100 µg) of mRNA-1273 vaccine administered in 2 doses 28 days apart (Section 3.1) to an adolescent population. Specifically, the primary immunogenicity objective of Study P203 is to infer effectiveness of mRNA-1273 (100 µg, 2 doses 28 days apart) 28 days after dose 2 of mRNA-1273 (Day 57) by comparison of immune responses in adolescents (aged ≥ 12 through < 18 years) to the young adult (≥ 18 to ≤ 25 year of age) cohort in Study P301, where efficacy was demonstrated.

Interim data from the current P203 study (based on a data snapshot of 08 May 2021) met the primary immunogenicity objective by demonstrating noninferiority of both the (i) geometric mean (GM) value of serum neutralizing Ab (nAb) and (ii) the seroconversion rate from adolescent participants compared with those from young adults (aged ≥18 to ≤ 25 years) enrolled in the ongoing Phase 3 efficacy study (Study P301). A request for an EUA in the adolescent population has been submitted to the US Food and Drug Administration (FDA) and global filings/approvals down to 12 years of age have occurred.

In addition, recent interim results of an ongoing Moderna Phase 2 study, mRNA-1273-P201 (Study P201) support administration of a booster dose (BD) of vaccine. Adult participants in Study mRNA-1273-P201 received 2 doses of either 50 µg or 100 µg of mRNA-1273 and were administered a 50µg booster of mRNA-1273 6 to 8 months after the second dose. Participants in mRNA-1273-P201 who received the BD demonstrated enhanced immune responses to SARS-CoV-2 compared to pre-boost levels and met the noninferiority criteria stipulated in the FDA Guidance on EUA for Vaccines to Prevent COVID-19. Additionally, no new safety signals emerged upon administration of the booster dose in Study P201. Based on cumulative evidence, the benefit-risk profile of a BD of mRNA-1273 is favorable, particularly in light of increasing breakthrough disease with the emergence of the Delta variant.

Thus, an additional objective of the current P203 study is to evaluate safety and immunogenicity of a third BD of 50 µg of mRNA-1273.

#### 1.2. Background and Overview

The Sponsor has developed a rapid-response, proprietary vaccine platform based on a messenger RNA (mRNA) delivery system. The platform is based on the principle and observations that cells in vivo can take up mRNA, translate it, and then display protein viral antigen(s) on the cell surface. The delivered mRNA does not enter the cellular nucleus or interact with the genome, is nonreplicating, and is expressed transiently. Messenger RNA vaccines have been used to induce immune responses against infectious pathogens such as SARS-CoV-2 (NCT04283461, NCT04405076), cytomegalovirus (NCT03382405), metapneumovirus and parainfluenza virus type 3 (NCT03392389), Zika virus (NCT04917861), and influenza virus (NCT03076385 and NCT03345043).

The Sponsor is using its mRNA-based platform to develop a novel lipid nanoparticle (LNP)-encapsulated mRNA-based vaccine against SARS-CoV-2 (mRNA-1273). mRNA-1273 encodes for the full-length spike (S) protein of SARS-CoV-2. The CoV S protein mediates attachment and entry of the virus into host cells (by fusion), making it a primary target for neutralizing antibodies (nAb) that prevent infection (Johnson et al 2016; Wang et al 2015; Wang et al 2018; Chen et al 2017; Corti et al 2015; Yu et al 2015; Kim et al 2019; Widjaja et al 2019; Corbett et al 2020a; Ju et al 2020; Robbiani et al 2020). It has been confirmed that the stabilized SARS-CoV-2 S protein (S2P) expresses well and is in the prefusion conformation (Wrapp et al 2020).

The development of the mRNA-1273 vaccine is being accelerated to address the current SARS-CoV-2 outbreak as a result of the uniquely rapid and scalable manufacturing process for mRNA-1273 vaccine.

##### 1.2.1. Nonclinical Studies

Nonclinical studies have demonstrated that CoV S proteins are immunogenic and S protein-based vaccines, including those based on mRNA delivery platforms, are protective in animals. Prior clinical studies of vaccines targeting related CoVs and other viruses have demonstrated that mRNA-based vaccines are safe and immunogenic. mRNA-1273 has shown preliminary evidence of protection against SARS-CoV-2 in studies in young mice (Corbett et al 2020a) and nonhuman primates (NHPs) (Corbett et al 2020b).

In support of the development of mRNA-1273 for prophylaxis against SARS-CoV-2 infection, nonclinical immunogenicity, biodistribution, and safety studies have been completed with similar mRNA-based vaccines formulated in LNPs containing SM-102 (heptadecane-9-yl 8-((2-hydroxyethyl)(6-oxo-6-(undecyloxy)hexyl)amino)octanoate), the novel proprietary lipid used in the mRNA-1273 LNP formulation.

A detailed review of nonclinical experience with mRNA-1273 vaccine is provided in the investigator's brochure (IB).

##### 1.2.2. Clinical Studies

The clinical development of mRNA-1273 vaccines to support its use in the adult population consists of 5 ongoing clinical trials being conducted in the US:

- Phase 1, open-label, dose ranging studies (Study 20-0003; NCT04283461; Study 21-0002, NCT04785144) sponsored by the National Institute of Allergy and Infectious Diseases
- Phase 2a, randomized, observer-blind, placebo-controlled, dose-confirmation study (Study mRNA-1273-P201; NCT04405076)
- Phase 3 randomized, stratified, observer-blind, placebo-controlled study (Study mRNA-1273-P301; NCT04470427) to evaluate the efficacy, safety, and immunogenicity of the vaccine
- Phase 3b, open-label study (Study mRNA-1273-P304, NCT04860297) to evaluate the safety and immunogenicity of the vaccine in adult solid organ transplant recipients and healthy controls.

In addition, 2 ongoing trials are being conducted in the pediatric population:

- Phase 2/3 randomized, placebo-controlled safety and immunogenicity study of mRNA-1273 in children 12 to less than 18 years of age (Study mRNA-1273-P203; NCT04649151)
- Phase 2/3, study to evaluate the safety and immunogenicity of mRNA-1273 in children less than 12 years of age (mRNA-1273-P204; NCT04796896).

The safety and immunogenicity of mRNA-1273 were evaluated in Phase 1 (DMID 20-0003) and Phase 2 (mRNA-1273-P201) studies that were important to select and to confirm the dose of the vaccine used in the pivotal Phase 3 study. In Study 20-0003 (Phase 1), 2 doses of 100 µg or higher generated the highest titers of nAb or binding antibody (bAb) with an acceptable safety profile, and this observation was the basis for selecting the 100-µg dose for use in the pivotal Phase 3 study. Importantly, the Ab levels after 2 doses of mRNA-1273 exceeded those from a pool of convalescent sera. Neutralizing activity was observed for the 100-µg mRNA-1273 dose as of Day 36 and was higher than that of the convalescent sera control group, and the median titers remained in the same range as the median titer in the convalescent sera control group at Day 119 across the age strata. In this study, the majority of the solicited ARs were mild or moderate. A higher incidence of severe solicited ARs was observed with the 250-µg dose (in the 18-55 year age cohort) compared with the lower doses (25 µg, 50 µg, and 100 µg); thus, the mRNA-1273 250-µg dose was not evaluated in participants ≥56 years of age. One severe unsolicited adverse event (AE) related to mRNA-1273 and 1 severe clinically meaningful elevation in serum lipase related to mRNA-1273 were also observed with the 250-µg dose (in the 18-55 year age cohort). Additionally, in Study 20-0003, T helper cell 1 (Th1)-directed CD4+ T-cells were observed to be induced across age groups, with limited indication of a T helper cell 2 (Th2)-directed response, and similar responses were observed among all age groups for the 100-µg dose. The predominance of a Th1-directed T cell profile helps mitigate concern of the risk of enhanced disease associated with Th2-driven pathophysiology.

mRNA-1273-P201 (Study P201) is an ongoing Phase 2a, safety, reactogenicity, and immunogenicity study in healthy adults that provided confirmation of the immunogenicity of both the 100 µg and 50 µg doses. The study was designed as a randomized, observer-blind, placebo-controlled dose confirmation study (Part A). Two mRNA-1273 dose levels, 50 µg and 100 µg, and placebo were evaluated in 2 age cohorts: Cohort 1 enrolled participants ≥ 18 to < 55 years old (300 participants), and Cohort 2 enrolled participants ≥ 55 years old (300 participants). A total of 600 participants received either mRNA-1273 or placebo according to a 1:1:1 randomization ratio; ie, within each age cohort, 100 participants each received mRNA-1273 50 µg, mRNA-1273 100 µg, or placebo.

An amendment to the Study 201 protocol adapted the study design to include open-label interventional phases (Part B and Part C). Part B allowed unblinding of participants and offered 2 injections of mRNA-1273 in an open-label manner, 28 days apart, to all participants who received placebo in Part A. Part B also offered a single BD of mRNA-1273 (50 µg) to participants who received 1 or 2 doses of mRNA-1273 (50 µg or 100 µg) in Part A. Part C was prompted by the need to proactively prepare for vaccination strategies that might induce broader protection, including against emerging variants of SARS-CoV-2 such as B.1.351. Part C enrolled participants from Study 301 who received 2 doses of mRNA-1273 100 µg at least 6 months prior. Part C participants received a single injection of mRNA-1273.351 (20 µg or 50 µg) or mRNA-1273.351 mixture (50 µg total – 25 µg of mRNA-1273 and 25 µg of mRNA-1273.351). mRNA-1273 demonstrated an acceptable safety profile in the participant population enrolled in this study at both dose levels and both age cohorts, as observed through 6 months after the second injection. Vaccination with mRNA-1273 in Study 201 resulted in robust immune responses to SARS-CoV-2 in participants 18 years and older at both dose levels, and persistence of immune response was observed up to 6 months after the second injection in Part A of the study. In Part A of Study 201, the time course and magnitude of antibody (both bAb and nAb) responses to mRNA-1273 was similar between 100 µg and 50 µg dose levels at each postbaseline time point (Days 29, 43, 57, and 209), although the 100 µg dose group had numerically greater responses (and was the dose selected for the pivotal Phase 3 efficacy study). In Part B of Study 201, administration of a 50 µg BD of mRNA-1273 6 months or more after the primary series improved the immune responses to

1.7-fold the peak achieved after the primary vaccination series in the current mRNA-1273-P301 study, where efficacy of mRNA-1273 against COVID-19 was demonstrated. Safety data based on preliminary results for 50 µg BDs of mRNA-1273 (selected participants in Part B) or mRNA-1273.351 (Part C cohort 1) are available. Percentages of participants with solicited local and systemic AEs were similar in the group who received mRNA-1273.351 as a BD compared to those who received mRNA-1273 vaccine as a BD. The majority of solicited local and systemic AEs were mild (grade 1) or moderate (grade 2). The frequency of any grade 3 solicited local or systemic AE was 15% (3 of 20 participants) after the BD of mRNA-1273 and 10.5% (2 of 19 participants) after the BD of mRNA-1273.351. There were no grade 4 solicited local or systemic AEs. The most common solicited local AE was injection site pain after injection in both groups (68.4% for the mRNA-1273.351 vaccine and 90.0% for the mRNA-1273 vaccine). The most common solicited systemic AEs after the BD of the mRNA-1273.351 vaccine were fatigue (36.8%), headache (36.8%), myalgia (31.6%), and arthralgia (21.1%). The most common solicited systemic AEs after the BD of the mRNA-1273 vaccine were fatigue (70.0%), headache (55.0%), arthralgia (50.0%), and myalgia (45.0%). Fever was reported after the BD of mRNA-1273 in 3 of 20 participants (15%) but not after the booster dose of mRNA-1273.351 (0 of 19 participants). There were no SAEs reported in this study.

Currently, a Phase 3 pivotal, randomized, placebo-controlled, observer-blind clinical study (mRNA-1273-P301; NCT04470427; Study P301) is being conducted in participants 18 years of age and older who are at an increased risk of COVID-19 disease. In addition, prespecified cohorts of participants who were either ≥ 65 years of age or 18 to < 65 years of age with comorbid medical conditions were included. A total of 30,351 participants were followed for a median of 92 days (range: 1-122) for the development of COVID-19 disease. Success criteria for early efficacy were met at first IA based on 95 adjudicated cases with a VE of 94.5% (95% CI: 86.5%, 97.8%; one-sided p-value < 0.0001). Study P301 is expected to provide immunogenicity data where an Ab threshold of protection against COVID-19 will be estimated.

In Study P301, solicited adverse reactions (ARs) were reported more frequently among vaccine participants than placebo participants. The most frequently reported ARs after any dose in the vaccine group were pain at the injection site (92.0% any grade; 6.1% grade ≥ 3), fatigue (70% any grade; 10.1% grade ≥ 3), headache (64.7% any grade; 5.7% grade ≥ 3), myalgia (61.5% any grade; 9.1% grade ≥ 3) and chills (45.4% any grade; 1.4% grade ≥ 3). The majority of local and systemic ARs had a median duration of 1 to 3 days. Overall, there was a higher reported rate of some ARs in younger age groups: the incidence of axillary swelling/tenderness, fatigue, headache, myalgia, arthralgia, chills, nausea/vomiting, and fever was higher in adults aged 18 to < 65 years than in those aged 65 years and above. Grade 3 solicited local ARs were more frequently reported after Dose 2 than after Dose 1. In the participants who received the vaccine, solicited systemic ARs were reported numerically more frequently by vaccine participants after Dose 2 than after Dose 1. Grade 3 systemic ARs (fatigue, myalgia, arthralgia, and headache) were reported more frequently after Dose 2 than after Dose 1. Unsolicited AEs and serious adverse events (SAEs) were reported at generally similar rates in participants who received mRNA-1273 and placebo from the first dose until the last observation. Unsolicited AEs that occurred in ≥ 1% of study participants who received mRNA-1273 and at a rate at least 1.5-fold higher rate than placebo, were lymphadenopathy-related events (1.1% of versus 0.6%). All of the lymphadenopathy events are similar to the axillary swelling/tenderness in the injected arm reported as solicited ARs. Hypersensitivity AEs were reported in 1.5% of vaccine recipients and 1.1% of placebo recipients. Hypersensitivity events in the vaccine group included injection site rash and injection site urticaria, which are likely related to vaccination. There have been no cases of severe hypersensitivity or anaphylactic reactions reported immediately after vaccination in the trial to date. There were 3 reports of Bell's palsy in the mRNA-1273 vaccine group (one of which was an SAE), which occurred 22, 28, and 32 days after vaccination, and one in the placebo group, which occurred 17 days after vaccination. The currently available information on Bell's palsy is insufficient to determine a causal relationship with the vaccine. There were 2 SAEs of facial swelling in vaccine recipients with a history of injection of dermatological fillers. The onset of swelling was reported 1 and 2 days, respectively, after vaccination and was likely related to vaccination. There was 1 SAE of intractable nausea and vomiting in a participant with prior history of severe headache and nausea requiring hospitalization. This event occurred 1 day after vaccination and was likely related to vaccination.

There were no other notable patterns or numerical imbalances between treatment groups for specific categories of AEs (including other neurologic, neuro-inflammatory, and thrombotic events) that would suggest a causal relationship to mRNA-1273.

The Sponsor submitted a data package to FDA on 30 November 2020 for consideration of Emergency Use Authorization (EUA). FDA granted an EUA for mRNA-1273 for use in adults 18 years of age and older on 18 December 2020.

Efficacy and safety results based on the final analysis of Part A with a database lock of 04 May 2021 are provided in the IB. The results are generally comparable to the IA. On 25 Aug 2021, Moderna announced that it had completed the rolling submission process for its Biologics License Application to the FDA for the full licensure of the Moderna COVID-19 vaccine (mRNA-1273) for active immunization to prevent COVID-19 in individuals 18 years of age and older. mRNA-1273 has been showing durable efficacy of 93% through 6 months after the second dose.

Interim data from the current P203 study (based on a data snapshot of 08 May 2021) met the primary immunogenicity objective by demonstrating noninferiority of both the (i) GM value of serum neutralizing Ab (nAb) and (ii) the seroconversion rate from adolescent participants compared with those from young adults (aged ≥18 to ≤ 25 years) enrolled in the ongoing Phase 3 efficacy study (Study P301). The geometric mean ratio (GMR) of adolescent (Study P203) to young adult (Study P301) nAb titers at Day 57 was 1.077 (95% CI: 0.939, 1.236), meeting the 1.5-fold noninferiority criterion (ie, lower bound of the 95% CI for GMR is > 0.67). The difference in adolescent to young adult nAb seroresponse rates at Day 57 was 0.2 (95% CI: -1.8, 2.4), meeting the 10% noninferiority criterion (lower bound of the 95% of the seroresponse rate difference is > –10%). In addition, results of VE against COVID-19, defined using either the "Study P301 case definition" or the "CDC case definition", and conducted starting 14 days after dose 1 or dose 2 were all similar to the 94.1% VE observed 14 days post dose 2 in the pivotal P301 study. Overall, local reactogenicity was higher and systemic reactogenicity was lower to mRNA-1273 in adolescents compared with that observed in the adult mRNA-1273-P301 study. Both solicited local and systemic ARs were more commonly reported by participants in the mRNA-1273 group and systemic ARs generally increased after dose 2. While the majority of these solicited ARs were grade 1 or grade 2, there was a higher frequency of grade 3 or higher solicited ARs in the mRNA-1273 group than in the placebo group after dose 1 and dose 2. The majority of the solicited ARs in participants who received mRNA-1273 occurred within the first 1 to 2 days after any dose and generally persisted for a median of 1 to 3 days. The frequency of reported solicited ARs were generally similar and there were no notable differences in the reported rates of unsolicited AEs observed between participants aged ≥ 12 to < 16 years and participants aged ≥ 16 to < 18 years.

Unsolicited treatment-emergent AEs (TEAEs) up to 28 days after any dose were more common in the mRNA-1273 group than in the placebo group. Imbalances in unsolicited TEAEs up to 28 days after any dose was primarily attributable to events related to reactogenicity. The incidence of medically-attended AEs (MAAEs) within 28 days of injection was generally similar between the mRNA-1273 and placebo groups. The incidence of unsolicited severe TEAEs and serious TEAEs in the 28 days after any dose was low overall and generally similar in participants who received mRNA-1273 and those who received placebo. Unsolicited TEAEs, including MAAEs, up to 28 days after any dose assessed by the Investigator as related to study treatment were more frequently reported in the mRNA-1273 group than in the placebo group.

A request for an EUA in the adolescent population has been submitted to the FDA and global filings/approvals down to 12 years of age have occurred. In the post-EUA period, anaphylaxis has been reported following mRNA-1273 administration. There have been very rare reports of myocarditis and pericarditis occurring after vaccination with COVID-19 mRNA vaccines; however, no cases of myocarditis were observed in the clinical program.

A detailed review of the clinical experience with LNPs containing SM-102 (mRNA vaccines and placebo) is provided in the IB.

#### 1.3. Benefit/Risk Assessment

##### 1.3.1. Potential Benefits from Participation

The following benefits may accrue to participants:

- The mRNA-1273 vaccine may be an effective vaccine against COVID-19.
- Participants will have a baseline (Day 1) evaluation for SARS-CoV-2 infection and ongoing surveillance for COVID-19 throughout the study.
- The study will contribute to the development of a vaccine against COVID-19 for adolescents.

##### 1.3.2. Risks to Study Participants and Risk Mitigation

Immediate systemic allergic reactions (eg, anaphylaxis) can occur following any vaccination. These reactions are very rare and are estimated to occur once per 450,000 vaccinations for vaccines that do not contain allergens such as gelatin or egg protein (Zent et al 2002). As a precaution, all participants will remain under observation at the study site for at least 30 minutes after injection.

Vasovagal syncope (fainting) can occur before or after any vaccination, is usually triggered by the pain or anxiety caused by the injection and is not related to the substance injected. Therefore, it is important that standard precautions and procedures are followed to avoid injury from fainting.

Intramuscular injection with other mRNA vaccines manufactured by the Sponsor containing the SM-102 lipid formulation commonly results in a transient and self-limiting local inflammatory reaction. This typically includes pain, erythema (redness), or swelling (hardness) at the injection site, which are mostly mild to moderate in severity and usually occur within 24 hours of injection.

The majority of local and systemic solicited ARs observed after injection with mRNA-1273 at the 100 µg dose level have been mild to moderate in severity (Section 1.2.2). The most commonly reported systemic ARs were headache, myalgia, fatigue, chills, and fever. In the majority of cases, the reactions resolved spontaneously within several days.

There is a theoretical risk that active vaccination to prevent SARS-CoV-2 infection may cause a paradoxical increase in the risk of COVID-19. This possibility is based on the rare phenomenon of vaccine-associated disease enhancement, which was first seen in the 1960s with 2 vaccines made in the same way (formalin-inactivated whole virus) and designed to protect children against infection with respiratory syncytial virus (Chin et al 1969) or measles virus (Fulginiti et al 1967). Disease enhancement has also been proposed as a possible explanation for cases of more serious disease associated with dengue vaccination (Thomas and Yoon 2019; WHO 2019).

In order to address this theoretical risk, animal studies were performed in young and aged wild-type mice and rhesus macaques (NHPs). These studies were designed to capture immunogenicity endpoints that would be predictive of enhanced respiratory disease (ERD) and also to evaluate if, at protective or subprotective dose levels of mRNA-1273, evidence of disease enhancement would be observed after challenge of the animals with SARS-CoV-2. These nonclinical studies demonstrated that mRNA-1273 is safe and well-tolerated in different animal species, is immunogenic; drives a robust SARS-CoV-2-specific Ab, neutralization, and Th1-directed cluster of differentiation (CD)4 T-cell response; fully protects animals from challenge at dose levels as low as 1 µg/dose in mice and 30 µg/dose in NHPs; and does not lead to ERD at protective or subprotective dose levels (Corbett et al 2020a; Corbett et al 2020b). Clinical immunogenicity data from the DMID Phase 1 study of mRNA-1273 demonstrated high levels of nAbs and Th1-polarized CD4+ T-cell responses (Jackson et al 2020), consistent with the immunogenicity observed in these nonclinical studies. These data suggest that a paradoxical increase in the risk of disease, while not eliminated, is likely to be low.

In the context of the EUA for individuals 18 years of age and older for mRNA-1273, there have been very rare reports of myocarditis and pericarditis occurring after vaccination with Moderna COVID-19 vaccine. Although causality has not been established, the majority of the cases have been reported in young males shortly after the second dose of the vaccine. These are typically mild cases and individuals tend to recover within a short time following standard treatment and rest (Gargano et al 2021).

The AEs after receiving a third dose of mRNA-1273 are described in Section 1.2.2. In general, solicited local and systemic adverse reactions after the BD (Study P201) were comparable to what had been observed with the primary series (Study P301).

##### 1.3.3. Overall Benefit/Risk Conclusion

All participants will receive a single dosage of 100 µg of mRNA-1273 vaccine or placebo administered in 2 doses 28 days apart (Section 3.1). Participants in Part A who received mRNA-1273 will be offered an option to receive a third BD of 50 µg of mRNA-1273 at least 6 months from second dose in Part C. Participants in Part A who received placebo will be offered to receive a single dose of 100 µg of mRNA-1273 vaccine administered in 2 doses 28 days apart in Part B and a third BD of 50 µg of mRNA-1273 at least 6 months from second dose in Part C.

Safety will be monitored throughout the study (Section 7.6).

Considering the limited number of approved vaccines for COVID-19, the participants' risk of COVID-19 outside the study during a pandemic, and the nonclinical and clinical data to date, the Sponsor considers the potential benefits of participation to exceed the risks.

### 2. OBJECTIVES AND ENDPOINTS

The objectives which will be evaluated in this study and endpoints associated with each objective are provided in Table 1.

**Table 1: Study Objectives and Endpoints**

| Objectives | Endpoints |
|---|---|
| **Primary Objectives** | **Primary Endpoints** |
| • To evaluate the safety and reactogenicity of 100 µg of mRNA-1273 vaccine administered in 2 doses 28 days apart | • Solicited local and systemic ARs through 7 days after each injection<br>• Unsolicited AEs through 28 days after each injection<br>• MAAEs through the entire study period<br>• SAEs through the entire study period<br>• AESIs through the entire study period<br>• Vital sign measurements<br>• Physical examination findings |
| • To infer efficacy of mRNA-1273 (100 µg, 2 doses 28 days apart), serum Ab responses obtained 28 days after the second injection of mRNA-1273 (Day 57) will be either:<br>– Evaluated against an accepted Ab threshold of protection against COVID-19 (if established in Study P301)<br>– Compared in primary vaccine response as measured by GM values of serum Ab and seroresponse rate in Study P203 with those obtained from young adult recipients (18-25 years of age) of mRNA-1273 in the clinical endpoint efficacy trial (Study P301) | • The proportion of participants with a serum Ab level at Day 57 ≥ an Ab threshold of protection[1]<br>• The primary vaccine response as measured by GM value of serum Ab level and seroresponse rate from Study P203 vaccine recipients at Day 57 compared with those obtained from young adult recipients (18-25 years of age) at Day 57 in the clinical endpoint efficacy trial (Study P301)[2]<br>1. If an accepted serum Ab threshold of vaccine protection against COVID-19 is available, this analysis will form the basis to infer efficacy<br>2. If a threshold is not available, efficacy will be inferred based on establishing noninferiority of adolescent (12 to < 18 years; this clinical study) to adult GM values of serum Ab and seroresponse rate obtained in Study P301 (GM value 12 to < 18 years / GM value 18-25 years) |

| Objectives | Endpoints |
|---|---|
| **Secondary Objectives** | **Secondary Endpoints** |
| • To evaluate the persistence of the immune response of mRNA-1273 vaccine (100 µg) administered in 2 doses 28 days apart, as assessed by the level of SARS-CoV-2 S2P-specific bAb through 1 year after Dose 2 | • The GM value of SARS-CoV-2 S2P-specific bAb on Day 1, Day 57 (1 month after Dose 2), Day 209 (6 months after Dose 2), and Day 394 (1 year after Dose 2) |
| • To evaluate the persistence of the immune response of mRNA-1273 vaccine (100 µg) administered in 2 doses 28 days apart, as assessed by the level of nAb through 1 year after Dose 2 | • The GM values of SARS-CoV-2-specific nAb on Day 1, Day 57 (1 month after Dose 2), Day 209 (6 months after Dose 2), and Day 394 (1 year after Dose 2) |
| • To evaluate the effect of mRNA-1273 on the incidence of SARS-CoV-2 infection compared with the incidence among placebo recipients | • The incidence of SARS-CoV-2 infection (symptomatic or asymptomatic infection) counted starting 14 days after the second dose of IP<br>• SARS-CoV-2 infection will be defined in participants with negative SARS-CoV-2 at baseline:<br>– bAb level against SARS-CoV-2 nucleocapsid protein negative at Day 1 that becomes positive (as measured by Roche Elecsys) at Day 57 or later, OR<br>– Positive RT-PCR counted starting 14 days after the second dose of IP |
| • To evaluate the incidence of asymptomatic SARS-CoV-2 infection after vaccination with mRNA-1273 or placebo | • The incidence of asymptomatic SARS-CoV-2 infection measured by RT-PCR and/or bAb levels against SARS-CoV-2 nucleocapsid protein (by Roche Elecsys) counted starting 14 days after the second dose of IP in participants with negative SARS-CoV-2 at baseline |

| Objectives | Endpoints |
|---|---|
| • To evaluate the incidence of COVID-19 after vaccination with mRNA-1273 or placebo. COVID-19 is defined as clinical symptoms consistent with SARS-CoV-2 infection AND positive RT-PCR for SARS-CoV-2 | • The incidence of the first occurrence of COVID-19 starting 14 days after the second dose of IP, where COVID-19 is defined as symptomatic disease based on the following criteria:<br>– The participant must have experienced at least TWO of the following systemic symptoms: Fever (≥ 38ºC/≥ 100.4ºF), chills, myalgia, headache, sore throat, new olfactory and taste disorder(s), OR<br>– The participant must have experienced at least ONE of the following respiratory signs/symptoms: cough, shortness of breath or difficulty breathing, OR clinical or radiographical evidence of pneumonia; AND<br>– The participant must have at least 1 NP swab, nasal swab, or saliva sample (or respiratory sample, if hospitalized) positive for SARS-CoV-2 by RT-PCR |
| **Exploratory Objectives** | **Exploratory Endpoints** |
| • To evaluate the genetic and/or phenotypic relationships of isolated SARS-CoV-2 strains to the vaccine sequence | • Alignment of genetic sequence of viral isolates with that of the vaccine sequence and comparison of bAb and nAb titers against isolated strain relative to prototype vaccine strain |
| • To describe the ratio or profile of specific bAb relative to nAb in serum | • Relative amounts or profiles of S protein-specific bAb and specific nAb levels/titers in serum |
| • To characterize the clinical profile and immune responses of participants with COVID-19 or with SARS-CoV-2 infection | • Description of clinical severity and immune responses of participants who are identified as infected by SARS-CoV-2 (COVID-19) |
| • To evaluate the incidence of asymptomatic SARS-CoV-2 infection after vaccination with mRNA-1273 or placebo in participants with serologic evidence of infection at baseline | • GM and GMFR of bAb levels against SARS-CoV-2 nucleocapsid protein (quantitative IgG) and % of participants with 2x, 3x, and 4x rise of bAb relative to baseline |

| Objectives | Endpoints |
|---|---|
| **Exploratory Objectives for Part B:** | **Exploratory Endpoints for Part B:** |
| • To evaluate the safety of 2 doses of mRNA-1273 | • MAAEs through the entire study period<br>• SAEs through the entire study period<br>• AESIs through the entire study period<br>• AEs leading to discontinuation from study participation through the last day of study participation |
| • To evaluate the incidence of SARS-CoV-2 or COVID-19 after vaccination with mRNA-1273 | • The incidence of SARS-CoV-2 infection (symptomatic or asymptomatic infection) counted starting 14 days after the second dose of mRNA-1273<br>• To evaluate the incidence of asymptomatic SARS-CoV-2 infection after vaccination with mRNA-1273 measured by RT-PCR and/or bAb levels against SARS-CoV-2 nucleocapsid protein (by Roche Elecsys) counted starting 14 days after the second dose in participants with negative SARS-CoV-2 at baseline<br>• The incidence of the first occurrence of symptomatic COVID-19 starting 14 days after the second dose of mRNA-1273 |
| **Objectives for Part C** | **Endpoints for Part C** |
| **Primary Objectives** | **Primary Endpoints** |
| • To evaluate the safety of the 50 µg BD of mRNA-1273 | • Solicited local and systemic ARs through 7 days after BD<br>• Unsolicited AEs through 28 days after BD injection<br>• MAAEs through the entire study period<br>• SAEs through the entire study period<br>• AESIs through the entire study period<br>• AEs leading to discontinuation from study participation post BD through the last day of study participation |

| Objectives | Endpoints |
|---|---|
| • To infer effectiveness of the 50 µg of mRNA-1273 booster by establishing noninferiority of Ab response after the booster dose compared to the primary series of mRNA-1273.GM values of serum Ab and seroresponse rate of post-booster in Study P203 compared with primary series from young adult (18-25 years of age) recipients of mRNA-1273 in the clinical endpoint efficacy trial (Study P301) | • Co-Primary endpoint(s):<br>– GM of post-booster (post-Dose 3) Ab against original strain in Study P203 as compared to post-primary series (post-Dose 2) against original strain in the young adults in Study P301<br>– Seroresponse rate of post-booster/Dose 3 from baseline (pre-Dose 1) as compared to post-Dose 2 from baseline (pre-Dose1) against original strain in the young adults in Study P301, using 4-fold rise definition<br>o Seroresponse is defined as a titer change from baseline (pre-Dose 1) below the LLOQ to (≥ 4 × LLOQ, or at least a 4-fold rise if baseline is ≥ LLOQ |
| **Key Secondary Objective** | **Key Secondary Endpoint** |
| • To evaluate immune response elicited by the 50 µg prototype booster of mRNA -1273 against variant(s) of interest | • GM of post-booster (post-Dose 3) Ab against circulating strain/Delta as compared to post-primary series (post-Dose 2) against circulating strain/Delta<br>• Seroresponse rate of post-booster/Dose 3 from baseline (pre-Dose 1) as compared to post-Dose 2 from baseline (pre-Dose1) against circulating strain/delta using 4-fold rise definition |
| • **Exploratory Objectives** | • **Exploratory Endpoints** |
| • To evaluate the persistence of the immune response of the BD of mRNA 1273 vaccine (50 µg) as assessed by the level of SARS-CoV-2 S2P specific bAb through 1 year after BD | • The GM value of SARS-CoV-2 S2P specific bAb on BD-Day 1, BD-Day 22 (1 month after BD), BD-Day 181 (6 months after BD), and BD-Day 361 (1 year after BD) |
| • To evaluate the persistence of the immune response of the BD of mRNA 1273 vaccine (50 µg) as | • The GM values of SARS-CoV-2-specific nAb on BD-Day 1, BD-Day 22 (1 month after BD), |

| Objectives | Endpoints |
|---|---|
| assessed by the level of nAb through 1 year after BD | BD-Day 181 (6 months after BD), and BD-Day 361 (1 year after BD) |
| • To evaluate the incidence of SARS-CoV-2 infection or COVID-19 after vaccination with mRNA-1273 | • The incidence of SARS-CoV-2 infection (symptomatic or asymptomatic infection) counted starting 14 days after BD of mRNA-1273<br>• To evaluate the incidence of asymptomatic SARS-CoV-2 infection after vaccination with mRNA-1273 measured by RT-PCR and/or bAb levels against SARS-CoV-2 nucleocapsid protein (by Roche Elecsys) counted starting 14 days after BD in participants with negative SARS-CoV-2 at baseline or pre-booster<br>• The incidence of the first occurrence of symptomatic COVID-19 starting 14 days after BD of mRNA-1273 |

Abbreviations: Ab = antibody; AE = adverse event; AESI = adverse event of special interest; AR = adverse reaction; bAb = binding antibody; BD = booster dose; COVID-19 = coronavirus disease 2019; GM = geometric mean; GMFR = geometric mean fold-rise; IP = investigational product; LLOQ = lower limit of quantification; LOD = limit of detection; MAAE = medically attended adverse event; nAb = neutralizing antibody; NP = nasopharyngeal; RT-PCR = reverse transcriptase polymerase chain reaction; S = spike; S2P = S protein; SAE = severe adverse event; SARS-CoV-2 = Severe Acute Respiratory Syndrome coronavirus 2.

### 3. STUDY DESIGN

#### 3.1. General Design

This is a three-part, Phase 2/3 study: Part A, Part B, and Part C. Participants in Part A, the Blinded Phase of the study, are blinded to their treatment assignment.

Part B, the Open-label Observational Phase of this study, is designed to offer participants who received placebo in Part A of this study and who meet the EUA eligibility criteria an option to receive mRNA-1273 in an open-label fashion (Figure 2). Participants who received mRNA-1273 (100 µg) in Part A of this study will proceed to Part B after they are unblinded and will continue to follow the Part A Schedule of Assessments (SoA). Part C, the Booster Dose Phase, is designed to offer participants in Part A and Part B, and who are at least 6 months from the last dose, the option to request a BD of (50 µg) of mRNA-1273.

Participants who received mRNA-1273 in Part A will be in the study approximately 25 months, which includes 1 month for screening (Day -28 to Day 1), up to 12 months for dosing (on Day 1, Day 29, and Day 209 for BD), and 12 months for follow-up.

Participants who received placebo in Part A will be in the study for approximately 25 months total, which includes approximately 9 months in Part A and approximately 4 months of follow-up following their second dose of mRNA-1273 in Part B before entering Part C or before BD is received; or approximately 12 months of follow-up.

Participants that decline to unblinding or decline to receive a BD will be in the study approximately 14 months total, which includes 1 month for screening (Day -28 to Day 1), 1 month for dosing (on Day 1 and Day 29), and 12 months for follow-up.

This study will be conducted in compliance with the protocol, Good Clinical Practice (GCP), and all applicable regulatory requirements.

##### 3.1.1. Part A, the Blinded Phase

The blinded phase of this study is a randomized, observer-blind, placebo-controlled study intended to infer the effectiveness of mRNA-1273 in an adolescent population aged 12 to < 18 years. The study includes 2 arms: (i) 100 µg of mRNA-1273, and (ii) placebo. Approximately, 3,000 participants between 12 to < 18 years of age will be randomly assigned in a 2:1 ratio to receive mRNA-1273 (n=2,000) or placebo (n=1,000).

The schematic of study arms and major study events for Part A is illustrated in Figure 1 and the SoA for Part A is located in Table 7.

The goal of the study is to seek an indication for use of mRNA-1273 (100 µg IM, given as 2 injections, 28 days apart) in the 12 to < 18 years age group. The basis for demonstrating vaccine effectiveness is proposed to be met by serum Ab response measured in this adolescent age group. The approach to inferring vaccine effectiveness will depend on whether an accepted serum Ab threshold conferring protection against COVID-19 has been established. If an Ab threshold of protection has been established, effectiveness will be inferred based on the proportion of adolescent study participants with serum Ab levels (on Day 57) that meet or exceed the Ab threshold. If an Ab threshold of protection has not been established, effectiveness will be inferred by demonstrating noninferiority of both (i) the GM value of serum nAb and (ii) the seroresponse rate from adolescent participants compared with those from young adults (18-25 years of age) enrolled in the ongoing clinical endpoint efficacy trial (Study P301). The statistical parameters to infer effectiveness are described in Section 2.

This study in adolescents will monitor all participants for a total of 12 months following the second dose of vaccine or placebo. Safety assessments will include solicited ARs (7 days after each injection), unsolicited AEs (28 days after each injection), MAAEs, SAEs, and adverse events of special interest (AESIs) (including multisystem inflammatory syndrome in children [MIS-C]) throughout the study period.

Blood samples will be collected from all participants at baseline (Day 1), Day 57 (28 days after Dose 2), Day 209 (6 months after Dose 2), and Day 394 for measurement of SARS-CoV-2-specific bAb and nAb responses. Blood samples will also be tested for the development of Ab directed against non-vaccine antigen (eg, Ab against the nucleocapsid protein), which will signify infection with SARS-CoV-2. The incidence of SARS-CoV-2 infection among vaccine recipients and placebo recipients will be compared to assess the potential for mRNA-1273 to reduce the rate of infection in vaccine recipients.

**Figure 1: Study Schema (Part A, Blinded Phase)**

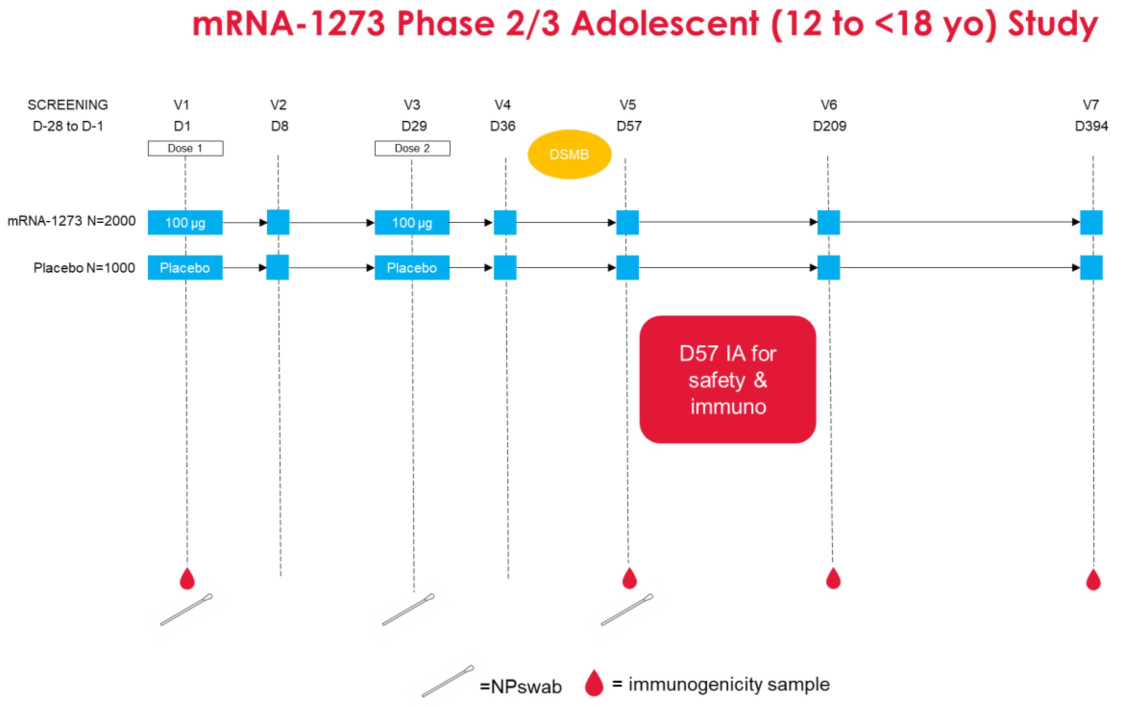

Abbreviation: D = day; DSMB = Data Safety Monitoring Board, IA = interim analysis, immuno = immunogenicity, NP = nasopharyngeal or nasal swab, V = visit, yo = years old.

Part A, the Blinded Phase of the study, comprises 8 scheduled visits including a screening visit and 7 scheduled visits, of which Visit 2 and Visit 4 will be virtual/telephone visits and the other visits will be in-clinic visits.

The study duration will be approximately 14 months, which includes 1 month for screening (Day -28 to Day 1), 1 month for dosing (on Day 1 and Day 29), and, for participants who received mRNA-1273 in Part A, 12 months of follow up after the second dose to monitor for safety, immunogenicity, and efficacy. Participants who received placebo in Part A will still be in the study for approximately 14 months total but will be followed for approximately 4 months following their second dose of mRNA-1273 in Part B before receiving a BD.

Note: Day 0 and Day 1 may be combined on the same day (Table 7).

After providing informed consent/assent, participants will undergo screening assessments to determine study eligibility. Screening assessments (Table 7) must be completed after signing the informed consent form (ICF)/assent form. The investigator will review study entry criteria to determine the participant eligibility during the Screening Period.

Eligible participants will enter the Treatment Period.

On Day 1, after the completion of the scheduled assessments (Table 7), participants will be administered a single IM dose of mRNA-1273 (100 µg) or placebo (procedures will be detailed in the mRNA-1273-P203 Pharmacy Manual). Participants will be closely monitored for safety and will remain at the study site for observation for at least 30 minutes after dosing. On Day 29, the second dose of investigational product (IP) will be administered. Participants will be monitored for 12 months after the second dose of IP for safety and immunogenicity assessments.

To test for the presence of SARS-CoV-2 by RT-PCR, nasopharyngeal (NP) or nasal swab samples will be collected on each day of injection prior to dosing and on Day 57 (28 days postdose 2), according to the SoA (Table 7).

During the course of the study, participants who meet prespecified disease criteria that suggest possible SARS-CoV-2 infection will be asked to contact the study site to arrange for a prompt, thorough, and careful assessment, including an NP or nasal swab sample to be tested for the presence of SARS-CoV-2 by RT-PCR. Confirmed, symptomatic cases of SARS-CoV-2 infection will be captured as MAAEs and reported in an expedited time frame to the Sponsor (Section 7.4.3).

All participants will be monitored for safety and reactogenicity and provide pre- and postdose blood specimens for immunogenicity through 12 months after the second dose of mRNA-1273.

Participants will be instructed on the day of the first dose (Day 1) and reminded on the day of the second dose (Day 29) how to document and report solicited local or systemic ARs in a provided electronic diary (eDiary). Solicited ARs, unsolicited AEs, MAAEs, AEs leading to withdrawal, AESIs, and SAEs will be assessed as described in Section 7.1, according to the time points in the SoA (Table 7).

Blood sampling for immunogenicity testing is scheduled throughout the study: on the day of injection before the first dose and 1, 6, and 12 months after the second dose.

Participants may experience AEs that necessitate an unscheduled visit, including situations when the investigator asks a participant to return to the study clinic for an unscheduled visit following the report of an AE. Additional examinations may be conducted at these visits as necessary to ensure the safety and well-being of participants during the study. Electronic case report forms (eCRFs) should be completed for each unscheduled visit.

##### 3.1.2. Part B, the Open-label Observational Phase

Part B, the Open-label Observational Phase of the study, will be prompted by the authorization of a COVID-19 vaccine under an EUA for any persons under the age of 18 years. Participants will be transitioned to Part B of the study as their age group becomes EUA-eligible. This transition permits all ongoing study participants to eventually be informed of the availability and eligibility criteria of any COVID-19 vaccine made available under an EUA and the option to offer all ongoing study participants an opportunity to schedule a Participant Decision Visit to know their original treatment assignment (placebo vs. mRNA-1273 100 µg vaccine).

Part B provides the opportunity for study participants to be informed regarding the EUA, to be unblinded to their original assignment (mRNA-1273 or placebo), and, for those who previously received placebo, to actively request to receive 2 doses of mRNA-1273 (100 µg) vaccine.

**Figure 2: Study Schema (Part B, Open-label Observational Phase)**

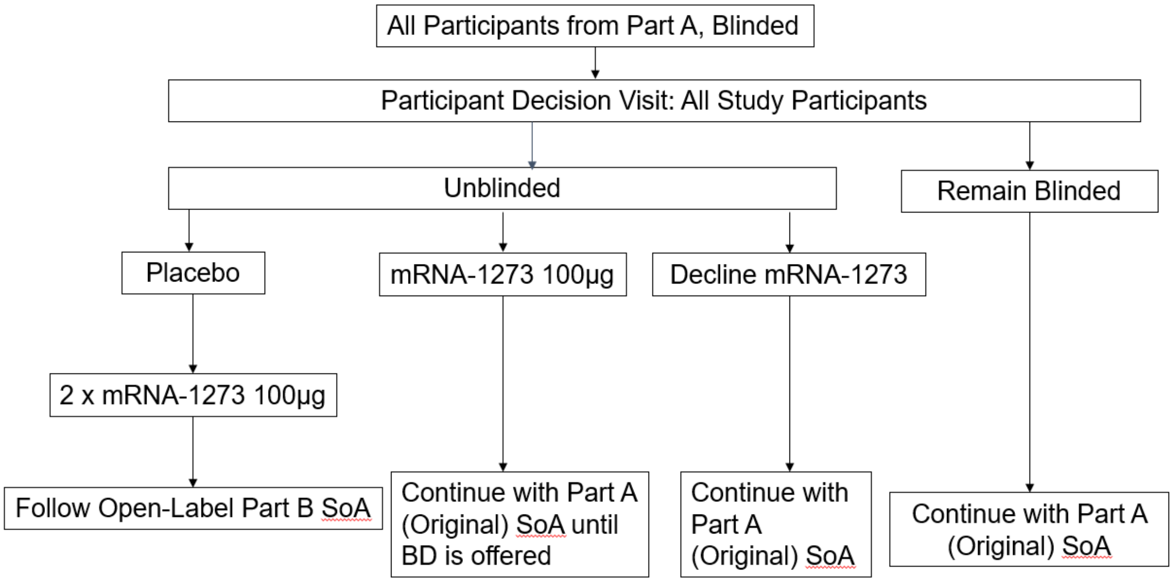

Abbreviation: BD = booster dose; SoA = Schedule of Assessments.

At the Participant Decision Clinic Visit (Table 8), EUA-eligible participants will:

- Be given the option to be unblinded as to their original group assignment (placebo vs. mRNA-1273 vaccine [100 µg]),
- Be counseled about the importance of continuing other public health measures to limit the spread of disease including social distancing, wearing a mask, and hand-washing,
- Sign a revised ICF and assent if not signed at a previous clinic visit, and
- Provide an NP or nasal swab for RT-PCR for SARS-CoV-2 and a blood sample for serology and immunogenicity.

Participants that decline unblinding will remain in Part A and follow the Part A SoA in Table 7.

After the Participant Decision Clinic Visit, participants that consent to unblinding will follow the Part A SoA (Table 7) or Part B SoA (Table 9) as follows:

- Participants received placebo in Part A and consent to unblinding and to receiving 2 doses of mRNA-1273 in Part B: These participants will proceed to Part B and follow the Part B SoA in Table 9 until BD is offered.
- Participants received 2 doses of mRNA-1273 in Part A and consent to unblinding: Due to statistical considerations, these participants will be considered in the Open-label Observational Phase but will continue to follow the Part A SoA in Table 7 until BD is offered.

##### 3.1.3. Part C, Booster Dose Phase

Part C is designed to offer participants in Part A and Part B, who are at least 6 months from the last dose, the option to request a BD (50 µg) of mRNA-1273.

If eligible, each study participant will receive a notification letter and will be asked to schedule a BD-1 visit at their study site. Principal Investigators should consider current local public health guidance for administration of COVID-19 vaccines under EUA and marketing authorization (if any) when determining the scheduling priority of participants.

**Figure 3: Study Schema (Part C, Open-label Booster Dose Observational Phase)**

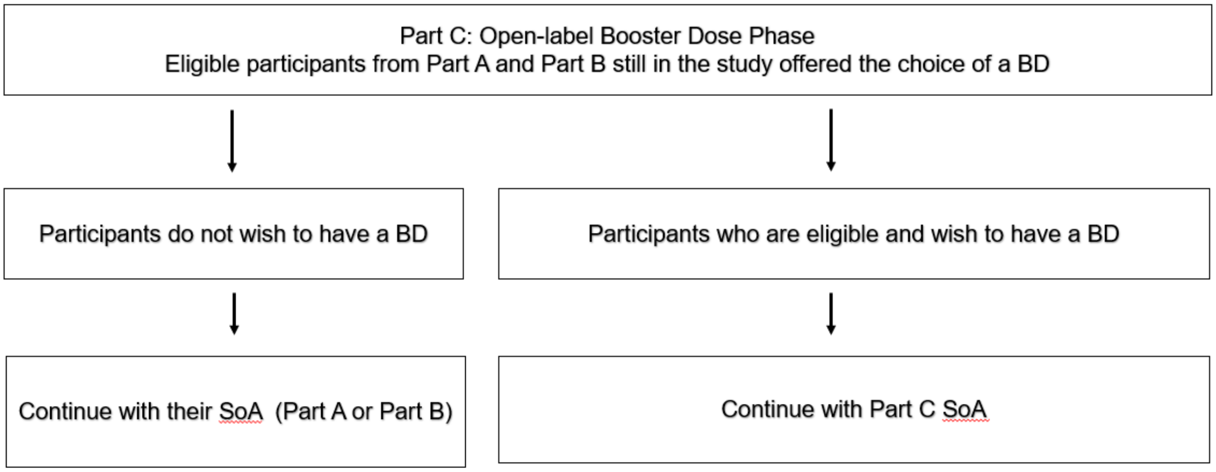

Abbreviations: BD = booster dose; D = day; SoA = Schedule of Assessments

At the BD-1 visit, each participant will:

- Be encouraged to remain in the ongoing study,
- Sign a revised ICF that includes both updated safety information relevant to the ongoing study and a BD, and the option to receive a BD,
- Be given the option to receive a BD consisting of a 50 µg dose of mRNA-1273,
- Be counselled about the importance of continuing other public health measures to limit the spread of disease including social distancing, wearing a mask, and hand-washing.

At the BD-1 visit, participants who request a BD and are eligible will have the following study site visits and complete scheduled activities (subject to investigational vaccine availability) according to the Part C Supplemental SoA:

- BD-1 visit: Participants will receive a single 50 µg dose of mRNA-1273
- BD-1a visit: All participants who chose to receive BD, Day 4, 3 days after BD on Day 1
- BD-2 visit: Day 29, 28 days after the BD on Day 1
- BD-3 visit: Day 181, 180 days after the BD on Day 1
- BD-4 visit: Day 361, 360 days after the BD on Day 1

Participants that crossed over from placebo to mRNA-1273 at and decline BD will continue with Part B SoA (Table 9).

Participants that received mRNA-1273 in Part A and decline BD will continue with in Part A SoA (Table 7).

The investigator is responsible for conducting all assessments as specified in the Part C Supplemental SoA, according to the schedule. As this Supplemental SoA is intended to occur in addition to the original SoAs being followed by all participants in Part A or Part B, there is a possibility for study visits to overlap. If visits overlap according to respective visit windows, a single visit may be done with the combined study procedures completed once.

#### 3.2. Scientific Rationale for Study Design

The single age cohort in this Phase 2/3 study, 12 to < 18 years of age, was established to understand the tolerability and immunogenicity of mRNA-1273 in an adolescent population. The lower age boundary used in this study is consistent with the definition of adolescence provided by the American Academy of Pediatrics (Hardin et al 2017). Knowledge of the tolerability of mRNA-1273 in adolescent participants will be critical before proceeding to future studies in younger children.

With SARS-CoV-2 expected to be circulating in the general population during the study, all participants will provide pre-injection blood samples and postinjection blood samples for Ab analysis through 12 months after the last dose of IP during Part A of the study. In addition, participants will have NP or nasal swab samples collected, before the injections on Day 1 and Day 29, and on Day 57. Furthermore, with any signs or symptoms or MAAE suggesting SARS-CoV-2 infection in a participant, an additional nasal or NP swab sample and a blood sample will be taken to confirm the diagnosis of SARS-CoV-2 via serology and RT-PCR. Additionally, clinical information will be carefully collected to evaluate the severity of the clinical case.

As it is possible that participants are naturally exposed to SARS-CoV-2 through community exposure, the NP or nasal swab samples collected before study injection and the serologic assays for Ab responses to nonvaccine antigen(s) may help discriminate between natural infection and vaccine-induced Ab responses, should such discrimination be needed.

The study design was amended to include a transition to Part B, the Open-Label Observational Phase. During Part B of the study, all ongoing study participants were permitted (a) to be informed of the availability and eligibility criteria of any COVID-19 vaccine made available under an EUA and (b) to schedule a study visit to know their original group assignment (placebo vs. mRNA-1273 vaccine). Part B also provided the opportunity for eligible study participants who previously received placebo to receive 2 doses of mRNA-1273 vaccine under open-label conditions.

Part C, the booster phase, was prompted by the recent interim results of an ongoing Moderna Phase 2 study (mRNA-1273-P201), in which participants who had received 2 doses of 50 µg or 100 µg of mRNA-1273 6-8 months prior were administered a 50 µg booster of mRNA-1273. Participants demonstrated enhanced immune responses compared to pre-boost levels and met the noninferiority criteria stipulated in the FDA Guidance on EUA for Vaccines to Prevent COVID-19. Additionally, no new safety signals emerged upon administration of the BD in Study mRNA-1273-P201. Based on cumulative evidence, the benefit-risk profile of a BD of mRNA-1273 is favorable, particularly in light of increasing breakthrough disease with the emergence of the Delta variant. Providing the option for a BD to all participants currently enrolled in Part B or who received mRNA-1273 in Part A is expected to generate booster data in the adolescent population.

#### 3.3. Justification for Dose, Control Product, and Choice of Study Population

The 100 µg dose level is currently being investigated in a large Phase 3 efficacy study in adults 18 years of age and older; therefore, based on this and the results of the studies described in Section 1.2.2, the Sponsor intends to study a single dose level of 100 µg in this Phase 2/3 study in the adolescents age group of 12 to < 18 years of age.

At the time of study initiation, there were no licensed SARS-CoV-2 vaccines available, and so 0.9% sodium chloride will be used as a placebo control for the safety and immunogenicity assessments. The mRNA-1273 vaccine and placebo injections look different, so administration was blinded in Part A of this study (Section 8.1).

Following potential EUA of a COVID-19 vaccine for persons under 18 years of age, this study amendment is designed to give EUA-eligible participants the opportunity to transition to Part B, the Open-label Observational Phase (Figure 2). Transitioning the study to Part B, Open-label Observational Phase, permits (a) all ongoing study participants to be informed of the availability and eligibility criteria of any COVID-19 vaccine made available under an EUA and (b) the option to offer all ongoing study participants who request unblinding an opportunity to schedule a study visit to know their original group assignment (placebo vs. mRNA-1273 [100 µg vaccine]). Part B, the Open-label Observational Phase, also provides the opportunity for study participants who previously received placebo to request to receive 2 doses of mRNA-1273 (100 µg) vaccine.

For Part C, provision for the option of a single BD for eligible participants during the Open-Label Observational Phase, the BD selected is 50 µg, based primarily on data from Study mRNA-1273-P201. In Part A of Study mRNA-1273-P201, the time course and magnitude of antibody (both bAb and nAb) responses to mRNA-1273 was similar between 100 µg and 50 µg dose levels at each postbaseline time point (Days 29, 43, 57, and 209), although the 100 µg dose group had numerically greater responses. In Part B of Study mRNA-1273-P201, administration of a 50 µg BD of mRNA-1273 6 months or more after the primary series, improved the immune responses to 1.7-fold the peak achieved after the primary vaccination series in the current mRNA-1273-P301 study, where efficacy of mRNA-1273 against COVID-19 was demonstrated.

#### 3.4. End-of-Study Definition

The end-of-study (EOS) for the full study is defined as completion of the last visit of the last participant in the study or the last scheduled procedure as shown in the Part C SoA (Table 10) or Part A, Table 7 (if remained blinded or decline to receive a BD) for the last participant in this study.

### 4. STUDY POPULATION (PART A: BLINDED PHASE AND PART B: OPEN LABEL OBSERVATIONAL PHASE)

Participants will be enrolled at 25 study sites in the United States or its territories.

Prospective approval of protocol deviations to recruitment and enrollment criteria, also known as protocol waivers or exemptions, is not permitted.

#### 4.1. Eligibility Criteria (Part A)

##### 4.1.1. Inclusion Criteria

Each participant must meet all of the following criteria at the Screening Visit (Day 0) or at Day 1, unless noted otherwise, to be enrolled in this study:

1. Male or female, 12 to < 18 years of age at the time of consent (Screening Visit, Day 0) who, in the opinion of the investigator, is in good general health based on review of medical history and screening physical examination.
2. Investigator assessment that the participant, in the case of an emancipated minor, or parent(s)/legally acceptable representative(s) [LAR(s)] understand and are willing and physically able to comply with protocol-mandated follow-up, including all procedures and provides written informed consent/assent.
3. Body mass index (BMI) at or above the third percentile according to WHO Child Growth Standards at the Screening Visit (Day 0); see Section 10.2.18.
4. Female participants of nonchildbearing potential may be enrolled in the study. Nonchildbearing potential is defined as premenarche or surgically sterile (history of bilateral tubal ligation, bilateral oophorectomy, hysterectomy).
5. Female participants of childbearing potential may be enrolled in the study if the participant fulfills all the following criteria:
    - Has a negative pregnancy test at Screening (Day 0), on the day of the first injection (Day 1), and on the day of the second injection (Day 29)
    - Has practiced adequate contraception or has abstained from all activities that could result in pregnancy for at least 28 days prior to the first injection (Day 1)
    - Has agreed to continue adequate contraception or abstinence through 3 months following the second injection (Day 29)

Adequate female contraception is defined as consistent and correct use of a US FDA-approved contraceptive method in accordance with the product label (Section 10.3).

##### 4.1.2. Exclusion Criteria

Participants who meet any of the following criteria at the Screening Visit (Day 0) or at Day 1, unless noted otherwise, will be excluded from the study:

1. Travel outside of the United States in the 28 days prior to the Screening Visit (Day 0).
2. Pregnant or breastfeeding.
3. Is acutely ill or febrile 24 hours prior to or at the Screening Visit (Day 0). Fever is defined as a body temperature ≥ 38.0°C/≥ 100.4°F. Participants who meet this criterion may have visits rescheduled within the relevant study visit windows. Afebrile participants with minor illnesses can be enrolled at the discretion of the investigator.
4. Prior administration of an investigational CoV (eg, SARS-CoV-2, SARS-CoV, MERS-CoV) vaccine.
5. Current treatment with investigational agents for prophylaxis against COVID-19.
6. Has a medical, psychiatric, or occupational condition that may pose additional risk as a result of participation, or that could interfere with safety assessments or interpretation of results according to the investigator's judgment.
7. Current use of any inhaled substance (eg, tobacco or cannabis smoke, nicotine vapors).
8. History of chronic smoking (≥ 1 cigarette a day) within 1 year of the Screening Visit (Day 0).
9. History of illegal substance use or alcohol abuse within the past 2 years. This exclusion does not apply to historical cannabis use that was formerly illegal in the participant's state but is legal at the time of screening.
10. History of a diagnosis or condition that, in the judgment of the investigator, may affect study endpoint assessment or compromise participant safety, specifically:
    - Congenital or acquired immunodeficiency, including human immunodeficiency virus (HIV) infection.
    - Suspected active hepatitis
    - Has a bleeding disorder that is considered a contraindication to IM injection or phlebotomy
    - Dermatologic conditions that could affect local solicited AR assessments
    - History of anaphylaxis, urticaria, or other significant AR requiring medical intervention after receipt of a vaccine

- Diagnosis of malignancy within the previous 10 years (excluding nonmelanoma skin cancer)
- Febrile seizures

11. Receipt of:
    - Any licensed vaccine within 28 days before the first dose of IP or plans for receipt of any licensed vaccine through 28 days following the last dose of IP
    - Systemic immunosuppressants or immune-modifying drugs for > 14 days in total within 6 months prior to the day of enrollment (for corticosteroids, ≥ 20 mg/day prednisone equivalent). Topical tacrolimus is allowed if not used within 14 days prior to the day of enrollment. Participants may have visits rescheduled for enrollment if they no longer meet this criterion within the Screening Visit window. Inhaled, nasal, and topical steroids are allowed.
    - Intravenous blood products (red cells, platelets, immunoglobulins) within 3 months prior to enrollment
12. Has donated ≥ 450 mL of blood products within 28 days prior to the Screening Visit (Day 0) or plans to donate blood products during the study.
13. Participated in an interventional clinical study within 28 days prior to the Screening Visit (Day 0) or plans to do so while participating in this study.
14. Is an immediate family member or has a household contact who is an employee of the research center or otherwise involved with the conduct of the study.

##### 4.2. Study Eligibility Criteria (Part B)

1. Participants must have been previously enrolled in the mRNA-1273-P203 study.
2. Female participants of childbearing potential may be enrolled in the study if the participant has a negative pregnancy test on the day of the first injection (OL-Day 1) and on the day of the second injection (OL-Day 29).

#### 4.3. Study Eligibility Criteria (Part C)

Inclusion Criteria:

1. Participants must have been previously enrolled in the mRNA-1273-P203 study, are actively participating in Part A or Part B and are least 6 months from the last dose.
2. Female participants of childbearing potential may be enrolled in the study if the participant has a negative pregnancy test on the day of the first injection (BD-Day 1).

Exclusion Criteria:

1. Pregnant or breastfeeding.
2. Is acutely ill or febrile 24 hours prior to or at the Screening Visit (Day 0). Fever is defined as a body temperature ≥ 38.0°C/≥ 100.4°F. Participants who meet this criterion may have visits rescheduled within the relevant study visit windows. Afebrile participants with minor illnesses can be enrolled at the discretion of the investigator.
3. Has a medical, psychiatric, or occupational condition that may pose additional risk as a result of participation, or that could interfere with safety assessments or interpretation of results according to the investigator's judgment.
4. History of a diagnosis or condition (after enrolment in Part A) that, in the judgment of the investigator, may affect study endpoint assessment or compromise participant safety:
    - Suspected active hepatitis
    - Has a bleeding disorder that is considered a contraindication to IM injection or phlebotomy
    - Dermatologic conditions that could affect local solicited AR assessments
    - History of anaphylaxis, urticaria, or other significant AR requiring medical intervention after receipt of a vaccine
    - Diagnosis of malignancy (excluding nonmelanoma skin cancer)
5. Receipt of:
    - Any authorized or licensed vaccine within 28 days before the first dose of IP or plans for receipt of any licensed vaccine through 28 days following the last dose of IP or any seasonal influenza vaccine within 14 days before the first dose of IP or plans for receipt of any seasonal vaccine 14 days following the last dose of IP.
6. Participated in an interventional clinical study, other than mRNA-1273-P203, within 28 days prior to the Screening Visit (Day 0) or plans to do so while participating in this study.

#### 4.4. Lifestyle Restrictions

Participants must not eat or drink anything hot or cold within 10 minutes before oral temperature is taken.

#### 4.5. Screen Failures (Part A: Blinded Phase Only)

Screen failures are defined as participants who consent to participate in the clinical study but are not subsequently randomly assigned to treatment. A minimum set of screen failure information is required to ensure transparent reporting of screen failures to meet the Consolidated Standards of Reporting Trials (CONSORT) publishing requirements and to respond to queries from regulatory authorities. Minimum information includes date of informed consent, demography, reason(s) for screen failure, eligibility criteria, and information on any SAE that may have occurred from Day 1 to the time of withdrawal.

### 5. STUDY TREATMENT

#### 5.1. Investigational Product Administered

The term IP refers to mRNA-1273 (100 µg for Part A and Part B, and 50 µg for Part C) vaccine or placebo (0.9% sodium chloride) in this study.

The mRNA-1273 is an LNP dispersion of an mRNA encoding the prefusion stabilized S protein of SARS-CoV-2 formulated in LNPs composed of 4 lipids (1 proprietary and 3 commercially available): the proprietary ionizable lipid SM-102; cholesterol; 1,2-distearoyl-sn-glycero-3-phosphocholine (DSPC); and 1-monomethoxypolyethyleneglycol-2,3-dimyristylglycerol with polyethylene glycol of average molecular weight 2000 (1-monomethoxypolyethyleneglycol-2,3-dimyristylglycerol polyethylene glycol 2000 [PEG2000-DMG]). mRNA-1273 injection is provided as a sterile liquid for injection, white to off-white dispersion in appearance, at a concentration of 0.5 mg/mL in 20 mM Tris buffer containing 87 mg/mL sucrose and 10.7 mM sodium acetate at pH 7.5.

#### 5.2. Randomization

Random assignment of participants will use a centralized interactive response technology, in accordance with pregenerated randomization schedules.

#### 5.3. Dosing and Management of mRNA-1273 Vaccine

##### 5.3.1. Preparation of Study Vaccine for Injection

Each dose of IP will be prepared for each participant based on the assigned treatment, as detailed in the mRNA-1273-P203 Pharmacy Manual. For Part A and Part B, the volume of IP injected will be 0.5 mL consisting of either 100 µg dose of mRNA-1273 or placebo (normal saline), as detailed in the mRNA-1273-P203 Pharmacy Manual. For Part C, each injection will have a volume of 0.25 mL and contain mRNA-1273 50 µg.

##### 5.3.2. Administration of Study Vaccine

In the Blinded Phase, Part A of the study, each participant will receive 2 doses of IP by IM injection, 28 days apart (ie, Day 1 and Day 29) into the deltoid muscle, according to their assigned regimen and according to the procedures specified in the mRNA-1273-P203 Pharmacy Manual. Preferably, both doses should be administered into the nondominant arm.

In the open-label Part B of the study, mRNA-1273 vaccine will be administered as an IM injection into the deltoid muscle following the injection schedule for each group based on the product received in Part A. Participants who received placebo in Part A will receive 2 doses of mRNA-1273 (100 µg) on open-label (OL) – Day 1 and OL-Day 29 of Part B (Table 9). Preferably, both doses should be administered into the nondominant arm.

For Part C, mRNA-1273 vaccine will be administered as an IM injection into the deltoid muscle. Preferably, the dose should be administered into the nondominant arm. Each injection will have a volume of 0.25 mL and contain mRNA-1273 50 µg.

The IP will be prepared for injection as a single 0.5 mL (Part A and Part B) or 0.25 mL (Part C) dose for each participant Per Protocol (PP), as detailed in the Pharmacy Manual.

At each visit when IP is administered, participants will be monitored for a minimum of 30 minutes after administration. Assessments will include vital sign measurements and monitoring for local or systemic reactions (SoA, Table 7 and Table 9).

Eligibility for a subsequent dose of IP will be determined by following the criteria outlined in Section 6.

The study sites will be appropriately staffed with individuals with basic cardiopulmonary resuscitation training/certification. Either on-site resuscitation equipment and personnel or appropriate protocols for the rapid transport of participant to a resuscitation area or facility are required.

##### 5.3.3. Study Vaccine Delivery and Receipt

The Sponsor or designee is responsible for the following:

- Supplying the IP
- Confirming the appropriate labeling of the IP, so that it complies with the legal requirements of the United States

The investigator is responsible for acknowledging the receipt of the IP by a designated staff member at the study site, including the following:

- Confirming that the IP was received in good condition
- Confirming that the temperature during shipment from the Sponsor to the investigator's designated storage location was appropriate
- Confirming that the Sponsor has authorized the IP for use
- Ensuring the appropriate dose level of IP is properly prepared using aseptic technique

Further description of the IP and instructions for the receipt, storage, preparation, administration, accountability, and destruction of the IP are described in the mRNA-1273-P203 Pharmacy Manual.

##### 5.3.4. Study Vaccine Packaging and Labeling

The Sponsor will provide the investigator (via the study site pharmacy) with adequate quantities of IP. The sterile IP is packaged in 10R glass vials with a 5.0-mL fill volume (Part A, Part B, and

Part C) or a 10R glass vial with an 8.0 mL fill volume (Part C only). The IP will have all required labeling per regulations and will be supplied to the pharmacy in an unblinded manner.

The IP will be packaged and labeled in accordance with the standard operating procedures of the Sponsor or of its designee, Code of Federal Regulations (CFR) Title 21, Good Manufacturing Practice guidelines, International Council for Harmonisation (ICH) GCP guidelines, guidelines for Quality System Regulations, and applicable regulations.

##### 5.3.5. Study Vaccine Storage

The IP must be stored as per the temperature conditions printed on the IP label in a secure area with limited access and protected from moisture and light until it is prepared for administration (Section 5.3.1). The refrigerator and/or freezer utilized for IP storage should have automated temperature recording and a 24-hour alert system in place that allows for rapid response in case of refrigerator malfunction. There must be an available backup refrigerator and freezer. The refrigerators and freezers must be connected to a backup generator(s). In addition, IP accountability study staff are required to keep a temperature log to establish a record of compliance with these storage conditions. The study site is responsible for reporting any IP that was not temperature controlled during shipment or during storage. Such IP will be retained for inspection by the monitor and disposed of according to approved methods.

##### 5.3.6. Study Vaccine Accountability

It is the investigator’s responsibility that the IP accountability study staff maintain accurate records in an IP accountability log of receipt of all IP, study site IP inventory, IP dispensing, IP injections, and return to the Sponsor or alternative disposition of used and unused IP vials.

A study site monitor will review the inventory and accountability log during study site visits and at the completion of the study. Additional details are found in the mRNA-1273-P203 Pharmacy Manual.

##### 5.3.7. Study Vaccine Handling and Disposal

A study site monitor will reconcile the IP inventory during the conduct and at the end of the study for compliance. Once fully reconciled at the study site at the end of the study, the IP can be destroyed at the investigational site or at a Sponsor-selected third party, as appropriate.

Vaccine may be destroyed at the study site only if permitted by local regulations and authorized by the Sponsor. A certificate of destruction must be completed and sent to the Sponsor or designee.

#### 5.4. Study Treatment Compliance

All doses of IP will be administered at the study site under direct observation of medically qualified study staff and appropriately recorded (date and time) in the eCRF. Qualified study site staff will confirm that the participant has received the entire dose of IP. If a participant does not receive IP or does not receive all of the planned dose, the reason for the missed dose will be recorded. Data will be reconciled with study site accountability records to assess compliance.

Participants who miss the second or third dose due to noncompliance with the visit schedule and not due to a safety pause will still be required to follow the original visit and testing schedule as described in the protocol and their regimen schedule. Unless consent is withdrawn, a participant who withdraws or is withheld from receiving the second dose will remain in the study and complete all safety and immunogenicity assessments required through the participant's last scheduled study visit.

The study site staff are responsible for ensuring that participants comply with the allowed study visit windows. If a participant misses a visit, every effort should be made to contact the participant and complete a visit within the defined visit window (Part A, Table 7; Part B, Table 9; and Part C, Table 10). If a participant does not complete a visit within the time window, that visit will be classified as a missed visit and the participant will continue with subsequent scheduled study visits. All safety requirements of the missed visit will be captured and included in the subsequent visit.

#### 5.5. Prior and Concomitant Medications

##### 5.5.1. Prior Medications and Therapies

Information about prior medications (including any prescription or over-the-counter medications, vaccines, or blood products) taken by the participant within the 28 days before providing informed consent/assent (or as designated in the inclusion/exclusion requirements) will be recorded in the participant's eCRF.

##### 5.5.2. Concomitant Medications and Therapies

At each study visit, study site staff must question the participant and/or the participants' parent(s)/LAR(s) regarding any medications taken and vaccinations received by the participant and record the following information in the eCRF:

- All nonstudy vaccinations administered within the period starting 28 days before the first dose of IP.
- All concomitant medications and nonstudy vaccinations taken through 28 days after each dose of IP. Antipyretics and analgesics taken prophylactically (ie, taken in the absence of any symptoms in anticipation of an injection reaction) will be recorded as such.
- Any concomitant medications relevant to or for the treatment of an SAE or an MAAE.
- Participants will be asked in the eDiary if they have taken any antipyretic or analgesic to treat or prevent fever or pain within 7 days after each dose of IP, including the day of

injection. Reported antipyretic or analgesic medications should be recorded in the source document by the study site staff during the postinjection study visits or via other participant interactions (eg, telephone calls).

##### 5.5.3. Recording of Concomitant Medications and Concomitant Vaccinations

Study site staff must question the participant regarding any medications taken and vaccinations received by the participant and record the following information in the eCRF:

- All nonstudy vaccinations administered within the period starting 28 days before the first dose of IP.
- Seasonal influenza vaccine administered for the current influenza season (typically October through April in the Northern Hemisphere).
- All concomitant medications and nonstudy vaccinations taken through 28 days after each dose of IP. Antipyretics and analgesics taken prophylactically (ie, taken in the absence of any symptoms in anticipation of an injection reaction) will be recorded as such.
- Any concomitant medications used to prevent or treat COVID-19 or its symptoms.
- Any concomitant medications relevant to or for the treatment of an SAE or an MAAE.
- Participants will be asked in the eDiary if they have taken any antipyretic or analgesic to treat or prevent fever or pain within 7 days after each IP dose, including on the day of dosing. Reported antipyretic or analgesic medications should be recorded in the source document by the study site staff during the postinjection study visits or via other participant interactions (eg, phone calls).

Concomitant medications (including vaccinations) will be coded using the WHO Drug Dictionary. If a participant takes a prohibited drug therapy, the investigator and the contract research organization's (CRO's) medical monitor will make a joint decision about continuing or withholding further injection of the participant based on the time the medication was administered, the drug's pharmacology and pharmacokinetics, and whether use of the medication will compromise the participant's safety or interpretation of the data. It is the investigator's responsibility to ensure that details regarding the concomitant medications are adequately recorded in the eCRF.

##### 5.5.4. Concomitant Medications and Vaccines that May Lead to the Elimination of a Participant from Per-Protocol Analyses

The use of the following concomitant medications and/or vaccines will not require withdrawal of the participant from the study (with the exception of planned or actual receipt of a non-study

COVID-19 vaccine either under EUA or licensed) but may determine a participant's eligibility to receive a second dose or evaluability in the PP analysis (analysis sets are described in Section 8.4):

- Any investigational or nonregistered product (drug or vaccine) other than the IP used during the study period.
- Immunosuppressants or other immune-modifying drugs administered chronically (ie, more than 14 days in total) during the study period. For corticosteroids, this will mean that prednisone ≥ 20 mg/day or the equivalent is not permitted. Inhaled, nasal, and topical steroids are allowed.
- Long-acting immune-modifying drugs administered at any time during the study period (eg, infliximab).
- Immunoglobulins and/or any blood products administered during the study period.

#### 5.6. Intervention After the End of the Study

Any SAE occurring after the end of the study and considered to be caused by the IP must be reported to the Sponsor.

### 6. DELAYING OR DISCONTINUING STUDY TREATMENT AND PARTICIPANT WITHDRAWAL FROM THE STUDY

#### 6.1. Criteria for Delay of Vaccine Administration

##### 6.1.1. Individual Participant Criteria for Delay of Study Vaccination

Body temperature (oral) must be measured on dosing visits before vaccine administration. The following events constitute criteria for delay of injection, and if either of these events occur at the time scheduled for dosing, the participant may receive the study injection at a later date within the time window specified in the relevant SoA (Table 7), or the participant may be discontinued from dosing at the discretion of the investigator (Section 6.2):

- Acute moderate or severe infection with or without fever at the time of dosing
- Fever, defined as body temperature ≥ 38.0°C/≥ 100.4°F at the time of dosing

Participants with a minor illness without fever, as assessed by the investigator, can be vaccinated. Participants with a fever of ≥ 38.0°C/≥ 100.4°F will be contacted within the time window acceptable for participation and re-evaluated for eligibility. If the investigator determines that the participant's health on the day of dosing temporarily precludes injection, the visit should be rescheduled within the allowed interval for that visit if possible or at a time the participant is clinically stable according to the judgement of the investigator.

If a participant takes a prohibited drug therapy, an injection could be delayed within the visit window based on the joint decision of the investigator and the CRO's medical monitor (Section 5.5.3).

#### 6.2. Discontinuing Study Vaccination

Participants can discontinue study injection (ie, refuse the second dose) for any reason, without prejudice to further treatment the participant may need to receive.

The investigator, in consultation with the Sponsor's medical monitor, may withhold a participant from further injection if the participant experiences any of the following:

- Becomes pregnant
- Withdrawal of consent (not related to COVID-19)
- Develops, during the course of the study, symptoms or conditions listed in the exclusion criteria (Section 4.1.2)
- Experiences an AE (other than reactogenicity) after injection that is considered by the investigator to be related to IP (Section 7.5.9) and is of Grade 3 (severe) or greater severity

- Experiences an AE or SAE that, in the judgment of the investigator, requires IP withdrawal due to its nature, severity, or required treatment, regardless of the causal relationship to vaccine
- Experiences an AESI
- Experiences a clinically significant change in vital sign measurements, or general condition that, in the judgment of the investigator, requires vaccine withdrawal
- Experiences anaphylaxis clearly related to IP
- Experiences generalized urticaria related to IP

The reason(s) for withdrawal from further injection will be recorded in the eCRF.

If a participant takes a prohibited drug therapy, the investigator could withhold the second dose based on a joint decision of the investigator and the CRO's medical monitor (Section 5.5.3).

Every reasonable attempt will be made to follow up with participants for safety throughout the entire scheduled study period according to their regimen, even if the participant does not receive the second dose or misses one or more visits. Unless participants withdraw consent, they are expected to remain in the study and complete all scheduled visits and assessments.

#### 6.3. Participant Discontinuation/Withdrawal from the Study

Participants who withdraw or are withdrawn from the study will not be replaced. A "withdrawal" from the study refers to a situation wherein a participant does not return for the final visit planned in the protocol. The statistical management of participant withdrawals is discussed in Section 8.

Participants can withdraw consent and withdraw from the study at any time, for any reason, without prejudice to further treatment the participant may need to receive. The investigator will request that the participant complete all study procedures pending at the time of withdrawal.

If participant desires to withdraw from the study because of an AE, the investigator will try to obtain agreement to follow up with the participant until the event is considered resolved or stable and will then complete the EOS eCRF.

Information related to the withdrawal will be documented in the eCRF. The investigator will document whether the decision to withdraw a participant from the study was made by the participant or by the investigator, as well as which of the following possible reasons was responsible for withdrawal:

- AE (specify)
- SAE (specify)

- Death
- Lost to follow-up (LTFU)
- Physician decision (specify)
- Pregnancy
- Protocol deviation
- Study terminated by Sponsor
- Withdrawal of consent by participant (specify)
- Other (specify)

Participants who are withdrawn from the study because of AEs (including SAEs) must be clearly distinguished from participants who are withdrawn for other reasons. Investigators will follow up with participants who are withdrawn from the study as result of an SAE or AE until resolution of the event.

A participant who withdraws from the study may request destruction of any samples taken and not tested, and the investigator must document this in the study site study records.

If the participant withdraws consent for disclosure of future information, the Sponsor may retain and continue to use any data collected before such a withdrawal of consent (Section 10.2.10).

The Sponsor will continue to retain and use all research data that have already been collected for the study evaluation, unless the participant has requested destruction of these samples. All biological samples that have already been collected may be retained and analyzed at a later date (or as permitted by local regulations).

#### 6.4. Study Pause Rules

During Part A of the study, the investigators, study medical monitor, and Sponsor will monitor for events that could trigger a study pause. Study pause rule criteria, events, and thresholds are described in Table 2. Although these pause rules are not applicable during Part B or Part C, participants will continue to be monitored for the events in Table 2, and the Sponsor will be notified if any of these events occur.

**Table 2: Pause Rule Criteria, Events, and Thresholds**

| Pause Rule Criterion | Event | Participant Threshold for Triggering Study Pause |
|---|---|---|
| 1 | Any death due to SARS-CoV-2 infection | ≥ 1 |
| 2 | Any related SAE or related Grade 4 AE | ≥ 1 |
| 3 | Hospitalization due to SARS-CoV-2 infection | ≥ 1 |
| 4[a] | Any Grade 3 or higher solicited local AR lasting ≥ 24 hours and occurring within 7-days of injection (Days 1-8) | ≥ 30 participants out of the first 300 participants enrolled |
| 5[a] | Any Grade 3 or higher solicited systemic AR lasting ≥ 24 hours and occurring within 7-days of injection (Days 1-8) | ≥ 30 participants out of the first 300 participants enrolled |
| 6[a] | Any ≥ Grade 3 or higher unsolicited AR that cannot be reasonably attributed to a cause other than vaccination | ≥ 30 participants out of the first 300 participants enrolled |

Abbreviations: AE = adverse event; AR = adverse reaction; ICU = intensive care unit; SAE = serious adverse event; SARS-CoV-2 = Severe Acute Respiratory Syndrome coronavirus.
[a.] Pause Rules 4, 5, and 6 apply only to the first 300 participants enrolled.

If any of the thresholds for a study pause is met during Part A, the Sponsor will immediately suspend further enrollment, pause study dosing, and notify all investigators. Such a suspension will remain in force until the threshold event(s) is (are) reviewed by the Data Safety Monitoring Board (DSMB) and a recommendation to continue is provided to the Sponsor.

The investigator or designee is responsible for reporting to the Sponsor, via the electronic data capture (EDC) system within 24 hours of observation, each event that potentially meets any pause rule criterion. The Sponsor will inform the DSMB of any event that potentially meets any pause rule criterion. The DSMB will review all available study data to adjudicate such events in accordance with the DSMB charter.

The Sponsor will notify the Center for Biologics Evaluation and Research (CBER) within 48 hours in the event of a study pause. In the event of a study pause, all safety and immunogenicity assessments will continue PP. The window allowance for injection visits may be extended by an additional 7 days (ie, +14 days) for affected participants at the discretion of the Sponsor.

During Part B and Part C of the study, the Sponsor will continue to inform the DSMB of the occurrence of any of the events in Table 2. The DSMB will review all available relevant study data to adjudicate such events in accordance with the DSMB charter.

#### 6.5. Lost to Follow-up

A participant will be considered LTFU if he or she repeatedly fails to return for scheduled visits without stating an intention to withdraw consent and is unable to be contacted by the study site. The following actions must be taken if a participant fails to return to the clinic for a required study visit:

- The study site staff must attempt to contact the participant and reschedule the missed visit as soon as possible, counsel the participant on the importance of maintaining the assigned visit schedule and ascertain whether the participant wishes to and/or should continue in the study.
- Before a participant is deemed LTFU, the investigator or designee must make every effort to regain contact with the participant (where possible, 3 telephone calls and, if necessary, a certified letter to the participant’s last known mailing address or local equivalent methods). These contact attempts (eg, dates of telephone calls and registered letters) should be documented in the participant’s medical record.
- A participant who continues to be unreachable or continues to be noncompliant with study visits or procedures will be considered to have withdrawn from the study.
- A participant should not be considered LTFU until due diligence has been completed.

### 7. STUDY ASSESSMENTS AND PROCEDURES

Before performing any study procedures, all potential participants and/or participants' parent/LAR will sign an ICF (as detailed in Section 10.2.6). Participants will undergo study procedures at the time points specified in the Part A SoA (Table 7), Part B SoA (Table 9), and/or Part C (Table 10).

After the participant proceeds to the Participant Decision Clinic Visit (Part A) of the study, participants who received mRNA-1273 in Part A will continue to follow the open-label Part A SoA (Table 7). Participants who received placebo in Part A will transition to the open-label Part B of the study (Figure 5) and will follow the Part B SoA (Table 9). Participants in Part A or Part B who transitioned to Part C, 6 months from the last dose, will follow open-label Part C SoA (Table 10) until study completion. Participants who remained blinded will follow Part A SoA. Participants who do not receive a BD will continue with their SoA (Part A or Part B).

A participant can also be seen for an unscheduled visit at any time during the study. An unscheduled visit may be prompted by reactogenicity issues, illness visit criteria for COVID-19, or new or ongoing AEs. The study site also has the discretion to make reminder telephone calls or send text messages to inform the participant about visits, review eDiary requirements, or follow-up on ongoing or outstanding issues.

In accordance with "FDA Guidance on Conduct of Clinical Trials of Medical Products during COVID-19 Public Health Emergency" (DHHS 2020), investigators may convert study site visits to home visits or telemedicine visits with the approval of the Sponsor. Such action should be taken to protect the safety and well-being of study participants and study site staff or to comply with state or municipal mandates.

General considerations for study assessments and procedures include the following:

- Protocol waivers or exemptions are not allowed. The study procedures and their timing must be followed as presented in Table 7 and Table 9. Adherence to the study design requirements is essential and required for study conduct.
- Immediate safety concerns should be discussed with the Sponsor immediately upon occurrence or awareness to determine if the participant should continue study treatment or participation in the study.
- All screening evaluations must be completed and reviewed to confirm that potential participants meet all eligibility criteria. The investigator will maintain a screening log to record details of all participants screened and to confirm eligibility or record reasons for screening failure, as applicable.
- Procedures conducted as part of the participant's routine clinical management (eg, blood count) and obtained before signing of the ICF may be utilized for screening or baseline

assessments provided the procedures met the protocol-specified criteria and were performed within the time frame defined in the SoA.

#### 7.1. Safety Assessments and Procedures

In Part A (blinded), Part B (open-label), and Part C (open-label), safety assessments will include monitoring and recording of the following for each participant, according to the SoA (Table 7, Table 9, and Table 10):

- Solicited local and systemic ARs (Section 7.5.3) that occur during the 7 days following each injection (ie, the day of injection and 6 subsequent days) in **Part A and Part C**. Solicited ARs will be recorded daily using eDiaries (Section 7.1.1).
- Unsolicited AEs observed or reported during the 28 days following each injection (ie, the day of injection and 27 subsequent days) in Parts A and C. Unsolicited AEs are defined in Section 7.5.1.
- AEs leading to discontinuation from dosing and/or withdrawal from study participation from Day 1 through the last day of study participation (ie, Day 394 per Part A SoA, OL-Day 178 in Part B, and BD-Day 361 in Part C).
- MAAEs from first dose on Day 1 through the entire study period (ie, Day 394 per Part A SoA, OL-Day 178 in Part B, and BD-Day 361 in Part C).
- SAEs from first dose on Day 1 through the entire study period (ie, Day 394 per Part A SoA, OL-Day 178 in Part B, and BD-Day 361 in Part C).
- AESIs through the entire study period (ie, Day 394 per Part A SoA, OL-Day 178 in Part B, and BD-Day 361 in Part C).
- Vital sign measurements (Section 7.1.4)
- Physical examination findings (Section 7.1.5).
- Assessments for SARS-CoV-2 infection from Day 1 through study completion (Section 7.1.6).
- Details of all pregnancies in female participants will be collected after the start of study treatment and until the end of their participation in the study (Section 7.5.6).

##### 7.1.1. Use of Electronic Diaries

At the time of consent/assent, participants or their caregivers must confirm they will be willing to complete an eDiary using either an application downloaded to their smartphone or using a device that is provided at the time of enrollment. Before enrollment on Day 1, participants or their caregivers will be instructed to download the eDiary application or will be provided an eDiary device to record solicited ARs (Section 7.5.3) on Day 1. Participants who were originally randomized to placebo and who opt to receive mRNA-1273 will enter the Open-label Observational Phase. They will receive Dose 1 at D209 (OL-Day 1), return for their second dose at OL-Day 29, and return again at OL-Day 57. Participants who entered the Open-label phase, may opt to receive a BD in Part C, and will follow Part C SoA until study completion. During the open-label period Part B, the eDiary will not be used. During the open-label period Part C, the eDiary will be used to record solicited ARs.

In Part A, the blinded phase of the study, at each injection visit, participants or their caregivers will be instructed (Day 1) or reminded (Day 29) on thermometer usage to measure body temperature, ruler usage to measure injection site erythema and swelling/induration (hardness), and self-assessment for localized axillary swelling or tenderness on the same side as the injection arm.

In Part A, the blinded phase of the study, at each injection visit, participants or their caregivers will record data into the eDiary starting approximately 30 minutes after injection under supervision of the study site staff to ensure successful entry of assessments. The study site staff will perform any retraining as necessary. Participants or their caregivers will continue to record data in the eDiary after they leave the study site, preferably in the evening and at the same time each day, on the day of injection and for 6 days following injection.

Participants or their caregivers will record the following data in the eDiary:

- Solicited local and systemic reactogenicity ARs, as defined in Section 7.5.3, that occur on the day of each vaccine administration and during the 7 days after vaccine administration (ie, the day of injection and 6 subsequent days). If a solicited local or systemic AR continues beyond Day 7 after vaccination, the participant will be prompted to capture details of the solicited local or systemic AR in the eDiary until it is resolves or the next IP injection occurs, whichever occurs first; capture of details of ARs in the eDiary should not exceed 28 days after each vaccination and not to exceed 28 days after each vaccination, whichever occurs first. Adverse reactions recorded in the eDiary beyond Day 7 should be reviewed by the study site staff either during the next scheduled telephone call or at the next study site visit.
- Daily oral body temperature measurement should be performed at approximately the same time each day using the thermometer provided by the study site. If body temperature is taken more than once in a given day, only the highest temperature reading should be recorded.

- Measurement, as applicable, for solicited local ARs (injection site erythema and swelling/induration); the size measurements will be performed using the ruler provided by the study site.
- Any medications taken to treat or prevent pain or fever on a day of injection or for the next 6 days.

The eDiary will be the only source documents allowed for solicited systemic or local ARs (including body temperature measurements). Participants or their caregivers will be instructed to complete eDiary entries daily. The participant or their caregiver will have a limited window on the following day to complete assessments for the previous day; quantitative temperature recordings and measurement of any injection site erythema or swelling/induration reported on the following day may be excluded from the analyses of solicited ARs.

Any new safety information reported during safety telephone calls or at study site visits (including a solicited reaction) that is not already captured in the eDiary will be described in the source documents as a verbally reported event. An event reported in this manner must be described as a solicited event and entered on the solicited AR eCRF.

Study site staff will review eDiary data with participants at visits 7 days after each injection.

The eDiary will also be used every 4 weeks, starting at Day 71 through Day 183 and again starting at Day 223 through Day 363 for participants in Part A (per Figure 5), to capture the occurrence of AEs, MAAEs, SAEs, AESI, or AEs leading to withdrawal. As specified in the applicable SoA (Table 7), the eDiary will prompt the participant to complete an eDiary questionnaire that collects the following data:

- Changes in health since last completing the questionnaire or since in contact with the study site
- Known exposure to someone with known COVID-19 or SARS-CoV-2 infection
- Any experience of symptoms of COVID-19
- Any MAAEs or SAEs

If an eDiary record results in identification of relevant safety events according to the study period, or of symptoms of COVID-19, a follow-up safety telephone call will be triggered.

Completion of eDiary questionnaires will alternate with safety telephone calls (Section 7.1.2) as the procedure for safety follow-up approximately every 4 weeks starting at Day 85 through Day 197 and again starting at Day 237 through Day 377 (Part A SoA, Table 7).

In Part C, diary entries will be recorded by the participant at approximately 30 minutes after BD injection while at the study site with instruction provided by study staff. Study participants will continue to record entries in the eDiary each day after they leave the study site, preferably in the evening, on the day of injection and for 6 days following injection. If a solicited local or systemic AR continues beyond Day 7 after vaccination, the participant will be prompted to capture details of the solicited local or systemic AR in the eDiary until the AR resolves; capture of details of ARs in the eDiary should not exceed 28 days after vaccination. Adverse reactions recorded in eDiaries beyond Day 7 should be reviewed either via telephone call or at the following study visit. Review of eDiary will occur on BD-Day 8.

###### 7.1.1.1. Ancillary Supplies for Participant Use

Study sites will distribute Sponsor-provided oral thermometers and rulers for use by participants in assessing body temperature and injection site reactions for recording solicited ARs in eDiaries. Based on availability, smartphone devices may be provided to those participants who do not have their own device to use for eDiary activities.

##### 7.1.2. Safety Telephone Calls

A safety telephone call is a telephone call made to the participant by trained study site personnel. This call will follow a script, which will facilitate the collection of relevant safety information.

Safety telephone calls follow a schedule for each participant as indicated in the Part A SoA (Table 7), Part B (Table 9), and Part C (Table 10).

The participant will be interviewed according to the script about occurrence of AEs, MAAEs, SAEs, AESI, AEs leading to study withdrawal, concomitant medications associated with those events, and any nonstudy vaccinations (Section 7.5.7). In addition, study personnel will collect information on known participant exposure to someone with known COVID-19 or SARS-CoV-2 infection and on participant experience of COVID-19 symptoms. All safety information collected from the telephone contact must be documented in source documents as described by the participant and not documented on the script used for the safety telephone contact. As noted in Section 7.1.1, an unscheduled follow-up safety telephone call may be triggered if an eDiary record results in identification of a relevant safety event.

##### 7.1.3. Safety Laboratory Assessments

No scheduled laboratory assessments for safety are planned. This is based on the absence of clinically significant abnormal laboratory findings in the Phase 1 and Phase 2 studies of mRNA-1273 in adults.

A point-of-care urine pregnancy test will be performed at the Screening Visit (Day 0) and before each vaccine dose. At any time, a pregnancy test either via blood or point-of-care urine can be performed, at the discretion of the investigator.

##### 7.1.4. Vital Sign Measurements

Vital sign measurements will include systolic and diastolic blood pressure, heart rate, respiratory rate, and body temperature (preferred route is oral). The participant will be seated for at least 5 minutes before all measurements are taken. Vital signs will be measured at the time points indicated in the SoAs (Table 7, Table 9, and Table 10). At dosing visits, vital sign measurements will be collected once before injection and at least 30 minutes post injection (before participants are discharged from the study site).

Febrile participants at dosing visits (fever is defined as a body temperature ≥ 38.0°C/≥ 100.4°F) may have visits rescheduled within the relevant study visit windows. Afebrile participants with minor illnesses may be injected at the discretion of the investigator.

When procedures overlap and are scheduled to occur at the same time point, the order of procedures should be vital sign measurements and then the blood collection.

##### 7.1.5. Physical Examinations

A full physical examination, including height and weight, will be performed at scheduled time points as indicated in the Part A, Part B, and Part C SoAs (Table 7, Table 9, and Table 10, respectively). The full examination will include assessment of skin, head, ears, eyes, nose, throat, neck, thyroid, lungs, heart, cardiovascular system, abdomen, lymph nodes, and musculoskeletal system/extremities. Any clinically significant finding identified during a study visit should be reported as an MAAE.

Symptom-directed physical examinations may be performed at other time points at the discretion of the investigator. On each injection day before injection and again 7 days after injection, the arm receiving the injection should be examined and the associated lymph nodes should be evaluated. Any clinically significant finding identified during a study visit should be reported as an MAAE.

Body mass index will be calculated at the Screening Visit (Day 0) only.

##### 7.1.6. Assessment for SARS-CoV-2 Infection

Study participants will have NP or nasal swab samples collected for SARS-CoV-2 testing at time points specified in the SoA (Part A, Table 7; Part B, Table 9; and Part C, Table 10).

For Part A, Part B, and Part C, a study illness visit or a consultation will be arranged within 72 hours or as soon as possible to collect an NP or nasal swab sample to ascertain the presence of SARS-CoV-2 via RT-PCR if a participant experiences any of the following:

- Signs or symptoms of SARS-CoV-2 infection as defined by the CDC (CDC 2020b)
- Exposure to an individual confirmed to be infected with SARS-CoV-2

- MAAE suggesting a SARS-CoV-2 infection

If the participant had known exposure to COVID-19 (eg, exposure to someone with confirmed COVID-19), it will be captured in the COVID-19 exposure form.

If scheduled, the study illness visit may collect additional clinical information, including assessments such as medical history, physical examination, blood sampling for clinical laboratory testing, and nasal, saliva, and/or NP swab sampling for viral PCR (including multiplex PCR for respiratory viruses including SARS-CoV-2) to evaluate the severity of the clinical case. Radiologic imaging studies may be conducted. All findings will be recorded in the eCRF.

If participants are confirmed to have SARS-CoV-2 infection, the investigator will notify the participant, and the participant's primary care physician, of the diagnosis. If the study participant does not have a primary care physician, the investigator will assist them to obtain one. The participant will also be instructed on infection prevention measures consistent with local public health guidance.

Any confirmed symptomatic SARS-CoV-2 infection occurring in participants will be captured as an MAAE along with relevant concomitant medications and details about severity, seriousness, and outcome. Additionally, a convalescent visit will be scheduled approximately 28 days (+7 days) after diagnosis (Section 7.4.3). At this visit, an NP or nasal swab sampling for viral PCR and a blood sample will be collected for potential immunologic assessment of SARS-CoV-2 infection.

#### 7.2. Immunogenicity Assessments

Blood samples for immunogenicity assessments will be collected at the time points indicated in the SoA (Part A, Table 7; Part B, Table 9; and Part C, Table 10): The following analytes will be measured:

- Serum nAb level against SARS-CoV-2 as measured by pseudovirus and/or live virus neutralization assays
- Serum bAb levels against SARS-CoV-2 as measured by ligand-binding assay specific to the SARS-CoV-2 S protein
- Serum bAb levels against SARS-CoV-2 nucleocapsid protein as measured by ligand-binding assay specific to the SARS-CoV-2 nucleocapsid protein

Serum collected from all participants will be tested for bAb against SARS-CoV-2 nucleocapsid protein at specified time points. In addition, serum samples from a selected subset of study participants who received mRNA-1273 will be selected for testing of nAb and bAb against the SARS-CoV-2 S protein. Sample aliquots will be designed to ensure that backup samples are available and that vial volumes are likely to be adequate for future testing needs. The actual time and date of each sample collected will be recorded in the eCRF, and unique sample identification will be utilized to maintain the blind at the laboratory at all times and to allow for automated sample tracking and housing. Handling and preparation of the samples for analysis, as well as shipping and storage requirements, will be provided in a separate study manual.

Measurement of bAb and nAb levels will be performed in a laboratory designated by the Sponsor.

According to the ICF (Section 10.2.6), serum from immunogenicity testing may be used for future research, which may be performed at the discretion of the Sponsor to further characterize the immune response to SARS-CoV-2, additional assay development, and the immune response across CoVs.

The maximum planned volume of blood sampled per participant for immunogenicity assessments in 1 day is 21 mL.

#### 7.3. Exploratory Assessments and Biomarkers

Exploratory assessments may include assessment of biomarkers for safety, reactogenicity, inflammatory and cardiac function. Serologic markers of disease severity, immune response to SARS-CoV-2, RT-PCR of NP or nasal swab or saliva samples, and genetic sequences of SARS-CoV-2 strains isolated from participants' samples may also be measured.

#### 7.4. Efficacy Assessments

##### 7.4.1. Vaccine Effectiveness Assessments

Vaccine effectiveness for adolescents of ages of 12 to < 18 years will be inferred based on serum Ab responses obtained on Day 57 (28 days after the second injection of mRNA-1273). Inference will be based on assessing the adolescent Ab responses against the following:

1. *If available at the time of analysis*, adolescent Ab responses will be assessed against an accepted serum nAb threshold conferring protection against COVID-19.
2. *If an accepted threshold of protection is not available*, adolescent Ab responses will be assessed by establishing noninferiority of the GM value and seroresponse rate of serum nAb from adolescent participants compared with those from young adults enrolled in the ongoing clinical endpoint efficacy trial (Study P301). The statistical parameters to infer effectiveness are described in Section 2.

**COVID-19:**

To be considered as a case of COVID-19 for the evaluation of the primary efficacy endpoint, the following case definition must be met:

- The participant must have experienced at least TWO of the following systemic symptoms: Fever (≥ 38ºC/≥ 100.4ºF), chills, myalgia, headache, sore throat, new olfactory and taste disorder(s), OR
- The participant must have experienced at least ONE of the following respiratory signs/symptoms: cough, shortness of breath or difficulty breathing, OR clinical or radiographical evidence of pneumonia; AND
- The participant must have at least 1 NP or nasal swab or saliva sample (or respiratory sample, if hospitalized) positive for SARS-CoV-2 by RT-PCR.

**Severe COVID-19:**

To be considered severe COVID-19, the following criteria must be met:

- Confirmed COVID-19 as per the primary efficacy endpoint case definition, plus any of the following:
  - Clinical signs indicative of severe systemic illness, respiratory rates ≥ 30 per minute, heart rate ≥ 125 beats per minute, SpO2 ≤ 93% on room air at sea level or PaO2/FIO2 < 300 mm Hg, OR
  - Respiratory failure or Acute Respiratory Distress Syndrome, (defined as needing high-flow oxygen, noninvasive or mechanical ventilation, or extracorporeal membrane oxygenation [ECMO]), evidence of shock (systolic blood pressure < 90 mmHg, diastolic BP < 60 mmHg or requiring vasopressors), OR
  - Significant acute renal, hepatic, or neurologic dysfunction, OR
  - Admission to an ICU or death.

The secondary case definition of COVID-19 is defined as the following systemic symptoms: fever (temperature > 38ºC/≥ 100.4ºF), or chills, cough, shortness of breath or difficulty breathing, fatigue, muscle aches, or body aches, headache, new loss of taste or smell, sore throat, congestion or runny nose, nausea, or vomiting or diarrhea AND a positive NP or nasal swab or saliva sample for SARS-CoV-2 by RT-PCR.

Death attributed to COVID-19 is defined as any participant who dies during the study with a cause directly attributed to a complication of COVID-19.

**SARS-CoV-2 Infection:**

- SARS-CoV-2 infection is defined in participants with SARS-CoV-2 negative at baseline:
  - bAb level against SARS-CoV-2 nucleocapsid protein negative at Day 1, which becomes positive post baseline, OR

  – Post-baseline Positive RT-PCR

##### 7.4.2. Surveillance for COVID-19 Symptoms

Surveillance for COVID-19 symptoms will be conducted via biweekly telephone calls or eDiary prompts as specified in Section 7.1.1 and Figure 4; starting after participant enrollment and throughout the study.

If there is no response to an eDiary prompt for 3 days, the study site staff will contact the study participant by phone.

According to the CDC as of 10 Jun 2020 (CDC 2020c), patients with COVID-19 have reported a wide range of symptoms ranging from mild symptoms to severe illness. Throughout the study, to survey for COVID-19, the following prespecified symptoms that meet the criteria for suspicion of COVID-19 will be elicited weekly from the participant and the presence of any one of these symptoms lasting at least 48 hours (except for fever and/or respiratory symptoms) will result in the study site staff arranging an illness visit to collect an NP or nasal swab within 72 hours:

- Fever (temperature ≥ 38ºC/≥ 100.4ºF) or chills (of any duration, including ≤ 48 hours)
- Shortness of breath or difficulty breathing (of any duration, including ≤ 48 hours)
- Cough (of any duration, including ≤ 48 hours)
- Fatigue
- Muscle or body aches
- Headache
- New loss of taste or smell
- Sore throat
- Congestion or runny nose
- Nausea or vomiting
- Diarrhea

**Figure 4: Surveillance for COVID-19 Symptoms and the Corresponding Clinical Data Pathways**

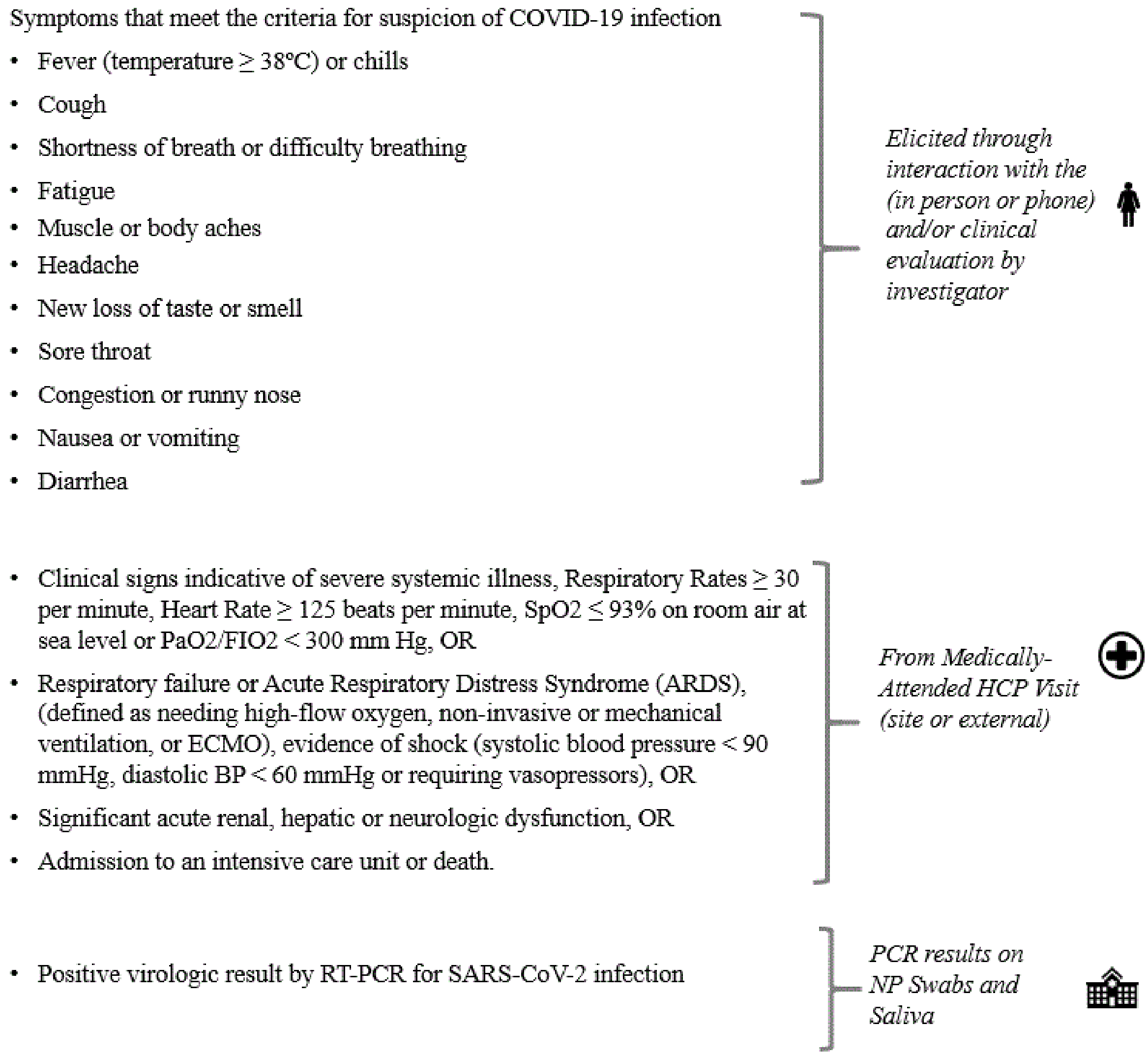

Abbreviations: BP = blood pressure, COVID-19 = coronavirus disease 2019, ECMO, SpO2 = oxygen saturation, PaO2 = partial pressure of oxygen, FIO2 = fraction of inspired oxygen, HCP = healthcare practitioner, RT-PCR = reverse transcriptase polymerase chain reaction, SARS-CoV-2 = Severe Acute Respiratory Syndrome coronavirus 2.

It is important to note that some of the symptoms of COVID-19 overlap with solicited systemic ARs that are expected after vaccination with mRNA-1273 (eg, myalgia, headache, fever, and chills). During the first 7 days after vaccination, when these solicited ARs are common, investigators should use their clinical judgment to decide if an NP or nasal swab should be collected. The collection of an NP or nasal swab prior to the Day 1 and Day 29 vaccination can help ensure that cases of COVID-19 are not overlooked. Any study participant who reports respiratory symptoms during the 7-day period after vaccination should be evaluated for COVID-19.

During the course of the study, participants with symptoms of COVID-19 will be asked to return within 72 hours or as soon as possible to the study site or medically qualified staff from the study site will conduct a home visit as soon as possible to collect an NP or nasal swab sample (for RT-PCR) for evaluation of COVID-19. Both study site visits and home visits are referred to as illness visits (Section 7.1.6). The NP or nasal swab sample will also be tested for the presence of other respiratory infections. Additionally, a convalescent visit will be scheduled approximately 28 days (+7 days) after diagnosis (Section 7.4.3). At this visit, an NP or nasal swab sampling for viral PCR and a blood sample will be collected for potential immunologic assessment of SARS-CoV-2 infection. In addition, the study site may collect an additional respiratory sample for SARS-CoV-2 testing to be able to render appropriate medical care for the study participant as determined by local standards of care.

Cases are defined as participants meeting clinical criteria based both on symptoms for COVID-19 and on RT-PCR detection of SARS-CoV-2 from samples collected within 72 hours of the study participant reporting symptoms meeting the definition of COVID-19. Participants who are hospitalized for COVID-19 without the opportunity for a clinic or home visit will also be considered cases, assuming that the symptomology criteria for COVID-19 are met and a respiratory sample is positive for SARS-CoV-2 by PCR at a clinical laboratory improvement amendments-certified laboratory. Investigators are encouraged to try to obtain a respiratory sample during the course of hospitalization for submission to the study central laboratory, if feasible. The investigator should determine if the criteria for severe COVID-19 has been met.

Severe COVID-19 is defined in Section 7.4.1.

All clinical findings will be recorded in the eCRF. All confirmed cases of COVID-19 will be captured as MAAEs, along with relevant concomitant medications and details about severity, seriousness, and outcome, and will be reported immediately to the Sponsor or designee (Section 7.5.4).

##### 7.4.3. Follow up/Convalescent Period After Diagnosis with COVID-19

Any confirmed COVID-19 occurring in a participant will be captured as an MAAE along with relevant concomitant medications and details about severity, seriousness, and outcome. Study participants will be monitored by medically qualified study site personnel for a 28-day period after diagnosis. Additionally, a convalescent visit will be scheduled approximately 28 days (+7 days) after diagnosis. At this visit, an NP or nasal swab sampling for viral PCR and a blood sample will be collected for potential immunologic assessment of SARS-CoV-2 infection. The investigator should determine if the criteria for severe COVID-19 has been met. If the participant is hospitalized, medically qualified study site personnel will try to obtain medical records and SARS-CoV-2 diagnostic results. If the participant is later discharged from the hospital during the

28-day period following diagnosis of COVID-19, the study site personnel will arrange for a resumption of the protocol schedule.

#### 7.5. Safety Definitions and Related Procedures

##### 7.5.1. Adverse Event

An AE is defined as any untoward medical occurrence associated with the use of a drug in humans, whether or not considered drug related.

**Events Meeting the Adverse Event Definition**

- Exacerbation of a chronic or intermittent pre-existing condition including either an increase in frequency and/or intensity of the condition
- New conditions detected or diagnosed after the first dose of IP even though they may have been present before the start of the study

**Events NOT Meeting the Adverse Event Definition**

- Medical or surgical procedure (eg, endoscopy, appendectomy): the condition that leads to the procedure should be the AE.
- Situations in which an untoward medical occurrence did not occur (social and/or convenience admission to a hospital).

An AR is any AE for which there is a reasonable possibility that the vaccine caused the AE (Section 7.5.9). For the purposes of investigational new drug safety reporting, “reasonable possibility” means that there is evidence to suggest a causal relationship between the vaccine and the AE.

An unsolicited AE is any AE reported by the participant that is not specified as a solicited AR in the protocol or is specified as a solicited AR but starts outside the protocol-defined period for reporting solicited ARs (ie, for the 7 days after each dose of vaccine).

##### 7.5.2. Serious Adverse Events

An AE (including an AR) is considered an SAE if, in the view of either the investigator or Sponsor, it results in any of the following outcomes:

- **Death**
  A death that occurs during the study or that comes to the attention of the investigator during the protocol-defined follow-up period must be reported to the Sponsor, whether or not it is considered related to the IP.

- **Is life-threatening**
  An AE is considered life-threatening if, in the view of either the investigator or the Sponsor, its occurrence places the participant at immediate risk of death. It does not include an AE that, had it occurred in a more severe form, might have caused death.
- **Inpatient hospitalization or prolongation of existing hospitalization**
  In general, inpatient hospitalization indicates the participant was admitted to the hospital or emergency ward for at least one overnight stay for observation and/or treatment that would not have been appropriate in the physician's office or outpatient setting. The hospital or emergency ward admission should be considered an SAE regardless of whether opinions differ as to the necessity of the admission. Complications that occur during inpatient hospitalization will be recorded as an AE; however, if a complication/AE prolongs hospitalization or otherwise fulfills SAE criteria, the complication/AE will be recorded as a separate SAE.
- **Persistent or significant incapacity or substantial disruption of the ability to conduct normal life functions**
  This definition is not intended to include experiences of relatively minor medical significance such as uncomplicated headache, nausea/vomiting, diarrhea, influenza, and accidental trauma (eg, sprained ankle), which may interfere with or prevent everyday life functions but do not constitute a substantial disruption.
- **Congenital anomaly or birth defect**
- **Medically important event**
  Medical judgment should be exercised in deciding whether SAE reporting is appropriate in other situations, such as important medical events that may not be immediately life-threatening or result in death or hospitalization but may jeopardize the participant or require medical or surgical intervention to prevent one of the other outcomes listed in the above definition. These events should usually be considered serious. Examples of such medical events include allergic bronchospasm requiring intensive treatment in an emergency room or at home, blood dyscrasias or convulsions that do not result in inpatient hospitalization, or the development of drug dependency or drug abuse.

##### 7.5.3. Solicited Adverse Reactions

The term "reactogenicity" refers to the occurrence and intensity of selected signs and symptoms (ARs) occurring after IP injection. The eDiary will solicit daily participant reporting of ARs using a structured checklist (Section 7.1.1). Participants will record such occurrences in an eDiary on the day of each dose injection and for the 6 days after the day of dosing (Part A and Part C only).

Severity grading of reactogenicity will occur automatically based on participant entry into the eDiary according to the grading scales presented in Table 3 modified from the Toxicity Grading Scale for Healthy Adult and Adolescent Volunteers Enrolled in Preventative Vaccine Clinical Trials (DHHS 2007).

If a solicited local or systemic AR continues beyond Day 7 after vaccination, the participant will be prompted to capture details of the solicited local or systemic AR in the eDiary until it resolves or the next IP injection occurs whichever occurs first; capture of details of ARs in the eDiary should not exceed 28 days after each vaccination. Adverse reactions recorded in the eDiary beyond Day 7 should be reviewed by the study site staff either during the next scheduled telephone call or at the next study site visit. All solicited ARs (local and systemic) will be considered causally related to dosing.

**Table 3: Solicited Adverse Reactions and Grades**

| Reaction | Grade 0 | Grade 1 | Grade 2 | Grade 3 | Grade 4[a] |
|---|---|---|---|---|---|
| Injection site pain | None | Does not interfere with activity | Repeated use of over-the-counter pain reliever > 24 hours or interferes with activity | Any use of prescription pain reliever or prevents daily activity | Requires emergency room visit or hospitalization |
| Injection site erythema (redness) | < 25 mm/ < 2.5 cm | 25 - 50 mm/ 2.5 - 5 cm | 51 - 100 mm/ 5.1 - 10 cm | > 100 mm/ > 10 cm | Necrosis or exfoliative dermatitis |
| Injection site swelling/induration (hardness) | < 25 mm/ < 2.5 cm | 25 - 50 mm/ 2.5 - 5 cm | 51 - 100 mm/ 5.1 - 10 cm | > 100 mm/ > 10 cm | Necrosis |
| Axillary (underarm) swelling or tenderness ipsilateral to the side of injection | None | No interference with activity | Repeated use of over-the-counter (non-narcotic) pain reliever > 24 hours or some interference with activity | Any use of prescription (narcotic) pain reliever or prevents daily activity | Emergency room visit or hospitalization |
| Headache | None | No interference with activity | Repeated use of over-the-counter pain reliever > 24 hours or some interference with activity | Significant; any use of prescription pain reliever or prevents daily activity | Requires emergency room visit or hospitalization |
| Fatigue | None | No interference with activity | Some interference with activity | Significant; prevents daily activity | Requires emergency room visit or hospitalization |
| Myalgia (muscle aches all over body) | None | No interference with activity | Some interference with activity | Significant; prevents daily activity | Requires emergency room visit or hospitalization |
| Arthralgia (joint aches in several joints) | None | No interference with activity | Some interference with activity | Significant; prevents daily activity | Requires emergency room visit or hospitalization |
| Nausea/vomiting | None | No interference with activity or 1-2 episodes/ 24 hours | Some interference with activity or > 2 episodes/24 hours | Prevents daily activity, requires outpatient intravenous hydration | Requires emergency room visit or hospitalization for hypotensive shock |

| Reaction | Grade 0 | Grade 1 | Grade 2 | Grade 3 | Grade 4[a] |
|---|---|---|---|---|---|
| Chills | None | No interference with activity | Some interference with activity not requiring medical intervention | Prevents daily activity and requires medical intervention | Requires emergency room visit or hospitalization |
| Fever (oral) | < 38.0°C<br>< 100.4°F | 38.0 – 38.4°C<br>100.4 - 101.1°F | 38.5 – 38.9°C<br>101.2 – 102.0°F | 39.0 – 40.0°C<br>102.1 - 104.0°F | > 40.0°C<br>> 104.0°F |

a. Grading for Grade 4 events per investigator assessment (with exception of fever).

Source: Guidance for industry - Toxicity Grading Scale for Healthy Adult and Adolescent Volunteers Enrolled in Preventative Vaccine Clinical Trials (DHHS 2007).

Any solicited AR that meets any of the following criteria must be entered into the participant's source document and must also be recorded by the study site staff on the solicited AR page of the participant's eCRF:

- Solicited local or systemic AR that results in a visit to a healthcare practitioner (HCP; otherwise meets the definition of an MAAE)
- Solicited local or systemic AR leading to the participant withdrawing from the study or the participant being withdrawn from the study by the investigator (AE leading to withdrawal)
- Solicited local or systemic AR lasting beyond 7 days after injection
- Solicited local or systemic AR that leads to participant withdrawal from IP
- Solicited local or systemic AR that otherwise meets the definition of an SAE

##### 7.5.4. Medically Attended Adverse Events

An MAAE is an AE that leads to an unscheduled visit to an HCP. This would include visits to a study site for unscheduled assessments (eg, abnormal laboratory test results follow-up, COVID-19 (Section 7.4.1) and visits to HCPs external to the study site (eg, urgent care, primary care physician). Investigators will review unsolicited AEs for the occurrence of any MAAE. Unsolicited AEs will be captured on the AE page of the eCRF.

All confirmed COVID-19 cases (Section 7.4.1) will be recorded as MAAEs and reported to the Sponsor or designee immediately and in all circumstances within 24 hours, using the SAE Mailbox, the SAE Hotline, or the SAE Fax line (Section 7.5.11). The investigator will submit any updated COVID-19 case data to the Sponsor within 24 hours of it being available.

###### 7.5.4.1. Anaphylaxis

All suspected cases of anaphylaxis should be recorded as MAAEs and reported as SAEs (Section 7.5.2) based on criteria for a medically important event, unless the event meets other serious criteria. As an SAE, the event should be reported to the Sponsor or designee immediately and in all circumstances within 24 hours as per Section 7.5.10 (Reporting SAEs). The investigator will submit any updated anaphylaxis case data to the Sponsor within 24 hours of it being available. For reporting purposes, a participant who displays signs/symptoms consistent with anaphylaxis as shown below should be reported as a potential case of anaphylaxis. This is provided as general guidance for Investigators and is based on the Brighton Collaboration case definition (Rüggeberg et al 2007).

Anaphylaxis is an acute hypersensitivity reaction with multi-organ-system involvement that can present as or rapidly progress to a severe life-threatening reaction. It may occur following exposure to allergens from a variety of sources. Anaphylaxis is a clinical syndrome characterized by:

- Sudden onset AND
- Rapid progression of signs and symptoms AND
- Involving two or more organ systems, as follows:
  - **Skin/mucosal**: urticaria (hives), generalized erythema, angioedema, generalized pruritus with skin rash, generalized prickle sensation, red and itchy eyes
  - **Cardiovascular**: measured hypotension, clinical diagnosis of uncompensated shock, loss of consciousness or decreased level of consciousness, evidence of reduced peripheral circulation
  - **Respiratory**: bilateral wheeze (bronchospasm), difficulty breathing, stridor, upper airway swelling (lip, tongue, throat, uvula, or larynx), respiratory distress, persistent dry cough, hoarse voice, sensation of throat closure, sneezing, rhinorrhea
  - **Gastrointestinal**: diarrhea, abdominal pain, nausea, vomiting

##### 7.5.5. Adverse Events of Special Interest

An AESI is an AE (serious or nonserious) of scientific and medical concern specific to the Sponsor's product or program, for which ongoing monitoring and immediate notification by the Investigator to the Sponsor is required and documentation is in the form of a case narrative. Such events may require further investigation to characterize and understand them. Refer to Section 10.4, Appendix 4 for a list of AESIs pertinent to this study. All AESIs will be collected throughout the entire study period and must be reported to the Sponsor or designee immediately and in all circumstances within 24 hours of becoming aware of the event via the EDC system. If a site receives a report of a new AESI from a study participant or receives updated data on a previously reported AESI and the eCRF has been taken offline, then the site can report this information on a paper AESI form using the SAE Mailbox, the SAE Hotline, or the SAE Fax line (Section 7.5.11).

**Acute Myocarditis and/or Pericarditis**

These definitions are intended to serve as a guide to help in the reporting of suspected cases of myocarditis and/or pericarditis; however, the diagnosis of suspected cases is left to the investigator's clinical judgement.

All suspected cases of probable and confirmed myocarditis, pericarditis, or myopericarditis should be recorded as an AESI,_and reported as an SAE, if the event meets seriousness criteria. As an SAE, the event should be reported to the Sponsor or designee immediately and in all circumstances within 24 hours as per Section 7.5.12. The investigator will submit any updated myocarditis, pericarditis, or myopericarditis case data to the Sponsor within 24 hours of it being available. For reporting purposes, a participant who displays signs/symptoms consistent with the CDC case definitions as described below (Gargano et al 2021), should be reported as a potential case of confirmed or probable myocarditis, pericarditis, or myopericarditis.

**Acute Myocarditis Case Definition**

Presence of ≥ 1 new or worsening of the following clinical symptoms (persons who lack the listed symptoms but who meet other criteria may be classified as subclinical myocarditis [probable or confirmed]):

- Chest pain/pressure/discomfort
- Dyspnea/shortness of breath/pain with breathing
- Palpitations
- Syncope

OR

Infants and children aged < 12 years might instead have ≥ 2 of the following symptoms:

- Irritability
- Vomiting
- Poor feeding
- Tachypnea
- Lethargy

AND

For PROBABLE CASE:

Presence of ≥ 1 new finding of the following:

- Troponin level above upper limit of normal (any type of troponin)
- Abnormal electrocardiogram (ECG or EKG) or rhythm monitoring findings consistent with myocarditis
  - To meet the ECG or rhythm monitoring criterion, a probable case must include at least one of the following:
    - ST segment or T-wave abnormalities
    - Paroxysmal or sustained atrial, supraventricular, or ventricular arrhythmias
    - AV nodal conduction delays or intraventricular conduction defects
- Abnormal cardiac function or wall motion abnormalities on echocardiogram
- Cardiac magnetic resonance imaging (cMRI) finding consistent with myocarditis (Ferreira et al 2018)

AND

- No other identifiable cause of the symptoms and findings

For CONFIRMED CASE:

- Histopathologic confirmation of myocarditis (using Dallas criteria [Aretz et al 1987])

OR

- cMRI findings consistent with myocarditis in the presence of troponin level above upper limit of normal (any type of troponin)

AND

- No other identifiable cause of the symptoms and findings

**Acute Pericarditis Case Definition**

Presence of ≥ 2 new or worsening of the following clinical features (Adler et al 2015):

- Acute chest pain (Typically described as pain made worse by lying down, deep inspiration, or cough; and relieved by sitting up or leaning forward, although other types of chest pain may occur)
- Pericardial rub on examination

- New ST-elevation or PR-depression on EKG
- New or worsening pericardial effusion on echocardiogram or magnetic resonance imaging

**Myopericarditis Case Definition**

Participants who meet criteria for both myocarditis and pericarditis may be described under myopericarditis.

**MIS-C Case Definition**

Investigators will also be asked to report, as AESI, clinical signs/symptoms consistent with the CDC case definition of MIS-C (https://emergency.cdc.gov/han/2020/han00432.asp):

- An individual aged < 21 years presenting with fever, laboratory evidence of inflammation, and evidence of clinically severe illness requiring hospitalization, with multisystem (> 2) organ involvement (cardiac, renal, respiratory, hematologic, gastrointestinal, dermatologic or neurological);

AND

- No alternative plausible diagnoses;

AND

- Positive for current or recent SARS-CoV-2 infection by RT-PCR, serology, or antigen test; or COVID-19 exposure within the 4 weeks prior to the onset of symptoms:
  1. Fever ≥ 38.0°C/≥ 100.4ºF for ≥ 24 hours, or report of subjective fever lasting ≥ 24 hours
  2. Including, but not limited to, one or more of the following: an elevated C-reactive protein (CRP), erythrocyte sedimentation rate (ESR), fibrinogen, procalcitonin, d-dimer, ferritin, lactic acid dehydrogenase (LDH), or interleukin 6 (IL-6), elevated neutrophils, reduced lymphocytes, and low albumin

Some individuals may fulfill full or partial criteria for Kawasaki disease but should be reported if they meet the case definition for MIS-C. Consider MIS-C in any pediatric death with evidence of SARS-CoV-2 infection.

All cases of MIS-C will be reported to the Sponsor or designee immediately and in all circumstances within 24 hours, using the SAE Mailbox, the SAE Hotline, or the SAE Fax line (Section 7.5.11).

##### 7.5.6. Recording and Follow-up of Pregnancy

Female individuals who have a positive pregnancy test at Screening should not be enrolled; participants who have a positive pregnancy test any time during the study should receive no further dosing with IP but should be asked to remain in the study and be monitored for safety.

Details of all pregnancies in female participants will be collected after the start of study treatment and until the end of their participation in the study.

- If a pregnancy is reported, the investigator should inform the Sponsor within 24 hours of learning of the pregnancy and should follow the procedures outlined in this section.
- Abnormal pregnancy outcomes (eg, spontaneous abortion, fetal death, stillbirth, congenital anomalies, ectopic pregnancy) are considered SAEs.

Pregnancies occurring in participants after enrollment must be reported to Sponsor or designee within 24 hours of the study site learning of its occurrence, using the SAE Mailbox, the SAE Hotline, or the SAE Fax line (Section 7.5.11). If the participant agrees to submit this information, the pregnancy must be followed to determine the outcome, including spontaneous or voluntary termination, details of the birth, and the presence or absence of any birth defects, congenital abnormalities, or maternal and/or newborn complications. This follow-up should occur even if intended duration of the safety follow-up for the study has ended. Pregnancy report forms will be distributed to the study site to be used for this purpose. The investigator must immediately (within 24 hours of awareness) report to the Sponsor any pregnancy resulting in an abnormal outcome according to the procedures described for SAEs.

##### 7.5.7. Eliciting and Documenting Adverse Events

The investigator is responsible for ensuring that all AEs and SAEs are recorded in the eCRF and reported to the Sponsor.

Solicited ARs will be collected from Day 1 through 7 days after each dose only in Part A and Part C. Other (unsolicited) AEs will be collected from Day 1 through 28 days after each dose in Part A and Part C.

Both MAAEs and SAEs will be collected from participants as specified in the SoA until the end of their participation in each of the study parts (ie, Day 394 in Part A, OL-Day 178 in Part B, and BD-Day 361 in Part C). Any AEs that occur before administration of IP will be analyzed separately from AEs.

At every study site visit or telephone contact, participants will be asked a standard question to elicit any medically related changes in their well-being (including COVID-19 symptoms) according to the scripts provided. Participants will also be asked if they have been hospitalized, had any accidents, used any new medications, changed concomitant medication regimens (both prescription and over-the-counter medications), or had any nonstudy vaccinations.

In addition to participant observations, physical examination findings, or data relevant to participant safety classified as an AE will be documented on the AE page of the eCRF.

After the initial AE/SAE report, the investigator is required to proactively follow each participant at subsequent visits/contacts. All AEs and SAEs will be treated as medically appropriate and followed until resolution, stabilization, the event is otherwise explained, or the participant is LTFU (as defined in Section 6.4).

##### 7.5.8. Assessment of Intensity

An event is defined as "serious" when it meets at least one of the predefined outcomes as described in the definition of an SAE (Section 7.5.2), NOT when it is rated as severe.

The severity (or intensity) of an AR or AE refers to the extent to which it affects the participant's daily activities. The Toxicity Grading Scale for Healthy Adult and Adolescent Volunteers Enrolled in Preventative Vaccine Clinical Trials (DHHS 2007) will be used to categorize local and systemic reactogenicity events (solicited ARs), and vital sign measurements observed during this study. Specific criteria for local and systemic reactogenicity events are presented in Section 7.5.3.

The determination of severity for all unsolicited AEs should be made by the investigator based upon medical judgment and the definitions of severity as follows:

- Mild: These events do not interfere with the participant's daily activities.
- Moderate: These events cause some interference with the participant's daily activities and require limited or no medical intervention.
- Severe: These events prevent the participant's daily activity and require intensive therapeutic intervention.

Study staff should elicit from the participant the impact of AEs on the participant's activities of daily living to assess severity and document appropriately in the participant's source documentation. Changes in the severity of an AE should be documented in the participant's source documentation to allow an assessment of the duration of the event at each level of intensity to be performed. An AE characterized as intermittent requires documentation of onset and duration of each episode. An AE that fluctuates in severity during the course of the event is reported once in the eCRF at the highest severity observed.

##### 7.5.9. Assessment of Causality

The investigator's assessment of an AE's relationship to IP is part of the documentation process but is not a factor in determining what is or is not reported in the study.

The investigator will assess causality (ie, whether there is a reasonable possibility that the IP caused the event) for all AEs and SAEs. The relationship will be characterized using the following classification:

**Not related:** There is not a reasonable possibility of a relationship to the IP. Participant did not receive the IP OR temporal sequence of the AE onset relative to administration of the IP is not reasonable OR the AE is more likely explained by another cause than the IP.

**Related:** There is a reasonable possibility of a relationship to the IP. There is evidence of exposure to the IP. The temporal sequence of the AE onset relative to the administration of the IP is reasonable. The AE is more likely explained by the IP than by another cause.

##### 7.5.10. Reporting Adverse Events

The investigator is responsible for reporting all AEs that are observed or reported during the study, regardless of their relationship to IP or their clinical significance. If there is any doubt as to whether a clinical observation is an AE, the event should be reported.

All unsolicited AEs reported or observed during the study will be recorded on the AE page of the eCRF. Information to be collected includes the type of event, time of onset, investigator-specified assessment of severity (impact on activities of daily living) and relationship to IP, time of resolution of the event, seriousness, as well as any required treatment or evaluations, and outcome. The unsolicited AEs resulting from concurrent illnesses, reactions to concurrent illnesses, reactions to concurrent medications, or progression of disease states must also be reported. All AEs will be followed until they are resolved or stable or judged by the investigator to be not clinically significant. The Medical Dictionary for Regulatory Activities (MedDRA) will be used to code all unsolicited AEs.

Any medical condition that is present at the time that the participant is screened but does not deteriorate should not be reported as an unsolicited AE. However, if it deteriorates at any time during the study, it should be recorded as an unsolicited AE.

##### 7.5.11. Reporting SAEs

Prompt notification by the investigator to the Sponsor of an SAE is essential so that legal obligations and ethical responsibilities towards the safety of participants and the safety of a study intervention under clinical investigation are met.

Any AE considered serious by the investigator or that meets SAE criteria (Section 7.5.2) must be reported to the Sponsor immediately (within 24 hours of becoming aware of the SAE) via the EDC system. The investigator will assess whether there is a reasonable possibility that the IP caused the SAE. The Sponsor will be responsible for notifying the relevant regulatory authorities of any SAE as outlined in the 21 US CFR Parts 312 and 320. The investigator is responsible for notifying the institutional review board (IRB) directly.

If the eCRF is unavailable at the time of the SAE, the following contact information is to be used for SAE reporting:

- SAE Mailbox:
- SAE Hotline (USA and Canada): +1-866-599-1341
- SAE Fax line (USA and Canada): +1-866-599-1342

Regulatory reporting requirements for SAE are described in Section 7.5.15.

The investigator and any qualified designees are responsible for detecting, documenting, and recording events that meet the definition of an AE, including SAEs, and remain responsible for following up AEs that are serious, considered related to IP or study procedures, or that caused the participant to discontinue the study.

##### 7.5.12. Time Period and Frequency for Collecting AE and SAE Information

Medical occurrences that begin before the start of IP dosing but after obtaining informed consent will be recorded in the Medical History/Current Medical Conditions section of the eCRF and not in the AE section; however, if the condition worsens at any time during the study, it will be recorded and reported as an AE.

Adverse events may be collected as follows:

- Observing the participant
- Receiving an unsolicited complaint from the participant
- Questioning the participant in an unbiased and nonleading manner

Solicited ARs will be collected from the day of injection through 6 days after each dose. Other (unsolicited) AEs will be collected from the day of injection through 28 days after each dose.

Serious AEs will be collected from the start of IP dosing until the last day of study participation.

All SAEs will be recorded and reported to the Sponsor or designee immediately and in all circumstances within 24 hours. The investigator will submit any updated SAE data to the Sponsor within 24 hours of it being available.

An abnormal value or result from a clinical or laboratory evaluation can also indicate an AE if it is determined by the investigator to be clinically significant (eg, leads to dose modification or study drug discontinuation, or meets any serious criteria). If this is the case, it must be recorded in the source document and as an AE on the appropriate AE form(s). The evaluation that produced the value or result should be repeated until that value or result returns to normal or is stabilized and the participant's safety is not at risk.

Investigators are not obligated to actively seek AEs or SAEs after EOS participation. However, if the investigator learns of any SAE (including a death) at any time after a participant has withdrawn from or completed the study, and the investigator considers the event to be reasonably related to the IP or study participation, the investigator must promptly notify the Sponsor.

###### 7.5.13. Method of Detecting AEs and SAEs

Electronic diaries have specifically been designed for this study by the Sponsor. The diaries will include prelisted AEs (solicited ARs) and intensity scales; they will also include blank space for the recording of information on other AEs (unsolicited AEs) and concomitant medications/vaccinations.

The investigator is responsible for the documentation of AEs regardless of treatment group or suspected causal relationship to IP. For all AEs, the investigator must pursue and obtain information adequate to determine the outcome of the AE and to assess whether the AE meets the criteria for classification as an SAE requiring immediate notification to the Sponsor or its designated representative.

Care will be taken not to introduce bias when detecting AEs and/or SAEs. Open-ended and nonleading verbal questioning of the participant is the preferred method to inquire about the occurrence of AE.

###### 7.5.14. Follow-up of AEs and SAEs

After the initial AE/SAE report, the investigator is required to proactively follow each participant at subsequent visits and contacts.

All AEs and SAEs will be treated as medically appropriate and followed until resolution, stabilization, the event is otherwise explained, or the participant is LTFU, as defined in Section 6.4.

###### 7.5.15. Regulatory Reporting Requirements for SAEs

- Prompt notification by the investigator to the Sponsor of an SAE is essential so that legal obligations and ethical responsibilities towards the safety of participants and the safety of a study intervention under clinical investigation are met.

- The Sponsor has a legal responsibility to notify both the local regulatory authority and other regulatory agencies about the safety of a study intervention under clinical investigation. The Sponsor will comply with country-specific regulatory requirements relating to safety reporting to the regulatory authority, IRBs, and investigators.
- Investigator safety reports must be prepared for suspected unexpected serious adverse reaction according to local regulatory requirements and Sponsor policy and forwarded to investigators as necessary.
- An investigator who receives an investigator safety report describing an SAE or other specific safety information (eg, summary or listing of SAEs) from the Sponsor will review and then file it along with the IB and will notify the IRB, if appropriate according to local requirements.

#### 7.6. Safety Monitoring

The CRO's medical monitor, the Sponsor's medical monitor, and the individual study site investigators will monitor safety throughout the study.

##### 7.6.1. Data Safety Monitoring Board

Safety oversight will be under the direction of a DSMB composed of external independent consultants with relevant expertise. Members of the DSMB will be independent from the study conduct and free of conflict of interest.

The DSMB will have separate meetings by teleconference to review unblinded safety data when half of the study population (1,500 randomized participants) have reached Day 8 (1 week after Dose 1) and again approximately when 25% (750), 50% (1,500), and 75% (2,250) of enrolled participants have reached Day 36 (1 week after Dose 2). Recruitment will continue, as applicable, during the DSMB review period. The DSMB will operate under the rules of an approved charter that will be written and reviewed at the organizational meeting of the DSMB. Details regarding the DSMB composition, responsibilities, procedures, and frequency of data review will be defined in its charter.

The DSMB will convene on an ad hoc basis if any of the pause rules, described in Section 6.4, are met. The DSMB will review all available unblinded study data to adjudicate any potential study pauses and make recommendations on further study conduct, including requesting additional information, recommending stopping the study, recommending changes to study conduct and/or the protocol, or recommending additional operational considerations due to safety issues that arise during the study.

##### 7.6.2. Independent Cardiac Event Adjudication Committee

An independent cardiac event adjudication committee (CEAC) that includes pediatric and adult cardiologists will review suspected cases of myocarditis and pericarditis to determine if they meet CDC criteria of "probable" or "confirmed" events, and assess severity (Gargano et al 2021). Any cases that the CEAC assesses as representing probable or confirmed cases of myocarditis or pericarditis will be referred to the Sponsor, who will then make a final decision on whether to suspend further enrollment and/or study dosing based on an assessment of the overall potential risk to study participants.

The CEAC will operate under the rules of an approved charter that will be written and reviewed at the organizational meeting of the CEAC. Details regarding the CEAC composition, responsibilities, procedures, and frequency of data review will be defined in its charter.

#### 7.7. Treatment of Overdose

As the study treatment is to be administered by a healthcare professional, it is unlikely that an overdose will occur. Dose deviations will be tracked as protocol deviations (Section 10.2.8).

#### 7.8. Pharmacokinetics

Pharmacokinetic parameters are not evaluated in this study.

#### 7.9. Pharmacodynamics

Pharmacodynamic parameters are not evaluated in this study.

#### 7.10. Biomarkers

Immunogenicity assessments are presented in Section 7.2. Biomarkers are not evaluated in this study.

#### 7.11. Health Economics

Health economics are not evaluated in this study.

### 8. STATISTICAL ANALYSIS PLAN

This section summarizes the planned statistical analysis strategy and procedures for the study. The details of statistical analysis will be provided in the statistical analysis plan (SAP), which will be finalized before the clinical database lock for the study. If changes are made to primary and/or key secondary objectives and hypotheses or the statistical methods related to those hypotheses after the study has begun but prior to any data unblinding, then the protocol will be amended (consistent with ICH Guideline E9). Changes to other secondary or exploratory analyses made after the protocol has been finalized, along with an explanation as to when and why they occurred, will be listed in the SAP or clinical study report (CSR) for the study. Ad hoc exploratory analyses, if any, will be clearly identified in the CSR.

#### 8.1. Blinding and Responsibility for Analyses

See Section 3.1.1 regarding the addition of a participant decision visit as part of this protocol amendment.

Part A of this study is observer-blind. The investigator, study staff, study participants, study site monitors, and Sponsor personnel (or its designees) will be blinded to the IP administered until study end or initiation of Part B or Part C, with the following exceptions:

- Unblinded pharmacy personnel (of limited number) will be assigned to vaccine accountability procedures and will prepare and administer mRNA-1273 (or placebo) to all participants. These pharmacy personnel will have no study functions other than study vaccine management, documentation, accountability, preparation, and administration. They will not be involved in participant evaluations and will not reveal the identity of IP to either the participant or the blinded study site personnel involved in the conduct of the study unless this information is necessary in the case of an emergency.
- Unblinded study site monitors, not involved in other aspects of monitoring, will be assigned as the IP accountability monitors. They will have responsibilities to ensure that study sites are following all proper IP accountability, preparation, and administration procedures.
- An unblinded statistical and programming team will perform the preplanned IA (Section 8.6.1). Sponsor team members will be prespecified to be unblinded to the IA results and will not communicate the results of IA to the blinded investigators, study site staff, clinical monitors, or participants. This is detailed in the study Data Blinding Plan.

In Part A, the dosing assignment will be concealed by having the unblinded pharmacy personnel prepare the IP in a secure location that is not accessible or visible to other study staff. An opaque sleeve over the syringe used for injection will maintain the blind at the time of injection, as the doses containing mRNA-1273 will look different to that of placebo. Only delegated unblinded study site staff will conduct the injection procedure. Once the injection is completed, only the blinded study staff will perform further assessments and interact with the participants. Access to the randomization code will be strictly controlled at the pharmacy.

As documented in the study Data Blinding Plan, the prespecified Sponsor and CRO personnel will be unblinded for the IA of immunogenicity, safety, and efficacy.

The purpose of the unblinding is to enable the group to develop regulatory submission documents and to address questions from regulatory agencies during the regulatory review of the submission. After unblinding, this unblinded team will not participate in the conduct or execution of the subsequent course of the study. The study Data Blinding Plan provides details of the blinding/unblinding process and personnel. The study site staff, investigators, study monitors, and participants will remain blinded until the initiation of Part B or Part C.

##### 8.1.1. Breaking the Blind

A participant or participants may be unblinded in the event of an SAE or other severe event, or if there is a medical emergency requiring the identity of the product to be known to properly treat a participant. If a participant becomes seriously ill or pregnant during the study, the blind will be broken if knowledge of the administered vaccine will affect that participant's dosing options. In the event of a medical emergency requiring identification of the vaccine administered to an individual participant, the investigator will make every attempt to contact the Sponsor medical lead to explain the need for opening the code within 24 hours of opening the code. The investigator will be responsible for documenting the time, date, reason for the code break, and the names of the personnel involved.

In addition to the aforementioned situations where the blind may be broken, the data will also be unblinded to a statistical team at specified time points for IA as outlined in Section 8.6.1.

#### 8.2. Statistical Hypothesis

##### 8.2.1. Part A

If an accepted serum Ab threshold of protection against COVID-19 is established for the primary immunogenicity objective, the null hypothesis is that the percentage of participants on mRNA-1273 with serum Ab equal to or above the established threshold at Day 57 is ≤ 70% (ie, $H_0$: percentage of participants on mRNA-1273 ≤ 70% with serum Ab at Day 57 equal to or above the established threshold).

The study would be considered as meeting the immunogenicity objective if the 95% confidence interval (CI) of percentage of participants on mRNA-1273 rules out 70% (lower bound of the 95% CI > 70%).

If an accepted serum Ab threshold of protection against COVID-19 is not available for the primary immunogenicity objective, the immunogenicity analysis of primary vaccine response will be performed using the noninferiority tests of the two null hypotheses based on the two coprimary endpoints, respectively.

Coprimary endpoint 1: Ab GM at Day 57

The null hypothesis:

$H^1_0$: immunogenicity response to mRNA-1273 as measured by Ab GM at Day 57 is inferior in adolescents (12 to < 18 years of age) compared with that in young adults (18-25 years of age) using mRNA-1273 Study P301 data.

The noninferiority in Ab GM in adolescents compared with that in young adults (18-25 years of age) is demonstrated by that the lower bound of the 95% CI of the GMR rules out 0.67 (lower bound > 0.67) using a noninferiority margin of 1.5. The GMR is the ratio of the GM value of adolescents on mRNA-1273 in this study, Study P203, at Day 57 compared with the GM value of young adults (18-25 years of age) on mRNA-1273 in Study P301.

Coprimary endpoint 2: Ab seroresponse rate at Day 57

A definition of seroresponse will be provided in the SAP based on forthcoming information about assay performance.

The null hypothesis:

$H^2_0$: immunogenicity response to mRNA-1273 as measured by seroresponse rate at Day 57 is inferior in adolescents (12 to < 18 years of age) compared with that in young adults (18-25 years of age) using mRNA-1273 Study P301 data.

The noninferiority in seroresponse rate in adolescents compared with that in young adults (18-25 years of age) is demonstrated by that the lower bound of the 95% CI of the seroresponse rate difference rules out -10% (ie, lower bound > -10%) using the noninferiority margin of 10%. The seroresponse rate difference is defined as the rate in adolescents receiving mRNA-1273 minus the rate in young adults (18-25 years of age) receiving mRNA-1273 from Study P301.

The study would be considered as meeting the primary immunogenicity objective if noninferiority is demonstrated based on both coprimary endpoints.

Details regarding the assay to be used to assess noninferiority will be provided in the SAP.

##### 8.2.2. Part C

The immunogenicity analysis of BD vaccine response against the original strain will be performed using the noninferiority tests of the two null hypotheses based on the two coprimary endpoints, respectively.

**Coprimary Endpoint 1: Ab GM at BD-Day 29**

Null hypothesis

$H^1_0$: immunogenicity response to mRNA-1273 BD as measured by Ab GM at BD-Day 29 in adolescents (12-<18 years of age) in Study P203 Part C is inferior compared with Ab GM that at Day 57 (28 days after Dose 2) in the primary series of mRNA-1273 in young adults (18-25 years of age) in Study P301

The noninferiority in Ab GM at BD-Day 29 in Study P203 Part C compared with Ab GM at Day 57 in the primary series in young adults (18-25 years of age) in Study P301 will be demonstrated by the GMR 95% CI lower bound >0.67 using a noninferiority margin of 1.5. The GMR is defined as the GM of Ab at BD-D29 in Study P203 Part C compared with Ab GM at Day 57 (28 days after Dose 2) following the primary series of mRNA-1273 in Study P301.

**Coprimary Endpoint 2: Ab Seroresponse Rate (SRR) at BD-Day 29**

Null hypothesis

$H^2_0$: immunogenicity response to mRNA-1273 BD as measured by SRR at BD-Day 29 in adolescents (12-<18 years of age) in Study P203 Part C is inferior compared with SRR at Day 57 (28 days after Dose 2) following the primary series of mRNA-1273 in young adults (18-25 years of age) in Study P301.

The noninferiority in SRR at BD-D29 compared with SRR at Day 57 (28 days after Dose 2) following the primary series of mRNA-1273 will be demonstrated by the SRR 95% CI lower bound > -10% using the noninferiority margin of 10%. The SRR difference is defined as the SRR at BD-Day 29 Study P203 Part C minus the rate at Day 57 (28 days after Dose 2) following the primary series of mRNA-1273 in young adults in Study P301. The SRR is defined as a titer change from baseline (pre-Dose 1) below the LLOQ to ≥ 4 × LLOQ, or at least a 4-fold rise if baseline is ≥ LLOQ.

The primary immunogenicity objective in Part C is met if the noninferiority is demonstrated based on both coprimary endpoints.

**Key Secondary Endpoint 1: GM of Ab Against Circulating Strain/Delta at BD-Day 29**

Null hypothesis

$H^1_0$: immunogenicity response to mRNA-1273 BD as measured by GM of Ab against circulating strain/Delta at BD-Day 29 in adolescents (12-<18 years of age) in Study P203 Part C is inferior compared with GM of Ab against circulating strain/Delta at Day 57 (28 days after Dose 2) in the primary series of mRNA-1273 in young adults (18-25 years of age) in Study P301.

The noninferiority in Ab GM at BD-Day 29 in Study P203 Part C compared with Ab GM at Day 57 in the primary series in young adults (18-25 years of age) in Study P301 will be demonstrated by the GMR 95% CI lower bound >0.67 using a noninferiority margin of 1.5. The GMR is defined as the GM of Ab at BD-Day 29 in Study P203 Part C compared with Ab GM at Day 57 (28 days after Dose 2) following the primary series of mRNA-1273 in Study P301.

**Key Secondary Endpoint 2: Seroresponse Rate (SRR) of Ab Against Circulating Strain/Delta at BD-Day 29**

Null hypothesis

$H^2_0$: immunogenicity response to mRNA-1273 BD as measured by SRR of Ab against circulating strain/Delta at BD-Day 29 in adolescents (12-<18 years of age) in Study P203 Part C is inferior compared with SRR of Ab against circulating strain/Delta at Day 57 (28 days after Dose 2) following the primary series of mRNA-1273 in young adults (18-25 years of age) in Study P301.

The noninferiority in SRR at BD-D29 compared with SRR at Day 57 (28 days after Dose 2) following the primary series of mRNA-1273 will be demonstrated by the SRR 95% CI lower bound > -10% using the noninferiority margin of 10%. The SRR difference is defined as the SRR at BD-Day 29 Study P203 Part C minus the rate at Day 57 (28 days after Dose 2) following the primary series of mRNA-1273 in young adults in Study P301. The SRR is defined as a titer change from baseline (pre-Dose 1) below the LLOQ to ≥ 4 × LLOQ, or at least a 4-fold rise if baseline is ≥ LLOQ.

The key secondary immunogenicity objective in Part C is met if the noninferiority is demonstrated based on both key secondary endpoints.

**Multiplicity Adjustment**

A sequential hypothesis testing (fixed-sequence) method will be used to adjust multiplicity to preserve the family-wise Type I error rate (alpha = 0.05). The hypothesis testing for the two coprimary endpoints (geometric mean titer [GMT] and SRR) for the primary series of mRNA-1273 in Part A was completed and statistically significant based on data snapshot dated 08 May 2021, and thus the alpha level of 0.05 can be passed to Part C hypothesis testing. In Part C, the hypothesis testing for the two coprimary endpoints (GMT and SRR against the original strain) after BD of mRNA-1273 will be tested first at alpha level of 0.05. If the testing of both the coprimary endpoints in Part C is statistically significant (meeting the noninferiority success criteria of the coprimary endpoints), the alpha level of 0.05 will be passed to the hypothesis testing of the key secondary endpoints (GMT and SRR against circulating strain/Delta). The testing will continue through the sequence only until an endpoint is not statistically significant (did not meet specified noninferiority success criteria), in which case the testing will stop.

#### 8.3. Power and Sample Size

##### 8.3.1. Part A

The sample size of this study is driven by safety. Approximately 3,000 participants will be randomly assigned in a 2:1 ratio to receive mRNA-1273 and placebo (Section 5.2). With 2,000 participants exposed to mRNA-1273, the study has at least 90% probability to observe at least 1 participant with an AE at a true 0.25% AE rate.

Serum samples from all participants will be collected and banked, a subset of participants will be selected, and their samples will be processed for immunogenicity testing (the Immunogenicity Subset).

Approximately 362 participants who receive mRNA-1273 will be selected for the Immunogenicity Subset, with a target of 289 participants receiving mRNA-1273 in the PP Immunogenicity Subset (adjusting for approximately 20% of participants who may be excluded from the PP Immunogenicity Subset, as they may not have immunogenicity results due to any reason). The sample size of the Immunogenicity Subset may be updated with data from other mRNA-1273 studies or external data especially regarding a threshold of protection. In such a situation, the final sample size of the Immunogenicity Subset will be documented in the SAP.

For the primary immunogenicity objective, with approximately 289 participants in the PP Immunogenicity Subset, the study will have > 90% power to rule out 70% with a 2-sided 95% CI for the percentage of mRNA-1273 participants exceeding the acceptable threshold if the true rate of participants exceeding the acceptable threshold is 80%.

If an acceptable Ab threshold of protection against COVID-19 is not available at the time of analysis, for the primary immunogenicity objective, noninferiority tests of two null hypotheses based on two coprimary endpoints, respectively, will be performed. The sample size calculation for each of the two noninferiority tests was performed, and the larger sample size was chosen for the study.

- With approximately 289 participants in the PP Immunogenicity Subset in Study P203 and 289 participants in the PP Immunogenicity Subset in young adults (18-25 years of age) from Study P301, there will be 90% power to demonstrate noninferiority of the immune response as measured by Ab GM in adolescents in Study P203 at a 2-sided alpha of 0.05, compared with that in young adults (18-25 years of age) from Study P301 receiving mRNA-1273, assuming an underlying GMR value of 1 and a noninferiority margin of 1.5. The standard deviation (SD) of the log-transformed levels is assumed to be 1.5.

- With approximately 289 participants in the PP Immunogenicity Subset in Study P203 and 289 participants in the PP Immunogenicity Subset in young adults (18-25 years of age) from Study P301, there will be at least 90% power to demonstrate noninferiority of the immune response as measured by the seroresponse rate in adolescents in Study P203 at a 2-sided alpha of 0.05, compared with that in young adults (18-25 years of age) from Study P301 receiving mRNA-1273, assuming a true seroresponse rate of 85% in young adults (18-25 years of age) from Study P301, and a true seroresponse rate of 85% in adolescents in Study P203 (ie, true rate difference is 0 compared to young adults from Study P301), and a noninferiority margin of 10%.

##### 8.3.2. Part C

All participants enrolled in Part A or Part B who meet the eligibility criteria for BD will be offered a BD of mRNA-1273 50 µg. With more than 1,000 participants expected to receive mRNA-1273 BD, the study Part C has a 90% probability to observe at least 1 participant with an AE at a true AE rate of 0.25%.

Serum samples from all participants will be collected and banked, a subset of participants will be selected, and their samples will be processed for immunogenicity testing (the Immunogenicity Subset) at specified timepoints.

Approximately 362 participants who receive mRNA-1273 BD will be selected for the Immunogenicity Subset for Part C, with a target of 289 participants receiving mRNA-1273 BD in the PP Immunogenicity Subset for Part C (adjusting for approximately 20% of participants who may be excluded from the PP Immunogenicity Subset, as they may not have immunogenicity results due to any reason or may have protocol deviations impacting critical data).

For the primary immunogenicity objective in Part C, noninferiority tests of two null hypotheses based on two coprimary endpoints, respectively, will be performed.

– With approximately 289 participants receiving mRNA-1273 BD in the PP Immunogenicity Subset in Study P203 Part C and 289 participants in the PP Immunogenicity Subset in young adults (18-25 years of age) in Study P301, there will be 90% power to demonstrate noninferiority of the immune response post BD as measured by Ab GM in adolescents in Study P203 Part C compared with Ab GM in young adults (18-25 years of age) following primary series of mRNA-1273 in Study P301, at a 2-sided alpha of 0.05, assuming an underlying GMR value of 1 and a noninferiority margin of 1.5. The SD of the log-transformed levels is assumed to be 1.5.

- With approximately 289 participants receiving mRNA-1273 BD in the PP Immunogenicity Subset in Study P203 Part C and 289 participants in the PP Immunogenicity Subset in young adults (18-25 years of age) in Study P301, there will be at least 90% power to demonstrate noninferiority of the immune response post BD as measured by SRR in adolescents in Study P203 Part C compared with SRR in young adults of (18-25 years of age) following primary series of mRNA-1273 in Study P301, at a 2-sided alpha of 0.05, assuming true SRR of 90% in young adults (18-25 years of age) following primary series of mRNA-1273 in Study P301, and a true SRR of 90% post BD in adolescents in Study P203 Part C (ie, true rate difference is 0 compared to young adults [18-25 years of age] from Study P301), and a noninferiority margin of 10%.

#### 8.4. Analysis Sets

The analysis sets are defined in Table 4.

**Table 4: Analysis Sets**

| Analysis Set | Description |
|---|---|
| Randomization Set | All participants who are randomized, regardless of the participants' treatment status in the study. |
| Full Analysis Set (FAS) | All randomized participants who received at least 1 injection of IP. |
| Immunogenicity Subset | A subset of participants in the FAS will be selected for immunogenicity testing. |
| Per-protocol (PP) Immunogenicity Subset | A subset of participants in the FAS will be selected for immunogenicity testing. The PP Immunogenicity Subset includes participants selected for the Immunogenicity Subset who received planned doses of study vaccination per schedule, complied with immunogenicity testing schedule, and have no major protocol deviations that impact key or critical data. Participants who are seropositive at baseline will be excluded from the PP Immunogenicity Subset. The PP Immunogenicity Subset will be used for analyses of immunogenicity unless specified otherwise. |
| PP Set for Efficacy | All participants in the FAS who received planned doses of study vaccination, had no immunologic or virologic evidence of prior COVID-19, and have no major protocol deviations that impact key or critical efficacy data. |
| Solicited Safety Set | The Solicited Safety Set consists of FAS participants who contributed any solicited AR data.<br>The Solicited Safety Set will be used for the analyses of solicited ARs. |
| Safety Set | All randomized participants who receive at least 1 dose of IP. The Safety Set will be used for all analyses of safety except for the solicited ARs. |
| Modified Intent-to-Treat (mITT) Set | All participants in the FAS who have no serologic or virologic evidence of prior SARS-CoV-2 infection before the first dose of IP (both negative RT-PCR test for SARS-CoV-2 and negative serology test based on bAb specific to SARS-CoV-2 nucleocapsid) at baseline |
| Modified Intent-to-Treat-1 (mITT1) Set | All participants in the mITT Set excluding those who received the wrong treatment (ie, at least 1 dose received in Part A is not as randomized). |

Abbreviations: AR = adverse reaction; bAb = binding antibody; COVID-19 = coronavirus disease 2019; FAS = full analysis set; IP = investigational product; mITT = modified intent-to-treat; PP = per protocol; RT-PCR = reverse transcription polymerase chain reaction.

#### 8.5. Statistical Methods

Data from Part A, Part B, and Part C will be presented separately as applicable.

##### 8.5.1. Baseline Characteristics and Demographics

Demographic variables (eg, age, race, sex, height, weight, and BMI) and baseline characteristics will be summarized by treatment group. Summary statistics (mean, SD for continuous variables, and number and percentage for categorical variables) will be provided.

##### 8.5.2. Safety Analyses

All safety analyses will be based on the Safety Set, except summaries of solicited ARs, which will be based on the Solicited Safety Set. All safety analyses will be provided by treatment group.

Safety and reactogenicity will be assessed by clinical review of all relevant parameters including solicited ARs (local and systemic events), unsolicited AEs, SAEs, MAAEs, AESI, AEs leading to discontinuation, vital sign measurements, and physical examination findings.

The number and percentage of participants with any solicited local AR, with any solicited systemic AR, and with any solicited AR during the 7-day follow-up period after each injection (except Part B) will be summarized.

The number and percentage of participants with any solicited local AR, with any solicited systemic AR, and with any solicited AR during the 7-day follow-up period after each dose (except Part B) will be provided. A 2-sided 95% exact CI using the Clopper-Pearson method will also be provided for the percentage of participants with any solicited AR.

The number and percentage of participants with unsolicited AEs, SAEs, MAAEs, Grade 3 or higher ARs and AEs, and AEs leading to discontinuation from IP or withdrawal from the study will be summarized. Unsolicited AEs will be presented by MedDRA preferred term and system organ class.

The number of events of solicited ARs, unsolicited AEs/SAEs, and MAAEs will be reported in summary tables accordingly.

For all other safety parameters, descriptive summary statistics will be provided, and Table 5 summarizes analysis strategy for safety parameters. Further details will be described in the SAP.

**Table 5: Analysis Strategy for Safety Parameters**

| Safety Endpoint | Number and Percentage of Participants, Number of Events | 95% CI |
|---|---|---|
| Any solicited AR (overall and by local, systemic) | X | X |
| Any unsolicited AE | X | — |
| Any SAE | X | — |
| Any AESI | X | — |
| Any unsolicited MAAE | X | — |
| Any unsolicited treatment-related AE | X | — |
| Any treatment-related SAE | X | — |
| Discontinuation due to AE | X | — |
| Any severe AE | X | — |
| Any treatment-related severe AE | X | — |

Abbreviations: AE = adverse event; AESI = adverse event of special interest; AR = adverse reaction; CI = confidence interval; MAAE = medically attended adverse event; PT = preferred term; SAE = serious adverse event; SOC = system organ class.
Notes: 95% CI using the Clopper-Pearson method, X = results will be provided. Unsolicited AEs will be summarized by SOC and PT coded by MedDRA.

##### 8.5.3. Immunogenicity Analyses

The SAP will describe the complete set of immunogenicity analyses, including the approach to sample participants into an Immunogenicity Subset for analysis of immunogenicity. The PP Immunogenicity Subset is the primary analysis set for immunogenicity unless otherwise specified. The primary immunogenicity objective of this study is to use the immunogenicity response to infer efficacy in adolescents (12 to < 18 years in this study). The primary immunogenicity analysis is performed in Part A.

If an accepted serum Ab threshold of protection against COVID-19 is available based on data from other mRNA-1273 studies or external data, the number and percentage of participants with Ab greater than or equal to the threshold at Day 57 will be provided with a 2-sided 95% CI using the Clopper-Pearson method. If the lower bound of the 95% CI on the mRNA-1273 group is > 70%, the primary immunogenicity objective of this study will be considered to be met.

The percentage of participants with serum Ab greater than or equal to the threshold with 95% CI will be provided at each postbaseline time point. The CI will be calculated using the Clopper-Pearson method.

If an accepted serum Ab threshold of protection against COVID-19 is not established, the noninferiority of primary vaccine response as measured by Ab GM and seroresponse rate in adolescents compared with those in young adults (18-25 years of age) receiving mRNA-1273 will be assessed. The study is considered as meeting the primary immunogenicity objective if the noninferiority of the immune response to mRNA-1273 as measured by both GM and seroresponse rate at Day 57 is demonstrated in adolescents in this study at a 2-sided alpha of 0.05, compared with that in young adults (18-25 years of age) in Study P301 receiving mRNA-1273.

An analysis of covariance (ANCOVA) model will be carried out with Ab value at Day 57 as a dependent variable and a group variable (adolescents in Study P203 and young adults in Study P301) as the fixed variable. The GM values of the adolescents at Day 57 will be estimated by the geometric least square mean (GLSM) from the model. The GMR (ratio of GM values) will be estimated by the ratio of GLSM from the model. A corresponding 2-sided 95% CI will be provided to assess the difference in immune response for the adolescents in Study P203 compared to the young adults (18-25 years of age) in Study P301 at Day 57. The noninferiority of immune response to mRNA-1273 as measured by GM will be considered demonstrated if the lower bound of the 95% CI of the GMR is > 0.67 based on the noninferiority margin of 1.5.

The number and percentage (rate) of participants achieving Ab seroresponse at Day 57 will be summarized. The difference of seroresponse rates between adolescents receiving mRNA-1273 in Study P203 and young adults (18-25 years of age) receiving mRNA-1273 in Study P301 will be calculated with 95% CI. The noninferiority in seroresponse rate of adolescents in Study P203 compared to young adults (18-25 years of age) in Study P301 will be considered demonstrated if the lower bound of the 95% of the seroresponse rate difference is > -10%, based on the noninferiority margin of 10%.

In addition, the GM level of specific nAb and bAb with corresponding 95% CI will be provided at each time point. The 95% CIs will be calculated based on the t-distribution of the log transformed values then back transformed to the original scale. The geometric mean fold-rise (GMFR) of nAb and bAb with corresponding 95% CI will be provided at each time point with Day 57 as the primary time point of interest. Descriptive summary statistics including median, minimum, and maximum will also be provided.

##### 8.5.4. Efficacy Analyses

To evaluate the incidence of COVID-19 after vaccination with mRNA-1273 or placebo, the incidence rate will be provided by vaccination group, calculated as the number of cases divided by the total person-time. The incidence rate ratio of mRNA-1273 versus placebo may be provided with its 95% CI computed using the exact method conditional upon the total number of cases adjusted by the total person-time.

For SARS-CoV-2 infection (serologically confirmed SARS-CoV-2 infection or COVID-19), regardless of symptomatology or severity, infection rate will be provided by vaccination group. The infection rate ratio of mRNA-1273 versus placebo may be provided with its 95% CI using the exact method conditional upon the total number of cases adjusted by the total person-time. The incidence rate of asymptomatic SARS-CoV-2 infection will also be provided.

The secondary efficacy analyses will be performed in the PP set, with sensitivity analyses in the FAS, mITT Set, and mITT1 Set.

##### 8.5.5. Long-term Analysis (including Part B)

Long-term analysis will be performed including data collected in the Open-label Observational Phase (Part B), and prior to BD if a BD is received. The analysis will include participants who are randomized to mRNA-1273 in Part A and remained in the study with or without unblinding, and participants who are originally randomized to placebo and later crossed over to receive mRNA-1273 in Part B after unblinding. Long-term analysis of applicable safety, efficacy, and immunogenicity endpoints will be summarized descriptively by treatment cohort as defined in Table 6 without treatment group comparison.

In the long-term safety analysis, unsolicited AEs (SAE, AESI, MAAE, and AE leading to discontinuation) and deaths will be summarized.

In the long-term immunogenicity analysis, nAb and bAb values will be summarized at specified timepoints.

In the long-term efficacy analysis, the incidence rates of COVID-19 and of SARS-CoV-2 infection cases will be counted starting 14 days after the second dose of IP for participants in treatment cohorts of mRNA-1273 and Placebo or starting 14 days after the second dose of mRNA-1273 for participants in the Placebo-mRNA-1273 cohort. Incidence rate with 95% CI adjusting for person-time will be provided. The incidence rate of asymptomatic SARS-CoV-2 infection will also be provided.

**Table 6: Treatment Cohorts for the Long-term Analysis**

| Long-term Treatment Cohort | Description |
|---|---|
| **mRNA-1273** | Participants randomized to mRNA-1273 in the Blinded Phase |
| **Placebo** | Participants randomized to Placebo in the Blinded Phase who did not cross over to mRNA-1273 in the Open-label Observational Phase |
| **Placebo-mRNA-1273** | Participants randomized to Placebo in the Blinded Phase who crossed over to mRNA-1273 in the Open-label Observational Phase |

##### 8.5.6. Booster Phase Analysis (Part C)

Booster phase analysis will be performed in participants who receive a BD in Part C. Safety, immunogenicity endpoints, and incidence of COVID-19 and SARS-CoV-2 infection will be summarized descriptively, and by pre-booster SARS-CoV-2 status if applicable.

In the booster phase safety analysis, the number and percentage of participants with unsolicited AEs, severe AEs, treatment-related AEs, SAEs, MAAEs, AESIs, and AEs leading to discontinuation from the study participation in Part C will be summarized.

The number and percentage of participants with any solicited local AR, with any solicited systemic AR, and with any solicited AR during the 7-day follow-up period after the BD will be provided.

In the Part C booster phase, for the coprimary endpoints (Ab GM and SRR against the original strain) and key secondary endpoints (Ab GM and SRR against the circulating strain/Delta), the following analyses will be performed for the respective hypothesis testing:

- The GM titers with 95% CI will be summarized using t-distribution of the log transferred values and then back transformed to the original scale. The GMR with 95% CI to compare post-booster GM at BD-Day 29 in adolescents in Study P203 with the primary series GM at Day 57 (28 days after Dose 2) in young adults (18-25 years of age) in Study P301 will be computed based on the t-distribution of mean difference in the log transferred values and then back transformed to the original scale.
- The SRR with 95% CI (using Clopper-Pearson method) will be summarized. The SRR difference with 95% CI (using Miettinen-Nurminen score method) to compare post-booster SRR at BD-Day 29 in adolescents in Study P203 with the primary series SRR at Day 57 (28 days after Dose 2) in young adults in Study P301 will be computed. The SRR is defined as a titer change from baseline (pre-Dose 1) below the LLOQ to $\geq 4 \times$ LLOQ, or at least a 4-fold rise if baseline is $\geq$ LLOQ.

In the booster phase immunogenicity analyses, the GM titers or levels for specific nAb and bAb, with corresponding 95% CI will be provided at BD-Day 1, BD-Day 29, BD-Day 181, and BD-Day 361 (Part C). GMFR relative to pre-Dose 1, BD-Day 1 and prior Day 57 in the phase of primary 2-dose series of mRNA-1273 will be summarized with 95% CIs at BD-Day 29, BD-Day 181, and BD-Day 361 (Part C). The 95% CIs will be calculated based on the t-distribution of the log transformed values then back transformed to the original scale. Descriptive summary statistics including median, minimum, and maximum will also be provided. The immunogenicity descriptive summaries will be provided in the participants who have available nAb and bAb assay results for the specified time points.

In the booster phase, the incidence rate of the first occurrence of symptomatic COVID-19 starting 14 days after the BD will be summarized by pre-booster SARS-CoV-2 status. The incidence rate of SARS-CoV-2 infection (symptomatic or asymptomatic infection) starting 14 days after the BD, and the incidence rate of asymptomatic SARS-CoV-2 infection starting 14 days after the BD will be summarized. The incidence rates with 95% CI will be computed adjusting for person-years starting from BD-Day 1 in Part C.

##### 8.5.7. Exploratory Analyses

Exploratory analyses will be described in the SAP before database lock.

##### 8.5.8. Subgroup Analyses

Subgroup analyses will be performed as described in the SAP.

#### 8.6. Study Analyses

##### 8.6.1. Interim Analyses

More than one IA may be performed.

- The IA of immunogenicity, safety, and efficacy will be performed after Day 57 immunogenicity data are available for the immunogenicity subset and at least 1,500 participants (1,000 participants receiving mRNA-1273) have completed Day 57 (1 month after Dose 2, Part A). This IA will be considered the primary analysis of immunogenicity.
- An IA of immunogenicity and safety may be performed after all or subset of participants who receive booster dose have completed BD-Day 29 after the booster dose.
- At the Sponsor’s discretion, a CSR may be developed for an IA.

##### 8.6.2. Final Analysis

The final analysis of all applicable endpoints will be performed after all participants have completed all planned study procedures. Results of this analysis will be presented in a final CSR, including individual listings.

Additional information about all study analyses may be provided in the SAP.

### 9. REFERENCES

Adler Y, Charron P, Imazio M, Badano L, Barón-Esquivias G, Bogaert J, et al. 2015 ESC guidelines for the diagnosis and management of pericardial diseases: the task force for the diagnosis and management of pericardial diseases of the european society of cardiology (ESC) endorsed by: The European Association for Cardio-Thoracic Surgery (EACTS). Eur Heart J. 2015 Nov 7;36(42):2921-64.

Aretz HT. Myocarditis: the Dallas criteria. Hum Pathol. 1987 Jun;18(6):619-24.

Boehmer TK, DeVies J, Caruso E, van Santen KL, Tang S, Black CL, et al. Changing age distribution of the COVID-19 pandemic — United States, May-August 2020. MMWR Morb Mortal Wkly Rep. 2020;69(39):1404-1409.

Centers for Disease Control and Prevention (CDC). 2020a. Coronavirus Disease 2019 (COVID-19). Retrieved from: https://covid.cdc.gov/covid-data-tracker/#cases. (accessed 2020 Sep 21).

Centers for Disease Control and Prevention (CDC). 2020b. Coronavirus Disease 2019 (COVID-19). Retrieved from: https://covid.cdc.gov/covid-data-tracker/#demographics. (accessed 2020 Sep 21).

Centers for Disease Control and Prevention (CDC). 2020c. Coronavirus Disease 2019 (COVID-19). Retrieved from: https://www.cdc.gov/coronavirus/2019-ncov/symptoms-testing/symptoms.html (accessed 2020 Jun 11).

Chen Y, Lu S, Jia H, Deng Y, Zhou J, Huang B, et al. A novel neutralizing monoclonal antibody targeting the N-terminal domain of the MERS-CoV spike protein. Emerg Microbes Infect. 2017;6(5):e37.

Chen N, Zhou M, Dong X, Qu J, Gong F, Han Y, et al. Epidemiological and clinical characteristics of 99 cases of 2019 novel coronavirus pneumonia in Wuhan, China: a descriptive study. Lancet. 2020;395(10223):507-513.

Chin J, Magoffin RL, Shearer LA, Schieble JH, Lennette EH. Field evaluation of a respiratory syncytial virus vaccine and a trivalent parainfluenza virus vaccine in a pediatric population. Am J Epidemiol. 1969;89(4):449–63.

ClinicalTrials.gov [Internet]. Bethesda (MD): National Library of Medicine (US). - Identifier NCT03076385, Safety, tolerability, and immunogenicity of VAL-506440 in healthy adult subjects. Retrieved from: https://clinicaltrials.gov/ct2/keydates/NCT03076385. (accessed 2020 Jun 01).

ClinicalTrials.gov [Internet]. Bethesda (MD): National Library of Medicine (US). - Identifier NCT04917861, A Study of Zika Vaccine mRNA-1893 in Adult Participants Living in Endemic and Non-Endemic Flavivirus. Retrieved from: https://clinicaltrials.gov/ct2/keydates/NCT04917861. (accessed 2021 Jul 22).

ClinicalTrials.gov [Internet]. Bethesda (MD): National Library of Medicine (US). - Identifier NCT03345043, Safety, tolerability, and immunogenicity of VAL-339851 in healthy adult subjects. Retrieved from: https://clinicaltrials.gov/ct2/keydates/NCT03345043. (accessed 2020 Jun 01).

ClinicalTrials.gov [Internet]. Bethesda (MD): National Library of Medicine (US). - Identifier NCT03382405, Safety, reactogenicity, and immunogenicity of cytomegalovirus vaccines mRNA-1647 and mRNA-1443 in healthy adults. Retrieved from: https://clinicaltrials.gov/ct2/keydates/NCT03382405. (accessed 2020 Jun 01).

ClinicalTrials.gov [Internet]. Bethesda (MD): National Library of Medicine (US). - Identifier NCT03392389, Safety, reactogenicity, and immunogenicity of mRNA-1653 in healthy adults. Retrieved from: https://clinicaltrials.gov/ct2/keydates/NCT03392389. (accessed 2020 Jun 01).

ClinicalTrials.gov [Internet]. Bethesda (MD): National Library of Medicine (US). - Identifier NCT04283461, Safety and immunologic protocol of SARS-CoV-2 vaccine (mRNA-1273) to treat novel coronavirus; 2020 Feb 22. Retrieved from: https://clinicaltrials.gov/ct2/keydates/NCT04283461. (accessed 2020 Jun 01).

ClinicalTrials.gov [Internet]. Bethesda (MD): National Library of Medicine (US). - Identifier NCT04405076, Dose-Confirmation Study to Evaluate the Safety, Reactogenicity, and Immunogenicity of mRNA-1273 COVID-19 Vaccine in Adults Aged 18 Years and Older; 2020 May 28. Retrieved from: https://clinicaltrials.gov/ct2/keydates/NCT04405076. (accessed 2020 Aug 24).

ClinicalTrials.gov [Internet]. Bethesda (MD): National Library of Medicine (US). - Identifier NCT04470427, A Study to Evaluate Efficacy, Safety, and Immunogenicity of mRNA-1273 Vaccine in Adults Aged 18 Years and Older to Prevent COVID-19; 2020 Jul 14. Retrieved from: https://clinicaltrials.gov/ct2/keydates/NCT04470427. (accessed 2020 Aug 24).

Corbett KS, Edwards DK, Leist SR, Abiona OM, Boyoglu-Barnum S, Gillespie RA, et al. SARS-CoV-2 mRNA vaccine development enabled by prototype pathogen preparedness. Nature. 2020a. doi:10.1038/s41586-020-2622-0.

Corbett KS, Flynn B, Foulds KE, Francica JR, Boyoglu-Barnum S, Werner AP, et al. Evaluation of the mRNA-1273 vaccine against SARS-CoV-2 in nonhuman primates. N Engl J Med. 2020b;383(16):1544-55. doi: 10.1056/NEJMoa2024671.

Corti D, Zhao J, Pedotti M, Simonelli L, Agnihothram S, Fett C, et al. Prophylactic and postexposure efficacy of a potent human monoclonal antibody against MERS coronavirus. Proc Natl Acad Sci U S A. 2015;112(33):10473-8.

Cruz AT, Zeichner SL. COVID-19 in children: initial characterization of the pediatric disease. Pediatrics. 2020;145(6):e20200834.

Department of Health and Human Services (DHHS), Food and Drug Administration, Center for Biologics Evaluation and Research (US). Guidance for industry: Toxicity grading scale for healthy adult and adolescent volunteers enrolled in preventative vaccine clinical trials. September 2007 [cited 2019 Apr 10] [10 screens]. Available from:

https://www.fda.gov/downloads/BiologicsBloodVaccines/GuidanceComplianceRegulatoryInformation/Guidances/Vaccines/ucm091977.pdf.

Department of Health and Human Services (DHHS), Food and Drug Administration, Center for Biologics Evaluation and Research (US). Guidance for Industry, Investigators, and Institutional Review Boards. FDA guidance on conduct of clinical trials of medical products during COVID-19 public health emergency. September 2020. Updated on April 16, 2020. [cited 2020 Apr 17]. Available from: https://www.fda.gov/media/136238/download.

Ferreira VM, Schulz-Menger J, Holmvang G, Kramer CM, Carbone I, Sechtem U, et al. Cardiovascular magnetic resonance in nonischemic myocardial inflammation: expert recommendations. J Am Coll Cardiol. 2018;72(24):3158-76.

Fontanet A, Grant R, Tondeur L, Madec Y, Grzelak L, Cailleau I, et al. SARS-CoV-2 infection in primary schools in northern France: A retrospective cohort study in an area of high transmission. medRxiv. 2020

Fulginiti VA, Eller JJ, Downie AW, Kempe CH. Altered reactivity to measles virus. Atypical measles in children previously immunized with inactivated measles virus vaccines. JAMA. 1967;202(12):1075-80.

Gargano JW, Wallace M, Hadler SC, Langley G, Su JR, Oster ME, et al. Use of mRNA COVID-19 vaccine after reports of myocarditis among vaccine recipients: update from the advisory committee on immunization practices - United States, June 2021. MMWR Morb Mortal Wkly Rep. 2021;70(27):977-82.

Hardin AP, Hackell JM, Committee on practice and ambulatory medicine. Age limit of pediatrics. Pediatrics. 2017;140(3):e20172151.

Jackson LA, Anderson EJ, Rouphael NG, Roberts PC, Makhene M, Coler RN, et al. An mRNA vaccine against SARS-CoV-2 – preliminary report. N Engl J Med. 2020 Jul 14. Online ahead of print.

Johnson RF, Bagci U, Keith L, Tang X, Mollura DJ, Zeitlin L, et al. 3B11-N, a monoclonal antibody against MERS-CoV, reduces lung pathology in rhesus monkeys following intratracheal inoculation of MERS-CoV Jordan-n3/2012. Virology. 2016;490:49-58.

Ju B, Zhang Q, Ge J, Wang R, Sun J, GE X, et al. Human neutralizing antibodies elicited by SARS-CoV-2 infection. Nature. 2020;584(7819)115-19. doi:10.1038/s41586-020-2380-z.

Kim Y, Lee H, Park K, Park S, Lim JH, So MK, et al. Selection and characterization of monoclonal antibodies targeting Middle East Respiratory Syndrome coronavirus through a human synthetic fab phage display library panning. Antibodies (Basel). 2019;8(3):42.

Leeb RT, Price S, Sliwa S, Kimball A, Szucs L, Caruso E, et al. COVID-19 trends among school-aged children — United States, March 1-September 19 2020. MMWR Morb Mortal Wkly Rep. 2020;69(39):1410-15.

Licciardi F, Giani T, Baldini L, Favalli EG, Caporali R, Cimaz R. COVID-19 and what pediatric rheumatologists should know: a review from a highly affected country. Pediatr Rheumatol Online J. 2020;18(1):35.

Robbiani DF, Gaebler C, Muecksch F, Lorenzi JC, Wang Z, Cho A, et al. Convergent antibody responses to SARS-CoV-2 in convalescent individuals. Nature. 2020;584(7821):437-42.

Rothan HA, Byrareddy SN. The epidemiology and pathogenesis of coronavirus disease (COVID 19) outbreak. J Autoimmun. 2020;109:102433.

Rüggeberg JU, Gold MS, Bayas JM, et al. Anaphylaxis: case definition and guidelines for data collection, analysis, and presentation of immunization safety data. Vaccine. 2007;25(31):5675-84.

Schwartz NG, Moorman AC, Makaretz A, Chang KT, Chu VT, Szablewski CM, et al. Adolescent with COVID-19 as the source of an outbreak at a 3-week family gathering — four states, June-July 2020. MMWR Morb Mortal Wkly Rep. 2020;69(40):1457–1459.

Shen KL, Yang YH, Jiang RM, Wang TY, Zhao DC, Jiang Y, Lu XX, et al. Updated diagnosis, treatment and prevention of COVID-19 in children: experts' consensus statement (condensed version of the second edition). World J Pediatr. 2020:16(3)232-39.

Thomas SJ, Yoon IK. A review of Dengvaxia®: development to deployment. Hum Vaccin Immunother. 2019;15(10), 2295-314.

Wang L, Shi W, Joyce MG, Modjarrad K, Zhang Y, Leung K, et al. Evaluation of candidate vaccine approaches for MERS-CoV. Nat Commun. 2015;6:7712.

Wang L, Shi W, Chappell JD, Joyce MG, Zhang Y, Kanekiyo M, et al. Importance of neutralizing monoclonal antibodies targeting multiple antigenic sites on the middle east respiratory syndrome coronavirus spike glycoprotein to avoid neutralization escape. J Virol. 2018;92(10):e02002-17.

Widjaja I, Wang C, van Haperen R, Gutiérrez-Álvarez J, van Dieren B, Okba NMA, et al. Towards a solution to MERS: protective human monoclonal antibodies targeting different domains and functions of the MERS-coronavirus spike glycoprotein. Emerg Microbes Infect. 2019;8(1):516-30.

Wrapp D, Wang N, Corbett KS, Goldsmith JA, Hseih CL, Abiona O, et al. Cryo-EM structure of the 2019-nCoV spike in the prefusion conformation. Science. 2020;367(6483):1260-3.

World Health Organization (WHO). Dengue vaccine: WHO position paper, September 2018 – recommendations. Vaccine. 2019;37(35):4848-49.

World Health Organization. (2020a, Jun 25). Coronavirus disease 2019 (COVID-19) Situation Report – 157. Retrieved from: https://www.who.int/docs/default-source/coronaviruse/situation-reports/20200625-covid-19-sitrep-157.pdf?sfvrsn=423f4a82_2. (accessed 2020 Jun 26).

World Health Organization. (2020b, Sep 28). WHO coronavirus disease (COVID-19) dashboard. Retrieved from: https://covid19.who.int/. (accessed 2020 Sep 28).

Wrapp D, Wang N, Corbett KS, Goldsmith JA, Hseih CL, Abiona O, et al. Cryo-EM structure of the 2019-nCoV spike in the prefusion conformation. Science. 2020;367(6483):1260-3.

Yu X, Zhang S, Jiang L, Cui Y, Li D, Wang D, et al. Structural basis for the neutralization of MERS-CoV by a human monoclonal antibody MERS-27. Sci Rep. 2015;5:13133.

Zent O, Arras-Reiter C, Broeker M, Hennig R. Immediate allergic reactions after vaccinations - a postmarketing surveillance review. Eur J Pediatr. 2002;161(1):21-5.

### 10. SUPPORTING DOCUMENTATION AND OPERATIONAL CONSIDERATIONS

#### 10.1. APPENDIX 1: Schedule of Assessments

The schedules of assessments are presented in Table 7 for Part A Table 9 for Part B, and Table 10 for Part C.

If a participant cannot attend a study site visit (scheduled or unscheduled) with the exception of Screening or Day 1, a home visit is acceptable if performed by appropriately delegated study site staff or a home healthcare service provided by the Sponsor (Section 7). If neither a participant visit to the study site nor a home visit to the participant is possible (with the aforementioned exceptions), a safety telephone call should be performed that includes the assessments scheduled for the safety telephone calls.

After the Participant Decision Clinic Visit, participants will continue to follow the Part A SoA (Table 7) or Part B SoA (Table 9), as described in Section 3.1.2 and as shown in Figure 5. All eligible participants will be offered a BD and those who choose to receive BD will follow Part C SoA (Table 10).

**Table 7: Schedule of Assessments Part A, Blinded Phase; Part B Open-Label Observational Phase for Participants who Received mRNA-1273 in Part A**

| **Visit Number** | 0 | 1 | 2 | 3 | 4 | 5 | - | | 6 | - | | 7 |
|---|---|---|---|---|---|---|---|---|---|---|---|---|
| **Type of Visit** | C | C | Virtual Call | C | Virtual Call | C | SFU | | C | SFU | | C |
| **Month Time Point** | | M0 | | M1 | | M2 | eDiary | SC | M7 | eDiary | SC | M13 |
| **Study Visit Day** | D0[1] (Screening) | D1 (Baseline) | D8[2] | D29[3] | D36[2, 3] | D57[2, 3] | Every 4 weeks D71 – D183[3, 4] | Every 4 weeks D85– D197[3, 5] | D209/ Participant Decision Visit[3,6] | Every 4 weeks D223– D363[3, 4] | Every 4 weeks D237– D377[3, 5] | D394[3] |
| **Window Allowance (Days)** | - 28 | | + 3 | + 7 | + 3 | + 7 | ±3 | ± 3 | - 56/+ 56 | ± 3 | ± 3 | ± 14 |
| **Days Since Most Recent Injection** | - | 0 | 7 | 28/0 | 7 | 28 | - | - | 180 | - | - | 365 |
| Informed consent/assent form, demographics, concomitant medications, medical history | X | | | | | | | | | | | |
| Revised informed consent/assent form | | | | | | | | | X | | | |
| Review of inclusion and exclusion criteria | X | X | | | | | | | | | | |
| Physical examination including vital signs, height, weight[7] | X | X | | X | | X | | | X | | | X |
| Pregnancy test[8] | X | X | | X | | | | | | | | |
| Randomization | | X | | | | | | | | | | |

| **Visit Number** | 0 | 1 | 2 | 3 | 4 | 5 | - | | 6 | - | | 7 |
|---|---|---|---|---|---|---|---|---|---|---|---|---|
| **Type of Visit** | C | C | Virtual Call | C | Virtual Call | C | SFU | | C | SFU | | C |
| **Month Time Point** | | M0 | | M1 | | M2 | eDiary | SC | M7 | eDiary | SC | M13 |
| **Study Visit Day** | D0[1] (Screening) | D1 (Baseline) | D8[2] | D29[3] | D36[2, 3] | D57[2, 3] | Every 4 weeks D71 – D183[3,4] | Every 4 weeks D85– D197[3, 5] | D209/ Participant Decision Visit[3,6] | Every 4 weeks D223– D363[3, 4] | Every 4 weeks D237– D377[3, 5] | D394[3] |
| **Window Allowance (Days)** | - 28 | | + 3 | + 7 | + 3 | + 7 | ±3 | ± 3 | - 56/+ 56 | ± 3 | ± 3 | ± 14 |
| **Days Since Most Recent Injection** | - | 0 | 7 | 28/0 | 7 | 28 | - | - | 180 | - | - | 365 |
| Study injection (including 30-minute postdose observation period) | | X | | X | | | | | | | | |
| Blood sample for vaccine immunogenicity[9] | | X | | | | X | | | X | | | X |
| Nasopharyngeal or nasal swab sample for SARS-CoV-2[10] | | X | | X | | X | | | X | | | |
| Surveillance for COVID-19/ Illness visit[11]/ Unscheduled visit | | X | X | X | X | X | X | X | X | X | X | X |
| Convalescent Visit[12] | | X | X | X | X | X | X | X | X | X | X | X |
| eDiary activation for recording solicited ARs (7 days)[13] | | X | | X | | | | | | | | |
| Review of eDiary data | | | X | | X | | | | | | | |
| Follow-up safety telephone calls[14] | | | | | | | | X | | X | | |

| **Visit Number** | 0 | 1 | 2 | 3 | 4 | 5 | - | | 6 | - | | 7 |
|---|---|---|---|---|---|---|---|---|---|---|---|---|
| **Type of Visit** | C | C | Virtual Call | C | Virtual Call | C | SFU | | C | SFU | | C |
| **Month Time Point** | | M0 | | M1 | | M2 | eDiary | SC | M7 | eDiary | SC | M13 |
| **Study Visit Day** | D0[1] (Screening) | D1 (Baseline) | D8[2] | D29[3] | D36[2, 3] | D57[2, 3] | Every 4 weeks D71 – D183[3,4] | Every 4 weeks D85– D197[3, 5] | D209/ Participant Decision Visit [3,6] | Every 4 weeks D223– D363[3, 4] | Every 4 weeks D237– D377[3, 5] | D394[3] |
| **Window Allowance (Days)** | - 28 | | + 3 | + 7 | + 3 | + 7 | ±3 | ± 3 | - 56/+ 56 | ± 3 | ± 3 | ± 14 |
| **Days Since Most Recent Injection** | - | 0 | 7 | 28/0 | 7 | 28 | - | - | 180 | - | - | 365 |
| Recording of unsolicited AEs | | X | X | X | X | X | | | | | | |
| Recording of MAAEs and concomitant medications relevant to or for the treatment of the MAAE[15] | | X | X | X | X | X | X | | X | X | | X |
| Recording of SAEs and concomitant medications relevant to or for the treatment of the SAE[15] | | X | X | X | X | X | X | | X | X | | X |
| Recording of AESI | | X | X | X | X | X | X | | X | X | | X |
| Recording of concomitant medications and non-study vaccinations[15] | | X | X | X | X | X | | | | | | |
| Study completion | | | | | | | | | | | | X |

Abbreviations: AE = adverse event; AESI = adverse event of special interest; AR = adverse reaction; C = clinic visit; COVID-19 = coronavirus disease 2019; D = day; eDiary = electronic diary; FDA = US Food and Drug Administration; ICF = informed consent form; IRB = institutional review board; M = month; MAAE = medically attended AE; SC = safety (telephone) call; SFU = safety follow-up; RT-PCR = reverse transcriptase polymerase chain reaction; SAE = serious adverse event; SARS-CoV-2 = Severe Acute Respiratory Syndrome coronavirus 2.

**Note:** In accordance with FDA Guidance on Conduct of Clinical Trials of Medical Products during COVID-19 Public Health Emergency (DHHS 2020), investigators may convert study site visits to home visits or telemedicine visits with the approval of the Sponsor.

**Note**: If during the participant decision visit (Table 8) the participant is unblinded and determined to have received 2 doses of mRNA-1273 in Part A, due to statistical considerations, they will be considered in the Open-label Observational Phase but will continue to follow the Part A Schedule of Assessments.

1. Day 0 and Day 1 may be combined on the same day. Additionally, the Day 0 visit may be performed over multiple visits if preformed within the 28-day screening window.
2. All scheduled study visits should be completed within the respective visit windows. If the participant is not able to attend a study site visit as a result of the COVID-19 pandemic (self-quarantine or disruption of study site activities following business continuity plans and/or local government mandates for “stay at home” or “shelter in place”), a safety telephone call to the participant should be made in place of the study site visit. The safety telephone call should encompass all scheduled visit assessments that can be completed remotely, such as assessment for AEs and concomitant medications (eg, as defined in scheduled safety telephone calls). Home visits will be permitted for all nondosing visits, with the exception of Screening, if a participant cannot visit the study site as a result of the COVID-19 pandemic. Home visits must be permitted by the study site IRB and the participant via informed consent/assent and have prior approval from the Sponsor (or its designee).
3. If the visit for the second dose (Day 29) is disrupted and cannot be completed at Day 29 +7 days as a result of the COVID-19 pandemic (self-quarantine or disruption of study site activities following business continuity plans and/or local government mandates for “stay at home” or “shelter in place”), the visit window may be extended to Day 29 + 21 days. When the extended visit window is used, the remaining study visits should be rescheduled to follow the inter-visit interval from the actual date of the second dose.
4. Safety follow-up via an eDiary questionnaire will be performed every 4 weeks from Day 71 to Day 183 and again from Day 223 to Day 363.
5. Safety follow-up via a safety telephone call will be performed every 4 weeks from Day 85 to Day 197 and again from Day 237 to Day 377.
6. The Participant Decision visit may be performed over multiple visits. Once the Participant Decision Visit has been initiated, all assessments must be completed within a 7-day period.
7. Physical examination: A full physical examination, including height and weight, will be performed at Day 1, Day 29, Day 57, Day 209, and Day 394. BMI will be calculated only at Screening Visit (Day 0). Symptom-directed physical examinations may be performed at other time points at the discretion of the investigator. On each injection day before injection and again 7 days after injection, the arm receiving the injection should be examined and the associated lymph nodes should be evaluated. Any clinically significant finding identified during a study visit should be reported as an MAAE. Vital signs are to be measured pre- and post dose on days of injection (Day 1 and Day 29). When applicable, vital signs should be measured before blood collection. Participants who are febrile (body temperature ≥ 38.0°C/100.4°F) before injection on Day 1 or Day 29 must have the visit rescheduled within the relevant visit window to receive the injection. Afebrile participants with minor illnesses can be enrolled at the discretion of the investigator.
8. Pregnancy test at Screening and Day 1 and before the second study injection will be a point-of-care urine test. At the discretion of the investigator, a pregnancy test either via blood or point-of-care urine test can be performed at any time.
9. Sample must be collected prior to dosing of injection on Day 1.
10. The nasopharyngeal or nasal swab sample will be used to ascertain the presence of SARS-CoV-2 via RT-PCR.
11. An unscheduled visit may be prompted by reactogenicity issues, illness visit criteria for Covid-19, or new or ongoing AEs. If a participant meets the prespecified criteria of suspicion for COVID-19 (Section 7.1.6), the participants will be asked to return within 72 hours or as soon as possible to the study site for an unscheduled illness visit, to include an NP or nasal swab sample (for RT-PCR testing) and other clinical evaluations. If a study site visit is not possible, a home visit may be arranged to collect the NP or nasal sample and conduct clinical evaluations. If a home visit is not possible, the participant will be asked to submit a saliva sample to the study site by a Sponsor-approved method. At this illness visit, the NP or nasal swab samples will be collected to evaluate for the presence of SARS-CoV-2 infection. NP or nasal swab samples will also be tested for the

presence of other respiratory pathogens. In addition, the study site may collect an additional NP or nasal sample for SARS-CoV-2 testing to be able to render appropriate medical care for the study participant as determined by local standards of care. Additionally, clinical information will be carefully collected to evaluate the severity of the clinical case.

12. A convalescent visit will be scheduled approximately 28 days (+7 days) after diagnosis. At this visit, an NP or nasal swab sampling for viral PCR and a blood sample will be collected for potential immunologic assessment of SARS-CoV-2 infection.

13. Diary entries will be recorded by the participant at approximately 30 minutes after injection while at the study site with instruction provided by study staff. Study participants will continue to record entries in the eDiary each day after they leave the study site, preferably in the evening, on the day of injection and for 6 days following injection. If a solicited local or systemic AR continues beyond Day 7 after vaccination, the participant will be prompted to capture details of the solicited local or systemic AR in the eDiary until the AR is resolves or the next IP injection occurs, whichever occurs first; capture of details of ARs in the eDiary should not exceed 28 days after each vaccination. Adverse reactions recorded in eDiaries beyond Day 7 should be reviewed either via telephone call or at the following study visit.

14. Trained study site personnel will call all participants to collect information relating to any AEs, MAAEs, SAEs, AEs leading to study withdrawal, information on concomitant medications associated with those events, and any nonstudy vaccinations. In addition, study personnel will collect information on known participant exposure to someone with known COVID-19 or SARS-CoV-2 infection and on participant experience of COVID-19 symptoms.

15. All concomitant medications and nonstudy vaccinations will be recorded through 28 days after each injection; all concomitant medications relevant to or for the treatment of an SAE or MAAE will be recorded from Day 1 through the final visit (Day 394).

##### Table 8: Participant Decision Clinic Visit

| | All Participants |
|---|---|
| Return to clinic for Participant Decision Clinic Visit | X |
| Sign revised Informed Consent Form | X |
| Confirm participant’s choice to be unblinded or not to be unblinded | X |
| Confirm participant’s choice to receive open-label mRNA-1273 | X |
| Nasopharyngeal or nasal swab | X |
| Blood for immunologic analysis | X |
| Counsel about public health measures to limit virus spread[1] | X |

1. All participants are counseled about the importance of continuing other public health measures to limit the spread of disease including physical-social distancing, wearing a mask, and hand-washing.

##### Figure 5: Schedule of Part A and Part B Participant Visits During the Part B Open-label Period

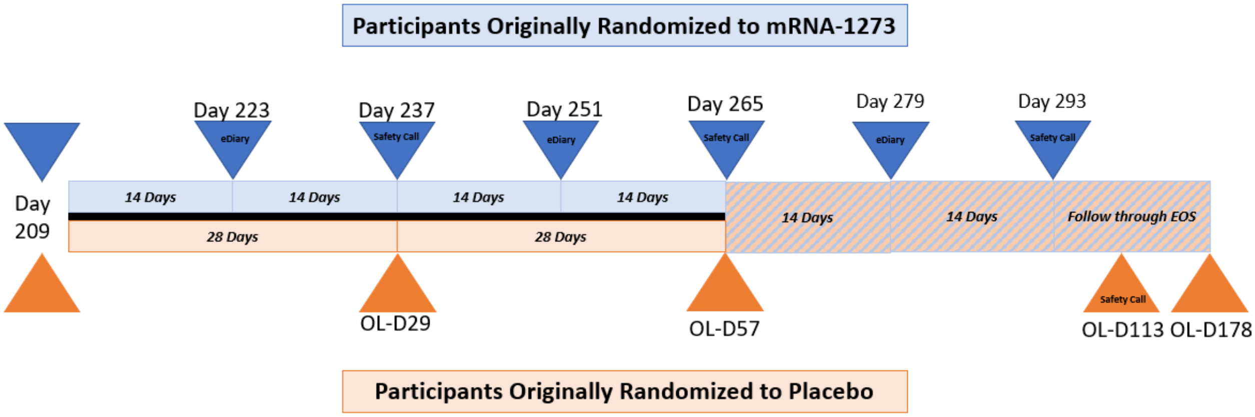

**Table 9: Schedule of Assessments Part B, Open-label Observational Phase for Participants Who Previously Received Placebo**

| **Visit Number** | 7 | 8 | 9 | 10 | 11 |
|---|---|---|---|---|---|
| **Type of Visit** | C | C | C | SC[8] | C |
| **Study Visit Day** | (~D209/ Participant Decision Visit) OL-D1 | OL-D29 | OL-D57[2] | OL-D113 | OL-D178[9] |
| **Window Allowance (Days)** | -56/+56[1] | -3/+7 | ±14 | ± 3 | ±14 |
| **Days Since Most Recent Injection** | 0 | 28 | 56 | 112 | 177 |
| Informed consent form | These assessments are already performed as part of the regular D209 visit. | | | | |
| Blood for vaccine immunogenicity | | | X | | X |
| Nasopharyngeal or nasal swab sample for SARS-CoV-2[3] | | X | X | | |
| Physical examination including vital signs[4] | PE including vitals performed as part of D209 visit. | X | X | | X |
| Physical examination including vital signs[4] (cont.) | X Vitals obtained post dose. | X | | | |
| Pregnancy testing | X | X | | | |
| Study injection (including 30-minute post-dosing observation period) | X | X | | | |
| Informed consent for booster dose | | | X[7] | | |
| Recording of MAAEs and concomitant medications relevant to or for the treatment of the MAAE[5] | X | X | X | X | X |
| Recording of SAEs and concomitant medications relevant to or for the treatment of the SAE[5] | X | X | X | X | X |
| Recording of AESI | X | X | X | X | X |
| Recording of concomitant medications and non-study vaccinations[5] | X | X | X | X | X |
| Surveillance for COVID-19/ Illness visit[6]/ Unscheduled visit | X | X | X | X | X |
| Study completion (for Part B) | | | | | X |

Abbreviations: AE = adverse event; AESI = adverse event of special interest; C = clinic visit; COVID-19 = coronavirus disease 2019; D = Day; M = month; MAAE = medically attended AE; NP = nasopharyngeal; OL = open-label; PCR = polymerase chain reaction; RT-PCR = reverse transcriptase polymerase chain reaction; SAE = serious adverse events; SARS-CoV-2 = Severe Acute Respiratory Syndrome coronavirus 2; SC = safety (telephone) call.

1. The Participant Decision visit may be performed over multiple visits. Once the Participant Decision Visit has been initiated, all assessments must be completed within a 7-day period.

2. After the OL-D57 visit, Part B participants who choose to receive BD will have one additional safety call at OL-D113 prior to entering Part C.

3. The NP or nasal swab sample, collected prior to vaccination on days of injection, will be used to ascertain the presence of SARS-CoV-2 via PCR.

4. Physical examination: A symptom-directed physical examination will be performed at OL-D1, OL-D29, OL-D57, and OL-D178. Symptom-directed physical examinations may be performed at other time points at the discretion of the investigator. Any clinically significant finding identified during a study visit should be reported as an MAAE. Vital signs are to be collected pre- and post-dosing on days of injection (OL-D1 and OL-D29). When applicable, vital sign measurements should be performed before blood collection. Participants who are febrile (body temperature ≥ 38.0°C/100.4°F) before injection on OL-D1 or OL-D29 must be rescheduled within the relevant window period to receive the injection. Afebrile participants with minor illnesses can be administered investigational product at the discretion of the investigator.

5. All concomitant medications and non-study vaccinations will be recorded through 28 days post-injection; all concomitant medications relevant to or for the treatment of an SAE or MAAE will be recorded from Day 1 through OL-D113 before entering Part C. If a participant declines a BD, all concomitant medications relevant to or for the treatment of an SAE or MAAE will be recorded from Day 1 through OL-D178.

6. If a participant meets the prespecified criteria of suspicion for COVID-19 (Section 7.1.6), the participants will be asked to return within 72 hours or as soon as possible to the study site for an unscheduled illness visit, to include an NP or nasal swab sample (for RT-PCR testing) and other clinical evaluations. If a study site visit is not possible, a home visit may be arranged to collect the NP or nasal sample and conduct clinical evaluations. If a home visit is not possible, the participant will be asked to submit a saliva sample to the study site by a Sponsor-approved method. At this illness visit, the NP or nasal swab samples will be collected to evaluate for the presence of SARS-CoV-2 infection. NP or nasal swab samples will also be tested for the presence of other respiratory pathogens. In addition, the study site may collect an additional NP or nasal sample for SARS-CoV-2 testing to be able to render appropriate medical care for the study participant as determined by local standards of care. Additionally, clinical information will be carefully collected to evaluate the severity of the clinical case.

7. During OL-D57, all participants will be given the option to receive a BD. Participants will choose whether to receive the BD or not and sign the consent form. Those who decline BD will continue with Part B. Those who choose to receive a BD will have one safety call prior to entering Part C.

8. All participants in Part B who choose to receive BD will have one additional safety call at OL-D113 prior to entering Part C.

9. OL-D178 is only for those who decline BD. Participants who choose to receive a BD will follow Part C SoA after the OL-D113 safety call.

#### Table 10: Part C – Booster Dose Phase

| **Visit Number** | BD-1 | BD-1a | | BD-2 | BD-3 | | BD-4 |
|---|---|---|---|---|---|---|---|
| **Type of Visit** | C | C | SC[11] | C | C | SC | C |
| **Study Visit Day** | BD-D1[1] (D209: mRNA-1273; D394:placebo-mRNA-1273) | BD-D4 | 7, 14, and 21 days after BD-D1 (BD-D8, BD-D15, BD-D22) | BD-D29 | BD-D181 (Day 390: mRNA-1273; Day 571: placebo-mRNA-1273) | 270 days after BD-1 (BD-D271) | BD-D361 (Day 571 mRNA-1273; Day 751 placebo-mRNA-1273) |
| **Window Allowance (Days)** | - | -2 | +3 | -3/+14 | -3/+14 | +3 | -3/+14 |
| **Days Since Most Recent Vaccination (in Part C)** | 0 | 3 | 7, 14, 21 | 28 | 180 | 270 | 360 |
| Confirm informed consent form signing | X | | | | | | |
| Physical examination[2] | X | | | X | X | | X |
| Pregnancy testing[3] | X | | | | | | |
| **Immunogenicity Assessment** | | | | | | | |
| Blood for immunologic analysis[4] | X | | | X | X | | X |
| **Biomarker Assessment** | | | | | | | |
| Blood sample for potential biomarker analysis[5] | | X | | | | | |
| **Dosing** | | | | | | | |
| Study injection (including 30-minute post-dosing observation period[6]) | X | | | | | | |
| **Efficacy Assessment** | | | | | | | |
| Surveillance for COVID-19/Unscheduled Visit[7] | X | | X | X | X | X | X |
| Nasal swab[8] | X | | | | | | |

| **Visit Number** | BD-1 | BD-1a | | BD-2 | BD-3 | | BD-4 |
|---|---|---|---|---|---|---|---|
| **Type of Visit** | C | C | SC[11] | C | C | SC | C |
| **Study Visit Day** | BD-D1[1] (D209: mRNA-1273; D394:placebo-mRNA-1273) | BD-D4 | 7, 14, and 21 days after BD-D1 (BD-D8, BD-D15, BD-D22) | BD-D29 | BD-D181 (Day 390: mRNA-1273; Day 571: placebo-mRNA-1273) | 270 days after BD-1 (BD-D271) | BD-D361 (Day 571 mRNA-1273; Day 751 placebo-mRNA-1273) |
| **Window Allowance (Days)** | - | -2 | +3 | -3/+14 | -3/+14 | +3 | -3/+14 |
| **Days Since Most Recent Vaccination (in Part C)** | 0 | 3 | 7, 14, 21 | 28 | 180 | 270 | 360 |
| **Safety Assessments** | | | | | | | |
| Follow-up safety[9] | | | X | | | X | X |
| eDiary activation for recording solicited ARs (7 days)[10] | X | | | | | | |
| Review of eDiary data[10] | | | X[10(BD-D8)] | | | | |
| Recording of unsolicited AEs[11] | X | X | X | X | | | |
| Recording of MAAEs and concomitant medications relevant to or for the treatment of the MAAE[12] | X | X | X | X | X | X | X |
| Recording of AE leading to withdrawal [12] | X | X | X | X | X | X | X |
| Recording of AESIs[12] | X | X | X | X | X | X | X |
| Recording of SAEs and concomitant medications relevant to or for the treatment of the SAE[12] | X | X | X | X | X | X | X |
| Recording of concomitant medications and non-study vaccinations[12] | X | X | X | X | X | X | X |

Abbreviations: AE = adverse event; AESI = adverse event of special interest; AR = adverse reaction; BD = booster dose; C = clinic visit; D = day; eDiary = electronic diary; ICF = informed consent form; MAAE = medically attended AE; NP = nasopharyngeal; SAE = serious adverse event; SC = safety (phone) call.
Note: In accordance with FDA Guidance on Conduct of Clinical Trials of Medical Products during COVID-19 Public Health Emergency (DHHS 2020), investigators may convert study site visits to home visits or telemedicine visits with the approval of the Sponsor.

1. A BD may be administered to all participants who are currently enrolled in Part A or Part B provided there are no current contraindications for further dosing (Section 6). A participant who is currently in the Convalescent Period may come in for a BD-1 visit and receive a BD as long they are no longer symptomatic, and this includes the possibility for a Convalescent Visit Day 28 to overlap and be combined with a BD-1 visit.
2. Symptom-directed physical examination will be performed at the BD-Day 1. On dosing day before injection, the arm receiving the injection should be examined and the associated lymph nodes should be evaluated. At visits BD-2 (BD-D29), BD-3 (BD-D181) and BD-4 (BD-D361), a symptom-directed physical examination may be performed at the discretion of the investigator. Any clinically significant finding identified during a study visit should be reported as a MAAE. Vital signs are to be collected pre- and post-dosing (participant will be seated for at least 5 minutes before all measurements are taken per Section 7.1.4) on the day of injection (BD-D1). Participants who are febrile (body temperature ≥ 38.0°C/100.4°F) before dosing (BD-D1) must be rescheduled to receive the injection. Afebrile participants with minor illnesses can be vaccinated at the discretion of the investigator.
3. The pregnancy test at the BD-1 visit will be a point-of-care urine test. At the discretion of the investigator, a pregnancy test either via blood or point-of-care urine test can be performed.
4. Sample should be collected prior to dosing on BD-D1.
5. All participants who chose to receive a BD. Serum sample from two ~4 mL blood draws. Biomarker plasma and biomarker serum samples will be stored for potential future biomarker assessment.
6. Post-dosing, participants will have a 30-minute observation period.
7. An unscheduled visit may be prompted by reactogenicity issues, illness visit criteria for COVID-19, or new or ongoing AEs. If a participant meets the prespecified criteria of suspicion for COVID-19 (Section 7.1.6), the participants will be asked to return within 72 hours or as soon as possible to the study site for an unscheduled illness visit, to include an NP or nasal swab sample (for RT-PCR testing) and other clinical evaluations. If a study site visit is not possible, a home visit may be arranged to collect the nasal swab sample and conduct clinical evaluations. If a home visit is not possible, the participant will be asked to submit a saliva sample to the study site by a Sponsor-approved method. At this illness visit, the NP or nasal swab samples will be collected to evaluate for the presence of SARS-CoV-2 infection. Nasal swab samples will also be tested for the presence of other respiratory pathogens. In addition, the study site may collect an additional nasal swab sample for SARS-CoV-2 testing to be able to render appropriate medical care for the study participant as determined by local standards of care. Additionally, clinical information will be carefully collected to evaluate the severity of the clinical case.
8. The nasal swab sample must be collected prior to injection at the BD-1 visit.
9. Trained study personnel will call all participants to collect information relating to any unsolicited AEs, MAAEs (including any signs and symptoms of COVID-19), AESIs, AEs leading to withdrawal, SAEs, information on concomitant medications associated with those events, and any non-study vaccinations.
10. Diary entries will be recorded by the participant at approximately 30 minutes after injection while at the study site with instruction provided by study staff. Study participants will continue to record entries in the eDiary each day after they leave the study site, preferably in the evening, on the day of injection and for 6 days following injection. If a solicited local or systemic AR continues beyond Day 7 after vaccination, the participant will be

prompted to capture details of the solicited local or systemic AR in the eDiary until the AR resolves; capture of details of ARs in the eDiary should not exceed 28 days after vaccination. Adverse reactions recorded in eDiaries beyond Day 7 should be reviewed either via telephone call or at the following study visit. Review of eDiary will occur on BD-D8.

11. Only for participants who chose to receive a BD.
12. All concomitant medications relevant to or for the treatment of an SAE or MAAE will be recorded from Screening through the final visit.

#### 10.2. APPENDIX 2: Study Governance Considerations

##### 10.2.1. Regulatory and Ethical Considerations

This study will be conducted in accordance with the protocol and with the following:

- Consensus ethical principles derived from international guidelines including the Declaration of Helsinki and Council for International Organizations of Medical Sciences International Ethical Guidelines.
- Applicable ICH GCP Guidelines.
- Applicable laws and regulatory requirements.
- The protocol, protocol amendments, ICF, IB, and other relevant documents (eg, advertisements) must be submitted to an IRB by the investigator and reviewed and approved by the IRB before the study is initiated.
- Any amendments to the protocol will require IRB approval before implementation of changes made to the study design, except for changes necessary to eliminate an immediate hazard to study participants.
- The investigator will be responsible for the following:
  - Providing written summaries of the status of the study to the IRB annually or more frequently in accordance with the requirements, policies, and procedures established by the IRB
  - Notifying the IRB of SAEs or other significant safety findings as required by IRB procedures
  - Providing oversight of the conduct of the study at the site and adherence to requirements of 21 CFR, ICH guidelines, the IRB, European regulation 536/2014 for clinical studies (if applicable), and all other applicable local regulations.

##### 10.2.2. Study Monitoring

Before an investigational site can enter a participant into the study, a representative of the Sponsor or its representatives will visit the investigational study site to do the following:

- Determine the adequacy of the facilities
- Discuss with the investigator(s) and other personnel their responsibilities with regard to protocol adherence, and the responsibilities of the Sponsor or its representatives. This

will be documented in a clinical study agreement between the Sponsor, the designated CRO, and the investigator.

According to ICH GCP guidelines, the Sponsor of the study is responsible for ensuring the proper conduct of the study with regard to protocol adherence and validity of data recorded on the eCRFs. The study monitor's duties are to aid the investigator and the Sponsor in the maintenance of complete, accurate, legible, well-organized, and easily retrievable data. The study monitor will advise the investigator of the regulatory necessity for study-related monitoring, audits, IRB review, and inspection by providing direct access to the source data/documents. In addition, the study monitor will explain to and interpret for the investigator all regulations applicable to the clinical evaluation of an IP as documented in ICH guidelines.

It is the study monitor's responsibility to inspect the eCRFs and source documentation throughout the study to protect the rights of the participants; to verify adherence to the protocol; to verify completeness, accuracy, and consistency of the data; and to confirm adherence of study conduct to any local regulations. Details will be outlined in the Clinical Monitoring Plan. During the study, a monitor from the Sponsor or a representative will have regular contacts with the investigational site, for the following:

- Provide information and support to the investigator(s).
- Confirm that facilities remain acceptable.
- Confirm that the investigational team is adhering to the protocol, that the data are being accurately recorded in the eCRFs, and that IP accountability checks are being performed.
- Perform source data verification. This includes a comparison of the data in the eCRFs with the participant's medical records at the hospital or practice, and other records relevant to the study. This will require direct access to all original records for each participant (eg, clinical charts or electronic medical record system).
- Record and report any protocol deviations not previously sent.
- Confirm AEs and SAEs have been properly documented on eCRFs and confirm any SAEs have been forwarded to the SAE Hotline, and those SAEs that met criteria for reporting have been forwarded to the IRB.

The monitor will be available between visits if the investigator(s) or other staff needs information or advice.

##### 10.2.3. Audits and Inspections

The Sponsor, their designee(s), the IRB, or regulatory authorities will be allowed to conduct study site visits to the investigational facilities for the purpose of monitoring or inspecting any aspect of the study. The investigator agrees to allow the Sponsor, their designee(s), the IRB, or regulatory authorities to inspect the IP storage area, IP stocks, IP records, participant charts and study source documents, and other records relative to study conduct.

Authorized representatives of the Sponsor, a regulatory authority, and any IRB may visit the study site to perform audits or inspections, including source data verification. The purpose of a Sponsor audit or inspection is to systematically and independently examine all study-related activities and documents to determine whether these activities were conducted, and data were recorded, analyzed, and accurately reported according to the protocol, ICH E6(R2) GCP, and any applicable regulatory requirements. The investigator should contact the Sponsor immediately if contacted by a regulatory agency about an inspection.

The principal investigator must obtain IRB approval for the investigation. Initial IRB approval, and all materials approved by the IRB for this study including the participant consent/assent form and recruitment materials, must be maintained by the investigator and made available for inspection.

##### 10.2.4. Financial Disclosure

The investigator is required to provide financial disclosure information to allow the Sponsor to submit the complete and accurate certification or disclosure statements required under 21 CFR 54. In addition, the investigator must provide the Sponsor with a commitment to promptly update this information if any relevant changes occur during the course of the investigation and for 1 year following the completion of the study.

The Sponsor, the CRO, and the study site are not financially responsible for further testing or treatment of any medical condition that may be detected during the screening process. In addition, in the absence of specific arrangements, the Sponsor, the CRO, and the study site are not financially responsible for further treatment of the disease under study.

##### 10.2.5. Recruitment Procedures

Advertisements to be used for the recruitment of study participants, and any other written information regarding this study to be provided to the participant should be submitted to the Sponsor for approval. All documents must be approved by the IRB.

##### 10.2.6. Informed Consent/Assent Process

The informed consent/assent document(s) must meet the requirements of 21 CFR 50, local regulations, ICH guidelines, Health Insurance Portability and Accountability Act, where applicable, and the IRB or study site. All consent documents will be approved by the appropriate IRB. The actual ICF used at each study site may differ, depending on local regulations and IRB requirements. However, all versions of the ICF must contain the standard information found in the sample ICF provided by the Sponsor. Any change to the content of the ICF must be approved by the Sponsor and the IRB prior to the ICF being used.

If new information becomes available that may be relevant to the participant's willingness to continue participation in the study, this will be communicated to them in a timely manner. Such information will be provided via a revised ICF or an addendum to the original ICF.

The investigator or his/her representative will explain the nature of the study to the participant and answer all questions regarding the study.

The investigator is responsible for ensuring that the participant fully understands the nature and purpose of the study. Information should be given in both oral and written form whenever possible.

No participant should be obliged to participate in the study. The participant must be informed that participation is voluntary. Participants, their relatives, guardians, or (if applicable) LARs must be given ample opportunity to inquire about details of the study. The information must make clear that refusal to participate in the study or withdrawal from the study at any stage is without any prejudice to the participant's subsequent care.

The participant must be allowed sufficient time to decide whether they wish to participate in the study.

The participant must be made aware of, and give consent to, direct access to his/her source medical records by study monitors, auditors, the IRB, and regulatory authorities. The participant should be informed that such access will not violate participant confidentiality or any applicable regulations. The participant and/or participants' parent(s)/LAR(s) should also be informed that he/she is authorizing such access by signing the ICF.

A copy of the ICF(s) must be provided to the participant and/or participants' parent(s)/LAR(s).

A participant who is rescreened is not required to sign another ICF if the rescreening occurs within 28 days from the previous ICF signature date (within the initial Screening Period).

The ICF will also explain that excess serum from immunogenicity testing may be used for future research, which may be performed at the discretion of the Sponsor to further characterize the immune response to SARS-CoV-2, additional assay development, and the immune response across CoVs.

##### 10.2.7. Protocol Amendments

No change or amendment to this protocol may be made by the investigator or the Sponsor after the protocol has been agreed to and signed by all parties unless such change(s) or amendment(s) has (have) been agreed upon by the investigator or the Sponsor. Any change agreed upon will be recorded in writing, and the written amendment will be signed by the investigator and the Sponsor. IRB approval is required prior to the implementation of an amendment, unless overriding safety reasons warrant immediate action, in which case the IRB(s) will be promptly notified.

Any modifications to the protocol or the ICF, which may impact the conduct of the study, potential benefit of the study, or may affect participant safety, including changes of study objectives, study design, participant population, sample sizes, study procedures, or significant administrative aspects will require a formal amendment to the protocol. Such an amendment will be released by the Sponsor, agreed by the investigator(s), and approved by the relevant IRB(s) prior to implementation. A signed and dated statement that the protocol, any subsequent relevant amended documents, and the ICF have been approved by relevant IRB(s) must to be provided to the Sponsor before the study is initiated.

Administrative changes of the protocol are minor corrections and/or clarifications that have no effect on the way the study is to be conducted. These administrative changes will be released by the Sponsor, agreed by the investigators, and notified to the IRB(s).

##### 10.2.8. Protocol Deviations

The noncompliance may be either on the part of the participant, the investigator, or the study site staff. As a result of protocol deviations, corrective actions are to be developed by the study site and implemented promptly.

After a participant proceeds to the Participant Decision Visit (Part A) of the study, participants who received mRNA-1273 will continue to follow the open-label Part A SoA. Participants who received placebo will transition to open-label Part B of the study and will follow the Part B SoA (Table 9). Participants who transitioned to Part C, 6 months after last dose, will follow the Part C SoA (Table 10).

It is the responsibility of the study site investigator to use continuous vigilance to identify and report protocol deviations to the Sponsor or its designee. All protocol deviations must be addressed in study source documents, reported to study monitor. Protocol deviations must be sent to the reviewing IRB per their policies. The study site investigator is responsible for knowing and adhering to the reviewing IRB requirements.

##### 10.2.9. Data Protection

Participants will be assigned a unique identifier by the Sponsor. Any participant records or datasets that are transferred to the Sponsor will contain the identifier only; participant names or any information which would make the participant identifiable will not be transferred.

The participant must be informed that his/her personal study-related data will be used by the Sponsor in accordance with local data protection law. The level of disclosure must also be explained to the participant.

The participant must be informed that his/her medical records may be examined by clinical quality assurance (QA) auditors or other authorized personnel appointed by the Sponsor, by appropriate IRB members, and by inspectors from regulatory authorities.

Individual participant medical information obtained as a result of this study is considered confidential, and disclosure to third parties is prohibited. Information will be accessible to authorized parties or personnel only. Medical information may be given to the participant's physician or to other appropriate medical personnel responsible for the participant's well-being. Each participant will be asked to complete a form allowing the investigator to notify the participant's primary health care provider of his/her participation in this study.

All laboratory specimens, evaluation forms, reports, and other records will be identified in a manner designed to maintain participant confidentiality. All records will be kept in a secure storage area with limited access. Clinical information will not be released without the written permission of the participant, except as necessary for monitoring and auditing by the Sponsor, its designee, relevant regulatory authority, or the IRB.

The investigator and all employees and coworkers involved with this study may not disclose or use for any purpose other than performance of the study, any data, record, or other unpublished, confidential information disclosed to those individuals for the purpose of the study. Prior written agreement from the Sponsor or its designee must be obtained for the disclosure of any confidential information to other parties.

##### 10.2.10. Sample Retention and Future Biomedical Research

The retention period of laboratory samples will be 20 years, or as permitted by local regulations, to address further scientific questions related to mRNA-1273 or antirespiratory virus immune response. In addition, identifiable samples can be destroyed at any time at the request of the participant. During the study, or during the retention period, in addition to the analysis outlined in the study endpoints, exploratory analysis may be conducted using other Ab-based methodologies on any remaining blood or serum samples, including samples from participants who are screened but are not subsequently enrolled. These analyses will extend the search for other potentially relevant biomarkers to investigate the effect of mRNA-1273, as well as to determine how changes in biomarkers may relate to exposure and clinical outcomes. A decision to perform such exploratory research may arise from new scientific findings related to the drug class or disease, as well as reagent and assay availability.

###### 10.2.11. Dissemination of Clinical Study Data

The Sponsor shares information about clinical trials and results on publicly accessible websites, based on international and local legal and regulatory requirements, and other clinical trial disclosure commitments established by pharmaceutical industry associations. These websites include clinicaltrials.gov, EU clinical trial register (eu.ctr), as well as some national registries.

In addition, results from clinical trials are required to be submitted to peer-reviewed journals following internal company review for accuracy, fair balance, and intellectual property. For those journals that request sharing of the analyzable data sets that are reported in the publication, interested researchers are directed to submit their request to clinicalstudydatarequest.com.

Individual participant data and supporting clinical documents are available for request at clinicalstudydatarequest.com. While making information available, the privacy of participants in clinical studies sponsored by the Sponsor is assured. Details on data sharing criteria and process for requesting access can be found at this web address: clinicalstudydatarequest.com.

###### 10.2.12. Data Quality Assurance and Quality Control

Data collection is the responsibility of the clinical study staff at the study site under the supervision of the study site investigator. The investigator is responsible for ensuring the accuracy, completeness, legibility, and timeliness of the data reported.

- All participant data relating to the study will be recorded in the eCRF unless transmitted to the Sponsor or designee electronically (eg, laboratory data). The investigator is responsible for verifying that data entries are accurate and correct by physically or electronically signing the eCRF.
- The investigator must maintain accurate documentation (source data) that supports the information entered in the eCRF.

- The investigator must permit study-related monitoring, audits, IRB review, and regulatory agency inspections and provide direct access to source data documents.
- Monitoring details describing strategy (eg, risk-based initiatives in operations and quality such as Risk Management and Mitigation Strategies and Analytical Risk-Based Monitoring), methods, responsibilities and requirements, including handling of noncompliance issues and monitoring techniques (central, remote, or on-site monitoring) are provided in the Clinical Monitoring Plan.
- The Sponsor or designee is responsible for the data management of this study including quality checking of the data.
- The Sponsor assumes accountability for actions delegated to other individuals (eg, CRO).
- Study monitors will perform ongoing source data verification to confirm that data entered into the CRF by authorized study site personnel are accurate, complete, and verifiable from source documents; that the safety and rights of participants are being protected; and that the study is being conducted in accordance with the currently approved protocol and any other study agreements, ICH GCP, and all applicable regulatory requirements.
- Records and documents, including signed ICFs, pertaining to the conduct of this study must be retained by the investigator for a period of at least 2 years after the last marketing application approval or, if not approved, 2 years following the discontinuance of the test article for investigation. If this requirement differs from any local regulations, the local regulations will take precedence unless the local retention policy is less than 2 years. No records may be destroyed during the retention period without the written approval of the Sponsor. No records may be transferred to another location or party without written notification to the Sponsor.

Quality assurance includes all the planned and systematic actions that are established to ensure that the clinical study is performed and the data are generated, documented (recorded), and reported according to ICH GCP and local/regional regulatory standards.

A QA representative from the Sponsor or a qualified designee, who is independent of and separated from routine monitoring, may periodically arrange inspections/audits of the clinical study by reviewing the data obtained and procedural aspects. These inspections may include on-site inspections/audits and source data checks. Direct access to source documents is required for the purpose of these periodic inspections/audits.

##### 10.2.13. Data Collection and Management

This study will be conducted in compliance with ICH CGP guidelines. This study will also be conducted in accordance with the most recent version of the Declaration of Helsinki.

This study will use electronic data collection to collect data directly from the study site using eCRFs. The investigator is responsible for ensuring that all sections of each eCRF are completed promptly and correctly and that entries can be verified against any source data.

Study monitors will perform source document verification to identify inconsistencies between the eCRFs and source documents. Discrepancies will be resolved in accordance with the principles of GCP. Detailed study monitoring procedures are provided in the Clinical Monitoring Plan.

Adverse events will be coded with MedDRA. Concomitant medications will be coded using WHO - Drug Dictionary.

##### 10.2.14. Source Documents

Source documents are original documents or certified copies, and include, but are not limited to, eDiaries, medical and hospital records, screening logs, informed consent/assent forms, telephone contact logs, and worksheets. Source documents provide evidence for the existence of the participant and substantiate the integrity of the data collected. Source documents are filed at the investigator’s study site.

Data reported on the CRF or entered in the eCRF that are transcribed from source documents must be consistent with the source documents or the discrepancies must be explained. The investigator may need to request previous medical records or transfer records, depending on the study. Also, current medical records must be available.

The Sponsor or its designee requires that the investigator prepare and maintain adequate and accurate records for each participant treated with the IP. Source documents such as any hospital, clinic, or office charts, and the signed ICFs are to be included in the investigator’s files with the participant’s study records.

##### 10.2.15. Retention of Records

The principal investigator must maintain all documentation relating to the study for a period of at least 2 years after the last marketing application approval or, if not approved, 2 years following the discontinuance of the test article for investigation. If this requirement differs from any local regulations, the local regulations will take precedence unless the local retention policy is > 2 years.

If it becomes necessary for the Sponsor or the regulatory authority to review any documentation relating to the study, the investigator must permit access to such records. No records will be destroyed without the written consent of the Sponsor, if applicable. It is the responsibility of the Sponsor to inform the investigator when these documents no longer need to be retained.

##### 10.2.16. Study and Site Closure

If the study is prematurely terminated or suspended, the Sponsor shall promptly inform the investigators, the IRBs, the regulatory authorities, and any CRO(s) used in the study of the reason for termination or suspension, as specified by the applicable regulatory requirements. The investigator shall promptly inform the participants and should assure appropriate participant therapy and/or follow-up.

The Sponsor or designee reserves the right to close the study site or terminate the study at any time for any reason at the sole discretion of the Sponsor.

The investigator may initiate study site closure at any time, provided there is reasonable cause and sufficient notice is given in advance of the intended termination.

Reasons for the early closure of a study site by the Sponsor or investigator may include but are not limited to:

- Continuation of the study represents a significant medical risk to participants
- Failure of the investigator to comply with the protocol, the requirements of the IRB or local health authorities, the Sponsor’s procedures, or GCP guidelines
- Inadequate recruitment of participants by the investigator
- Discontinuation of further mRNA-1273 development

Study sites will be closed upon study completion. A study site is considered closed when all required documents and study supplies have been collected and a study site closure visit has been performed.

##### 10.2.17. Publication Policy

The results of this study may be published or presented at scientific meetings. If this is foreseen, the investigator agrees to submit all manuscripts or abstracts to the Sponsor before submission. This allows the Sponsor to protect proprietary information and to provide comments.

The Sponsor will comply with the requirements for publication of study results. In accordance with standard editorial and ethical practice, the Sponsor will generally support publication of multicenter studies only in their entirety and not as individual study site data. In this case, a coordinating investigator will be designated by mutual agreement.

Authorship will be determined by mutual agreement and in line with International Committee of Medical Journal Editors authorship requirements.

The clinical study plan and the results of the study will be published on www.ClinicalTrials.gov in accordance with 21 CFR 50.25(c). The results of and data from this study belong to the Sponsor.

##### 10.2.18. Body Mass Index (BMI) Charts for Boys and Girls

**For boys aged 5 through 19 years:**

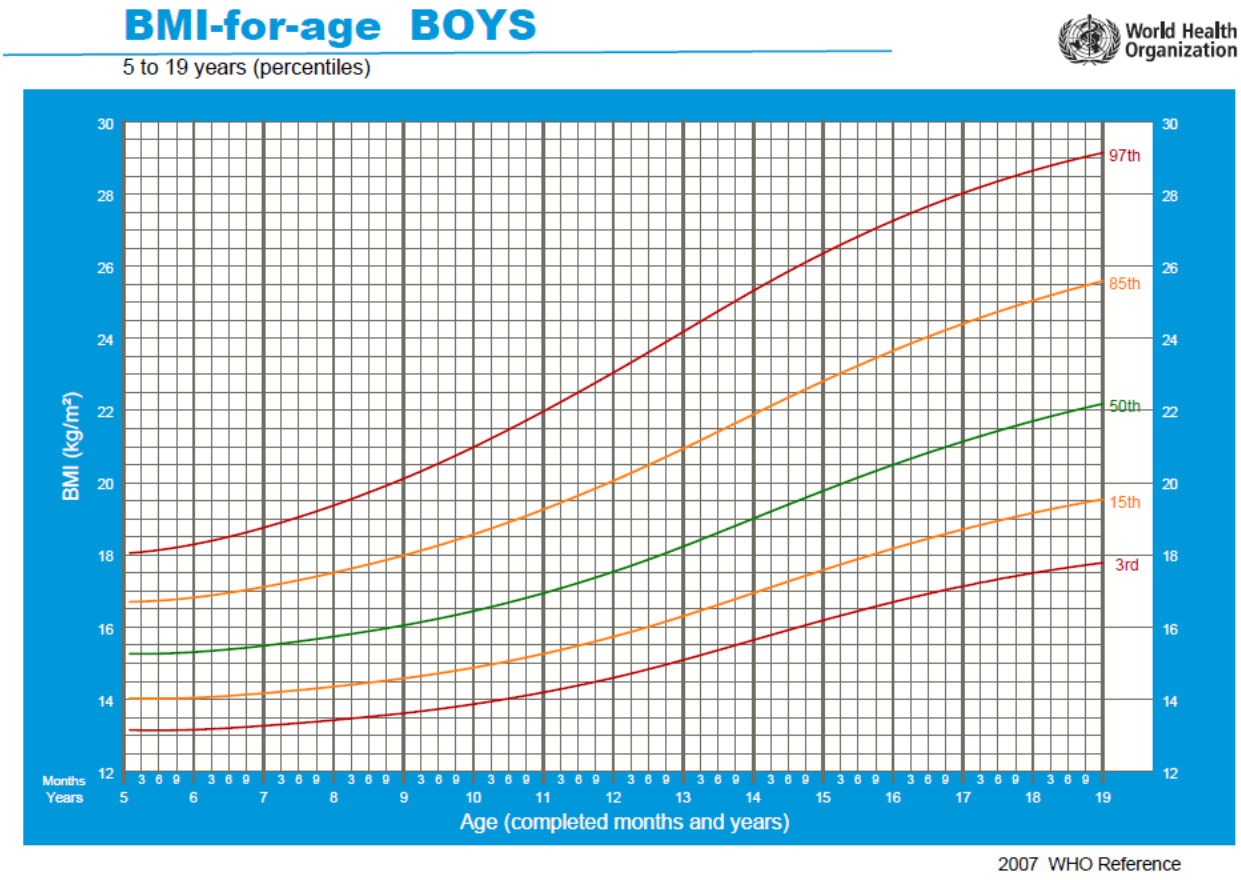

**For girls aged 5 through 19 years:**

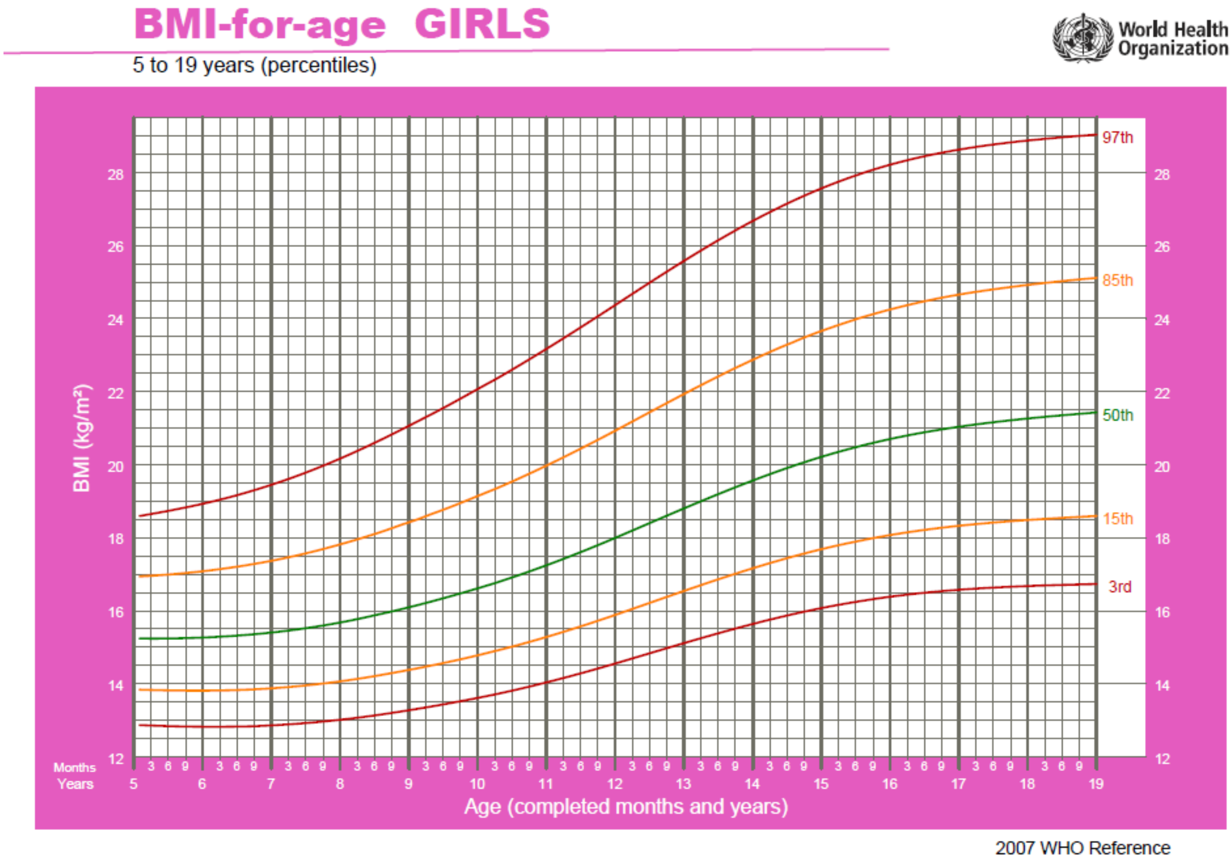

#### 10.3. APPENDIX 3: Contraceptive Guidance

**Woman of Childbearing Potential (WOCBP)**

Females of childbearing potential are those who are considered fertile following menarche and until becoming postmenopausal unless permanently sterile (see below). If fertility is unclear (eg, amenorrhea in adolescents or athletes) and a menstrual cycle cannot be confirmed before first dose of study treatment, additional evaluation should be considered.

Women in the following categories are not considered WOCBP:

1. Premenarchal
2. Premenopausal, surgically sterile female with 1 of the following:
   a. Documented complete hysterectomy
   b. Documented surgical sterilization

   For individuals with permanent infertility due to an alternate medical cause other than the above, (eg, Mullerian agenesis, androgen insensitivity), investigator discretion should be applied in determining study entry.

   Note: Documentation can come from the study site personnel's review of the participant's medical records, medical examination, or medical history interview.

**Contraception Guidance:**

Adequate female contraception is defined as consistent and correct use of an FDA-approved contraceptive method in accordance with the product label. For example:

- Barrier method (such as condoms, diaphragm, or cervical cap) used in conjunction with spermicide
- Intrauterine device
- Prescription hormonal contraceptive taken or administered via oral (pill), transdermal (patch), subdermal, or IM route
- Sterilization of a female participant's monogamous male partner prior to entry into the study

Note that periodic abstinence (eg, calendar, ovulation, symptothermal, postovulation methods) and withdrawal are not acceptable methods of contraception.

#### 10.4. APPENDIX 4: Adverse Event of Special Interest Terms

Investigators should report all events which fall into the following categories as an AESI per the reporting processes specified in Section 7.5.5. The following AESIs are medical concepts that may be related to COVID-19 or are of interest in COVID-19 vaccine safety surveillance. Even if the events below occur in the setting of a COVID infection, the event should still be reported as an AESI if it is one of the medical concepts below.

| | |
|---|---|
| **Anosmia, Ageusia** | • New onset COVID associated or idiopathic events without other etiology excluding congenital etiologies or trauma |
| **Subacute thyroiditis** | • Including but not limited to events of: atrophic thyroiditis, autoimmune thyroiditis, immune-mediated thyroiditis, silent thyroiditis, thyrotoxicosis and thyroiditis |
| **Acute pancreatitis** | • Including but not limited to events of: autoimmune pancreatitis, immune-mediated pancreatitis, ischemic pancreatitis, edematous pancreatitis, pancreatitis, acute pancreatitis, hemorrhagic pancreatitis, necrotizing pancreatitis, viral pancreatitis, and subacute pancreatitis<br>• Excluding known etiologic causes of pancreatitis (alcohol, gallstones, trauma, recent invasive procedures) |
| **Appendicitis** | • Include any event of appendicitis |
| **Rhabdomyolysis** | • New onset rhabdomyolysis without known etiology such as excessive exercise or trauma |
| **Acute respiratory distress syndrome (ARDS)** | • Including but not limited to new events of ARDS and respiratory failure. |
| **Coagulation disorders** | • Including but not limited to thromboembolic and bleeding disorders, disseminated intravascular coagulation, pulmonary embolism, deep vein thrombosis |
| **Acute cardiovascular injury** | • Including but not limited to myocarditis, pericarditis, microangiopathy, coronary artery disease, arrhythmia, stress cardiomyopathy, heart failure, or acute myocardial infarction |
| **Acute kidney injury** | • Include events with idiopathic or autoimmune etiologies<br>• Exclude events with clear alternate etiology (trauma, infection, tumor, or iatrogenic causes such as medications or radiocontrast etc)<br>• Include all cases that meet the following criteria<br>○ Increase in serum creatinine by ≥ 0.3 mg/dl (≥26.5 umol/l) within 48 hours;<br>OR |

| | |
|---|---|
| | ○ Increase in serum creatinine to ≥ 1.5 times baseline, known or presumed to have occurred within prior 7 days<br>OR<br>○ Urine volume ≤0.5 ml/ kg/ hour for 6 hours |
| **Acute liver injury** | • Include events with idiopathic or autoimmune etiologies<br>• Exclude events with clear alternate etiology (trauma, infection, tumor, etc)<br>• Include all cases that meet the following criteria<br>○ 3-fold elevation above the upper normal limit for ALT or AST<br>OR<br>○ > 2-fold elevation above the upper normal limit for total serum bilirubin or GGT or ALP |
| **Dermatologic findings** | • Chilblain-like lesions<br>• Single organ cutaneous vasculitis<br>• Erythema multiforme<br>• Bullous rashes<br>• Severe cutaneous adverse reactions including but not limited to: Stevens-Johnson Syndrome (SJS), Toxic Epidermal Necrolysis (TEN), Drug Reaction with Eosinophilia and Systemic Symptoms (DRESS) and fixed drug eruptions |
| **Multisystem inflammatory disorders** | • Multisystem inflammatory syndrome in adults (MIS-A)<br>• Multisystem inflammatory syndrome in children (MIS-C)<br>• Kawasaki's disease |
| **Thrombocytopenia** | • Platelet counts < 150 x10^9<br>• Including but not limited to immune thrombocytopenia, platelet production decreased, thrombocytopenia, thrombocytopenic purpura, thrombotic thrombocytopenic purpura, or HELLP syndrome |
| **Acute aseptic arthritis** | • New onset aseptic arthritis without clear alternate etiology (eg, gout, osteoarthritis, and trauma) |
| **New onset of or worsening of neurologic disease** | • Including but not limited to:<br>○ Guillain-Barre Syndrome<br>○ Acute disseminated encephalomyelitis (ADEM)<br>○ Peripheral facial nerve palsy (Bell's palsy)<br>○ Transverse myelitis<br>○ Encephalitis/Encephalomyelitis<br>○ Aseptic meningitis<br>○ Febrile seizures<br>○ Generalized seizures/convulsions |

| | |
|---|---|
| | ○ Stroke (Hemorrhagic and non-hemorrhagic)<br>○ Narcolepsy |
| **Anaphylaxis** | • Anaphylaxis as defined per protocol.<br>• Follow reporting procedures in protocol Section 7.5.5 |
| **Other syndromes** | • Fibromyalgia<br>• Postural Orthostatic Tachycardia Syndrome<br>• Chronic Fatigue Syndrome (Includes Myalgic encephalomyelitis and Post viral fatigue syndrome)<br>• Myasthenia gravis |

#### 10.5. APPENDIX 5: Protocol Amendment History

**Amendment 2, 27 Jul 2021**

**Main Rationale for the Amendment:**

The main rationale for this amendment is to increase awareness of possible cases and to add a case definition for myocarditis and pericarditis as well as guidance for reporting and assessing suspected cases for this study, given the recent reports in the Emergency Use Authorization (EUA) experience of cases with a temporal association between COVID-19 messenger RNA (mRNA) vaccine administration and signs and symptoms of myocarditis and pericarditis.

The summary of changes table provided below describes the major changes made in Amendment 2 relative to Amendment 1, including the sections modified and the corresponding rationales. The synopsis of Amendment 2 has been modified to correspond to changes in the body of the protocol. Minor grammar and formatting corrections were made throughout the document to enhance clarity and readability, and new references were added in support of the definition of myocarditis and pericarditis (which did not affect the conduct of the study).

**Summary of Major Changes in Protocol Amendment 2:**

| Section # and Name | Description of Change | Brief Rationale |
|---|---|---|
| Title Page, Signature page, Protocol Amendment Summary of Changes, and Header | • Updated the protocol version and date.<br>• Added Protocol Amendment Summary of Changes for Amendment 2. | Updated to reflect the new version and date. |
| Section 1.3.2 (Risks to Study Participation and Risk Mitigation) | Added paragraph on rare reports of myocarditis and pericarditis occurring after vaccination with Moderna coronavirus disease 2019 (COVID-19) vaccine under Emergency Use Authorization in adults 18 years of age and older. | To reflect addendum made to Investigator’s Brochure. |
| Section 7.4.5 (Adverse Events of Special Interest) | Added Center for Disease Control and Prevention (CDC) case definitions for myocarditis and pericarditis. | To provide guidance to the investigators regarding assessing and reporting myocarditis and pericarditis for this study population. |

| | | |
|---|---|---|
| Section 7.5.2 (Independent Cardiac Event Adjudication Committee) | Added new section. | To describe the proposed mechanism to assess risk of myocarditis and pericarditis in the study population (to address Center for Biologics Evaluation and Research request to describe how the risk of myocarditis and pericarditis will be assessed in the study population receiving mRNA-1273). |
| Section 8.1 (Blinding and Responsibility for Analyses) and Section 8.6.1 (Interim Analysis) | Updated these sections to align with the revised Statistical Analysis Plan (SAP) V2.0, which states the interim analysis will be performed for immunogenicity, safety, and efficacy. | To align with the updated SAP. |
| Section 9 (References) | Updated reference list. | To support the text added to the protocol for myocarditis and pericarditis. |
| Appendix 1, Schedule of Assessments, Table 7 and Table 9 | Increased D209 Participant Decision Visit window to -56/+56. | Minus window expanded to accommodate participants whose D209 visit is expected to occur after Emergency Use Authorization is granted in this age group. |
| Appendix 1, Schedule of Assessments, Table 9 | New footnote was added. | To clarify that D209/Participant Decision Visit may occur over multiple visits but must be completed within 7 days. |
| Section 10.4 (Appendix 4: Adverse Event of Special Interest Terms) | Added new appendix to the protocol. | To include AESI list in the protocol instead of as a separate document. |
| Section 10.5 (Appendix 5: Protocol Amendment History) | Moved the Summary of Changes for Amendment 1 to Appendix 5: Protocol Amendment History. | To reflect the Summary of Changes for the new version of the amendment. |

**Amendment 1, 23 Mar 2021**

**Main Rationale for the Amendment:**

The primary rationales for this amendment are as follows:

- To update the primary endpoints and null hypotheses (or success criteria) for establishing immunobridging and assumptions for the sample size calculations, as recommended by Health Authorities.
- Following the authorization of a COVID-19 vaccine under an Emergency Use Authorization (EUA), this study amendment is designed to transition to Part B, the Open-Label Observational Phase. Transitioning the study to Part B, the Open-label Observational Phase permits all ongoing study participants (a) to be informed of the availability and eligibility criteria of any COVID-19 vaccine made available under an EUA and (b) for EUA-eligible study participants, the opportunity to schedule a study visit to know their original group assignment (placebo versus mRNA-1273).
- Part B, the Open-label Observational Phase also provides the opportunity for study participants who previously received placebo to request to receive 2 doses of the mRNA-1273 vaccine.

The summary of changes table provided here describes the major changes made in Amendment 1 relative to the original protocol, including the sections modified and the corresponding rationales. The synopsis of Amendment 1 has been modified to correspond to changes in the body of the protocol.

#### Summary of Major Changes in Protocol Amendment 1:

| Section # and Name | Description of Change | Brief Rationale |
|---|---|---|
| Title Page, Protocol Approval Page, Headers, Protocol Amendment Summary of Changes | Updated the protocol version and date. | To reflect the new version and date of the protocol. |
| Protocol Synopsis – Objectives and Section 2 (Objectives and Endpoints) | The primary endpoints were redefined. Secondary and exploratory endpoints were refined. An additional secondary objective and corresponding endpoint were added. A new exploratory objective and endpoint were added. | If an accepted threshold of protection is not available, using coprimary endpoints based on nAb geometric mean and seroresponse rate (added) in the noninferiority test was recommended by Health Authorities, as it is considered standard in immunobridging. A secondary objective/endpoint to evaluate asymptomatic SARS-CoV-2 infection was added to be separate from the objective/endpoint of SARS-CoV-2 infection in participants with SARS-CoV-2 negative at baseline. An exploratory objective/endpoint was added to evaluate asymptomatic SARS-CoV-2 infection in participants seropositive at baseline. |
| Protocol Synopsis – Overall Study Design, Section 3.1 (General Study Design), Section 3.1.1 (Part A, the Blinded Phase), Section 3.1.2 (Part B, the Open-label Observational Phase), and Section 7 (Study Assessments and Procedures) | The study design was updated to describe the updated crossover design of the study, including Part A (the Blinded Phase) and Part B (the Open-label Observational Phase). | The addition of the crossover design provides the opportunity for study participants to be informed regarding the EUA of mRNA-1273 for any persons under the age of 18 years, be unblinded to their original assignment (mRNA-1273 or placebo), and for those who previously received placebo to actively request to receive 2 doses of mRNA-1273 (100 µg) vaccine. |
| Protocol Synopsis – Study Duration and Section 3.1.1 (Part A, the Blinded Phase) | The study duration was clarified to reflect the differing study duration for participants who were initially randomly assigned to mRNA-1273 versus patients who were initially randomized to placebo but who opt to receive mRNA-1273 after unblinding. | The addition of the crossover design was one of the primary purposes of this amendment. |
| Protocol Synopsis – Inclusion Criteria and Section 4.1.1 (Inclusion Criteria) | Updated to allow abstinence as a contraception option and remove the criteria that participants not be currently breastfeeding | Updated to be consistent with the current standard of care in the United States. |
| Protocol Synopsis – Exclusion Criteria and Section 4.1.2 (Exclusion Criteria) | Updated to remove exclusion of participants with a known history of SARS-CoV-2 infection or | Updated to be consistent with the current standard of care in the United States, and to address the increasing prevalence of |

| Section # and Name | Description of Change | Brief Rationale |
|---|---|---|
| | contact with a confirmed case of SARS-CoV-2 infection. | SARS-CoV-2 infection and make the study more consistent with the "real world" (EUA) setting. |
| Protocol Synopsis – Study Eligibility Criteria (Part B) and Section 4.2 (Study Eligibility Criteria, Part B) | Section added for Part B of the study to specify that participants participating in Part B must have been previously enrolled in the mRNA-1273-P203 study; and that female participants of childbearing potential may enroll in Part B if they have negative pregnancy tests at OL-Day 1 and OL-Day 29. | To establish separate eligibility criteria for Part B of the study. |
| Protocol Synopsis – Procedures and Assessments (Safety Assessments) and Section 7.1 (Safety Assessments and Procedures) | Updated to clarify the collection periods for ARs, AEs leading to discontinuation from dosing and/or study participation, MAAEs, SAEs, and AESIs. | To clarify based on the new crossover design of the study. |
| Protocol Synopsis – Procedures and Assessments (Immunogenicity Assessments) and Section 7.2 (Immunogenicity Assessments) | Updated to note the assessments for Parts A and B of the study, to clarify the analytes to be measured, and to add testing of serum for nAb and bAb against the SARS-CoV-2 S protein. | Revised the assessments and tests plan for the unbinding or participant decision visit and subsequent visits to be used for long-term follow-up analyses. |
| Protocol Synopsis – Procedures and Assessments (Efficacy Assessments) and Section 7.3.1 (Vaccine Effectiveness Assessments) | Updated to refine the Ab response assessment if an accepted threshold of protection is not available. | To define the coprimary endpoints required for the noninferiority test to establish immunobridging. |
| Protocol Synopsis – Statistical Methods (Hypothesis Testing) and Section 8.2 (Statistical Hypothesis) | Updated to redefine the study hypotheses based on updates to the coprimary endpoints. | To specify the hypotheses for the updated coprimary endpoints using noninferiority test. |
| Protocol Synopsis – Statistical Methods (Power and Sample Size) and Section 8.3 (Power and Sample Size) | Updated to reflect increases in the size of the Immunogenicity Subset for the purposes of establishing acceptable noninferiority margins. | To detail the sample size and power calculations for the updated primary endpoints and hypotheses. |
| Protocol Synopsis – Statistical Methods (Analysis Sets) and Section 8.4 (Analysis Sets) | Added mITT and miTT1 analysis sets. | To define mITT and mITT1 to be used in the sensitivity analyses of secondary efficacy endpoints. |
| Protocol Synopsis – Statistical Methods (Immunogenicity Analyses) and Section 8.5.3 (Immunogenicity Analyses) | Updated the analyses of the coprimary endpoints | To describe the analysis methods for the updated coprimary endpoints. |
| Protocol Synopsis – Statistical Methods (Efficacy Analyses) and Section 8.5.4 (Efficacy Analyses) | Updated to clarify the secondary efficacy analyses. | To clarify the secondary efficacy analyses. |
| Protocol Synopsis – Statistical Methods (Long-term Analysis) and Section 8.5.5 (Long-term Analysis) | Added section to describe long-term analyses of safety, efficacy, and immunogenicity data to include data collected during Part B of the study. | To describe long-term analyses including Part B data. |

| Section # and Name | Description of Change | Brief Rationale |
|---|---|---|
| Protocol Synopsis – Study Analyses (Interim Analyses) and Section 8.6.1 (Interim Analyses) | Updated to indicate that more than one interim analysis may be performed, and to describe the timepoints at which the analyses will be performed. | To support potential EUA in the adolescent age group. |
| Section 1.2.2 (Clinical Studies) | Revised section to provide data for ongoing mRNA-1273 studies. | To harmonize section content with current status of development in the program. |
| Section 1.3.2 (Risks from Study Participation and Their Mitigation), 3.1.1 (Part A, the Blinded Phase), 5.3.2 (Administration of Study Vaccine), 7.1.1 (Use of Electronic Diaries), and 7.1.4 (Vital Sign Measurements) | Updated post-IP administration observation period from 60 minutes to 30 minutes. | Pursuant to cross-functional discussion and observation period of 15 minutes under the EUA. |
| Section 3.3 (Justification for Dose, Control Product, and Choice of Study Population) | Updated section to include justification for the crossover design. | The addition of the crossover design was one of the primary purposes of this amendment. |
| Section 5.3.2 (Administration of Study Vaccine) | Updated IP administration to include the IP administered in Parts A and B of the study. | The addition of the crossover design was one of the primary purposes of this amendment. |
| Section 6.2 (Discontinuing Study Vaccination) | Updated to remove the allowance for study removal for serology or RT-PCR testing positive for SARS-CoV2 for either Day 1 or for an illness visit. | Updated to be consistent with current standard of care in the US. |
| Section 6.4 (Study Pause Rules) | Updated to clarify that study pause rules are only applicable to Part A of the study but that participants will be monitored for the events leading to study pause during Part B of the study. Updated to clarify actions to be taken if thresholds for any pause rules are met.<br>Updated to clarify that any Grade 3 or higher AEs meet event criteria. | To clarify safety assessments after crossover. |
| Section 7.1.1. (Use of Electronic Diaries) | Updated to clarify that eDiaries will only be used in Part A of the study. | The solicited AR profile will be sufficiently demonstrated in Part A; therefore, it is no longer necessary to collect additional data on solicited ARs following injection in the open-label phase. |
| Section 7.3.1 (Vaccine Effectiveness Assessments) | Updated to further define criteria for SARS-CoV-2 infection. | To clarify the definition of SARS-CoV-2 infection in participants who are SARS-CoV-2 negative at baseline. |

| Section # and Name | Description of Change | Brief Rationale |
|---|---|---|
| Section 7.4.4.1 (Anaphylaxis) | Added section to characterize anaphylaxis and provide reporting requirements. | This text is being added to all mRNA-1273 protocols based on recent reports of anaphylaxis in the post-Emergency Use Authorization setting. |
| Section 7.4.5 (Adverse Events of Special Interest) | Updated to define an AESI and provide reporting requirements. | To provide more specific guidance to sites. |
| Section 7.4.7 (Eliciting and Documenting Adverse Events) | Updated to clarify that solicited ARs will be collected only in Part A, and to define the end of study participation for the purposes of collection MAAEs and SAEs. | To provide more specific guidance to sites. |
| Section 8.1 (Blinding and Responsibility for Analyses) | Updated to describe unblinding procedures relevant to the interim analyses. | To clarify the blinding/unblinding plan and process. |
| Section 10.1 (Appendix 1: Schedule of Assessments) | Table 7 title updated to reflect Part A of the study.<br><br>Updated study visit day D209 to "D209/Participant Decision Visit"<br><br>Increased visit window for D209 to ±56 days.<br><br>Increased visit window for the safety follow-ups (D223-D363) to ±3 days.<br><br>Row added for revised informed consent/assent form.<br><br>Added note to clarify the SoA to be followed after unblinding for participants who previously received mRNA-1273.<br><br>Added footnote for time frame for Participant Decision Clinic Visit.<br><br>Revised footnote to indicate that e-Diary recording will start approximately 30 minutes after injection. | To clarify as related to crossover design. |
| | Added table (Table 8) for the Participant Decision Clinic Visit. | To allow for participant unblinding and decision to receive mRNA-1273 if previously randomized to placebo. |

| Section # and Name | Description of Change | Brief Rationale |
|---|---|---|
| | Added new SoA (Table 9) for Part B of the study. | To provide instructions for participants previously randomized to placebo who have elected to receive mRNA-1273 in Part B of the study. |
| | Added flow chart (Figure 4) between Parts A and B of the study. | To demonstrate participants' movement between Part A and Part B, and back to Part A. |
| Section 10.2.8 (Protocol Deviations) | Added guidance for participants' movement between Part A and Part B. | The addition of the crossover design was one of the primary purposes of this amendment. |

Abbreviations: AESI = adverse event of special interest; AR = adverse reaction; bAb = binding antibody; CBER = Center for Biologics Evaluation and Research; D = day; eDiary = electronic diary; EUA = Emergency Use Authorization; IP = investigational product; MAAE = medically attended adverse event; mITT = modified intent-to-treat; nAb = neutralizing antibody; SAE = serious adverse event; RT-PCR = reverse transcriptase polymerase chain reaction; SARS-CoV2 = Severe Acute Respiratory Syndrome coronavirus 2; SoA = Schedule of Assessments.

Signature Page for VV-CLIN-003517 v4.0

| | |
|---|---|
| Approval | Rituparna Das<br>Clinical<br>09-Nov-2021 18:48:19 GMT+0000 |

| | |
|---|---|
| Approval | Charbel Haber<br>Regulatory<br>09-Nov-2021 19:12:48 GMT+0000 |

| | |
|---|---|
| Approval | Deborah Manzo<br>Clinical<br>09-Nov-2021 22:18:06 GMT+0000 |

| | |
|---|---|
| Approval | Jacqueline Miller<br>Clinical<br>10-Nov-2021 02:01:26 GMT+0000 |

| | |
|---|---|
| Approval | Walter Straus<br>Pharmacovigilance<br>10-Nov-2021 02:10:07 GMT+0000 |

Signature Page for VV-CLIN-003517 v4.0

### CLINICAL STUDY PROTOCOL

| | |
|---|---|
| **Protocol Title:** | A Phase 2/3, Two-Part, Open-Label, Dose-Escalation, Age De-escalation and Randomized, Observer-Blind, Placebo-Controlled Expansion Study to Evaluate the Safety, Tolerability, Reactogenicity, and Effectiveness of mRNA-1273 SARS-CoV-2 Vaccine in Healthy Children 6 Months to Less Than 12 Years of Age |
| **Protocol Number:** | mRNA-1273-P204 |
| **Sponsor Name:** | ModernaTX, Inc. |
| **Legal Registered Address:** | 200 Technology Square<br>Cambridge, MA 02139 |
| **Sponsor Contact and Medical Monitor:** | Sabine Schnyder Ghamloush, MD<br>ModernaTX, Inc.<br>200 Technology Square<br>Cambridge, MA 02139<br>Telephone: 1-617-758-9453<br> |
| **Regulatory Agency Identifier Number(s):** | IND: 019745 |
| **Amendment Number:** | 5 |
| **Date of Amendment 5:** | 29 Sep 2021 |
| **Date of Amendment 4:** | 25 Aug 2021 |
| **Date of Amendment 3:** | 23 Jul 2021 |
| **Date of Amendment 2:** | 17 Jun 2021 |
| **Date of Amendment 1:** | 30 Apr 2021 |
| **Date of Original Protocol:** | 24 Feb 2021 |

**CONFIDENTIAL**

All financial and nonfinancial support for this study will be provided by ModernaTX, Inc. The concepts and information contained in this document or generated during the study are considered proprietary and may not be disclosed in whole or in part without the expressed written consent of ModernaTX, Inc. The study will be conducted according to the *International Council for Harmonisation (ICH) of Technical Requirements for Registration of Pharmaceuticals for Human Use, E6(R2) Good Clinical Practice (GCP) Guidance.*

#### PROTOCOL APPROVAL – SPONSOR SIGNATORY

| | |
|---|---|
| **Study Title:** | A Phase 2/3, Two-Part, Open-Label, Dose-Escalation, Age De-escalation and Randomized, Observer-Blind, Placebo-Controlled Expansion Study to Evaluate the Safety, Tolerability, Reactogenicity, and Effectiveness of mRNA-1273 SARS-CoV-2 Vaccine in Healthy Children 6 Months to Less Than 12 Years of Age |
| **Protocol Number:** | mRNA-1273-P204 |
| **Amendment Number:** | 5 |
| **Date of Amendment:** | 29 Sep 2021 |

Protocol accepted and approved by:

| | |
|---|---|
| **Please see eSignature and date in the last page of the document.** | |
| Rituparna Das, MD, PhD<br>Clinical Development – COVID-19 Vaccines<br>ModernaTX, Inc.<br>200 Technology Square<br>Cambridge, MA 02139<br>Telephone: 617-710-9794 | Date |

### DECLARATION OF INVESTIGATOR

I have read and understood all sections of the protocol titled "A Phase 2/3, Two-Part, Open-Label, Dose-Escalation, Age De-escalation and Randomized, Observer-Blind, Placebo-Controlled Expansion Study to Evaluate the Safety, Tolerability, Reactogenicity, and Effectiveness of mRNA-1273 SARS-CoV-2 Vaccine in Healthy Children 6 Months to Less Than 12 Years of Age" and the most recent version of the investigator's brochure.

I agree to supervise all aspects of the protocol and to conduct the clinical investigation in accordance with the current protocol, the *International Council for Harmonisation (ICH) of Technical Requirements for Registration of Pharmaceuticals for Human Use, E6(R2) Good Clinical Practice (GCP) Guidance*, and all applicable government regulations. I will not make changes to the protocol before consulting with ModernaTX, Inc. or implement protocol changes without institutional review board (IRB) approval except to eliminate an immediate risk to participants.

I agree to administer study treatment only to participants under my personal supervision or the supervision of a subinvestigator. I will not supply study treatment to any person not authorized to receive it. I also agree that persons debarred from conducting or working on clinical studies by any court or regulatory agency will not be allowed to conduct or work on studies for the Sponsor or a partnership in which the Sponsor is involved. I will immediately disclose it in writing to the Sponsor if any person who is involved in the study is debarred, or if any proceeding for debarment is pending, or, to the best of my knowledge, threatened.

I will not disclose confidential information contained in this document, including participant information, to anyone other than the recipient study staffs and members of the IRB. I agree to ensure that this information will not be used for any purpose other than the evaluation or conduct of the clinical investigation without prior written consent from ModernaTX, Inc. I will not disclose information regarding this clinical investigation or publish results of the investigation without authorization from ModernaTX, Inc.

The signature below provides the necessary assurance that this study will be conducted according to all stipulations of the protocol, including statements regarding confidentiality, and according to local legal and regulatory requirements, US federal regulations, and ICH E6(R2) GCP guidelines.

______________________________ ______________________

Signature of principal investigator Date

______________________________

Printed name of principal investigator

### PROTOCOL AMENDMENT SUMMARY OF CHANGES

| DOCUMENT HISTORY | |
|---|---|
| **Document** | **Date** |
| Amendment 5 | 29 Sep 2021 |
| Amendment 4 | 25 Aug 2021 |
| Amendment 3 | 23 Jul 2021 |
| Amendment 2 | 17 Jun 2021 |
| Amendment 1 | 30 Apr 2021 |
| Original Protocol | 24 Feb 2021 |

**Amendment 5, (29 Sep 2021): Current Amendment**

**Main Rationale for the Amendment:**

This amendment is considered to be non-substantial based on the criteria set forth in Article 10(a) of Directive 2001/20/European Commission (EC) of the European Parliament and the Council of the European Union.

The main rationale for this amendment is to simplify the process for potential cross-over vaccination given the increased sample size of the study and to ensure retention in the study for safety follow-up, incorporate Clarification Memo #8, align with the Cardiac Event Adjudication Committee (CEAC) charter, and clarify the Data Safety Monitoring Board (DSMB) safety data review process for the younger age groups (2 to < 6 years; 6 months to < 2 years) to match the process in the older age group (6 to < 12 years) and allow DSMB review of Part 1 (open-label phase) data before start of Part 2 (blinded phase) for the younger age groups. Additional updates were made to clarify the statistical analysis plan, including the inferred efficacy analysis and interim analyses.

The summary of changes table provided below describes the major changes made to Amendment 5 relative to Amendment 4, including the sections modified and corresponding rationales. The synopsis of Amendment 5 has been modified to correspond to changes in the body of the protocol. Minor copy edits and administrative updates were made throughout the protocol to align with new content as well as for clarity, readability, and/or accuracy.

**Summary of Major Changes in Protocol Amendment 5:**

| Section # and Name | Description of Change | Brief Rationale |
|---|---|---|
| Section 3.1 (General Design; Figure 1), Section 3.1.1.2 (Treatment and Follow-up Period), Section 7.2 (Blood Collections for Immunogenicity | Made updates to explain that blood sample collection for participants in Cohort D will be prior to randomization and the first dose at Day 1 and within 4 | For clarification. |

| | | |
|---|---|---|
| Assessments and Biomarker Samples), Appendix 1 (Schedule of Assessments; Table 10) | days of receiving Dose 2 at Day 30 (+ 3 days). | |
| Section 3.3 (Justification for Dose, Control Product, and Choice of Study Population), Appendix 1 (Schedule of Assessments) | Made updates to Section 3.3 to clarify the justification for the choice of study population, including unblinding of eligible study participants, placebo recipient cross-over vaccination, booster dose eligibility for participants who received a lower dose in Part 1 than was ultimately approved for their respective age group in Part 2, and study discontinuation for participants receiving an EUA vaccine outside the protocol.<br><br>In accordance with changes made to Section 3.3, updated text and Table 11 in Appendix 1 to clarify placebo recipient cross-over vaccination. | Given the increase in sample size, unblinding via clinic visit is no longer feasible and will instead be performed over the phone, if desired by the family. To ensure retention in the study for safety follow-up, cross-over vaccination will be offered to all placebo recipients per their request if any COVID-19 vaccine (mRNA-1273 or other) becomes authorized or licensed for their age group. |
| Section 4.2 (Exclusion Criteria) | Added text to exclusion criteria #8 to provide context related to the influenza vaccine. | For incorporation of Clarification Memo #8. |
| Section 6.1.1 (Individual Participant Criteria for Delay of Study Vaccination) | Provided text in Section 6.1.1 to describe additional reasons to delay study vaccination for participants. | For incorporation of Clarification Memo #8. |
| Section 7.3.3 (Follow-up/Convalescent Period After Diagnosis with COVID-19) | Deleted text, "Study participants will be monitored by trained study site personnel for a 28 day period after diagnosis." | For accuracy. |
| Section 7.4.5 (Adverse Events of Special Interest), Section 7.5.2 (Data Safety Monitoring Board) | Clarified that the CEAC will review suspected cases of myocarditis/pericarditis to determine if they meet CDC criteria of "probable" or "confirmed" event and to assess severity, but recommendations to the Sponsor to continue vaccine dosing will be made by DSMB. | To align with CEAC charter. |
| Section 7.5.2 (Data Safety Monitoring Board) | Changed description of timing of DSMB review and scope for younger age groups (2 to < 6 years; 6 months to < 2 years) from separate reviews in each age group with data from Part 1 and Part 2 during the conduct of | To allow the DSMB to review data from Part 1 (open-label phase) for all doses used in < 6 year old participants before starting Part 2 for 2 to < 6 years and 6 months to < 2 years age groups. |

| | | |
|---|---|---|
| | Part 2 to one review of all available Part 1 safety data for all doses used in < 6 years before start of Part 2 for each age group. | |
| Section 8.5.4 (Inferred Efficacy Analysis) | Added text to explain the following: 1) analyses of efficacy endpoints in Part 2 will be performed for the randomized blinded phase, and 2) additional exploratory analyses will be conducted in the blinded and unblinded phases for participants randomized to mRNA-1273 in Part 2, and in the unblinded phase for participants who are originally randomized to the placebo arm and cross-over to mRNA-1273 after use of any COVID-19 vaccine is authorized or licensed for the participant's age group. | For clarification. |
| Section 8.6.1 (Interim Analyses) | Updated description of Part 2 to indicate that an interim analysis of immunogenicity and safety will be performed after all or a subset of participants have completed Day 57 (1 month after the second dose) in Part 1 or Part 2 within an age group. | For clarification. |
| Appendix 1 (Schedule of Assessments; Table 10 and Table 11) | Updated footnote #2 in Table 10 and added footnote #2 to Table 11 to include cross-references for Section 6.1.1. | For alignment. |

Abbreviations: CDC = Centers for Disease Control and Prevention; CEAC = Cardiac Event Adjudication Committee; COVID-19 = coronavirus disease 2019; DSMB = Data Safety Monitoring Board; EUA = Emergency Use Authorization; mRNA = messenger RNA; SoA = Schedule of Assessments.

#### PROTOCOL SYNOPSIS

**Name of Sponsor/Company:** ModernaTX, Inc.

**Name of Investigational Product:** mRNA-1273 for injection

**Name of Active Ingredient:** mRNA-1273

**Protocol Title:** A Phase 2/3, Two-Part, Open-Label, Dose-Escalation, Age De-escalation and Randomized, Observer-Blind, Placebo-Controlled Expansion Study to Evaluate the Safety, Tolerability, Reactogenicity, and Effectiveness of mRNA-1273 SARS-CoV-2 Vaccine in Healthy Children 6 Months to Less Than 12 Years of Age

**Protocol Number:** mRNA-1273-P204

**Study Period (years):** Approximately 14 months

**Phase of Development:** Phase 2/3

**Estimated date first participant enrolled:** 15 Mar 2021

**Estimated date last participant completed:** 12 Jun 2023

**Total Number of Sites:** Approximately 75 to 100 study sites in the United States and Canada.

**Objectives and Endpoints**

| Objectives | Endpoints |
|---|---|
| **Primary Objectives** | **Primary Endpoints** |
| • To evaluate the safety and reactogenicity of up to 3 dose levels (25, 50, and 100 µg) of mRNA-1273 vaccine administered as 2 doses 28 days apart in 3 age groups | • Solicited local and systemic ARs through 7 days after each injection<br>• Unsolicited AEs through 28 days after each injection<br>• MAAEs through the entire study period<br>• SAEs through the entire study period<br>• AESIs, including MIS-C and myocarditis and/or pericarditis, through the entire study period |
| • To infer the efficacy of mRNA-1273 (25, 50, and 100 µg, administered as | • The proportion of participants with a serum antibody level at Day 57 ≥ antibody threshold of protection |

| | |
|---|---|
| 2 doses 28 days apart) based on immunogenicity in 3 age groups | – If an accepted serum antibody threshold of vaccine protection against COVID-19 is available, this analysis will form the basis to infer efficacy<br>• The GM value of serum antibody level and seroresponse rate from Study P204 vaccine recipients at Day 57 compared with those from young adult (18 to 25 years of age) vaccine recipients (Day 57) in the clinical endpoint efficacy trial (Study P301)<br>– If a threshold is not available, efficacy will be inferred by establishing noninferiority for each age group (6 to < 12 years, 2 to < 6 years, and 6 months to < 2 years in Study P204) compared to 18- to 25-year old participants (Study P301) by both GM value of serum antibody levels and seroresponse rate. A definition of seroresponse will be provided in the statistical analysis plan based on forthcoming information about assay performance |
| **Secondary Objectives** | **Secondary Endpoints** |
| • To evaluate the persistence of the immune response to mRNA-1273 vaccine (25, 50, and 100 μg) administered as 2 doses 28 days apart | • The GM values of SARS-CoV-2 S protein-specific bAb on Day 1, Day 57 (1 month after Dose 2), Day 209 (6 months after Dose 2), and Day 394 (1 year after Dose 2)<br>• The GM values of SARS-CoV-2-specific nAb on Day 1, Day 57 (1 month after Dose 2), Day 209 (6 months after Dose 2), and Day 394 (1 year after Dose 2) |
| • To evaluate the incidence of SARS-CoV-2 infection after vaccination with mRNA-1273 or placebo | • The incidence of SARS-CoV-2 infection including symptomatic and asymptomatic infection (by serology and/or RT-PCR) postbaseline |

| | |
|---|---|
| | • SARS-CoV-2 infection will be defined in participants with negative SARS-CoV-2 at baseline:<br>– bAb level against SARS-CoV-2 nucleocapsid protein negative at Day 1 that becomes positive (as measured by Roche Elecsys) postbaseline, OR<br>– Positive RT-PCR postbaseline |
| • To evaluate the incidence of asymptomatic SARS-CoV-2 infection after vaccination with mRNA-1273 or placebo | • The incidence of SARS-CoV-2 infection measured by RT-PCR and/or bAb levels against SARS-CoV-2 nucleocapsid protein (by Roche Elecsys) postbaseline in participants with negative SARS-CoV-2 at baseline, in the absence of any COVID-19 symptoms. |
| • To evaluate the incidence of COVID-19 after vaccination with mRNA-1273 or placebo. COVID-19 is defined as clinical symptoms consistent with COVID-19 AND positive RT-PCR for SARS-CoV-2 | • The incidence of the first occurrence of COVID-19 postbaseline, where COVID-19 is defined as symptomatic disease based on CDC case definition.[1] |
| **Exploratory Objectives** | **Exploratory Endpoints** |
| • To evaluate the genetic and/or phenotypic relationships of isolated SARS-CoV-2 strains to the vaccine sequence | • Alignment of genetic sequence of viral isolates with that of the vaccine sequence |
| • To describe the ratio or profile of specific S protein bAb relative to nAb in serum | • Relative amounts or profiles of S protein-specific bAb and specific nAb titers in serum |
| • To characterize the clinical profile and immune responses of participants with COVID-19 or with SARS-CoV-2 infection | • Description of clinical severity and immune responses of participants who are identified as infected by SARS-CoV-2 (COVID-19) |
| • To assess, in a subset of participants, the SARS-CoV-2 S protein-specific T-cell responses | • Magnitude, phenotype, and percentage of cytokine-producing S protein-specific T cells, as measured by flow cytometry at different time points after vaccination relative to baseline |

| | |
|---|---|
| • To explore asymptomatic SARS-CoV-2 infection after vaccination with mRNA-1273 or placebo in participants with serologic evidence of infection at baseline | • GM and GMFR of bAb levels against SARS-CoV-2 nucleocapsid protein (quantitative IgG) |

Abbreviations: AE = adverse event; AESI = adverse event of special interest; AR = adverse reaction; bAb = binding antibody; CDC = Center for Disease Control; COVID-19 = coronavirus disease 2019; GM = geometric mean; GMFR = geometric mean fold-rise; IgG = immunoglobin G; IP = investigational product; LOD = limit of detection; MAAE = medically attended adverse event; MIS-C = multisystem inflammatory syndrome in children; mRNA = messenger RNA; nAb = neutralizing antibody; RT-PCR = reverse transcriptase polymerase chain reaction; S = spike; SAE = severe adverse event; SARS-CoV-2 = severe acute respiratory syndrome coronavirus 2.

[1] The case definition of COVID-19 includes at least one of the following systemic symptoms: fever (temperature > 38ºC/≥ 100.4ºF) or chills (of any duration, including ≤ 48 hours), cough (of any duration, including ≤ 48 hours), shortness of breath or difficulty breathing (of any duration, including ≤ 48 hours), fatigue, headache, myalgia, nasal congestion or rhinorrhea, new loss of taste or smell, sore throat, abdominal pain, diarrhea, nausea or vomiting, poor appetite or poor feeding, AND a positive test for SARS-CoV-2 by RT-PCR.

**Overall Study Design**

This is a Phase 2/3, two-part, open-label, dose-escalation, age de-escalation and randomized, observer-blind, placebo-controlled expansion study intended to infer the effectiveness of messenger RNA (mRNA)-1273 in children aged 6 months to < 12 years. The study population will be divided into 3 age groups (6 to < 12 years, 2 to < 6 years, and 6 months to < 2 years), and up to 3 dose levels (25, 50, and 100 µg) of mRNA-1273 will be evaluated.

The study will be conducted in 2 parts. Part 1 of the study will be open-label and consist of dose-escalation and age de-escalation in approximately 1,275 participants to select the dose for each age group with the highest number enrolled in the oldest age group (the table below provides the approximate number of participants in each age group). Part 2 of the study will be a placebo-controlled observer-blind evaluation of the selected dose in up to 12,000 participants (up to 4,000 participants in each of the 6 to < 12 years, 2 to < 6 years, and the 6 months to < 2 years age groups). No participants in Part 1 will participate in Part 2 of the study.

In order to expedite the study of the safety of mRNA-1273 in school-aged children 6 to < 12 years of age; Part 1 will enroll a total of approximately 375 participants per dose (both at the 50 and 100 µg dose level) in this age group (6 to < 12 years). The first 75 of these participants per dose will be included in the safety evaluation for dose-escalation and age de-escalation as well as the immunogenicity assessment needed for dose selection, as applicable. Additionally, approximately 300 participants per dose will be enrolled to assess for any adverse event (AE) occurring at 1% or higher. Conversely, approximately 75 participants in the middle age group (2 to < 6 years) will be enrolled per dose in Part 1 for safety and dose selection and Part 2 will utilize the dose selected in Part 1 to inform on AEs occurring at a frequency of 1% or greater. In the youngest age group (6 months to < 2 years), approximately 150 participants will be enrolled per dose in Part 1 for safety and dose selection and Part 2 will utilize the dose selected in Part 1 to inform on AEs occurring at a frequency of 1% or greater. For more details, please refer to Safety Oversight.

The study will begin with the oldest age group (6 to < 12 years) and age de-escalate as described under Study Progression. Each age group will begin with Part 1 and advance to Part 2 independently.

The mRNA-1273 investigational vaccine or placebo will be administered as 2 intramuscular (IM) injections approximately 28 days apart.

The mRNA-1273 dose levels that will be evaluated in each age group in Part 1 and Part 2 of the study are given in the following table.

##### Planned Age Groups and mRNA-1273 Dose Levels in Part 1 and Part 2 of the Study

| Age Group | Part 1: mRNA-1273 25 µg | Part 1: mRNA-1273 50 µg | Part 1: mRNA-1273 100 µg | Part 2: Selected Dose Level of mRNA-1273 From Part 1 | Part 2: Placebo |
|---|---|---|---|---|---|
| 6 to < 12 years | | Study Arm 1 (n=375) | Study Arm 2 (n=375) | Study Arm 8 (n= 3,000) | Study Arm 9 (n= 1,000) |
| 2 to < 6 years | Study Arm 7 (Optional) (n=75) | Study Arm 3 (n=75) | Study Arm 4 (n=75) | Study Arm 10 (n= up to 3,000) | Study Arm 11 (n=up to 1,000) |
| 6 months to < 2 years | Study Arm 5 (n=150) | Study Arm 6 (n=150) | | Study Arm 12 (n= up to 3,000) | Study Arm 13 (n=up to 1,000) |

##### Study Progression

Part 1 of the study will be open-label. The study will begin with enrollment of approximately 375 participants in the 6 to < 12 years age group (Study Arm 1), and dosing with 50 µg of mRNA-1273. After at least 75 participants in Study Arm 1 have completed Day 8 (1 week after Dose 1 of mRNA-1273 50 µg), an internal safety team (IST) will review the available safety data and provide a recommendation whether to escalate the dose to 100 µg in the 6 to < 12 years age group (Study Arm 2; n = 375) and independently whether to begin dosing at the 50-µg dose level in the 2 to < 6 years age group (Study Arm 3; n = 75). After at least 75 participants in Study Arm 1 reach Day 36 (1 week after Dose 2 of mRNA-1273 50 µg), the IST will again review the available safety data and provide a recommendation whether to administer Dose 2 of mRNA-1273 100 µg in Study Arm 2 and whether to administer Dose 2 of mRNA-1273 50 µg in Study Arm 3. Simultaneously, the enrollment of the remaining 300 planned participants in both Arms 1 and 2 will be ongoing. A preliminary safety and immunogenicity data review of Arm 1, and Arm 2 as applicable, will aid in the selection of a dose level for Part 2. Once all or a subset of participants from each of Study Arms 1 and 2 reach Day 57 (1 month after Dose 2 of mRNA-1273), an interim analysis may be conducted to review the safety and immunogenicity data. Cumulative safety data for approximately 300 participants at the selected dose level will be reviewed by the Data Safety Monitoring Board (DSMB) before enrollment in Part 2, and the DSMB safety review recommendation will enable the expansion of the 6 to < 12 years age group (Part 2) to receive either mRNA-1273 at the selected dose level (Study Arm 8; n = 3,000) or placebo (Study Arm 9; n = 1,000). The primary analysis of immunogenicity in Part 2 for Study Arm 8 (mRNA-1273 recipients) will be conducted after a pre-specified Immunogenicity Subset of participants reaches Day 57, and safety analysis for Arms 8 and 9 will be conducted after a subset or all participants reach Day 57.

For the middle age group (2 to < 6 years) progression in Part 1 will be as follows: After 75 participants in Study Arm 3 have completed Day 8 (1 week after Dose 1 of mRNA-1273 50 µg), an IST will review the available safety data and provide a recommendation whether to escalate the dose to 100 µg in the 2 to < 6 years age group (Study Arm 4; n = 75) and whether to begin dosing in the 6 months to < 2 years age group at the 25 µg-dose level (Study Arm 5; n = 150). After the 75 participants in Study Arm 3 reach Day 36 (1 week after Dose 2 of mRNA-1273 50 µg), the IST will review the available safety data and provide a recommendation whether to administer Dose 2 of mRNA-1273 100 µg in Study Arm 4 and whether to administer Dose 2 of mRNA-1273 25 µg in Study Arm 5. An optional Arm 7 may be enrolled in this age group (approximately 75 participants) at the 25-µg dose if the 100-µg dose is eliminated at any point during dose escalation process, to maintain dose ranging for this age group. A preliminary safety and immunogenicity data review of all applicable Arms will aid in the selection of a dose level for Part 2. Once all or a subset of participants in all applicable Arms reach Day 57 (1 month after Dose 2 of mRNA-1273), an interim analysis may be conducted to review safety and immunogenicity data. A preliminary safety and immunogenicity data review of all applicable Arms will aid in the selection of a dose level for Part 2 and for expansion of the 2 to < 6 years age group (Part 2) to receive either mRNA-1273 at the selected dose level (Study Arm 10; n = up to 3,000) or placebo (Study Arm 11; n = up to 1,000). The primary analysis of immunogenicity in Part 2 for Study Arm 10 (mRNA-1273 recipients) will be conducted after a pre-specified Immunogenicity Subset of participants reaches Day 57, and safety analysis for Arms 10 and 11 will be conducted after a subset or all participants in Arms 10 and 11 reach Day 57.

For the youngest age group (6 months to < 2 years) progression in Part 1 will be as follows: In Study Arm 5, after 150 participants have completed Day 8 (1 week after Dose 1 of mRNA-1273 25 µg), an IST will review the available safety data and provide a recommendation whether to escalate the dose to 50 µg (Study Arm 6; n = 150). After the 150 participants in Study Arm 5 reach Day 36 (1 week after Dose 2 of mRNA-1273 25 µg), the IST will review the available safety data and provide a recommendation whether to administer Dose 2 of mRNA-1273 50 µg in Study Arm 6. Once all or a subset of participants in Study Arms 5 and 6 reach Day 57 (1 month after Dose 2 of mRNA-1273), an interim analysis may be conducted to review the tolerability and immunogenicity data at each dose level. A preliminary safety and immunogenicity data review of Arm 5 and Arm 6, as applicable, will aid in the selection of a dose level for Part 2 and allow for expansion of the 6 months to < 2 years age group (Part 2) to receive either mRNA-1273 at the selected dose level (Study Arm 12; n = up to 3,000) or placebo (Study Arm 13; n = up to 1,000). The primary analysis of immunogenicity in Part 2 for Study Arm 12 (mRNA-1273 recipients) will be conducted after the pre-specified Immunogenicity Subset of participants reaches Day 57, and safety analysis will be conducted after a subset or all participants in Arm 12 and 13 reach Day 57.

In general, if a decision is made not to proceed with administration of a higher dose and/or the second injection at a given dose level (eg, due to safety concerns), the participants scheduled to receive the higher dose may receive a lower dose, and the participants who had received the higher dose level as their first injection will likely be given the next lower dose level that was tolerated for their second injection. The dose level not tolerated in the older age group will not be administered in any of the younger age groups. Within an age group, if one of the dose levels is found to have an unacceptable tolerability profile, the age group may progress to Part 2 at a lower dose level.

The final analysis will be performed when participants (Study Arms 1 to 13) conclude the safety follow-up at 12 months.

The goal of the study is to support an indication for use of mRNA-1273 50 or 100 µg IM, given as 2 injections, approximately 28 days apart in the 6 to < 12 years age group; mRNA-1273 25, 50, or 100 µg IM, given as 2 injections, approximately 28 days apart in the 2 to < 6 years age group; and mRNA-1273 25 or 50 µg IM, given as 2 injections, approximately 28 days apart in the 6 months to < 2 years age group. The basis for demonstrating vaccine effectiveness is proposed to be met by measuring serum antibody responses in the study participants. The approach to inferring vaccine effectiveness will depend on whether an accepted serum antibody threshold conferring protection against coronavirus disease 2019 (COVID-19) has been established. If an antibody threshold of protection has been established, effectiveness will be inferred based on the proportion of study participants with serum antibody levels (on Day 57) that meet or exceed the antibody threshold. If an antibody threshold of protection has not been established, effectiveness will be inferred based on demonstrating noninferiority for each age group (6 to < 12 years, 2 < 6 years, and 6 months to < 2 years in Study P204) compared to young adult (18 to 25 years of age) participants enrolled in the ongoing clinical endpoint efficacy trial (Study P301) by both geometric mean (GM) values and seroresponse rate.

This study in children 6 months to < 12 years of age will monitor all participants for a total of 12 months following the second dose of vaccine or placebo. If a COVID-19 vaccine is authorized for a specific age group before the end of the study, please refer to the Justification for Dose, Control Product, and Choice of Study Population Section in the protocol for unblinding and/or cross-over plans. Safety assessments will include solicited adverse reactions (ARs) (the day of injection and 6 subsequent days), unsolicited adverse events (AEs; 28 days after each injection), and medically attended adverse events (MAAEs), serious AEs (SAEs), and adverse events of special interest (AESIs; including multisystem inflammatory syndrome in children [MIS-C] and myocarditis and/or pericarditis) throughout the study period.

Blood samples will be collected from participants in Part 1 and Part 2 of the study for assessment of immunogenicity. If a Day 1 (baseline) blood sample cannot be obtained in either Part 1 or Part 2, the participant will be considered either a screen failure due to inability to satisfy inclusion criterion 3 or could be rescheduled for rescreening. Participants may be rescreened 1 time, either within the same screening period for Part 1 or Part 2 or in a new screening period of Part 2 if the initial screening was in Part 1. If the participant is rescreened and a blood sample still cannot be obtained, then he/she will be considered a screen failure due to inability to satisfy inclusion criterion 3. All participants in Part 1 of the study will provide blood specimens for immunogenicity on Day 1 (prior to the first dose) and will also provide additional samples after the second dose of mRNA-1273 at Day 57, Day 209, and Day 394, except for the expansion of Arms 1 and 2 (N= ~300 participants per arm) who have a scheduled Day 1 blood draw and a voluntary blood draw on Day 57 only. Participants in each age group in Part 2 will be assigned to 1 of 5 phlebotomy cohorts. Participants in 3 of these 5 cohorts will provide blood specimens for immunogenicity at the following time points: Day 1 (prior to randomization and before the first dose), Day 57, and one of Day 29 (prior to the second dose), Day 209, or Day 394 (such that each participant provides a total of 3 blood samples only). Participants in the Cohort D (remainder of the age group) will provide a blood sample at Day 1 (prior to randomization and before the first dose) and within 4 days of receiving Dose 2 at Day 30 (+3 days) for storage and potential future biomarker testing. A fifth cohort of participants (selected Vaccine and Treatment Evaluation Units [VTEU] sites only) will provide blood samples for assessment of exploratory serology and cell-mediated immunity (CMI; spike [S]-protein-specific T cell responses) on Day 1 (prior to randomization and before the first dose), Day 43, Day 209, and Day 394. Blood samples will also be tested for the development of antibodies directed against nonvaccine antigen (eg, antibodies against the nucleocapsid protein), which will signify infection with severe acute respiratory syndrome coronavirus 2 (SARS-CoV-2). The incidence of SARS-CoV-2 infection among vaccine recipients and placebo recipients will be compared to assess the potential for mRNA-1273 to reduce the rate of infection in vaccine recipients. In addition, all participants will be monitored for symptoms of COVID-19 and scheduled for illness visits if concerning symptoms occur, and a nasal swab will be collected at the illness visit.

**Safety Oversight:**

The contract research organization's medical monitor, the Sponsor's medical monitor, and the individual study site investigators will monitor safety throughout the study.

Internal Safety Team

An IST will review safety data throughout the study. For Part 1 dose-escalation and age de-escalation, based on the review of all available safety data through at least Day 8 (1 week after Dose 1 of mRNA-1273) for at least 75 participants at each dose level within the 6 to < 12 years and 2 to < 6 years age group, the IST will recommend whether dose-escalation and age de-escalation are appropriate. This process will then be repeated for all participants in the 25 and 50 µg dose levels within the lower age group of 6 months to < 2 years of age. An IST review of all available safety data through at least Day 36 (1 week after Dose 2 of mRNA-1273 at each dose level) of at least 75 participants will be required prior to the administration of the second injection of the next higher dose. In addition, the IST will escalate any safety concerns to the DSMB. The frequency of IST meetings will be described in more detail in the IST charter.

Data Safety Monitoring Board

Safety oversight will be under the direction of a DSMB composed of external independent consultants with relevant expertise. Members of the DSMB will be independent from the study conduct and free of conflict of interest. The DSMB will meet at pre-specified timepoints during the study to assess safety throughout the study conduct. For the 6 to < 12 years age group, the DSMB will review cumulative safety data for approximately 300 participants enrolled at the selected dose level in Part 1 before enrollment begins in Part 2. For both the middle age group (2 to < 6 years) and the youngest age group (6 months to < 2 years) the DSMB will review cumulative safety data in both younger age groups (2 to < 6 years; 6 months to < 2 years) combined and at all dose levels administered in Part 1, before start of Part 2 (blinded phase) for each age group. The DSMB will operate under the rules of an approved charter that will be written and reviewed at the organizational meeting of the DSMB. Details regarding the DSMB composition, responsibilities, procedures, and frequency of data review will be defined in its charter.

If any of the study pause rules are met, the Sponsor will immediately suspend further enrollment and/or study dosing and the DSMB will convene on an ad hoc basis. The DSMB will review all available unblinded study data (as applicable) to help adjudicate any potential study pauses and make recommendations on further study conduct, including requesting additional information, recommending stopping the study, recommending changes to study conduct and/or the protocol, or recommending additional operational considerations due to safety issues that arise during the study.

A Cardiac Event Adjudication Committee will adjudicate any suspected cases of myocarditis, pericarditis, or myopericarditis and enable the DSMB to make recommendations to the Sponsor.

**Study Duration:** The study duration for each participant will be approximately 14 months, which includes 1 month for screening (Day -28 to Day -1), 1 month for dosing (on Day 1 and Day 29), and 12 months of follow-up after the second dose of investigational product (IP).

**Number of Participants:**

Part 1: Approximately 1,275 participants (approximately 750 participants in the 6 to < 12 years age group, approximately 225 participants 2 to < 6 years age group, and approximately 300 participants in the 6 months to < 2 years age group).

Part 2: Up to 12,000 participants (up to 3,000 participants each exposed to mRNA-1273 and 1,000 participants exposed to placebo in the 6 to < 12 years, 2 to < 6 years, and 6 months to < 2 years age groups).

**Study Eligibility Criteria:**

**Inclusion Criteria:**

Participants are eligible to be included in the study only if all the following criteria apply:

1. The participant is male or female, 6 months to < 12 years of age at the time of consent/assent (Screening Visit), who is in good general health, in the opinion of the investigator, based on review of medical history and screening physical examination.
2. If the participant has a chronic disease (eg, asthma, diabetes mellitus, cystic fibrosis, human immunodeficiency virus [HIV] infection), the disease should be stable, per investigator assessment, so that the participant can be considered eligible for inclusion. Stable diseases are those which have had no change in their status or in the medications required to control them in the 6 months prior to Screening Visit.

   Note: a change in medication for dose optimization (eg, insulin dose changes), change within class of medication, or reduction in dose are not considered signs of instability.
3. In the investigator's opinion, the parent(s)/legally acceptable representative(s) understand and are willing and physically able to comply with protocol-mandated follow-up, including all procedures, and provide written informed consent and participants are willing to provide assent.
4. The participant is 2 years or older and has a body mass index at or above the third percentile according to WHO Child Growth Standards at the Screening Visit.

   The participant is less than 2 years of age and the participant's height and weight are both at or above the 3$^{rd}$ percentile according to WHO Child Growth Standard at the Screening Visit.
5. Female participants of nonchildbearing potential may be enrolled in the study. Nonchildbearing potential is defined as premenarche.

**Special inclusion criteria for female participants who have reached menarche:**

6. Female participants of childbearing potential may be enrolled in the study if the participant fulfills all of the following criteria:
   - Has a negative pregnancy test at Screening. Pregnancy test will be performed if deemed appropriate by the investigator.
   - Has practiced adequate contraception or has abstained from all activities that could result in pregnancy for at least 28 days prior to the first injection (Day 1).
   - Has agreed to continue adequate contraception or abstinence through 3 months

following the second injection (Day 29).

- Is not currently breastfeeding.

Adequate female contraception is defined as abstinence or consistent and correct use of a US Food and Drug Administration-approved contraceptive method in accordance with the product label.

**Special inclusion criteria for children 6 months to < 12 months of age**

7. The participant was born at full-term (≥ 37 weeks gestation) with a minimum birth weight of 2.5 kg.

**Exclusion Criteria:**

Participants will be excluded from the study if any of the following criteria apply:

1. Has a known history of SARS-CoV-2 infection within 2 weeks prior to administration of IP or known close contact with anyone with laboratory-confirmed SARS-CoV-2 infection or COVID-19 within 2 weeks prior to administration of IP.
2. Is acutely ill or febrile 24 hours prior to or at the Screening Visit. Fever is defined as a body temperature ≥ 38.0°C/≥ 100.4°F. Participants who meet this criterion may have visits rescheduled within the relevant study visit windows. Afebrile participants with minor illnesses can be enrolled at the discretion of the investigator.
3. Has previously been administered an investigational or approved CoV (eg, SARS-CoV-2, SARS-CoV, Middle East respiratory syndrome-CoV) vaccine.
4. Has undergone treatment with investigational or approved agents for prophylaxis against COVID-19 (eg, receipt of SARS-CoV-2 monoclonal antibodies) within 6 months prior to enrollment.
5. Has a known hypersensitivity to a component of the vaccine or its excipients. Hypersensitivity includes, but is not limited to, anaphylaxis or immediate allergic reaction of any severity to a previous dose of messenger RNA COVID-19 vaccine or any of its components (including polyethylene glycol [PEG] or immediate allergic reaction of any severity to polysorbate).
6. Has a medical or psychiatric condition that, according to the investigator's judgment, may pose additional risk as a result of participation, interfere with safety assessments, or interfere with interpretation of results.
7. Has a history of a diagnosis or condition that, in the judgment of the investigator, may affect study endpoint assessment or compromise participant safety, specifically the following:
   - Congenital or acquired immunodeficiency, excluding HIV infection, as

described in Inclusion Criteria 2

- Chronic hepatitis or suspected active hepatitis
- A bleeding disorder that is considered a contraindication to IM injection or phlebotomy
- Dermatologic conditions that could affect local solicited AR assessments
- Any prior diagnosis of malignancy (excluding nonmelanoma skin cancer)
- Febrile seizures*

  *In Part 2 of the study, a history of a single, simple febrile seizure is allowed for children 6 years and older.

8. Has received the following:
   - Any routine vaccination with inactivated or live vaccine(s) within 14 days prior to first vaccination or plans to receive such a vaccine through 14 days following the last study vaccination.
     - Note: This excludes influenza vaccine that may be given, however, not within 14 days prior to or post Dose 1 or Dose 2. If a participant receives an influenza vaccine, this should be captured within the concomitant medication electronic case report form (eCRF) (Section 5.5.2).
   - Systemic immunosuppressants or immune-modifying drugs for > 14 days in total within 6 months prior to the day of enrollment (for corticosteroids, ≥ 1 mg/kg/day or ≥ 10 mg/day prednisone equivalent, if participant weighs > 10 kg). Participants may have visits rescheduled for enrollment if they no longer meet this criterion within the Screening Visit window. Inhaled, nasal, and topical steroids are allowed.
   - Intravenous or subcutaneous blood products (red cells, platelets, immunoglobulins) within 3 months prior to enrollment.
9. Has participated in an interventional clinical study within 28 days prior to the Screening Visit or plans to do so while participating in this study.
10. Is an immediate family member, or household contact, of an employee of the study site or Moderna or someone otherwise directly involved with the conduct of the study. As applicable, family members/household contacts of employees of the larger institution or affiliated private practice not part of the study site may be enrolled.

**Study Treatment:**

**Investigational Product:**

The term IP refers to mRNA-1273 (25, 50, and 100 μg) vaccine or placebo (0.9% sodium chloride) in this study.

mRNA-1273 is a lipid nanoparticle (LNP) dispersion of an mRNA that encodes the prefusion stabilized S protein of SARS-CoV-2 formulated in LNPs composed of 4 lipids (1 proprietary and 3 commercially available): the proprietary ionizable lipid heptadecan-9-yl 8-((2hydroxyethyl)(6-oxo-6-(undecyloxy)hexyl)amino)octanoate (SM102); cholesterol; 1,2-distearoyl-sn-glycero-3-phosphocholine (DSPC); and 1-monomethoxypolyethyleneglycol-2,3-dimyristylglycerol with PEG of average molecular weight 2000 (1-monomethoxypolyethyleneglycol-2,3-dimyristylglycerol polyethylene glycol 2000 [PEG2000-DMG]). mRNA-1273 injection is provided as a sterile liquid for injection and is a white to off-white dispersion at a concentration of 0.2 mg/mL in 20 mM Tris buffer containing 87 mg/mL sucrose and 4.3 mM sodium acetate at pH 7.5.

**Mode of Administration:**

Each participant will receive 2 doses of IP by IM injection approximately 28 days apart (Day 1 and Day 29) into the deltoid muscle or anterolateral thigh (per investigator's discretion).

**Procedures and Assessments:**

**Safety Assessments:**

Safety assessments will include monitoring and recording of the following for each participant:

- Solicited local and systemic ARs that occur during the 7 days following each injection (ie, the day of injection and 6 subsequent days). Solicited ARs will be recorded daily using electronic diaries (eDiary).
- Unsolicited AEs observed or reported during the 28 days following each injection (ie, the day of injection and 27 subsequent days).
- AEs leading to discontinuation from dosing and/or study participation from Day 1 through the last day of study participation.
- MAAEs from first dose on Day 1 through the entire study period.
- SAEs from first dose on Day 1 through the entire study period.
- AESIs including MIS-C and myocarditis and/or pericarditis, through the entire study period.
- Physical examination findings.
- Assessments for SARS-CoV-2 infection from Day 1 through study completion.
- Details of all pregnancies in female participants will be collected after the start of study treatment and until the end of their participation in the study.

**Blood Collections for Immunogenicity Assessments and Biomarker Samples:**

The following analytes will be measured in blood samples for immunogenicity assessments and biomarker samples:

- Serum neutralizing antibody (nAb) titer against SARS-CoV-2 as measured by pseudovirus and/or live virus neutralization assays.
- Serum binding antibody (bAb) titer as measured by enzyme-linked immunosorbent assay specific to the SARS-CoV-2 S protein.
- For Part 1, testing for serologic markers for SARS-CoV-2 infection using a nonvaccine antigen-based blood test at Day 1 (prior to the first dose), Day 57, Day 209, and Day 394.

For Part 2, participants in each age group will be assigned to 5 phlebotomy cohorts. Blood sampling will be performed in 3 of these 5 cohorts for immunogenicity at 3 of the following time points: Day 1 (prior to randomization and the first dose), Day 57, and one of Day 29 (prior to the second dose), Day 209, and Day 394. Participants in the Cohort D (remainder of the age group) will provide a blood sample at Day 1 (prior to randomization and the first dose) and within 4 days of receiving Dose 2 at Day 30 (+3 days) for storage and potential future biomarker testing (may include cardiac biomarkers). For participants already enrolled in Cohort D prior to amendment 4, this blood draw will be optional and confirmed at time of re-consenting; for all participants newly enrolled into Cohort D under amendment 4, it will be mandatory. A fifth cohort of participants at selected VTEU sites only, will provide blood samples for assessment of exploratory serology and CMI on Day 1 (prior to randomization and the first dose), Day 43, Day 209, and Day 394.

**Inferred Efficacy Assessments:**

Vaccine effectiveness for children 6 months to < 12 years of age will be inferred based on serum antibody responses obtained on Day 57 (28 days after the second injection of mRNA-1273). Inference will be based on assessing the antibody responses against the following:

1. *If available at the time of analysis*, antibody responses will be assessed against an accepted serum antibody threshold conferring protection against COVID-19.
2. *If an accepted threshold of protection is not available*, noninferiority of the geometric mean (GM) value of serum antibody and seroresponse rate of children 6 months old to < 12 years old (Study P204) compared with the GM value of serum antibody and seroresponse rate from young adults (18 to 25 years of age) enrolled in the ongoing clinical endpoint efficacy trial (Study P301) will be assessed.

**Statistical Methods:**

**Hypothesis Testing:**

If an accepted antibody threshold of protection against COVID-19 is established for the primary immunogenicity endpoint, the null hypothesis is that the percentage of participants on mRNA-1273 with serum antibody above the established threshold at Day 57 is ≤ 70% (ie, $H_0$: percentage of participants on mRNA-1273 with serum antibody at Day 57 above the established threshold ≤ 70%).

For each age group, the study will be considered to meet the immunogenicity endpoint if the 95% confidence interval (CI) of percentage of participants on mRNA-1273 rules out 70% (lower bound of the 95% CI is > 70%).

The null hypotheses may be updated when the information on an acceptable antibody threshold becomes available. In this case, the null hypothesis update will be provided in the statistical analysis plan (SAP).

If an accepted serum antibody threshold of protection against COVID-19 is not available for the primary immunogenicity endpoint, immune response as measured by GM value and seroresponse rate in each age group based on Day 57 antibody levels will be compared to

Day 57 antibody levels from young adults (18 to 25 years of age) in Study P301. Noninferiority tests of 2 null hypotheses based on 2 coprimary endpoints will be performed, respectively.

Coprimary endpoint 1: antibody GM value at Day 57

The null hypothesis $H^1_0$: immunogenicity response to mRNA-1273, as measured by antibody GM value at Day 57, is inferior in children (in age groups 6 months to < 2 years, 2 to < 6 years, and 6 to < 12 years) compared with that in young adults (18 to 25 years of age) using mRNA-1273 Study P301 data.

The noninferiority in antibody GM value in an age group in children compared with that in young adults (18 to 25 years of age) is demonstrated by meeting both success criteria:

- The lower bound of the 95% CI of the geometric mean ratio (GMR) ruling out 0.67 (lower bound > 0.67) using a noninferiority margin of 1.5, AND
- The GMR point estimate > 0.8 (minimum threshold).

The GMR is the ratio of the GM value of SARS-CoV-2-specific antibody in children in an age group receiving mRNA-1273 in this Study P204 compared with the GM value of young adults (18 to 25 years of age) receiving mRNA-1273 in Study P301 at Day 57.

Coprimary endpoint 2: antibody seroresponse at Day 57

A definition of seroresponse will be provided in the statistical analysis plan (SAP) based on forthcoming information about assay performance.

The null hypothesis $H^2_0$: immunogenicity response to mRNA-1273 as measured by seroresponse rate at Day 57 is inferior in children compared with that in young adults (18 to 25 years of age) in Study P301.

The noninferiority in seroresponse rate in an age group in children compared with that in young adults (18 to 25 years of age) is demonstrated by meeting both success criteria:

- The lower bound of the 95% CI of the seroresponse rate difference ruling out -10% (ie, lower bound > -10%) using the noninferiority margin of 10%, AND
- The seroresponse rate difference point estimate > -5% (minimum threshold)

The seroresponse rate difference is defined as the seroresponse rate in children receiving mRNA-1273 minus the seroresponse rate in young adults of 18 to 25 years of age receiving mRNA-1273 from Study P301.

The study would be considered to meet the primary immunogenicity endpoint in an age group if the noninferiority in the age group compared with the young adults (18 to 25 years of age) is demonstrated based on both coprimary endpoints.

**Power and Sample Size:**

The initial age groups in Part 1 are for estimation purposes. With 750 participants (approximately 375 participants at each dose level) in the initial 6 to < 12 years age group, there is at least a 90% probability to observe at least 1 participant with an AE at a true AE rate of 1% for a given dose level. In each of the younger age groups (2 to < 6 years and 6 months to < 2 years), the safety assessment will occur during the conduct of Part 2 after approximately 375 participants have been exposed to mRNA-1273 at the dose level selected for Part 2. For further details, please refer to the DSMB Section in this synopsis.

The sample size in the expansion (Part 2) is considered to be sufficient to support a safety database in the pediatric participants 6 months to < 12 years of age. With approximately 3,000 participants each in the 6 to < 12 years, 2 to < 6 years, and 6 months to < 2 years of age groups exposed to mRNA-1273 at a given dose level in Part 2, the study has at least a 95% probability to observe at least 1 participant with an AE at a true 0.1% AE rate for a given dose level.

A subset of Full Analysis Set (FAS) participants (Immunogenicity Subset) in each age group will be selected for measuring immunogenicity data. The immunogenicity samples of the Immunogenicity Subset will be processed, and the analysis of primary immunogenicity endpoint will be based on the Immunogenicity Per-Protocol (PP) Subset.

If a threshold of protection is available, for the primary immunogenicity endpoint, with approximately 289 participants on mRNA-1273 in the Immunogenicity PP Subset of an age group, there will be at least 90% power to rule out 70% with a 2-sided 95% CI (lower bound of the 95% CI > 70%) for the percentage of mRNA-1273 participants exceeding the accepted threshold if the true rate of participants exceeding the threshold is 80%.

If an acceptable antibody threshold of protection against COVID-19 is not available at the time of analysis for the primary immunogenicity endpoint, noninferiority tests of the 2 null hypotheses based on the 2 coprimary endpoints, respectively, will be performed. The sample size calculation for each of the 2 noninferiority tests was performed, and the larger sample size was chosen for the study.

- With approximately 289 participants receiving mRNA-1273 in the Immunogenicity PP Subset of each age group in Study P204 and young adults (18 to 25 years of age) in Study P301, there will be 90% power to demonstrate noninferiority of the immune response, as measured by the antibody GM value, in pediatric population at a 2-sided alpha of 0.05, compared with that in young adults (18 to 25 years of age) in Study P301 receiving mRNA-1273, assuming an underlying GMR value of 1, a noninferiority margin of 0.67 (or 1.5) and a point estimate minimum threshold of 0.8. The standard deviation of the natural log-transformed levels is assumed to be 1.5.

- With approximately 289 participants receiving mRNA-1273 in the Immunogenicity PP Subset of each age group in Study P204 and young adults (18 to 25 years of age) in Study P301, there will be at least 90% power to demonstrate noninferiority of the immune response as measured by seroresponse rate in children at a 2-sided alpha of 0.05, compared with that in young adults 18 to 25 years of age in Study P301 receiving mRNA-1273, assuming seroresponse rate of 85% in young adults of 18 to 25 years of age from Study P301, true seroresponse rate of 85% in children (or true rate difference is 0 compared to young adults from Study P301), a noninferiority margin of 10% and a point estimate minimum threshold of -5% in seroresponse rate difference.
- In the 1273-P203 interim analysis (data snapshot on 08 May 2021), the observed seroresponse rates at Day 57 were high in both 18 to 25 years of age group from Study P301 (98.6% with 95% CI: 96.6, 99.6) and 12 to17 years of age group (98.8% with 95% CI: 97.0, 99.7), based on pseudovirus nAb ID50 titers. For this Study P204, if the true seroresponse rates were assumed to be 95% or higher in both 18 to 25 years of age group from Study P301 and an age group in children, with between-group true difference within 4%, there will be a >90% power to demonstrate noninferiority by seroresponse rate in children compared with 18 to 25 years of age in Study P301, at a 2-sided alpha of 0.05

Assuming approximately 25% of participants in the Immunogenicity Subset will not meet the criteria to be included in the Immunogenicity PP Subset, approximately 528 participants in Part 2 (~396 receiving mRNA-1273 and ~132 receiving placebo) will be selected for the Immunogenicity Subset from which approximately 289 participants on mRNA-1273 will be suitable for the Immunogenicity PP Subset.

**Analysis Sets:**

The analysis sets are defined in the following table:

| Analysis Set | Description |
|---|---|
| Randomization Set | Part 2: All participants who are randomly assigned, regardless of the participants' treatment status in the study. |
| Full Analysis Set (FAS) | Part 1: All enrolled participants in Part 1 who receive at least 1 injection of IP<br>Part 2: All randomly assigned participants who receive at least 1 injection of IP |

| | |
|---|---|
| Per-Protocol (PP) Set | All participants in the FAS who receive planned doses of IP per schedule, comply with the immunogenicity schedule, and have no major protocol deviations that impact key or critical data. Participants who are RT-PCR positive or seropositive at baseline will be excluded from the PP Set. The PP Set in Part 1 will be used for all analyses of immunogenicity in Part 1 unless specified otherwise. |
| Immunogenicity Subset | A subset of participants in the FAS will be selected for immunogenicity sampling and testing. |
| Per-Protocol Immunogenicity Subset | A subset of participants in the FAS will be selected for immunogenicity testing. The PP Immunogenicity Subset includes participants selected for the Immunogenicity Subset who receive planned doses of IP per schedule, comply with immunogenicity testing schedule, and have no major protocol deviations that impact key or critical data. Participants who are RT-PCR positive or seropositive at baseline will be excluded from the PP Immunogenicity Subset, in addition to participants with HIV who are receiving highly active anti-retroviral therapy (HAART). The PP Immunogenicity Subset will be used for all analyses of immunogenicity unless specified otherwise. |
| Safety Set | All enrolled participants (in Part 1) and all randomly assigned participants (in Part 2) who receive at least 1 dose of IP. The Safety Set will be used for all analyses of safety except for solicited ARs. |
| Solicited Safety Set | The Solicited Safety Set consists of participants in the Safety Set who contributed any solicited AR data.<br>The Solicited Safety Set will be used for the analyses of solicited ARs. |
| Modified Intent-to-Treat (mITT) Set | All participants in the FAS who have no serologic or virologic evidence of prior SARS-CoV-2 infection before the first dose of IP (both negative RT-PCR test for SARS-CoV-2 and negative serology test based on bAb specific to SARS-CoV-2 nucleocapsid) at Baseline |
| Modified Intent-to-Treat-1 (mITT1) Set | All participants in the mITT Set excluding those who received the wrong treatment (ie, at least 1 dose received that is not as randomized) |

Abbreviations: AR = adverse reaction; bAb = binding antibody; COVID-19 = coronavirus disease 2019; IP = investigational product; RT-PCR = reverse transcriptase polymerase chain reaction; SARS-CoV-2 = severe acute respiratory syndrome coronavirus 2.

**Safety Analyses:**

All safety analyses will be based on the Safety Set, except summaries of solicited ARs, which will be based on the Solicited Safety Set. All safety analyses will be provided by vaccination group (dose levels of mRNA-1273 and placebo) and by age group. Participants in Part 1 and Part 2 of the study and in different age groups who receive the same mRNA-1273 dose level may be combined for safety analysis.

Safety and reactogenicity will be assessed by clinical review of all relevant parameters including solicited ARs (local and systemic events), unsolicited AEs, SAEs, MAAEs, AESIs, AEs leading to discontinuation, and physical examination findings.

The number and percentage of participants with any solicited local AR, with any solicited systemic AR, and with any solicited AR during the 7-day follow-up period after each injection by toxicity grade will be provided. A 2-sided 95% exact CI using the Clopper-Pearson method will also be provided for the percentage of participants with any solicited AR.

The number and percentage of participants with unsolicited AEs, SAEs, MAAEs, severe AEs, and AEs leading to discontinuation from IP or withdrawal from the study will be summarized. Unsolicited AEs will be presented by Medical Dictionary for Regulatory Activities preferred term and system organ class.

The number of events of solicited ARs, unsolicited AEs/SAEs, and MAAEs will be reported in summary tables accordingly.

For all other safety parameters, descriptive summary statistics will be provided. Further details will be described in the SAP.

**Immunogenicity Analyses:**

The primary analysis population for immunogenicity will be the Immunogenicity PP Subset, unless specified otherwise. The primary objective of this study is to use the immunogenicity response to infer efficacy in participants aged 6 months to < 12 years at the dose level selected for expansion. For this objective, analyses of immunogenicity will be performed for each pediatric age group separately at the selected dose level based on the participants in the Immunogenicity PP Subset. For each pediatric age group, participants in Part 2 in the Immunogenicity PP Subset may be used for immunogenicity primary analysis. Participants from Part 1 and Part 2 who receive the same mRNA-1273 dose level selected for expansion may be combined for immunogenicity analyses. If the same dose level is selected for expansion for more than one age group, these age groups may be combined for immunogenicity analyses.

An accepted antibody threshold of protection against COVID-19 may be available based on data from other mRNA-1273 studies or external data. If such a threshold of protection against

COVID-19 is available, the number and percentage of participants with antibody greater than or equal to the threshold at Day 57 will be provided with a 2-sided 95% CI using the Clopper-Pearson method. For an age group, if the lower bound of the 95% CI on the mRNA-1273 group is > 70%, the primary immunogenicity endpoint of this study will be considered to be met for that age group.

The number and percentage of participants with serum antibody greater than or equal to the threshold with 2-sided 95% CI will be provided at each postbaseline time point. The CI will be calculated using the Clopper-Pearson method.

If an accepted serum antibody threshold of protection against COVID-19 is not established, immune response as measured by GM value and seroresponse rate in each age group based on Day 57 antibody levels will be compared to that in young adults (18 to 25 years of age) using data from Study P301. An analysis of covariance model will be carried out with antibody at Day 57 as dependent variable and a group variable (a pediatric age group in Study P204 versus young adults [18 to 25 years of age] in Study P301) as the fixed variable for each pediatric age group. The GM values of the pediatric age group at Day 57 will be estimated by the geometric least square mean (GLSM) from the model. The GMR (ratio of GM values) will be estimated by the ratio of GLSM from the model. The corresponding 2-sided 95% CI will be provided to assess the difference in immune response between the pediatric age group (Study P204) compared to the young adults (18 to 25 years of age) in Study P301 at Day 57. For each pediatric age group, the noninferiority of GM value will be considered demonstrated if:

- The lower bound of the 95% CI of the GMR is > 0.67 based on the noninferiority margin of 1.5, AND
- The GMR point estimate > 0.8 (minimum threshold).

The number and percentage of participants with seroresponse due to vaccination will be provided with 2-sided 95% CI using the Clopper-Pearson method at each postbaseline time point with Day 57 being the primary interest. The seroresponse rate difference with 95% CI at Day 57 will be provided between children receiving mRNA-1273 in Study P204 and young adults of 18 to 25 years of age receiving mRNA-1273 from Study P301. For each pediatric age group, the noninferiority of seroresponse rate will be considered demonstrated if:

- The lower bound of the 95% CI of the seroresponse rate difference is > -10% based on the noninferiority margin of 10%, AND
- The seroresponse rate difference point estimate > -5% (minimum threshold).

In addition, the GM value of anti-SARS-CoV-2-specific antibody with corresponding 95% CI will be provided at each time point. The 95% CIs will be calculated based on the t-distribution of the log-transformed values then back transformed to the original scale. For each age group, the geometric mean fold-rise of specific nAb and bAb with corresponding 95% CI at each postbaseline time point over preinjection baseline at Day 1 will be provided. Descriptive summary statistics including median, minimum, and maximum will also be provided.

**Multiplicity adjustment between age groups:**

A sequential hypothesis testing (fixed-sequence method) will be used to adjust multiplicity to preserve the family-wise Type I error rate (alpha = 0.05), starting with the oldest age group, followed by the middle age group, and then followed by the youngest age group. The immunogenicity coprimary endpoint hypotheses for the oldest age group (6 to < 12 years of age) will be tested first at alpha level of 0.05. If the testing in the oldest age group is statistically significant (meeting the noninferiority success criteria of the coprimary endpoints), the alpha level of 0.05 will be passed to the testing of the coprimary endpoint hypotheses in the middle age group (2 to < 6 years of age). If the testing in the middle age group is statistically significant at alpha level of 0.05, then the alpha 0.05 is preserved for testing the coprimary endpoint hypotheses in the youngest age group (6 months to < 2 years of age). The testing will continue through the sequence only until an endpoint in an age group is not statistically significant (did not meet noninferiority success criteria of any primary endpoint), in which case the testing will stop.

**Inferred Efficacy Analyses:**

To evaluate the incidence of COVID-19 after vaccination with mRNA-1273 or placebo, the incidence rate will be provided by vaccination group, dose level, and age group, calculated as the number of cases divided by the total person-time. Participants in Part 1 and Part 2 of the study and in different age groups who receive the same mRNA-1273 dose level (if applicable) may be combined in the analysis.

For serologically confirmed SARS-CoV-2 infection or COVID-19, regardless of symptomatology or severity, infection rate will be provided by vaccination group, dose level, and age group. The same analysis will be conducted for asymptomatic SARS-CoV-2 infection.

The secondary efficacy analyses will be performed in the PP Set, with sensitivity analyses in FAS, mITT Set, and mITT1 Set. Analyses of the efficacy endpoints in Part 2 will be performed for the randomized blinded phase. Additional exploratory analyses will be conducted in the blinded and unblinded phases for participants randomized to mRNA-1273 in Part 2, and in the unblinded phase for participants who are originally randomized to the placebo arm and cross-over to mRNA-1273 after use of any COVID-19 vaccine is authorized or licensed for the participant’s age group.

**Study Analyses:**

**Interim Analyses:**

Part 1: Interim analyses may be performed after all or a subset of participants (with immunogenicity samples collected for D1 and D57) have completed Day 57 within an age group in Part 1 (one optional interim analysis for each age group, 3 interim analyses total for the 3 age groups in Part 1). Analyses of safety and immunogenicity may be conducted at each interim analysis.

Part 2: An interim analysis of immunogenicity and safety will be performed after all or a subset of participants have completed Day 57 (1 month after the second dose) in Part 1 or Part 2 within an age group. This interim analysis will be considered the primary analysis of immunogenicity for a given age group. Another interim analysis on safety may be performed after a different subset or all participants have completed Day 57 in an age group.

**Final Analysis:**

The final analysis of all endpoints will be performed after participants have completed all planned study procedures. Results of this analysis will be presented in a final clinical study report, including individual listings.

Additional information about all study analyses may be provided in the SAP.

#### LIST OF FIGURES

Figure 1: Study Schema....55

Figure 2: Surveillance for COVID-19 Symptoms and the Corresponding Clinical Data Pathways ....................................................................................................................91

### LIST OF ABBREVIATIONS AND TERMS

The following abbreviations and terms are used in this study protocol.

| Abbreviation or Specialist Term | Definition |
|---|---|
| AE | adverse event |
| AESI | adverse event of special interest |
| AR | adverse reaction |
| bAb | binding antibody |
| BMI | body mass index |
| CD | cluster of differentiation |
| CDC | US Centers for Disease Control and Prevention |
| CEAC | Cardiac Event Adjudication Committee |
| CFR | Code of Federal Regulations |
| CI | confidence interval |
| CMI | cell-mediated immunity |
| CoV | coronavirus |
| COVID-19 | coronavirus disease 2019 |
| CRF | case report form |
| CRO | contract research organization |
| CSR | clinical study report |
| DMID | Division of Microbiology and Infectious Diseases |
| DSMB | Data Safety Monitoring Board |
| DSPC | 1,2-distearoyl-sn-glycero-3-phosphocholine |
| eCRF | electronic case report form |
| EDC | electronic data capture |
| eDiary | electronic diary |
| ERD | enhanced respiratory disease |
| FAS | Full Analysis Set |
| FDA | US Food and Drug Administration |
| GCP | Good Clinical Practice |

| Abbreviation or Specialist Term | Definition |
|---|---|
| GLSM | geometric least squares mean |
| GM | geometric mean |
| GMR | geometric mean ratio |
| GMT | geometric mean titer |
| HCP | healthcare practitioner |
| HIV | human immunodeficiency virus |
| IB | investigator’s brochure |
| ICF | informed consent form |
| ICH | International Council for Harmonisation |
| ICU | intensive care unit |
| IgG | immunoglobulin G |
| IM | intramuscular(ly) |
| IND | investigational new drug |
| IP | investigational product |
| IRB | institutional review board |
| IST | internal safety team |
| LAR | legally acceptable representative |
| LNP | lipid nanoparticle |
| LOD | limit of detection |
| LTFU | lost to follow-up |
| MAAE | medically attended adverse event |
| MedDRA | Medical Dictionary for Regulatory Activities |
| MERS | Middle East respiratory syndrome |
| MIS-C | multisystem inflammatory syndrome in children |
| mRNA | messenger RNA |
| nAb | neutralizing antibody(ies) |
| NHP | nonhuman primate |
| NIAID | National Institute of Allergy and Infectious Diseases |

| **Abbreviation or Specialist Term** | **Definition** |
|---|---|
| PEG | polyethylene glycol |
| PEG2000-DMG | 1-monomethoxypolyethyleneglycol-2,3-dimyristylglycerol polyethylene glycol 2000 |
| PP | per protocol |
| QA | quality assurance |
| RT-PCR | reverse transcriptase polymerase chain reaction |
| S | spike |
| S2P | S protein stabilized with 2 proline mutations |
| SAE | serious adverse event |
| SAP | statistical analysis plan |
| SARS | severe acute respiratory syndrome |
| SARS-CoV-2 | severe acute respiratory syndrome coronavirus 2 |
| SM-102 | heptadecan-9-yl 8-((2-hydroxyethyl)(6-oxo-6-(undecyloxy)hexyl)amino)octanoate |
| SoA | schedule of assessments |
| Study P301 | Study mRNA-1273-P301; NCT04470427 |
| Th | T helper cell |
| VTEU | Vaccine and Treatment Evaluation Units |
| WHO | World Health Organization |
| WOCBP | woman of childbearing potential |

### 1. INTRODUCTION

#### 1.1. Study Rationale

Coronaviruses (CoVs) are a large family of viruses that cause illness ranging from the common cold to more severe diseases, such as Middle East respiratory syndrome (MERS) and severe acute respiratory syndrome (SARS). Coronaviruses are zoonotic, meaning that they are transmitted between animals and people. An outbreak of the coronavirus disease 2019 (COVID-19) caused by severe acute respiratory syndrome coronavirus 2 (SARS-CoV-2) began in Wuhan, Hubei Province, China in December 2019 and has spread throughout China and to over 215 other countries, territories, and areas, including the United States (WHO 2020a). On 11 Mar 2020, the World Health Organization (WHO) officially declared COVID-19 a pandemic. As of 30 Jan 2021, the WHO dashboard (WHO 2020b) reported that there have been more than 2.1 million COVID-19 deaths worldwide. As of 29 Jan 2021, the US Centers for Disease Control and Prevention (CDC) reported over 25 million cases of COVID-19 in all 50 states and 5 jurisdictions, with 431,619 attributed and probable deaths (CDC 2020a). While the CDC reports that the highest risk of disease burden is in older adults and populations with certain underlying comorbid conditions such as heart disease, diabetes, and lung disease, a substantial burden in children is now being recognized. Evidence is emerging (described below) to suggest that children < 18 years of age, particularly adolescents, may be disproportionately contributing to the number of new cases as schools re-open at varying degrees of in-person learning. As of 29 Jan 2021, the CDC reported 2,125,186 cases of COVID-19 in children less than 18 years of age (11.1% of all US cases) and 267 deaths (approximately 0.1% of all US deaths; CDC 2020b).

During incubation, those infected can also transmit the virus before developing symptoms (Chen et al 2020). Person-to-person transmission occurs primarily via direct contact or through droplets spread by coughing or sneezing from an infected individual, whether symptomatic or not (Chen et al 2020; Licciardi et al 2020; Rothan and Byrareddy 2020; Shen et al 2020). SARS-CoV-2 can also be transmitted via the fecal-oral pathway (Cruz and Zeichner 2020).

During the COVID-19 pandemic, children throughout much of the world have had school attendance limited in an attempt to control infection. Therefore, the main source of infection for SARS-CoV-2 in children, with or without clinical symptoms, is infected household contacts. Indeed, a retrospective cohort study of high school students, parents and siblings of students, teachers, and staff conducted in France in early April 2020 suggests that there was little to no transmission from infected students to other students or school staff. Rather, a high prevalence of antibodies against SARS-CoV-2 among families suggests familial clustering of COVID-19 cases (Fontanet et al 2020).

A recent report of COVID-19 trends in school-aged children in the United States from 01 Mar 2020 to 19 Sep 2020 indicates that 37% of laboratory-confirmed cases of COVID-19 in school-aged children occurred in children 5 to 11 years of age while 63% occurred in adolescents 12 to 17 years of age (Leeb et al 2020). The weekly incidence among adolescents was 37.4 cases per 100,000 compared with 19.0 cases per 100,000 for younger children. Among school-aged children with laboratory-confirmed COVID-19, 58% reported at least one symptom and 5% reported no symptoms, although information on symptoms was missing or unknown for 37%. Overall, 1.2% of school-aged children with COVID-19 were hospitalized, 0.1% required intensive care unit (ICU) admission, and < 0.01% died of COVID-19. Furthermore, at least one underlying condition was reported in 3% of adolescents and 2% of younger children. Chronic lung disease, including asthma, was most commonly reported (55%), followed by disability (neurologic or neurodevelopmental disorders, intellectual or physical disability, and vision or hearing impairment; 9%), immunosuppressive conditions (7%), diabetes (6%), psychological conditions (6%), cardiovascular disease (5%), and severe obesity (4%) (Leeb et al 2020). Based on the COVID-NET report, in 42.3% of children with at least 1 underlying condition, the most prevalent conditions were obesity (37.8%), chronic lung disease (18.0%), and prematurity (15.4%) (Kim et al 2020). Of particular interest is the phenomenon known as multisystem inflammatory syndrome in children (MIS-C) (PICS 2020). Since those early reports, a number of articles have been published describing a hyperinflammatory syndrome with features of Kawasaki disease in children and adolescents infected with SARS-CoV-2 (Belhadjer et al 2020; Dufort et al 2020; Verdoni et al 2020). Targeted surveillance in the United States from March through May 2020 revealed 186 patients across 26 states who met a pre-specified case definition of MIS-C (Feldstein et al 2020). The median age was 8.3 years (interquartile range: 3.3 to 12.5 years). Most (73%) were previously healthy, and 70% were positive for SARS-CoV-2 by reverse transcriptase polymerase chain reaction (RT-PCR) or antibody testing. The condition affected a variety of organ systems, most commonly the gastrointestinal, cardiovascular, hematologic, mucocutaneous, and respiratory systems. More recently, the Brighton Collaboration has drafted a manuscript proposing case definitions of MIS-C based on 5 levels of diagnostic certainty (Vogel et al 2020).

Evidence suggests that there is substantial burden of COVID-19 in younger age groups. Another study examined the age distribution of COVID-19 in the United States from May to August 2020 based on 3 indicators: COVID-19-like illness-related emergency department visits, positive RT-PCR results for SARS-CoV-2, and confirmed COVID-19 cases (Boehmer et al 2020). These authors report an estimated mean COVID-19 incidence during this time period of 179.3 cases per 100,000 in individuals 10 to 19 years of age. Finally, a recent report describes an adolescent (13-year-old female), whose only symptom was nasal congestion, yet she was the index case in an outbreak of COVID-19 linked to a family gathering that ultimately crossed 4 states and included 5 households and 11 individuals (Schwartz et al 2020). A June COVID-19 outbreak in a Georgia overnight camp demonstrated that children 6 to 19 years of age are susceptible to SARS-CoV-2 infection and transmission (Szablewski et al 2020). In addition, in the second half of July, Rhode Island childcare programs reported 52 confirmed and probable childcare-associated COVID-19 cases, 30 (58%) cases of which were among children with a median age of 5 years (Link-Gelles et al 2020). This suggests that adolescents and children can serve as the source of COVID-19 outbreaks, even when their symptoms are mild, as in these cases.

There is currently no approved vaccine against SARS-CoV-2 for children. In December 2020, following review of safety and efficacy data observed to date, the Food and Drug Administration (FDA) granted Emergency Use Authorization (EUA) to 2 messenger RNA (mRNA)-based SARS-CoV-2 vaccines, including mRNA-1273, for adults. To address prevention of pediatric COVID-19 as well as to potentially help curb SARS-CoV-2 transmission, there is an urgent public health need for rapid development of SARS-CoV-2 vaccines in children.

The objective for this Phase 2/3 study is to evaluate the safety, tolerability, reactogenicity, and effectiveness of up to 3 dose levels (25, 50, and 100 µg) of mRNA-1273 vaccine administered as 2 doses 28 days apart (Section 3.1) to healthy children 6 months to < 12 years of age divided into 3 age groups (6 to < 12 years, 2 to < 6 years, and 6 months to < 2 years). Another Phase 2/3 study to evaluate the safety and reactogenicity of a single dose level (100 µg) of mRNA-1273 vaccine administered as 2 doses 28 days apart to an adolescent population (12 to < 18 years of age) is ongoing.

#### 1.2. Background and Overview

The Sponsor has developed a rapid-response proprietary vaccine platform based on a mRNA delivery system. The platform is based on the principle and observations that cells in vivo can take up mRNA, translate it, and then present viral antigen(s) on the cell surface. The delivered mRNA does not enter the cellular nucleus or interact with the genome, is nonreplicating, and is expressed transiently. Messenger RNA vaccines have been used to induce immune responses against infectious pathogens such as cytomegalovirus (NCT03382405), human metapneumovirus and parainfluenza virus type 3 (NCT03392389), and influenza virus (NCT03076385 and NCT03345043).

The Sponsor is using its mRNA-based platform to develop a novel lipid nanoparticle (LNP)-encapsulated mRNA-based vaccine against SARS-CoV-2 (mRNA-1273). mRNA-1273 encodes for the full-length spike (S) protein of SARS-CoV-2, modified to introduce 2 proline residues to stabilize the S protein (S2P). It has been confirmed that the stabilized SARS-CoV-2 S2P expresses well and is in the prefusion conformation (Wrapp et al 2020). The CoV S protein mediates attachment and entry of the virus into host cells (by fusion), making it a primary target for neutralizing antibodies (nAb) that prevent infection (Corti et al 2015; Wang et al 2015;

Yu et al 2015; Johnson et al 2016; Chen et al 2017; Wang et al 2018; Kim et al 2019; Widjaja et al 2019; Corbett et al 2020a; Ju et al 2020; Robbiani et al 2020).

##### 1.2.1. Nonclinical Studies

The National Institute of Allergy and Infectious Diseases (NIAID) Vaccine Research Center and the Sponsor performed nonclinical studies in young and aged wild-type mice, Syrian Golden hamsters, and rhesus macaques (nonhuman primates [NHPs]) to evaluate dose-ranging responses to mRNA-1273 (immunogenicity) and high-dose virus SARS-CoV-2 challenge (protection) and to address the theoretical concern of enhanced respiratory disease (ERD) mediated by vaccine-induced antibody responses and/or T helper cell (Th) 2-directed T-cell responses observed with other vaccines against viral respiratory diseases (Graham 2020).

Nonclinical animal studies demonstrated that mRNA-1273 is immunogenic in all species assessed, with a dose-dependent response in immunoglobulin G (IgG) binding antibody (bAb) titers and a correlation that is statistically significant between bAb and (nAb) activity. In addition, antigen-specific T-cell responses were observed in studies in mice and NHPs. The Th1-directed cluster of differentiation (CD) 4 and CD8 T-cell responses were measured after boost in animals that were vaccinated with mRNA-1273. In various animal models, immunological measurements suggested that Th1 responses predominated, IgG2a/c/IgG1 ratios were favorable, and high levels of SARS-CoV-2 nAb were observed, suggesting that ERD after mRNA-1273 administration would be unlikely. In addition to measuring the immune response, mice, hamsters, and NHPs were challenged with high-dose SARS-CoV-2 virus. In these studies, dose levels were included that were predicted to be optimal (fully protective) and suboptimal (subprotective). At higher doses, mice and NHPs were fully protected from viral replication in both lungs and nasal passages. At subprotective dose levels, animals either remained fully protected or had reduced viral lung burden post challenge versus control animals (Corbett et al 2020a; Corbett et al 2020b).

Overall, nonclinical animal studies demonstrated that mRNA-1273 is safe and well tolerated, is immunogenic, fully protects animals from challenge at optimal dose levels, and does not result in ERD at protective or subprotective dose levels.

In support of the development of mRNA-1273 for prophylaxis against SARS-CoV-2 infection, nonclinical immunogenicity, biodistribution, and safety studies have been completed with similar mRNA-based vaccines formulated in LNPs containing heptadecan-9-yl 8-((2-hydroxyethyl)(6-oxo-6-(undecyloxy)hexyl)amino)octanoate (SM-102), the novel proprietary lipid used in the mRNA-1273 LNP formulation.

A detailed review of nonclinical experience with mRNA-1273 vaccine is provided in the investigator's brochure (IB).

##### 1.2.2. Clinical Studies

The mRNA-1273 vaccine is currently being evaluated in 4 ongoing trials.

The first is a safety and immunogenicity Phase 1 study (NCT04283461) sponsored and conducted by the Division of Microbiology and Infectious Diseases (DMID; investigational new drug [IND] application 019635) of the NIAID. The Phase 1 study is an open-label dose-ranging study of mRNA-1273 in healthy adult male and nonpregnant female participants in 3 age groups: 18 to 55 years, inclusive (60 participants); 56 to 70 years, inclusive (30 participants); and ≥ 71 years (30 participants). Participants were randomly assigned to 1 of 4 dose levels of mRNA-1273: 25 µg, 50 µg, 100 µg, and 250 µg. Each participant received the same dose by intramuscular (IM) injection (0.5 mL) of mRNA-1273 on Days 1 and 29 in the deltoid muscle. Blood samples were obtained at baseline, Days 8, 15, 29 (prior to Dose 2), 36, 43, and 57, as well as 3, 6, and 12 months after the second vaccination (Days 119, 209, and 394, respectively). Safety monitoring is ongoing for 12 months after the second injection.

On 14 Jul 2020, a preliminary report of findings in this Phase 1 study through Day 57 for the 18- to 55-year age-cohort (25, 100, and 250 µg dosage groups) was published (Jackson et al 2020). After the second injection, serum viral neutralizing activity was detected by 2 methods in all 42 participants evaluated (of 45 enrolled), with values that were comparable to or greater than the geometric mean titers (GMT) measured in the convalescent serum samples. Regarding safety, no serious adverse events (SAEs) were reported and no study-halting rules were triggered. In general, solicited systemic adverse reactions (ARs) were more common after the second injection. Solicited ARs that occurred in more than half the participants included fatigue, chills, headache, myalgia, and pain at the injection site. While none of the participants (N= 45) at any dose level experienced fever following the first dose, mild fever was observed in 5 participants (33%), moderate fever was observed in 1 participant (6.7%), and no participants experienced severe fever at 100 µg following the second dose. Data on 40 older adults > 55 years of age who received 2 doses of either 25 or 100 µg in the same Phase 1 DMID study were recently published (Anderson et al 2020). After the second injection, serum neutralizing activity was detected in all participants by multiple methods with binding and neutralizing antibody titers similar to those reported in adults 18 to 55 years of age and above the median for convalescent serum. Solicited ARs were predominantly mild or moderate in severity and most frequently included fatigue, chills, headache, myalgia, and pain at the injection site. Specifically, regarding fever, no participant reported fever of any severity following the first injection. Following the second injection, 2 participants in the 100-µg dose group reported fever categorized as mild. All participants have been vaccinated, and safety and immunogenicity follow-up is ongoing. Indeed, immunogenicity data through 119 days after the first vaccination for 34 participants revealed binding and neutralizing GMTs that exceeded the median GMTs in a panel of convalescent sera from

41 controls. No SAEs were noted, no study-halting rules were met, and no new related adverse events (AEs) were reported after Day 57 (Widge et al 2021).

Additionally, an ongoing, placebo-controlled, dose-finding Phase 2a study (mRNA-1273-P201; NCT04405076) conducted by the Sponsor under IND 19745 aims to evaluate the safety, reactogenicity, and immunogenicity of mRNA-1273 administered as 2 doses 28 days apart at dose levels of 50 and 100 µg. The study is being conducted in 600 healthy adults in 2 age cohorts: 18 to 54 years of age (300 participants) and at least 55 years of age (300 participants). All participants were randomly assigned in a ratio of 1:1:1 to receive either placebo or mRNA-1273 at 1 of 2 doses, either 50 µg or 100 µg. The study was designed to begin with parallel enrollment of all 300 participants in Cohort 1 (≥ 18 to < 55 years old) and a sentinel group of 50 participants in Cohort 2 (≥ 55 years old). An independent safety monitoring committee (SMC) reviewed all blinded and unblinded safety data through Day 57 (1 month after Dose 2) for both cohorts and found the vaccine tolerable, saw no safety concerns, and recommended continuing the study as planned. The study is now completely enrolled, and all dosing is complete; participants are undergoing additional serologic testing and safety follow-up.

The 100 µg dose level is currently being investigated in a large Phase 3 efficacy study (mRNA-1273-P301; NCT04470427) in approximately 30,000 adults 18 years of age and older, randomly assigned 1:1 to receive either vaccine or placebo. Recently published data show a vaccine efficacy of 94.1% against symptomatic COVID-19 illness with onset at least 14 days after the second injection (Baden et al 2021). Although moderate transient reactogenicity occurred more frequently in the mRNA-1273 group, SAEs were rare and the incidence was similar between the 2 groups. On 18 Dec 2020, the Moderna COVID-19 vaccine was authorized by the US FDA for emergency use in individuals ≥ 18 years of age. The participants in this Phase 3 study are currently undergoing assessments for long-term safety and durability of vaccine efficacy.

Finally, a Phase 2/3, randomized, observer-blind, placebo-controlled study (mRNA-1273-P203) conducted by the Sponsor under IND 19745 is evaluating the safety, reactogenicity, and effectiveness of mRNA-1273 in healthy adolescents 12 to < 18 years of age. This study began enrolling in December 2020 and will enroll 3,000 participants randomly assigned 2:1 to receive mRNA-1273 100 µg or placebo administered as 2 doses 28 days apart. Participants will be followed for 12 months after the second dose.

A detailed review of the clinical experience with LNPs containing SM-102 (mRNA vaccines and placebo) is provided in the IB.

#### 1.3. Benefit/Risk Assessment

##### 1.3.1. Potential Benefits From Participation

The following benefits may accrue to participants:

- The mRNA-1273 vaccine may be an effective vaccine against COVID-19 in the study population.
- Participants will have a baseline (Day 1) evaluation for SARS-CoV-2 infection and ongoing surveillance for COVID-19 throughout the study.
- The study will contribute to the development of a vaccine against COVID-19 for children 6 months to < 12 years of age.

##### 1.3.2. Risks From Study Participation and Their Mitigation

Immediate systemic allergic reactions (eg, anaphylaxis) can occur following any vaccination. These reactions are very rare and are estimated to occur once per 450,000 vaccinations for vaccines that do not contain allergens such as gelatin or egg protein (Zent et al 2002). As a precaution, all participants will remain under observation at the study site for at least 30 minutes after injection.

Vasovagal syncope (fainting) can occur before or after any vaccination, is usually triggered by the pain or anxiety caused by the injection and is not related to the substance injected. Therefore, it is important that standard precautions and procedures are followed to avoid injury from fainting.

Intramuscular injection with other mRNA vaccines manufactured by the Sponsor containing the SM-102 LNP commonly results in a transient and self-limiting local inflammatory reaction. This typically includes pain, erythema (redness), or swelling (hardness) at the injection site, which are mostly mild to moderate in severity and usually occur within 24 hours of injection. In the large Phase 3 efficacy study of mRNA-1273 described above, delayed injection site reactions occurred with an incidence of approximately 1%. These reactions typically began 8 days or more following injection, were mostly not severe, and were rarely observed after the second dose.

The majority of local and systemic solicited ARs observed after injection with mRNA-1273 at the 100-µg dose level have been mild to moderate in severity (Section 1.2.2). The most commonly reported systemic ARs were headache, myalgia, fatigue, chills, and fever. In the majority of cases, the reactions resolved spontaneously within several days.

Laboratory abnormalities (including increases in hepatic enzymes and serum lipase levels) following injection were observed in clinical studies with similar mRNA-based vaccines. These abnormalities were without clinical symptoms or signs and returned toward baseline (Day 1) values over time. The clinical significance of these observations is unknown. Further details are provided in the current IB.

There is a theoretical risk that active vaccination to prevent SARS-CoV-2 infection may cause a paradoxical increase in the risk of severe COVID-19. This possibility is based on the rare phenomenon of vaccine-associated ERD, which was first seen in the 1960s with 2 vaccines made in the same way (formalin-inactivated whole virus) and designed to protect children against infection with respiratory syncytial virus (Chin et al 1969) or measles virus (Fulginiti et al 1967). It is noteworthy that these vaccines were the result of a completely different formulation and with an entirely different mechanism of action than mRNA-based vaccines such as mRNA-1273. Disease enhancement has also been proposed as a possible explanation for cases of more serious disease associated with dengue vaccination (Thomas and Yoon 2019; WHO 2019). It is not known if mRNA-1273 will increase the risk of enhanced disease; however, preliminary data from the ongoing Phase 3 study suggests no evidence of enhanced disease, as fewer cases of severe COVID-19 and COVID-19 were observed in participants who received mRNA-1273 than in those who received placebo.

In order to address this theoretical risk, animal studies have been performed in young and aged wild-type mice and rhesus macaques (NHPs). These studies were designed to capture immunogenicity endpoints that would be predictive of ERD and also to evaluate if, at protective or subprotective dose levels of mRNA-1273, evidence of disease enhancement would be observed after challenge of the animals with SARS-CoV-2. These nonclinical studies demonstrated that mRNA-1273 is safe and well tolerated in different animal species; is immunogenic; drives a robust SARS-CoV-2-specific Ab, neutralization, and Th1-directed CD4 T-cell response; fully protects animals from challenge at dose levels as low as 1 µg/dose in mice and 30 µg/dose in NHPs; and does not lead to ERD at protective or subprotective dose levels (Corbett et al 2020a; Corbett et al 2020b). Clinical immunogenicity data from the DMID Phase 1 study of mRNA-1273 demonstrated high levels of nAbs and Th1-polarized CD4 T-cell responses (Jackson et al 2020), consistent with the immunogenicity observed in the nonclinical studies. These data suggest that the risk of paradoxical ERD, while not eliminated, is likely to be low.

In the context of the EUA of mRNA-1273 for individuals aged 18 years and older, there have been very rare reports of myocarditis and pericarditis occurring after vaccination with Moderna COVID-19 Vaccine. Although causality has not been established, the majority of the cases have been reported in young males shortly after the second dose of the vaccine. These are typically mild cases and individuals tend to recover within a short time following standard treatment and rest (Gargano et al, 2021).

##### 1.3.3. Overall Benefit/Risk Conclusion

Based on the Phase 3 data and the results of the Phase 1 and Phase 2 studies described above, the Sponsor intends to study 3 dose levels (25, 50, and 100 µg) in the proposed Phase 2/3 study in participants 6 months to < 12 years of age. Briefly, the proposed study is designed to dose escalate and age de-escalate through 3 sequential age groups (6 to < 12 years, 2 to < 6 years, and 6 months to < 2 years). Participants at each dose level will receive 2 doses at that dose level approximately 28 days apart. The mRNA-1273 dose levels that will be evaluated in each age group are given in Table 1.

**Table 1: Age Groups and mRNA-1273 Dose Levels**

| Age Group | mRNA-1273 Dose Levels Planned to be Evaluated |
|---|---|
| 6 to < 12 year | 50 and 100 µg |
| 2 to < 6 year | 25[1], 50, and 100 µg |
| 6 months to < 2 year | 25 and 50 µg |

[1] The 25-µg dose will be evaluated if 100 µg is eliminated at any point during the dose escalation process, to maintain dose ranging for this age group.

The study will be conducted in 2 parts. Part 1 of the study will be open-label and consist of dose-escalation and age de-escalation to select the dose for each age group. Part 2 of the study will be a placebo-controlled observer-blind evaluation of the selected dose in each age group.

Immunogenicity data from participants who receive the mRNA-1273 vaccine at the selected dose level will be used to infer vaccine effectiveness. All participants will be followed up for 12 months after receipt of the second injection.

Safety will be monitored throughout the study (Section 7.5).

Given that the preliminary data from Phase 1 to 3 studies have shown no significant safety concerns and robust immunogenicity, mRNA-1273 may be used to address the current COVID-19 outbreak as a result of its uniquely rapid and scalable manufacturing process. In particular, a safe and effective vaccine against SARS-CoV-2 in children will help facilitate a return to school as an additional step towards normalization of daily activities.

Considering the lack of approved vaccines for COVID-19, the participants' risk of COVID-19 outside the study during a pandemic, and the nonclinical and clinical data to date, the Sponsor considers the potential benefits of participation to exceed the risks.

### 2. OBJECTIVES AND ENDPOINTS

The objectives that will be evaluated in this study and the endpoints associated with each objective are provided in Table 2.

**Table 2: Study Objectives and Endpoints**

| Objectives | Endpoints |
|---|---|
| **Primary Objectives** | **Primary Endpoints** |
| • To evaluate the safety and reactogenicity of up to 3 dose levels (25, 50, and 100 µg) of mRNA-1273 vaccine administered as 2 doses 28 days apart in 3 age groups | • Solicited local and systemic ARs through 7 days after each injection<br>• Unsolicited AEs through 28 days after each injection<br>• MAAEs through the entire study period<br>• SAEs through the entire study period<br>• AESIs, including MIS-C and myocarditis and/or pericarditis, through the entire study period |
| • To infer the efficacy of mRNA-1273 (25, 50, and 100 µg, administered as 2 doses 28 days apart) based on immunogenicity in 3 age groups | • The proportion of participants with a serum antibody level at Day 57 ≥ antibody threshold of protection<br>– If an accepted serum antibody threshold of vaccine protection against COVID-19 is available, this analysis will form the basis to infer efficacy<br>• The GM value of serum antibody level and seroresponse rate from Study P204 vaccine recipients at Day 57 compared with those from young adult (18 to 25 years of age) vaccine recipients (Day 57) in the clinical endpoint efficacy trial (Study P301)<br>– If a threshold is not available, efficacy will be inferred by establishing noninferiority for each age group (6 to < 12 years, 2 to < 6 years, and 6 months to < 2 years in Study P204) compared to 18- to 25-year old participants (Study P301) by both GM value of serum antibody levels and seroresponse rate. A definition of seroresponse will be provided in the |

| | |
|---|---|
| | statistical analysis plan based on forthcoming information about assay performance |
| **Secondary Objectives** | **Secondary Endpoints** |
| • To evaluate the persistence of the immune response to mRNA-1273 vaccine (25, 50, and 100 µg) administered as 2 doses 28 days apart | • The GM values of SARS-CoV-2 S protein-specific bAb on Day 1, Day 57 (1 month after Dose 2), Day 209 (6 months after Dose 2), and Day 394 (1 year after Dose 2)<br>• The GM values of SARS-CoV-2-specific nAb on Day 1, Day 57 (1 month after Dose 2), Day 209 (6 months after Dose 2), and Day 394 (1 year after Dose 2) |
| • To evaluate the incidence of SARS-CoV-2 infection after vaccination with mRNA-1273 or placebo | • The incidence of SARS-CoV-2 infection including symptomatic and asymptomatic infection (by serology and/or RT-PCR) postbaseline<br>• SARS-CoV-2 infection will be defined in participants with negative SARS-CoV-2 at baseline:<br>– bAb level against SARS-CoV-2 nucleocapsid protein negative at Day 1, that becomes positive (as measured by Roche Elecsys) postbaseline, OR<br>– Positive RT-PCR postbaseline |
| • To evaluate the incidence of asymptomatic SARS-CoV-2 infection after vaccination with mRNA-1273 or placebo | • The incidence of SARS-CoV-2 infection measured by RT-PCR and/or bAb levels against SARS-CoV-2 nucleocapsid protein (by Roche Elecsys) postbaseline in participants with negative SARS-CoV-2 at baseline, in the absence of any COVID-19 symptoms |
| • To evaluate the incidence of COVID-19 after vaccination with mRNA-1273 or placebo. COVID-19 is defined as clinical symptoms consistent with COVID-19 AND positive RT-PCR for SARS-CoV-2 | • The incidence of the first occurrence of COVID-19 postbaseline, where COVID-19 is defined as symptomatic disease based on CDC case definition[1] |

| Exploratory Objectives | Exploratory Endpoints |
|---|---|
| • To evaluate the genetic and/or phenotypic relationships of isolated SARS-CoV-2 strains to the vaccine sequence | • Alignment of genetic sequence of viral isolates with that of the vaccine sequence |
| • To describe the ratio or profile of specific S protein bAb relative to nAb in serum | • Relative amounts or profiles of S protein-specific bAb and specific nAb titers in serum |
| • To characterize the clinical profile and immune responses of participants with COVID-19 or with SARS-CoV-2 infection | • Description of clinical severity and immune responses of participants who are identified as infected by SARS-CoV-2 (COVID-19) |
| • To assess, in a subset of participants, the SARS-CoV-2 S protein-specific T-cell responses | • Magnitude, phenotype, and percentage of cytokine-producing S protein-specific T cells, as measured by flow cytometry at different time points after vaccination relative to baseline |
| • To explore asymptomatic SARS-CoV-2 infection after vaccination with mRNA-1273 or placebo in participants with serologic evidence of infection at baseline | • GM and GMFR of bAb levels against SARS-CoV-2 nucleocapsid protein (quantitative IgG) |

Abbreviations: AE = adverse event; AESI = adverse event of special interest; AR = adverse reaction; bAb = binding antibody; COVID-19 = coronavirus disease 2019; GM = geometric mean; GMFR = geometric mean fold-rise; IgG = immunoglobin; IP = investigational product; LOD = limit of detection; MAAE = medically attended adverse event; MIS-C = multisystem inflammatory syndrome in children; nAb = neutralizing antibody; RT-PCR = reverse transcriptase polymerase chain reaction; S = spike; SAE = severe adverse event; SARS-CoV-2 = severe acute respiratory syndrome coronavirus 2.

[1] The case definition of COVID-19 includes at least one of the following systemic symptoms: fever (temperature > 38ºC/≥ 100.4ºF) or chills (of any duration, including ≤ 48 hours), cough (of any duration, including ≤ 48 hours), shortness of breath or difficulty breathing (of any duration, including ≤ 48 hours), fatigue, headache, myalgia, nasal congestion or rhinorrhea, new loss of taste or smell, sore throat, abdominal pain, diarrhea, nausea or vomiting, poor appetite or poor feeding, AND a positive test for SARS-CoV-2 by RT-PCR.

### 3. STUDY DESIGN

#### 3.1. General Design

This is a Phase 2/3, two-part, open-label, dose-escalation, age de-escalation and randomized, observer-blind, placebo-controlled expansion study intended to infer the effectiveness of mRNA-1273 in children aged 6 months to < 12 years. The study population will be divided into 3 age groups (6 to < 12 years, 2 to < 6 years, and 6 months to < 2 years), and up to 3 dose levels (25, 50, and 100 µg) of mRNA-1273 will be evaluated.

The study will be conducted in 2 parts. Part 1 of the study will be open-label and consist of dose-escalation and age de-escalation in approximately 1,275 participants to select the dose for each age group with the highest number enrolled in the oldest age group (Table 3). Part 2 of the study will be a placebo-controlled observer-blind evaluation of the selected dose in up to 12,000 participants (up to 4,000 participants each in the 6 to < 12 years, 2 to < 6 years, and the 6 months to < 2 years of age groups). No participants in Part 1 will participate in Part 2 of the study.

In order to expedite the study of the safety of mRNA-1273 in school-aged children 6 to < 12 years of age; Part 1 will enroll a total of approximately 375 participants per dose (both at the 50 and 100 µg dose levels) in this age group (6 to < 12 years). The first 75 of these participants per dose will be included in the safety evaluation for dose-escalation and age de-escalation as well as the immunogenicity assessment needed for dose selection, as applicable. Additionally, approximately 300 participants per dose will be enrolled to assess for any AE occurring at 1% or higher. The sample size for Part 2 in this age group (6 to < 12 years) will be adjusted to approximately 1700 participants. Conversely, approximately 75 participants in the middle age group (2 to < 6 years) will be enrolled per dose in Part 1 for safety and dose selection and Part 2 will utilize the dose selected in Part 1 to inform on AEs occurring at a frequency of 1% or greater. In the youngest age group (6 months to < 2 years), approximately 150 participants will be enrolled per dose in Part 1 for safety and dose selection and Part 2 will utilize the dose selected in Part 1 to inform on AEs occurring at a frequency of 1% or greater. For details, please refer to Section 7.5.

The study will begin with the oldest age group (6 to < 12 years) and age de-escalate as described under Study Progression. Each age group will begin with Part 1 and advance to Part 2 independently. For details, please refer to the study schematic in Figure 1.

The mRNA-1273 investigational vaccine or placebo will be administered as 2 intramuscular (IM) injections approximately 28 days apart.

The mRNA-1273 dose levels that will be evaluated in each age group in Part 1 and Part 2 of the study are given in Table 3.

##### Table 3: Planned Age Groups and mRNA-1273 Dose Levels in Part 1 and Part 2 of the Study

| | Part 1 | | | Part 2 | |
|---|---|---|---|---|---|
| **Age Group** | **mRNA-1273 25 µg** | **mRNA-1273 50 µg** | **mRNA-1273 100 µg** | **Selected Dose Level of mRNA-1273 From Part 1** | **Placebo** |
| 6 to < 12 years | | Study Arm 1 (n=375) | Study Arm 2 (n=375) | Study Arm 8 (n=3,000) | Study Arm 9 (n=1,000) |
| 2 to < 6 years | Study Arm 7 (Optional) (n=75) | Study Arm 3 (n=75) | Study Arm 4 (n=75) | Study Arm 10 (n= up to 3,000) | Study Arm 11 (n= up to 1,000) |
| 6 months to < 2 years | Study Arm 5 (n=150) | Study Arm 6 (n=150) | | Study Arm 12 (n= up to 3,000) | Study Arm 13 (n= up to 1,000) |

The schematic of Study Arms and major study events is provided in Figure 1.

##### Figure 1: Study Schema

###### (a) Part 1: Dose Escalation, Age De-escalation

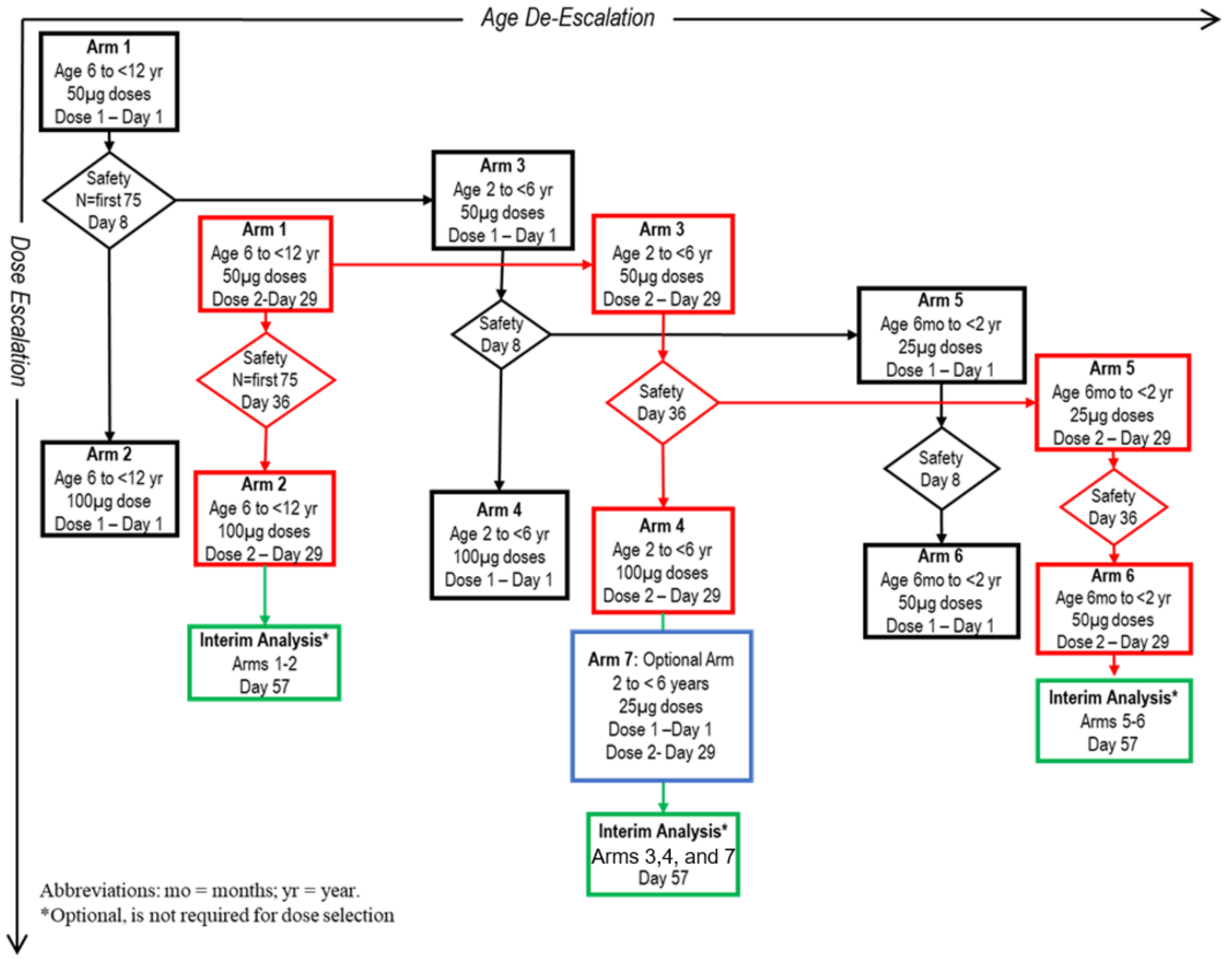

**(b) Part 2: Expansion**

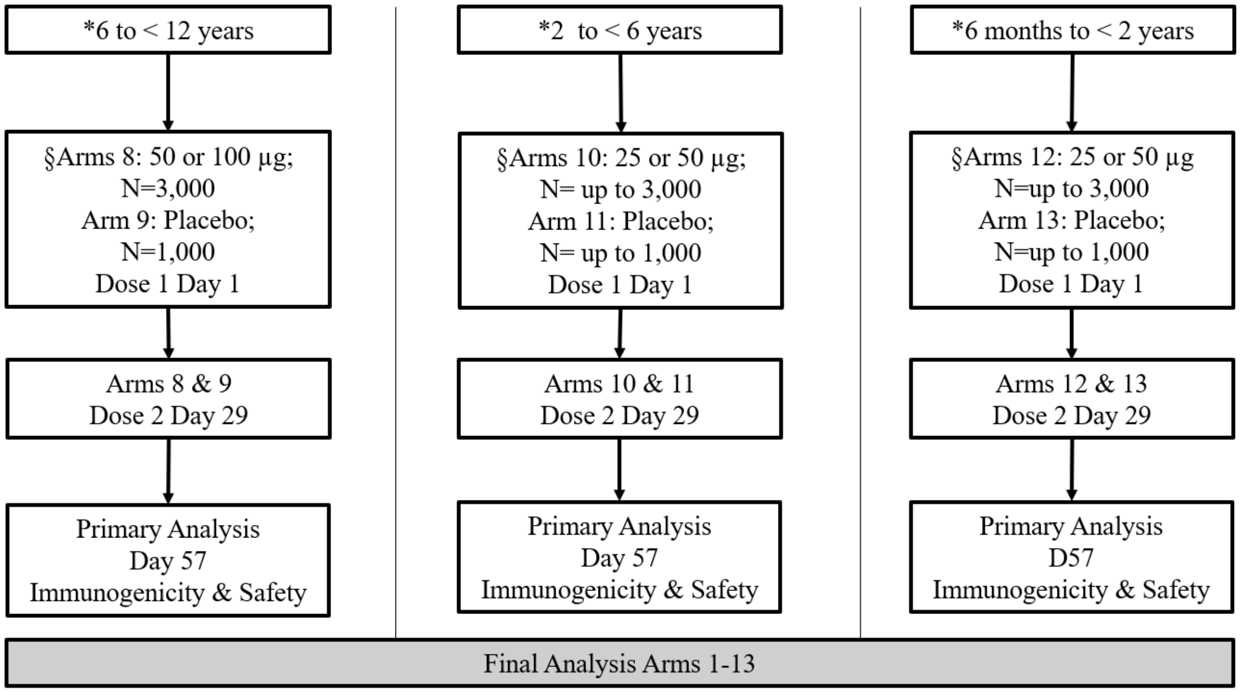

Abbreviations: CMI = cell-mediated immunity; D = Day; S = spike; VTEU = Vaccine and Treatment Evaluation Units.
*Expansion and primary analysis for each age group may occur at different times.
§Participants in each age group will be assigned to one of 5 phlebotomy cohorts. Participants in 3 of these 5 cohorts will provide blood specimens for immunogenicity on D1 (prior to randomization and first dose), D57, and one of D29 (prior to the second dose), D209, or D394 (such that each participant provides a total of 3 blood samples only). Participants in the Cohort D (remainder of the age group) will provide a blood sample on D1 (prior to randomization and before the first dose) and within 4 days of receiving Dose 2 at D30 (+3 days) for storage and potential future biomarker testing. A fifth cohort of participants (selected VTEU sites only) will provide blood samples for assessment of exploratory serology and CMI (S protein-specific T-cell responses) on D1 (prior to randomization and before the first dose), D43, D209, and D394.

#### Study Progression

Part 1 of the study will be open-label. The study will begin with enrollment of approximately 375 participants in the 6 to < 12 years age group (Study Arm 1) and dosing with 50 µg of mRNA-1273. After at least 75 participants in Study Arm 1 have completed Day 8 (1 week after Dose 1 of mRNA-1273 50 µg), an internal safety team (IST) will review the available safety data and provide a recommendation whether to escalate the dose to 100 µg in the 6 to < 12 years age group (Study Arm 2; n = 375) and independently whether to begin dosing at the 50-µg dose level in the 2 to < 6 years age group (Study Arm 3; n = 75). After at least 75 participants in Study Arm 1 reach Day 36 (1 week after Dose 2 of mRNA-1273 50 µg), the IST will again review the available safety data and provide a recommendation whether to administer Dose 2 of mRNA-1273 100 µg in Study Arm 2 and whether to administer Dose 2 of mRNA-1273 50 µg in Study Arm 3. Simultaneously, the enrollment of the remaining 300 planned participants for both Arm 1 and Arm 2 will be ongoing. A preliminary safety and immunogenicity data review of Arm 1, and Arm

2 as applicable, will aid in the selection of a dose level for Part 2. Once all or a subset of participants from each of Study Arms 1 and 2 reach Day 57 (1 month after Dose 2 of mRNA-1273), an interim analysis may be conducted to review the safety and immunogenicity data. Cumulative safety data for approximately 300 participants at the selected dose level will be reviewed by the Data Safety Monitoring Board (DSMB) before enrollment in Part 2, and the DSMB safety review recommendation will enable the expansion of the 6 to < 12 years age group (Part 2) to receive either mRNA-1273 at the selected dose level (Study Arm 8; n = 3,000) or placebo (Study Arm 9; n = 1,000). The primary analysis of immunogenicity in Part 2 for Study Arm 8 (mRNA-1273 recipients) will be conducted after the pre-specified Immunogenicity Subset of participants reaches Day 57, and safety analysis will be conducted after a subset or all participants in Arms 8 and 9 reach Day 57.

For the middle age group (2 to < 6 years), progression in Part 1 will be as follows: After 75 participants in Study Arm 3 have completed Day 8 (1 week after Dose 1 of mRNA-1273 50 µg), an IST will review the available safety data and provide a recommendation whether to escalate the dose to 100 µg in the 2 to < 6 years age group (Study Arm 4; n = 75) and whether to begin dosing in the 6 months to < 2 years age group at the 25-µg dose level (Study Arm 5; n = 150). After the 75 participants in Study Arm 3 reach Day 36 (1 week after Dose 2 of mRNA-1273 50 µg), the IST will review the available safety data and provide a recommendation whether to administer Dose 2 of mRNA-1273 100 µg in Study Arm 4 and whether to administer Dose 2 of mRNA-1273 25 µg in Study Arm 5. An optional Arm 7 may be enrolled in this age group (approximately 75 participants) at the 25-µg dose if the 100-µg dose is eliminated at any point during dose escalation process, to maintain dose ranging for this age group. A preliminary safety and immunogenicity data review of all applicable Arms will aid in the selection of a dose level for Part 2. Once all or a subset of participants in all applicable Arms reach Day 57 (1 month after Dose 2 of mRNA-1273), an interim analysis may be conducted to review safety and immunogenicity data. A preliminary safety and immunogenicity data review of all applicable Arms will aid in the selection of a dose level for Part 2 and for expansion of the 2 to < 6 years age group (Part 2) to receive either mRNA-1273 at the selected dose level (Study Arm 10; n = up to 3,000) or placebo (Study Arm 11; n = up to 1,000). The primary analysis of immunogenicity in Part 2 for Study Arm 10 (mRNA-1273 recipients) will be conducted after a pre-specified Immunogenicity Subset of participants reaches Day 57, and safety analysis will be conducted after a subset or all participants in Arms 10 and 11 reach Day 57.

For the youngest age group (6 months to < 2 years) progression in Part 1 will be as follows: In Study Arm 5, after 150 participants have completed Day 8 (1 week after Dose 1 of mRNA-1273 25 µg), an IST will review the available safety data and provide a recommendation whether to escalate the dose to 50 µg (Study Arm 6; n = 150). After the 150 participants in Study Arm 5 reach

Day 36 (1 week after Dose 2 of mRNA-1273 25 µg), the IST will review the available safety data and provide a recommendation whether to administer Dose 2 of mRNA-1273 50 µg in Study Arm 6. Once all or a subset of participants in Study Arms 5 and 6 reach Day 57 (1 month after Dose 2 of mRNA-1273), an interim analysis may be conducted to review the tolerability and immunogenicity data at each dose level. A preliminary safety and immunogenicity data review of Arm 5 and Arm 6, as applicable, will aid in the selection of a dose level for Part 2 and allow for expansion of the 6 months to < 2 years age group (Part 2) to receive either mRNA-1273 at the selected dose level (Study Arm 12; n = up to 3,000) or placebo (Study Arm 13; n = up to 1,000). The primary analysis of immunogenicity in Part 2 for Study Arm 12 (mRNA-1273 recipients) will be conducted after the pre-specified Immunogenicity Subset of participants reaches Day 57, and safety analysis will be conducted after a subset or all participants in Arm 12 and 13 reach Day 57.

In general, if a decision is made not to proceed with administration of the higher dose and/or the second injection at a given dose level (eg, due to safety concerns), the participants scheduled to receive the higher dose may receive a lower dose, and the participants who had received the higher dose level as their first injection will likely be given the next lower dose level that was tolerated for their second injection. The dose level not tolerated in the older age group will not be administered in any of the younger age groups. Within an age group, if one of the dose levels is found to have an unacceptable tolerability profile, the age group may progress to Part 2 at a lower dose level.

The final analysis will be performed when participants (Study Arms 1 to 13) conclude the safety follow-up at 12 months.

The goal of the study is to support an indication for use of mRNA-1273 50 or 100 µg IM, given as 2 injections, approximately 28 days apart in the 6 to < 12 years age group; mRNA-1273 25, 50, or 100 µg IM, given as 2 injections, approximately 28 days apart in the 2 to < 6 years age group; and mRNA-1273 25 or 50 µg IM, given as 2 injections, approximately 28 days apart in the 6 months to < 2 years age group. The basis for demonstrating vaccine effectiveness is proposed to be met by measuring serum antibody responses in the study participants. The approach to inferring vaccine effectiveness will depend on whether an accepted serum antibody threshold conferring protection against COVID-19 has been established. If an antibody threshold of protection has been established, effectiveness will be inferred based on the proportion of study participants with serum antibody levels (on Day 57) that meet or exceed the antibody threshold. If an antibody threshold of protection has not been established, effectiveness will be inferred by demonstrating noninferiority for each age group (6 to < 12 years, 2 < 6 years, and 6 months to < 2 years in Study P204) compared to young adult (18 to 25 years of age) participants enrolled in the ongoing clinical endpoint efficacy trial (Study P301) by both geometric mean (GM) values and seroresponse rate. The statistical parameters to infer effectiveness are described in Section 2.

This study in children 6 months to < 12 years of age will monitor all participants for a total of 12 months following the second dose of vaccine or placebo. If a COVID-19 vaccine is authorized or licensed for a specific age group before the end of the study, please refer to Section 3.3 for unblinding and/or cross-over plans. Safety assessments will include solicited ARs 7 days after each injection (ie, the day of injection and 6 subsequent days), unsolicited AEs (28 days after each injection), and medically attended adverse events (MAAEs), SAEs, and adverse events of special interest (AESIs) (including MIS-C and myocarditis and/or pericarditis) through the entire study period.

Blood samples will be collected from participants in Part 1 and Part 2 of the study for assessment of immunogenicity as specified in Section 3.1.1.2. If a Day 1 (baseline) blood sample cannot be obtained in either Part 1 or Part 2, the participant will be considered either a screen failure due to inability to satisfy inclusion criterion 3 or could be rescheduled for rescreening. Participants may be rescreened 1 time, either within the same screening period for Part 1 or Part 2 or in a new screening period of Part 2 if the initial screening was in Part 1. If the participant is rescreened and a blood sample still cannot be obtained, then he/she will be considered a screen failure due to inability to satisfy inclusion criterion 3. Blood samples will also be tested for the development of antibodies directed against nonvaccine antigen (eg, antibodies against the nucleocapsid protein), which will signify infection with SARS-CoV-2. The incidence of SARS-CoV-2 infection among vaccine recipients and placebo recipients will be compared to assess the potential for mRNA-1273 to reduce the rate of infection in vaccine recipients. In addition, all participants will be monitored for symptoms of COVID-19 and scheduled for illness visits if concerning symptoms occur, and a nasal swab will be collected at the illness visit.

##### 3.1.1. Study Periods

This study involves up to 8 scheduled visits including up to 6 in-person visits (if screening and baseline are done separately) and 2 telemedicine visits (Visit 2 and Visit 4; remote visit by means of telecommunication technology). An additional in clinic visit (Visit 4S) will be conducted for a cohort of participants from selected Vaccine and Treatment Evaluation Units (VTEU) sites on Day 43 (2 weeks after Dose 2 of mRNA-1273 or placebo) for exploratory serology and cell-mediated immunity (CMI; S protein-specific T-cell responses) in Part 2 of the study.

The study duration for each participant will be approximately 14 months, which includes 1 month for screening (Day -28 to Day -1), 1 month for dosing (on Day 1 and Day 29), and 12 months of follow-up after the second dose to monitor for safety, tolerability, reactogenicity, immunogenicity, and efficacy.

All scheduled study visits should be completed within the respective visit windows. If the participant is not able to attend a study site visit as a result of the COVID-19 pandemic (self-quarantine or disruption of study site activities following business continuity plans and/or local government mandates for “stay at home” or “shelter in place”), a telemedicine visit should be conducted in place of the study site visit. The telemedicine visit should encompass all scheduled visit assessments that can be completed remotely, such as assessment for AEs and concomitant medications (eg, as defined in scheduled safety telephone calls). Home visits will be permitted for all nondosing visits, with the exception of screening, if a participant cannot visit the study site as a result of the COVID-19 pandemic. Home visits must be permitted by the study site institutional review board (IRB) and the participant’s parent(s)/legally acceptable representatives (LAR[s]) via informed consent and have prior approval from the Sponsor (or its designee).

This study will be conducted in compliance with the protocol, Good Clinical Practice (GCP), and all applicable regulatory requirements (Section 10.2.1).

###### 3.1.1.1. Screening Period

After providing informed consent/assent, participants will undergo screening assessments to determine study eligibility. Screening assessments (Table 10) must be completed after the participant’s parent(s)/LAR(s) signs the informed consent form (ICF) and the participant, where applicable, signs the assent form. The investigator will review study entry criteria to determine participant eligibility during the Screening Period.

Screening Visit and Day 1 may be combined on the same day. Additionally, the Screening Visit may be performed over multiple visits if performed within the 28-day screening window (Table 10).

Eligible participants will enter the Treatment Period.

###### 3.1.1.2. Treatment and Follow-up Period

On Day 1, after the completion of the scheduled assessments (Table 10), participants will be administered a single IM dose of the investigational product (IP) mRNA-1273 (25, 50, or 100 µg) or placebo. Placebo will be administered only to those participating in Part 2 of the study. The procedures for IP administration will be detailed in the mRNA-1273-P204 Pharmacy Manual. Participants will be closely monitored for safety and will remain at the study site for observation for at least 30 minutes after dosing. On Day 29, the second dose of IP will be administered. If the visit for the second dose (Day 29) is disrupted and cannot be completed at Day 29 (+7 days) as a result of the COVID-19 pandemic (self-quarantine or disruption of study site activities following business continuity plans and/or local government mandates for “stay at home” or “shelter in place”), the visit window may be extended to Day 29 + 21 days. When the extended visit window is used, the remaining study visits should be rescheduled to follow the intervisit interval from the actual date of the second dose. Participants will be monitored for 12 months after the second dose of IP for safety, immunogenicity assessments, and biomarker samples.

To test for the presence of SARS-CoV-2 by RT--PCR, nasal swab samples will be collected on each day of injection prior to dosing and on Day 43 (if visit is applicable), Day 57 (1 month after Dose 2), Day 209 (6 months after Dose 2), and Day 394 (12 months after Dose 2) according to the schedule of assessments (SoA; Table 10). Participants who test positive at baseline may receive Dose 2 as long as they remain asymptomatic at the time of dosing.

During the course of the study, participants who meet pre-specified disease criteria that suggest possible SARS-CoV-2 infection will be asked to contact the study site to arrange for a prompt, thorough, and careful assessment, including a nasal swab sample to be tested for the presence of SARS-CoV-2 by RT-PCR. Confirmed, symptomatic cases of SARS-CoV-2 infection will be captured as MAAEs and reported in an expedited time frame to the Sponsor (Section 7.4.4).

All participants will be monitored for safety and reactogenicity. All participants in Part 1 of the study will provide blood specimens for immunogenicity on Day 1 (prior to the first dose) and will also provide additional samples after the second dose of mRNA-1273 at Day 57, Day 209, and Day 394, except for the expansion of Arms 1 and 2 (N= ~300 participants per arm) who have a scheduled Day 1 blood draw and a voluntary blood draw on Day 57 only. Participants in each age group in Part 2 will be assigned to 1 of 5 phlebotomy cohorts. Participants in 3 of these 5 cohorts will provide blood specimens for immunogenicity at the following time points: Day 1 (prior to randomization and before the first dose), Day 57, and one of Day 29 (prior to the second dose), Day 209, or Day 394 (such that each participant provides a total of 3 blood samples only). Participants in the Cohort D (remainder of the age group) will provide a blood sample at Day 1 (prior to randomization and before the first dose) and within 4 days of receiving Dose 2 at Day 30 (+3 days) for storage and potential future biomarker testing. For participants already enrolled in Cohort D prior to protocol amendment 4, this blood draw will be optional and confirmed at time of re-consenting; for all participants newly enrolled into Cohort D under amendment 4, it will be mandatory.

A fifth cohort of participants (selected VTEU sites only) will provide blood samples for assessment of exploratory serology and CMI (S protein-specific T-cell responses) on Day 1 (prior to randomization and before the first dose), Day 43, Day 209, and Day 394. Table 12 provides the blood sampling schedule for Part 2 of the study.

Participants and their parent(s)/LAR(s) will be instructed on the day of the first dose (Day 1) and reminded on the day of the second dose (Day 29) how to document and report solicited local or systemic ARs in a provided electronic diary (eDiary). Solicited ARs, unsolicited AEs, MAAEs,

AEs leading to withdrawal, AESIs, and SAEs will be assessed as described in Section 7.1, according to the time points in the SoA (Table 10).

An unscheduled visit may be prompted by reactogenicity issues, illness visit criteria for COVID-19, or new or ongoing AEs. If a participant meets the pre-specified criteria of suspicion for COVID-19, the participant will be asked to return within 72 hours or as soon as possible to the study site for an unscheduled illness visit, to include a nasal swab sample (for RT-PCR testing to evaluate for the presence of SARS-CoV-2 infection) and other clinical evaluations. If a study site visit is not possible, a home visit may be arranged to collect the nasal swab sample and conduct clinical evaluations. The study site may collect an additional nasal swab sample for SARS-CoV-2 testing to be able to render appropriate medical care for the study participant as determined by local standards of care. Additionally, clinical information will be carefully collected to evaluate the severity of the clinical case.

A convalescent visit will be scheduled approximately 28 days (+7 days) after diagnosis of COVID-19. At this visit, a nasal swab sample for viral RT-PCR and a blood sample for potential immunologic assessment of SARS-CoV-2 infection will be collected.

For treatment and follow-up assessments of placebo participants in the case that a COVID-19 vaccine is authorized or licensed for a specific age group during conduct of the trial, please refer to Section 3.3.

#### 3.2. Scientific Rationale for Study Design

This Phase 2/3 study in children 6 months to < 12 years of age is planned to understand the tolerability and immunogenicity of mRNA-1273 in a pediatric population. This study follows a pivotal Phase 3 study (Study P301) in 30,000 adults 18 years and older to demonstrate the tolerability, safety, and high efficacy of mRNA-1273 (100 µg on Days 1 and 29) against COVID-19. This pediatric study is intended to confirm safety in children between 6 months and 12 years of age and bridge immunogenicity between children and young adults (18 to 25 years of age) enrolled in the pivotal adult Phase 3 study (Study P301). It is necessary to demonstrate noninferiority of the induced immune response in children compared with that in adults to infer vaccine effectiveness in this age group.

Part 1 of the study is designed to dose escalate and age de-escalate through 3 age groups (6 to < 12 years, 2 to < 6 years, and 6 months to < 2 years). Each age group will begin dosing with the lowest dose planned for that group. Dose-escalation and age de-escalation will progress only after confirming the safety of a dose level in each age group after each IP injection. The placebo-controlled dose-expansion Part 2 of the study will begin only after an interim analysis is performed for safety and immunogenicity in each of the age groups at the selected dose level.

With SARS-CoV-2 expected to be circulating in the general population during the study, a pre-defined subset of participants will provide blood samples for antibody analysis starting on Day 29 and continuing through 12 months after the last dose of IP. In addition, participants will have nasal swab samples collected, before the injections on Day 1, Day 29, Day 43 (if applicable), Day 57, Day 209, and Day 394. Furthermore, an additional nasal swab sample and a blood sample will be taken to confirm the diagnosis of SARS-CoV-2 via RT-PCR and serology, respectively, if there are any signs or symptoms or an MAAE suggesting SARS-CoV-2 infection in a participant. Additionally, clinical information will be carefully collected to evaluate the severity of the clinical case.

As it is possible that participants are naturally exposed to SARS-CoV-2 through community exposure, the nasal swab samples collected before study injection and the serologic assays for antibody responses to nonvaccine antigen(s) may help discriminate between natural infection and vaccine-induced antibody responses, should such discrimination be needed.

#### 3.3. Justification for Dose, Control Product, and Choice of Study Population

Based on the Phase 3 data and the results of the Phase 1 and 2 studies described in Section 1.2.2, the Sponsor intends to study 3 dose levels (25, 50, and 100 µg) in the Phase 2/3 study in children 6 months to < 12 years of age. On 18 Dec 2020, the mRNA-1273 vaccine (100 µg dose) was authorized by the US FDA for emergency use in individuals ≥ 18 years of age.

As there are currently no licensed SARS-CoV-2 vaccines available for children 6 months to 12 years of age, 0.9% sodium chloride will be used as a placebo control for the safety and immunogenicity assessments in Part 2. Consequently, the mRNA-1273 vaccine and placebo injections will look different, so administration will be blinded (Section 8.1).

If a COVID-19 vaccine (mRNA-1273 or other) is authorized or licensed, eligible study participants (by virtue of their age) will be offered the opportunity to unblind via a phone call and learn what treatment they received, ideally after they have reached at least study Day 57. Participants who learn that they have received mRNA-1273 will continue in the study and will be followed per protocol. If a participant previously received placebo the participant will be offered cross-over vaccination with mRNA-1273 (first injection), followed by a second mRNA-1273 injection 28 days thereafter. The remainder of study visits will subsequently be performed as shown in the SoA for cross-over vaccination with mRNA-1273 if any COVID-19 vaccine is authorized or licensed for participant's age group (Table 11). All participants will continue to be followed until their Day 394 end-of-study visit. In addition, participants who received a lower dose in Part 1 than was ultimately approved for their respective age group in Part 2, will be eligible for a booster dose with the optimal dose once authorization or approval of mRNA-1273 has been granted for their age group. If a decision is made to go outside the protocol to receive an EUA vaccine, the participant will be discontinued from the study.

#### 3.4. End-of-Study Definition

The end of the study for the full study is defined as completion of the last visit of the last participant in the study or the last scheduled procedure as shown in the SoA (Table 10) for the last participant in this study.

### 4. STUDY POPULATION

Participants will be enrolled at approximately 75 to 100 study sites in the United States and Canada.

Prospective approval of protocol deviations to recruitment and enrollment criteria, also known as protocol waivers or exemptions, is not permitted.

#### 4.1. Inclusion Criteria

Participants are eligible to be included in the study only if all the following criteria apply:

1. The participant is male or female, 6 months to < 12 years of age at the time of consent/assent (Screening Visit), who is in good general health, in the opinion of the investigator, based on review of medical history and screening physical examination.
2. If the participant has a chronic disease (eg, asthma, diabetes mellitus, cystic fibrosis, human immunodeficiency virus [HIV] infection), the disease should be stable, per investigator assessment, so that the participant can be considered eligible for inclusion. Stable diseases are those which have had no change in their status or in the medications required to control them in the 6 months prior to Screening Visit.

   Note: a change in medication for dose optimization (eg, insulin dose changes), change within class of medication, or reduction in dose are not considered signs of instability.
3. In the investigator's opinion, the parent(s)/LAR(s) understand and are willing and physically able to comply with protocol-mandated follow-up, including all procedures, and provide written informed consent and participants are willing to provide assent.
4. The participant is 2 years or older and has a body mass index (BMI) at or above the third percentile according to WHO Child Growth Standards at the Screening Visit (Section 10.2.18).

   The participant is less than 2 years of age and the participant's height and weight are both at or above the 3rd percentile according to WHO Child Growth Standard at the Screening Visit (Section 10.2.18).
5. Female participants of nonchildbearing potential may be enrolled in the study. Nonchildbearing potential is defined as premenarche.

**Special inclusion criteria for female participants who have reached menarche:**

6. Female participants of childbearing potential may be enrolled in the study if the participant fulfills all of the following criteria:

- Has a negative pregnancy test at Screening. Pregnancy test will be performed if deemed appropriate by the investigator.
- Has practiced adequate contraception or has abstained from all activities that could result in pregnancy for at least 28 days prior to the first injection (Day 1).
- Has agreed to continue adequate contraception or abstinence through 3 months following the second injection (Day 29).
- Is not currently breastfeeding.

- Adequate female contraception is defined as abstinence or consistent and correct use of a US FDA-approved contraceptive method in accordance with the product label (Section 10.3).

**Special inclusion criteria for children 6 months to < 12 months of age**

7. The participant was born at full-term (≥ 37 weeks gestation) with a minimum birth weight of 2.5 kg.

#### 4.2. Exclusion Criteria

Participants will be excluded from the study if any of the following criteria apply:

1. Has a known history of SARS-CoV-2 infection within 2 weeks prior to administration of IP or known close contact with anyone with laboratory-confirmed SARS-CoV-2 infection or COVID-19 within 2 weeks prior to administration of IP.
2. Is acutely ill or febrile 24 hours prior to or at the Screening Visit. Fever is defined as a body temperature ≥ 38.0°C/≥ 100.4°F. Participants who meet this criterion may have visits rescheduled within the relevant study visit windows. Afebrile participants with minor illnesses can be enrolled at the discretion of the investigator.
3. Has previously been administered an investigational or approved CoV (eg, SARS-CoV-2, SARS-CoV, MERS-CoV) vaccine.
4. Has undergone treatment with investigational or approved agents for prophylaxis against COVID-19 (eg, receipt of SARS-CoV-2 monoclonal antibodies) within 6 months prior to enrollment.
5. Has a known hypersensitivity to a component of the vaccine or its excipients. Hypersensitivity includes, but is not limited to, anaphylaxis or immediate allergic reaction of any severity to a previous dose of an mRNA COVID-19 vaccine or any of its components (including polyethylene glycol [PEG] or immediate allergic reaction of any severity to polysorbate).

6. Has a medical or psychiatric condition that, according to the investigator's judgment, may pose additional risk as a result of participation, interfere with safety assessments, or interfere with interpretation of results.
7. Has a history of diagnosis or condition that, in the judgment of the investigator, may affect study endpoint assessment or compromise participant safety, specifically the following:
   - Congenital or acquired immunodeficiency, excluding HIV infection, as described in Inclusion Criteria 2
   - Chronic hepatitis or suspected active hepatitis
   - A bleeding disorder that is considered a contraindication to IM injection or phlebotomy
   - Dermatologic conditions that could affect local solicited AR assessments
   - Any prior diagnosis of malignancy (excluding nonmelanoma skin cancer)
   - Febrile seizures*

     *In Part 2 of the study, a history of a simple, single febrile seizure is allowed for children 6 years and older.
8. Has received the following:
   - Any routine vaccination with inactivated or live vaccine(s) within 14 days prior to first vaccination or plans to receive such a vaccine through 14 days following the last study vaccination.
     - Note: This excludes influenza vaccine that may be given, however, not within 14 days prior to or post Dose 1 or Dose 2. If a participant receives an influenza vaccine, this should be captured within the concomitant medication electronic case report form (eCRF) (Section 5.5.2).
   - Systemic immunosuppressants or immune-modifying drugs for > 14 days in total within 6 months prior to the day of enrollment (for corticosteroids, ≥ 1 mg/kg/day or ≥ 10 mg/day prednisone equivalent, if participant weighs > 10 kg). Participants may have visits rescheduled for enrollment if they no longer meet this criterion within the Screening Visit window. Inhaled, nasal, and topical steroids are allowed.
   - Intravenous or subcutaneous blood products (red cells, platelets, immunoglobulins) within 3 months prior to enrollment.

9. Has participated in an interventional clinical study within 28 days prior to the Screening Visit or plans to do so while participating in this study.
10. Is an immediate family member, or household contact, of an employee of the study site or Moderna or someone otherwise directly involved with the conduct of the study. As applicable, family members/household contacts of employees of the larger institution or affiliated private practice not part of the study site may be enrolled.

#### 4.3. Lifestyle Restrictions

Participants must not eat or drink anything hot or cold within 10 minutes before their temperature is taken.

#### 4.4. Screen Failures

Screen failures are defined as participants whose parent(s)/LAR(s) provide consent and, where applicable, participants provide the assent to participate in the clinical study but are not subsequently assigned (Part 1) or randomly assigned (Part 2) to treatment. A minimum set of screen failure information is required to ensure transparent reporting of screen failures to meet the Consolidated Standards of Reporting Trials publishing requirements and to respond to queries from regulatory authorities. Minimum information includes date of informed consent/assent, demography, reason(s) for screen failure, eligibility criteria, and information on any SAE that may have occurred from Day 1 at or after dosing to the time of withdrawal.

### 5. STUDY TREATMENT

#### 5.1. Investigational Product Administered

The term IP refers to mRNA-1273 (25, 50, and 100 µg) vaccine or placebo (0.9% sodium chloride) in this study.

mRNA-1273 is an LNP dispersion of an mRNA that encodes the prefusion stabilized S protein of SARS-CoV-2 formulated in LNPs composed of 4 lipids (1 proprietary and 3 commercially available): the proprietary ionizable lipid SM-102; cholesterol; 1,2-distearoyl-sn-glycero-3-phosphocholine (DSPC); and 1-monomethoxypolyethyleneglycol-2,3-dimyristylglycerol with PEG of average molecular weight 2000 (1-monomethoxypolyethyleneglycol-2,3-dimyristylglycerol polyethylene glycol 2000 [PEG2000-DMG]). mRNA-1273 injection is provided as a sterile liquid for injection and is a white to off-white dispersion at a concentration of 0.2 mg/mL in 20 mM Tris buffer containing 87 mg/mL sucrose and 4.3 mM sodium acetate at pH 7.5.

#### 5.2. Randomization

Random assignment of participants in Part 2 of the study will use a centralized interactive response technology, in accordance with pregenerated randomization schedules. Up to 4,000 participants each in the 6 to < 12 years, 2 to < 6 years, and 6 months to < 2 years age groups will be randomized in a 3:1 ratio to the mRNA-1273 arm (n = up to 3,000 participants in each group) or placebo arm (n = up to 1,000 participants in each group).

#### 5.3. Dosing and Management of mRNA-1273 Vaccine

##### 5.3.1. Preparation of Study Vaccine for Injection

Each dose of IP will be prepared for each participant based on the assigned treatment, as detailed in the mRNA-1273-P204 Pharmacy Manual. The volume of IP injected will be 0.5 mL consisting of a 25-µg dose of mRNA-1273, a 50-µg dose of mRNA-1273, a 100-µg dose of mRNA-1273, or placebo (normal saline), as detailed in the mRNA-1273-P204 Pharmacy Manual.

##### 5.3.2. Administration of Study Vaccine

Each participant will receive 2 doses of IP by IM injection approximately 28 days apart (Day 1 and Day 29) into the deltoid muscle or anterolateral thigh (per investigator's discretion), according to their assigned regimen and according to the procedures specified in the mRNA-1273-P204 Pharmacy Manual.

At each visit when IP is administered, participants will be monitored for a minimum of 30 minutes after administration. Assessments will include body temperature measurements (oral preferred for participants > 4 years of age, tympanic preferred for participants ≤ 4 years of age, but other methods acceptable in context of COVID-19 precautions) and monitoring for local or systemic reactions (Table 10).

Eligibility for the subsequent dose of IP will be determined by following the criteria outlined in Section 6.

The study sites will be appropriately staffed with individuals with basic cardiopulmonary resuscitation training/certification. Either on-site resuscitation equipment and personnel or appropriate protocols for the rapid transport of participants to a resuscitation area or facility are required.

##### 5.3.3. Study Vaccine Delivery and Receipt

The Sponsor or designee is responsible for the following:

- Supplying the IP
- Confirming the appropriate labeling of the IP, so that it complies with the legal requirements of the United States and Canada.

The investigator is responsible for acknowledging the receipt of the IP by a designated staff member at the study site, including the following:

- Confirming that the IP was received in good condition
- Confirming that the temperature during shipment from the Sponsor to the investigator's designated storage location was appropriate
- Confirming that the Sponsor has authorized the IP for use
- Ensuring the appropriate dose level of IP is properly prepared using aseptic technique

Further description of the IP and instructions for the receipt, storage, preparation, administration, accountability, and destruction of the IP are described in the mRNA-1273-P204 Pharmacy Manual.

##### 5.3.4. Study Vaccine Packaging and Labeling

The Sponsor will provide the investigator (via the study site pharmacy) with adequate quantities of IP. The sterile IP is packaged in 10R glass vials with a 6.3-mL fill volume. The IP will have all required labeling per regulations and will be supplied to the pharmacy in an unblinded manner.

The IP will be packaged and labeled in accordance with the standard operating procedures of the Sponsor or its designee, Code of Federal Regulations (CFR) Title 21, Good Manufacturing Practice guidelines, International Council for Harmonisation (ICH) GCP guidelines, guidelines for Quality System Regulations, and applicable regulations.

##### 5.3.5. Study Vaccine Storage

The IP must be stored at -15°C to -25°C in a secure area with limited access and be protected from moisture and light until it is prepared for administration (Section 5.3.1). The freezer should have automated temperature recording and a 24-hour alert system in place that allows for rapid response in case of freezer malfunction. There must be an available backup freezer. The freezer must be connected to a back-up generator, or an alternate plan must be in place in the event of a power failure. In addition, IP accountability study staff are required to keep a temperature log to establish a record of compliance with these storage conditions. The study site is responsible for reporting any IP that was not temperature controlled during shipment or during storage. Such IP will be retained for inspection by the monitor and disposed of according to approved methods.

##### 5.3.6. Study Vaccine Accountability

It is the investigator's responsibility that the IP accountability study staff maintain accurate records in an IP accountability log of receipt of all IP, study site IP inventory, IP dispensing, IP injections, and return to the Sponsor or alternative disposition of used and unused IP vials.

A study site monitor will review the inventory and accountability log during study site visits and at the completion of each part of the study. Additional details are found in the mRNA-1273-P204 Pharmacy Manual.

##### 5.3.7. Study Vaccine Handling and Disposal

A study site monitor will reconcile the IP inventory during the conduct and at the end of each part of the study for compliance. Once fully reconciled after each monitoring visit at the study site, and completion of each age cohort or at the end of the study, the IP can be destroyed at the investigational site or by a Sponsor-selected third party, as appropriate.

Vaccine may be destroyed at the study site only if permitted by local regulations and authorized by the Sponsor. A certificate of destruction must be completed and sent to the Sponsor or designee.

#### 5.4. Study Treatment Compliance

All doses of IP will be administered at the study site under direct observation of medically qualified study staff and appropriately recorded (date and time) in the eCRF. Qualified study site staff will confirm that the participant has received the entire dose of IP. If a participant does not receive IP or does not receive all of the planned doses, the reason for the missed dose will be recorded. Data will be reconciled with study site accountability records to assess compliance.

Participants who miss the second dose due to noncompliance with the visit schedule and not due to a safety pause will still be required to follow the original visit and testing schedule as described in the protocol and their regimen schedule. Unless consent/assent is withdrawn, a participant who withdraws or is withheld from receiving the second dose will remain in the study and complete all safety and immunogenicity assessments required through the participant's last scheduled study visit.

The study site staff are responsible for ensuring that participants comply with the allowed study visit windows. If a participant misses a visit, every effort should be made to contact the participant and complete a visit within the defined visit window (Table 10). If a participant does not complete a visit within the time window, that visit will be classified as a missed visit and the participant will continue with subsequent scheduled study visits. All safety requirements of the missed visit will be captured and included in the subsequent visit.

#### 5.5. Prior and Concomitant Medications

##### 5.5.1. Prior Medications and Therapies

Information about prior medications (including any prescription or over-the-counter medications, vaccines, or blood products) taken by the participant within the 28 days before his/her parent(s)/LAR(s) provided informed consent and the participant provided assent (or as designated in the inclusion/exclusion requirements) will be recorded in the participant's eCRF.

##### 5.5.2. Concomitant Medications and Therapies

At each study visit, study site staff must question the participant and/or the participant's parent(s)/LAR(s) regarding any medications taken and vaccinations received by the participant and record the following information in the eCRF:

- All nonstudy vaccinations administered within the period starting 14 days before the first dose of IP and through 14 days after the last dose of IP.
- Seasonal influenza vaccine administered for the current influenza season (typically October through April in the Northern Hemisphere).
- All concomitant medications taken through 28 days after each dose of IP. Antipyretics and analgesics taken prophylactically (ie, taken in the absence of any symptoms in anticipation of an injection reaction) will be recorded as such.
- Any concomitant medications used to prevent or treat COVID-19 or its symptoms.
- Any concomitant medications relevant to or for the treatment of an SAE or an MAAE.

Participants will be asked in the eDiary if they have taken any antipyretic or analgesic to treat or prevent fever or pain within 7 days after each IP dose, including on the day of dosing. Reported antipyretic or analgesic medications should be recorded in the source document by the study site staff during the postinjection period of the study visits or via other participant interactions (eg, telephone calls).

Data regarding nutritional supplements, eg, vitamins, probiotics, and herbal supplements, will not be collected.

Concomitant medications (including vaccinations) will be coded using the WHO Drug Dictionary. If a participant takes a prohibited drug therapy, the investigator and the contract research organization's (CRO's) medical monitor will make a joint decision about continuing or withholding further injection of the participant based on the time the medication was administered, the drug's pharmacology and pharmacokinetics, and whether use of the medication will compromise the participant's safety or interpretation of the data. It is the investigator's responsibility to ensure that details regarding concomitant medications are adequately recorded in the eCRF.

##### 5.5.3. Concomitant Medications and Vaccines That May Lead to the Elimination of a Participant From Per-Protocol Analyses

The use of the following concomitant medications and/or vaccines will not require withdrawal of the participant from the study but may determine a participant's eligibility to receive a second dose or be included in the per-protocol (PP) analysis (analysis sets are described in Section 8.4):

- Any investigational or nonregistered product (drug or vaccine) other than the IP used during the study period.
- Immunosuppressants or other immune-modifying drugs administered chronically (ie, > 14 days in total) during the study period. For corticosteroids, receipt of prednisone or the equivalent at a dose of ≥ 1 mg/kg/day (or ≥ 10 mg/day if participant weighs > 10 kg) is not permitted. Inhaled, nasal, and topical steroids are allowed.
- Long-acting immune-modifying drugs administered at any time during the study period (eg, infliximab).
- Immunoglobulins and/or any Ab-containing blood products administered during the study period.

#### 5.6. Intervention After the End of the Study

Any SAE, including death, occurring after the end of the study, and considered to be caused by the IP must be reported to the Sponsor.

### 6. DELAYING OR DISCONTINUING STUDY TREATMENT AND PARTICIPANT WITHDRAWAL FROM THE STUDY

#### 6.1. Criteria for Delay of Vaccine Administration

##### 6.1.1. Individual Participant Criteria for Delay of Study Vaccination

Body temperature (oral preferred for participants > 4 years of age or tympanic preferred for participants ≤ 4 years of age, but other methods are acceptable in context of COVID-19 precautions) must be measured on dosing visits before vaccine administration. The following events constitute criteria for delay of injection, and if any of these events occur at the time scheduled for dosing, the participant may receive the study injection at a later date within the time window specified in the SoA (Table 10), or the participant may be discontinued from dosing at the discretion of the investigator (Section 6.2):

- Acute moderate or severe infection with or without fever at the time of dosing
- Fever, defined as body temperature ≥ 38.0°C/≥ 100.4°F at the time of dosing

Participants with a minor illness without fever, as assessed by the investigator, can be vaccinated.

Participants with a fever of ≥ 38.0°C/≥ 100.4°F will be contacted within the time window acceptable for participation and re-evaluated for eligibility. If the investigator determines that the participant's health on the day of dosing temporarily precludes injection, the visit should be rescheduled within the allowed interval for that visit.

If a participant was exposed to a household contact (excluding school or other exposures) who tested positive for SARS-CoV-2 after the participant received the first dose, the administration of the second dose may be delayed to allow for at least 14 days between the positive test or the last day of symptoms (if present) of the household contact and Dose 2 as long as the participant remains asymptomatic.

If a participant tests positive for SARS-CoV-2 after having received Dose 1 but remains asymptomatic and the second dose is due before 14 days after the positive test, administration of the second dose may be delayed to allow for at least 14 days between the participant's positive test and Dose 2 as long as participant remains asymptomatic.

If a participant tests positive for SARS-CoV-2 after having received Dose 1 and develops symptoms of COVID-19, the participant may still receive a second dose, but the second dose should be delayed until approximately 90 days after diagnosis with COVID-19.

If a participant takes a prohibited drug therapy, an injection could be delayed within the visit window based on the joint decision of the investigator and the CRO's medical monitor (Section 5.5.2).

If unforeseen circumstances out of the control of the participant's family or the Study Site (eg, extreme weather, such as hurricanes) render it impossible for the participant to receive the second dose within the specified window, Dose 2 may be delayed at the principal investigator's discretion but should be administered as soon as possible thereafter.

#### 6.2. Discontinuing Study Vaccination

Participants can discontinue study injection (ie, refuse the second dose) for any reason, without prejudice to further treatment the participant may need to receive.

The investigator, in consultation with the Sponsor's medical monitor, may withhold a participant from further injection under the following circumstances:

- The participant becomes pregnant.
- The participant's parent(s)/LAR(s) withdraw consent or the participant withdraws assent.
- An RT-PCR result from an illness visit (Section 7.1.5) is positive for SARS-CoV-2 and the participant is symptomatic at the time of next injection.
- The participant develops, during the course of the study, symptoms or conditions listed in the exclusion criteria (Section 4.2).
- The participant experiences an AE (other than solicited reactogenicity) after injection that is considered by the investigator to be related to IP (Section 7.4.9) and is of Grade 3 (severe) or greater severity.
- The participant experiences an AE or SAE that, in the judgment of the investigator, requires IP withdrawal due to its nature, severity, or required treatment, regardless of the causal relationship to vaccine.
- The participant experiences an AESI (eg, MIS-C and myocarditis and/or pericarditis).
- The participant experiences a clinically significant change in general condition that, in the judgment of the investigator, requires vaccine withdrawal.
- The participant experiences anaphylaxis (described in Section 7.4.4) clearly related to IP.
- The participant experiences generalized urticaria related to IP.

The reason(s) for withdrawal from further injection will be recorded in the eCRF.

If a participant takes a prohibited drug therapy, the investigator could withhold the second dose based on a joint decision of the investigator and the CRO's medical monitor (Section 5.5.2).

Every reasonable attempt should be made to follow up with participants for safety throughout the entire scheduled study period according to their regimen, even if the participant does not receive the second dose or misses one or more visits. Unless the participant's parent(s)/LAR(s) withdraw consent or the participant withdraws assent, they are expected to remain in the study and complete all scheduled visits and assessments.

#### 6.3. Participant Discontinuation/Withdrawal From the Study

Participants who withdraw or are withdrawn from the study will not be replaced. A "withdrawal" from the study refers to a situation wherein a participant does not return for the final visit planned in the protocol.

The participant's parent(s)/LAR(s) can withdraw consent or the participant can withdraw assent and withdraw from the study at any time, for any reason, without prejudice to further treatment the participant may need to receive. The investigator will request that the participant complete all study procedures pending at the time of withdrawal.

If a participant desires to withdraw from the study because of an AE, the investigator will try to obtain agreement to follow up with the participant until the event is considered resolved or stable and will then complete the end of the study eCRF.

Information related to the withdrawal will be documented in the eCRF. The investigator will document whether the decision to withdraw a participant from the study was made by the participant or by the investigator, as well as which of the following possible reasons was the cause for withdrawal:

- AE (specify)
- SAE (specify)
- Death
- Lost to follow-up (LTFU)
- Physician decision (specify)
- Pregnancy
- Protocol deviation
- Study terminated by Sponsor
- Withdrawal of consent by participant's parent(s)/LAR(s) or withdrawal of assent by the participant (specify)
- Other (specify)

Participants who are withdrawn from the study because of AEs (including AESIs and SAEs) must be clearly distinguished from participants who are withdrawn for other reasons. Investigators will follow up with participants who are withdrawn from the study as result of an SAE or AE until resolution of the event.

A participant who withdraws from the study may request destruction of any samples taken and not tested, and the investigator must document this in the study site study records.

If the participant's parent(s)/LAR(s) withdraw consent or the participant withdraws assent for disclosure of future information, the Sponsor may retain and continue to use any data collected before such a withdrawal of consent/assent (Section 10.2.10).

The Sponsor will continue to retain and use all research data that have already been collected for the study evaluation, unless the participant has requested destruction of these samples. All biological samples that have already been collected may be retained and analyzed at a later date (or as permitted by local regulations).

#### 6.4. Study Pause Rules

The investigators, study medical monitor, and Sponsor will monitor for events that could trigger a study pause. Study pause rule criteria, events, and thresholds are described in Table 4.

**Table 4: Pause Rule Criteria, Events, and Thresholds**

| Pause Rule Criterion | Event | Participant Threshold for Triggering Study Pause |
|---|---|---|
| 1 | Any death due to SARS-CoV-2 infection | ≥ 1 |
| 2 | Any SAE or Grade 4 AE that cannot be reasonably attributed to a cause other than vaccination | ≥ 1 |
| 3 | ICU Admission due to COVID-19 | ≥ 1 |
| 4[1] | Individual Grade 3 or higher solicited local AR lasting ≥ 24 hours and occurring within 7 days of injection (Days 1-7) | At least 2 participants and ≥ 5% of the dosed participants in an age group |
| 5[1] | Individual Grade 3 or higher solicited systemic AR lasting ≥ 24 hours and occurring within 7 days of injection (Days 1-7) | At least 2 participants and ≥ 10% of the enrolled participants in an age group |
| 6[1] | Any ≥ Grade 3 or higher unsolicited AE that cannot be reasonably attributed to a cause other than vaccination | At least 2 participants and ≥ 5% of the enrolled participants in an age group |

Abbreviations: AE = adverse event; AR = adverse reaction; COVID-19 = coronavirus disease 2019; ICU = intensive care unit; SAE = serious adverse event; SARS-CoV-2 = severe acute respiratory syndrome coronavirus 2.
[1] "Individual AR" is defined as 1 AR type, eg, pain, erythema, or headache could each be an "individual AR."

If any of the thresholds for a study pause is met, the Sponsor will immediately suspend further enrollment and/or study dosing by notifying all investigators. Such a suspension will remain in force until the threshold events are reviewed by the DSMB and a recommendation to continue is provided to the Sponsor.

The investigator or designee is responsible for reporting to the Sponsor, via the electronic data capture (EDC) system, each event that potentially meets any pause rule criterion within 24 hours of observation. The Sponsor will inform the DSMB of any event that potentially meets any pause rule criterion. The DSMB will review all available study data to help adjudicate such events in accordance with the DSMB charter.

The Sponsor will notify the Center for Biologics and Evaluation Research within 48 hours in the event of a study pause. In the event of a study pause, all safety and immunogenicity assessments will continue per protocol. The window allowance for injection visits may be extended by an additional 7 days (ie, + 21 days) for affected participants at the discretion of the Sponsor.

#### 6.5. Lost to Follow-up

A participant will be considered LTFU if he or she repeatedly fails to return for scheduled visits without stating an intention to withdraw consent/assent and is unable to be contacted by the study site. The following actions must be taken if a participant fails to return to the clinic for a required study visit:

- The study site staff must attempt to contact the participant and reschedule the missed visit as soon as possible, counsel the participant and his/her parent(s)/LAR(s) on the importance of maintaining the assigned visit schedule and ascertain whether the participant wishes to and/or should continue in the study.
- Before a participant is deemed LTFU, the investigator or designee must make every effort to regain contact with the participant (where possible, 3 telephone calls and, if necessary, a certified letter to the participant's last known mailing address or local equivalent methods). These contact attempts (eg, dates of telephone calls and registered letters) should be documented in the participant's medical record.
- A participant who continues to be unreachable or continues to be noncompliant with study visits or procedures will be considered to have withdrawn from the study.
- A participant should not be considered LTFU until due diligence has been completed.

### 7. STUDY ASSESSMENTS AND PROCEDURES

Before performing any study procedures, all potential participants and/or participants' parent(s)/LAR(s) will sign an ICF (as detailed in Section 10.2.6). Participants will undergo study procedures at the time points specified in the SoA (Table 10). A participant can also be seen for an unscheduled visit at any time during the study. An unscheduled visit may be prompted by reactogenicity issues, illness visit criteria for COVID-19, or new or ongoing AEs. The study site also has the discretion to make reminder telephone calls or send text messages to inform the participant and/or participant's parent(s)/LAR(s) about visits, review eDiary requirements, or follow-up on ongoing or outstanding issues.

In accordance with "FDA Guidance on Conduct of Clinical Trials of Medical Products during COVID-19 Public Health Emergency" (DHHS 2020), investigators may convert study site visits to home visits or telemedicine visits with the approval of the Sponsor. Such action should be taken to protect the safety and well-being of study participants and study site staff or to comply with state or municipal mandates.

General considerations for study assessments and procedures include the following:

- Protocol waivers or exemptions are not allowed. The study procedures and their timing must be followed as presented in Table 10. Adherence to the study design requirements is essential and required for study conduct.
- Immediate safety concerns should be discussed with the Sponsor immediately upon occurrence or awareness to determine if the participant should continue study treatment or participation in the study.
- All screening evaluations must be completed and reviewed to confirm that potential participants meet all eligibility criteria. The investigator will maintain a screening log to record details of all participants screened and to confirm eligibility or record reasons for screening failure, as applicable.
- Procedures conducted as part of the participant's routine clinical management (eg, pregnancy test) and obtained before signing of the ICF may be utilized for screening or baseline assessments provided the procedures met the protocol-specified criteria and were performed within the time frame defined in the SoA (Table 10).

#### 7.1. Safety Assessments and Procedures

Safety assessments will include monitoring and recording of the following for each participant, according to the SoA (Table 10):

- Solicited local and systemic ARs (Section 7.4.3) that occur during the 7 days following each injection (ie, the day of injection and 6 subsequent days). Solicited ARs will be recorded daily using eDiary (Section 7.1.1).
- Unsolicited AEs observed or reported during the 28 days following each injection (ie, the day of injection and 27 subsequent days). Unsolicited AEs are defined in Section 7.4.1.
- AEs leading to discontinuation from dosing and/or study participation from Day 1 through the last day of study participation.
- MAAEs (Section 7.4.4) from first dose on Day 1 through the entire study period.
- SAEs (Section 7.4.2) from first dose on Day 1 through the entire study period.
- AESIs (Section 7.4.5) including MIS-C and myocarditis and/or pericarditis, through the entire study period.
- Physical examination findings (Section 7.1.4).
- Assessments for SARS-CoV-2 infection from Day 1 through study completion (Section 7.1.5).
- Details of all pregnancies in female participants after the start of study treatment and until the end of their participation in the study (Section 7.4.6).

##### 7.1.1. Use of Electronic Diaries

At the time of consent, participants’ parent(s)/LAR(s) must confirm they will be willing to complete an eDiary using either an application downloaded to their smartphone or using a device that is provided at the time of enrollment. Before enrollment on Day 1, participants’ parent(s)/LAR(s) will be instructed to download the eDiary application or will be provided an eDiary device to record solicited ARs (Section 7.4.3) on Day 1. Based on availability, smartphone devices may be provided to those participants’ parent(s)/LAR(s) who do not have their own device to use for eDiary activities.

At each injection visit, participants’ parent(s)/LAR(s) will be instructed (Day 1) or reminded (Day 29) on thermometer (oral/tympanic) usage to measure body temperature, ruler usage to measure injection site erythema and swelling/induration (hardness), and assessment for localized axillary swelling or tenderness on the same side as the injection arm/thigh.

At each injection visit, participants' parent(s)/LAR(s) will record data into the eDiary starting approximately 30 minutes after injection under the supervision of the study site staff to ensure successful entry of assessments. The study site staff will perform any retraining as necessary. Participants' parent(s)/LAR(s) will continue to record data in the eDiary after they leave the study site, preferably in the evening and at the same time each day, on the day of injection and for 6 days following injection.

Participants' parent(s)/LAR(s) will record the following data in the eDiary:

- Solicited local and systemic reactogenicity ARs, as defined in Section 7.4.3, that occur on the day of each vaccine administration and during the 7 days after vaccine administration (ie, the day of injection and 6 subsequent days). If a solicited local or systemic AR continues beyond Day 7 after vaccination, the participant's parent(s)/LAR(s) will be prompted to capture details of the solicited local or systemic AR in the eDiary until it resolves or the next IP injection occurs, whichever occurs first. Capture of details of ARs in the eDiary should not exceed 28 days after each vaccination. Adverse reactions recorded in the eDiary beyond Day 7 should be reviewed by the study site staff either during the next scheduled telephone call or at the next study site visit.
- Daily oral (for participants > 4 years of age) or tympanic (for participants ≤ 4 years of age) body temperature measurement should be performed at approximately the same time each day using the thermometer provided by the study site. If body temperature is taken more than once in a given day, only the highest temperature reading should be recorded.
- Measurement, as applicable, for solicited local ARs (injection site erythema and swelling/induration); the size measurements will be performed using the ruler provided by the study site.
- Any medications taken to treat or prevent pain or fever on a day of injection or for the next 6 days.

The eDiary will be the only source document allowed for solicited systemic or local ARs (including body temperature measurements). Participants' parent(s)/LAR(s) will be instructed to complete eDiary entries daily. The participant's parent(s)/LAR(s) will have a limited window on the following day to complete assessments for the previous day; quantitative temperature recordings and measurement of any injection site erythema or swelling/induration reported on the following day may be excluded from the analyses of solicited ARs.

Any new safety information reported during safety telephone calls or at study site visits (including a solicited reaction) that is not already captured in the eDiary will be described in the source documents as a verbally reported event. An event reported in this manner must be described as a solicited event and entered on the solicited AR eCRF.

Study site staff will review eDiary data with participants' parent(s)/LAR(s) at telemedicine visits 7 days after each injection.

The eDiary will also be used every 4 weeks, starting at Day 71 through Day 183 and again starting at Day 223 through Day 363, to capture the occurrence of AEs, MAAEs, SAEs, AESI, or AEs leading to withdrawal. As specified in the SoA (Table 10), the eDiary will prompt the participant's parent(s)/LAR(s) to complete an eDiary questionnaire that collects the following data:

- Changes in health since last completing the questionnaire or since in contact with the study site
- Any MAAEs, AESIs, or SAEs
- Known close contact with someone in the household who has known COVID-19 or SARS-CoV-2 infection. Per the CDC, "close contact" to someone with COVID-19 is defined as follows:
  - Being within 6 feet for a total of 15 minutes or more
  - Providing care at home
  - Having direct physical contact (hugged or kissed them)
  - Sharing eating or drinking utensils
  - Being sneezed or coughed upon or getting respiratory droplets on the participant

Any experience of symptoms of COVID-19

If an eDiary record results in identification of relevant safety events according to the study period or of symptoms of COVID-19, a follow-up safety telephone call will be triggered.

Completion of eDiary questionnaires will alternate with safety telephone calls (Section 7.1.2) as the procedure for safety follow-up approximately every 4 weeks; these safety telephone calls will take place from Day 85 through Day 197 and again from Day 237 through Day 377 (Table 10).

###### 7.1.1.1. Ancillary Supplies for Participant Use

Study sites will distribute Sponsor-provided oral or tympanic thermometers and rulers for use by participants' parent(s)/LAR(s) in assessing body temperature and injection site reactions for recording solicited ARs in electronic diaries (eDiary). Based on availability, smartphone devices may be provided to those participants' parent(s)/LAR(s) who do not have their own device to use for eDiary activities.

##### 7.1.2. Safety Telephone Calls

A safety telephone call is a telephone call made to the participants' parent(s)/LAR(s) by trained study site personnel. This call will follow a script, which will facilitate the collection of relevant safety information. Safety telephone calls follow a schedule for each participant as indicated in the SoA (Table 10). The participants' parent(s)/LAR(s) will be interviewed according to the script about the occurrence of AEs, MAAEs, SAEs, AESI, AEs leading to study withdrawal, concomitant medications associated with those events, and any nonstudy vaccinations (Section 7.4.7). In addition, study personnel will collect information on known participant exposure to someone with known COVID-19 or SARS-CoV-2 infection and on participant experience of COVID-19 symptoms. All safety information collected from the telephone contact must be documented in source documents as described by the participant's parent(s)/LAR(s) and not documented on the script used for the safety telephone contact. As noted in Section 7.1.1, an unscheduled follow-up safety telephone call may be triggered if an eDiary record results in identification of a relevant safety event.

##### 7.1.3. Safety Laboratory Assessments

No scheduled laboratory assessments for safety are planned. This is based on the absence of clinically significant abnormal laboratory findings in the Phase 1 and Phase 2 studies of mRNA-1273 in adults.

A point-of-care urine pregnancy test will be performed, if deemed appropriate by the investigator, at the Screening Visit and before each vaccine dose in female participants of childbearing potential. At any time during the study, a pregnancy test either via blood or point-of-care urine can be performed, at the discretion of the investigator.

Febrile participants at dosing visits (fever is defined as a body temperature ≥ 38.0°C/≥ 100.4°F) may have visits rescheduled within the relevant study visit windows. Afebrile participants with minor illnesses may receive the IP at the discretion of the investigator.

##### 7.1.4. Physical Examinations

A full physical examination, including body temperature (oral preferred for participants > 4 years of age, tympanic preferred for participants ≤ 4 years of age, but other methods are acceptable in context of COVID-19 precautions), length/height and weight, will be performed at the Screening Visit or on Day 1. The full examination will include assessment of skin, head, ears, eyes, nose, throat, neck, lungs, heart, abdomen, lymph nodes, and musculoskeletal system/extremities. Any clinically significant finding identified during a study visit should be reported as an MAAE.

Symptom-directed physical examinations will be performed on Day 1, Day 29, Day 43 (if the visit is applicable), Day 57, Day 209, and Day 394 and may be performed at other time points at the discretion of the investigator. Body temperature (oral/tympanic) should be measured on each injection day prior to injection. On each injection day before injection and again 7 days after injection (ie, the day of injection and 6 subsequent days), the injection site should be examined. Any clinically significant finding identified during a study visit should be reported as an MAAE.

Participants who are febrile (body temperature ≥ 38.0°C/≥ 100.4°F) before injection on Day 1 or Day 29 must have the visit rescheduled within the relevant visit window to receive the injection. Afebrile participants with minor illnesses may be injected at the discretion of the investigator.

Body mass index will be calculated for children ≥ 2 years of age at the Screening Visit only.

##### 7.1.5. Assessment for SARS-CoV-2 Infection

Study participants will have nasal swab samples collected for SARS-CoV-2 testing at the time points specified in the SoA (Table 10).

A study illness visit (study site visit or home visit) will be arranged within 72 hours or as soon as possible if a participant experiences any of the following:

- Signs or symptoms of SARS-CoV-2 infection as defined by the CDC (CDC 2020b)
- MAAE suggesting a SARS-CoV-2 infection

A study illness visit (study site visit or home visit) will be arranged within 4 to 6 days after last exposure* if a participant experiences the following:

- Exposure (close contact [definition in Section 7.1.1]) to an individual in the household confirmed to be infected with SARS-CoV-2

*Last exposure is defined as the last day a child was in close contact with a symptomatic person in the household (if the child was then isolated from that person) or the last day of the quarantine of the person the child was exposed to, if that person was asymptomatic and/or the child was unable to isolate from that person.

If the participant had a known exposure (close contact) to COVID-19 (eg, exposure to someone with confirmed COVID-19 in the household), it will be captured in the COVID-19 exposure form.

Additionally, clinical information will be carefully collected to evaluate the severity of the clinical case.

At each study illness visit, an initial assessment will be performed to determine general appearance. This initial assessment may be performed by physicians, advanced practice nurses, physician assistants, or registered nurses. If indicated, a physical examination by a study clinician (MD, DO, NP, or PA experienced in pediatric examination) may occur.

The study illness visit (study site visit or home visit) may collect additional clinical information, including assessments such as updated medical history, physical examination, blood sampling for clinical laboratory testing, and nasal swab sampling for viral RT-PCR to evaluate the severity of the clinical case. Radiologic imaging studies may be conducted. Study site may also collect an additional nasal swab sample for SARS-CoV-2 testing to be able to render appropriate medical care for the study participant as determined by local standards of care. All findings will be recorded in the eCRF.

If participants are confirmed to have SARS-CoV-2 infection, the investigator will notify the participant's parent(s)/LAR(s) and the participant's primary care physician of the diagnosis via fax or other Health Information Portability and Accountability Act compliant electronic data transfer (eg, e-mail if applicable). If the study participant does not have a primary care physician, the investigator will assist them to obtain one. The participant will also be instructed on infection prevention measures consistent with local public health guidance. Laboratory test results for SARS-CoV-2 infection in study participants should be submitted to state or local public health departments according to local policy. The investigator must either directly report to state or local public health department or obtain confirmation from the participant's primary care physician that the positive test was reported.

Any confirmed symptomatic SARS-CoV-2 infection that occurs in participants will be captured as an MAAE along with relevant concomitant medications and details about severity, seriousness, and outcome. Additionally, a convalescent visit will be scheduled approximately 28 days (+7 days) after diagnosis (Section 7.3.3). At this visit, a nasal swab sample for viral RT-PCR and a blood sample for potential immunologic assessment of SARS-CoV-2 infection will be collected.

If a participant tests positive at baseline but remains asymptomatic, Day 29 visit can be used as the convalescent visit to obtain a convalescent blood sample; the second dose may still be administered as long as participant remains asymptomatic and no other concerns arise.

#### 7.2. Blood Collections for Immunogenicity Assessments and Biomarker Samples

Blood samples for immunogenicity assessments and biomarker samples will be collected at the time points indicated in the SoA (Table 10, Table 11, and Table 12). The following analytes will be measured:

- Serum nAb titer against SARS-CoV-2 as measured by pseudovirus and/or live virus neutralization assays.
- Serum bAb titer as measured by enzyme-linked immunosorbent assay specific to the SARS-CoV-2 S protein.

- For Part 1, testing for serologic markers for SARS-CoV-2 infection using a nonvaccine antigen-based blood test at Day 1 (prior to the first dose), Day 57, Day 209, and Day 394.

For Part 2, participants in each age group will be assigned to 5 phlebotomy cohorts. Blood sampling will be performed in 3 of these 5 cohorts for immunogenicity at 3 of the following time points: Day 1 (prior to randomization and the first dose), Day 57, and one of Day 29 (prior to the second dose), Day 209, and Day 394. Participants in the Cohort D (remainder of the age group) will provide a blood sample at Day 1 (prior to randomization and the first dose) and within 4 days of receiving Dose 2 at Day 30 (+3 days) for storage and potential future biomarker testing (may include cardiac biomarkers). For participants already enrolled in Cohort D prior to amendment 4, this blood draw will be optional and confirmed at time of re-consenting; for all participants newly enrolled into Cohort D under amendment 4, it will be mandatory. A fifth cohort of participants at selected VTEU sites only, will provide blood samples for assessment of exploratory serology and CMI on Day 1 (prior to randomization and the first dose), Day 43, Day 209, and Day 394.

Sample aliquots will be designed to have backup samples, if possible; vial volumes will likely be adequate for future testing needs. The actual time and date of each sample collected will be recorded in the eCRF, and unique sample identification will be utilized to maintain the blind at the laboratory at all times and to allow for automated sample tracking and housing. Handling and preparation of the samples for analysis, as well as shipping and storage requirements, will be provided in a separate study laboratory manual.

Measurement of bAb and nAb levels will be performed in a laboratory designated by the Sponsor.

According to the ICF section (Section 10.2.6), serum from immunogenicity testing may be used for future research, which may be performed at the discretion of the Sponsor to further characterize the immune response to SARS-CoV-2, additional assay development, and the immune response across CoVs.

The planned volume of blood to be sampled per participant in Part 1 and Part 2 (serology cohorts) for each age group for immunogenicity assessments in 1 day is as follows:

- 6 months to < 2 years: approximately 8 mL
- 2 to < 6 years: approximately 16 mL
- 6 to < 12 years: approximately 24 mL

The planned volume of blood to be sampled per participant at the Day 30 (+3 days) blood draw for storage and potential future biomarker testing for Cohort D in Part 2 is as follows:

- All ages: approximately 4 mL

The planned volume of blood to be sampled per participant in the exploratory serology and CMI cohort for each age group in 1 day is as follows:

- 6 months to < 2 years: approximately 9 mL
- 2 to < 6 years: approximately 9 mL
- 6 to < 12 years: approximately 17 mL

Note: If less than 8 mL of blood is obtained for any given child at baseline, the child cannot be enrolled.

#### 7.3. Inferred Efficacy Assessments

##### 7.3.1. Vaccine Effectiveness Assessments

Vaccine effectiveness for children 6 months to < 12 years of age will be inferred based on serum antibody responses obtained on Day 57 (28 days after the second injection of mRNA-1273). Inference will be based on assessing the antibody responses against the following:

1. *If available at the time of analysis*, antibody responses will be assessed against an accepted serum antibody threshold conferring protection against COVID-19.
2. *If an accepted threshold of protection is not available*, noninferiority of the GM value of serum antibody and seroresponse rate of children 6 months to < 12 years of age (Study P204) compared with the GM value of serum antibody and seroresponse rate from young adults (18 to 25 years of age) enrolled in the ongoing clinical endpoint efficacy trial (Study P301) will be assessed. The statistical parameters to infer effectiveness are described in Section 2.

**COVID-19:**

To be considered as a case of COVID-19 for the evaluation of the efficacy endpoint, the following case definition must be met:

- The participant must have at least 1 nasal swab (or respiratory sample, if hospitalized) positive for SARS-CoV-2 by RT-PCR AND
- ONE of the following:
  - Fever (temperature ≥ 38ºC/≥100.4ºF) or chills (of any duration, including ≤ 4 hours)
  - Shortness of breath or difficulty breathing (of any duration, including ≤ 48 hours)
  - Cough (of any duration, including ≤ 48 hours)
  - Fatigue

- Muscle or body aches
- Headache
- New loss of taste or smell
- Sore throat
- Congestion or runny nose
- Abdominal pain
- Nausea or vomiting
- Diarrhea
- Poor appetite/poor feeding

**Severe COVID-19:**

To be considered severe COVID-19, the following criteria must be met:

- Confirmed COVID-19 as per the COVID-19 case definition, plus any of the following:
  - Meeting criteria for systemic inflammatory response syndrome based on age -specific variables (Table 5) OR
  - Respiratory failure or acute respiratory distress syndrome (defined as needing high-flow oxygen, noninvasive or mechanical ventilation, or extracorporeal membrane oxygenation), medical intervention for shock (intravenous fluids, vasopressors, etc), OR
  - Significant acute renal, hepatic, or neurologic dysfunction (Table 6), OR
  - Admission to an ICU or death.

**Table 5: Age-Specific Cut-Offs for Vital Signs and Laboratory Variables**

| Age Group | Heart Rate, Beats/Min | | Respiratory Rate, Breaths/Min | Leukocyte Count, Leukocytes × $10^3/mm^3$ | Systolic Blood Pressure, mm Hg |
|---|---|---|---|---|---|
| | Tachycardia | Bradycardia | | | |
| 1 month to 1 year | > 180 | < 90 | > 34 | > 17.5 or < 5 | < 70 |
| 2 to 5 year | > 140 | NA | > 22 | > 15.5 or < 6 | < 70 + (age in years x 2) |
| 6 to 10 year | > 130 | NA | > 18 | > 13.5 or < 4.5 | < 70 + (age in years x 2) |
| >10 years | > 130 | NA | >18 | > 13.5 or < 4.5 | < 90 |

Abbreviations: AHA = American Heart Association; NA = not applicable.

Note: Lower values for heart rate, leukocyte count, are for the 5th percentile and upper values for heart rate, respiratory rate, or leukocyte count are for the 95th percentile for that age group. Systolic blood pressure values are based on AHA definitions of hypotensive shock for pediatric population.
Source: Goldstein et al 2005 and Topjian et al 2020.

##### Table 6: Definition of Renal-, Liver- and Neurological Dysfunction for Pediatric Population (< 12 years of age)

| | |
|---|---|
| **Acute Renal Dysfunction** | • Increase in serum creatinine by ≥ 0.3 mg/dL (≥26.5 µmol/L) within 48 hours;<br>OR<br>• Increase in serum creatinine to ≥ 1.5 times baseline, known or presumed to have occurred within prior 7 days<br>OR<br>• Urine volume ≤ 0.5 mL/ kg/ hour for 6 hours |
| **Acute Liver Dysfunction** | ○ > 3-fold elevation above the upper normal limit for ALT or AST<br>OR<br>• > 2-fold elevation above the upper normal limit for total serum bilirubin or GGT or ALP |
| **Acute Neurological Dysfunction**[a] | ANY of the following:<br>• Loss of sense of smell or taste<br>• Seizures or status epilepticus<br>• Severe headache (preventing normal daily activities)<br>• Persistent difficulty walking or crawling (if crawling/walking before)<br>• Persistent altered awareness or confusion (preventing normal daily activities)<br>• Persistent severe fatigue or weakness (preventing normal daily activities) |

Abbreviations: ALT = alanine aminotransferase; ALP = alkaline phosphatase; AST = aspartate aminotransferase; GGT = gamma-glutamyl transferase.
Source: [a]LaRovere et al.

Death attributed to COVID-19 is defined as any participant who dies during the study with a cause directly attributed to a complication of COVID-19.

**SARS-CoV-2 Infection:**

- SARS-CoV-2 infection is defined in participants with SARS-CoV-2 negative at baseline:
  - bAb level against SARS-CoV-2 nucleocapsid protein negative at Day 1 that becomes positive (as measured by Roche Elecsys) postbaseline, OR
  - Positive RT-PCR postbaseline

##### 7.3.2. Surveillance for COVID-19 Symptoms

Surveillance for COVID-19 symptoms will be conducted via biweekly telephone calls or eDiary prompts as specified in Section 7.1.1 and Figure 2; starting after participant enrollment and continuing throughout the study.

If there is no response to an eDiary prompt for 2 days, the study site staff will contact the study participant by telephone.

According to the CDC, as of 22 Dec 2020 (CDC 2020c), patients with COVID-19 have reported a wide range of symptoms ranging from mild symptoms to severe illness. Throughout the study, to survey for COVID-19, the following pre-specified symptoms that meet the criteria for suspicion of COVID-19 will be elicited weekly from the participant, and the presence of any one of these symptoms (in the absence of an alternative diagnosis) lasting at least 48 hours (except for fever and/or respiratory symptoms) will result in the study site staff arranging an illness visit to collect a nasal swab for SARS-CoV-2 within 72 hours.

- Fever (temperature ≥ 38ºC/≥ 100.4ºF) or chills (of any duration, including ≤ 48 hours)
- Shortness of breath or difficulty breathing (of any duration, including ≤ 48 hours)
- Cough (of any duration, including ≤ 48 hours)
- Fatigue
- Muscle or body aches
- Headache
- New loss of taste or smell
- Sore throat
- Congestion or runny nose
- Abdominal pain
- Nausea or vomiting
- Diarrhea
- Poor appetite/poor feeding

**Figure 2: Surveillance for COVID-19 Symptoms and the Corresponding Clinical Data Pathways**

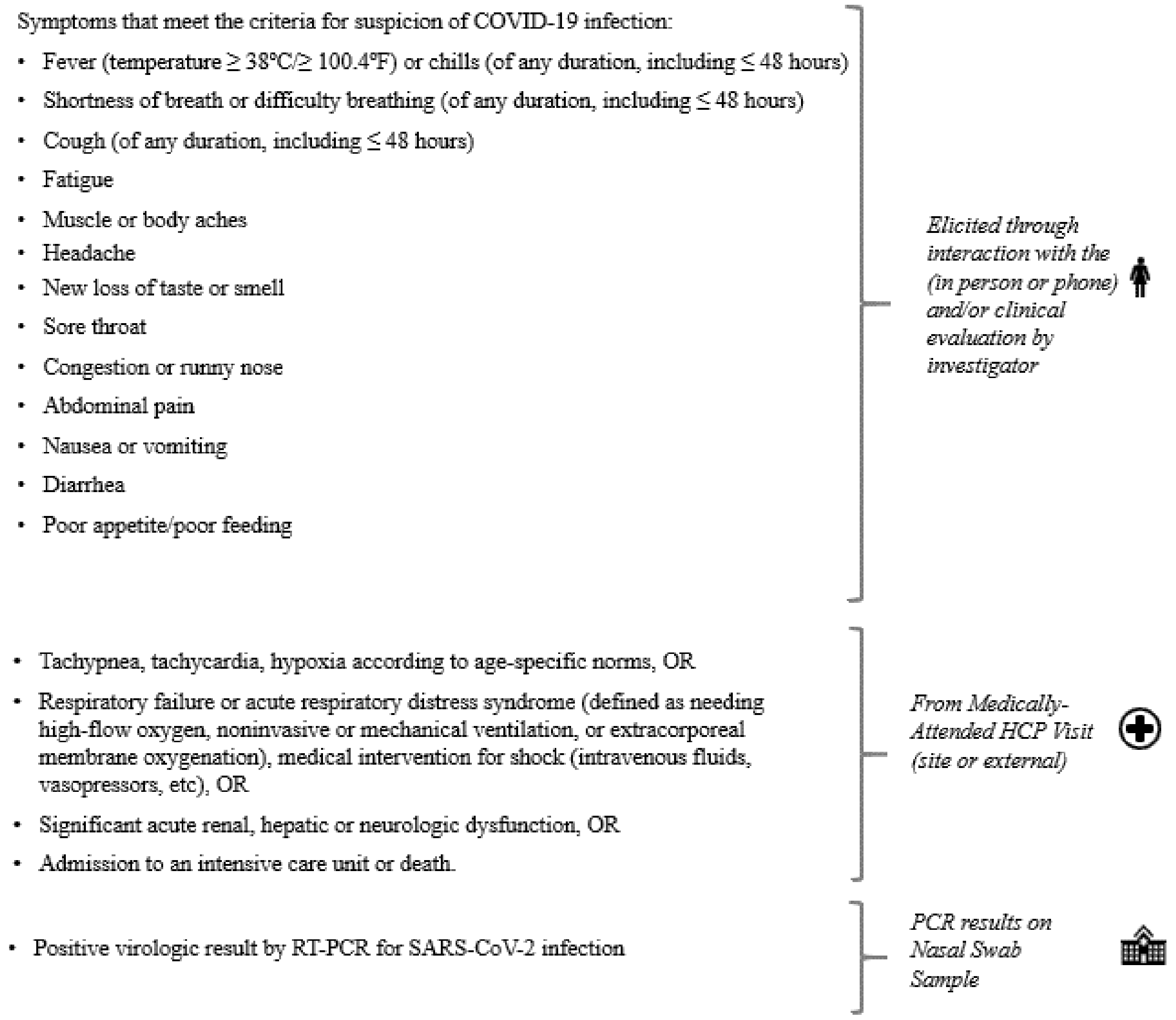

Abbreviations: COVID-19 = coronavirus disease 2019, HCP = healthcare practitioner, RT-PCR = reverse transcriptase polymerase chain reaction, SARS-CoV-2 = severe acute respiratory syndrome coronavirus 2.

It is important to note that some of the symptoms of COVID-19 overlap with solicited systemic ARs that are expected after vaccination with mRNA-1273 (eg, myalgia, headache, fever, and chills). During the first 7 days after vaccination, when these solicited ARs are common, investigators should use their clinical judgment to decide if a nasal swab should be collected. The collection of a nasal swab prior to the first dose on Day 1, prior to the second dose on Day 29, and then at all subsequent study visits (Day 43 [if visit is applicable], Day 57, Day 209, and Day 394) can help ensure that cases of COVID-19 are not overlooked. Any study participant who reports respiratory symptoms during the 7-day period after vaccination without an alternative diagnosis should be evaluated for COVID-19.

For children with febrile illnesses, if an alternative diagnosis is identified (eg, positive urine culture, streptococcal pharyngitis, cellulitis), the investigator may decide to omit the illness visit. Identification of an alternative viral agent (such as RSV or influenza by rapid testing) does not satisfy this requirement, as co-infections with SARS-CoV-2 may occur.

An investigator may elect to omit the illness visit if standard of care testing prior to the illness visit reveals a negative COVID-19 nucleic acid amplification test performed at a CLIA-certified or CLIA-waived laboratory and the principal investigator can obtain a copy of the negative test. Home testing kits cannot be used to satisfy this testing requirement. Standard of care evaluation of household contacts that reveals a negative COVID-19 is not sufficient to omit an illness visit in the study participant.

During the course of the study, participants with symptoms of COVID-19 will be asked to return within 72 hours or as soon as possible to the study site or trained staff from the study site will conduct a home visit as soon as possible to collect a nasal swab sample (for RT-PCR) for evaluation of COVID-19. Both study site visits and home visits are referred to as illness visits (Section 7.1.5). Additionally, a convalescent visit will be scheduled approximately 28 days (+7 days) after diagnosis (Section 7.3.3). At this visit, a nasal swab sample for viral RT-PCR and a blood sample will be collected for potential immunologic assessment of SARS-CoV-2 infection.

Cases are defined as participants meeting clinical criteria based both on symptoms for COVID-19 and on RT-PCR detection of SARS-CoV-2 from samples collected within 72 hours of the study participant reporting symptoms meeting the definition of COVID-19. Participants who are hospitalized for COVID-19 without the opportunity for a clinic or home visit will also be considered cases, assuming that the symptomology criteria for COVID-19 are met and a respiratory sample is positive for SARS-CoV-2 by RT-PCR at a certified laboratory.

Investigators are encouraged to try to obtain a respiratory sample during the course of hospitalization or immediately after hospital discharge as an unscheduled visit for submission to the study central laboratory, if feasible. The investigator should determine if the criteria for severe COVID-19 have been met.

Severe COVID-19 is defined in Section 7.3.1.

All clinical findings will be recorded in the eCRF. All confirmed cases of COVID-19 will be captured as MAAEs, along with relevant concomitant medications and details about severity, seriousness, and outcome, and will be reported immediately to the Sponsor or designee (Section 7.4.4).

##### 7.3.3. Follow-up/Convalescent Period After Diagnosis with COVID-19

Any confirmed COVID-19 occurring in a participant will be captured as an MAAE along with relevant concomitant medications and details about severity, seriousness, and outcome. Additionally, a convalescent visit will be scheduled approximately 28 days (+7 days) after diagnosis. At this visit, a nasal swab sample for viral RT-PCR and a blood sample for potential immunologic assessment of SARS-CoV-2 infection will be collected (Table 10). The investigator should determine if the criteria for severe COVID-19 have been met. If the participant is hospitalized, study site personnel will try to obtain medical records and SARS-CoV-2 diagnostic results. If the participant is later discharged from the hospital during the 28 day period following diagnosis of COVID-19, the study site personnel will arrange for a resumption of the protocol schedule.

Children hospitalized with possible or confirmed MIS-C will also have the same convalescent visit scheduled approximately 28 days (+7 days) after onset of hospitalization. At this visit, a nasal swab sample for viral RT-PCR and a blood sample for potential immunologic assessment of SARS-CoV-2 infection will be collected (see Table 10).

#### 7.4. Safety Definitions and Related Procedures

##### 7.4.1. Adverse Event

An AE is defined as any untoward medical occurrence associated with the use of a drug in humans, whether or not considered drug related.

**Events Meeting the Adverse Event Definition**

- Exacerbation of a chronic or intermittent pre-existing condition including either an increase in frequency and/or intensity of the condition.
- New conditions detected or diagnosed after the first dose of IP even though they may have been present before the start of the study.

**Events NOT Meeting the Adverse Event Definition**

- Medical or surgical procedure (eg, endoscopy, appendectomy): the condition that leads to the procedure should be the AE.
- Situations in which an untoward medical occurrence did not occur (social and/or convenience admission to a hospital).
- Planned procedures (eg, tonsillectomy or pressure-equalization tubes) that occur during the study period but were planned prior to enrollment will not be considered AE unless complications arise.

An AR is any AE for which there is a reasonable possibility that the vaccine caused the AE (Section 7.4.9). For the purposes of investigational new drug safety reporting, “reasonable possibility” means that there is evidence to suggest a causal relationship between the vaccine and the AE.

An unsolicited AE is any AE reported by the participant that is not specified as a solicited AR in the protocol or is specified as a solicited AR but starts outside the protocol-defined period for reporting solicited ARs (ie, the day of each dose of injection and the 6 days after the day of dosing).

##### 7.4.2. Serious Adverse Events

An AE (including an AR) is considered an SAE if, in the view of either the investigator or Sponsor, it results in any of the following outcomes:

- **Death**
  A death that occurs during the study or that comes to the attention of the investigator during the protocol-defined follow-up period must be reported to the Sponsor, whether or not it is considered related to the IP.
- **Is life-threatening**
  An AE is considered life-threatening if, in the view of either the investigator or the Sponsor, its occurrence places the participant at immediate risk of death. It does not include an AE that, had it occurred in a more severe form, might have caused death.
- **Inpatient hospitalization or prolongation of existing hospitalization**
  In general, inpatient hospitalization indicates the participant was admitted to the hospital or emergency ward for at least one overnight stay for treatment that would not have been appropriate in the physician’s office or outpatient setting. The hospital or emergency ward admission should be considered an SAE regardless of whether opinions differ as to the necessity of the admission. Complications that occur during inpatient hospitalization will be recorded as AEs; however, if a complication/AE prolongs hospitalization or otherwise fulfills SAE criteria, the complication/AE will be recorded as a separate SAE. Note: 24-hour observation admissions, typically used to extend the period of observation from an emergency department or urgent care visit, will not be considered inpatient hospitalization unless they are converted to hospital admission after the 24 hours of observation have expired.
- **Persistent or significant incapacity or substantial disruption of the ability to conduct normal life functions**
  This definition is not intended to include experiences of relatively minor medical significance such as uncomplicated headache, nausea/vomiting, diarrhea, influenza, and

accidental trauma (eg, sprained ankle) that may interfere with or prevent everyday life functions but do not constitute a substantial disruption.

- **Congenital anomaly or birth defect**
- **Medically important event**
  Medical judgment should be exercised in deciding whether SAE reporting is appropriate in other situations, such as important medical events that may not be immediately life-threatening or result in death or hospitalization but that jeopardize the participant or require medical or surgical intervention to prevent one of the other outcomes listed in the above definition. These events should usually be considered serious. Examples of such medical events include allergic bronchospasm requiring intensive treatment in an emergency room or at home, blood dyscrasias or convulsions that do not result in inpatient hospitalization, or the development of drug dependency or drug abuse.

##### 7.4.3. Solicited Adverse Reactions

The term “reactogenicity” refers to the occurrence and intensity of selected signs and symptoms (ARs) that occur after IP injection. The eDiary will solicit daily participant reporting of ARs using a structured checklist (Section 7.1.1). Participant’s parent(s)/LAR(s) will record such occurrences in an eDiary on the day of each dose of injection and for the 6 days after the day of dosing.

Severity grading of reactogenicity will occur automatically based on participant entry into the eDiary according to the grading scales presented in Table 6 and Table 7, modified from the Toxicity Grading Scale for Healthy Adult and Adolescent Volunteers Enrolled in Preventative Vaccine Clinical Trials (DHHS 2007).

If a solicited local or systemic AR continues beyond Day 7 after vaccination, the participant’s parent(s)/LAR(s) will be prompted to capture details of the solicited local or systemic AR in the eDiary until it resolves or the next IP injection occurs, whichever occurs first. Capture of details of ARs in the eDiary should not exceed 28 days after each vaccination. Adverse reactions recorded in the eDiary beyond Day 7 should be reviewed by the study site staff either during the next scheduled telephone call or at the next study site visit. All solicited ARs (local and systemic) will be considered related to the IP.

**Table 6: Solicited Adverse Reactions and Grades: Age 37 Months to < 12 Years[1]**

| Reaction | Grade 0 | Grade 1 | Grade 2 | Grade 3 | Grade 4[2] |
|---|---|---|---|---|---|
| Injection site pain | None | Does not interfere with activity | Interferes with activity | Prevents daily activity | Requires emergency room visit[3] or hospitalization |
| Injection site erythema (redness) | < 25 mm/ < 2.5 cm | 25-50 mm/ 2.5-5 cm | 51-100 mm/ 5.1-10 cm | > 100 mm/ > 10 cm | Necrosis or exfoliative dermatitis |
| Injection site swelling/induration (hardness) | < 25 mm/ < 2.5 cm | 25-50 mm/ 2.5-5 cm | 51-100 mm/ 5.1-10 cm | > 100 mm/ > 10 cm | Necrosis |
| Axillary (underarm) swelling or tenderness ipsilateral to the side of injection | None | No interference with activity | Some interference with activity | Prevents daily activity | Emergency room visit[3] or hospitalization |
| Headache | None | No interference with activity | Some interference with activity | Significant; Prevents daily activity | Requires emergency room visit[3] or hospitalization |
| Fatigue | None | No interference with activity | Some interference with activity | Significant; prevents daily activity | Requires emergency room visit[3] or hospitalization |
| Myalgia (muscle aches all over body) | None | No interference with activity | Some interference with activity | Significant; prevents daily activity | Requires emergency room visit[3] or hospitalization |
| Arthralgia (joint aches in several joints) | None | No interference with activity | Some interference with activity | Significant; prevents daily activity | Requires emergency room visit[3] or hospitalization |
| Nausea/vomiting | None | No interference with activity or 1-2 episodes/ 24 hours | Some interference with activity or > 2 episodes/ 24 hours | Prevents daily activity, | Requires emergency room visit[3] or hospitalization for hypotensive shock |
| Chills | None | No interference with activity | Some interference with activity not requiring medical intervention | Prevents daily activity and requires medical intervention | Requires emergency room visit[3] or hospitalization |
| Fever | < 38.0°C < 100.4°F | 38.0-38.4°C 100.4-101.1°F | 38.5-38.9°C 101.2-102.0°F | 39.0-40.0°C 102.1-104.0°F | > 40.0°C > 104.0°F |

| Reaction | Grade 0 | Grade 1 | Grade 2 | Grade 3 | Grade 4[2] |
|---|---|---|---|---|---|

[1] Age at time of enrollment determines the scale to be used.
[2] Grading for Grade 4 events per investigator assessment (with exception of fever).
[3] Emergency room visit includes urgent care visit.
Source: Guidance for industry - Toxicity Grading Scale for Healthy Adult and Adolescent Volunteers Enrolled in Preventative Vaccine Clinical Trials (DHHS 2007).

#### Table 7: Solicited Adverse Reactions and Grades: Age 6 to ≤ 36 Months[1]

| Reaction | Grade 0 | Grade 1 | Grade 2 | Grade 3 | Grade 4[2] |
|---|---|---|---|---|---|
| **Local Reaction** | | | | | |
| Injection site pain/tenderness | None | Mild discomfort to touch or some pain but no interference with normal daily activities | Cries when limb is moved/refuses to move limb or pain interferes with normal daily activities | Significant pain at rest or pain prevents normal daily activities | Requires emergency room visit[3] or hospitalization |
| Injection site erythema (redness) | < 5 mm/ < 0.5 cm | 5-20 mm/ 0.5-2.0 cm | > 20-50 mm > 2.0-5.0 cm | > 50 mm/ > 5 cm | Necrosis or exfoliative dermatitis |
| Injection site swelling/induration (hardness) | < 5 mm/ < 0.5 cm | 5-20 mm/ 0.5-2.0 cm | > 20-50 mm > 2.0-5.0 cm | > 50 mm/ > 5 cm | Necrosis |
| Groin or underarm swelling or tenderness ipsilateral to the side of injection | None | Some swelling or tenderness but no interference with normal daily activities | Swelling or tenderness that interferes with normal daily activities | Swelling or tenderness that prevents normal daily activities | Emergency room visit[3] or hospitalization |
| **Systemic Reaction** | | | | | |
| Fever | < 38.0°C | 38.0-38.4°C 100.4-101.1°F | 38.5-39.5°C 101.2-103.1°F | 39.6-40.0°C 103.2-104.0°F | > 40.0°C |
| Irritability/crying | None | Lasting < 1 hour or easily consolable | Lasting 1-3 hours or requiring increased attention | Lasting > 3 hours or inconsolable | Requires emergency room visit[3] or hospitalization |
| Sleepiness | None | Sleepier than usual or less interested in surroundings | Not interested in surroundings or sleeps through meals | Sleeps most of the time, hard to arouse | Inability to arouse |

| Reaction | Grade 0 | Grade 1 | Grade 2 | Grade 3 | Grade 4[2] |
|---|---|---|---|---|---|
| Loss of appetite | None | Eating less than normal for 1-2 feeds/meals | Missed 1-2 feeds/ meals completely | Missed > 2 feeds/meals completely or refuses most feeds/meals | Requires emergency room visit[3] or hospitalization |

[1] Age at time of enrollment determines the scale to be used.
[2] Grading for Grade 4 events per investigator assessment (with exception of fever).
[3] Emergency room visit includes urgent care visit.
Source: Guidance for industry - Toxicity Grading Scale for Healthy Adult and Adolescent Volunteers Enrolled in Preventative Vaccine Clinical Trials (DHHS 2007).

Any solicited AR that meets any of the following criteria must be entered into the participant's source document and must also be recorded by the study site staff on the solicited AR page of the participant's eCRF:

- Solicited local or systemic AR that results in a visit to a healthcare practitioner (HCP; otherwise meets the definition of an MAAE)
- Solicited local or systemic AR leading to the participant withdrawing from the study or the participant being withdrawn from the study by the investigator (AE leading to withdrawal)
- Solicited local or systemic AR lasting beyond 7 days after injection
- Solicited local or systemic AR that leads to participant withdrawal from IP
- Solicited local or systemic AR that otherwise meets the definition of an SAE

##### 7.4.4. Medically Attended Adverse Events

An MAAE is an AE that leads to an unscheduled visit to an HCP. This would include visits to a study site for unscheduled assessments (eg, abnormal laboratory test result follow-up, COVID-19 [Section 7.3.1]) and visits to HCPs external to the study site (eg, urgent care, primary care physician). Investigators will review unsolicited AEs for the occurrence of any MAAE. Unsolicited AEs will be captured on the AE page of the eCRF.

All confirmed COVID-19 cases (Section 7.3.1) will be recorded as MAAEs and reported to the Sponsor or designee immediately and in all circumstances within 24 hours, using the SAE Mailbox, the SAE Hotline, or the SAE Fax line (Section 7.4.11). The investigator will submit any updated COVID-19 case data to the Sponsor within 24 hours of it being available.

All suspected cases of anaphylaxis should be recorded as MAAEs and reported as an SAE, based on criteria for a medically important event, unless the event meets other serious criteria. As an SAE, the event should be reported to the Sponsor or designee immediately and in all circumstances within 24 hours as per Section 7.4.11. The investigator will submit any updated anaphylaxis case data to the Sponsor within 24 hours of it being available. For reporting purposes, a participant who displays signs/symptoms consistent with anaphylaxis as described below should be reported as a potential case of anaphylaxis. This is provided as general guidance for investigators and is based on the Brighton Collaboration case definition (Rüggeberg et al 2007).

Anaphylaxis is an acute hypersensitivity reaction with multi-organ-system involvement that can present as, or rapidly progress to, a severe life-threatening reaction. It may occur following exposure to allergens from a variety of sources.

Anaphylaxis is a clinical syndrome characterized by:

Sudden onset AND

Rapid progression of signs and symptoms AND

Involves 2 or more organ systems, as follows:

- **Skin/mucosal:** urticaria (hives), generalized erythema, angioedema, generalized pruritus with skin rash, generalized prickle sensation, red and itchy eyes
- **Cardiovascular:** measured hypotension, clinical diagnosis of uncompensated shock, loss of consciousness or decreased level of consciousness, evidence of reduced peripheral circulation
- **Respiratory:** bilateral wheeze (bronchospasm), difficulty breathing, stridor, upper airway swelling (lip, tongue, throat, uvula, or larynx), respiratory distress, persistent dry cough, hoarse voice, sensation of throat closure, sneezing, rhinorrhea
- **Gastrointestinal:** diarrhea, abdominal pain, nausea, vomiting

##### 7.4.5. Adverse Events of Special Interest

An AESI is an AE (serious or nonserious) of scientific and medical concern specific to the Sponsor’s product or program for which ongoing monitoring and immediate notification by the investigator to the Sponsor is required and documentation in the form of a case narrative. Such events may require further investigation to characterize and understand them. Refer to Section 10.4, Appendix 4 for a list of AESIs pertinent to this study. All AESIs will be collected through the entire study period and must be reported to the Sponsor or designee immediately and in all circumstances within 24 hours of becoming aware of the event via the EDC system. If a site receives a report of a new AESI from a study participant or receives updated data on a previously reported AESI and the eCRF has been taken offline, then the site can report this information on a paper AESI form using the SAE Mailbox, the SAE Hotline, or the SAE Fax line (Section 7.4.11).

**Acute Myocarditis and/or Pericarditis**

All suspected cases of probable and confirmed myocarditis, pericarditis, or myopericarditis should be recorded as an AESI and reported as an SAE, if the event meets seriousness criteria. As an SAE, the event should be reported to the Sponsor or designee immediately and in all circumstances within 24 hours as per Section 7.4.11. The investigator will submit any updated myocarditis, pericarditis or myopericarditis case data to the Sponsor within 24 hours of it being available. For reporting purposes, a participant who displays signs/symptoms consistent with the CDC case definition as described below (Gargano et al, 2021), should be reported as a potential case of confirmed or probable myocarditis, pericarditis, or myopericarditis.

**Acute Myocarditis Case Definition**

Presence of ≥ 1 new or worsening of the following clinical symptoms (persons who lack the listed symptoms but who meet other criteria may be classified as subclinical myocarditis [probable or confirmed]):

- Chest pain/pressure/discomfort
- Dyspnea/shortness of breath/pain with breathing
- Palpitations
- Syncope

OR

Infants and children aged < 12 years might instead have ≥ 2 of the following symptoms:

- Irritability
- Vomiting
- Poor feeding
- Tachypnea
- Lethargy

AND

<u>For PROBABLE CASE:</u>

Presence of ≥ 1 new finding of the following:

- Troponin level above upper limit of normal (any type of troponin)
- Abnormal electrocardiogram ECG or EKG or rhythm monitoring findings consistent with myocarditis

- To meet the ECG or rhythm monitoring criterion, a probable case must include at least one of the following:
  - ST segment or T-wave abnormalities
  - Paroxysmal or sustained atrial, supraventricular, or ventricular arrhythmias
  - AV nodal conduction delays or intraventricular conduction defects

- Abnormal cardiac function or wall motion abnormalities on echocardiogram
- cMRI finding consistent with myocarditis (Ferreira et al, 2018)

AND

- No other identifiable cause of the symptoms and findings

<u>For CONFIRMED CASE:</u>

- Histopathologic confirmation of myocarditis (using Dallas criteria [Aretz, 1987])

OR

- cMRI findings consistent with myocarditis in the presence of troponin level above upper limit of normal (any type of troponin)

AND

- No other identifiable cause of the symptoms and findings

**Acute Pericarditis Case Definition**

Presence of ≥ 2 new or worsening of the following clinical features (Adler, et al 2015):

- Acute chest pain (Typically described as pain made worse by lying down, deep inspiration, or cough; and relieved by sitting up or leaning forward, although other types of chest pain may occur)
- Pericardial rub on examination
- New ST-elevation or PR-depression on EKG
- New or worsening pericardial effusion on echocardiogram or MRI

**Myopericarditis Case Definition**

Participants who meet criteria for both myocarditis and pericarditis may be described under myopericarditis.

An independent Cardiac Event Adjudication Committee (CEAC) consisting of pediatric and adult cardiologists will review suspected cases of myocarditis, pericarditis, or myopericarditis to determine if they meet CDC criteria of “probable” or “confirmed” event, and to assess severity and enable the DSMB to make recommendations to the Sponsor to continue vaccine dosing. The CEAC will operate under the rules of an approved charter that will be written and reviewed at the organizational meeting of the CEAC. Details regarding the CEAC composition, responsibilities, procedures, and frequency of data review will be defined in its charter.

**MIS-C Case Definition**

Investigators will also be asked to report, as an AESI, clinical signs/symptoms consistent with the CDC case definition of MIS-C (CDC 2020d):

- An individual aged < 21 years presenting with fever, laboratory evidence of inflammation, and evidence of clinically severe illness requiring hospitalization, with multisystem (> 2) organ involvement (cardiac, renal, respiratory, hematologic, gastrointestinal, dermatologic, or neurological)

AND

- No alternative plausible diagnoses

WITH OR WITHOUT

- Positive for current or recent SARS-CoV-2 infection by RT-PCR, serology (non-S protein-based), or antigen test or COVID-19 exposure within the 4 weeks prior to the onset of symptoms:
    - Fever ≥ 38.0°C/≥ 100.4ºF for ≥ 24 hours, or report of subjective fever lasting ≥ 24 hours
    - Including, but not limited to, one or more of the following: an elevated C-reactive protein, erythrocyte sedimentation rate, fibrinogen, procalcitonin, d-dimer, ferritin, lactic acid dehydrogenase, or interleukin 6; elevated neutrophils; reduced lymphocytes; or low albumin

Some participants may fulfill full or partial criteria for Kawasaki disease but it should be reported if they meet the case definition for MIS-C. Consider MIS-C in any pediatric death with evidence of SARS-CoV-2 infection.

##### 7.4.6. Recording and Follow-up of Pregnancy

Female participants who have a positive pregnancy test at Screening should not be enrolled; participants who have a positive pregnancy test any time during the study should receive no further dosing with IP but should be asked to remain in the study and be monitored for safety.

Details of all pregnancies in female participants will be collected after the start of study treatment and until the end of their participation in the study.

- If a pregnancy is reported, the investigator should inform the Sponsor within 24 hours of learning of the pregnancy and should follow the procedures outlined in this section.
- Abnormal pregnancy outcomes (eg, spontaneous abortion, fetal death, stillbirth, congenital anomalies, ectopic pregnancy) are considered SAEs.

Pregnancies occurring in participants after enrollment must be reported to the Sponsor or designee within 24 hours of the study site learning of its occurrence, using the SAE Mailbox, the SAE Hotline, or the SAE Fax line (Section 7.4.11). If the participant agrees to submit this information, the pregnancy must be followed to determine the outcome, including spontaneous or voluntary termination, details of the birth, and the presence or absence of any birth defects, congenital abnormalities, or maternal and/or newborn complications. This follow-up should occur even if the intended duration of the safety follow-up for the study has ended. Pregnancy report forms will be distributed to the study site to be used for this purpose. The investigator must immediately (within 24 hours of awareness) report to the Sponsor any pregnancy resulting in an abnormal outcome according to the procedures described for SAEs.

##### 7.4.7. Eliciting and Documenting Adverse Events

The investigator is responsible for ensuring that all AEs and SAEs are recorded in the eCRF and reported to the Sponsor.

Solicited ARs will be collected from Day 1 through 7 days after each dose (ie, the day of injection and 6 subsequent days). Other (unsolicited) AEs will be collected from Day 1 through 28 days after each dose.

Both MAAEs and SAEs will be collected from participants as specified in the SoA (Table 10) until the end of their participation in the study. Any AEs that occur before administration of IP will be analyzed separately from AEs that occur after vaccine administration.

At every study site visit or telephone contact, participants or parent(s)/LAR(s) will be asked a standard question to elicit any medically related changes in the participant’s well-being (including COVID-19 symptoms). Participants or parent(s)/LAR(s) will also be asked if the participant has been hospitalized, had any accidents, used any new medications, changed concomitant medication regimens (both prescription and over-the-counter medications), or had any nonstudy vaccinations.

In addition to participant observations, physical examination findings or data relevant to participant safety classified as an AE will be documented on the AE page of the eCRF.

After the initial AE/SAE report, the investigator is required to proactively follow each participant at subsequent visits/contacts. All AEs and SAEs will be treated as medically appropriate and followed until resolution, stabilization, the event is otherwise explained, or the participant is LTFU (as defined in Section 6.5).

##### 7.4.8. Assessment of Intensity

An event is defined as “serious” when it meets at least one of the predefined outcomes as described in the definition of an SAE (Section 7.4.2), NOT when it is rated as severe.

The severity (or intensity) of an AR or AE refers to the extent to which it affects the participant’s daily activities. The Toxicity Grading Scale for Healthy Adult and Adolescent Volunteers Enrolled in Preventative Vaccine Clinical Trials (DHHS 2007), modified for use in children 37 months to < 12 years of age (Table 6) and 6 to ≤ 36 months of age (Table 7), will be used to categorize local and systemic reactogenicity events (solicited ARs) and body temperature measurements observed during this study. Specific criteria for local and systemic reactogenicity events are presented in Section 7.4.3.

The determination of severity for all unsolicited AEs should be made by the investigator based upon medical judgment and the definitions of severity, as follows:

- Mild: These events do not interfere with the participant’s daily activities.
- Moderate: These events cause some interference with the participant’s daily activities and require limited or no medical intervention.
- Severe: These events prevent the participant’s daily activity and require intensive therapeutic intervention.

Study staff should elicit from the participant or the parent(s)/LAR(s) the impact of AEs on the participant’s activities of daily living to assess severity and document it appropriately in the participant’s source documentation. Changes in the severity of an AE should be documented in the participant’s source documentation to allow an assessment of the duration of the event at each level of intensity to be performed. An AE characterized as intermittent requires documentation of onset and the duration of each episode. An AE that fluctuates in severity during the course of the event is reported once in the eCRF at the highest severity observed.

##### 7.4.9. Assessment of Causality

The investigator’s assessment of an AE’s relationship to IP is part of the documentation process but is not a factor in determining what is or is not reported in the study.

The investigator will assess causality (ie, whether there is a reasonable possibility that the IP caused the event) for all AEs and SAEs. The relationship will be characterized using the following classifications:

**Not related:** There is not a reasonable possibility of a relationship to the IP. Participant did not receive the IP OR the temporal sequence of AE onset relative to administration of the IP is not reasonable OR the AE is more likely explained by a cause other than the IP.

**Related:** There is a reasonable possibility of a relationship to the IP. There is evidence of exposure to the IP. The temporal sequence of AE onset relative to the administration of the IP is reasonable. The AE is more likely explained by the IP than by another cause.

##### 7.4.10. Reporting Adverse Events

The investigator is responsible for reporting all AEs that are observed or reported at the times specified in Table 10, regardless of their relationship to IP or their clinical significance. If there is any doubt as to whether a clinical observation is an AE, the event should be reported.

All unsolicited AEs reported or observed during the study will be recorded on the AE page of the eCRF. Information to be collected includes type of event, time of onset, investigator-specified assessment of severity (impact on activities of daily living) and relationship to IP, time of resolution of the event, seriousness, any required treatments or evaluations, and outcome. The unsolicited AEs resulting from concurrent illnesses, reactions to concurrent illnesses, reactions to concurrent medications, or progression of disease states must also be reported. All AEs will be followed until they are resolved or stable or judged by the investigator to be not clinically significant, including ongoing SAEs after study completion. The Medical Dictionary for Regulatory Activities (MedDRA) will be used to code all unsolicited AEs.

Any medical condition that is present at the time that the participant is screened but does not deteriorate should not be reported as an unsolicited AE. However, if it deteriorates at any time during the study, it should be recorded as an unsolicited AE.

##### 7.4.11. Reporting SAEs

Prompt notification by the investigator to the Sponsor of an SAE is essential so that legal obligations and ethical responsibilities towards the safety of participants and the safety of a study intervention under clinical investigation are met.

Any AE considered serious by the investigator or that meets SAE criteria (Section 7.4.2) must be reported to the Sponsor immediately (within 24 hours of becoming aware of the SAE) via the EDC system. The investigator will assess whether there is a reasonable possibility that the IP caused the SAE. The Sponsor will be responsible for notifying the relevant regulatory authorities of any SAE as outlined in the 21 US CFR Parts 312 and 320. The investigator is responsible for notifying the IRB directly.

If the eCRF is unavailable at the time of the SAE, the following contact information is to be used for SAE reporting:

- SAE Mailbox:
- SAE Hotline (United States and Canada): +1-866-599-1341

- SAE Fax line (United States and Canada): +1-866-599-1342

Regulatory reporting requirements for SAEs are described in Section 7.4.15.

The investigator and any qualified designees are responsible for detecting, documenting, and recording events that meet the definition of an AE, including SAEs, and remain responsible for following up AEs that are serious, considered related to IP or study procedures, or that caused the participant to discontinue the study.

##### 7.4.12. Time Period and Frequency for Collecting AE, AESI, and SAE Information

Medical occurrences that begin before the start of IP dosing but after obtaining informed consent/assent will be recorded in the Medical History/Current Medical Conditions section of the eCRF and not in the AE section; however, if the condition worsens at any time during the study, it will be recorded and reported as an AE.

Adverse events may be collected as follows:

- Observing the participant
- Receiving an unsolicited complaint from the participant or participant's parent(s)/LAR(s)
- Questioning the participant or participant's parent(s)/LAR(s) in an unbiased and nonleading manner

Solicited ARs will be collected from the day of injection through 6 days after each dose. Other (unsolicited) AEs will be collected from the day of injection through 28 days after each dose.

Serious AEs (including AESIs) will be collected from the start of IP dosing until the last day of study participation.

All SAEs and AESIs will be recorded and reported to the Sponsor or designee immediately and in all circumstances within 24 hours of becoming aware of the event via the EDC system. If a site receives a report of a new SAE or AESI from a study participant or receives updated data on a previously reported SAE or AESI and the eCRF has been taken offline, then the site can report this information on a paper SAE or AESI form using the SAE Mailbox, the SAE Hotline, or the SAE Fax line (Section 7.4.11).

An abnormal value or result from a clinical or laboratory evaluation can also indicate an AE if it is determined by the investigator to be clinically significant (eg, leads to dose modification or study drug discontinuation, or meets any serious criteria). If this is the case, it must be recorded in the source document and as an AE on the appropriate AE form(s). The evaluation that produced the value or result should be repeated until that value or result returns to normal or is stabilized and the participant's safety is not at risk.

Investigators are not obligated to actively seek AEs or SAEs after end of the study participation. However, if the investigator learns of any SAE (including a death) at any time after a participant has withdrawn from or completed the study, and the investigator considers the event to be reasonably related to the IP or study participation, the investigator must promptly notify the Sponsor.

##### 7.4.13. Method of Detecting AEs and SAEs

Electronic diaries have specifically been designed for this study by the Sponsor. The diaries will include prelisted AEs (solicited ARs) and intensity scales; they will also include blank space for the recording of information on other AEs (unsolicited AEs) and concomitant medications/vaccinations.

The investigator is responsible for the documentation of AEs regardless of treatment group or suspected causal relationship to IP. For all AEs, the investigator must pursue and obtain information adequate to determine the outcome of the AE and to assess whether the AE meets the criteria for classification as an SAE requiring immediate notification to the Sponsor or its designated representative.

Care will be taken not to introduce bias when detecting AEs and/or SAEs. Open-ended and nonleading verbal questioning of the participant and participant's parent(s)/LAR(s) is the preferred method to inquire about the occurrence of AE.

##### 7.4.14. Follow-up of AEs and SAEs

After the initial AE/SAE report, the investigator is required to proactively follow each participant at subsequent visits and contacts.

All AEs and SAEs will be treated as medically appropriate and followed until resolution, stabilization, the event is otherwise explained, or the participant is LTFU, as defined in Section 6.5.

##### 7.4.15. Regulatory Reporting Requirements for SAEs

- Prompt notification by the investigator to the Sponsor of an SAE is essential so that legal obligations and ethical responsibilities towards the safety of participants and the safety of a study intervention under clinical investigation are met.
- The Sponsor has a legal responsibility to notify both the local regulatory authority and other regulatory agencies about the safety of a study intervention under clinical investigation. The Sponsor will comply with country-specific regulatory requirements relating to safety reporting to the regulatory authority, IRBs, and investigators.

- Investigator safety reports must be prepared for suspected unexpected serious adverse reactions according to local regulatory requirements and Sponsor policy and forwarded to investigators as necessary.
- An investigator who receives an investigator safety report describing an SAE or other specific safety information (eg, summary or listing of SAEs) from the Sponsor will review and then file it along with the IB and will notify the IRB, if appropriate according to local requirements.

#### 7.5. Safety Monitoring

The CRO's medical monitor, the Sponsor's medical monitor, and the individual study site investigators will monitor safety throughout the study.

##### 7.5.1. Internal Safety Team

An IST will review safety data throughout the study. For Part 1 dose-escalation and age de-escalation, based on the review of all available safety data through at least Day 8 (1 week after Dose 1 of mRNA-1273) for at least 75 participants at each dose level within the 6 to < 12 years and 2 to < 6 years age group, the IST will recommend whether dose-escalation and age de-escalation are appropriate. This process will then be repeated for all participants in the 25 and 50 µg dose levels within the lower age group of 6 months to < 2 years of age. An IST review of all available safety data through at least Day 36 (1 week after Dose 2 of mRNA-1273 at each dose level) of at least 75 participants will be required prior to the administration of the second injection of the next higher dose. In addition, the IST will escalate any safety concerns to the DSMB. The frequency of IST meetings will be described in more detail in the IST charter.

##### 7.5.2. Data Safety Monitoring Board

Safety oversight will be under the direction of a DSMB composed of external independent consultants with relevant expertise. Members of the DSMB will be independent from study conduct and free of conflict of interest. The DSMB will meet at pre-specified timepoints during the study to assess safety throughout study conduct. For the 6 to < 12 years age group, the DSMB will review cumulative safety data for approximately 300 participants enrolled at the selected dose level in Part 1 before enrollment begins in Part 2. For both the middle age group (2 to < 6 years) and the youngest age group (6 months to < 2 years) the DSMB will review cumulative safety data in both younger age groups (2 to < 6 years; 6 months to < 2 years) combined and at all dose levels administered in Part 1 before start of Part 2 (blinded phase) for each age group. The DSMB will operate under the rules of an approved charter that will be written and reviewed at the organizational meeting of the DSMB. Details regarding the DSMB composition, responsibilities, procedures, and frequency of data review will be defined in its charter.

If any of the study pause rules, described in Section 6.4, are met, the Sponsor will immediately suspend further enrollment and/or study dosing and the DSMB will convene on an ad hoc basis. The DSMB will review all available unblinded study data (as applicable) to help adjudicate any potential study pauses and make recommendations on further study conduct, including requesting additional information, recommending stopping the study, recommending changes to study conduct and/or the protocol, or recommending additional operational considerations due to safety issues that arise during the study.

The CEAC consisting of pediatric and adult cardiologists will review suspected cases of myocarditis, pericarditis, or myopericarditis to determine if they meet CDC criteria of "probable" or "confirmed" event, and to assess severity and enable the DSMB to make recommendations to the Sponsor to continue vaccine dosing. The CEAC will operate under the rules of an approved charter that will be written and reviewed at the organizational meeting of the CEAC. Details regarding the CEAC composition, responsibilities, procedures, and frequency of data review will be defined in its charter.

#### 7.6. Treatment of Overdose

As the study treatment is to be administered by a healthcare professional, it is unlikely that an overdose will occur. Dose deviations will be tracked as protocol deviations (Section 10.2.8).

#### 7.7. Pharmacokinetics

Pharmacokinetic parameters will not be evaluated in this study.

#### 7.8. Pharmacodynamics

Pharmacodynamic parameters will not be evaluated in this study.

#### 7.9. Biomarkers

Immunogenicity assessments are presented in Section 7.2. Biomarkers will not be evaluated in this study.

#### 7.10. Health Economics

Health economics will not be evaluated in this study.

### 8. STATISTICAL ANALYSIS PLAN

This section summarizes the planned statistical analysis strategy and procedures for the study. The details of statistical analysis will be provided in the statistical analysis plan (SAP), which will be finalized before the clinical database lock for the study. If changes are made to primary and/or key secondary objectives and hypotheses or the statistical methods related to those hypotheses after the study has begun but prior to any data unblinding, then the protocol will be amended (consistent with ICH Guideline E9). Changes to other secondary or exploratory analyses made after the protocol has been finalized, along with an explanation as to when and why they occurred, will be listed in the SAP or clinical study report (CSR) for the study. Ad hoc exploratory analyses, if any, will be clearly identified in the CSR.

#### 8.1. Blinding and Responsibility for Analyses

Part 1 of this study will be open-label, blinding procedures will not be applicable.

Part 2 of this study will be conducted in an observer-blind manner. The investigator, study staff, study participants, study site monitors, and Sponsor personnel (or its designees) will be blinded to the IP administered until study end, with the following exceptions:

- Unblinded pharmacy personnel (of limited number) will be assigned to vaccine accountability procedures and will prepare and administer mRNA-1273 (or placebo) to all participants. These pharmacy personnel will have no study functions other than study vaccine management, documentation, accountability, preparation, and administration. They will not be involved in participant evaluations and will not reveal the identity of IP to either the participant or the blinded study site personnel involved in the conduct of the study unless this information is necessary in the case of an emergency.
- Unblinded study site monitors, not involved in other aspects of monitoring, will be assigned as the IP accountability monitors. They will have responsibilities to ensure that study sites are following all proper IP accountability, preparation, and administration procedures.
- An unblinded statistical and programming team will perform the primary analyses in Part 2 (Section 8.6.2). Sponsor team members will be pre-specified to be unblinded to the primary analysis results and will not communicate the results of primary analysis to the blinded investigators, study site staff, clinical monitors, or participants.

The dosing assignment will be concealed by having the unblinded pharmacy personnel prepare the IP in a secure location that is not accessible or visible to other study staff. An opaque sleeve over the syringe used for injection will maintain the blind at the time of injection, as the doses containing mRNA-1273 will look different from those of placebo. Only delegated unblinded study site staff will conduct the injection procedure. Once the injection is completed, only the blinded study staff will perform further assessments and interact with the participants. Access to the randomization code will be strictly controlled at the pharmacy.

The planned study analyses are described in Section 8.6.

##### 8.1.1. Breaking the Blind

A participant's treatment assignment may be unblinded (Part 2 of the study) in the event of an SAE or other severe event, or if there is a medical emergency requiring the identity of the product to be known to properly treat a participant. If a participant becomes seriously ill or pregnant during the study, the blind will be broken if knowledge of the administered vaccine will affect that participant's dosing options. In the event of a medical emergency requiring identification of the vaccine administered to an individual participant, the investigator will make every attempt to contact the Sponsor medical lead to explain the need for opening the code within 24 hours of opening the code. The investigator will be responsible for documenting the time, date, reason for the code break, and the names of the personnel involved. If a COVID-19 vaccine is authorized or licensed for a specific age group before the end of the study, please refer to Section 3.3 for unblinding and/or cross-over plans.

In addition to the aforementioned situations where the blind may be broken, the data will also be unblinded to a statistical team at specified time points for interim analyses as outlined in Section 8.6.1.

#### 8.2. Statistical Hypothesis

If an accepted antibody threshold of protection against COVID-19 is established for the primary immunogenicity endpoint, the null hypothesis is that the percentage of participants on mRNA-1273 with serum antibody above the established threshold at Day 57 is ≤ 70% (ie, $H_0$: percentage of participants on mRNA-1273 with serum antibody at Day 57 above the established threshold ≤ 70%).

For each age group, the study will be considered to meet the immunogenicity endpoint if the 95% confidence interval (CI) of percentage of participants on mRNA-1273 rules out 70% (lower bound of the 95% CI is > 70%).

The null hypotheses may be updated when the information on an acceptable antibody threshold becomes available. In this case, the null hypothesis update will be provided in the SAP.

If an accepted serum antibody threshold of protection against COVID-19 is not available for the primary immunogenicity endpoint, immune response as measured by GM value and seroresponse rate in each age group based on Day 57 antibody levels will be compared to Day 57 antibody levels from young adults (18 to 25 years of age) in Study P301. Noninferiority tests of 2 null hypotheses based on 2 coprimary endpoints will be performed, respectively.

Coprimary endpoint 1: antibody GM value at Day 57

The null hypothesis $H^1_0$: immunogenicity response to mRNA-1273, as measured by antibody GM value at Day 57, is inferior in children (in age groups 6 months to < 2 years, 2 to < 6 years, and 6 to < 12 years) compared with that in young adults (18 to 25 years of age) using mRNA-1273 Study P301 data.

The noninferiority in antibody GM value in an age group in children compared with that in young adults (18 to 25 years of age) is demonstrated by meeting both success criteria:

- The lower bound of the 95% CI of the geometric mean ratio (GMR) ruling out 0.67 (lower bound > 0.67) using a noninferiority margin of 1.5, AND
- The GMR point estimate > 0.8 (minimum threshold).

The GMR is the ratio of the GM value of SARS-CoV-2-specific antibody in children in an age group receiving mRNA-1273 in this Study P204 compared with the GM value of young adults (18 to 25 years of age) receiving mRNA-1273 in Study P301 at Day 57.

Coprimary endpoint 2: antibody seroresponse rate at Day 57

A definition of seroresponse will be provided in the SAP based on forthcoming information about assay performance.

The null hypothesis $H^2_0$: immunogenicity response to mRNA-1273 as measured by seroresponse rate at Day 57 is inferior in children compared with that in young adults (18 to 25 years of age) in Study P301.

The noninferiority in seroresponse rate in an age group in children compared with that in young adults (18 to 25 years of age) is demonstrated by meeting both success criteria:

- The lower bound of the 95% CI of the seroresponse rate difference ruling out -10% (ie, lower bound > -10%) using the noninferiority margin of 10%, AND
- The seroresponse rate difference point estimate > -5% (minimum threshold)

The seroresponse rate difference is defined as the seroresponse rate in children receiving mRNA-1273 minus the seroresponse rate in young adults of 18 to 25 years of age receiving mRNA-1273 from Study P301.

The study would be considered to meet the primary immunogenicity endpoint in an age group if the noninferiority in the age group compared with the young adults (18 to 25 years of age) is demonstrated based on both coprimary endpoints.

#### 8.3. Power and Sample Size

The initial age groups in Part 1 are for estimation purposes. With approximately 750 participants (approximately 375 participants at each dose level of mRNA-1273) in the initial 6 to < 12 years age group, there is at least a 90% probability to observe at least 1 participant with an AE at a true AE rate of 1% for a given dose level. In each of the younger age groups (2 to < 6 years and 6 months to < 2 years), the safety assessment will occur during the conduct of Part 2 after approximately 375 participants have been exposed to mRNA-1273 at the dose level selected for Part 2. For further details, please refer to Section 7.5.2.

The sample size in the expansion (Part 2) is considered to be sufficient to support a safety database in the pediatric participants 6 months to < 12 years of age. With up to 3,000 participants each in the 6 to < 12 years, 2 to < 6 years, and 6 months to < 2 years of age groups exposed to mRNA-1273 at a given dose level in Part 2, the study has at least a 95% probability to observe at least 1 participant with an AE at a true 0.1% AE rate for a given dose level.

A subset of Full Analysis Set (FAS) participants (Immunogenicity Subset) in each age group will be selected for measuring immunogenicity data. The immunogenicity samples of the Immunogenicity Subset will be processed, and the analysis of primary immunogenicity endpoint will be based on the Immunogenicity PP Subset.

If a threshold of protection is available, for the primary immunogenicity endpoint, with approximately 289 participants on mRNA-1273 in the Immunogenicity PP Subset of an age group, there will be at least 90% power to rule out 70% with a 2-sided 95% CI (lower bound of the 95% CI > 70%) for the percentage of mRNA-1273 participants exceeding the accepted threshold if the true rate of participants exceeding the threshold is 80%.

If an acceptable antibody threshold of protection against COVID-19 is not available at the time of analysis for the primary immunogenicity endpoint, noninferiority tests of the 2 null hypotheses based on the 2 coprimary endpoints will be performed, respectively. The sample size calculation for each of the 2 noninferiority tests was performed, and the larger sample size was chosen for the study.

- With approximately 289 participants receiving mRNA-1273 in the Immunogenicity PP Subset of each age group in Study P204 and young adults (18 to 25 years of age) in Study P301, there will be 90% power to demonstrate noninferiority of the immune response, as measured by the antibody GM value, in pediatric population at a 2-sided alpha of 0.05, compared with that in young adults (18 to 25 years of age) in Study P301 receiving mRNA-1273, assuming an underlying GMR value of 1, a noninferiority margin of 0.67 (or 1.5), and a point estimate minimum threshold of 0.8. The standard deviation of the natural log-transformed levels is assumed to be 1.5.

- With approximately 289 participants receiving mRNA-1273 in the Immunogenicity PP Subset of each age group in Study P204 and young adults (18 to 25 years of age) in Study P301, there will be at least 90% power to demonstrate noninferiority of the immune response as measured by seroresponse rate in children at a 2-sided alpha of 0.05, compared with that in young adults 18 to 25 years of age in Study P301 receiving mRNA-1273, assuming seroresponse rate of 85% in young adults of 18 to 25 years of age from Study P301, true seroresponse rate of 85% in children (or true rate difference is 0 compared to young adults from Study P301), a noninferiority margin of 10% and a point estimate minimum threshold of -5% in seroresponse rate difference.

In the 1273-P203 interim analysis (data snapshot on 08 May 2021), the observed seroresponse rates at Day 57 were high in both 18 to 25 years of age group from Study P301 (98.6% with 95% CI: 96.6, 99.6) and 12 to17 years of age group (98.8% with 95% CI: 97.0, 99.7), based on pseudovirus nAb ID50 titers. For this Study P204, if the true seroresponse rates were assumed to be 95% or higher in both 18 to 25 years of age group from Study P301 and an age group in children, with between-group true difference within 4%, there will be a >90% power to demonstrate noninferiority by seroresponse rate in children compared with 18 to 25 years of age in Study P301, at a 2-sided alpha of 0.05.Assuming approximately 25% of participants in the Immunogenicity Subset will not meet the criteria to be included in the Immunogenicity PP Subset, approximately 528 participants in Part 2 (~396 receiving mRNA-1273 and ~132 receiving placebo) will be selected for the Immunogenicity Subset from which approximately 289 participants on mRNA-1273 will be suitable for the Immunogenicity PP Subset.

#### 8.4. Analysis Sets

The analysis sets are defined in Table 8. The analysis sets may be defined for Part 1 and Part 2 separately.

**Table 8: Analysis Sets**

| Analysis Set | Description |
|---|---|
| Randomization Set | Part 2: All participants who are randomly assigned, regardless of the participants' treatment status in the study. |
| Full Analysis Set (FAS) | Part 1: All enrolled participants in Part 1 who receive at least 1 injection of IP.<br>Part 2: All randomly assigned participants who receive at least 1 injection of IP. |
| Per-Protocol (PP) Set | All participants in the FAS who receive planned doses of IP per schedule, comply with the immunogenicity schedule, and have no major protocol deviations that impact key or critical data. |

| Analysis Set | Description |
|---|---|
| | Participants who are RT-PCR positive or seropositive at baseline will be excluded from the PP Set. The PP Set in Part 1 will be used for all analyses of immunogenicity in Part 1 unless specified otherwise. |
| Immunogenicity Subset | A subset of participants in the FAS will be selected for immunogenicity sampling and testing. |
| Per-Protocol Immunogenicity Subset | A subset of participants in the FAS will be selected for immunogenicity testing. The PP Immunogenicity Subset includes participants selected for the Immunogenicity Subset who receive planned doses of IP per schedule, comply with immunogenicity testing schedule, and have no major protocol deviations that impact key or critical data. Participants who are RT-PCR positive or seropositive at baseline will be excluded from the PP Immunogenicity Subset, in addition to participants with HIV who are receiving highly active anti-retroviral therapy (HAART). The PP Immunogenicity Subset will be used for all analyses of immunogenicity unless specified otherwise. |
| Safety Set | All enrolled participants (in Part 1) and all randomly assigned participants (in Part 2) who receive at least 1 dose of IP. The Safety Set will be used for all analyses of safety except for solicited ARs. |
| Solicited Safety Set | The Solicited Safety Set consists of participants in the Safety Set who contributed any solicited AR data.<br>The Solicited Safety Set will be used for the analyses of solicited ARs. |
| Modified Intent-to-Treat (mITT) Set | All participants in the FAS who have no serologic or virologic evidence of prior SARS-CoV-2 infection before the first dose of IP (both negative RT-PCR test for SARS-CoV-2 and negative serology test based on bAb specific to SARS-CoV-2 nucleocapsid) at baseline. |
| Modified Intent-to-Treat-1 (mITT1) Set | All participants in the mITT Set excluding those who received the wrong treatment (ie, at least 1 dose received that is not as randomized). |

Abbreviations: AR = adverse reaction; bAb = binding antibody; COVID-19 = coronavirus disease 2019; HIV = human immunodeficiency virus; IP = investigational product; RT-PCR = reverse transcriptase polymerase chain reaction; SARS-CoV-2 = severe acute respiratory syndrome coronavirus 2.

#### 8.5. Statistical Methods

##### 8.5.1. Baseline Characteristics and Demographics

Demographic variables (eg, age, race, ethnicity) and baseline characteristics (eg, length/height, weight, and BMI) will be summarized by treatment group. Summary statistics (mean, standard deviation for continuous variables, and number and percentage for categorical variables) will be provided.

##### 8.5.2. Safety Analyses

All safety analyses will be based on the Safety Set, except summaries of solicited ARs, which will be based on the Solicited Safety Set. All safety analyses will be provided by vaccination group (dose levels of mRNA-1273 and placebo) and by age group. Participants in Part 1 and Part 2 of the study and in different age groups who receive the same mRNA-1273 dose level may be combined for safety analysis.

Safety and reactogenicity will be assessed by clinical review of all relevant parameters including solicited ARs (local and systemic events), unsolicited AEs, SAEs, MAAEs, AESIs, AEs leading to discontinuation, and physical examination findings.

The number and percentage of participants with any solicited local AR, with any solicited systemic AR, and with any solicited AR during the 7-day follow-up period after each injection by toxicity grade will be provided. A 2-sided 95% exact CI using the Clopper-Pearson method will also be provided for the percentage of participants with any solicited AR.

The number and percentage of participants with unsolicited AEs, SAEs, MAAEs, severe AEs, and AEs leading to discontinuation from IP or withdrawal from the study will be summarized. Unsolicited AEs will be presented by MedDRA preferred term and system organ class.

The number of events of solicited ARs, unsolicited AEs/SAEs, and MAAEs will be reported in summary tables accordingly.

For all other safety parameters, descriptive summary statistics will be provided, and Table 9 summarizes the analysis strategy for safety parameters. Further details will be described in the SAP.

**Table 9: Analysis Strategy for Safety Parameters**

| Safety Endpoint | Number and Percentage of Participants, Number of Events | 95% CI |
|---|---|---|
| Any solicited AR (overall and by local, systemic) | X | X |
| Any unsolicited AE | X | — |
| Any SAE | X | — |
| Any unsolicited MAAE | X | — |
| Any unsolicited treatment-related AE | X | — |
| Any treatment-related SAE | X | — |
| Discontinuation due to AE | X | — |
| Any severe AE | X | — |
| Any treatment-related severe AE | X | — |

Abbreviations: AE = adverse event; AR = adverse reaction; CI = confidence interval; MAAE = medically attended adverse event; MedDRA = Medical Dictionary for Regulatory Activities; PT = preferred term; SAE = serious adverse event; SOC = system organ class.

Note: 95% CI using the Clopper-Pearson method. X = results will be provided. Solicited ARs and unsolicited AEs will be summarized by SOC and PT coded by MedDRA.

##### 8.5.3. Immunogenicity Analyses

The primary analysis population for immunogenicity will be the Immunogenicity PP Subset, unless specified otherwise. The primary objective of this study is to use the immunogenicity response to infer efficacy in participants aged 6 months to < 12 years at the dose level selected for expansion. For this objective, analyses of immunogenicity will be performed for each pediatric age group separately at the selected dose level based on the participants in the Immunogenicity PP Subset. For each pediatric age group, participants in Part 2 in the Immunogenicity PP Subset may be used for immunogenicity primary analysis. Participants from Part 1 and Part 2 who receive the same mRNA-1273 dose level selected for expansion may be combined for immunogenicity analyses. If the same dose level is selected for expansion for more than one age group, these age groups may be combined for immunogenicity analyses.

An accepted antibody threshold of protection against COVID-19 may be available based on data from other mRNA-1273 studies or external data. If such a threshold of protection against COVID-19 is available, the number and percentage of participants with antibody greater than or equal to the threshold at Day 57 will be provided with a 2-sided 95% CI using the Clopper-Pearson method. For an age group, if the lower bound of the 95% CI on the mRNA-1273 group is > 70%, the primary immunogenicity endpoint of this study will be considered to be met for that age group.

The number and percentage of participants with serum antibody greater than or equal to the threshold with 2-sided 95% CI will be provided at each postbaseline time point. The CI will be calculated using the Clopper-Pearson method.

If an accepted serum antibody threshold of protection against COVID-19 is not established, immune response as measured by GM value and seroresponse rate in each age group based on Day 57 antibody levels will be compared to that in young adults (18 to 25 years of age) using data from Study P301. An analysis of covariance model will be carried out with antibody at Day 57 as dependent variable and a group variable (a pediatric age group in Study P204 versus young adults [18 to 25 years of age] in Study P301) as the fixed variable for each pediatric age group. The GM values of the pediatric age group at Day 57 will be estimated by the geometric least square mean (GLSM) from the model. The GMR (ratio of GM values) will be estimated by the ratio of GLSM from the model. The corresponding 2-sided 95% CI will be provided to assess the difference in immune response between the pediatric age group (Study P204) compared to the young adults (18 to 25 years of age) in Study P301 at Day 57. For each pediatric age group, the noninferiority of GM value will be considered demonstrated if:

- The lower bound of the 95% CI of the GMR is > 0.67 based on the noninferiority margin of 1.5, AND
- The GMR point estimate > 0.8 (minimum threshold).

The number and percentage of participants with seroresponse due to vaccination will be provided with 2-sided 95% CI using the Clopper-Pearson method at each postbaseline time point with Day 57 being of the primary interest. The seroresponse rate difference with 95% CI at Day 57 will be provided between children receiving mRNA-1273 in Study P204 and young adults of 18 to 25 years of age receiving mRNA-1273 from Study P301. For each pediatric age group, the noninferiority of seroresponse rate will be considered demonstrated if:

- The lower bound of the 95% CI of the seroresponse rate difference is > -10% based on the noninferiority margin of 10%, AND
- The seroresponse rate difference point estimate > -5% (minimum threshold).

In addition, the GM value of anti-SARS-CoV-2-specific antibody with corresponding 95% CI will be provided at each time point. The 95% CIs will be calculated based on the t-distribution of the log-transformed values then back transformed to the original scale. For each age group, the geometric mean fold-rise of specific nAb and bAb with corresponding 95% CI at each postbaseline time point over preinjection baseline at Day 1 will be provided. Descriptive summary statistics including median, minimum, and maximum will also be provided.

**Multiplicity Adjustment Between Age Groups:**

A sequential hypothesis testing (fixed-sequence method) will be used to adjust multiplicity to preserve the family-wise Type I error rate (alpha = 0.05), starting with the oldest age group, followed by the middle age group, and then followed by the youngest age group. The immunogenicity coprimary endpoint hypotheses for the oldest age group (6 to < 12 years of age) will be tested first at alpha level of 0.05. If the testing in the oldest age group is statistically significant (meeting the noninferiority success criteria of the coprimary endpoints), the alpha level of 0.05 will be passed to the testing of the coprimary endpoint hypotheses in the middle age group (2 to < 6 years of age). If the testing in the middle age group is statistically significant at alpha level of 0.05, then the alpha 0.05 is preserved for testing the coprimary endpoint hypotheses in the youngest age group (6 months to < 2 years of age). The testing will continue through the sequence only until an endpoint in an age group is not statistically significant (did not meet noninferiority success criteria of any primary endpoint), in which case the testing will stop.

##### 8.5.4. Inferred Efficacy Analysis

To evaluate the incidence of COVID-19 after vaccination with mRNA-1273 or placebo, the incidence rate will be provided by vaccination group, dose level, and age group, calculated as the number of cases divided by the total person-time. Participants in Part 1 and Part 2 of the study and in different age groups who receive the same mRNA-1273 dose level (if applicable) may be combined in the analysis.

For serologically confirmed SARS-CoV-2 infection or COVID-19, regardless of symptomatology or severity, infection rate will be provided by vaccination group, dose level, and age group. The same analysis will be conducted for asymptomatic SARS-CoV-2 infection.

The secondary efficacy analyses will be performed on the PP Set, with sensitivity analyses in FAS, mITT Set, and mITT1 Set. Analyses of the efficacy endpoints in Part 2 will be performed for the randomized blinded phase. Additional exploratory analyses will be conducted in the blinded and unblinded phases for participants randomized to mRNA-1273 in Part 2, and in the unblinded phase for participants who are originally randomized to the placebo arm and cross-over to mRNA-1273 after use of any COVID-19 vaccine is authorized or licensed for the participant’s age group.

##### 8.5.5. Exploratory Analyses

Exploratory analyses will be described in the SAP before database lock.

##### 8.5.6. Subgroup Analyses

Subgroup analyses will be performed as described in the SAP.

#### 8.6. Study Analyses

##### 8.6.1. Interim Analyses

Part 1: Interim analyses may be performed after all or a subset of participants (with immunogenicity samples collected for D1 and D57) have completed Day 57 within an age group in Part 1 (one optional interim analysis for each age group, 3 interim analyses total for the 3 age groups in Part 1). Analyses of safety and immunogenicity may be conducted at each interim analysis.

Part 2: An interim analysis of immunogenicity and safety will be performed after all or a subset of participants have completed Day 57 (1 month after the second dose) in Part 1 or Part 2 within an age group. This interim analysis will be considered the primary analysis of immunogenicity for a given age group. Another interim analysis on safety may be performed after a different subset or all participants have completed Day 57 in an age group.

##### 8.6.2. Final Analysis

The final analysis of all endpoints will be performed after participants have completed all planned study procedures. Results of this analysis will be presented in a final CSR, including individual listings.

Additional information about all study analyses may be provided in the SAP.

#### 9. REFERENCES

Adler Y, Charron P, Imazio M, Badano L, Barón-Esquivias G, Bogaert J, et al. 2015 ESC guidelines for the diagnosis and management of pericardial diseases: the task force for the diagnosis and management of pericardial diseases of the European society of cardiology (ESC) endorsed by: The European Association for Cardio-Thoracic Surgery (EACTS). Eur Heart J. 2015 Nov 7;36(42):2921-64.

Anderson EJ, Rouphael NG, Widge AT, Jackson LA, Roberts PC, Makhene M, et al; mRNA-1273 Study Group. Safety and immunogenicity of SARS-CoV-2 mRNA-1273 vaccine in older adults. N Engl J Med. 2020;383(25):2427-38.

Aretz HT. Myocarditis: the Dallas criteria. Hum Pathol. 1987 Jun;18(6):619-24.

Baden LR, El Sahly HM, Essink B, Kotloff K, Frey S, Novak R, et al; COVE Study Group. Efficacy and safety of the mRNA-1273 SARS-CoV-2 vaccine. N Engl J Med. 2021;384(5):403-16.

Belhadjer Z, Méot M, Bajolle F, Khraiche D, Legendre A, Abakka S, et al. Acute heart failure in multisystem inflammatory syndrome in children in the context of global SARS-CoV-2 pandemic. Circulation. 2020;142(5):429-36.

Boehmer TK, DeVies J, Caruso E, van Santen KL, Tang S, Black CL, et al. Changing age distribution of the COVID-19 pandemic — United States, May–August 2020. MMWR Morb Mortal Wkly Rep. 2020;69(39):1404-9.

Centers for Disease Control and Prevention (CDC). COVID data tracker: United States COVID-19 cases and deaths by state [Internet]. 2020a [updated 2021 Jan 29; cited 2021 Jan 29]. Available from: https://covid.cdc.gov/covid-data-tracker/#cases.

Centers for Disease Control and Prevention (CDC). COVID data tracker: demographic trends of COVID-19 cases and deaths in the US reported to CDC [Internet]. 2020b [updated 2021 Jan 29; cited 2021 Jan 29]. Available from: https://covid.cdc.gov/covid-data-tracker/#demographics.

Centers for Disease Control and Prevention (CDC). COVID-19: symptoms of coronavirus [Internet]. 2020c [updated 2020 Dec 22; cited 2021 Jan 29]. Available from: https://www.cdc.gov/coronavirus/2019-ncov/symptoms-testing/symptoms.html.

Centers for Disease Control and Prevention (CDC). Multisystem inflammatory syndrome in children (MIS-C) associated with coronavirus disease 2019 (COVID-19) [Internet]. 2020d [updated 2020 Mar 27; cited 2021 Feb 16]. Available from: https://emergency.cdc.gov/han/2020/han00432.asp

Centers for Disease Control and Prevention (CDC). Coronavirus Disease 2019 Case Surveillance - United States, January 22 - May 30, 2020 [Internet]. 2020d [updated 2020 June 19; cited 2020

Jun 20]. Available from: https://www.cdc.gov/mmwr/volumes/69/wr/mm6924e2.htm?s_cid=mm6924e2_w

Chen Y, Lu S, Jia H, Deng Y, Zhou J, Huang B, et al. A novel neutralizing monoclonal antibody targeting the N-terminal domain of the MERS-CoV spike protein. Emerg Microbes Infect. 2017;6(5):e37.

Chen N, Zhou M, Dong X, Qu J, Gong F, Han Y, et al. Epidemiological and clinical characteristics of 99 cases of 2019 novel coronavirus pneumonia in Wuhan, China: a descriptive study. Lancet. 2020;395(10223):507-13.

Chin J, Magoffin RL, Shearer LA, Schieble JH, Lennette EH. Field evaluation of a respiratory syncytial virus vaccine and a trivalent parainfluenza virus vaccine in a pediatric population. Am J Epidemiol. 1969;89(4):449-63.

ClinicalTrials.gov [Internet]. Bethesda (MD): National Library of Medicine (US). Identifier NCT03076385: Safety, tolerability, and immunogenicity of VAL-506440 in healthy adult subjects. [cited 2020 Jun 01]. Available from: https://clinicaltrials.gov/ct2/keydates/NCT03076385.

ClinicalTrials.gov [Internet]. Bethesda (MD): National Library of Medicine (US). Identifier NCT03345043: Safety, tolerability, and immunogenicity of VAL-339851 in healthy adult subjects. [cited 2020 Jun 01]. Available from: https://clinicaltrials.gov/ct2/keydates/NCT03345043.

ClinicalTrials.gov [Internet]. Bethesda (MD): National Library of Medicine (US). Identifier NCT03382405: Safety, reactogenicity, and immunogenicity of cytomegalovirus vaccines mRNA-1647 and mRNA-1443 in healthy adults. [cited 2020 Jun 01]. Available from: https://clinicaltrials.gov/ct2/keydates/NCT03382405.

ClinicalTrials.gov [Internet]. Bethesda (MD): National Library of Medicine (US). Identifier NCT03392389: Safety, reactogenicity, and immunogenicity of mRNA-1653 in healthy adults. [cited 2020 Jun 01]. Available from: https://clinicaltrials.gov/ct2/keydates/NCT03392389.

ClinicalTrials.gov [Internet]. Bethesda (MD): National Library of Medicine (US). Identifier NCT04283461: Safety and immunogenicity study of 2019-nCoV vaccine (mRNA-1273) for prophylaxis of SARS-CoV-2 infection (COVID-19). [cited 2020 Jun 01]. Available from: https://clinicaltrials.gov/ct2/keydates/NCT04283461.

ClinicalTrials.gov [Internet]. Bethesda (MD): National Library of Medicine (US). Identifier NCT04405076: Dose-confirmation study to evaluate the safety, reactogenicity, and immunogenicity of mRNA-1273 COVID-19 vaccine in adults aged 18 years and older. [cited 2020 Aug 24]. Available from: https://clinicaltrials.gov/ct2/keydates/NCT04405076.

ClinicalTrials.gov [Internet]. Bethesda (MD): National Library of Medicine (US). Identifier NCT04470427: A study to evaluate efficacy, safety, and immunogenicity of mRNA-1273 vaccine in adults aged 18 years and older to prevent COVID-19; 2020 Jul 14. [cited 2020 Aug 24]. Available from: https://clinicaltrials.gov/ct2/keydates/NCT04470427.

Corbett KS, Edwards DK, Leist SR, Abiona OM, Boyoglu-Barnum S, Gillespie RA, et al. SARS-CoV-2 mRNA vaccine design enabled by prototype pathogen preparedness. Nature. 2020a;586(7830):567-71.

Corbett KS, Flynn B, Foulds KE, Francica JR, Boyoglu-Barnum S, Werner AP, et al. Evaluation of the mRNA-1273 vaccine against SARS-CoV-2 in nonhuman primates. N Engl J Med. 2020b;383(16):1544-55.

Corti D, Zhao J, Pedotti M, Simonelli L, Agnihothram S, Fett C, et al. Prophylactic and postexposure efficacy of a potent human monoclonal antibody against MERS coronavirus. Proc Natl Acad Sci U S A. 2015;112(33):10473-8.

Cruz AT, Zeichner SL. COVID-19 in children: initial characterization of the pediatric disease. Pediatrics. 2020;145(6):e20200834.

Department of Health and Human Services (DHHS), Food and Drug Administration, Center for Biologics Evaluation and Research (US). Guidance for industry: Toxicity grading scale for healthy adult and adolescent volunteers enrolled in preventative vaccine clinical trials. September 2007 [cited 2019 Apr 10] [10 screens]. Available from: https://www.fda.gov/downloads/BiologicsBloodVaccines/GuidanceComplianceRegulatoryInformation/Guidances/Vaccines/ucm091977.pdf.

Department of Health and Human Services (DHHS), Food and Drug Administration (US). Guidance for Industry, Investigators, and Institutional Review Boards. Guidance for industry, investigators, and institutional review boards: Conduct of clinical trials of medical products during the COVID-19 public health emergency. March 2020 [updated 2021 Jan 27; cited 2021 Jan 29] [38 screens]. Available from: https://www.fda.gov/media/136238/download.

Dufort EM, Koumans EH, Chow EJ, Rosenthal EM, Muse A, Rowlands J, et al; New York State and Centers for Disease Control and Prevention Multisystem Inflammatory Syndrome in Children Investigation Team. Multisystem inflammatory syndrome in children in New York State. N Engl J Med 2020;383(4):347-58.

Feldstein LR, Rose EB, Horwitz SM, Collins JP, Newhams MM, Son MB, et al; Overcoming COVID-19 Investigators; CDC COVID-19 Response Team. Multisystem inflammatory syndrome in U.S. children and adolescents. N Engl J Med 2020;383(4):334-46.

Ferreira VM, Schulz-Menger J, Holmvang G, Kramer CM, Carbone I, Sechtem U, et al. Cardiovascular magnetic resonance in nonischemic myocardial inflammation: expert recommendations. J Am Coll Cardiol. 2018 Dec 18;72(24):3158-76.

Fontanet A, Grant R, Tondeur L, Madec Y, Grzelak L, Cailleau I, et al. SARS-CoV-2 infection in primary schools in northern France: a retrospective cohort study in an area of high transmission. medRxiv. 2020;2020.06.25.20140178. Epub 2020 Jun 29.

Fulginiti VA, Eller JJ, Downie AW, Kempe CH. Altered reactivity to measles virus: atypical measles in children previously immunized with inactivated measles virus vaccines. JAMA. 1967;202(12):1075-80.

Gargano JW, Wallace M, Hadler SC, Langley G, Su JR, Oster ME, et al. Use of mRNA COVID-19 vaccine after reports of myocarditis among vaccine recipients: update from the advisory committee on immunization practices - United States, June 2021. MMWR Morb Mortal Wkly Rep. 2021 Jul 9;70(27):977-82.

Goldstein B, Giroir B, Randolph A; Members of the International Consensus Conference on Pediatric Sepsis. International pediatric sepsis consensus conference: definitions for sepsis and organ dysfunction in pediatrics. Pediatr Crit Care Med. 2005;6(1):2-8.

Graham BS. Rapid COVID-19 vaccine development. Science. 2020;368(6494):945-6.

Jackson LA, Anderson EJ, Rouphael NG, Roberts PC, Makhene M, Coler RN, et al; mRNA-1273 Study Group. An mRNA vaccine against SARS-CoV-2 — preliminary report. N Engl J Med. 2020;383(20):1920-31.

Johnson RF, Bagci U, Keith L, Tang X, Mollura DJ, Zeitlin L, et al. 3B11-N, a monoclonal antibody against MERS-CoV, reduces lung pathology in rhesus monkeys following intratracheal inoculation of MERS-CoV Jordan-n3/2012. Virology. 2016;490:49-58.

Ju B, Zhang Q, Ge J, Wang R, Sun J, Ge X, et al. Human neutralizing antibodies elicited by SARS-CoV-2 infection. Nature. 2020;584(7819)115-9.

Kim Y, Lee H, Park K, Park S, Lim JH, So MK, et al. Selection and characterization of monoclonal antibodies targeting Middle East respiratory syndrome coronavirus through a human synthetic Fab phage display library panning. Antibodies (Basel). 2019;8(3):42.

Kim L, Whitaker M, O’Halloran A, Kambhampati A, Chai SJ, Reingold A, et al. Hospitalization rates and characteristics of children aged <18 years hospitalized with laboratory-confirmed COVID-19 — COVID-NET, 14 states, March 1–July 25, 2020. MMWR Morb Mortal Wkly Rep 2020;69(32):1081-8.

LaRovere KL, Riggs BJ, Poussaint TY, Young CC, Newhams MM, Maamari M, et al. Neurologic involvement in children and adolescents hospitalized in the United States for COVID-19 or multisystem inflammatory syndrome. *JAMA Neurol*. 2021;e210504.

Leeb RT, Price S, Sliwa S, Kimball A, Szucs L, Caruso E, et al. COVID-19 trends among school-aged children — United States, March 1–September 19 2020. MMWR Morb Mortal Wkly Rep. 2020;69(39):1410-5.

Licciardi F, Giani T, Baldini L, Favalli EG, Caporali R, Cimaz R. COVID-19 and what pediatric rheumatologists should know: a review from a highly affected country. Pediatr Rheumatol Online J. 2020;18:35.

Link-Gelles R, DellaGrotta AL, Molina C, Clyne A, Campagna K, Lanzieri TM, et al. Limited secondary transmission of SARS-CoV-2 in child care programs — Rhode Island, June 1–July 31, 2020. MMWR Morb Mortal Wkly Rep. 2020;69(34):1170-2.

Paediatric Intensive Care Society (PICS). PICS statement: Increased number of reported cases of novel presentation of multi-system inflammatory disease [Internet]. 27 April 2020 [cited 2021 Feb 01] [2 screens] Available from: https://pccsociety.uk/wp-content/uploads/2020/08/PICS-statement-re-novel-KD-C19-presentation-v2-27042020.pdf

Robbiani DF, Gaebler C, Muecksch F, Lorenzi JC, Wang Z, Cho A, et al. Convergent antibody responses to SARS-CoV-2 in convalescent individuals. Nature. 2020;584(7821):437-42.

Rothan HA, Byrareddy SN. The epidemiology and pathogenesis of coronavirus disease (COVID-19) outbreak. J Autoimmun. 2020;109:102433.

Rüggeberg JU, Gold MS, Bayas JM, Blum MD, Bonhoeffer J, Friedlander S, et al. Anaphylaxis: case definition and guidelines for data collection, analysis, and presentation of immunization safety data. Vaccine. 2007;25(31):5675-84.

Schwartz NG, Moorman AC, Makaretz A, Chang KT, Chu VT, Szablewski CM, et al. Adolescent with COVID-19 as the source of an outbreak at a 3-week family gathering — four states, June–July 2020. MMWR Morb Mortal Wkly Rep. 2020;69(40):1457-9.

Shen KL, Yang YH, Jiang RM, Wang TY, Zhao DC, Jiang Y, Lu XX, et al. Updated diagnosis, treatment and prevention of COVID-19 in children: experts' consensus statement (condensed version of the second edition). World J Pediatr. 2020:16(3)232-9.

Szablewski CM, Chang KT, Brown MM, Chu VT, Yousaf AR, Anyalechi N, et al. SARS-CoV-2 transmission and infection among attendees of an overnight camp — Georgia, June 2020. MMWR Morb Mortal Wkly Rep 2020;69(31):1023-5.

Thomas SJ, Yoon IK. A review of Dengvaxia®: development to deployment. Hum Vaccin Immunother. 2019;15(10):2295-314.

Topjian AA, Raymond TT, Atkins D, Chan M, Duff JP, Joyner BL Jr, et al. Part 4: Pediatric basic and advanced life support: 2020 American Heart Association Guidelines for cardiopulmonary resuscitation and emergency cardiovascular care. Circulation. 2020;142(suppl 2):S469-S523.

Verdoni L, Mazza A, Gervasoni A, Martelli L, Ruggeri M, Ciuffreda M, et al. An outbreak of severe Kawasaki-like disease at the Italian epicentre of the SARS-CoV-2 epidemic: an observational cohort study. Lancet 2020;395(10239):1771-8.

Vogel TP, Top KA, Karatzios C, Hilmers DC, Tapia LI, Moceri P, et al. Multisystem inflammatory syndrome in children and adults (MIS-C/A): case definition and guidelines for data collection, analysis, and presentation of immunization safety data. Vaccine. Submitted 2020. Available from: https://brightoncollaboration.us/wp-content/uploads/2020/12/submitted-pdf-build.pdf

Wang L, Shi W, Joyce MG, Modjarrad K, Zhang Y, Leung K, et al. Evaluation of candidate vaccine approaches for MERS-CoV. Nat Commun. 2015;6:7712.

Wang L, Shi W, Chappell JD, Joyce MG, Zhang Y, Kanekiyo M, et al. Importance of neutralizing monoclonal antibodies targeting multiple antigenic sites on the Middle East respiratory syndrome coronavirus spike glycoprotein to avoid neutralization escape. J Virol. 2018;92(10):e02002-17.

Widge AT, Rouphael NG, Jackson LA, Anderson EJ, Roberts PC, Makhene M, et al. Durability of responses after SARS-CoV-2 mRNA-1273 vaccination. N Engl J Med. 2021;384(1):80-2.

Widjaja I, Wang C, van Haperen R, Gutiérrez-Álvarez J, van Dieren B, Okba NM, et al. Towards a solution to MERS: protective human monoclonal antibodies targeting different domains and functions of the MERS-coronavirus spike glycoprotein. Emerg Microbes Infect. 2019;8(1):516-30.

World Health Organization (WHO). Dengue vaccine: WHO position paper, September 2018 – recommendations. Vaccine. 2019;37(35):4848-9.

World Health Organization (WHO). Coronavirus disease 2019 (COVID-19): situation report – 157. 2020a June 25 (cited 2020 Jun 26). Available from: https://www.who.int/docs/default-source/coronaviruse/situation-reports/20200625-covid-19-sitrep-157.pdf?sfvrsn=423f4a82_2.

World Health Organization (WHO). WHO coronavirus disease (COVID-19) dashboard. 2020b Sep 28 (cited 2021 Jan 30). Retrieved from: https://covid19.who.int/.

Wrapp D, Wang N, Corbett KS, Goldsmith JA, Hseih CL, Abiona O, et al. Cryo-EM structure of the 2019-nCoV spike in the prefusion conformation. Science. 2020;367(6483):1260-3.

Yu X, Zhang S, Jiang L, Cui Y, Li D, Wang D, et al. Structural basis for the neutralization of MERS-CoV by a human monoclonal antibody MERS-27. Sci Rep. 2015;5:13133.

Zent O, Arras-Reiter C, Broeker M, Hennig R. Immediate allergic reactions after vaccinations – a post-marketing surveillance review. Eur J Pediatr. 2002;161:21-5.

### 10. SUPPORTING DOCUMENTATION AND OPERATIONAL CONSIDERATIONS

#### 10.1. APPENDIX 1: Schedule of Assessments

The SoA is presented in Table 10. The SoA for placebo recipient cross-over vaccination with mRNA-1273 if any COVID-19 vaccine is authorized or licensed for participant's age group is presented in Table 11.

If a participant cannot attend a study site visit (scheduled or unscheduled), with the exception of Screening, Day 1, and Day 29, a home visit is acceptable if performed by appropriately delegated study site staff or a home healthcare service provided by the Sponsor (Section 7). If neither a participant visit to the study site nor a home visit to the participant is possible (with the aforementioned exceptions), a safety telephone call should be performed that includes the assessments scheduled for the safety telephone calls.

The blood sampling schedule for exploratory serology and CMI in Part 2 of the study is presented in Table 12.

#### Table 10: Schedule of Assessments

| Visit Number | 0 | 1 | 2 | 3 | 3A | 4 | 4S | 5 | | | 6 | | | 7 |
|---|---|---|---|---|---|---|---|---|---|---|---|---|---|---|
| **Type of Visit** | C | C | TMV | C | C | TMV | C | C | SFU | | C | SFU | | C |
| **Month Time Point** | | M0 | | M1 | | | | M2 | eDiary | SC | M7 | eDiary | SC | M13 |
| **Study Visit Day** | D-28 to D-1 (Screening)[1] | D1 (Baseline) | D8 | D29[2] | D30 | D36[2] | D43[2,3] | D57[2, 4] | Every 4 weeks D71 – D183[2, 5] | Every 4 weeks D85– D197[2, 6] | D209[2, 4] | Every 4 weeks D223– D363[2, 5] | Every 4 weeks D237– D377[2, 6] | D394[2, 4] |
| **Window Allowance (Days)** | - | - | + 3 | + 7 | +3 | + 3 | ± 2 | + 7 | ± 3 | ± 3 | ± 14 | ± 3 | ± 3 | ± 14 |
| **Days Since Most Recent Injection** | - | 0 | 7 | 28 | 1 | 7 | 14 | 28 | - | - | 180 | - | - | 365 |
| Informed consent/assent form, demographics, concomitant medications, medical history | X | | | | | | | | | | | | | |
| Review of inclusion and exclusion criteria | X | X | | | | | | | | | | | | |
| Physical examination including body temperature, length/height, weight, and BMI[7] | X | X | | X | | | X | X | | | X | | | X |
| Pregnancy test[8] | X | X | | X | | | | | | | | | | |
| Randomization | | X | | | | | | | | | | | | |
| Study injection (including 30-minute postdose observation period) | | X | | X | | | | | | | | | | |
| Blood sample for vaccine immunogenicity (Part 1)[9] | | X | | | | | | X[19] | | | X[19] | | | X[19] |
| Blood sample for vaccine immunogenicity (Part 2)[10] | | X | | X | | | | X | | | X | | | X |
| Blood sample for exploratory serology and cell-mediated immunity (Part 2)[3,10] | | X | | | | | X | | | | X | | | X |
| Blood sample for potential biomarker analysis (Part 2)[10,11] | | | | | X[11] | | | | | | | | | |
| Nasal swab sample for SARS-CoV-2[12] | | X | | X | | | X | X | | | X | | | X |

| **Visit Number** | 0 | 1 | 2 | 3 | 3A | 4 | 4S | 5 | | | 6 | | | 7 |
|---|---|---|---|---|---|---|---|---|---|---|---|---|---|---|
| **Type of Visit** | C | C | TMV | C | C | TMV | C | C | SFU | | C | SFU | | C |
| **Month Time Point** | | M0 | | M1 | | | | M2 | eDiary | SC | M7 | eDiary | SC | M13 |
| **Study Visit Day** | D-28 to D-1 (Screening)[1] | D1 (Baseline) | D8 | D29[2] | D30 | D36[2] | D43[2,3] | D57[2, 4] | Every 4 weeks D71 – D183[2, 5] | Every 4 weeks D85– D197[2, 6] | D209[2, 4] | Every 4 weeks D223– D363[2, 5] | Every 4 weeks D237– D377[2, 6] | D394[2, 4] |
| **Window Allowance (Days)** | - | - | + 3 | + 7 | +3 | + 3 | ± 2 | + 7 | ± 3 | ± 3 | ± 14 | ± 3 | ± 3 | ± 14 |
| **Days Since Most Recent Injection** | - | 0 | 7 | 28 | 1 | 7 | 14 | 28 | - | - | 180 | - | - | 365 |
| Surveillance for COVID-19/illness visit/ unscheduled visit[13] | | | X | X | | X | X | X | X | X | X | X | X | X |
| Convalescent visit[14] | | | | | | X | X | X | X | X | X | X | X | X |
| eDiary activation for recording solicited ARs (7 days)[15] | | X | | X | | | | | | | | | | |
| Review of eDiary data | | | X | | | X | | | | | | | | |
| Follow-up safety telephone calls[16] | | | | | | | | | | X | | | X | |
| Recording of unsolicited AEs | | X | X | X | X | X | X | X | | | | | | |
| Recording of MAAEs and concomitant medications relevant to or for the treatment of the MAAE[17] | | X | X | X | X | X | X | X | X | | X | X | | X |
| Recording of SAEs and concomitant medications relevant to or for the treatment of the SAE[17,18] | | X | X | X | X | X | X | X | X | | X | X | | X |
| Recording of AESIs (eg, MIS-C, myocarditis/pericarditis)[18] | | X | X | X | X | X | X | X | X | | X | X | | X |
| Recording of concomitant medications and nonstudy vaccinations[17] | | X | X | X | X | X | X | X | | | | | | |
| Study completion | | | | | | | | | | | | | | X |

Abbreviations: AE = adverse event; AESI = adverse event of special interest; AR = adverse reaction; BMI = body mass index; C = clinic visit; COVID-19 = coronavirus disease 2019; D = day; eCRF = electronic case report form; EDC = electronic data capture; eDiary = electronic diary; FDA = Food and Drug Administration; IP = investigational product; IRB = institutional review board; LAR = legally acceptable representative; M = month; MAAE = medically attended adverse event; MIS-C = multisystem inflammatory syndrome in children; RT-PCR = reverse transcriptase polymerase chain reaction; SAE = serious adverse event; SARS-CoV-2 = severe acute respiratory syndrome coronavirus 2; SC = safety (telephone) call; SFU = safety follow-up; TMV = telemedicine visit; VTEU = Vaccine and Treatment Evaluation Units.

**Note:** In accordance with FDA Guidance on Conduct of Clinical Trials of Medical Products during COVID-19 Public Health Emergency (FDA March 2020), investigators may convert study site visits to home visits or telemedicine visits (with the exception of Screening, Day 1, and Day 29) with the approval of the Sponsor.

1. Screening Visit and Day 1 may be combined on the same day. Additionally, the Screening Visit may be performed over multiple visits if performed within the 28-day screening window.
2. If the visit for the second dose (Day 29) is disrupted and cannot be completed at Day 29 (+7 days) as a result of the COVID-19 pandemic (self-quarantine or disruption of study site activities following business continuity plans and/or local government mandates for "stay at home" or "shelter in place"), the visit window may be extended to Day 29 + 21 days. When the extended visit window is used, the remaining study visits should be rescheduled to follow the intervisit interval from the actual date of the second dose. Refer to Section 6.1.1 for individual participant criteria for delay of study vaccination.
3. To be conducted during Part 2 of the study in a cohort of participants at selected VTEU sites only.
4. All scheduled study visits should be completed within the respective visit windows. If the participant is not able to attend a study site visit as a result of the COVID-19 pandemic (self-quarantine or disruption of study site activities following business continuity plans and/or local government mandates for "stay at home" or "shelter in place"), a telemedicine visit should be conducted in place of the study site visit. The telemedicine visit should encompass all scheduled visit assessments that can be completed remotely, such as assessment for AEs and concomitant medications (eg, as defined in scheduled safety telephone calls). Home visits will be permitted for all nondosing visits, with the exception of Screening, if a participant cannot visit the study site as a result of the COVID-19 pandemic. Home visits must be permitted by the study site IRB and the participant's parent(s)/LAR(s) via informed consent and have prior approval from the Sponsor (or its designee).
5. Safety follow-up via an eDiary questionnaire will be performed every 4 weeks from Day 71 to Day 183 and again from Day 223 to Day 363.
6. Safety follow-up via a safety telephone call will be performed every 4 weeks from Day 85 to Day 197 and again from Day 237 to Day 377.
7. A full physical examination, including body temperature (oral [for participants > 4 years of age] or tympanic [for participants ≤ 4 years of age]), length/height, and weight, will be performed at the Screening Visit or on Day 1. Symptom-directed physical examination will be performed on Day 1, Day 29, Day 43 (if the visit is applicable), Day 57, Day 209, and Day 394 and may be performed at other time points at the discretion of the investigator. Body mass index will be calculated only at Screening Visit. Body temperature (oral/tympanic) should be measured on each injection day prior to injection. On each injection day before injection and again 7 days after injection, the injection site should be examined. Any clinically significant finding identified during a study visit should be reported as an MAAE. Participants who are febrile (body temperature ≥ 38.0°C/≥ 100.4°F) before injection on Day 1 or Day 29 must have the visit rescheduled within the relevant visit window to receive the injection. Afebrile participants with minor illnesses may be injected at the discretion of the investigator.
8. Pregnancy test at Screening and Day 1 and before the second study injection will be a point-of-care urine test and will be performed if deemed appropriate by the investigator. At the discretion of the investigator, a pregnancy test either via blood or point-of-care urine test can be performed at any time during the study.
9. On Day 1, sample must be collected prior to randomization and dosing. If a Day 1 (baseline) blood sample cannot be obtained in Part 1, the participant will be considered either a screen failure due to inability to satisfy inclusion criterion 3 or could be scheduled for rescreening. Participants may be rescreened 1 time, either within the same screening period for Part 1 or for a new screening period for Part 2 later in the study. If the participant is rescreened and a blood sample still cannot be obtained, then he/she will be considered a screen failure due to inability to satisfy inclusion criterion 3.
10. On Day 1, sample must be collected prior to randomization and dosing. On Day 29, sample must be collected prior to dosing. In Part 2, participants in each age group will be assigned to 1 of 5 phlebotomy cohorts (Table 12). Participants in 3 of these 5 cohorts will provide blood specimens for immunogenicity at the following time points: Day 1, Day 57, and one of Day 29, Day 209, or Day 394 (such that each participant provides a total of 3 blood samples only). Participants in the Cohort D (remainder of the age group) will provide a blood sample at Day 1 (prior to randomization and the first dose) and within 4 days of receiving Dose 2 at Day 30 (+3 days) for storage and potential future biomarker testing. A fifth cohort of participants (selected VTEU sites only) will provide blood samples for assessment of exploratory serology and CMI (S protein-specific T-cell responses) on Day 1, Day 43, Day 209, and D394. Table 12 provides the blood sampling schedule in Part 2 of the study. If a Day 1 (baseline) blood sample cannot be obtained in either Part 1 or Part 2, the participant will be considered either a screen failure due to inability to satisfy inclusion criterion 3 or could be rescheduled for rescreening. Participants may be rescreened 1 time, either within the same screening period for Part 1 or Part 2 or in a new screening period of Part 2 if the initial screening was in Part 1. If the participant is rescreened and a blood sample still cannot be obtained, then he/she will be considered a screen failure due to inability to satisfy inclusion criterion 3.
11. Part 2, Cohort D participants only, one ~4 mL blood draw. For participants already enrolled in Cohort D prior to protocol amendment 4, this blood draw will be optional and confirmed at time of re-consenting; for all participants newly enrolled into Cohort D under amendment 4, it is mandatory.
12. The nasal swab sample (collected before dosing on injection day) will be used to ascertain the presence of SARS-CoV-2 via RT-PCR.

13. An unscheduled visit may be prompted by reactogenicity issues, illness visit criteria for COVID-19, or new or ongoing AEs. If a participant meets the pre-specified criteria of suspicion for COVID-19 (Section 7.1.5), the participant will be asked to return within 72 hours or as soon as possible to the study site for an unscheduled illness visit to include a nasal swab sample (for RT-PCR testing to evaluate for the presence of SARS-CoV-2 infection) and other clinical evaluations. If a study site visit is not possible, a home visit may be arranged to collect the nasal swab sample and conduct clinical evaluations. The study site may collect an additional nasal swab sample for SARS-CoV-2 testing to be able to render appropriate medical care for the study participant as determined by local standards of care. Additionally, clinical information will be carefully collected to evaluate the severity of the clinical case.

14. A convalescent visit will be scheduled approximately 28 days (+7 days) after diagnosis of COVID-19. At this visit, a nasal swab sample for viral RT-PCR and a blood sample for potential immunologic assessment of SARS-CoV-2 infection will be collected.

15. At each injection visit, participants' parent(s)/LAR(s) will record data into the eDiary starting approximately 30 minutes after injection under the supervision of the study site staff to ensure successful entry of assessment. Participants' parent(s)/LAR(s) will continue to record entries in the eDiary after they leave the study site, preferably in the evening and at the same time each day, on the day of injection and for 6 days following injection. If a solicited local or systemic AR continues beyond Day 7 after vaccination, the participant's parent(s)/LAR(s) will be prompted to capture details of the solicited local or systemic AR in the eDiary until the AR is resolved or the next IP injection occurs, whichever occurs first. Capturing details of ARs in the eDiary should not exceed 28 days after each vaccination. Adverse reactions recorded in eDiaries beyond Day 7 should be reviewed by the study site staff either during the next scheduled telephone call or at the next study site visit.

16. Trained study site personnel will call all participants to collect information relating to any MAAEs, SAEs, AEs leading to study withdrawal and information on concomitant medications associated with those events and any nonstudy vaccinations. In addition, study personnel will collect information on known participant exposure to someone with known COVID-19 or SARS-CoV-2 infection and on any COVID-19 symptoms the participant may experience.

17. All concomitant medications and nonstudy vaccinations will be recorded through 28 days after each injection; all concomitant medications relevant to or for the treatment of an SAE or MAAE will be recorded from Day 1 through the final visit (Day 394).

18. In addition to MIS-C and myocarditis and/or pericarditis, a list of AESIs pertinent to this study may be referenced in the list of AESIs that is maintained by the Sponsor and provided to each investigator. All SAEs and AESIs will be reported to the Sponsor or designee immediately and in all circumstances within 24 hours of becoming aware of the event via the EDC system. If a site receives a report of a new SAE or AESI from a study participant or receives updated data on a previously reported SAE or AESI and the eCRF has been taken offline, then the site can report this information on a paper SAE or AESI form using the SAE Mailbox, the SAE Hotline, or the SAE Fax line (Section 7.4.11).

19. For Part 1, only the first approximately 75 participants in Arm 1 and Arm 2 will have postbaseline scheduled blood draws; the 300 participants in each of the expansion part of Arm 1 and Arm 2 may have an optional blood draw on Day 57. All participants in Arm 3, 4, 5, 6 and 7 will have postbaseline blood draws on Day 57, Day 209 and Day 394.

**Table 11: Schedule of Assessments for Placebo Recipient Cross-Over Vaccination with mRNA-1273 if any COVID-19 Vaccine is Authorized or Licensed for Participant's Age Group[1]**

| Schedule of Assessments for Placebo Recipient Cross-Over Vaccination | Cross-Over D1 | D29 (+ 7)[2] | D36 (+ 3) | D57 (+ 7) | Remainder of Study Visits |
|---|---|---|---|---|---|
| Study injection (including 30-minute postdose observation period) | X | X | | | |
| Safety follow-up call | | | X | X | |
| Recording of unsolicited AEs | X | X | X | X | |
| Recording of MAAEs and concomitant medications relevant to or for the treatment of the MAAE | X | X | X | X | X |
| Recording of SAEs and concomitant medications relevant to or for the treatment of the SAE | X | X | X | X | X |
| Recording of AESIs (eg, MIS-C and myocarditis and/or pericarditis) | X | X | X | X | X |

Abbreviations: AE = adverse event; AESI = adverse event of special interest; D = day; MAAE = medically attended adverse event; MIS-C = multisystem inflammatory syndrome in children; SAE = serious adverse event; SARS-CoV-2 = severe acute respiratory syndrome coronavirus 2.

1. Authorized or licensed in participant's age group.
2. Refer to Section 6.1.1 for individual participant criteria for delay of study vaccination.

**Table 12: Phlebotomy Schedule for Serology, Biomarker Sample, and Cell-Mediated Immunity for Part 2 (Expansion) of the Study**

| Cohort | Number of subjects | Study Visit Day | | | | | | |
|---|---|---|---|---|---|---|---|---|
| | | D1[1,2] | D29[1] | D30 (+3)[3] | D43 | D57 | D209 | D394 |
| **Phlebotomy Schedule for Serology: To be Executed Within Age Group** | | | | | | | | |
| A | First 176 (132 mRNA-1273: 44 placebo) | X | X | | | X | | |
| B | Next 176 (132 mRNA-1273: 44 placebo) | X | | | | X | X | |
| C | Next 176 (132 mRNA-1273: 44 placebo) | X | | | | X | | X |
| D | Remainder of the age group | X | | X[3] | | | | |
| **Phlebotomy Schedule for Cell-Mediated Immunity: To be Executed Within Each Age Group at Selected VTEU Sites only** | | | | | | | | |
| E (CMI with exploratory serology) | 24 (18 mRNA-1273: 6 placebo) | X | | | X | | X | X |

Abbreviations: CMI = cell-mediated immunity; D = day; VTEU = Vaccine and Treatment Evaluation Units.

1. On Day 1, sample must be collected prior to randomization and dosing. On Day 29, sample must be collected prior to dosing.
2. If a Day 1 (baseline) blood sample cannot be obtained in either Part 1 or Part 2, the participant will be considered either a screen failure due to inability to satisfy inclusion criterion 3 or could be scheduled for rescreening. Participants may be rescreened 1 time, either within same screening period for Part 1 or Part 2 or in a new screening period for Part 2 if the initial screening was in Part 1. If the participant is rescreened and a blood sample still cannot be obtained, then he/she will be considered a screen failure due to inability to satisfy inclusion criterion 3.
3. Serum sample from ~4 ml of blood only, to be stored for potential future use for biomarker assessment.

#### 10.2. APPENDIX 2: Study Governance Considerations

##### 10.2.1. Regulatory and Ethical Considerations

This study will be conducted in accordance with the protocol and with the following:

Consensus ethical principles derived from international guidelines, including the Declaration of Helsinki and Council for International Organizations of Medical Sciences International Ethical Guidelines.

Applicable ICH GCP guidelines.

Applicable laws and regulatory requirements.

The protocol, protocol amendments, ICF, IB, and other relevant documents (eg, advertisements) must be submitted to an IRB by the investigator and reviewed and approved by the IRB before the study is initiated.

Any amendments to the protocol will require IRB approval before implementation of changes made to the study design, except for changes necessary to eliminate an immediate hazard to study participants.

The investigator will be responsible for the following:

Providing written summaries of the status of the study to the IRB annually or more frequently in accordance with the requirements, policies, and procedures established by the IRB

Notifying the IRB of SAEs or other significant safety findings as required by IRB procedures

Providing oversight of the conduct of the study at the site and adherence to requirements of 21 CFR, ICH guidelines, the IRB, European regulation 536/2014 for clinical studies (if applicable), and all other applicable local regulations.

##### 10.2.2. Study Monitoring

Before an investigational site can enter a participant into the study, a representative of the Sponsor or its representatives will visit the investigational study site to do the following:

Determine the adequacy of the facilities.

Discuss with the investigator(s) and other personnel their responsibilities with regard to protocol adherence, and the responsibilities of the Sponsor or its representatives. This will be documented in a clinical study agreement between the Sponsor, the designated CRO, and the investigator.

According to ICH GCP guidelines, the Sponsor of the study is responsible for ensuring the proper conduct of the study with regard to protocol adherence and validity of data recorded on the eCRFs.

The study monitor's duties are to aid the investigator and the Sponsor in the maintenance of complete, accurate, legible, well-organized, and easily retrievable data. The study monitor will advise the investigator of the regulatory necessity for study-related monitoring, audits, IRB review, and inspection by providing direct access to the source data/documents. In addition, the study monitor will explain to and interpret for the investigator all regulations applicable to the clinical evaluation of an IP as documented in ICH guidelines.

It is the study monitor's responsibility to inspect the eCRFs and source documentation throughout the study to protect the rights of the participants; to verify adherence to the protocol; to verify the completeness, accuracy, and consistency of the data; and to confirm adherence of study conduct to any local regulations. Details will be outlined in the Clinical Monitoring Plan. During the study, a monitor from the Sponsor or a representative will have regular contacts with the investigational site, for the following purposes:

Provide information and support to the investigator(s).

Confirm that facilities remain acceptable.

Confirm that the investigational team is adhering to the protocol, that the data are being accurately recorded in the eCRFs, and that IP accountability checks are being performed.

Perform source data verification. This includes a comparison of the data in the eCRFs with the participant's medical records at the hospital or practice and other records relevant to the study. This will require direct access to all original records for each participant (eg, clinical charts or electronic medical record system).

Record and report any protocol deviations not previously sent.

Confirm that AEs and SAEs have been properly documented on eCRFs, that any SAEs have been forwarded to the SAE Hotline, and that those SAEs that meet criteria for reporting have been forwarded to the IRB.

The monitor will be available between visits if the investigator(s) or other staff need information or advice.

##### 10.2.3. Audits and Inspections

The Sponsor, their designee(s), the IRB, or regulatory authorities will be allowed to conduct study site visits to the investigational facilities for the purpose of monitoring or inspecting any aspect of the study. The investigator agrees to allow the Sponsor, their designee(s), the IRB, or regulatory authorities to inspect the IP storage area, IP stocks, IP records, participant charts and study source documents, and other records relative to study conduct.

Authorized representatives of the Sponsor, a regulatory authority, and any IRB may visit the study site to perform audits or inspections, including source data verification. The purpose of a Sponsor audit or inspection is to systematically and independently examine all study-related activities and documents to determine whether these activities were conducted and data were recorded, analyzed, and accurately reported according to the protocol, ICH E6(R2) GCP, and any applicable regulatory requirements. The investigator should contact the Sponsor immediately if contacted by a regulatory agency about an inspection.

The principal investigator must obtain IRB approval for the investigation. Initial IRB approval, and all materials approved by the IRB for this study, including the informed consent/assent forms and recruitment materials, must be maintained by the investigator and made available for inspection.

##### 10.2.4. Financial Disclosure

The investigator is required to provide financial disclosure information to allow the Sponsor to submit the complete and accurate certification or disclosure statements required under 21 CFR 54. In addition, the investigator must provide the Sponsor with a commitment to promptly update this information if any relevant changes occur during the course of the investigation and for 1 year following the completion of the study.

The Sponsor, the CRO, and the study site are not financially responsible for further testing or treatment of any medical condition that may be detected during the screening process. In addition, in the absence of specific arrangements, the Sponsor, the CRO, and the study site are not financially responsible for further treatment of the disease under study.

##### 10.2.5. Recruitment Procedures

Advertisements to be used for the recruitment of study participants and any other written information regarding this study to be provided to the participant's parent(s)/LAR(s) should be submitted to the Sponsor for approval. All documents must be approved by the IRB.

##### 10.2.6. Informed Consent/Assent Process

The informed consent and assent document(s) must meet the requirements of 21 CFR 50, local regulations, ICH guidelines, the Health Insurance Portability and Accountability Act, where applicable, and the IRB or study site. All consent/assent documents will be approved by the appropriate IRB. The actual ICF used at each study site may differ, depending on local regulations and IRB requirements. However, all versions of the ICF must contain the standard information found in the sample ICF provided by the Sponsor. Any change to the content of the ICF must be approved by the Sponsor and the IRB prior to the ICF being used.

If new information becomes available that may be relevant to the participant's willingness to continue participation in the study, this will be communicated to them in a timely manner. Such information will be provided via a revised ICF or an addendum to the original ICF.

The investigator or his/her representative will explain the nature of the study to the participant's parent(s)/LAR(s) and answer all questions regarding the study.

The investigator is responsible for ensuring that the participant's parent(s)/LAR(s) fully understands the nature and purpose of the study. Information should be given in both oral and written form whenever possible.

No participant should be obliged to participate in the study. The participant's parent(s)/LAR(s) must be informed that participation is voluntary. The participant's relatives, guardians, or (if applicable) LARs must be given ample opportunity to inquire about details of the study. The information must make clear that refusal to participate in the study or withdrawal from the study at any stage is without any prejudice to the participant's subsequent care.

The participant's parent(s)/LAR(s) must be allowed sufficient time to decide whether they wish to let their child participate in the study.

The participant's parent(s)/LAR(s) must be made aware of, and give consent to, direct access to participant's source medical records by study monitors, auditors, the IRB, and regulatory authorities. The participant's parent(s)/LAR(s) should be informed that such access will not violate participant confidentiality or any applicable regulations. The participant's parent(s)/LAR(s) should also be informed that he/she is authorizing such access by signing the ICF.

A copy of the ICF(s) must be provided to the participants' parent(s)/LAR(s).

Parent(s)/LAR(s) of a participant who is rescreened (allowed once) are not required to sign another ICF if the rescreening occurs within 28 days from the previous ICF signature date (within the initial Screening Period).

The ICF will also explain that excess serum from immunogenicity testing may be used for future research, which may be performed at the discretion of the Sponsor to further characterize the immune response to SARS-CoV-2, additional assay development, and the immune response across CoVs.

##### 10.2.7. Protocol Amendments

No change or amendment to this protocol may be made by the investigator or the Sponsor after the protocol has been agreed to and signed by all parties unless such change(s) or amendment(s) has (have) been agreed upon by the investigator or the Sponsor. Any change agreed upon will be recorded in writing, and the written amendment will be signed by the investigator and the Sponsor.

Approval of the IRB is required prior to the implementation of an amendment, unless overriding safety reasons warrant immediate action, in which case the IRB(s) will be promptly notified.

Any modifications to the protocol or the ICF that may impact the conduct of the study or potential benefit of the study or may affect participant safety, including changes of study objectives, study design, participant population, sample sizes, study procedures, or significant administrative aspects, will require a formal amendment to the protocol. Such an amendment will be released by the Sponsor, agreed to by the investigator(s), and approved by the relevant IRB(s) prior to implementation. A signed and dated statement that the protocol, any subsequent relevant amended documents, and the ICF have been approved by relevant IRB(s) must be provided to the Sponsor before the study is initiated.

Administrative changes of the protocol are minor corrections and/or clarifications that have no effect on the way the study is to be conducted. These administrative changes will be released by the Sponsor, agreed to by the investigators, and notified to the IRB(s).

##### 10.2.8. Protocol Deviations

The noncompliance may be on the part of the participant, the investigator, or the study site staff. As a result of protocol deviations, corrective actions are to be developed by the study site and implemented promptly.

It is the responsibility of the study site investigator to use continuous vigilance to identify and report protocol deviations to the Sponsor or its designee. All protocol deviations must be addressed in study source documents and reported to study monitor. Protocol deviations must be sent to the reviewing IRB per their policies. The study site investigator is responsible for knowing and adhering to the reviewing IRB requirements.

##### 10.2.9. Data Protection

Participants will be assigned a unique identifier by the Sponsor. Any participant records or datasets that are transferred to the Sponsor will contain the identifier only; participant names or any information that would make the participant identifiable will not be transferred.

The participant’s parent(s)/LAR(s) must be informed that the participant’s personal study-related data will be used by the Sponsor in accordance with local data protection law. The level of disclosure must also be explained to the participant’s parent(s)/LAR(s).

The participant’s parent(s)/LAR(s) must be informed that the participant’s medical records may be examined by clinical quality assurance (QA) auditors or other authorized personnel appointed by the Sponsor, by appropriate IRB members, and by inspectors from regulatory authorities.

Individual participant medical information obtained as a result of this study is considered confidential, and disclosure to third parties is prohibited. Information will be accessible to authorized parties or personnel only. Medical information may be given to the participant's physician or to other appropriate medical personnel responsible for the participant's well-being. Each participant's parent(s)/LAR(s) will be asked to complete a form allowing the investigator to notify the participant's primary health care provider of his/her participation in this study.

All laboratory specimens, evaluation forms, reports, and other records will be identified in a manner designed to maintain participant confidentiality. All records will be kept in a secure storage area with limited access. Clinical information will not be released without the written permission of the participant's parent(s)/LAR(s), except as necessary for monitoring and auditing by the Sponsor, its designee, the relevant regulatory authority, or the IRB.

The investigator and all employees and coworkers involved with this study may not disclose or use for any purpose other than performance of the study, any data, record, or other unpublished confidential information disclosed to those individuals for the purpose of the study. Prior written agreement from the Sponsor or its designee must be obtained for the disclosure of any confidential information to other parties.

##### 10.2.10. Sample Retention and Future Biomedical Research

The retention period of laboratory samples will be 20 years, or as permitted by local regulations, to address further scientific questions related to mRNA-1273 or anti-respiratory virus immune response. In addition, identifiable samples can be destroyed at any time at the request of the participant. During the study, or during the retention period, in addition to the analysis outlined in the study endpoints, exploratory analysis may be conducted using other Ab-based methodologies on any remaining blood or serum samples, including samples from participants who are screened but are not subsequently enrolled and samples collected and stored from Cohort D in Part 2. These analyses will extend the search for other potentially relevant biomarkers to investigate the effect of mRNA-1273, as well as to determine how changes in biomarkers may relate to exposure and clinical outcomes. A decision to perform such exploratory research may arise from new scientific findings related to the drug class or disease, as well as reagent and assay availability.

##### 10.2.11. Dissemination of Clinical Study Data

The Sponsor shares information about clinical trials and results on publicly accessible websites, based on international and local legal and regulatory requirements and other clinical trial disclosure commitments established by pharmaceutical industry associations. These websites include clinicaltrials.gov, EU clinical trial register (eu.ctr), and some national registries.

In addition, results from clinical trials are required to be submitted to peer-reviewed journals following internal company review for accuracy, fair balance, and intellectual property. For those journals that request sharing of the analyzable data sets that are reported in the publication, interested researchers are directed to submit their request to clinicalstudydatarequest.com.

Individual participant data and supporting clinical documents are available for request at clinicalstudydatarequest.com. While making information available, the privacy of participants in clinical studies sponsored by the Sponsor is ensured. Details on data sharing criteria and the process for requesting access can be found at this web address: clinicalstudydatarequest.com.

##### 10.2.12. Data Quality Assurance and Quality Control

Data collection is the responsibility of the clinical study staff at the study site under the supervision of the study site investigator. The investigator is responsible for ensuring the accuracy, completeness, legibility, and timeliness of the data reported.

- All participant data relating to the study will be recorded in the eCRF unless transmitted to the Sponsor or designee electronically (eg, laboratory data). The investigator is responsible for verifying that data entries are accurate and correct by physically or electronically signing the eCRF.
- The investigator must maintain accurate documentation (source data) that supports the information entered in the eCRF.
- The investigator must permit study-related monitoring, audits, IRB review, and regulatory agency inspections and provide direct access to source data documents.
- Monitoring details describing strategy (eg, risk-based initiatives in operations and quality such as Risk Management and Mitigation Strategies and Analytical Risk-Based Monitoring), methods, responsibilities and requirements, including handling of noncompliance issues and monitoring techniques (central, remote, or on-site monitoring) are provided in the Clinical Monitoring Plan.
- The Sponsor or designee is responsible for the data management of this study, including quality checks of the data.
- The Sponsor assumes accountability for actions delegated to other individuals (eg, CRO).

- Study monitors will perform ongoing source data verification to confirm that data entered into the CRF by authorized study site personnel are accurate, complete, and verifiable from source documents; that the safety and rights of participants are being protected; and that the study is being conducted in accordance with the currently approved protocol and any other study agreements, ICH GCP, and all applicable regulatory requirements.
- Records and documents, including signed ICFs, pertaining to the conduct of this study must be retained by the investigator for a period of at least 2 years after the last marketing application approval or, if not approved, 2 years following the discontinuance of the test article for investigation. If this requirement differs from any local regulations, the local regulations will take precedence unless the local retention policy is less than 2 years. No records may be destroyed during the retention period without the written approval of the Sponsor. No records may be transferred to another location or party without written notification to the Sponsor.

Quality assurance includes all the planned and systematic actions that are established to ensure that the clinical study is performed and the data are generated, documented (recorded), and reported according to ICH GCP and local/regional regulatory standards.

A QA representative from the Sponsor or a qualified designee, who is independent of and separated from routine monitoring, may periodically arrange inspections/audits of the clinical study by reviewing the data obtained and procedural aspects. These inspections may include on-site inspections/audits and source data checks. Direct access to source documents is required for the purpose of these periodic inspections/audits.

##### 10.2.13. Data Collection and Management

This study will be conducted in compliance with ICH CGP guidelines. This study will also be conducted in accordance with the most recent version of the Declaration of Helsinki.

This study will use electronic data collection to collect data directly from the study site using eCRFs. The investigator is responsible for ensuring that all sections of each eCRF are completed promptly and correctly and that entries can be verified against any source data.

Study monitors will perform source document verification to identify inconsistencies between the eCRFs and source documents. Discrepancies will be resolved in accordance with the principles of GCP. Detailed study monitoring procedures are provided in the Clinical Monitoring Plan.

Adverse events will be coded with MedDRA. Concomitant medications will be coded using WHO - Drug Dictionary.

##### 10.2.14. Source Documents

Source documents are original documents or certified copies, and include, but are not limited to, eDiaries, medical and hospital records, screening logs, ICFs, telephone contact logs, and worksheets. Source documents provide evidence for the existence of the participant and substantiate the integrity of the data collected. Source documents are filed at the investigator’s study site.

Data reported on the CRF or entered in the eCRF that are transcribed from source documents must be consistent with the source documents or the discrepancies must be explained. The investigator may need to request previous medical records or transfer records, depending on the study. Also, current medical records must be available.

The Sponsor or its designee requires that the investigator prepare and maintain adequate and accurate records for each participant treated with the IP. Source documents such as any hospital, clinic, or office charts, and the signed ICFs are to be included in the investigator’s files with the participant’s study records.

##### 10.2.15. Retention of Records

The principal investigator must maintain all documentation relating to the study for a period of at least 2 years after the last marketing application approval or, if not approved, 2 years following the discontinuance of the test article for investigation. If this requirement differs from any local regulations, the local regulations will take precedence unless the local retention policy is > 2 years.

If it becomes necessary for the Sponsor or the regulatory authority to review any documentation relating to the study, the investigator must permit access to such records. No records will be destroyed without the written consent of the Sponsor, if applicable. It is the responsibility of the Sponsor to inform the investigator when these documents no longer need to be retained.

##### 10.2.16. Study and Site Closure

If the study is prematurely terminated or suspended, the Sponsor shall promptly inform the investigators, the IRBs, the regulatory authorities, and any CRO(s) used in the study of the reason for termination or suspension, as specified by the applicable regulatory requirements. The investigator shall promptly inform the participants and should ensure appropriate participant therapy and/or follow-up.

The Sponsor or designee reserves the right to close the study site or terminate the study at any time for any reason at the sole discretion of the Sponsor.

The investigator may initiate study site closure at any time, provided there is reasonable cause and sufficient notice is given in advance of the intended termination.

Reasons for the early closure of a study site by the Sponsor or investigator may include but are not limited to the following:

- Continuation of the study represents a significant medical risk to participants
- Failure of the investigator to comply with the protocol, the requirements of the IRB or local health authorities, the Sponsor's procedures, or GCP guidelines
- Inadequate recruitment of participants by the investigator
- Discontinuation of further mRNA-1273 development

Study sites will be closed upon study completion. A study site is considered closed when all required documents and study supplies have been collected and a study site closure visit has been performed.

##### 10.2.17. Publication Policy

The results of this study may be published or presented at scientific meetings. If this is foreseen, the investigator agrees to submit all manuscripts or abstracts to the Sponsor before submission. This allows the Sponsor to protect proprietary information and to provide comments.

The Sponsor will comply with the requirements for publication of study results. In accordance with standard editorial and ethical practice, the Sponsor will generally support publication of multicenter studies only in their entirety and not as individual study site data. In this case, a coordinating investigator will be designated by mutual agreement.

Authorship will be determined by mutual agreement and in line with International Committee of Medical Journal Editors authorship requirements.

The clinical study plan and the results of the study will be published on www.ClinicalTrials.gov in accordance with 21 CFR 50.25(c). The results and data from this study belong to the Sponsor.

##### 10.2.18. Body Mass Index Charts for Boys and Girls

**For boys from birth to 5 years:**

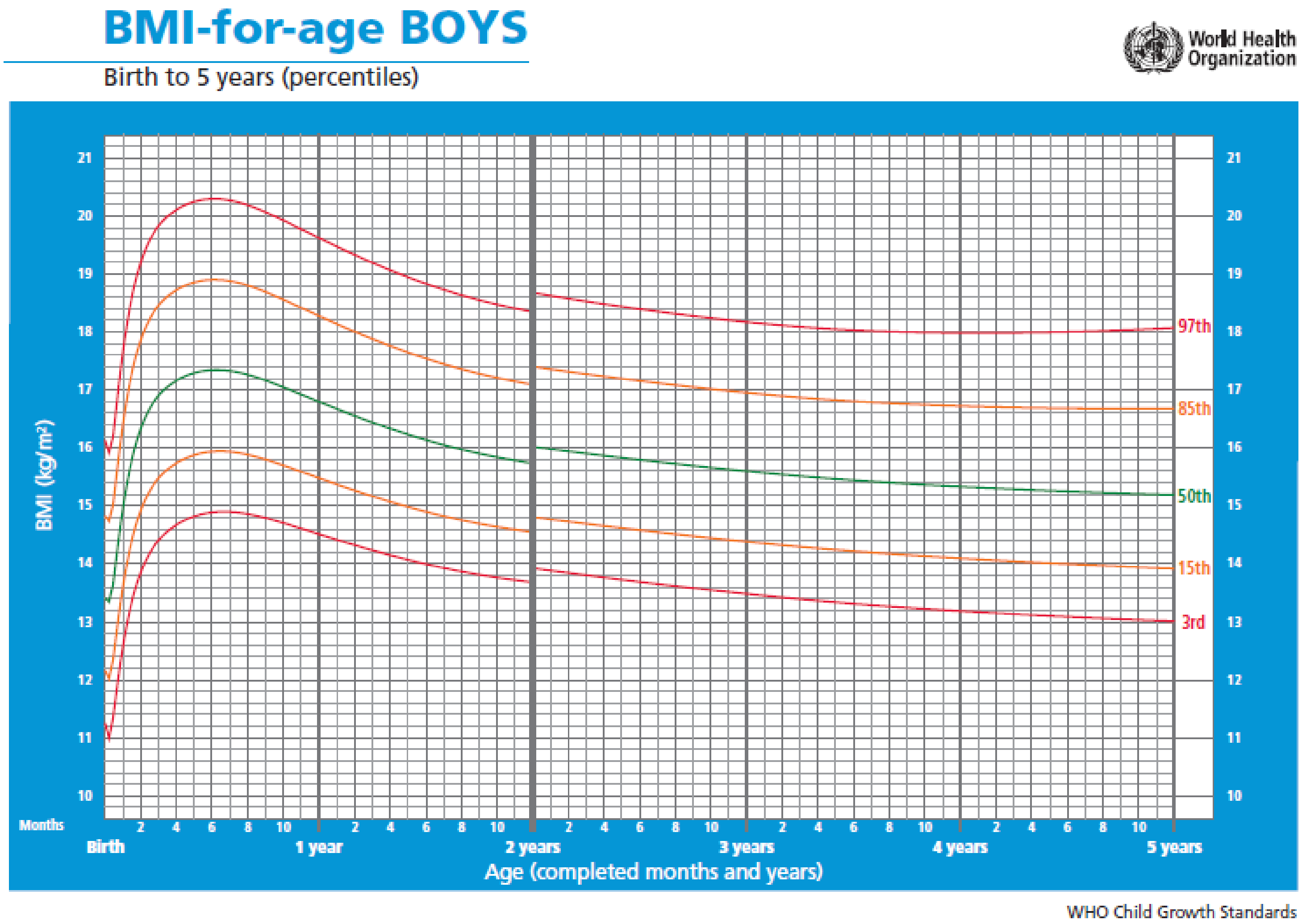

**For boys aged 5 through 19 years:**

#### BMI-for-age BOYS

5 to 19 years (percentiles)

World Health Organization

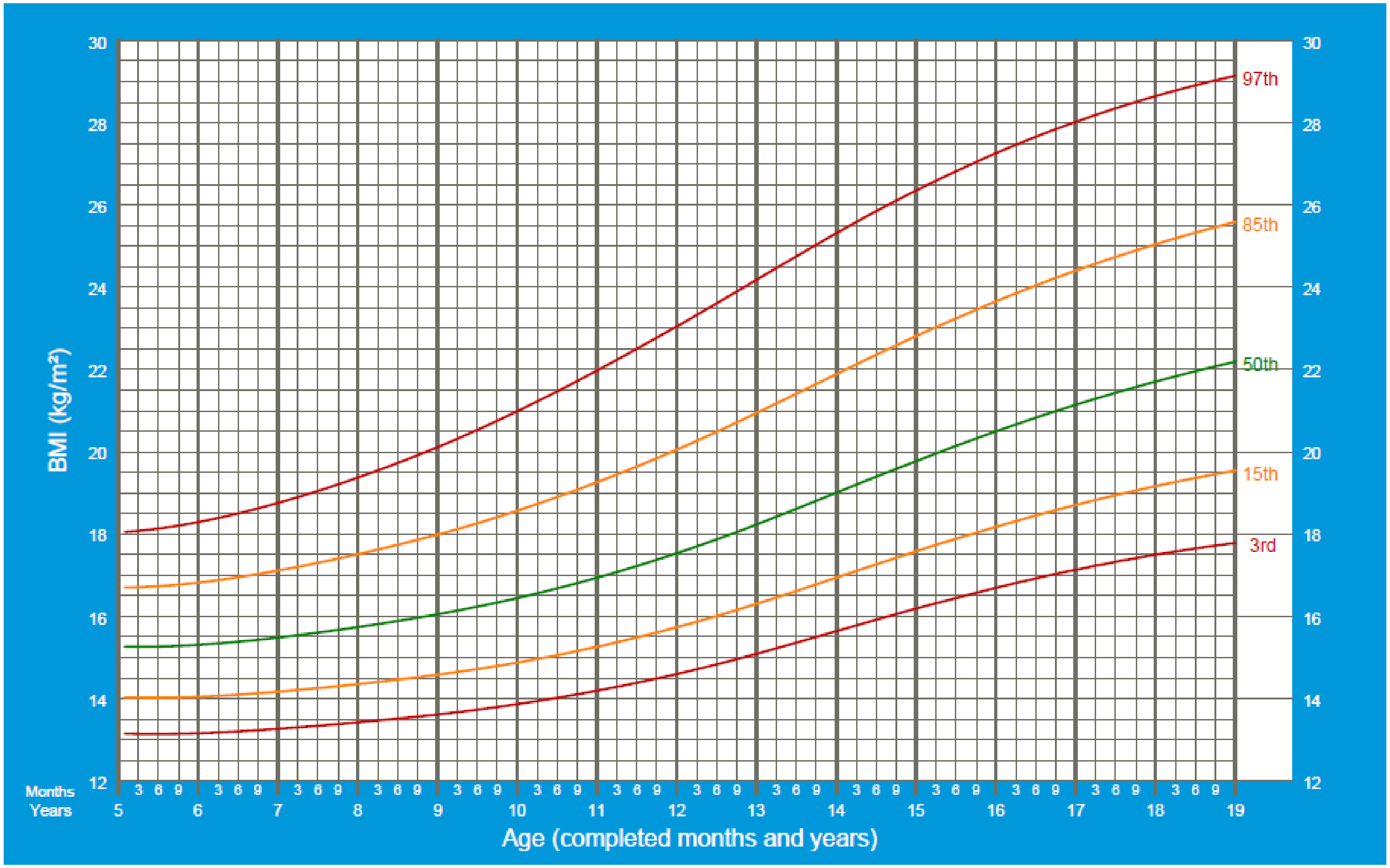

2007 WHO Reference

**For girls from birth to 5 years:**

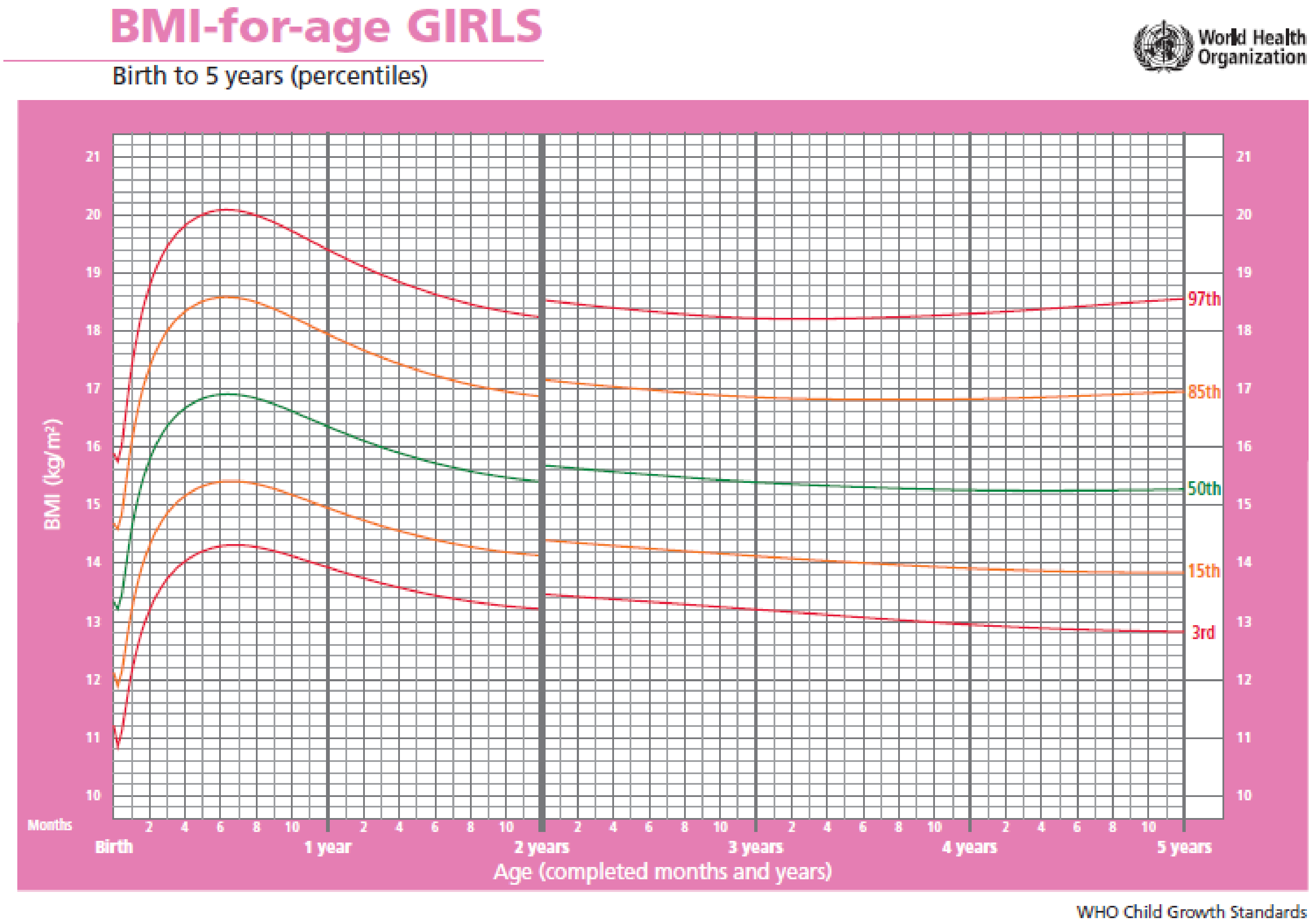

**For girls aged 5 through 19 years:**

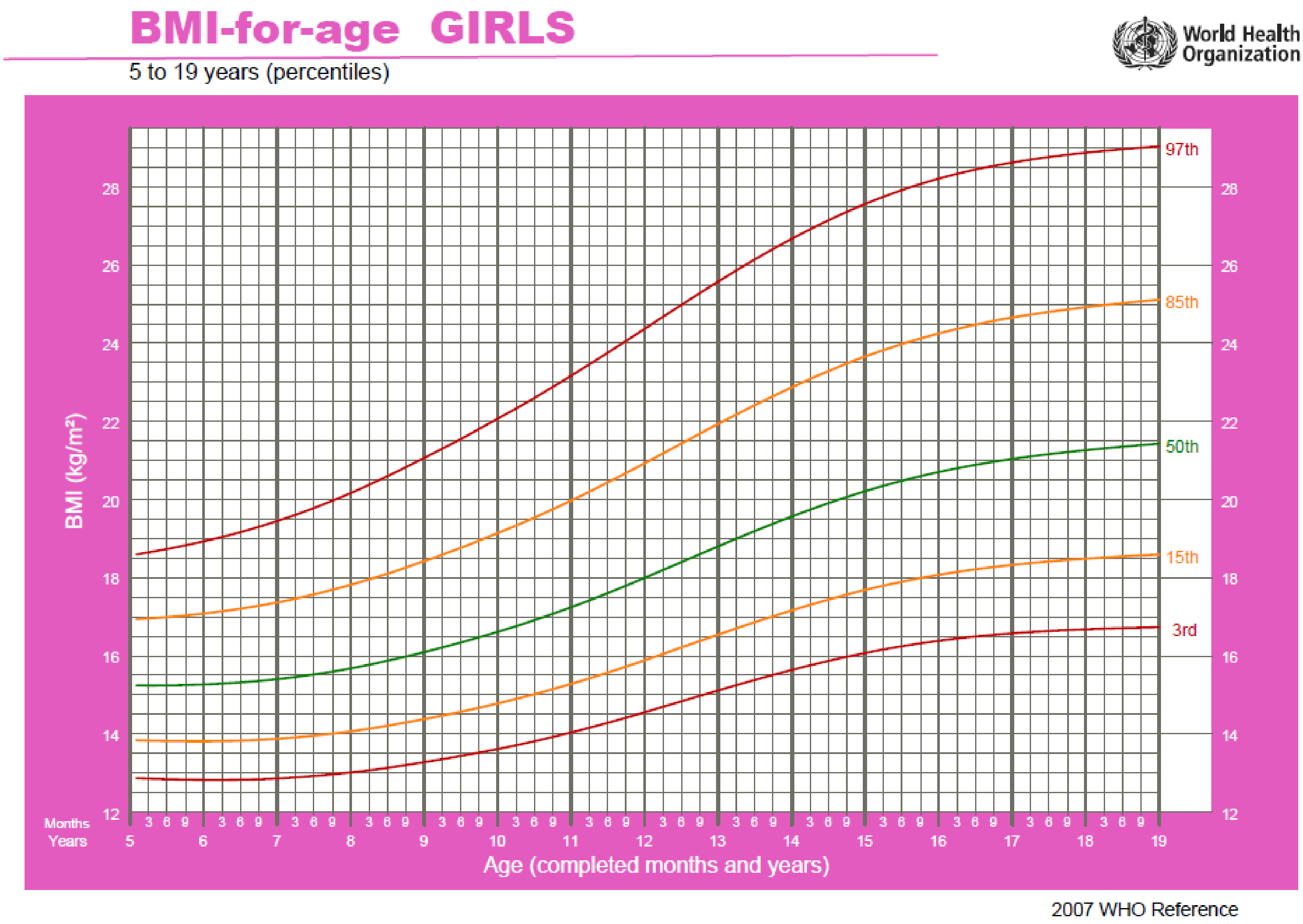

**For boys from 6 months to 2 years**

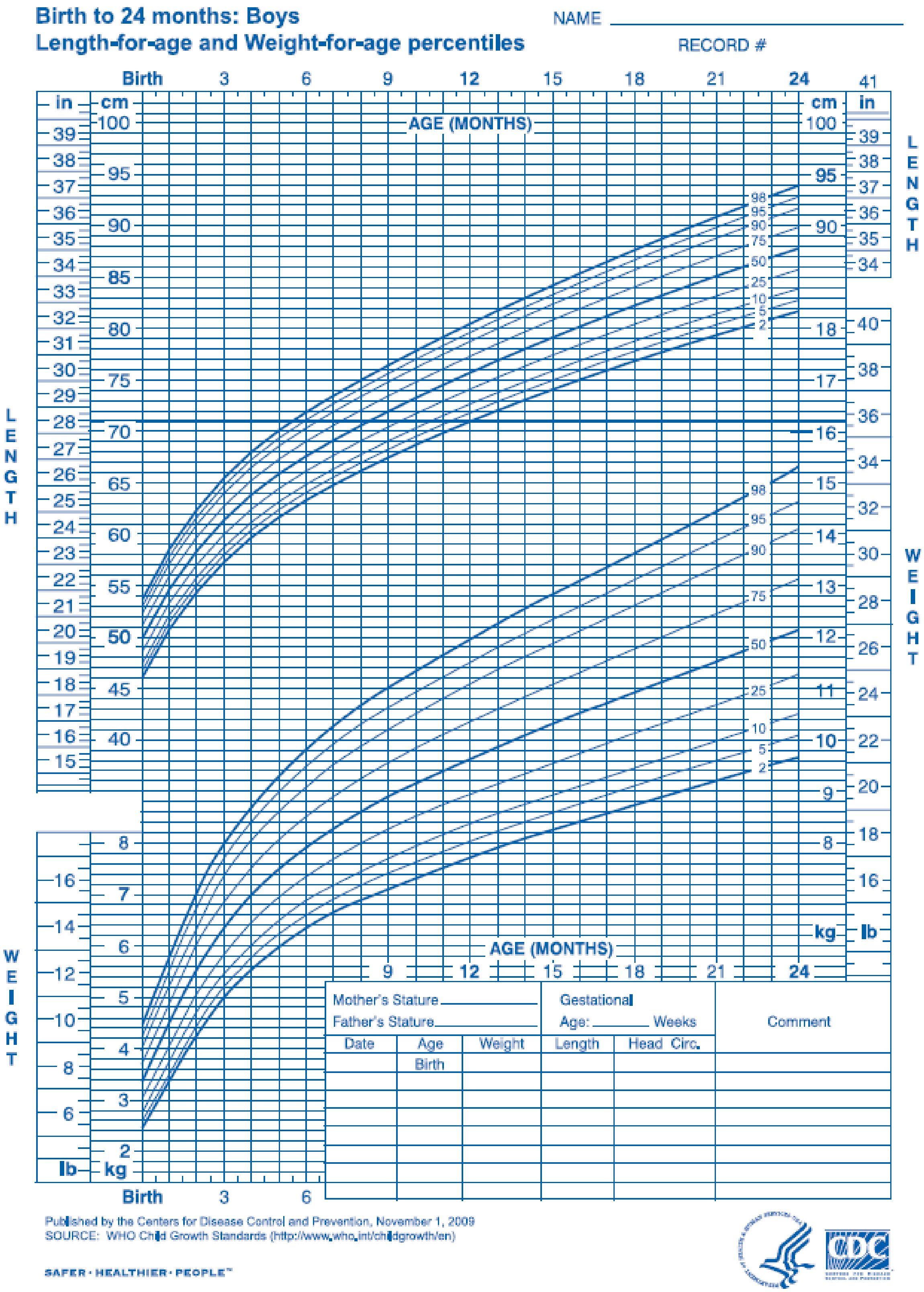

**For boys from 6 months to 2 years**

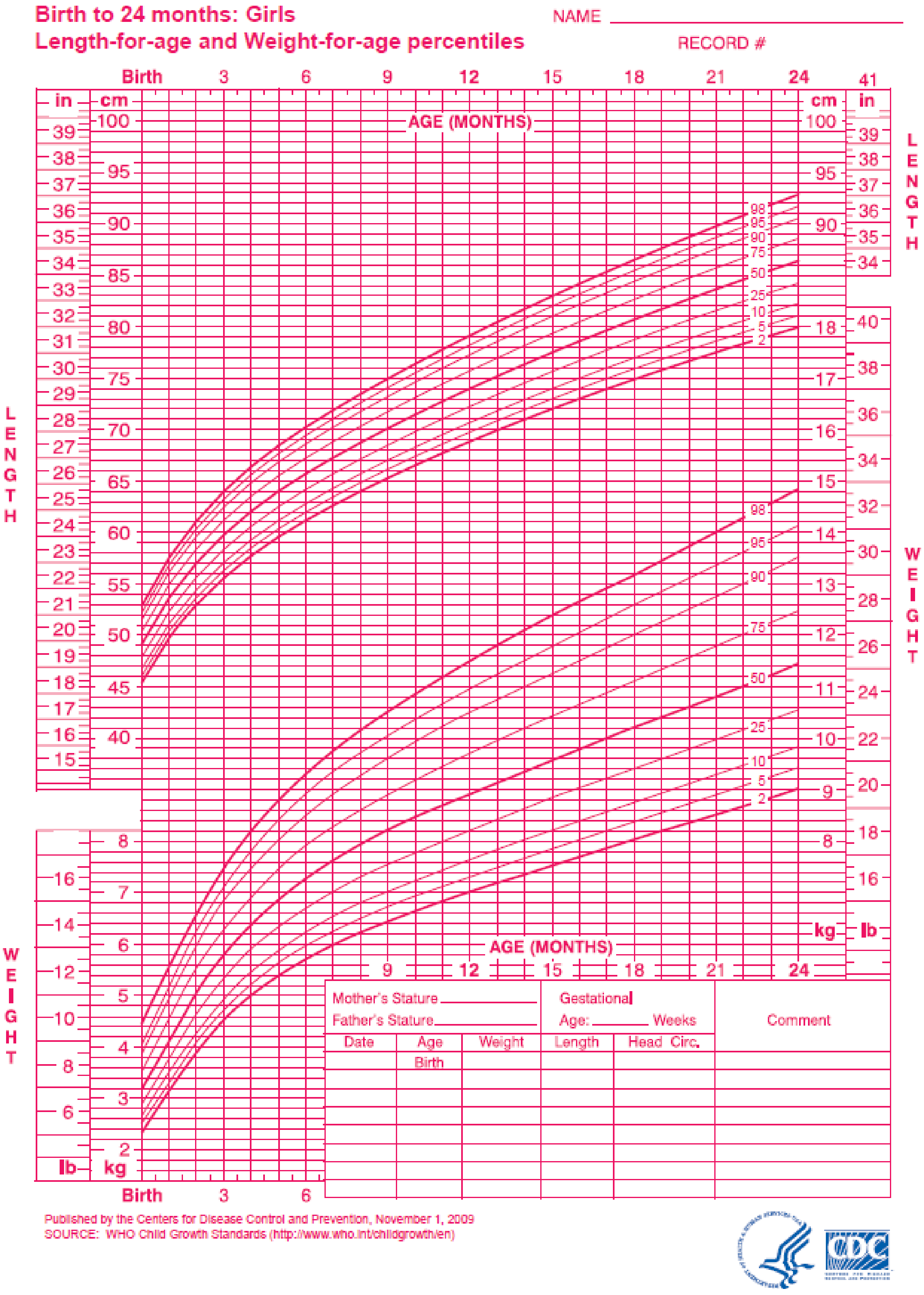

#### 10.3. APPENDIX 3: Contraceptive Guidance

##### Woman of Childbearing Potential (WOCBP)

Females of childbearing potential are those who are considered fertile following menarche and unless permanently sterile (see below). If fertility is unclear (eg, amenorrhea in adolescents or athletes) and a menstrual cycle cannot be confirmed before first dose of IP, additional evaluation should be considered.

Women in the following categories are not considered WOCBP:

1. Premenarchal
2. Surgically sterile female with one of the following:
   a. Documented complete hysterectomy
   b. Documented surgical sterilization

   For individuals with permanent infertility due to an alternate medical cause other than the above (eg, Mullerian agenesis, androgen insensitivity), investigator discretion should be applied in determining study entry.

   Note: Documentation can come from the study site personnel's review of the participant's medical records, medical examination, or medical history interview.

##### Contraception Guidance:

Adequate female contraception is defined as consistent and correct use of an FDA-approved contraceptive method in accordance with the product label, for example:

Barrier method (such as condoms, diaphragm, or cervical cap) used in conjunction with spermicide

Intrauterine device

Prescription hormonal contraceptive taken or administered via the oral (pill), transdermal (patch), subdermal, or IM route

Note: While **complete abstinence is accepted** as adequate female contraception in this age group, periodic abstinence (eg, calendar, ovulation, symptothermal, postovulation methods) and withdrawal are not acceptable methods of contraception.

#### 10.4. APPENDIX 4: Adverse Event of Special Interest Terms

Investigators should report all events which fall into the following categories as an AESI per the reporting processes specified in Section 7.4.5. The following AESIs are medical concepts that may be related to COVID-19 or are of interest in COVID-19 vaccine safety surveillance. Even if the events below occur in the setting of a COVID infection, the event should still be reported as an AESI if it is one of the medical concepts below.

| Medical Concept | Additional Notes |
|---|---|
| **Anosmia, Ageusia** | • New onset COVID associated or idiopathic events without other etiology excluding congenital etiologies or trauma |
| **Subacute thyroiditis** | • Including but not limited to events of: atrophic thyroiditis, autoimmune thyroiditis, immune-mediated thyroiditis, silent thyroiditis, thyrotoxicosis and thyroiditis |
| **Acute pancreatitis** | • Including but not limited to events of: autoimmune pancreatitis, immune-mediated pancreatitis, ischemic pancreatitis, edematous pancreatitis, pancreatitis, acute pancreatitis, hemorrhagic pancreatitis, necrotizing pancreatitis, viral pancreatitis, and subacute pancreatitis<br>• Excluding known etiologic causes of pancreatitis (alcohol, gallstones, trauma, recent invasive procedures) |
| **Appendicitis** | • Include any event of appendicitis |
| **Rhabdomyolysis** | • New onset rhabdomyolysis without known etiology such as excessive exercise or trauma |
| **Acute respiratory distress syndrome (ARDS)** | • Including but not limited to new events of ARDS and respiratory failure. |
| **Coagulation disorders** | • Including but not limited to thromboembolic and bleeding disorders, disseminated intravascular coagulation, pulmonary embolism, deep vein thrombosis |
| **Acute cardiovascular injury** | • Including but not limited to myocarditis, pericarditis, microangiopathy, coronary artery disease, arrhythmia, stress cardiomyopathy, heart failure, or acute myocardial infarction |
| **Acute kidney injury** | • Include events with idiopathic or autoimmune etiologies<br>• Exclude events with clear alternate etiology (trauma, infection, tumor, or iatrogenic causes such as medications or radiocontrast etc)<br>• Include all cases that meet the following criteria<br>○ Increase in serum creatinine by ≥ 0.3 mg/dl (≥26.5 µmol/l) within 48 hours;<br>OR |

| | |
|---|---|
| | ○ Increase in serum creatinine to ≥ 1.5 times baseline, known or presumed to have occurred within prior 7 days<br>OR<br>○ Urine volume ≤0.5 mL/ kg/ hour for 6 hours |
| **Acute liver injury** | • Include events with idiopathic or autoimmune etiologies<br>• Exclude events with clear alternate etiology (trauma, infection, tumor, etc)<br>• Include all cases that meet the following criteria<br>○ 3-fold elevation above the upper normal limit for ALT or AST<br>OR<br>○ > 2-fold elevation above the upper normal limit for total serum bilirubin or GGT or ALP |
| **Dermatologic findings** | • Chilblain-like lesions<br>• Single organ cutaneous vasculitis<br>• Erythema multiforme<br>• Bullous rashes<br>• Severe cutaneous adverse reactions including but not limited to: Stevens-Johnson Syndrome (SJS), Toxic Epidermal Necrolysis (TEN), Drug Reaction with Eosinophilia and Systemic Symptoms (DRESS) and fixed drug eruptions |
| **Multisystem inflammatory disorders** | • Multisystem inflammatory syndrome in adults (MIS-A)<br>• Multisystem inflammatory syndrome in children (MIS-C)<br>• Kawasaki's disease |
| **Thrombocytopenia** | • Platelet counts < 150 x10^9<br>• Including but not limited to immune thrombocytopenia, platelet production decreased, thrombocytopenia, thrombocytopenic purpura, thrombotic thrombocytopenic purpura, or HELLP syndrome |
| **Acute aseptic arthritis** | • New onset aseptic arthritis without clear alternate etiology (eg, gout, osteoarthritis, and trauma) |
| **New onset of or worsening of neurologic disease** | • Including but not limited to:<br>○ Guillain-Barre Syndrome<br>○ Acute disseminated encephalomyelitis (ADEM)<br>○ Peripheral facial nerve palsy (Bell's palsy)<br>○ Transverse myelitis<br>○ Encephalitis/Encephalomyelitis<br>○ Aseptic meningitis<br>○ Febrile seizures<br>○ Generalized seizures/convulsions<br>○ Stroke (Hemorrhagic and non-hemorrhagic) |

| | |
|---|---|
| | ○ Narcolepsy |
| **Anaphylaxis** | • Anaphylaxis as defined per protocol.<br>• Follow reporting procedures in protocol Section 7.4.5 |
| **Other syndromes** | • Fibromyalgia<br>• Postural Orthostatic Tachycardia Syndrome<br>• Chronic Fatigue Syndrome (Includes Myalgic encephalomyelitis and Post viral fatigue syndrome)<br>• Myasthenia gravis |

### 10.5. APPENDIX 5: Protocol Amendment History

#### 10.5.1. Amendment 4, 25 Aug 2021:

##### Main Rationale for the Amendment:

The main rationale for this amendment is to introduce an additional blood draw within 4 days after the second dose for participants in Cohort D in Part 2 in each age group. The samples will be stored for potential future analysis per a request from the FDA. For participants already enrolled in Cohort D prior to protocol amendment 4 implementation at the site, this blood draw will be optional and confirmed at time of re-consenting; for all participants newly enrolled into Cohort D under amendment 4, it will be mandatory.

##### Summary of Major Changes in Protocol Amendment 4:

| Section # and Name | Description of Change | Brief Rationale |
|---|---|---|
| Title Page, Signature page, Synopsis, and Header | Updated the protocol version and date. | Updated to reflect the new version and date. |
| Global | Minor grammar and formatting corrections were made throughout the document. | Updates were made for clarity and readability. |
| Synopsis, Section 3.1.1.2 (Treatment and Follow-up period), and Section 10.1 (Appendix 1 Schedule of Assessments - Table 10) | Language was added to clarify that the 300 participants in the Arm 1 and 2 expansions will have a scheduled Day 1 blood draw and a voluntary blood draw on Day 57. | To clarify that only expansion portion of Arms 1 and 2 have a voluntary Day 57 blood draw, but that blood draws are mandatory for all other Arms in part 1 (as per Clarification Memo # 7) |
| Synopsis, Section 3.1 (General Design - Figure 1b), Section 3.1.1.2 (Treatment and Follow-up period), Section 7.2 (Blood Collections for Immunogenicity Assessments and Biomarker Samples), and Section 10.1 | Day 30 (+3 days) blood draw was added to indicate that participants in Cohort D (remainder of the age group) will provide a blood sample at Day 1 and at Day 30 (+3 days) for storage and potential future biomarker testing. | Day 30 (+3 days) blood draw was added per the request from the FDA. |

| | | |
|---|---|---|
| (Appendix 1 Schedule of Assessments - Table 10) | | |
| Synopsis, Section 7.2 (Blood Collections for Immunogenicity Assessments and Biomarker Samples), Section 10.1 (Appendix 1 Schedule of Assessments) - Table 10 (Schedule of Assessments), and Table 12 (Phlebotomy Schedule for Serology, Biomarker Samples, and Cell-Mediated Immunity for Part 2 [Expansion] of the Study) | Header in Section 7.2 was updated from "Immunogenicity Assessments" to "Blood Collections for Immunogenicity Assessments and Biomarker Samples"<br><br>A note was added to Section 7.2 indicating that if less than 8 mL of blood is drawn at baseline then the child cannot be enrolled.<br><br>Table 12 title in Section 10.1 was updated to include "Biomarker Sample."<br><br>Language and/or column was added to each section to indicate that there will be a Day 30 (+3 days) blood draw for storage and potential future biomarker testing for Cohort D in Part 2, and that the volume will be ~4 mL for all age groups. | Headings and titles were updated for clarity and consistency with content under heading.<br><br>Blood volume was indicated as this is a pediatric study.<br><br>Note language was added for clarity on enrollment procedure. |
| Synopsis and Section 7.4.5 (Adverse Events of Special interest) | The Cardiac Endpoint Adjudication Committee's name was updated to the officially recognized name as the "Cardiac Event Adjudication Committee" | This change was to align the committee language across all relevant Moderna studies. |
| Section 7.1.5 (Assessment for SARS-CoV-2 Infection) | Language was added to specify that exposure to an individual in the household confirmed to be infected with SARS-CoV-2 will require a study illness visit. | This clarification was made to capture highest risk exposure only. |

| | | |
|---|---|---|
| Section 7.3.2 (Surveillance for COVID-19 Symptoms) | Language was added to specify when an illness visit would be required. | Clarification of circumstances that require an illness visit. |
| Section 10.2.10 (Sample Retention and Future Biomedical Research) | Language was added to indicate that the samples collected and stored from Cohort D in Part 2 may undergo additional exploratory analysis using Ab-based methodologies. | To reflect the addition of a Day 30 blood sample that will be stored but only analyzed if an appropriate biomarker is identified. |

##### 10.5.2. Amendment 3, 23 Jul 2021

**Main Rationale for the Amendment:**

The main rationale for this amendment is to add a case definition for myocarditis and pericarditis as well as guidance for reporting and assessing suspected cases for this study, given the recent emergence of a temporal association between mRNA vaccine administration and signs and symptoms of myocarditis/pericarditis.

In addition, the sample size for each age group in Part 2 (blinded part) is increased to allow for a 95% probability to detect a rare adverse event occurring at a rate of 1 in 1,000.

**Summary of Major Changes in Protocol Amendment 3:**

| Section # and Name | Description of Change | Brief Rationale |
|---|---|---|
| Title Page, Signature page, Synopsis, and Header | Updated the protocol version and date. | Updated to reflect the new version and date. |
| Global | Minor grammar and formatting corrections were made throughout the document. | Updates were made for clarity and readability. |
| Synopsis, Section 3.1 (General Design), Section 5.2 (Randomization), and Section 8.3 (Power and Sample Size) | The overall sample size for Part 2 was updated to approximately 12,000 participants.<br><br>The overall sample size for each age group was updated to up to 4,000 participants. | Samples sizes were increased to allow for a 95% probability to detect a rare adverse event occurring at a rate of 1 in 1,000. |

| | | |
|---|---|---|
| | The sample size for Study Arms 8, 10, and 12 were each updated to up to 3,000 participants.<br><br>The sample size for Study Arms 9, 11, and 13 were each updated to up to 1,000 participants. | |
| Synopsis, Section 2 (Objectives and Endpoints),<br>Section 3.1 (General Design) - Study Progression,<br>Section 6.2 (Discontinuing Study Vaccination),<br>Section 7.1 (Safety Assessments and Procedures), and<br>Section 10.1 (Appendix 1: Schedule of Assessments - Table 10 and Table 11) | AESIs of Myocarditis and/or pericarditis were added (in addition to MIS-C). | To reflect addition of case definition for myocarditis and pericarditis in AESI section. |
| Synopsis, Section 7.4.5 (Adverse Events of Special Interest), and Section 7.5.2 (Data Safety Monitoring Board) | Included language for the addition of an external clinical endpoint adjudication committee, that will adjudicate any suspected cases of myocarditis, pericarditis, or myopericarditis and make recommendations in consultation with the DSMB to the Sponsor. | To reflect the addition of an external clinical endpoint adjudication committee for cases of myocarditis/pericarditis to the safety oversight of the study. |
| Section 1.3.2 (Risks to Study Participation and Their Mitigation) | Added paragraph on very rare reports of myocarditis and pericarditis occurring after vaccination with Moderna COVID-19 Vaccine under Emergency Use Authorization in adults aged 18 years and older. | To reflect addendum made to Investigator’s Brochure. |

| | | |
|---|---|---|
| Section 7.4.4 (Medically Attended Adverse Events) | Language was updated to Unsolicited AEs will be captured on the AE page of the eCRF. | To reflect the correct method of collecting unsolicited AEs. |
| Section 7.4.5 (Adverse Events of Special Interest) | Added CDC case definitions for myocarditis and pericarditis. | To provide guidance to the investigators regarding assessing and reporting myocarditis and pericarditis for this study population. |
| Section 10.4 (Appendix 4: Adverse Event of Special Interest Terms) | Appendix 4 was added to the protocol. | To include AESI list in the protocol instead of a separate document. |

##### 10.5.3. Amendment 2, 17 Jun 2021

**Main Rationale for the Amendment:**

The main rationale for this amendment is to add an optional blood collection on Day 57 for participants in the expansion part of Arm 1 and Arm 2 to gather additional data on immunogenicity. The handling of potential unblinding requests in Part 2 is further clarified. In addition, the decision in Part 1 to not evaluate the 100-µg dose in participants less than 2 years old is integrated into this amendment by removing Arm 7 (6 months to < 2 years, 100 µg dose) from this age group. The change in dose level is based on moderate, increased reactogenicity observed in Arm 2 (6 to < 12 years of age, 100 µg), which led to the decision to not evaluate the 100-µg dose in the 2 to < 6 years age group. In order to maintain dose-ranging in the 2 to < 6 years age group, Arm 7 may instead enroll participants in this age group to evaluate the 25-µg dose if the 100-µg dose is eliminated at any point during the dose escalation process.

The Summary of Major Changes table describes the major changes made in Amendment 2, including the sections modified and the corresponding rationales. The synopsis of Amendment 2 has been modified to correspond to changes in the body of the protocol.

**Summary of Major Changes in Protocol Amendment 2:**

| **Section # and Name** | **Description of Change** | **Brief Rationale** |
|---|---|---|
| Protocol Synopsis and Section 3.1 General Design | Clarified that preliminary safety and immunogenicity data of Arm 1 (6 to < 12 years of age, 50 µg) and Arm 2 (6 to < 12 years of age, 100 µg), as applicable, will aid in the selection of a dose level for Part 2. | Tolerability and safety data will be available from both doses (50 and 100 µg) and will be an essential metric for dose selection. Additionally, immunogenicity data from the 50-µg dose (a dose lower than the 100-µg dose assessed in the P301 adult efficacy trial) will allow for assessment of the likelihood of the 50-µg dose to meet noninferiority criteria. These data will allow an informed dose selection decision for Part 2. |
| Protocol Synopsis and Section 3.1 General Design | Revised the age group and dose level of Arm 7 (6 months to < 2 years of age, 100 µg) to 2 to < 6 years and 25 µg of mRNA-1273. Arm 7 was made optional, the 25-µg dose will be evaluated if 100-µg dose is eliminated at any point during the dose escalation process, to maintain dose ranging. | The 100-µg dose was not evaluated in participants aged < 2 years based on the internal safety team recommendation. The dose level and age group of Arm 7 was revised to maintain dose-ranging in the 2 to < 6 years age group by adding a lower dose than originally planned (25 µg). |

| | | |
|---|---|---|
| Section 3.3 Justification for Dose, Control Product, and Choice of Study Population | Clarified that if a COVID-19 vaccine (mRNA-1273 or other) is authorized or licensed, eligible study participants will be offered the opportunity to unblind, and a nasal swab and a blood sample will be collected. Participants who received mRNA-1273 will continue in the study. If a participant previously received placebo and mRNA-1273 is authorized for use in the participant's age group, the participant will be offered unblinding followed by a cross-over vaccination with mRNA-1273. If mRNA-1273 is not yet authorized for the relevant age group, but an alternative vaccine is authorized, previous placebo recipients may seek the alternative vaccine and withdraw from study. | To clarify plans for unblinding requests. |
| Section 7.4.3 Solicited Adverse Reactions Table 7 | Revised the temperature ranges for fever in Table 7: Solicited Adverse Reactions and Grades: Age 6 to ≤ 36 Months. | To correct the temperature cut-offs for consistency with internal Moderna standard for the 6 to ≤ 36 months age group. |
| Protocol Synopsis and Section 7.5.2 Data Safety Monitoring Board | Clarified that the DSMB will review safety data from a subset of approximately 300 participants at the selected dose level in the 6 to < 12 years age group rather than the full safety set for Arm 1 and Arm 2 from Part 1 (N = 750 participants) before allowing the start of enrollment in Part 2. | Dose selection will be based on safety data from approximately 300 participants. Approximately 300 participants will have reached Day 43. This will expedite expansion of the study. The safety review will still allow for detection of an AE occurring at a rate ~ 1% (N = ~ 375 participants per group remains the same). |

| | | |
|---|---|---|
| Protocol Synopsis and Section 7.5.2 Data Safety Monitoring Board | For both the middle age group (2 to < 6 years) and the youngest age group (6 months to < 2 years) the DSMB will review cumulative safety data after approximately 400 participants have been exposed to mRNA-1273 at selected dose in each age group, combining participants from Part 1 and Part 2, before further expansion in each respective age group. | To reflect changes made to dose escalation for < 6 year olds in Part 1 (elimination of 100 µg group), and to moderately increase the subset of participants exposed at chosen dose level before final expansion in Part 2 (N= ~375 participants per group was updated to N= ~400 participants per group) |
| Protocol Synopsis and Section 8.6.1 Interim Analyses | Clarified that the interim analyses in Part 1 will be optional and will be performed after all or a subset of participants have completed Day 57. Clarified that the interim analysis in Part 2 will be performed after all or a subset of participants have completed Day 57. | To reflect changes made to dose selection plans that will expedite the expansion of the study. |
| Section 10.1 Appendix 1: Schedule of Assessments | Added an optional blood collection for immunogenicity on Day 57 for the expansion part of Arm 1 and Arm 2. | An optional blood collection was added for the expansion part of Arm 1 and Arm 2 to gather additional immunogenicity data in Part 1. |

##### 10.5.4. Amendment 1, 30 Apr 2021

**Main Rationale for the Amendment:**

The main purpose of this amendment is to allow a Data Safety Monitoring Board (DSMB) safety review when approximately 375 children have received mRNA-1273 in each age group, before expanding enrollment to each full age cohort. This review will allow assessment of less frequent adverse events (AEs), occurring at a rate of approximately 1 in 100. Per this amendment, this will be achieved in different ways for each of the 3 age groups:

1. For the 6 to < 12 years of age group: To expedite this formal safety review in school-aged children, an additional 300 participants will be enrolled to each dose level in Part 1 (ie, to have n = 375 per dose group), and the DSMB will review Day 57 safety data for all 375 participants in each dose group before advancing this age group to Part 2.
2. For the 2 to < 6 years and 6 months to < 2 years of age groups: DSMB safety review will occur within Part 2, at a time when it is anticipated that a total of approximately 375 participants (accounting for the 3:1 randomization to vaccine or placebo in the blinded Part 2) have been exposed to mRNA-1273 at the dose level selected for Part 2. DSMB will occur when Day 57 safety data from this subset is available for review.

The Summary of Changes table provided here describes the major changes made in Amendment 1 relative to the original protocol, including the sections modified and the corresponding rationales. The synopsis of Amendment 1 has been modified to correspond to changes in the body of the protocol.

**Summary of Major Changes in Protocol Amendment 1:**

| Section # and Name | Description of Change | Brief Rationale |
|---|---|---|
| Title Page, Protocol Approval Page, and Protocol Amendment Summary of Changes | Updated protocol version and date. Added Protocol Amendment Summary of Changes.<br><br>Medical monitor/sponsor contact information was updated. | Updated to reflect new version, date of protocol, and new medical monitor/sponsor contact. Protocol Summary of Changes added to be in line with Moderna guidelines. |
| Global | Minor grammar and formatting corrections were made throughout the document. | Updates were made for clarity and readability. |
| Global | Primary endpoint language was updated to use 'antibody' rather than 'nAb.' | To reflect potential use of binding antibody at time of analysis, if available. |
| Protocol Synopsis and Section 2 (Objectives and Endpoints) | The Secondary Objective "to evaluate the incidence of SARS-CoV-2 infection after vaccination with mRNA-1273 or placebo" Endpoint language was updated. | To maintain consistency across all Moderna protocols. |
| Protocol Synopsis and Section 2 (Objectives and Endpoints) | The secondary objective "To evaluate the incidence of asymptomatic SARS-CoV-2 infection after vaccination with mRNA-1273 or placebo" Endpoint language was updated. | To maintain consistency with other mRNA-1273 related protocols. Additionally, this language was updated to clarify that only participants without evidence of prior infection at baseline will be included in the analysis of the secondary objective of asymptomatic infection with SARS-CoV-2. |

| | | |
|---|---|---|
| Protocol Synopsis and Section 2 (Objectives and Endpoints) | The following exploratory objective was added, "To explore asymptomatic SARS-CoV-2 infection after vaccination with mRNA-1273 or placebo in participants with serologic evidence of infection at baseline."<br>The corresponding exploratory endpoint was added, "GM and GMFR of bAb levels against SARS-CoV-2 nucleocapsid protein (quantitative IgG)"<br><br>New abbreviations were added to the footnote. | To explore serological evidence of asymptomatic infections in seropositive participants. |
| Synopsis and Section 2 (Objectives and Endpoints) | The definition of COVID-19 infection was updated to match the CDC guidelines and the details of this guideline was moved to the footer. | To reflect the generally milder and more varied presentation of SARS-CoV-2 infection in the pediatric population, matching the secondary case definition in adult study (P301/COVE study). |
| Section 3.1 (General Design) Figure 1 (Study Schema) | Study Schema was updated to correct typographical error. | Figure updated to ensure consistency throughout protocol. |
| Protocol Synopsis and Section 4.1 (Inclusion Criteria) | Language was added to clarify that the height and weight of children < 2 years of age must both meet or exceed the 3rd percentile according to WHO Child Growth Standard at the Screening Visit. | Height and weight are better measures of growth than BMI for children < 2 years of age. |
| Protocol Synopsis and Section 4.1 (Inclusion Criteria) | The heading before criterion 7, "Special inclusion criteria for children 6 months to < 2 years of age" was updated to "Special inclusion criteria for children 6 months to < 12 months of age" | Exclusion of prematurity is considered relevant only for children under 12 months of age in the context of mRNA vaccines (not live vaccines). |

| | | |
|---|---|---|
| Protocol Synopsis and Section 4.2 (Exclusion Criteria) | A note was added to the exclusion of febrile seizures to indicate that a history of a simple, single febrile seizure is allowed for children 6 years and older in Part 2 of the study. | The risk of febrile seizures is most relevant for children up to 6 years of age. Once the safety data has been reviewed in Part 1, children 6 years of age and older can be included even with a history of a single, simple febrile seizures. |
| Protocol Synopsis, Section 3.1 (General Design), Section 5.2 (Randomization), and Section 8.3 (Power and Sample Size) | The sample size was increased from 750 to 1,350 participants in Part 1.<br><br>Sample size in Part 2 was adjusted in the 6 to < 12 years of age group from 2,000 to 1,700 (mRNA-1273 arm ~1,275 participants; placebo arm ~425 participants). | To reflect changes per main rationale for Amendment. |
| Protocol Synopsis, Section 3.1 (General Design), Section 8.5.3 (Immunogenicity Analyses), and Section 8.6.1 (Interim Analyses) | Language was updated to reflect when the primary immunogenicity analysis will be done for Part 2. | To reflect adjustment in the safety oversight plan. |
| Synopsis and Section 7.5.2 (Data Safety Monitoring Board) | Language was added to update the plan for the review process. | To update and clarify the changes made to the process based on main reason for amendment. |
| Protocol Synopsis, Section 8.2 (Statistical Hypothesis), and Section 8.3 (Power and Sample Size) | Coprimary endpoint 1 and 2 language updated. | To reflect updates to the statistical plan. |
| Protocol Synopsis and Section 8.5.3 (Immunogenicity Analyses) | Language for multiplicity adjustment between age groups was added. | To reflect updates for the statistical plan. |

| | | |
|---|---|---|
| Section 3.3 (Justification for Dose, Control Product, and Choice of Study Population) | Language was updated to indicate that if immunogenicity criteria are successfully met in P204 age cohorts, blinded placebo participants will be given the opportunity to receive the mRNA-1273 vaccine, and participants who received a lower dose than the dose receiving Emergency Use Authorization will be provided a booster with the 'optimal dose' for a given age group. | To describe the planned approach to dosing placebo recipients and lower dose recipients in case of Emergency Use Authorization of mRNA-1273 for each age group during the conduct of the trial. |
| Section 7.1.5 (Assessment for SARSCoV2 infection) | Language was updated to include further clarification on procedures to follow when participant is exposed to an individual with confirmed SARS-CoV-2 infection. A definition of last exposure was also added.<br><br>Language was added to clarify that an initial assessment will be performed at a study illness visit to determine general appearance, and to provide details as to whom may perform the assessment. | To provide a clear definition of last exposure, and the processes and timing to follow when a participant is exposed to a confirmed SARS-CoV-2 infected person.<br><br>Clarification of which providers can perform initial assessments for illness visits. |
| Section 7.3.1 (Vaccine Effectiveness Assessments) Table 5 (Age-Specific Cut-Offs for Vital Signs and Laboratory Variables | Removed irrelevant ages (rows). Systolic Blood pressure parameter updated to include hypotension parameters for pediatric population. | To ensure age-appropriate information is included in definition of Severe COVID-19. |
| Synopsis, Section 2 (Objectives and Endpoints), and Section 7.3.1 (Vaccine Effectiveness Assessments) | The definition of a SARS-CoV-2 infection was updated. | To reflect pediatric manifestations of SARS-CoV-2 infection and match CDC case definition. |

| | | |
|---|---|---|
| Section 7.3.1 (Vaccine Effectiveness Assessments) Table 6 (Definition of renal-, liver-, and neurological dysfunction for Pediatric Population (< 12 years of age) | Table 6 was added to define renal, liver, and neurological dysfunction for this study. | To define severe COVID-19 with age-appropriate definitions and objective measures. |
| Section 7.3.2 (Surveillance for COVID-19 Symptoms:) | Language was added to provide guidance for omitting or conducting an illness visit for febrile children that have an alternative diagnosis identified. | To match the American Academy of Pediatrics guidance on COVID-19 testing in children. |
| Section 7.3.3 (Follow-up/Convalescent Period After Diagnosis with COVID-19) | Language was added to provide guidance for children hospitalized with possible or confirmed MIS-C. | Clarification of sample collection for suspected MIS-C. |
| Section 10.1 (APPENDIX 1: Schedule of Assessments); Table 10: Schedule of Assessments | A footnote was added to clarify the blood sample for vaccine immunogenicity in Part 1 and Part 2. | Clarification of subset of participants in each Arm that will have samples collected for immunogenicity postbaseline. |
| Section 10.2.18 (Body Mass Index Charts for Boys and Girls) | CDC charts based on WHO data for use in the US in children < 2 years old added for reference. | Height and weight charts were added for use in children < 2 years of age. |

Signature Page for VV-CLIN-003244 v1.0

| | |
|---|---|
| Approval | Rituparna Das<br>Clinical<br>29-Sep-2021 23:51:06 GMT+0000 |

| | |
|---|---|
| Approval | Deborah Manzo<br>Clinical<br>30-Sep-2021 13:22:08 GMT+0000 |

Signature Page for VV-CLIN-003244 v1.0
